# Comparative transmissibility of SARS-CoV-2 variants Delta and Alpha in New England, USA

**DOI:** 10.1101/2021.10.06.21264641

**Authors:** Rebecca Earnest, Rockib Uddin, Nicholas Matluk, Nicholas Renzette, Katherine J. Siddle, Christine Loreth, Gordon Adams, Christopher H. Tomkins-Tinch, Mary E. Petrone, Jessica E. Rothman, Mallery I. Breban, Robert Tobias Koch, Kendall Billig, Joseph R. Fauver, Chantal B.F. Vogels, Sarah Turbett, Kaya Bilguvar, Bony De Kumar, Marie L. Landry, David R. Peaper, Kevin Kelly, Greg Omerza, Heather Grieser, Sim Meak, John Martha, Hannah H. Dewey, Susan Kales, Daniel Berenzy, Kristin Carpenter-Azevedo, Ewa King, Richard C. Huard, Sandra C. Smole, Catherine M. Brown, Timelia Fink, Andrew S. Lang, Glen R. Gallagher, Pardis C. Sabeti, Stacey Gabriel, Bronwyn L. MacInnis, New England Variant Investigation Team, Ryan Tewhey, Mark D. Adams, Daniel J. Park, Jacob E. Lemieux, Nathan D. Grubaugh

## Abstract

The severe acute respiratory syndrome coronavirus 2 (SARS-CoV-2) Delta variant quickly rose to dominance in mid-2021, displacing other variants, including Alpha. Studies using data from the United Kingdom and India estimated that Delta was 40-80% more transmissible than Alpha, allowing Delta to become the globally dominant variant. However, it was unclear if the ostensible difference in relative transmissibility was due mostly to innate properties of Delta’s infectiousness or differences in the study populations. To investigate, we formed a partnership with SARS-CoV-2 genomic surveillance programs from all six New England US states. By comparing logistic growth rates, we found that Delta emerged 37-163% faster than Alpha in early 2021 (37% Massachusetts, 75% New Hampshire, 95% Maine, 98% Rhode Island, 151% Connecticut, and 163% Vermont). We next computed variant-specific effective reproductive numbers and estimated that Delta was 58-120% more transmissible than Alpha across New England (58% New Hampshire, 68% Massachusetts, 76% Connecticut, 85% Rhode Island, 98% Maine, and 120% Vermont). Finally, using RT-PCR data, we estimated that Delta infections generate on average ∼6 times more viral RNA copies per mL than Alpha infections. Overall, our evidence indicates that Delta’s enhanced transmissibility could be attributed to its innate ability to increase infectiousness, but its epidemiological dynamics may vary depending on the underlying immunity and behavior of distinct populations.

## Introduction

The evolution and emergence of severe acute respiratory syndrome coronavirus 2 (SARS-CoV-2) variants associated with increased transmissibility, more severe disease, and/or decreased vaccine effectiveness continues to exacerbate the coronavirus disease 2019 (COVID-19) pandemic (CDC, 2021a). In particular, two SARS-CoV-2 variants with enhanced transmissibility substantially altered the pandemic’s trajectory: Alpha (B.1.1.7 lineage) and Delta (B.1.617.2 and AY.x sub-lineages). Alpha, defined in part by a N501Y amino acid substitution in the spike gene, was first detected in the United Kingdom in late 2020 and became the dominant global variant by early 2021 (Alpert et al., 2021; GISAID, 2021; Rambaut et al., 2020a). Delta, which contains spike L452R and P681H, was first detected in India in early 2021 and displaced Alpha as the dominant variant by mid-2021. This shift led to a significant resurgence in COVID-19 cases in many countries (Bolze et al., 2021; CDC, 2021b; Challen et al., 2021; GISAID, 2021; WHO, 2021).

Viral transmissibility can be affected by two main factors: innate attributes of the variant itself and the specific population in which it spreads. Variants have defining sets of mutations that may lead to innately increased transmissibility (e.g., increased viral loads, longer infection duration, decreased infectious doses) (Grubaugh et al., 2021). The rapid spread of Delta in many locations around the world suggests that it is innately more transmissible than Alpha and other SARS-CoV-2 variants. However, estimates of Delta’s transmissibility may also vary between populations due to differences in underlying immunity, control measures, behaviors, and demographics. For example, a variant that is more likely to cause vaccine breakthroughs may have a larger observed transmissibility advantage in populations with higher vaccination rates because it can spread to more individuals. Studies conducted in the United Kingdom estimated that Delta is 40-80% more transmissible than Alpha, which itself was more transmissible than the SARS-CoV-2 lineages previously in circulation (SAGE, 2021). The World Health Organization similarly estimated a 55% increase in Delta transmissibility based on data from India and the United Kingdom (Campbell et al., 2021). To understand whether these estimates are applicable elsewhere, it is critical to compare the relative transmissibility of SARS-CoV-2 variants in different locations to test the sensitivity of estimates to population-specific conditions. Accurate variant transmissibility estimates ultimately inform local and regional control measures.

In this study, we posed several important questions that arose with Delta: (***1***) how much more transmissible was Delta than Alpha, (***2***) were the differences in relative transmissibility consistent across state populations, and (***3***) was Delta more transmissible because it caused higher viral loads during infection? To investigate each, we partnered with SARS-CoV-2 genomic surveillance programs from six New England US states: Connecticut, Maine, Massachusetts, New Hampshire, Rhode Island, and Vermont. Using logistic growth rates and estimated effective reproductive numbers (Alpert et al., 2021; Davies et al., 2021; Petrone et al., 2021), we found that Delta was consistently more transmissible than Alpha, but the relative comparison varied substantially across states. Furthermore, we found on average ∼6 times more viral RNA copies per mL from samples collected from Delta infections compared to Alpha infections, supporting the hypothesis that Delta may be more transmissible because it generates higher viral loads. Overall, we estimated that Delta is 58-120% more transmissible than Alpha in New England. We attributed the overall transmissibility advantage of Delta to its innate ability to enhance infections and the range of estimates to the variation in underlying characteristics of different populations.

## Results

### Genomic surveillance revealed similar variant frequency trajectories across New England

In response to emerging SARS-CoV-2 variants, all states within the New England region of the US (Connecticut, Maine, Massachusetts, New Hampshire, Rhode Island, and Vermont) increased their virus sequencing capacity through local and regional partnerships (**Figure 1A**). By late March 2021, at least 5% of the weekly reported COVID-19 cases were being sequenced from each state (with the exception of two weeks in June for Vermont where weekly coverage dropped to 3%), and the maximum daily sequencing coverage ranged from 16% (New Hampshire) to 46% (Maine). From these state-level sequencing data, we tracked the frequencies of the SARS-CoV-2 Alpha variant (B.1.1.7 lineage), the Delta variant (B.1.617.2 and AY.x sub-lineages), and all other lineages (referred to as ‘Other’) (**Figure 1B**). We observed similar trajectories in variant frequencies, with Other declining as Alpha increased in March/April 2021. Beginning in June 2021, Delta rapidly displaced the Alpha and Other lineages. We also observed that the emergence of Delta resulted in a “selective sweep” and more fully dominated the variant landscape compared to Alpha. By the final week in July 2021, Delta comprised the vast majority of sequenced samples in all states (range 94-100%). In contrast, while Alpha was the main variant in early 2021, we still observed other lineages maintained in the population.

**Figure 1.**
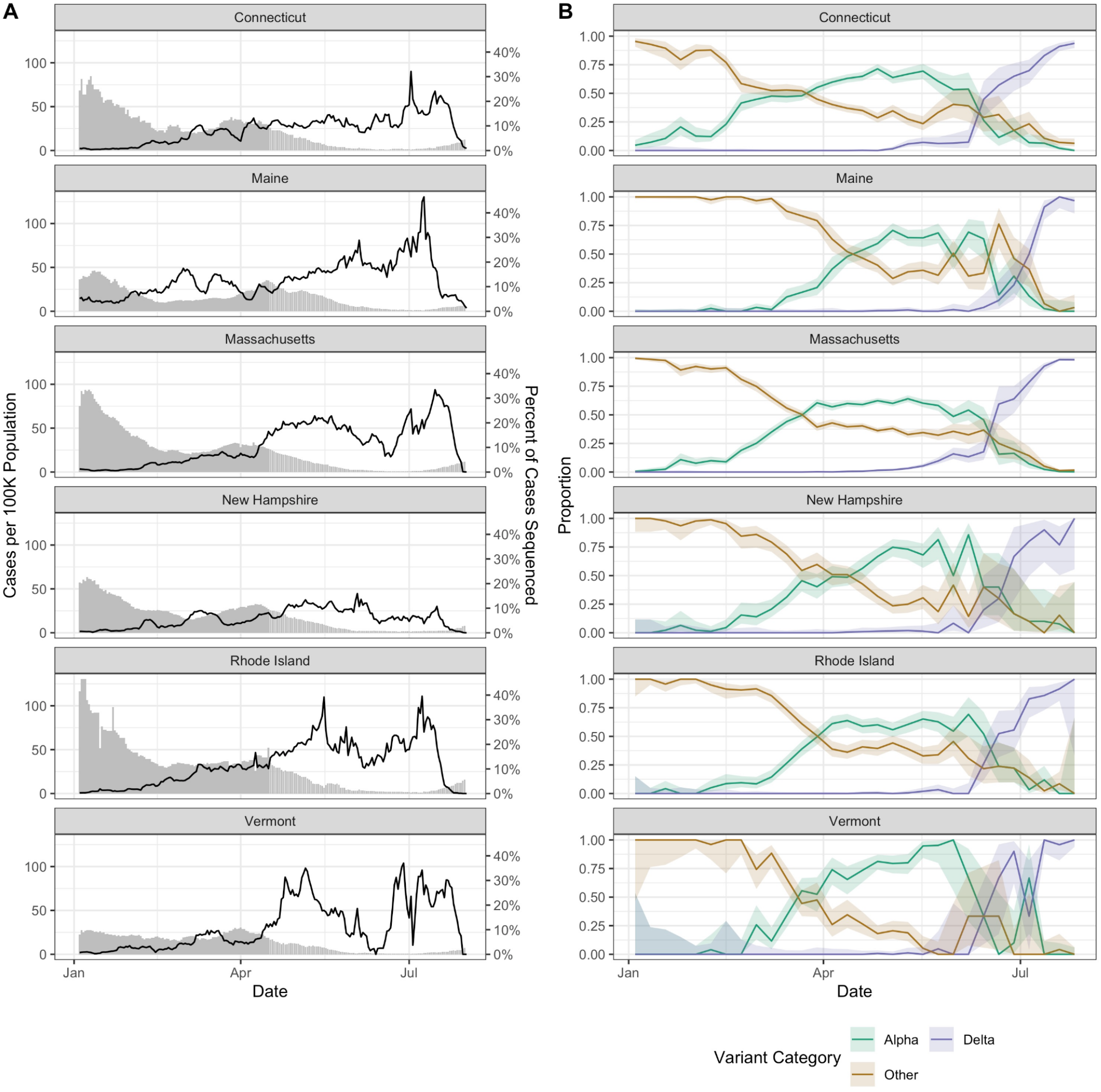
SARS-CoV-2 sequencing coverage and variant frequency tracking. **(A)** Confirmed cases per 100K population (bars) and percent of cases sequenced (lines) by state (7 day rolling average), January-August 2021. The variability in percent of cases sequenced represents changing sample availability and suitability for sequencing. The drop in percent sequenced at the end of August does not reflect real decreases in sequencing coverage but instead (1) the 1-3 week delays between sample collection and sequence availability and (2) how the data are plotted using 7 day rolling average. **(B)** Weekly proportion of sequenced genomes belonging to each variant category with 95% confidence intervals, January-August 2021. A breakdown of the number of genomes by state and lineage is included in **Tables S1-S3**. Complete lists of genomes used from GISAID are available in **Data S1-S7**.

### Delta emerged faster than Alpha and dominated the variant landscape

While the dominance of Delta made it clear that it was more transmissible than the other SARS-CoV-2 variants (**Figure 1**), it was unknown by how much transmissibility estimates varied across populations. We first addressed this knowledge gap by comparing the initial growth rates of Delta and Alpha across New England. Since Alpha and Delta emerged at different times, we defined their emergence periods as the 90 days following their initial detection in each state (**Figure 2A**). We then estimated the logistic growth rate of Alpha and Delta during their respective state-specific 90-day emergence periods (**Figure 2B**). We found that Delta emerged faster than Alpha despite higher vaccination rates during mid-2021.

**Figure 2.**
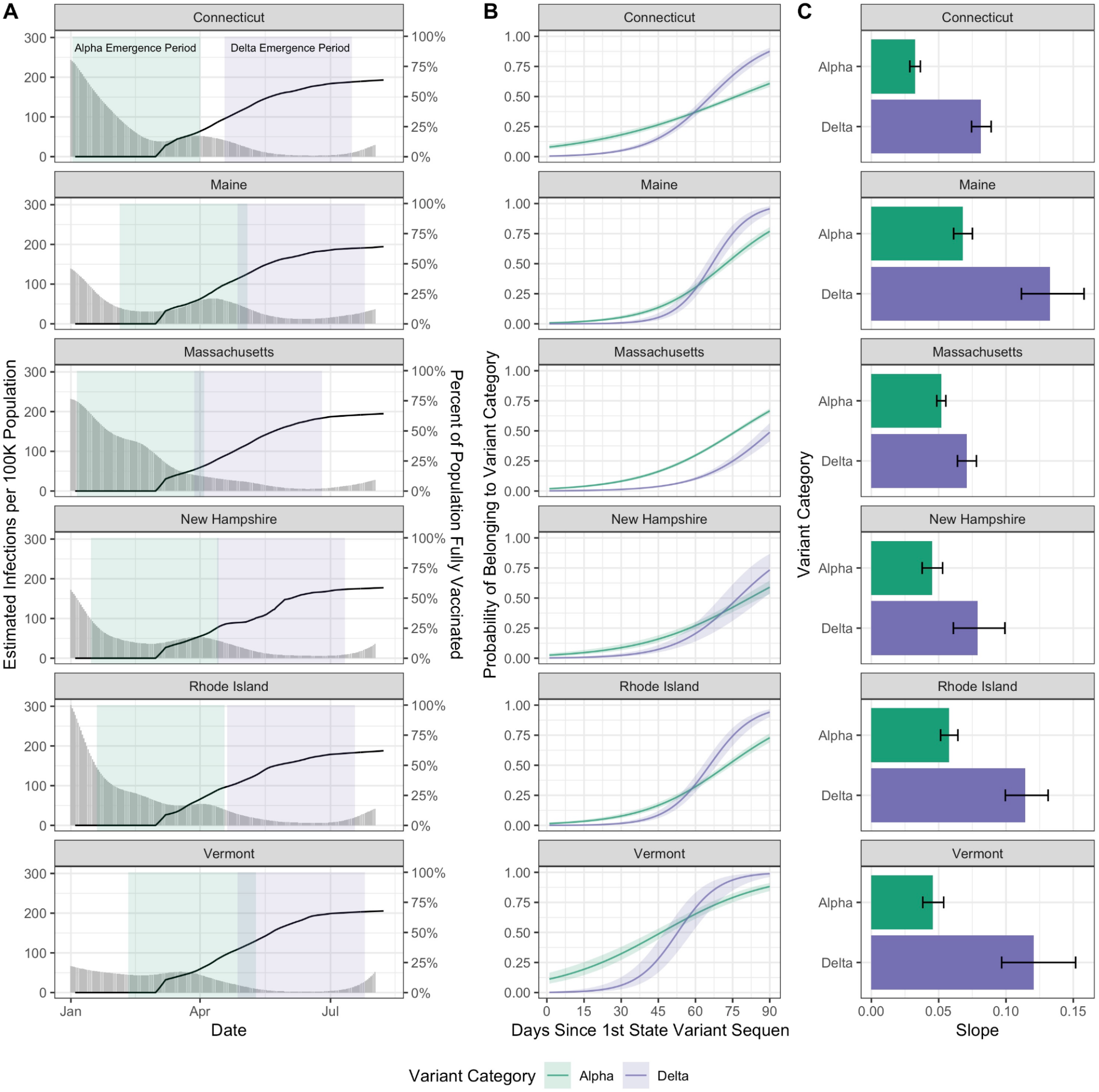
Variant logistic growth rates during their respective emergence periods in the context of infections and vaccination. **(A)** Estimated infections per 100K population (grey bars, left axis) and percent of the population fully vaccinated (black lines, right axis) (7 day rolling average), with the colored rectangles indicating the 90-day emergence periods for each variant. **(B)** We ran a binomial logistic regression with 95% confidence intervals for the variant category as the outcome and the number of days since the first detection as the predictor to estimate the logistic growth rate for Alpha versus Delta. Shown as the predicted probability of a given sequence belonging to each variant category over time. The analysis is restricted to the first 90 days of emergence in each state as shown in (A). **(C)** The regression coefficients (slopes) of the logistic growth rate from (B) with 95% confidence intervals. An initial exploration of the logistic growth rates for Delta compared to Alpha to the vaccination rates or estimated infections per state at the start of the Delta 90-day emergence period can be found in **Figure S1**.

While Alpha appeared to initially outpace Delta, we hypothesized that this was due to gaps in surveillance programs that impeded detection of Alpha but were addressed before Delta emerged. As noted previously (**Figure 1A**), sequencing coverage improved over time in all states as incident cases declined. The probability of a given sequenced sample belonging to Alpha at the start of its emergence period was 11% in Vermont, 8% in Connecticut, and between 1-2% in the remaining states, indicating Alpha likely was circulating for some time before its first detection (**Figure 2B**). In contrast, the probability of a given sequenced sample belonging to Delta was 0% at the start of the emergence period in all states. We estimated that the logistic growth rate for Delta was 163% greater than Alpha in Vermont, 151% in Connecticut, 98% in Rhode Island, 95% in Maine, 75% in New Hampshire, and 37% in Massachusetts (**Figure 2C**). From first sequenced detection, it took Delta on average 71 days (range 54-92 days) to become dominant (surpass 50% predicted frequency; **Figure 2B**). Given that the Alpha and Delta variants circulating across New England are all genetically similar, the differences in the growth rates between states are likely due in part to population-specific factors. As an initial exploration, we compared the increase in the logistic growth rate for Delta compared to Alpha (**Figure S1**) to the vaccination rates or estimated infections (**Figure 2A**) per state at the start of the Delta 90-day emergence period. We noted an association between the relative emergence speed of Delta with state vaccination rates (**Figure S1A);** however, states with earlier Delta detection dates, like Massachusetts, necessarily have lower vaccination rates during the Delta emergence period (**Figure S1C**). We did not note an association between the relative emergence speed of Delta and estimated infections per 100K population in each state (**Figure S1B**).

### Delta was more transmissible than Alpha in all New England states

We showed that Delta emerged faster in New England than Alpha had previously (**Figure 2**) and rose to higher levels of dominance, almost completely displacing Alpha and other lineages (**Figure 1**). However, we still do not know how much more transmissible Delta was than Alpha when they were co-circulating. To answer this question, we used our previously developed framework to estimate the variant-specific effective reproductive number (*R*_t_) (Chitwood et al., 2021; Petrone et al., 2021). Our *R*_t_ estimates for each variant approximate the time-varying average number of secondary cases from a primary infection within a population. *R*_t_ estimates greater than 1 imply that COVID-19 cases associated with a variant will increase in the future. Delta had a *R*_t_ > 1 for the majority of the time period following its emergence that exceeded the *R*_t_ estimates for Alpha and Other (**Figure 3**).

**Figure 3.**
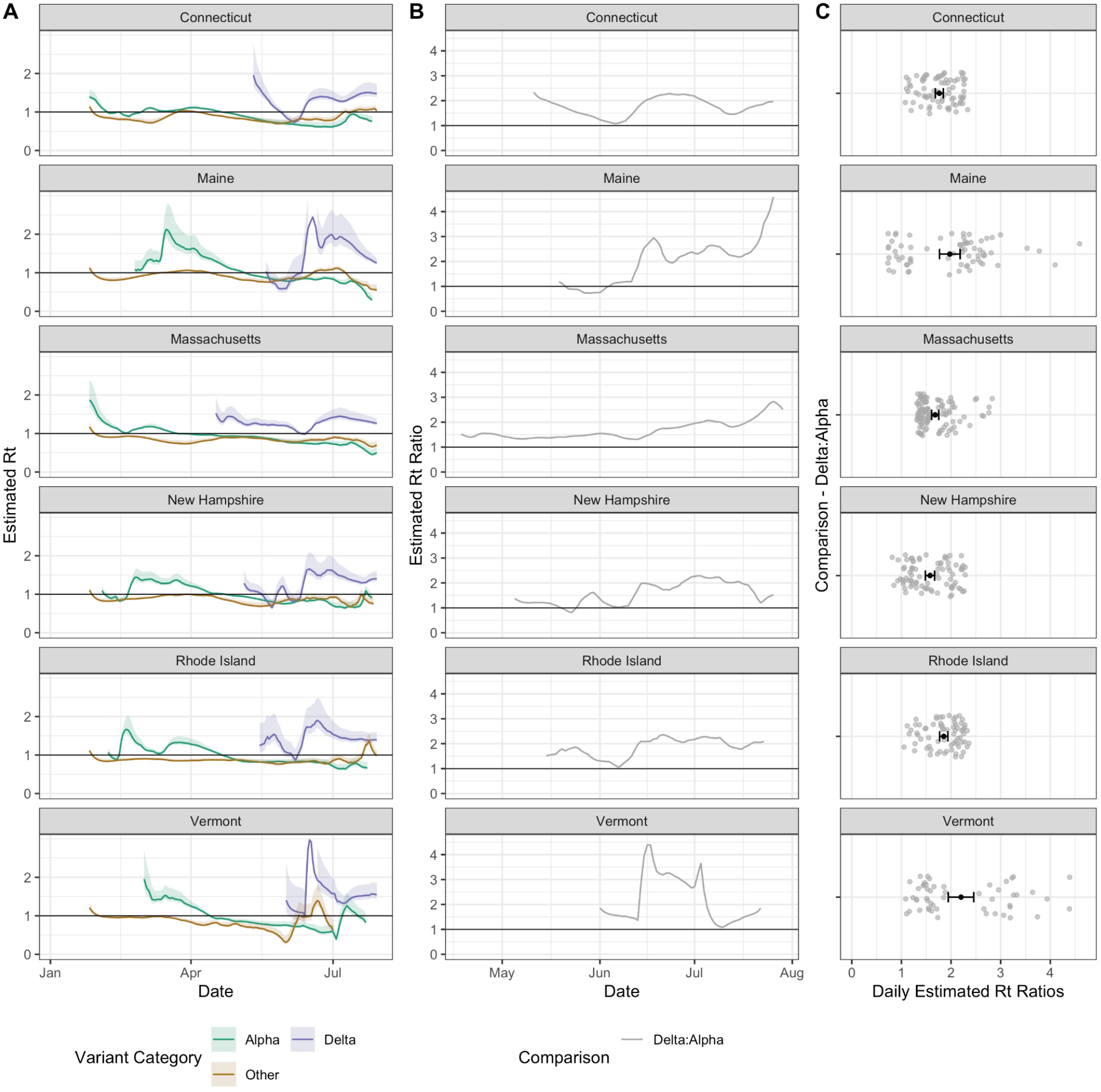
Comparison of variant effective reproductive numbers to estimate relative transmissibility. **(A)** Estimated effective reproductive number *R*_t_ over time for each variant category. We multiplied estimated infections from *Covidestim* by variant sequencing frequencies over time (each using a 7 day rolling average), and then calculated the variant-specific *R*_t_ using *EpiEstim* (Chitwood et al., 2021; Cori et al., 2013). Note that the y-axis differs from that in (B). The upper limit of Delta’s confidence intervals in Maine and Vermont are not plotted but reach a maximum value of 3.57 and 4.51, respectively. The multiplicative increases in *R*_t_ versus the strength of support by variant category and state is shown in **Figure S2**. **(B)** Daily ratios of *R*_t_ values for Delta compared to Alpha from (A). **(C)** Daily estimated *R*_t_ ratios for Delta compared to Alpha over the period displayed in (B) with the mean and 95% confidence intervals.

We computed *R*_t_ for each variant category during January-August 2021 (**Figure 3A**) by combining the frequency estimates from our genomic surveillance data (**Figure 1B**) with daily estimated SARS-CoV-2 infections (**Figure 2A**). Our mean *R*_t_ estimates for Other was < 1 for all states, ranging from 0.84 (Massachusetts) to 0.90 (Maine). Prior to the emergence of Delta, our mean *R*_t_ for Alpha was 1.22 across states, dropping to a mean *R*_t_ of 0.81 as vaccination increased during the period following Delta’s emergence. Our mean *R*_t_ estimate for Delta was 1.36, ranging from 1.25 (New Hampshire) to 1.57 (Vermont). We found that the *R*_t_ for Delta exceeded that of Alpha in all states for the majority of the time following its initial detection (**Figure 3B**). We then estimated that the mean *R*_t_ ratio of Delta to Alpha was 2.20 in Vermont, 1.98 in Maine, 1.85 in Rhode Island, 1.76 in Connecticut, 1.68 in Massachusetts, and 1.58 in New Hampshire (**Figure 3C**), suggesting that Delta was on average 58-120% more transmissible than Alpha depending on the state.

In addition, we calculated the multiplicative increase in *R*_t_ for Delta versus Alpha, another measure of relative transmissibility (Davies et al., 2021) (**Figure S2**). For this estimate we exponentiated the coefficients from the binomial logistic regression and multiplied them by the mean generation interval to estimate the change in the probability of a given sequence belonging to a lineage over a generation interval. The multiplicative increase in *R*_t_ for Delta was greater than that for Alpha in all states. Across states, we observed a mean 1.30 increase in the probability of a sample belonging to Alpha over a generation interval, compared to 1.69 for Delta. Our multiplicative increase in *R*_t_ estimates suggests that Delta had the greatest advantage in Maine (1.99x increase) and Vermont (1.87x increase), and the lowest advantage in New Hampshire (1.51x) and Massachusetts (1.45x), corroborating our *R*_t_ ratio estimates (**Figure 3C**).

### Delta infections on average had a higher number of viral RNA copies than Alpha infections

One potential mechanism for Delta’s increased transmissibility relative to Alpha (**Figures 2 and 3**) is that infections with the Delta variant could lead to higher virus titers than those with Alpha. To test this hypothesis, we compared the RT-qPCR cycle threshold (CT) values of sequence-confirmed Alpha and Delta infections (nasal swabs) from four institutes in New England: Yale University (Connecticut), Jackson Laboratory (Connecticut), Mass General Brigham (Massachusetts), and the Health and Environmental Testing Laboratory (Maine) (**Figure 4**). PCR CT values are a metric of virus RNA copies, and lower CT values indicate that there are more copies. We consistently found lower CT values for Delta infections across all institutes, but some small sample sizes meant that not all comparisons showed significant differences.

**Figure 4.**
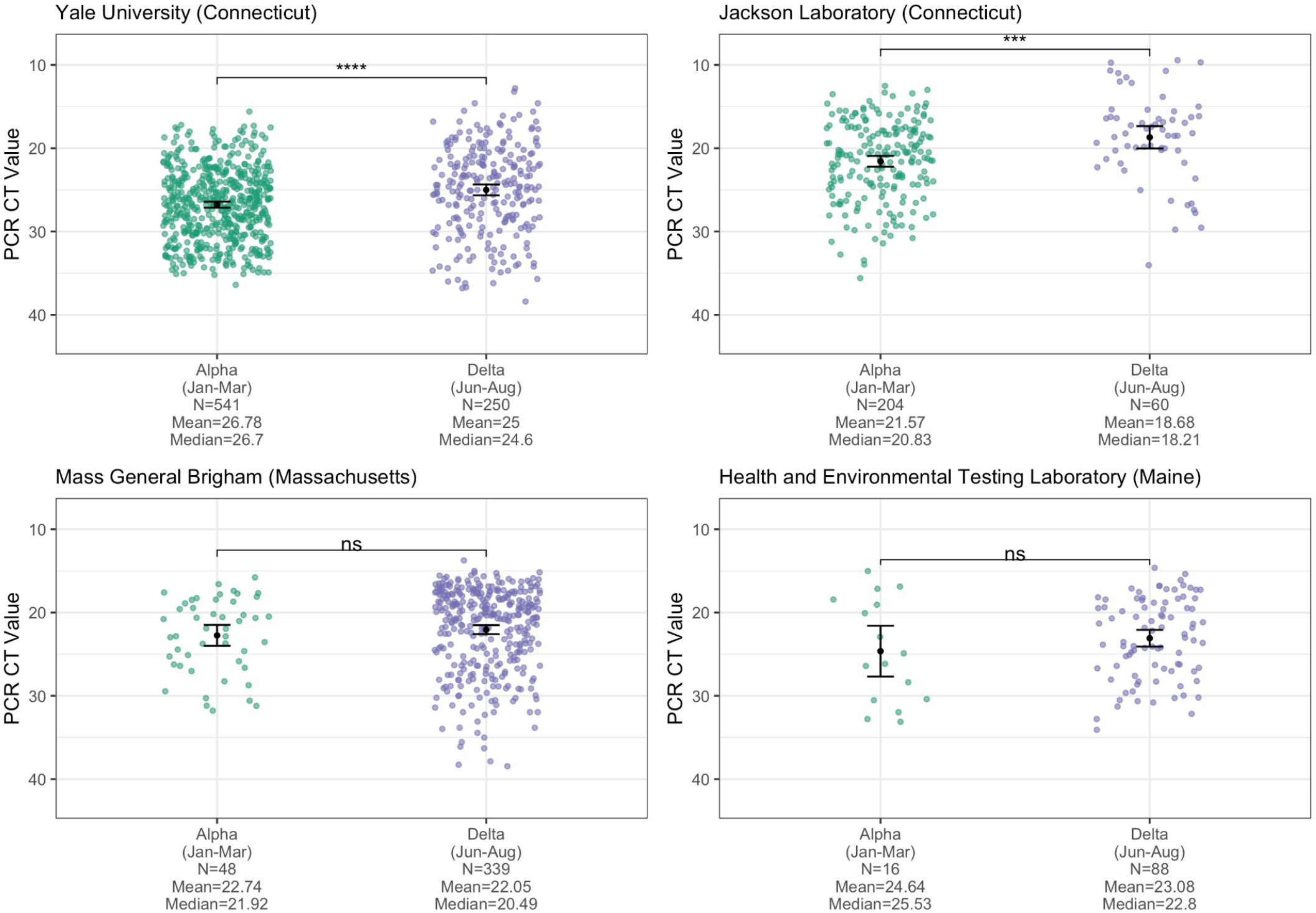
Cross-sectional PCR data from Alpha and Delta samples. PCR CT values (inverted y-axis) plotted by institute and variant category, limiting Alpha samples to January-March 2021 and Delta samples to June-August 2021 to account for their respective emergence periods. Monthly CT values for Alpha are shown in **Figure S3**. For each institute, the means of the two variant categories were compared using a t-test, with statistical significance symbols corresponding to the following values: ns (p > 0.05), *** (p ≤ 0.001), and **** (p ≤ 0.0001). Data from the full month of August were not available for most of the institutes at the time of analysis. The ‘Yale University (Connecticut)’ data are from the N1 primer/probe set (originally from the “CDC assay”) of a research use only RT-PCR assay to discriminate among variants (Vogels et al., 2021) (data shown as virus RNA copies/mL in **Figure S4**). The ‘Jackson Laboratory (Connecticut)’ data are from the N gene primer/probe set of the TaqPath COVID-19 Combo Kit (ThermoFisher). The Mass General Brigham (Massachusetts) data are from the E gene of the Roche Cobas 6800 test (ORF1a data shown in **Figure S5**). The ‘Health and Environmental Testing Laboratory (Maine)’ data are from the N1 primer/probe set (originally from the “CDC assay”) of the OPTI SARS-CoV-2 RT-PCR Test (OPTI Medical Systems).

Importantly, PCR CT values from cross-sectional tests can be biased by the epidemic period because viral loads are dynamic and tend to decrease with time (Hay et al., 2021). During the emergence phase of an epidemic, most PCR tests come from recent infections, while the opposite is true when the epidemic is declining. The result is that PCR CT values could be higher (meaning less virus detected) during the declining phase even though the infection dynamics are the same throughout the epidemic. We first investigated if PCR CT values for a variant change during different epidemic phases by comparing the monthly CT values from Alpha generated by one of the institutes (Yale University), and did not find a clear pattern of mean CT values for Alpha increasing as the variant declined in May-July 2021 (**Figure S3**). Still, to account for any effects that the epidemic period may have on our comparisons, we limited our analysis to the approximate emergence phase of each variant: January-March 2021 for Alpha and June-August for Delta. Furthermore, the PCR data that we used from the four institutes are from different assays and some target different genes (though most target the nucleocapsid [N] gene). The Yale University data are from the N1 primer/probe set (originally from the “CDC assay”) of a ‘research use only’ assay (Vogels et al., 2021); the Health and Environmental Testing Laboratory data are from the same N1 primer/probe set at Yale but from the OPTI SARS-CoV-2 RT-PCR Test; the Jackson Laboratory data are from the N gene primer/probe set of the TaqPath COVID-19 Combo Kit; and the Mass General Brigham data are from the envelope (E) and open reading frame 1a (ORF1a) gene primer/probe set of the Roche Cobas 6800 test. Therefore, we analyzed the PCR CT values independently for each institute and gene target.

Assessing cross-sectional PCR data from the four institutions in New England, we consistently found lower mean CT values (more viral RNA copies) from Delta compared to Alpha nasal swab samples (**Figure 4**). The differences were significant from the Yale University (*p* ≤ 0.0001) and Jackson Laboratory data (*p* ≤ 0.001), but not from Mass General Brigham and the Health and Environmental Testing Laboratory (each *p* > 0.05), likely due to small sample sizes and/or changing CT cutoff criteria for sequencing (e.g. the Alpha CT values from Mass General Brigham do not extend above 31, suggesting a stringent cutoff). In addition, for the Yale University samples, we used a standardized PCR curve to translate the CT values into viral RNA copies per mL (Vogels et al., 2020). We found 6.06 times more RNA copies per mL (non-log scale) on average in Delta nasal swab samples compared to Alpha samples (**Figure S4**). Thus, during their respective emergence periods, nasal swabs collected from individuals infected with Delta on average had higher viral copies than from Alpha infections, possibly contributing to enhanced transmissibility.

## Discussion

In mid-2021, the SARS-CoV-2 variant Delta emerged across the US, displacing previous variants, including Alpha. Determining how much more transmissible Delta was than Alpha across different settings remains an important public health question. With more than 3 million SARS-CoV-2 genomes available on public repositories, large scale and globally diverse assessments can be conducted. However, there is always a risk when analyzing diverse sources of data without input from the data submitters on possible sampling biases that may not be apparent in the repositories. Here we directly partnered with SARS-CoV-2 genomic surveillance programs from across six states in the New England region of the US to confidently assess relative transmissibility at the state level (**Figure 1**). We found that the logistic growth rates (**Figure 2**) and effective reproductive numbers (**Figure 3**) of Delta were greater than Alpha across all New England states, although there was considerable variation between states. Our estimates of the transmission advantage (measured as the mean *R*_t_ ratio of Delta to Alpha) for New Hampshire, Massachusetts, and Connecticut (range 58-76%) were within the 40-80% estimate range provided by the UK (SAGE, 2021). We estimated a greater transmission advantage for Delta in Rhode Island (85%), Maine (98%), and Vermont (120%). This variation may be driven by differences in the underlying state populations, such as population density, vaccination rates, travel patterns, control measures, behaviors, and competing variants in circulation.

The conditions under which each variant emerged could have influenced their initial growth rates. At the time of Alpha’s emergence in January/February 2021, 0% of the population across New England was reported as being fully vaccinated (**Figure 2A**). In comparison, when Delta first emerged in March/April 2021, 18-37% of the population was fully vaccinated. Estimated infections per 100K population were also substantially lower during Delta’s emergence period (**Figure 2A**). However, with rising vaccination rates, all of the states began relaxing capacity constraints from late February to late March 2021. States continued rolling back COVID-19 mitigation measures with the majority lifted by the end of May 2021, although some states maintained indoor masking for unvaccinated individuals (Ballotpedia, 2021). Finally, the emergence of Delta occurred within the background of other variants, including Alpha (**Figure 1B**). Thus, the fitness landscape for SARS-CoV-2 variants may have changed dramatically during 2021, potentially playing out differently across the states and explaining the large range in relative growth rates between Delta and Alpha (**Figure 2C**).

Our study adds to the growing evidence that Delta may be more transmissible in part by causing higher viral loads (often approximated by PCR CT values) during acute infections (Despres et al., 2021; Kislaya et al., 2021; Kissler et al., 2021; Li et al., 2021; Teyssou et al., 2021). We found significantly lower mean PCR CT values (corresponding to more viral RNA copies) for Delta versus Alpha infections from nasal swab samples tested by Yale University and Jackson Laboratory in Connecticut (**Figure 4**). For the Yale data, we used a standardized curve to translate the CT values into virus copies and found that Delta samples had on average ∼6 times more viral RNA copies per mL than Alpha samples (**Figure S4**). The overall pattern of lower CT values for Delta versus Alpha held for the Massachusetts and Maine data, although the differences in mean values were not significant. This may be due to relatively small sample sizes for Alpha in those locations and/or changing CT cutoff criteria for sequencing.

An important limitation to our comparative analyses was that sequencing coverage improved in states over time (**Figure 1A**), and thus there may have been a greater delay in the time to the first sequenced detection for Alpha versus Delta. In addition, due to the uncertainty around the serial interval, we selected an uncertain serial interval approach to explore various possible distributions when calculating *R*_t_ (**Methods**). While Vermont had the highest relative growth rates and mean *R*_t_ ratio comparing Delta and Alpha, it also had more variable sequencing coverage due to relatively low case counts (**Figure 1A**). Variant frequencies based on a relatively small number of sequences likely are driving some of the variability and uncertainty around its *R*_t_ estimates (**Figure 3A**), and therefore results for Vermont should be interpreted more cautiously. For the PCR CT analysis, we necessarily used only confirmed variant sequence data, which represents a fraction of the total PCR CT data and is biased against higher CT values that lack sufficient virus RNA for sequencing. It is also important to be cautious in interpreting CT values among institutes due to differences in the diagnostic PCR test platforms and subsequent CT cut-off values. For this reason, we do not aim to directly compare institutes, but rather sought to establish whether an overall pattern of lower CT values for Delta versus Alpha samples held. Mean CT values can also be influenced by the epidemic trajectory, as discussed previously. To assess whether we observe this in our data, we plotted Alpha CT values by month for the Yale data (**Figure S3**) and did not find a pattern in monthly mean CT values indicative of a bias due to the epidemic trajectory. Lastly, we conducted our analysis at the state level, however there could be within-state heterogeneity in our estimates. Drivers of heterogeneity could include a lack of even sequencing coverage across each state. We attempted to assess the within-state location of each sequence used in our analysis, but only the state location was typically available among publicly available data. In addition, there could be differences in population demographics, behavior, immunity, and/or control measures at more local levels that could lead to more heterogeneous estimates.

In conclusion, while we determined that the Delta SARS-CoV-2 variant was more transmissible than Alpha in New England, there was substantial spatial heterogeneity in our estimates across states (range 58-120%). Our analysis demonstrated that, in addition to possible innate differences between variants, there were likely other factors at play such as vaccination levels, demographics, behavior, control measures, etc. that affected each variant’s transmissibility within a given population. The exact mechanisms driving differences in variant transmissibility between states are unclear, but these factors remain important when considering how another variant might rise to dominance in the future. Considering Delta’s current apparent dominance in most locations with an active SARS-CoV-2 genomic surveillance system, the next variant to displace Delta may evolve from within the B.1.617.2 clade. It is impossible to predict when and where the next variant of concern will emerge, and thus it is important to enhance SARS-CoV-2 genomic surveillance and further our understanding of how different population characteristics impact variant transmissibility.

## Methods

### Ethics

#### Yale University (Connecticut)

The Institutional Review Board from the Yale University Human Research Protection Program determined that the RT-qPCR testing and sequencing of de-identified remnant COVID-19 clinical samples obtained from clinical partners conducted in this study is not research involving human subjects (IRB Protocol ID: 2000028599).

#### Jackson Laboratory (Connecticut)

The Institutional Review Board of The Jackson Laboratory determined that use of de-identified residual COVID-19 clinical samples obtained from the Clinical Genomics Laboratory for RT-qPCR testing and sequencing for this study is not research involving human subjects (IRB Determination: 2020-NHSR-021).

#### Mass General Brigham (Massachusetts)

The Institutional Review Board of Partners Human Research determined that the use of excess, de-identified COVID-19 clinical specimens obtained within the Partners Healthcare network for RT-qPCR testing and genomic sequencing for this study is not research involving human subjects (IRB Protocol ID: 2019P003305). In addition, the Institutional Review Board of the Massachusetts Department of Public Health has reviewed and approved this study to perform genomic sequencing of coronaviruses (IRB Protocol ID: 1603078).

#### Health and Environmental Testing Laboratory (Maine)

A Memorandum of Understanding between The State of Maine Department of Health and Human Services and The Jackson Laboratory determined that extracted viral RNA from human respiratory specimens which have tested positive for SARS-CoV-2 and are used for sequencing will not be used in human subjects, in clinical trials, or for diagnostic purposes involving human subjects.

### Genomic surveillance data

We obtained SARS-CoV-2 genomic sequence data from GISAID as of August 13, 2021. We restricted the dataset to our states of interest (Connecticut, Maine, Massachusetts, New Hampshire, Rhode Island, and Vermont) and removed sequences that lacked a lineage assignment (N=4977), had mismatched metadata for the sample collection location (N=81), or were outside of our January 1, 2021 - August 1, 2021 time period of interest (N=4260). This yielded a final dataset of 33,408 genomes. We categorized the GISAID SARS-CoV-2 genomes into 3 mutually exclusive categories: Alpha (B.1.1.7), Delta (B.1.617.2 and all AY.x sub-lineages existing as of August 13, 2021), and Other (any sequences not included in the prior categories that had a lineage assignment). A breakdown of the number of genomes by state and lineage is included in **Tables S1-S3**. Complete lists of genomes used from GISAID are available in **Data S1-S7**.

### Transmissibility estimates

All figures were plotted using RStudio (v 1.4.1106).

#### Confirmed cases per 100K population

We obtained confirmed case data from the Johns Hopkins Center for Systems Science and Engineering. We used state population estimates for 2019 from the United States Census Bureau to calculate confirmed cases per 100K population (**Figure 1A**).

#### Sequencing coverage

We calculated sequencing coverage per state as the number of curated sequenced genomes made publicly available on GISAID divided by the number of confirmed cases, and plotted the data based on the sample collection date (**Figure 1A**).

#### Variant frequencies among sequenced samples

We divided the number of sequences belonging to each variant category (Delta, Alpha, or Other) by the total number of sequences to estimate variant frequencies. To reflect the uncertainty in sequencing frequencies, we calculated a Jeffreys’ interval, using the resulting 0.025 and 0.975 quantiles to form a 95% confidence interval (**Figure 1B**).

#### Percent of the population fully vaccinated

We obtained the percent of the state populations that were fully vaccinated from the Centers for Disease Control and Prevention (dataset downloaded August 11, 2021) (**Figure 2A**).

#### Infections per 100K Population

We obtained estimated infections for each state from *Covidestim*, a Bayesian nowcasting approach that accounts for differences in diagnosis and reporting by anchoring its estimates to death data, which are generally more reliable than case data (dataset downloaded August 15, 2021) (Chitwood et al., 2021). We used state population estimates for 2019 from the United States Census Bureau to calculate estimated infections per 100K population (**Figure 2A**).

#### Logistic growth rates

We first defined the emergence period for Alpha versus Delta in each state as the time from each variant category’s first phylogenetic detection in the GISAID data to 90 days afterward. We ran a binomial logistic regression for Alpha and Delta, separately, with the variant category as the outcome and the number of days since the first detection as the predictor. We plotted the smoothed fitted curves for the emergence periods with their 95% confidence intervals (**Figure 2B**), which shows the probability of a given sequence belonging to a specific variant category over time. We also report the logistic regression coefficients as the log odds of a given sequence belonging to a specific variant category **(Figure 2C**).

#### Effective reproductive number R_t_ estimates and R_t_ ratios

Following a previous approach used to estimate the comparative transmissibility of SARS-CoV-2 variants (Petrone et al., 2021), we multiplied daily estimated infections from *Covidestim* by the variant frequencies (calculated on a 7 day rolling average, with any negative values induced by the rolling average set to 0) to get daily estimated numbers of infections for each variant category. Within the *EpiEstim* R package (Cori et al., 2013), we used an uncertain serial interval to estimate *R*_t_ over 21 day sliding windows, with the mean serial interval of 5.2 days (allowed to vary between 2.2 and 8.2 days) and a standard deviation of 4 days (allowed to vary between 2.5 and 5.5 days) based on estimates available in the literature (Alene et al., 2021; Du et al., 2020; Li et al., 2020). For Vermont, we removed a single ‘Other’ variant category sequence on July 21, 2021 (after the ‘Other’ variant category had died out) that was leading to highly inflated *R*_t_ estimates. We began *R*_t_ estimation when there were at least 12 cumulative estimated infections to achieve a posterior coefficient of variation of 0.3 (Cori et al., 2013) and restricted estimation to before August 1, 2021 as variant frequencies became less certain after that time due to delays in sequencing and reporting. We further truncated the variant-specific *R*_t_ estimates in states where estimated variant-specific infections went consistently to 0 (i.e. died out). EpiEstim assumes an uninformative prior for mean *R*_t_ of 5, ensuring that when there are few infections (e.g., 0 variant-specific infections for the majority of the 21-day estimation window) the prior becomes disproportionately weighted relative to the data and *R*_t_ estimates erroneously seem to increase. Therefore, we truncated the *R*_t_ estimates at the date when they reached their lowest value after variant-specific infections died out. For Alpha *R*_t_ estimates, we truncated the following states: Connecticut, Maine and New Hampshire (July 26, 2021); Rhode Island (July 23, 2021);and Vermont (July 22, 2021). For Other *R*_t_ estimates, we truncated the following states: New Hampshire (July 27, 2021) and Vermont (July 1, 2021). We did not truncate any of the Delta *R*_t_ estimates as Delta infections grew in all states. We used the same serial interval parameters for all variant categories due to the lack of consensus regarding differences (Pung et al., 2021; Ryu et al., 2021). We reported the resulting mean *R*_t_ estimates and the 95% confidence intervals (**Figure 3A**). We plotted the daily *R*_t_ ratio for Delta versus Alpha over time (**Figure 3B**) and as a dotplot with the mean and 95% confidence intervals (**Figure 3C**).

#### Multiplicative reproductive number estimates

We calculated the multiplicative increase in *R*_t_ for Delta versus Alpha, a measure of relative transmissibility by multiplying the coefficients from the previously described binomial logistic regression by the mean generation interval, 5.2 days, and exponentiating to get an estimate of the increase in the probability of a given sequence belonging to a lineage over a generation interval (Davies et al., 2021). We plotted this against the log-transformed *p*-value associated with each coefficient (**Figure S2**) to provide the level of support for the multiplicative value estimate.

### RT-qPCR and lineage identification

All figures were plotted using RStudio (v 1.4.1106).

#### Yale University (Connecticut)

Clinical samples (nasal swabs in viral transport media) were received from confirmed SARS-CoV-2 positive individuals from routine testing provided by Yale New Haven Hospital. The samples were primarily from Connecticut and were collected for a variety of inpatient and outpatient testing programs. Nucleic acid was extracted from 300µL of the original sample using the MagMAX viral/pathogen nucleic acid isolation kit, eluting in 75µl of elution buffer. The extracted nucleic acid was tested for SARS-CoV-2 RNA using a “research use only” (RUO) RT-qPCR assay (Vogels et al., 2021). For this analysis, only the CT values from the CDC N1 primer/probe set were used (**Figure 4**). N1 CT values were also converted into SARS-CoV-2 RNA copies using a standard curve, as previously described (Vogels et al., 2020) (**Figure S4**).

To determine the SARS-CoV-2 lineage, samples with CT values ≤ 35 were sequenced using the Illumina COVIDSeq Test RUO version. Amplicons were pooled and cleaned before quantification with Qubit High Sensitivity dsDNA kit. The resulting libraries were sequenced using a 2×150 approach on an Illumina NovaSeq at the Yale Center for Genomic Analysis. Each sample was given at least 1 million reads. Samples were typically processed in sets of 93 or 94 with negative controls incorporated during the RNA extraction, cDNA synthesis, and amplicon generation steps. The reads were aligned to the Wuhan-Hu-1 reference genomes (GenBank MN908937.3) using BWA-MEM v.0.7.15 (Li, 2013). Adaptor sequences were trimmed, primer sequences were masked, and consensus genomes were called (simple majority >60% frequency) using iVar v1.3.1 (Grubaugh et al., 2019) and SAMtools (Danecek et al., 2021). An ambiguous ‘N’ was used when fewer than 20 reads were present at a site. In all cases, negative controls were analyzed and confirmed to consist of at least 99% Ns. Pangolin v.2.4.2 (O’Toole et al., 2021) was used to assign lineages (Rambaut et al., 2020b).

#### Jackson Laboratory (Connecticut)

Clinical samples were received in The Jackson Laboratory Clinical Genomics Laboratory (CGL) as part of a statewide (Connecticut) COVID-19 surveillance program, with the majority of samples representing asymptomatic screening of nursing home and assisted living facility residents and staff. Total nucleic acids were extracted from anterior nares swabs in viral transport media or saline (200µL) using the MagMAX Viral RNA Isolation kit (ThermoFisher) on a KingFisher Flex purification system. Samples were tested for the presence of SARS-CoV-2 RNA using the TaqPath COVID-19 Combo Kit (ThermoFisher). For this analysis, only the CT values from the N gene primer/probe set were used (**Figure 4**).

Samples with CT values ≤ 30 for the N gene target were prepared for sequencing using the Illumina COVIDSeq Test kit. Sequencing was performed on an Illumina NovaSeq or NextSeq in the CGL. Data analysis was performed using the DRAGEN COVID Lineage App in BaseSpace Sequence Hub. Sequences with >80% of bases with non-N basecalls and ≥1500-fold median coverage were considered successful and were submitted to GISAID. Lineages were assigned using pangolin v.2.4.2 (O’Toole et al., 2021) and the most current version of the pangoLEARN assignment algorithm.

#### Mass General Brigham (Massachusetts)

Clinical samples were received from confirmed SARS-CoV-2 positive tests collected at Massachusetts General Brigham testing facilities during routine testing, via nasal swabs in viral transport media. Clinical samples were tested for the presence of SARS-CoV-2 RNA using the Roche Cobas 6800 test, targeting both the E and ORF1a genes. CT values were analyzed independently for both the E (**Figure 4**) and ORF1a (**Figure S5**) gene primer/probe sets.

Genomic sequencing was conducted on clinical samples with CT values ≤ 30 using the Illumina COVIDSeq Test protocol. The resulting libraries were pooled, cleaned, and quantified using the Qubit High Sensitivity dsDNA kit. Sequencing was performed at Massachusetts General Hospital or at the Broad Institute of MIT and Harvard using a 2×150 approach on an Illumina NovaSeq SP, an Illumina NextSeq 550, or an Illumina NextSeq 2000. Sequences were analyzed through the Broad Institute Data Analysis Platform using the viral-ngs 2.1.28 on the Terra platform (app.terra.bio). All of the workflows named below are publicly available via the Dockstore Tool Registry Service (dockstore.org/organizations/BroadInstitute/collections/pgs). Sequences with an assembly length >24000 non-N bases were considered complete genomes. Lineages were assigned using the most up-to-date version of the pangoLEARN assignment algorithm.

#### Health and Environmental Testing Laboratory (Maine)

Clinical samples (nasal swabs in viral transport media) were received from confirmed SARS-CoV-2 positive tests collected at the State of Maine Department of Health and Human Services testing facilities (Health and Environmental Testing Laboratory (HETL)) during routine testing. Viral RNA were extracted on a Thermofisher KingFisher Flex purification using the MagMAX Viral/Pathogen II Nucleic Acid Isolation Kit. Extracted samples were tested for the presence of SARS-CoV-2 RNA using the OPTI SARS-CoV-2 RT-PCR Test (OPTI Medical Systems) with the ABI 7500fast DX thermocycler. For this analysis, only the CT values from the N1 gene primer/probe set were used (**Figure 4**).

Prior to sequencing, samples were not reanalyzed for the presence of SARS-CoV-2 RNA. HETL utilized the QIAseq DIRECT SARS-CoV-2 Kit from Qiagen for targeted whole genome library preparation. Samples were sequenced on an Illumina MiSeq using a MiSeq® Reagent Kit v2 (300 cycle) kit. QIAGEN CLC Genomics Workbench was used to assemble viral genomes. Consensus sequences were uploaded to Nextstrain and Pango for quality control, strain and clade identification.

Additional SARS-CoV-2 positive samples from HETL were sequenced by the Jackson Laboratory (Maine) using the Illumina COVID-Seq protocol modified to include 6e2 copies/μL of a unique SDSI control spiked into each cDNA sample. Samples were sequenced on a NextSeq500 using paired 75 bp reads by the Genome Technology group on Jackson Laboratory’s Bar Harbor campus. Sequence reads were analyzed using an in-house pipeline (https://github.com/tewhey-lab/SARS-CoV-2-Consensus) that leverages minimap2, samtools and iVar for read mapping and variant calling. Sequences with >80% non-N bases were considered complete genomes for analysis.

### RT-qPCR CT value and virus RNA comparisons

We binned the RT-qPCR CT and virus RNA data (separately) for Alpha (January-March 2021) and Delta (June-August 2021) samples and compared the group means for each institute using a t-test, with statistical significance symbols corresponding to the following values: ns (p > 0.05), * (p ≤ 0.05), ** (p ≤ 0.01), *** (p ≤ 0.001), **** (p ≤ 0.0001) (**Figure 4, Figure S4-S5**).

## Supporting information

DataS1-7

## Data Availability

A summary of the SARS-CoV-2 lineages used from GISAID are available in Tables S1-S3 and a complete list of GISAID acknowledgements are available in Data S1-S7. All other publicly available data and code used for this paper are available on github (https://github.com/grubaughlab/2021_paper_Delta-v-Alpha).

## Data and code availability

A summary of the SARS-CoV-2 lineages used from GISAID are available in **Tables S1-S3** and a complete list of GISAID acknowledgements are available in **Data S1-S7**. All other data and code used for this paper are available on github (https://github.com/grubaughlab/2021_paper_Delta-v-Alpha).

## New England Variant Investigation Team Authors

Ahmad Altajar, Alexandra DeJesus, Anderson Brito, Anne E. Watkins, Anthony Muyombwe, Caleb Neal, Chaney C. Kalinich, Chen Liu, Christine Loreth, Christopher Castaldi, Claire Pearson, David Ferguson, Erika Buzby, Eva Laszlo, Gina Vicente, Heidi Munger, Hillary Johnson, Irina R. Tikhonova, Isabel M. Ott, Jafar Razeq, Jessica Brown, Jianhui Wang, Johanna Vostok, John P Beauchamp, Joshua Hall, Lawrence C. Madoff, Lori Webber, Luc Gagne, Mary Barter, Niall Lennon, Nicholas Fitzgerald, Nicholas Kerantzas, Pei Hui, Rachel Harrington, Randy Downing, Rashida Haye, Ryan Lynch, Scott Hennigan, Seana Cofsky, Selina Clancy, Shrikant Mane, Stacey Gabriel, Stephanie Baez, Stephanie Ash, Steve Fleming, Steven Murphy, Tara Alpert, Trevor Rivard, Wade Schulz

## Acknowledgments

We thank The Broad Genomics Platform, The Broad Clinical Research Sequencing Program, and The Broad Data Sciences Platform for their tremendous work making data available for surveillance and research, members of the New England ‘variant’ group that meet weekly to discuss and share data regarding emerging SARS-CoV-2 variants, the groups that continuously make their sequencing data available to the public, the frontline and essential workers for their service during the pandemic, D. MacCannell for his ongoing support and permission to use the data generated by the CDC, Y. Grad for technical advice, and our friends and family - particularly V. Parsons, P. Jack, and S. Taylor - for their support. This work was funded by CTSA Grant Number TL1 TR001864 (R.E. and M.E.P.), Fast Grant from Emergent Ventures at the Mercatus Center at George Mason University (N.D.G.), the Centers for Disease Control and Prevention (CDC) Broad Agency Announcement # 75D30120C09570 (N.D.G.) and # 75D30120C09605 (B.L.M), and CDC Baseline Surveillance Contract # 75D30121C10501 (Clinical Research Sequencing Platform, LLC).

## Author Contributions

Conceptualization, R.E., J.E.L., N.D.G.;

Performed genomic surveillance, all authors;

Provided PCR data, R.U., N.M., N.R., M.I.B., R.T.B., K.B., C.B.F.V., S.T., M.L.L., D.R.P., K.K., G.O., H.G., S.M., J.M., R.T., M.D.A., J.E.L., N.D.G., New England Variant Investigation Team;

Methods development, M.E.P., J.E.R., R.T.K., J.E.L., N.D.G;

Data analysis and interpretation, R.E., J.E.L., N.D.G.;

Supervision, R.T., M.D.A., D.J.P., J.E.L., N.D.G.;

Writing - Original Draft, R.E., M.E.P., C.B.F.V., N.D.G;

Writing - Review & Editing, all authors.

## Supplementary Information

**Table S1.**
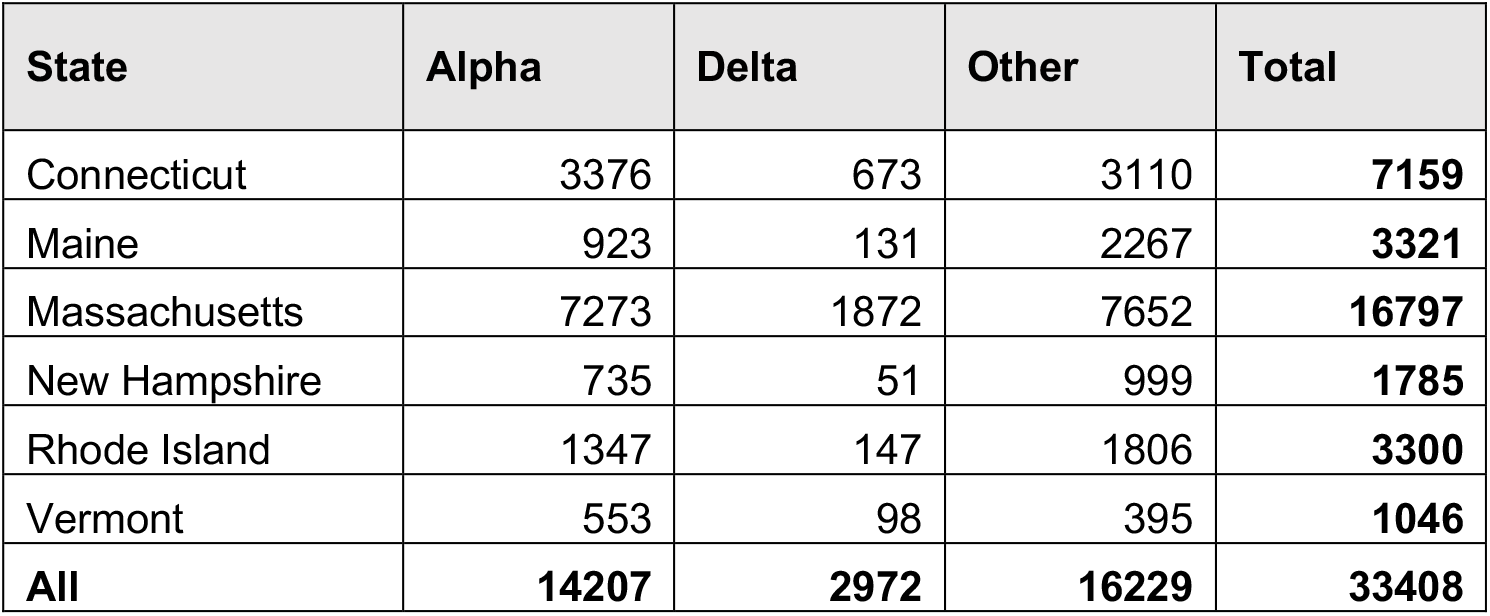
Number of genomes used in this study by state and variant category. (pdf)

**Table S2.**
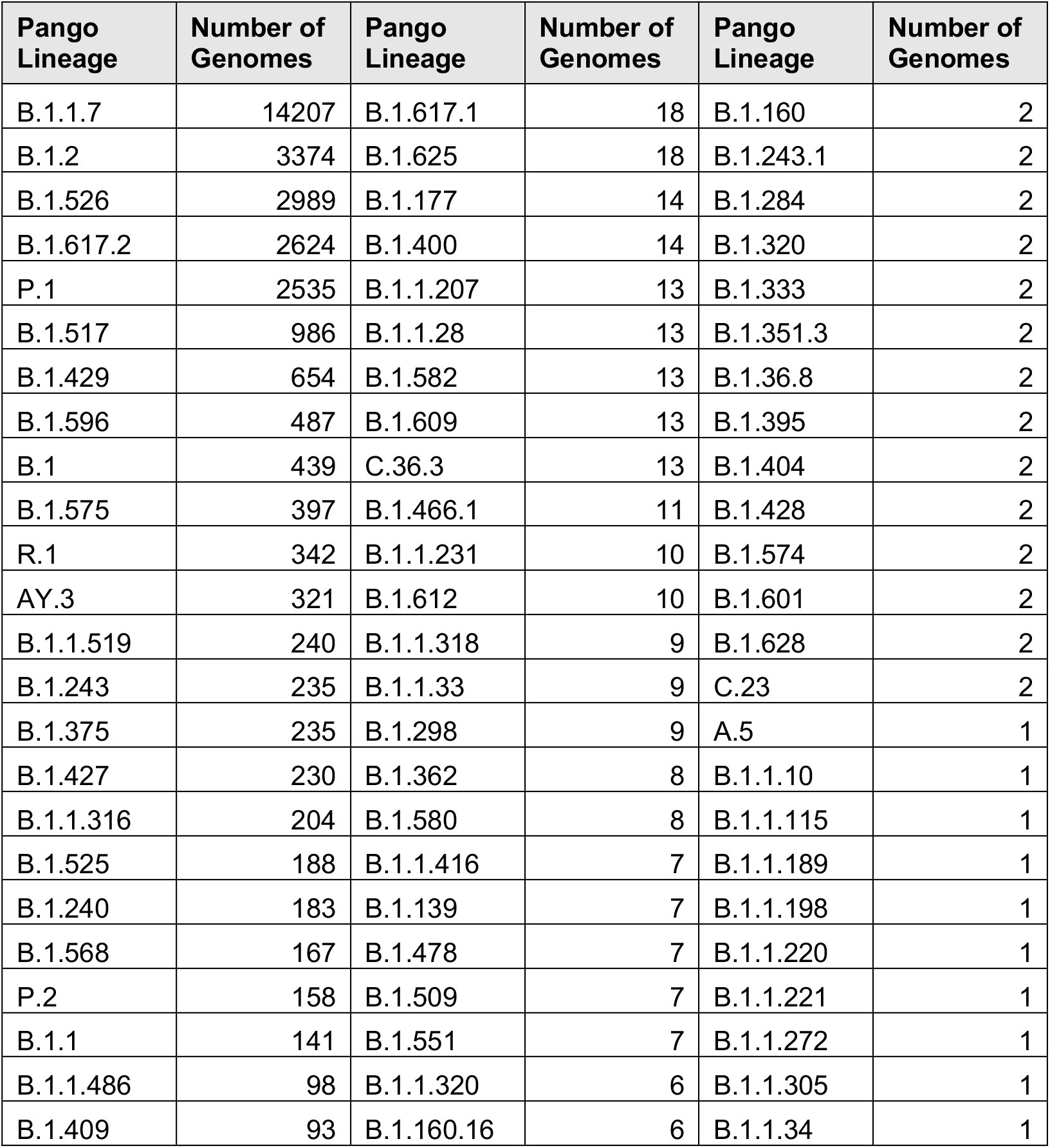
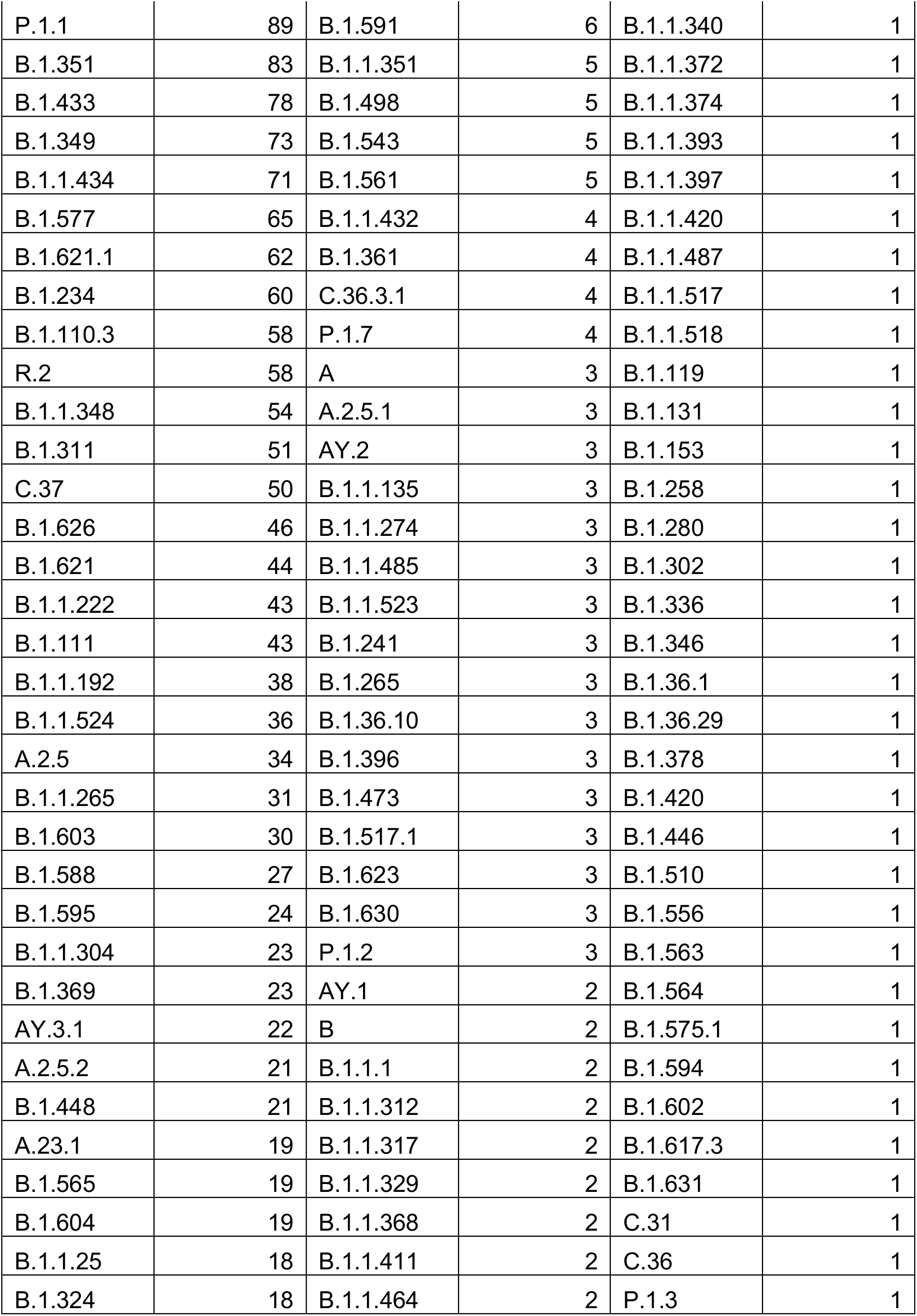
Number of genomes used in this study by pango lineage. (pdf)

**Table S3.**
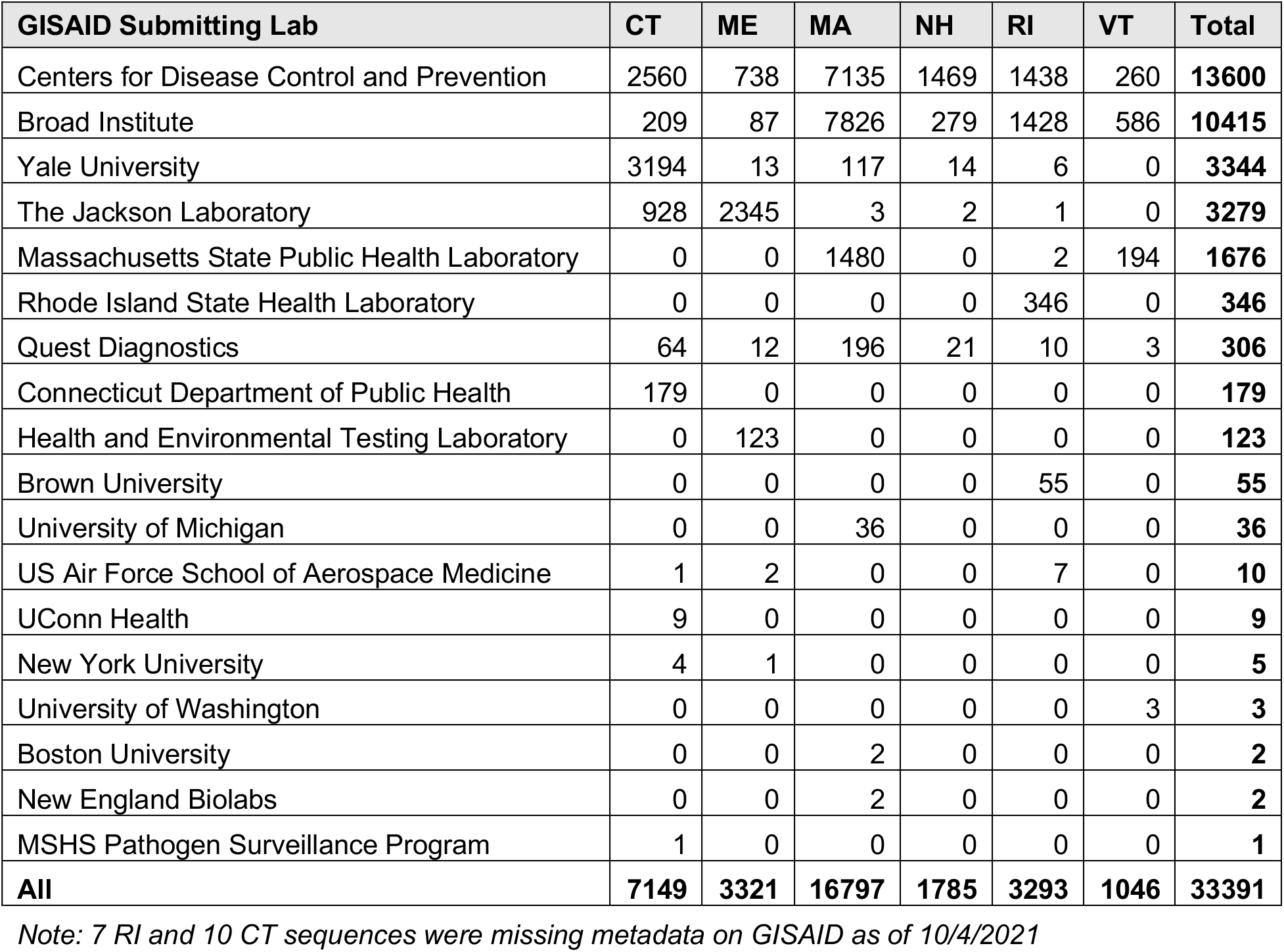
Number of genomes used in this study by submitting lab and state. (pdf)

**Data S1**. List of SARS-CoV-2 sequences from Connecticut used in this study and author acknowledgments. (pdf)

**Data S2**. List of SARS-CoV-2 sequences from Maine used in this study and author acknowledgments. (pdf)

**Data S3**. List of SARS-CoV-2 sequences from Massachusetts (part 1) used in this study and author acknowledgments. (pdf)

**Data S4**. List of SARS-CoV-2 sequences from Massachusetts (part 2) used in this study and author acknowledgments. (pdf)

**Data S5**. List of SARS-CoV-2 sequences from New Hampshire used in this study and author acknowledgments. (pdf)

**Data S6**. List of SARS-CoV-2 sequences from Rhode Island used in this study and author acknowledgments. (pdf)

**Data S7**. List of SARS-CoV-2 sequences from Vermont used in this study and author acknowledgments. (pdf)

**Figure S1.**
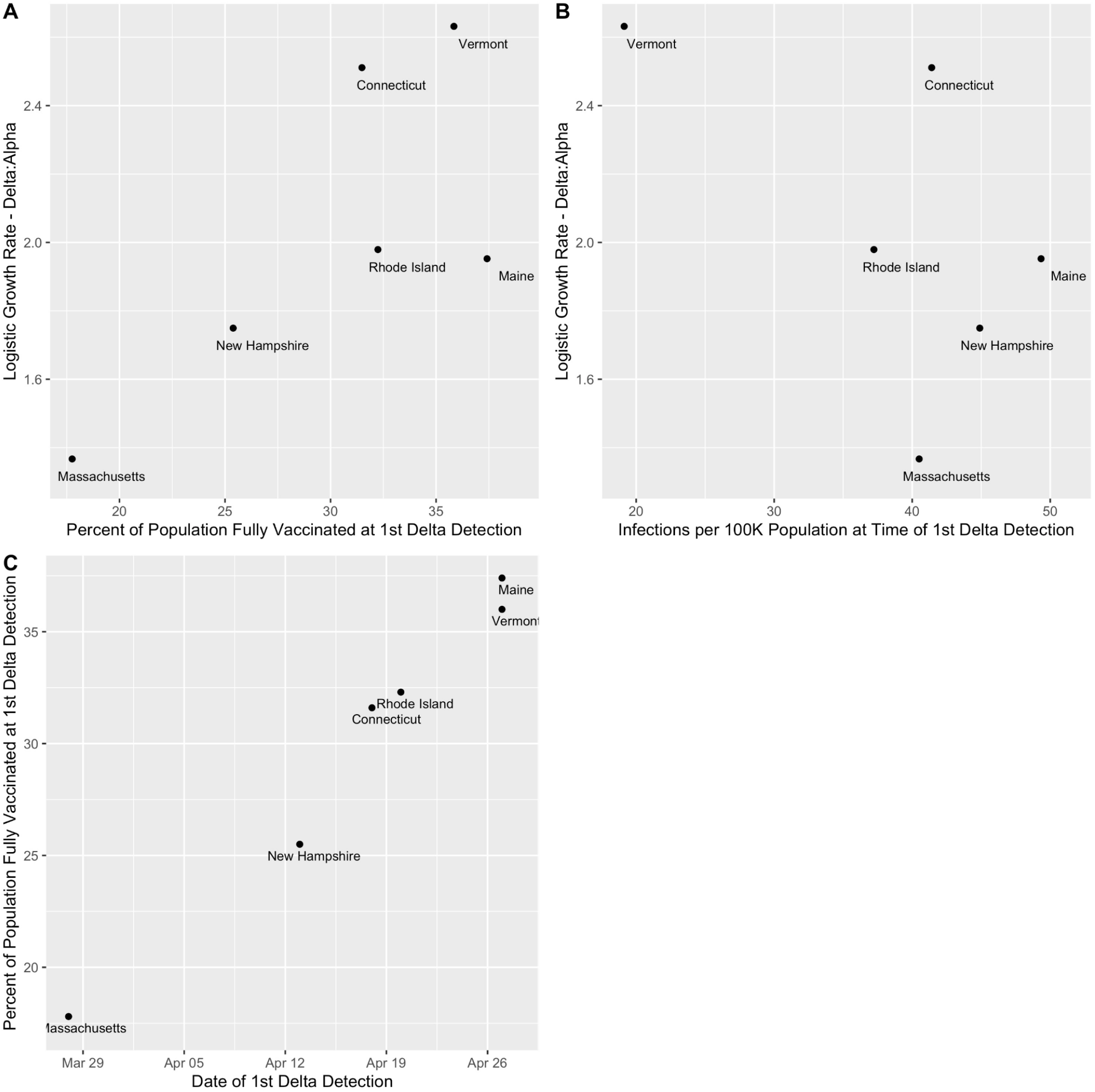
Associations between variant logistic growth rates, vaccination status, estimated infections, and date of first Delta detection, related to Figure 2. **(A)** Percent of the population fully vaccinated (7 day rolling average) at the time of the initial Delta detection in each state versus the relative logistic growth rates of Delta versus Alpha (calculated over the 90 day emergence period). **(B)** Estimated infections per 100K population at the time of the initial Delta detection in each state versus the relative logistic growth rates of Delta versus Alpha (calculated over the 90 day emergence period). **(C)** Date of the initial Delta detection in each state versus the percent of the population fully vaccinated (7 day rolling average) at that time.

**Figure S2.**
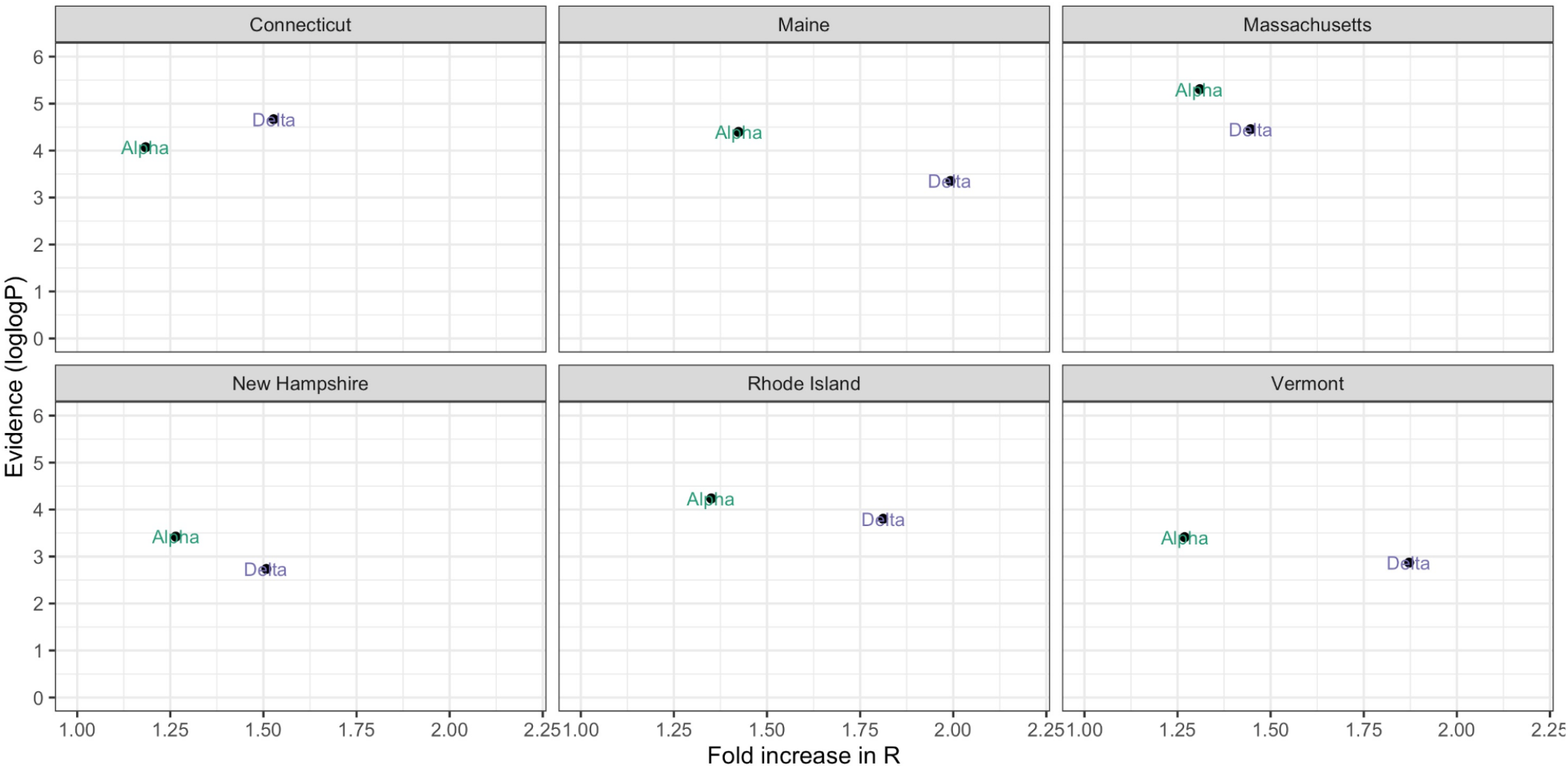
Multiplicative increase in *R*_*t*_ versus the strength of support by variant category and state during the initial 90 day emergence period following 1st detection, related to Figure 3. We ran a binomial logistic regression with the variant category as the outcome and the number of days since the 1st detection as the predictor. The multiplicative increase is calculated by multiplying the regression coefficient by the mean generation interval of 5.2 days and exponentiating. The strength of support is the logged p-value for the coefficient.

**Figure S3.**
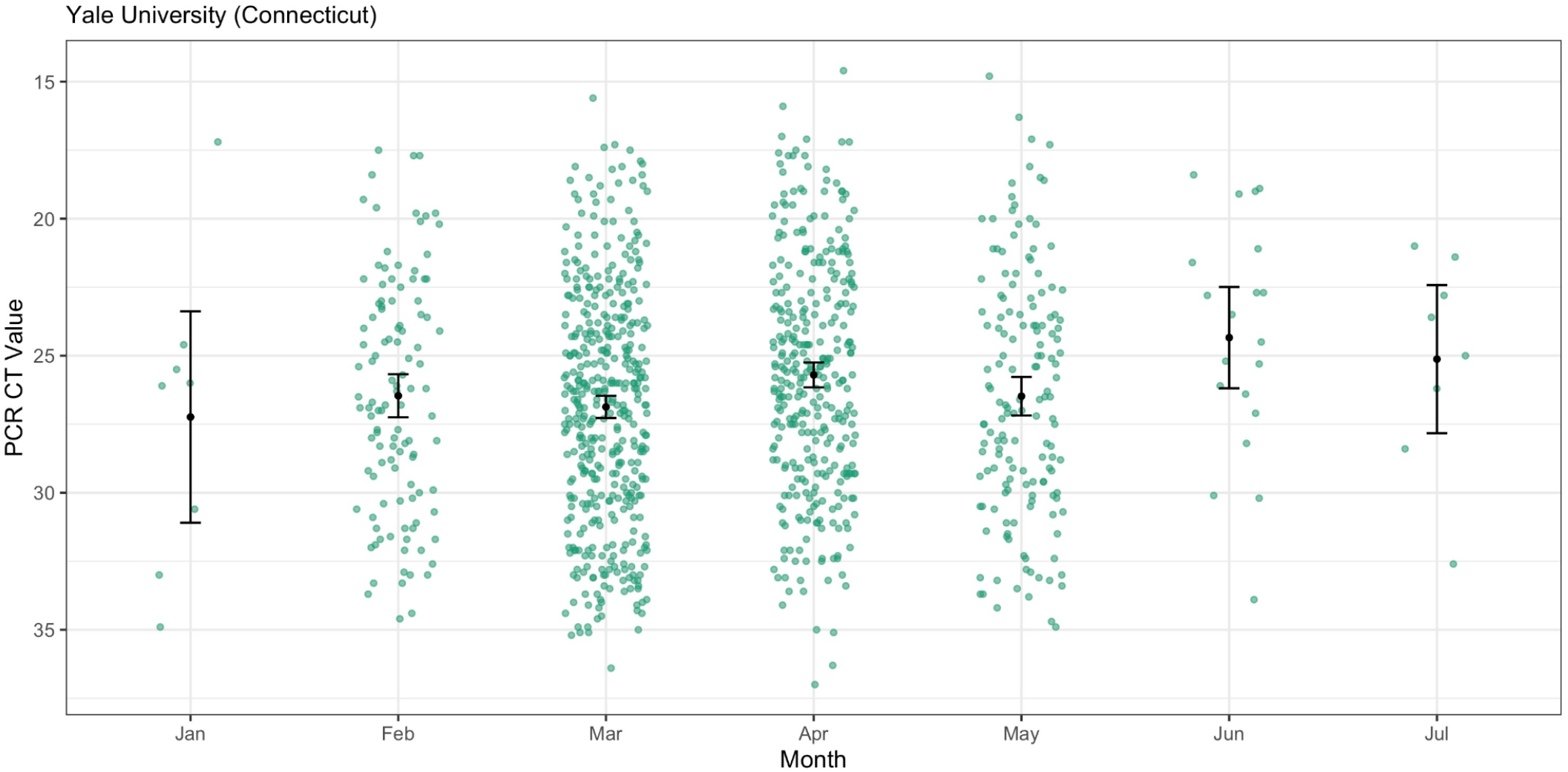
Monthly Alpha PCR CT values, related to Figure 4. We plotted the CT values of Yale University Alpha samples from January-July 2021 with the mean and 95% confidence intervals. There were no Alpha samples for August 2021.

**Figure S4.**
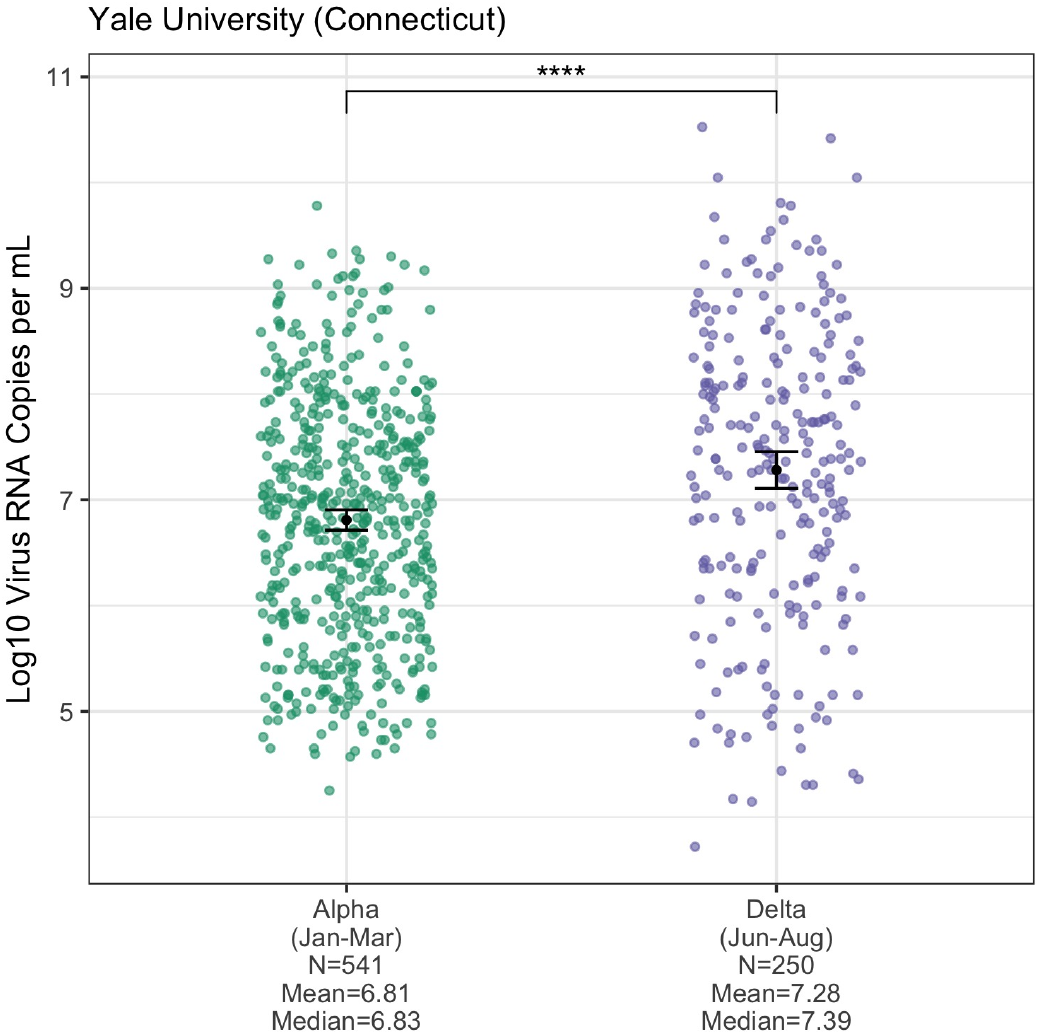
Virus copy calculation from PCR CT values, related to Figure 4. We used a standardized curve to translate the Yale PCR CT values into log10 virus RNA copies per mL and added the mean and 95% confidence intervals. We removed 4 Alpha samples from early January 2021 that were tested using a different PCR assay. Alpha samples were again limited to January-March and Delta samples to June-August. The means were compared using a t-test.

**Figure S5.**
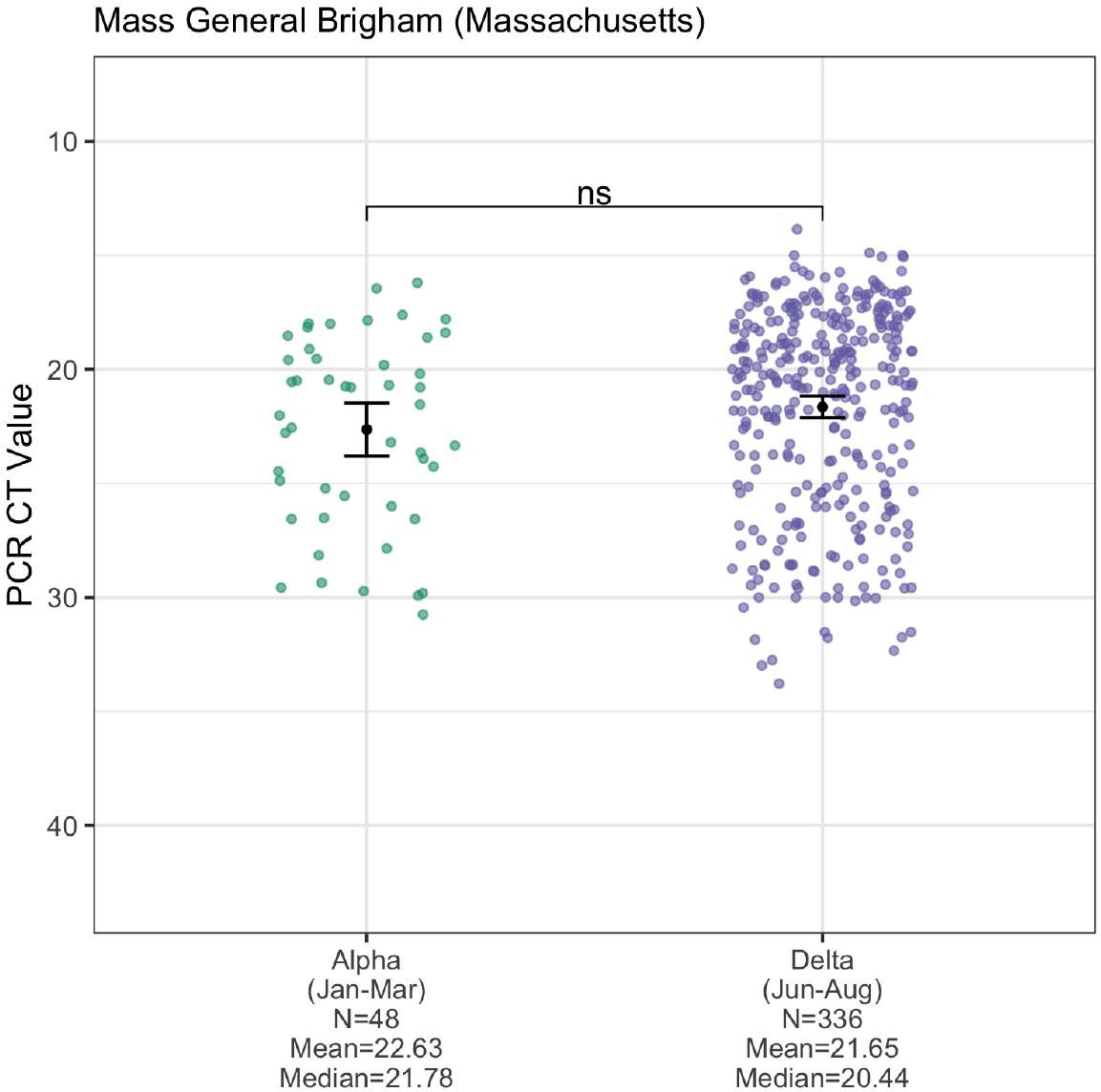
ORF1a target CT values, related to Figure 4. PCR CT values (inverted y-axis) plotted by variant category, limiting Alpha samples to January-March 2021 and Delta samples to June-August 2021 to account for their respective emergence periods. The means of the two variant categories were compared using a t-test. The Mass General Brigham (Massachusetts) data are from the ORF1 gene of the Roche Cobas 6800 test (E gene data shown in **Figure 4**). Three samples were dropped from the ORF1a analysis due to NA values.

