## Supplementary material for "Comparative transmissibility of SARS-CoV-2 variants Delta and Alpha in New England, USA": TableS1

| State | Alpha | Delta | Other | Total |
|---|---|---|---|---|
| Connecticut | 3376 | 673 | 3110 | **7159** |
| Maine | 923 | 131 | 2267 | **3321** |
| Massachusetts | 7273 | 1872 | 7652 | **16797** |
| New Hampshire | 735 | 51 | 999 | **1785** |
| Rhode Island | 1347 | 147 | 1806 | **3300** |
| Vermont | 553 | 98 | 395 | **1046** |
| **All** | **14207** | **2972** | **16229** | **33408** |
