## Supplementary material for "Comparative transmissibility of SARS-CoV-2 variants Delta and Alpha in New England, USA": TableS2

| Pango Lineage | Number of Genomes | Pango Lineage | Number of Genomes | Pango Lineage | Number of Genomes |
|---|---|---|---|---|---|
| B.1.1.7 | 14207 | B.1.617.1 | 18 | B.1.160 | 2 |
| B.1.2 | 3374 | B.1.625 | 18 | B.1.243.1 | 2 |
| B.1.526 | 2989 | B.1.177 | 14 | B.1.284 | 2 |
| B.1.617.2 | 2624 | B.1.400 | 14 | B.1.320 | 2 |
| P.1 | 2535 | B.1.1.207 | 13 | B.1.333 | 2 |
| B.1.517 | 986 | B.1.1.28 | 13 | B.1.351.3 | 2 |
| B.1.429 | 654 | B.1.582 | 13 | B.1.36.8 | 2 |
| B.1.596 | 487 | B.1.609 | 13 | B.1.395 | 2 |
| B.1 | 439 | C.36.3 | 13 | B.1.404 | 2 |
| B.1.575 | 397 | B.1.466.1 | 11 | B.1.428 | 2 |
| R.1 | 342 | B.1.1.231 | 10 | B.1.574 | 2 |
| AY.3 | 321 | B.1.612 | 10 | B.1.601 | 2 |
| B.1.1.519 | 240 | B.1.1.318 | 9 | B.1.628 | 2 |
| B.1.243 | 235 | B.1.1.33 | 9 | C.23 | 2 |
| B.1.375 | 235 | B.1.298 | 9 | A.5 | 1 |
| B.1.427 | 230 | B.1.362 | 8 | B.1.1.10 | 1 |
| B.1.1.316 | 204 | B.1.580 | 8 | B.1.1.115 | 1 |
| B.1.525 | 188 | B.1.1.416 | 7 | B.1.1.189 | 1 |
| B.1.240 | 183 | B.1.139 | 7 | B.1.1.198 | 1 |
| B.1.568 | 167 | B.1.478 | 7 | B.1.1.220 | 1 |
| P.2 | 158 | B.1.509 | 7 | B.1.1.221 | 1 |
| B.1.1 | 141 | B.1.551 | 7 | B.1.1.272 | 1 |
| B.1.1.486 | 98 | B.1.1.320 | 6 | B.1.1.305 | 1 |
| B.1.409 | 93 | B.1.160.16 | 6 | B.1.1.34 | 1 |
| P.1.1 | 89 | B.1.591 | 6 | B.1.1.340 | 1 |
| B.1.351 | 83 | B.1.1.351 | 5 | B.1.1.372 | 1 |
| B.1.433 | 78 | B.1.498 | 5 | B.1.1.374 | 1 |
| B.1.349 | 73 | B.1.543 | 5 | B.1.1.393 | 1 |
| B.1.1.434 | 71 | B.1.561 | 5 | B.1.1.397 | 1 |
| B.1.577 | 65 | B.1.1.432 | 4 | B.1.1.420 | 1 |
| B.1.621.1 | 62 | B.1.361 | 4 | B.1.1.487 | 1 |
| B.1.234 | 60 | C.36.3.1 | 4 | B.1.1.517 | 1 |
| B.1.110.3 | 58 | P.1.7 | 4 | B.1.1.518 | 1 |
| R.2 | 58 | A | 3 | B.1.119 | 1 |
| B.1.1.348 | 54 | A.2.5.1 | 3 | B.1.131 | 1 |
| B.1.311 | 51 | AY.2 | 3 | B.1.153 | 1 |
| C.37 | 50 | B.1.1.135 | 3 | B.1.258 | 1 |
| B.1.626 | 46 | B.1.1.274 | 3 | B.1.280 | 1 |
| B.1.621 | 44 | B.1.1.485 | 3 | B.1.302 | 1 |
| B.1.1.222 | 43 | B.1.1.523 | 3 | B.1.336 | 1 |

| | | | | | |
|---|---|---|---|---|---|
| B.1.111 | 43 | B.1.241 | 3 | B.1.346 | 1 |
| B.1.1.192 | 38 | B.1.265 | 3 | B.1.36.1 | 1 |
| B.1.1.524 | 36 | B.1.36.10 | 3 | B.1.36.29 | 1 |
| A.2.5 | 34 | B.1.396 | 3 | B.1.378 | 1 |
| B.1.1.265 | 31 | B.1.473 | 3 | B.1.420 | 1 |
| B.1.603 | 30 | B.1.517.1 | 3 | B.1.446 | 1 |
| B.1.588 | 27 | B.1.623 | 3 | B.1.510 | 1 |
| B.1.595 | 24 | B.1.630 | 3 | B.1.556 | 1 |
| B.1.1.304 | 23 | P.1.2 | 3 | B.1.563 | 1 |
| B.1.369 | 23 | AY.1 | 2 | B.1.564 | 1 |
| AY.3.1 | 22 | B | 2 | B.1.575.1 | 1 |
| A.2.5.2 | 21 | B.1.1.1 | 2 | B.1.594 | 1 |
| B.1.448 | 21 | B.1.1.312 | 2 | B.1.602 | 1 |
| A.23.1 | 19 | B.1.1.317 | 2 | B.1.617.3 | 1 |
| B.1.565 | 19 | B.1.1.329 | 2 | B.1.631 | 1 |
| B.1.604 | 19 | B.1.1.368 | 2 | C.31 | 1 |
| B.1.1.25 | 18 | B.1.1.411 | 2 | C.36 | 1 |
| B.1.324 | 18 | B.1.1.464 | 2 | P.1.3 | 1 |
