## Supplementary material for "Comparative transmissibility of SARS-CoV-2 variants Delta and Alpha in New England, USA": TableS3

| GISAID Submitting Lab | CT | ME | MA | NH | RI | VT | Total |
|---|---|---|---|---|---|---|---|
| Centers for Disease Control and Prevention | 2560 | 738 | 7135 | 1469 | 1438 | 260 | **13600** |
| Broad Institute | 209 | 87 | 7826 | 279 | 1428 | 586 | **10415** |
| Yale University | 3194 | 13 | 117 | 14 | 6 | 0 | **3344** |
| The Jackson Laboratory | 928 | 2345 | 3 | 2 | 1 | 0 | **3279** |
| Massachusetts State Public Health Laboratory | 0 | 0 | 1480 | 0 | 2 | 194 | **1676** |
| Rhode Island State Health Laboratory | 0 | 0 | 0 | 0 | 346 | 0 | **346** |
| Quest Diagnostics | 64 | 12 | 196 | 21 | 10 | 3 | **306** |
| Connecticut Department of Public Health | 179 | 0 | 0 | 0 | 0 | 0 | **179** |
| Health and Environmental Testing Laboratory | 0 | 123 | 0 | 0 | 0 | 0 | **123** |
| Brown University | 0 | 0 | 0 | 0 | 55 | 0 | **55** |
| University of Michigan | 0 | 0 | 36 | 0 | 0 | 0 | **36** |
| US Air Force School of Aerospace Medicine | 1 | 2 | 0 | 0 | 7 | 0 | **10** |
| UConn Health | 9 | 0 | 0 | 0 | 0 | 0 | **9** |
| New York University | 4 | 1 | 0 | 0 | 0 | 0 | **5** |
| University of Washington | 0 | 0 | 0 | 0 | 0 | 3 | **3** |
| Boston University | 0 | 0 | 2 | 0 | 0 | 0 | **2** |
| New England Biolabs | 0 | 0 | 2 | 0 | 0 | 0 | **2** |
| MSHS Pathogen Surveillance Program | 1 | 0 | 0 | 0 | 0 | 0 | **1** |
| **All** | **7149** | **3321** | **16797** | **1785** | **3293** | **1046** | **33391** |

*Note: 7 RI and 10 CT sequences were missing metadata on GISAID as of 10/4/2021*
