## Supplementary material for "Comparative transmissibility of SARS-CoV-2 variants Delta and Alpha in New England, USA": DataS1-7: DataS1_Connecticut.pdf

We gratefully acknowledge the following Authors from the Originating laboratories responsible for obtaining the specimens, as well as the Submitting laboratories where the genome data were generated and shared via GISAID, on which this research is based.

All Submitters of data may be contacted directly via www.gisaid.org

Authors are sorted alphabetically.

| Accession ID | Originating Laboratory | Submitting Laboratory | Authors |
|---|---|---|---|
| EPI_ISL_1445343, EPI_ISL_1445498, EPI_ISL_1445634, EPI_ISL_1445944, EPI_ISL_1446005, EPI_ISL_1446150, EPI_ISL_1479558, EPI_ISL_1479578, EPI_ISL_1479818, EPI_ISL_1479863, EPI_ISL_1491861, EPI_ISL_1513212, EPI_ISL_1513213, EPI_ISL_1513235, EPI_ISL_1513565, EPI_ISL_1513568, EPI_ISL_1525106, EPI_ISL_1550457, EPI_ISL_1550530, EPI_ISL_1550577, EPI_ISL_1550683, EPI_ISL_1550734, EPI_ISL_1550756, EPI_ISL_1550759, EPI_ISL_1551106, EPI_ISL_1551108, EPI_ISL_1551200, EPI_ISL_1551233, EPI_ISL_1551248, EPI_ISL_1551590, EPI_ISL_1560083, EPI_ISL_1560109, EPI_ISL_1560110, EPI_ISL_1560898, EPI_ISL_1561465, EPI_ISL_1561466, EPI_ISL_1562757, EPI_ISL_1563133, EPI_ISL_1563134, EPI_ISL_1563285, EPI_ISL_1648973, EPI_ISL_1648993, EPI_ISL_1648994, EPI_ISL_1648996, EPI_ISL_1649003, EPI_ISL_1649168, EPI_ISL_1649363, EPI_ISL_1649725, EPI_ISL_1650261, EPI_ISL_1650688, EPI_ISL_1667481, EPI_ISL_1667505, EPI_ISL_1667600, EPI_ISL_1668126, EPI_ISL_1668350, EPI_ISL_1685564, EPI_ISL_1685565, EPI_ISL_1685567, EPI_ISL_1685795, EPI_ISL_1686416, EPI_ISL_1686747, EPI_ISL_1686819, EPI_ISL_1687194, EPI_ISL_1687198, EPI_ISL_1687201, EPI_ISL_1687233, EPI_ISL_1687234, EPI_ISL_1687236, EPI_ISL_1687377, EPI_ISL_1687537, EPI_ISL_1687553, EPI_ISL_1687577, EPI_ISL_1687699, EPI_ISL_1687961, EPI_ISL_1688345, EPI_ISL_1688777, EPI_ISL_1688779, EPI_ISL_1689886, EPI_ISL_1689892, EPI_ISL_1690049, EPI_ISL_1695466, EPI_ISL_1701916, EPI_ISL_1701992, EPI_ISL_1702783, EPI_ISL_1702842, EPI_ISL_1736902, EPI_ISL_1737179, EPI_ISL_1737368, EPI_ISL_1737462, EPI_ISL_1834933, EPI_ISL_1835007, EPI_ISL_1835047, EPI_ISL_1835226, EPI_ISL_1835244, EPI_ISL_1835421, EPI_ISL_1835574, EPI_ISL_1835575, EPI_ISL_1835576, EPI_ISL_1835690, EPI_ISL_1835716, EPI_ISL_1835719, EPI_ISL_1835720, EPI_ISL_1835736, EPI_ISL_1835737, EPI_ISL_1835833, EPI_ISL_1836050, EPI_ISL_1836065, EPI_ISL_1836191, EPI_ISL_1836259, EPI_ISL_1836720, EPI_ISL_1836760, EPI_ISL_1837204, EPI_ISL_1837278, EPI_ISL_1837710, EPI_ISL_1838011, EPI_ISL_1900340, EPI_ISL_1925182, EPI_ISL_1925184, EPI_ISL_1925185, EPI_ISL_1925220, EPI_ISL_1925475, EPI_ISL_1925490, EPI_ISL_1925549, EPI_ISL_1995768, EPI_ISL_1996474, EPI_ISL_1996575, EPI_ISL_1996583, EPI_ISL_1996587, EPI_ISL_1996592, EPI_ISL_1996593, EPI_ISL_1996595, EPI_ISL_1996599, EPI_ISL_1997546, EPI_ISL_1997553, EPI_ISL_1997569, EPI_ISL_1997577, EPI_ISL_1997581, EPI_ISL_1997682, EPI_ISL_1997750, EPI_ISL_1997771, EPI_ISL_1998334, EPI_ISL_1998343, EPI_ISL_1998344, EPI_ISL_1998348, EPI_ISL_1998349, EPI_ISL_1998406, EPI_ISL_1998464, EPI_ISL_1998583, EPI_ISL_1998584, EPI_ISL_1998676, EPI_ISL_1998764, EPI_ISL_1998794, EPI_ISL_1998822, EPI_ISL_1999444, EPI_ISL_1999574, EPI_ISL_1999583, EPI_ISL_2000243, EPI_ISL_2000469, EPI_ISL_2000490, EPI_ISL_2035153, EPI_ISL_2039316, EPI_ISL_2039555, EPI_ISL_2039618, EPI_ISL_2039677, EPI_ISL_2039790, EPI_ISL_2039972, EPI_ISL_2039973, EPI_ISL_2040021, EPI_ISL_2040643, EPI_ISL_2040904, EPI_ISL_2041276, EPI_ISL_2041743, EPI_ISL_2042217, EPI_ISL_2043215, EPI_ISL_2043348, EPI_ISL_2043479, EPI_ISL_2043510, EPI_ISL_2089598, EPI_ISL_2089751, EPI_ISL_2089753, EPI_ISL_2089755, EPI_ISL_2089988, EPI_ISL_2145994, EPI_ISL_2146969, EPI_ISL_2148741, EPI_ISL_2149467, EPI_ISL_2150180, EPI_ISL_2150957, EPI_ISL_2150964, EPI_ISL_2158959, EPI_ISL_2159411, EPI_ISL_2185761, EPI_ISL_2185912, EPI_ISL_2185919, EPI_ISL_2185954, EPI_ISL_2186191, EPI_ISL_2186219, EPI_ISL_2186220, EPI_ISL_2202649, EPI_ISL_2241977, EPI_ISL_2242476, EPI_ISL_2242672, EPI_ISL_2242751, EPI_ISL_2242812, EPI_ISL_2242907, EPI_ISL_2242924, EPI_ISL_2242982, EPI_ISL_2243078, EPI_ISL_2243091, EPI_ISL_2243235, EPI_ISL_2243236, EPI_ISL_2243255, EPI_ISL_2243301, EPI_ISL_2243542, EPI_ISL_2243624, EPI_ISL_2243880, EPI_ISL_2243884, EPI_ISL_2243990, EPI_ISL_2244162, EPI_ISL_2244196, EPI_ISL_2244274, EPI_ISL_2244322, EPI_ISL_2244323, EPI_ISL_2244328, EPI_ISL_2244389, EPI_ISL_2244396, EPI_ISL_2244490, EPI_ISL_2244743, EPI_ISL_2280315, EPI_ISL_2280317, EPI_ISL_2280664, EPI_ISL_2280667, EPI_ISL_2283846, EPI_ISL_2321261, EPI_ISL_2370286, EPI_ISL_2372378, EPI_ISL_2421854, EPI_ISL_2422002, EPI_ISL_2482337, EPI_ISL_2489813, EPI_ISL_2600087, EPI_ISL_2784893, EPI_ISL_2785260, EPI_ISL_2877684, EPI_ISL_3220460, EPI_ISL_3321823, EPI_ISL_3323088, EPI_ISL_3323814, EPI_ISL_3323868, EPI_ISL_3323904, EPI_ISL_3323909, EPI_ISL_3324307, EPI_ISL_3324417, EPI_ISL_3324747, EPI_ISL_3325020, EPI_ISL_3325059, EPI_ISL_3325061, EPI_ISL_3325064, EPI_ISL_3325066, EPI_ISL_3325228, EPI_ISL_3325247 | | | |
| see above | Aegis Sciences Corporation | Centers for Disease Control and Prevention Division of Viral Diseases, Pathogen Discovery | Adrian Paskey; Alec Vest; Benjamin Rambo-Martin; Christopher Gulvick; Clinton R. Paden; Cyndi Clark; Dakota Howard; Darlene Wagner; Dhwani Batra; Dillon Nall; Duncan MacCannell; Ethan Sanders; Holly Houdeshell; Jason Caravas; Kara Moser; Matthew Hardison; Matthew Schmerer; Ola Kvalvaag; Patrick Campbell; Peter W. Cook; Rob Case; Scott Sammons; Shatavia Morrison; Shaun Westlund; Vikramsinha Ghorpade; Yvette Unoarumhi |
| EPI_ISL_2991501 | Broad Institute Clinical Research Sequencing Platform | Infectious Disease Program, Broad Institute of Harvard and MIT | A.E.; Adams, G.; Anahtar, M.; B.L.; B.W.; Bauer, M.; Birren; Branda, J.; Carter, A.; Cerrato, F.; Chaluvadi, S.; Chapman; Cusick, C.; D.J.; DeRuff, K.; Flowers, K.; Gallagher, G.; Gladden-Young, A.; Gnirke, A.; Harris, J.; J.E.; K.J.; LaRocque, R.; Lagerborg, K.; Lemieux; Lin; Loreth, C.; MacInnis; Neumann, A.; Normandin, E.; P.C.; Park; Pierce, V.; Reilly, S.; Rosenberg, E.; Rudy, M.; Ryan, E.; S.B.; Sabeti; Shaw, B.; Siddle; Slater, D.; Smole, S.; Tomkins-Tinch, C.; Turbett, S. |
| EPI_ISL_1413154, EPI_ISL_1413209, EPI_ISL_1413212, EPI_ISL_1413218, EPI_ISL_1413221, EPI_ISL_1413251, EPI_ISL_1413254, EPI_ISL_1413316, EPI_ISL_1413328, EPI_ISL_1413330, EPI_ISL_1413341, EPI_ISL_1413344, EPI_ISL_1413352, EPI_ISL_1413355, EPI_ISL_1413358, EPI_ISL_1413361, EPI_ISL_1413363, EPI_ISL_1516474, EPI_ISL_1516499, EPI_ISL_1516500, EPI_ISL_1516501, EPI_ISL_1516523, EPI_ISL_1516529, EPI_ISL_1516535, EPI_ISL_1516554, EPI_ISL_1516564, EPI_ISL_1516568, EPI_ISL_1516570, EPI_ISL_1516572, EPI_ISL_1516575, EPI_ISL_1516577, EPI_ISL_1516585, EPI_ISL_1516590, EPI_ISL_1578211, EPI_ISL_1578223, EPI_ISL_1578226, EPI_ISL_1578285, EPI_ISL_1578380, EPI_ISL_1578382, EPI_ISL_1578399, EPI_ISL_1578403, EPI_ISL_1578404, EPI_ISL_1578435, EPI_ISL_1578439, EPI_ISL_1710049, EPI_ISL_1710051, EPI_ISL_1710057, EPI_ISL_1710061, EPI_ISL_1710065, EPI_ISL_1710087, EPI_ISL_1710092, EPI_ISL_1710093, EPI_ISL_1710097, EPI_ISL_1710107, EPI_ISL_1710130, EPI_ISL_1710134, EPI_ISL_1710135, EPI_ISL_1743718, EPI_ISL_1757047, EPI_ISL_1757048, EPI_ISL_1757049, EPI_ISL_1757050, EPI_ISL_1757051, EPI_ISL_1757054, EPI_ISL_1757075, EPI_ISL_1757268, EPI_ISL_1757269, EPI_ISL_1757270, EPI_ISL_1757271, EPI_ISL_1757272, EPI_ISL_1757274, EPI_ISL_1757277, EPI_ISL_1786720, EPI_ISL_1786721, EPI_ISL_1786722, EPI_ISL_1786723, EPI_ISL_1786724, EPI_ISL_1786725, EPI_ISL_1786726, EPI_ISL_1786727, EPI_ISL_1786728, EPI_ISL_1786729, EPI_ISL_1786730, EPI_ISL_1786731, EPI_ISL_1786732, EPI_ISL_1825141, EPI_ISL_1825142, EPI_ISL_1825143, EPI_ISL_1825214, EPI_ISL_1825216, EPI_ISL_1825218, EPI_ISL_1825219, EPI_ISL_1825220, EPI_ISL_1825222, EPI_ISL_1825223, EPI_ISL_1825225, EPI_ISL_1825228, EPI_ISL_1825242, EPI_ISL_1825248, EPI_ISL_1825249, EPI_ISL_1825250, EPI_ISL_1825254, EPI_ISL_1825372, EPI_ISL_1825373, EPI_ISL_1825518, EPI_ISL_1918405, EPI_ISL_1918406, EPI_ISL_1918407, EPI_ISL_1918409, EPI_ISL_1918410, EPI_ISL_1918996, EPI_ISL_1918997, EPI_ISL_1918998, EPI_ISL_1919000, EPI_ISL_1919001, EPI_ISL_1919003, EPI_ISL_1919004, EPI_ISL_1919005, EPI_ISL_1919006, EPI_ISL_1919007, EPI_ISL_1919008, EPI_ISL_1919009, EPI_ISL_1919010, EPI_ISL_1967942, EPI_ISL_1967943, EPI_ISL_1967944, EPI_ISL_1967945, EPI_ISL_1967947, EPI_ISL_1967948, EPI_ISL_1967950, EPI_ISL_1967951, EPI_ISL_1967952, EPI_ISL_1967953, EPI_ISL_1967955, EPI_ISL_1967956, EPI_ISL_1971507, EPI_ISL_1971508, EPI_ISL_1971509, EPI_ISL_1971510, EPI_ISL_1971511, EPI_ISL_1971512, EPI_ISL_1971513, EPI_ISL_1971514, EPI_ISL_1971515, EPI_ISL_1971516, EPI_ISL_1971517, EPI_ISL_1971518, EPI_ISL_1971519, EPI_ISL_1971520, EPI_ISL_1971521, EPI_ISL_1971522, EPI_ISL_1971523, EPI_ISL_1971524, EPI_ISL_1971525, EPI_ISL_1971526, EPI_ISL_1971527, EPI_ISL_1971528, EPI_ISL_1971529, EPI_ISL_1971530, EPI_ISL_1971531, EPI_ISL_1971533, EPI_ISL_1971534, EPI_ISL_1971535, EPI_ISL_1971536, EPI_ISL_2096217, EPI_ISL_2096218, EPI_ISL_2096219, EPI_ISL_2096220, EPI_ISL_2096221, EPI_ISL_2096222, EPI_ISL_2096223, EPI_ISL_2096224, EPI_ISL_2096225, EPI_ISL_2096226, EPI_ISL_2096227, EPI_ISL_2103825, EPI_ISL_2103826, EPI_ISL_2103827, EPI_ISL_2103828, EPI_ISL_2103829, EPI_ISL_2103830, EPI_ISL_2103831, EPI_ISL_2103832, EPI_ISL_2103833, EPI_ISL_2103834, EPI_ISL_2192589, EPI_ISL_2192590, EPI_ISL_2192591, EPI_ISL_2192592, EPI_ISL_2192593, EPI_ISL_2192594, EPI_ISL_2311863, EPI_ISL_2311864, EPI_ISL_2311865, EPI_ISL_2311866, EPI_ISL_2311867, EPI_ISL_3007307, EPI_ISL_3084597, EPI_ISL_3127201, EPI_ISL_3134282, EPI_ISL_3134283, EPI_ISL_3134284, EPI_ISL_3152938, EPI_ISL_3152939, EPI_ISL_3152940, EPI_ISL_3155729, EPI_ISL_3155730, EPI_ISL_3155731 | | | |
| see above | Broad Institute Clinical Research Sequencing Platform | Infectious Disease Program, Broad Institute of Harvard and MIT | Adams, G.; B.L.; B.W.; Bauer, M.; Birren; Blumenstiel, B.; Brown, C.; Carter, A.; Chaluvadi, S.; D.J.; DeFelice, M.; DeRuff, K.; Dodge, S.; Gabriel, S.; Gallagher, G.; Gladden-Young, A.; Granger, B.; J.E.; K.J.; Lagerborg, K.; Larkin, K.; Lee, M.; Lemieux; Lennon, N.; Loreth, C.; Madoff, L.; McGovern, S.; Meldrim, J.; Normandin, E.; P.C.; Park; Pearlman, L.; Reilly, S.; Rudy, M.; Sabeti; Siddle; Smole, S.; Tomkins-Tinch, C.; Vicente, G.; and MacInnis |
| EPI_ISL_2036747, EPI_ISL_2036748, EPI_ISL_2036749, EPI_ISL_2036750, EPI_ISL_2036751, EPI_ISL_2036752, EPI_ISL_2036753, EPI_ISL_2036754, EPI_ISL_2036755, EPI_ISL_2036756, EPI_ISL_2036757, EPI_ISL_2036758, EPI_ISL_2036761, EPI_ISL_2228088, EPI_ISL_2228089, EPI_ISL_2228090, EPI_ISL_2500422, EPI_ISL_2500423, EPI_ISL_2500424, EPI_ISL_2500425, EPI_ISL_2500426, EPI_ISL_2500427, EPI_ISL_2500428, EPI_ISL_2500429, EPI_ISL_2500431, EPI_ISL_2500432, EPI_ISL_2500433, EPI_ISL_2500435, EPI_ISL_2500436, EPI_ISL_2501533, EPI_ISL_2501534, EPI_ISL_2501535, EPI_ISL_2501536, EPI_ISL_2501537, EPI_ISL_2501538, EPI_ISL_2501539, EPI_ISL_2501540, EPI_ISL_2501541, EPI_ISL_2626591, EPI_ISL_2626592, EPI_ISL_2626593, EPI_ISL_2710379, EPI_ISL_2710381, EPI_ISL_2710382, EPI_ISL_3023765, EPI_ISL_3023766, EPI_ISL_3023767, EPI_ISL_3023768, EPI_ISL_3023769, EPI_ISL_3023770, EPI_ISL_3023771, EPI_ISL_3023772, EPI_ISL_3023773, EPI_ISL_3023774, EPI_ISL_3023777, EPI_ISL_3023779, EPI_ISL_3023780, EPI_ISL_3023781, EPI_ISL_3023782, EPI_ISL_3023783, EPI_ISL_3023784, EPI_ISL_3023785, EPI_ISL_3023786, EPI_ISL_3023787, EPI_ISL_3023788, EPI_ISL_3023789, EPI_ISL_3056762, EPI_ISL_3061867, EPI_ISL_3061868, EPI_ISL_3061869, EPI_ISL_3061870, EPI_ISL_3061871, EPI_ISL_3151561, EPI_ISL_3151562, EPI_ISL_3151563, EPI_ISL_3151564, EPI_ISL_3151565, EPI_ISL_3151566, EPI_ISL_3151567, EPI_ISL_3151568, EPI_ISL_3151569, EPI_ISL_3151570, EPI_ISL_3151571, EPI_ISL_3151572, EPI_ISL_3151573, EPI_ISL_3151574, EPI_ISL_3151575, EPI_ISL_3151576, EPI_ISL_3151577, EPI_ISL_3151578, EPI_ISL_3151579, EPI_ISL_3151580, EPI_ISL_3151581, EPI_ISL_3151582, EPI_ISL_3151583, EPI_ISL_3151584, EPI_ISL_3151585, EPI_ISL_3151586, EPI_ISL_3151587, EPI_ISL_3151588, EPI_ISL_3151589, EPI_ISL_3151590, EPI_ISL_3151591, EPI_ISL_3151592, EPI_ISL_3151593, EPI_ISL_3151594, EPI_ISL_3151595, EPI_ISL_3151596, EPI_ISL_3151597, EPI_ISL_3151598, EPI_ISL_3151599, EPI_ISL_3151600, EPI_ISL_3320650, EPI_ISL_3320651, EPI_ISL_3320652, EPI_ISL_3320653, EPI_ISL_3320654, EPI_ISL_3320655, EPI_ISL_3320656, EPI_ISL_3320657, EPI_ISL_3320658, EPI_ISL_3320659, EPI_ISL_3320660, EPI_ISL_3320661, EPI_ISL_3320662, EPI_ISL_3320663, EPI_ISL_3320664, EPI_ISL_3320665, EPI_ISL_3320666, EPI_ISL_3320667, EPI_ISL_3320668, EPI_ISL_3320669, EPI_ISL_3320670, EPI_ISL_3320671, EPI_ISL_3320672, EPI_ISL_3320673, EPI_ISL_3320674, EPI_ISL_3320675, EPI_ISL_3320676, EPI_ISL_3320677, EPI_ISL_3320678, EPI_ISL_3320679, EPI_ISL_3320680, EPI_ISL_3320681, EPI_ISL_3320682, EPI_ISL_3320683, EPI_ISL_3320684, EPI_ISL_3320685, EPI_ISL_3320686, EPI_ISL_3320687, EPI_ISL_3320688, EPI_ISL_3320689, EPI_ISL_3320690, EPI_ISL_3320691, EPI_ISL_3320692, EPI_ISL_3320693, EPI_ISL_3320694, EPI_ISL_3320695, EPI_ISL_3320696, EPI_ISL_3320697, EPI_ISL_3320698, EPI_ISL_3320699, EPI_ISL_3320700, EPI_ISL_3320701, EPI_ISL_3320702, EPI_ISL_3320703, EPI_ISL_3320704, EPI_ISL_3320705, EPI_ISL_3320706, EPI_ISL_3320707, EPI_ISL_3320708, EPI_ISL_3320709, EPI_ISL_3320710, EPI_ISL_3320711, EPI_ISL_3320712, EPI_ISL_3320713, EPI_ISL_3320714, EPI_ISL_3320715, EPI_ISL_3320716 | | | |
| see above | CT Department of Public Health | CT Department of Public Health | Claire Pearson; Claire_Pearson; Tu N. Nguyen; Tu_N_Nguyen |
| EPI_ISL_3347367, EPI_ISL_3347368, EPI_ISL_3347369, EPI_ISL_3347370, EPI_ISL_3347371, EPI_ISL_3347372, EPI_ISL_3347373, EPI_ISL_3347374, EPI_ISL_3347375, EPI_ISL_3347376, EPI_ISL_3347377, EPI_ISL_3347378, EPI_ISL_3347379, EPI_ISL_3347380 | | | |
| see above | CT Department of Public Health | Grubaugh Lab - Yale School of Public Health | Anderson Brito; Annie Watkins; Chaney Kalinich; Chantal Vogels; Claire Pearson; Isabel Ott; Jessica Rothman; Joseph Fauver; Kendall Billig; Mallery Breban; Mary Petrone; Nathan Grubaugh; Tara Alpert; Tobias Koch; Tu N. Nguyen |
| EPI_ISL_1272720, EPI_ISL_1272721, EPI_ISL_1272722, EPI_ISL_1272723, EPI_ISL_1272724, EPI_ISL_1272725, EPI_ISL_1272726, EPI_ISL_1318465, EPI_ISL_1318466, EPI_ISL_1318467, EPI_ISL_1446620, EPI_ISL_1711711, EPI_ISL_1711712, EPI_ISL_1711713, EPI_ISL_1711716, EPI_ISL_1798336, EPI_ISL_1798382, EPI_ISL_1798388, EPI_ISL_1798396, EPI_ISL_1798400, EPI_ISL_1798417, EPI_ISL_1798466, EPI_ISL_1798468, EPI_ISL_2105160, EPI_ISL_2105171, EPI_ISL_2105204, EPI_ISL_2105205, EPI_ISL_2105213, EPI_ISL_2229112, EPI_ISL_2229113, EPI_ISL_2229114, EPI_ISL_2229115, EPI_ISL_2229116, EPI_ISL_2383894, EPI_ISL_2383895, EPI_ISL_2384064, EPI_ISL_2384065, EPI_ISL_2384066, EPI_ISL_2384067, EPI_ISL_2384068, EPI_ISL_2384069, EPI_ISL_2451739, EPI_ISL_2451741, EPI_ISL_2451743, EPI_ISL_2451744, EPI_ISL_2451746, EPI_ISL_2451748, EPI_ISL_2645335, EPI_ISL_2787514, EPI_ISL_2840429, EPI_ISL_3128000, EPI_ISL_3128001, EPI_ISL_3128002, EPI_ISL_3241919, EPI_ISL_3242017, EPI_ISL_3242018, EPI_ISL_3242019, EPI_ISL_3242020, EPI_ISL_3305769, EPI_ISL_3354023, EPI_ISL_3354024, EPI_ISL_3354028, EPI_ISL_3354170, EPI_ISL_3354234, EPI_ISL_3354242, EPI_ISL_3354491, EPI_ISL_3354494, EPI_ISL_3354496, EPI_ISL_3354509, EPI_ISL_3355119, EPI_ISL_3355123 | | | |
| see above | CT-Dr. Katherine A. Kelley State Public Health Lab | Centers for Disease Control and Prevention Division of Viral Diseases, Pathogen Discovery | Alex Burgin; Alison Laufer Halpin; Anna Montmayeur; Anna Uehara; Ben L. Rambo-Martin; Ben Rambo-Martin; Clinton Paden; Clinton R. Paden; Dakota Howard; Darlene Wagner; Dave Wentworth; Dhwani Batra; Haibin Wang; Jasmine Padilla; Jing Zhang; Justin Lee; Katie Dillon; Krista Queen; Kristen Knipe; Kristine Lacek; Lori Rowe; Mark Burroughs; Matthew Schmerer; Meghan Bentz; Mili Sheth; Peter Cook; Peter W. Cook; Rachel Marine; Sam Shepard; Sarah Nobles; Shoshona Le; Suxiang Tong; Vivien Dugan; Yan Li; Ying Tao; Yvette Unoarumhi |
| EPI_ISL_1226212, EPI_ISL_1226213 | CT-Dr. Katherine A. Kelley State Public Health Lab | Genomics and Discovery, Respiratory Viruses Branch, Division of Viral Diseases, Centers for Disease Control and Prevention | Anna Montmayeur; Anna Uehara; Ben L. Rambo-Martin; Clinton R. Paden; Dhwani Batra; Haibin Wang; Jasmine Padilla; Jing Zhang; Justin Lee; Katie Dillon; Krista Queen; Kristen Knipe; Kristine Lacek; Lori Rowe; Mark Burroughs; Matthew Schmerer; Mili Sheth; Peter W. Cook; Rachel Marine; Sam Shepard; Sarah Nobles; Shoshona Le; Suxiang Tong; Yan Li; Ying Tao |
| EPI_ISL_1182027, EPI_ISL_1182028, EPI_ISL_1182029, EPI_ISL_1182030, EPI_ISL_1225640, EPI_ISL_1225654, EPI_ISL_1225661, EPI_ISL_1225722, EPI_ISL_1225728, EPI_ISL_1292971, EPI_ISL_1292972, EPI_ISL_1292973, EPI_ISL_1292974, EPI_ISL_1292975, EPI_ISL_1292976, EPI_ISL_1292977, EPI_ISL_1292978, EPI_ISL_1293140 | | | |
| see above | Connecticut DPH | Grubaugh Lab - Yale School of Public Health | Anderson Brito; Annie Watkins; Anthony Muyombwe; Chaney Kalinich; Chantal Vogels; Isabell Ott; Jafar Razeq; Jessica Rothman; Joseph Fauver; Mallery Breban; Mary Petrone; Nathan Grubaugh; Randy Downing; Stephen M. Bart; Tara Alpert |
| EPI_ISL_2034694, EPI_ISL_2034695, EPI_ISL_2034696, EPI_ISL_2034697, EPI_ISL_2034698, EPI_ISL_2034699, EPI_ISL_2034700, EPI_ISL_2034701, EPI_ISL_2034702, EPI_ISL_2034703, EPI_ISL_2034704, EPI_ISL_2034705, EPI_ISL_2034706, EPI_ISL_2034707, EPI_ISL_2034708, EPI_ISL_2034710, EPI_ISL_2034711, EPI_ISL_2034713, EPI_ISL_2034715, EPI_ISL_2034717, EPI_ISL_2034719, EPI_ISL_2034721, EPI_ISL_2034723, EPI_ISL_2034725, EPI_ISL_2034726, EPI_ISL_2034728, EPI_ISL_2034730, EPI_ISL_2034731, EPI_ISL_2034733, EPI_ISL_2034734, EPI_ISL_2034735, EPI_ISL_2034737, EPI_ISL_2034738, EPI_ISL_2034741, EPI_ISL_2034743, EPI_ISL_2034745, EPI_ISL_2034747, EPI_ISL_2034748, EPI_ISL_2034750, EPI_ISL_2034752, EPI_ISL_2034753, EPI_ISL_3105429, EPI_ISL_3105466, EPI_ISL_3105469, EPI_ISL_3105471, EPI_ISL_3105507, EPI_ISL_3105508 | | | |
| see above | Connecticut DPH | Yale Center for Genomic Analysis | Anderson Brito; Annie Watkins; Anthony Muyombwe; Brooke Sullivan; Chaney Kalinich; Chantal Vogels; Curt Scharfe; Irina Tikhonova; Isabel Ott; Jafar Razeq; Jessica Rothman; Joseph Fauver; Kaya Bilguvar; Mallery Breban; Mary Petrone; Nathan Grubaugh; Randy Downing; Shrikant Mane; Stephen M. Bart; Tara Alpert |
| EPI_ISL_912163, EPI_ISL_912164, EPI_ISL_912185, EPI_ISL_1168737 | Connecticut Department of Health | Grubaugh Lab - Yale School of Public Health | Anderson Brito; Anne Wyllie; Annie Watkins; Chaney Kalinich; Chantal Vogels; Isabel Ott; Joseph Fauver; Mallery Breban; Mary Petrone; Nathan Grubaugh; Tara Alpert |
| EPI_ISL_1091805, EPI_ISL_1091808, EPI_ISL_1091809, EPI_ISL_1091821, EPI_ISL_1091822, EPI_ISL_1091823, EPI_ISL_1091828, EPI_ISL_1091829, EPI_ISL_1091835, EPI_ISL_1091838, EPI_ISL_1091840, EPI_ISL_1091841, EPI_ISL_1091844, EPI_ISL_1091845, EPI_ISL_1091846, EPI_ISL_1091847, EPI_ISL_1091848, EPI_ISL_1091854, EPI_ISL_1091855, EPI_ISL_1091856, EPI_ISL_1091857, EPI_ISL_1091865, EPI_ISL_1112933, EPI_ISL_1112940, EPI_ISL_1112941, EPI_ISL_1112942, EPI_ISL_1112943, EPI_ISL_1112945, EPI_ISL_1239849 | | | |
| see above | Connecticut Department of Public Health | Grubaugh Lab - Yale School of Public Health | Anderson Brito; Anne Wyllie; Annie Watkins; Chaney Kalinich; Chantal Vogels; Isabel Ott; Joseph Fauver; Mallery Breban; Mary Petrone; Nathan Grubaugh; Tara Alpert |
| EPI_ISL_2112524, EPI_ISL_2112525, EPI_ISL_2112527 | Epidemiology of Microbial Diseases, Yale School of Public Health | Epidemiology of Microbial Diseases, Yale School of Public Health | A.E.; A.F.; Alpert, T.; Breban; Brito; C.B.; C.C.; Fauver; Grubaugh; I.M.; Iwasaki, A.; J.E.; J.R.; Kalinich; Landry; Lucas, C.; M.E.; M.I.; M.L.; N.D.; Ott; Petrone; Rothman; Vogels; Watkins |
| EPI_ISL_1038997 | Fair Haven CHC | Grubaugh Lab - Yale School of Public Health | Anderson Brito; Anne Wyllie; Annie Watkins; Chaney Kalinich; Chantal Vogels; Isabel Ott; Joseph Fauver; Mallery Breban; Mary Petrone; Nathan Grubaugh; Tara Alpert |
| EPI_ISL_2280241, EPI_ISL_2785871, EPI_ISL_3305600 | Fulgent Genetics | Centers for Disease Control and Prevention Division of Viral Diseases, Pathogen | Adrian Paskey; Becky Tsai; Benafsh Sapra; Benjamin Rambo-Martin; Christopher Gulvick; Clinton R. Paden; Dakota Howard; Darlene Wagner; Dhwani Batra; Doreen Ng; Duncan MacCannell; Harry Gao; James Xie; Jason Caravas; John Gao; Joseph Fierro; Kara Moser; Matthew Schmerer; Mickey Li; Peter W. Cook; Scott Sammons; Shatavia Morrison; Yan Meng; Yvette Unoarumhi |

| | | | |
|---|---|---|---|
| | | Discovery | |
| EPI_ISL_1168736, EPI_ISL_1225677, EPI_ISL_1293217, EPI_ISL_1378726, EPI_ISL_1378753, EPI_ISL_1378830, EPI_ISL_1378831, EPI_ISL_1378832, EPI_ISL_1447673, EPI_ISL_1447674, EPI_ISL_1447675, EPI_ISL_1447676, EPI_ISL_1509270, EPI_ISL_1509271, EPI_ISL_1587428, EPI_ISL_1674706, EPI_ISL_1674707, EPI_ISL_1674708, EPI_ISL_1674709, EPI_ISL_1793096, EPI_ISL_2159676, EPI_ISL_2598692 | | | |
| see above | Grubaugh Lab - Yale School of Public Health | Grubaugh Lab - Yale School of Public Health | Anderson Brito; Anne Wyllie; Annie Watkins; Chaney Kalinich; Chantal Vogels; Isabel Ott; Isabell Ott; Jessica Rothman; Joseph Fauver; Mallery Breban; Marie L. Landry; Mary Petrone; Nathan Grubaugh; Rebecca Earnest; Tara Alpert |
| EPI_ISL_850712, EPI_ISL_850893 | Helix / Illumina | Genomics and Discovery, Respiratory Viruses Branch, Division of Viral Diseases, Centers for Disease Control and Prevention | Alexandre Bolze; Ary Ascencio; Ben L. Rambo-Martin Eileen de Feo; Brad Sickler; Charlotte Rivera-Garcia; Christine Tran; Clinton R. Paden; David Becker; Dhwani Batra; Duncan MacCannell; Efren Sandoval; Elizabeth Cirulli; Eric Allen; Geraint Levan; James Lu; Jan Antico; Jason Nguyen; Jimmy Ramirez; Jingtao Liu; Kelly Schiabor Barrett; Kim Gietzen; Magnus Isaksson; Marc Laurent; Matthew Tolentino; Nicole L. Washington; Peter W. Cook; Phil Febbo; Ryan Cho; Shannon Wickline; Sherry Wang; Simon White; Summer Galloway; Suxiang Tong; Tyler Cassens; William Lee |
| EPI_ISL_1088709, EPI_ISL_1089040, EPI_ISL_1089371 | Helix / Illumina | Respiratory Viruses Branch, Division of Viral Diseases, Centers for Disease Control and Prevention | Alexandre Bolze; Ary Ascencio; Ben L. Rambo-Martin; Brad Sickler; Charlotte Rivera-Garcia; Christine Tran; Clinton R. Paden; Dakota Howard; David Becker; Dhwani Batra; Duncan MacCannell; Efren Sandoval; Eileen de Feo; Elizabeth Cirulli; Eric Allen; Geraint Levan; James Lu; Jan Antico; Jason Nguyen; Jimmy Ramirez; Jingtao Liu; Kelly Schiabor Barrett; Kim Gietzen; Magnus Isaksson; Marc Laurent; Matthew Tolentino; Nicole L. Washington; Peter W. Cook; Phil Febbo; Ryan Cho; Shannon Wickline; Sherry Wang; Simon White; Summer Galloway; Suxiang Tong; Tyler Cassens; William Lee |
| EPI_ISL_1267529, EPI_ISL_1338248, EPI_ISL_1340444, EPI_ISL_1444251, EPI_ISL_1460972, EPI_ISL_1480433, EPI_ISL_1511963, EPI_ISL_1554119, EPI_ISL_1554509, EPI_ISL_1581122, EPI_ISL_1592827, EPI_ISL_1593000, EPI_ISL_1679012, EPI_ISL_1679226, EPI_ISL_1679237, EPI_ISL_1679503, EPI_ISL_1679709, EPI_ISL_1701836, EPI_ISL_1734801, EPI_ISL_1734963, EPI_ISL_1735196, EPI_ISL_1735342, EPI_ISL_1796658, EPI_ISL_1797373, EPI_ISL_1797387, EPI_ISL_1803536, EPI_ISL_1907176, EPI_ISL_1907177, EPI_ISL_1907387, EPI_ISL_1907534, EPI_ISL_1907767, EPI_ISL_1907994, EPI_ISL_1923182, EPI_ISL_1923510, EPI_ISL_1923536, EPI_ISL_1924154, EPI_ISL_1942166, EPI_ISL_2097517, EPI_ISL_2098039, EPI_ISL_2159878, EPI_ISL_2247352 | | | |
| see above | Helix/Illumina | Centers for Disease Control and Prevention Division of Viral Diseases, Pathogen Discovery | Adrian Paskey; Alexandre Bolze; Ary Ascencio; Ben L. Rambo-Martin; Benjamin Rambo-Martin; Brad Sickler; Charlotte Rivera-Garcia; Christine Tran; Christopher Gulvick; Clinton R. Paden; Dakota Howard; Darlene Wagner; David Becker; Dhwani Batra; Duncan MacCannell; Efren Sandoval; Eileen de Feo; Elizabeth Cirulli; Eric Allen; Geraint Levan; James Lu; Jan Antico; Jason Caravas; Jason Nguyen; Jimmy Ramirez; Jingtao Liu; Kara Moser; Kelly Schiabor Barrett; Kim Gietzen; Magnus Isaksson; Marc Laurent; Matthew Schmerer; Matthew Tolentino; Nicole L. Washington; Peter W. Cook; Phil Febbo; Ryan Cho; Scott Sammons; Shannon Wickline; Shatavia Morrison; Sherry Wang; Simon White; Summer Galloway; Suxiang Tong; Tyler Cassens; William Lee; Yvette Unoarumhi |
| EPI_ISL_876678, EPI_ISL_876679 | Helix/Illumina | Genomics and Discovery, Respiratory Viruses Branch, Division of Viral Diseases, Centers for Disease Control and Prevention | ; Alexandre Bolze; Ary Ascencio; Ben L. Rambo-Martin; Brad Sickler; Charlotte Rivera-Garcia; Christine Tran; Clinton R. Paden; David Becker; Dhwani Batra; Duncan MacCannell; Efren Sandoval; Eileen de Feo; Elizabeth Cirulli; Eric Allen; Geraint Levan; James Lu; Jan Antico; Jason Nguyen; Jimmy Ramirez; Jingtao Liu; Kelly Schiabor Barrett; Kim Gietzen; Magnus Isaksson; Marc Laurent; Matthew Tolentino; Nicole L. Washington; Peter W. Cook; Phil Febbo; Ryan Cho; Shannon Wickline; Sherry Wang; Simon White; Summer Galloway; Suxiang Tong; Tyler Cassens; William Lee |
| EPI_ISL_966866, EPI_ISL_967080, EPI_ISL_967157, EPI_ISL_967249, EPI_ISL_967328, EPI_ISL_967402, EPI_ISL_967440, EPI_ISL_978782, EPI_ISL_1016581, EPI_ISL_1016681, EPI_ISL_1016688 | | | |
| see above | Helix/Illumina | Respiratory Viruses Branch, Division of Viral Diseases, Centers for Disease Control and Prevention | ; Alexandre Bolze; Ary Ascencio; Ben L. Rambo-Martin; Brad Sickler; Charlotte Rivera-Garcia; Christine Tran; Clinton R. Paden; Dakota Howard; David Becker; Dhwani Batra; Duncan MacCannell; Efren Sandoval; Eileen de Feo; Elizabeth Cirulli; Eric Allen; Geraint Levan; James Lu; Jan Antico; Jason Nguyen; Jimmy Ramirez; Jingtao Liu; Kelly Schiabor Barrett; Kim Gietzen; Magnus Isaksson; Marc Laurent; Matthew Tolentino; Nicole L. Washington; Peter W. Cook; Phil Febbo; Ryan Cho; Shannon Wickline; Sherry Wang; Simon White; Summer Galloway; Suxiang Tong; Tyler Cassens; William Lee |
| EPI_ISL_907064, EPI_ISL_1233921 | Infectious Diseases, Quest Diagnostics | Infectious Diseases, Quest Diagnostics | Anderson, B.; Bernstein; D.F.; Gerasimova, A.; Hua, M.; K.E.; Kagan; L.E.; Lacbawan, F.; Liu, Y.; Livingston; Owen, R.; R.M.; Rosenthal; S.H.; Shalhout |
| EPI_ISL_1512912, EPI_ISL_1512919, EPI_ISL_1513109, EPI_ISL_1692079, EPI_ISL_1692104, EPI_ISL_1692174, EPI_ISL_1692265, EPI_ISL_1692342, EPI_ISL_1693324, EPI_ISL_1795596 | | | |
| see above | Infinity Biologix | Centers for Disease Control and Prevention Division of Viral Diseases, Pathogen Discovery | Adrian Paskey; Benjamin Rambo-Martin; Chirayu Goswami; Christian Bixby; Christopher Gulvick; Clinton R. Paden; Dakota Howard; Darlene Wagner; Dhwani Batra; Duncan MacCannell; Jason Caravas; Jonathan Schultz; Kara Moser; Matthew Schmerer; Peter W. Cook; Robin Grimwood; Russ Hager; Scott Sammons; Shatavia Morrison; Yihe Wang; Yvette Unoarumhi |
| EPI_ISL_886249, EPI_ISL_886252, EPI_ISL_886290, EPI_ISL_886322, EPI_ISL_886346, EPI_ISL_886362, EPI_ISL_886379, EPI_ISL_886399, EPI_ISL_886444, EPI_ISL_886531, EPI_ISL_886543, EPI_ISL_886561, EPI_ISL_886562, EPI_ISL_886566, EPI_ISL_886694, EPI_ISL_886705, EPI_ISL_886748, EPI_ISL_886839, EPI_ISL_886846, EPI_ISL_886878, EPI_ISL_886963, EPI_ISL_886980, EPI_ISL_887079 | | | |
| see above | Labcorp | Genomics and Discovery, Respiratory Viruses Branch, Division of Viral Diseases, Centers for Disease Control and Prevention | ; Amanda Douglas; Amanda Suchanek; Andrea Throop; Ayla Burns; Ben L. Rambo-Martin; Bobbi Croy; Brian Krueger; Brian Norvell; Christos Petropoulos; Clinton R. Paden; Craig Lukasik; Debbie Boles; Dhwani Batra; Duncan MacCannell; Eyad Almasri; Goran Stevovic; Howard Engler; Hrushikesh Deshmukh; Jake Humphrey; Jana Schroth; Joe Voshell; John Pruitt; Jonathan Meltzer; Jonathan Williams; Kimberly Wagner; Lax Iyer; Lyndon Tilson; Manoj Jain; Marcia Eisenberg; Mary Ann Cristobal; Mary Williamson; Michael Levandoski; Mike Sapeta; Mindy Nye; Minoo Agarwal; Mohan Kolli; Nuthawin Charoensri; Oren Cohen; Peter W. Cook; Prashant Gupta; Qian Zeng; Rama Ghatti; Scott Parker; Scott Ryan; Stanley Letovsky; Steven Ragan; Summer Galloway; Suresh Babu Selvaraju; Susan Countryman; Susan Hicks; Suxiang Tong; Suzanne Dale; Thomas Urban; Tim Kuphal; Tricia Zwiefelhofer; Vincent Drouillon |
| EPI_ISL_1193355, EPI_ISL_1193369, EPI_ISL_1220950, EPI_ISL_1220954, EPI_ISL_1220963, EPI_ISL_1220965, EPI_ISL_1221012, EPI_ISL_1221013, EPI_ISL_1221016, EPI_ISL_1221173, EPI_ISL_1255098, EPI_ISL_1291199, EPI_ISL_1291251, EPI_ISL_1291301, EPI_ISL_1298249, EPI_ISL_1298255, EPI_ISL_1319903, EPI_ISL_1319904, EPI_ISL_1320454, EPI_ISL_1320455, EPI_ISL_1320456, EPI_ISL_1338858, EPI_ISL_1339218, EPI_ISL_1339530, EPI_ISL_1481403, EPI_ISL_1514217, EPI_ISL_1514218, EPI_ISL_1514219, EPI_ISL_1514581, EPI_ISL_1515331, EPI_ISL_1548189, EPI_ISL_1549012, EPI_ISL_1549013, EPI_ISL_1549656, EPI_ISL_1609503, EPI_ISL_1609869, EPI_ISL_1609956, EPI_ISL_1611115, EPI_ISL_1611550, EPI_ISL_1680470, EPI_ISL_1680685, EPI_ISL_1680686, EPI_ISL_1680687, EPI_ISL_1680689, EPI_ISL_1680690, EPI_ISL_1680694, EPI_ISL_1680695, EPI_ISL_1680696, EPI_ISL_1680697, EPI_ISL_1680698, EPI_ISL_1680699, EPI_ISL_1680700, EPI_ISL_1680701, EPI_ISL_1680772, EPI_ISL_1680773, EPI_ISL_1680774, EPI_ISL_1680775, EPI_ISL_1680776, EPI_ISL_1680777, EPI_ISL_1680778, EPI_ISL_1680779, EPI_ISL_1680781, EPI_ISL_1680782, EPI_ISL_1680783, EPI_ISL_1680784, EPI_ISL_1680789, EPI_ISL_1680790, EPI_ISL_1680791, EPI_ISL_1680792, EPI_ISL_1680797, EPI_ISL_1680821, EPI_ISL_1680850, EPI_ISL_1680865, EPI_ISL_1680868, EPI_ISL_1681199, EPI_ISL_1681200, EPI_ISL_1681201, EPI_ISL_1681203, EPI_ISL_1681204, EPI_ISL_1681205, EPI_ISL_1681208, EPI_ISL_1681224, EPI_ISL_1681225, EPI_ISL_1681255, EPI_ISL_1681256, EPI_ISL_1681257, EPI_ISL_1681258, EPI_ISL_1681259, EPI_ISL_1681260, EPI_ISL_1681261, EPI_ISL_1681264, EPI_ISL_1681265, EPI_ISL_1681266, EPI_ISL_1681267, EPI_ISL_1681269, EPI_ISL_1681273, EPI_ISL_1681274, EPI_ISL_1681276, EPI_ISL_1681298, EPI_ISL_1681332, EPI_ISL_1681333, EPI_ISL_1681334, EPI_ISL_1681335, EPI_ISL_1681337, EPI_ISL_1681339, EPI_ISL_1681340, EPI_ISL_1681341, EPI_ISL_1681342, EPI_ISL_1681343, EPI_ISL_1681344, EPI_ISL_1681345, EPI_ISL_1681346, EPI_ISL_1681347, EPI_ISL_1681349, EPI_ISL_1681351, EPI_ISL_1681358, EPI_ISL_1681406, EPI_ISL_1681421, EPI_ISL_1681422, EPI_ISL_1681425, EPI_ISL_1681426, EPI_ISL_1681427, EPI_ISL_1681428, EPI_ISL_1681429, EPI_ISL_1681430, EPI_ISL_1681440, EPI_ISL_1681454, EPI_ISL_1681455, EPI_ISL_1681456, EPI_ISL_1681457, EPI_ISL_1681458, EPI_ISL_1681459, EPI_ISL_1681460, EPI_ISL_1681473, EPI_ISL_1681475, EPI_ISL_1681476, EPI_ISL_1681506, EPI_ISL_1681520, EPI_ISL_1681564, EPI_ISL_1681565, EPI_ISL_1681567, EPI_ISL_1681569, EPI_ISL_1681602, EPI_ISL_1681658, EPI_ISL_1681904, EPI_ISL_1682377, EPI_ISL_1683108, EPI_ISL_1683110, EPI_ISL_1683113, EPI_ISL_1683114, EPI_ISL_1683345, EPI_ISL_1683412, EPI_ISL_1683640, EPI_ISL_1684280, EPI_ISL_1684306, EPI_ISL_1798731, EPI_ISL_1799981, EPI_ISL_1800504, EPI_ISL_1801297, EPI_ISL_1801298, EPI_ISL_1801299, EPI_ISL_1801300, EPI_ISL_1801301, EPI_ISL_1801335, EPI_ISL_1801336, EPI_ISL_1801338, EPI_ISL_1801339, EPI_ISL_1801341, EPI_ISL_1801342, EPI_ISL_1801343, EPI_ISL_1801344, EPI_ISL_1801345, EPI_ISL_1801347, EPI_ISL_1801368, EPI_ISL_1801369, EPI_ISL_1801395, EPI_ISL_1801396, EPI_ISL_1801421, EPI_ISL_1801424, EPI_ISL_1801425, EPI_ISL_1801430, EPI_ISL_1801431, EPI_ISL_1801433, EPI_ISL_1801435, EPI_ISL_1801436, EPI_ISL_1801438, EPI_ISL_1801439, EPI_ISL_1801529, EPI_ISL_1801530, EPI_ISL_1801531, EPI_ISL_1801532, EPI_ISL_1801534, EPI_ISL_1801537, EPI_ISL_1801540, EPI_ISL_1801542, EPI_ISL_1801547, EPI_ISL_1801548, EPI_ISL_1801584, EPI_ISL_1801585, EPI_ISL_1801586, EPI_ISL_1801587, EPI_ISL_1801588, EPI_ISL_1801589, EPI_ISL_1801590, EPI_ISL_1801591, EPI_ISL_1801594, EPI_ISL_1801598, EPI_ISL_1801599, EPI_ISL_1801607, EPI_ISL_1801608, EPI_ISL_1801613, EPI_ISL_1801614, EPI_ISL_1801615, EPI_ISL_1801616, EPI_ISL_1801617, EPI_ISL_1801618, EPI_ISL_1801619, EPI_ISL_1801620, EPI_ISL_1801621, EPI_ISL_1801622, EPI_ISL_1801623, EPI_ISL_1801628, EPI_ISL_1801714, EPI_ISL_1801716, EPI_ISL_1801719, EPI_ISL_1801720, EPI_ISL_1801723, EPI_ISL_1801725, EPI_ISL_1801726, EPI_ISL_1801727, EPI_ISL_1801761, EPI_ISL_1801843, EPI_ISL_1801844, EPI_ISL_1801845, EPI_ISL_1801847, EPI_ISL_1801851, EPI_ISL_1801879, EPI_ISL_1801880, EPI_ISL_1801884, EPI_ISL_1801885, EPI_ISL_1801886, EPI_ISL_1801888, EPI_ISL_1801889, EPI_ISL_1801890, EPI_ISL_1801891, EPI_ISL_1801896, EPI_ISL_1801936, EPI_ISL_1801937, EPI_ISL_1801938, EPI_ISL_1801939, EPI_ISL_1801940, EPI_ISL_1801941, EPI_ISL_1801943, EPI_ISL_1801992, EPI_ISL_1801993, EPI_ISL_1801995, EPI_ISL_1801997, EPI_ISL_1801998, EPI_ISL_1801999, EPI_ISL_1802000, EPI_ISL_1802118, EPI_ISL_1802302, EPI_ISL_1802303, EPI_ISL_1802308, EPI_ISL_1802311, EPI_ISL_1802313, EPI_ISL_1925840, EPI_ISL_1926408, EPI_ISL_1926409, EPI_ISL_1926450, EPI_ISL_1926453, EPI_ISL_1926455, EPI_ISL_1926457, EPI_ISL_1926458, EPI_ISL_1926461, EPI_ISL_1926462, EPI_ISL_1926463, EPI_ISL_1926464, EPI_ISL_1926465, EPI_ISL_1926466, EPI_ISL_1926467, EPI_ISL_1926468, EPI_ISL_1926469, EPI_ISL_1926470, EPI_ISL_1926473, EPI_ISL_1926474, EPI_ISL_1926598, EPI_ISL_1926600, EPI_ISL_1926601, EPI_ISL_1926602, EPI_ISL_1927844, EPI_ISL_1927874, EPI_ISL_1927880, EPI_ISL_1928694, EPI_ISL_1929344, EPI_ISL_1929403, EPI_ISL_1929406, EPI_ISL_1929421, EPI_ISL_1929466, EPI_ISL_1929923, EPI_ISL_1930068, EPI_ISL_1930097, EPI_ISL_1930165, EPI_ISL_1930181, EPI_ISL_1930187, EPI_ISL_1930270, EPI_ISL_1930278, EPI_ISL_1930298, EPI_ISL_1930301, EPI_ISL_1930304, EPI_ISL_1930307, EPI_ISL_1930324, EPI_ISL_1930365, EPI_ISL_1930368, EPI_ISL_1930374, EPI_ISL_1930377, EPI_ISL_1930379, EPI_ISL_1930383, EPI_ISL_1930458, EPI_ISL_2045502, EPI_ISL_2045507, EPI_ISL_2045509, EPI_ISL_2045510, EPI_ISL_2045511, EPI_ISL_2045512, EPI_ISL_2045515, EPI_ISL_2045517, EPI_ISL_2045518, EPI_ISL_2045557, EPI_ISL_2045577, EPI_ISL_2045578, EPI_ISL_2045580, EPI_ISL_2045582, EPI_ISL_2045583, EPI_ISL_2045590, EPI_ISL_2045733, EPI_ISL_2045810, EPI_ISL_2045825, EPI_ISL_2045828, EPI_ISL_2045830, EPI_ISL_2045831, EPI_ISL_2045832, EPI_ISL_2045863, EPI_ISL_2045866, EPI_ISL_2045872, EPI_ISL_2045873, EPI_ISL_2045923, EPI_ISL_2046552, EPI_ISL_2181710, EPI_ISL_2182413, EPI_ISL_2182416, EPI_ISL_2182419, EPI_ISL_2182426, EPI_ISL_2182429, EPI_ISL_2182622, EPI_ISL_2182628, EPI_ISL_2182631, EPI_ISL_2182642, EPI_ISL_2182805, EPI_ISL_2182828, EPI_ISL_2182831, EPI_ISL_2182842, EPI_ISL_2183064, EPI_ISL_2183077, EPI_ISL_2183085, EPI_ISL_2184243, EPI_ISL_2306643, EPI_ISL_2306648, EPI_ISL_2306758, EPI_ISL_2306859, EPI_ISL_2306860, EPI_ISL_2306914, EPI_ISL_2306915, EPI_ISL_2306917, EPI_ISL_2306927, EPI_ISL_2306973, EPI_ISL_2306974, EPI_ISL_2306982, EPI_ISL_2307004, EPI_ISL_2307007, EPI_ISL_2398988, EPI_ISL_2399020, EPI_ISL_2399042, EPI_ISL_2399043, EPI_ISL_2399048, EPI_ISL_2399107, EPI_ISL_2399109, EPI_ISL_2399291, EPI_ISL_2399350, EPI_ISL_2399351, EPI_ISL_2399353, EPI_ISL_2399354, EPI_ISL_2399355, EPI_ISL_2399356, EPI_ISL_2480483, EPI_ISL_2480484, EPI_ISL_2480509, EPI_ISL_2611752, EPI_ISL_2872162, EPI_ISL_3016831, EPI_ISL_3111155, EPI_ISL_3111861, EPI_ISL_3111862, EPI_ISL_3112229, EPI_ISL_3112333, EPI_ISL_3112495, EPI_ISL_3329472, EPI_ISL_3330478, EPI_ISL_3330480, EPI_ISL_3330482, EPI_ISL_3330492, EPI_ISL_3330499, EPI_ISL_3330501, EPI_ISL_3330510, EPI_ISL_3330567, EPI_ISL_3330603 | | | |
| see above | Laboratory Corporation of America | Centers for Disease Control and Prevention Division of Viral Diseases, Pathogen Discovery | Adrian Paskey; Amanda Douglas; Amanda Suchanek; Andrea Throop; Ayla Burns; Ben L. Rambo-Martin; Benjamin Rambo-Martin; Bobbi Croy; Brian Krueger; Brian Norvell; Christopher Gulvick; Christos Petropoulos; Clinton R. Paden; Craig Lukasik; Dakota Howard; Darlene Wagner; Debbie Boles; Dhwani Batra; Duncan MacCannell; Eyad Almasri; Eyad Almasri Debbie Boles; Goran Stevovic; Howard Engler; Hrushikesh Deshmukh; Jake Humphrey; Jana Schroth; Jason Caravas; Joe Voshell; John Pruitt; Jonathan Meltzer; Jonathan Williams; Kara Moser; Kimberly Wagner; Lax Iyer; Lisa Pfefferle; Lyndon Tilson; Manoj Jain; Marcia Eisenberg; Mary Ann Cristobal; Mary Williamson; Matthew Robinson; Matthew Schmerer; Michael Levandoski; Mike Sapeta; Mindy Nye; Minoo Agarwal; Mohan Kolli; Nuthawin Charoensri; Oren Cohen; Peter W. Cook; Prashant Gupta; Qian Zeng; Rama Ghatti; Scott Parker; Scott Ryan; Scott Sammons; Shatavia Morrison; Stanley Letovsky; Steven Ragan; Suresh Babu Selvaraju; Susan Countryman; Susan Hicks; Suxiang Tong; Suzanne Dale; Thomas Urban; Tim Kuphal; Tricia Zwiefelhofer; Vincent Drouillon; Yvette Unoarumhi |
| EPI_ISL_1020777, EPI_ISL_1020804, EPI_ISL_1020820, EPI_ISL_1020840, EPI_ISL_1020878, EPI_ISL_1021001, EPI_ISL_1021036, EPI_ISL_1021065, EPI_ISL_1021076, EPI_ISL_1021089, EPI_ISL_1021153, EPI_ISL_1021156, EPI_ISL_1021169, EPI_ISL_1021170, EPI_ISL_1021188, EPI_ISL_1021189, EPI_ISL_1021202, EPI_ISL_1021230, EPI_ISL_1021231, EPI_ISL_1021232, EPI_ISL_1021258, EPI_ISL_1021259, EPI_ISL_1021306, EPI_ISL_1021396, EPI_ISL_1025964, EPI_ISL_1026438, EPI_ISL_1026462, EPI_ISL_1026471, EPI_ISL_1026511, EPI_ISL_1026535, EPI_ISL_1026599, EPI_ISL_1026601, EPI_ISL_1026618, EPI_ISL_1026625, EPI_ISL_1026652, EPI_ISL_1026714, EPI_ISL_1026730, EPI_ISL_1026770, EPI_ISL_1026812, EPI_ISL_1026835, EPI_ISL_1027930, EPI_ISL_1030603, EPI_ISL_1030634, EPI_ISL_1030659, EPI_ISL_1030660, EPI_ISL_1030802, EPI_ISL_1030878, EPI_ISL_1031404, EPI_ISL_1031442, EPI_ISL_1031464, EPI_ISL_1031793, EPI_ISL_1031794, EPI_ISL_1031796, EPI_ISL_1032158, EPI_ISL_1032308, EPI_ISL_1032309, EPI_ISL_1032351, EPI_ISL_1032352, EPI_ISL_1032353, EPI_ISL_1032356, EPI_ISL_1032430, EPI_ISL_1036961, EPI_ISL_1037066, EPI_ISL_1037070, EPI_ISL_1037072, EPI_ISL_1037091, EPI_ISL_1037140, EPI_ISL_1037264, EPI_ISL_1037269, EPI_ISL_1037669, EPI_ISL_1037689, EPI_ISL_1037717, EPI_ISL_1037732, EPI_ISL_1037736, EPI_ISL_1037748, EPI_ISL_1037776, EPI_ISL_1037795, EPI_ISL_1037869, EPI_ISL_1037896, EPI_ISL_1037935, EPI_ISL_1037945, EPI_ISL_1037970, EPI_ISL_1038026, EPI_ISL_1038045, EPI_ISL_1038142, EPI_ISL_1038157, EPI_ISL_1038166, EPI_ISL_1038579, EPI_ISL_1038580, EPI_ISL_1081209, EPI_ISL_1159925, EPI_ISL_1160028, EPI_ISL_1160149, EPI_ISL_1160158, EPI_ISL_1160183, EPI_ISL_1160430, EPI_ISL_1160452, EPI_ISL_1160477, EPI_ISL_1160520, EPI_ISL_1160541, EPI_ISL_1160554, EPI_ISL_1160570, EPI_ISL_1160627, EPI_ISL_1160662, EPI_ISL_1160668, EPI_ISL_1160693, EPI_ISL_1160713, EPI_ISL_1160789, EPI_ISL_1160852, EPI_ISL_1160854, EPI_ISL_1160949, EPI_ISL_1160950, EPI_ISL_1160951, EPI_ISL_1160952, EPI_ISL_1161148, EPI_ISL_1161150, EPI_ISL_1161151, EPI_ISL_1161176, EPI_ISL_1161182, EPI_ISL_1161183, EPI_ISL_1161184, EPI_ISL_1161185, EPI_ISL_1161417, EPI_ISL_1161418, EPI_ISL_1161455, EPI_ISL_1161475, EPI_ISL_1161554, EPI_ISL_1161588, EPI_ISL_1161646, EPI_ISL_1161681, EPI_ISL_1161684, EPI_ISL_1161702, EPI_ISL_1161755, EPI_ISL_1161768, EPI_ISL_1161794, EPI_ISL_1161807, EPI_ISL_1161809, EPI_ISL_1161810, EPI_ISL_1161814, EPI_ISL_1161840, EPI_ISL_1161841, EPI_ISL_1161877, EPI_ISL_1161884, EPI_ISL_1161893, EPI_ISL_1162316, EPI_ISL_1162318, EPI_ISL_1162325, EPI_ISL_1162327, EPI_ISL_1162328, EPI_ISL_1162338, EPI_ISL_1162468, EPI_ISL_1162579, EPI_ISL_1162580, EPI_ISL_1162602, EPI_ISL_1162605, EPI_ISL_1162694, EPI_ISL_1162714, EPI_ISL_1162718, EPI_ISL_1162753, EPI_ISL_1162786, EPI_ISL_1162788, EPI_ISL_1162795, EPI_ISL_1162803, EPI_ISL_1162804, EPI_ISL_1162825, EPI_ISL_1162830, EPI_ISL_1162837, EPI_ISL_1162886, EPI_ISL_1162892, EPI_ISL_1162895, EPI_ISL_1162896, EPI_ISL_1162917, EPI_ISL_1162932, EPI_ISL_1162971, EPI_ISL_1162975, EPI_ISL_1162977, EPI_ISL_1163001, EPI_ISL_1163004, EPI_ISL_1163005, EPI_ISL_1163013, EPI_ISL_1163014, EPI_ISL_1163015, EPI_ISL_1163054, EPI_ISL_1163100, EPI_ISL_1163163, EPI_ISL_1163226, EPI_ISL_1163252, EPI_ISL_1163255, EPI_ISL_1163256, EPI_ISL_1163283, EPI_ISL_1163284, EPI_ISL_1163297, EPI_ISL_1163308, EPI_ISL_1163342, EPI_ISL_1163394, EPI_ISL_1163444, EPI_ISL_1163455, EPI_ISL_1163518, EPI_ISL_1163519, EPI_ISL_1255296 | | | |
| see above | Laboratory Corporation of America | Respiratory Viruses Branch, Division of Viral Diseases, Centers for Disease Control and Prevention | Alexandre Bolze; Amanda Douglas; Amanda Suchanek; Andrea Throop; Ary Ascencio; Ayla Burns; Ben L. Rambo-Martin; Bobbi Croy; Brad Sickler; Brian Krueger; Brian Norvell; Charlotte Rivera-Garcia; Christine Tran; Christos Petropoulos; Clinton R. Paden; Craig Lukasik; Dakota Howard; David Becker; Dhwani Batra; Duncan MacCannell; Efren Sandoval; Eileen de Feo; Elizabeth Cirulli; Eric Allen; Eyad Almasri Debbie Boles; Geraint Levan; Goran Stevovic; Howard Engler; Hrushikesh Deshmukh; Jake Humphrey; James Lu; Jan Antico; Jana Schroth; Jason Nguyen; Jimmy Ramirez; Jingtao Liu; Joe Voshell; John Pruitt; Jonathan Meltzer; Jonathan Williams; Kelly Schiabor Barrett; Kim Gietzen; Kimberly Wagner; Lax Iyer; Lyndon Tilson; Magnus Isaksson; Manoj Jain; Marc Laurent; Marcia Eisenberg; Mary Ann Cristobal; Mary Williamson; Matthew Tolentino; Michael Levandoski; Mike Sapeta; Mindy Nye; Minoo Agarwal; Mohan Kolli; Nicole L. Washington; Nuthawin Charoensri; Oren Cohen; Peter W. Cook; Phil Febbo; Prashant Gupta; Qian Zeng; Rama Ghatti; Ryan Cho; Scott Parker; Scott Ryan; Shannon Wickline; Sherry Wang; Simon White; Stanley Letovsky; Steven Ragan; Summer Galloway; Suresh Babu Selvaraju; Susan Countryman; Susan Hicks; Suxiang Tong; Suzanne Dale; Thomas Urban; Tim Kuphal; Tricia Zwiefelhofer; Tyler Cassens; Vincent Drouillon; William Lee |
| EPI_ISL_1709946 | MSHS Clinical Microbiology Laboratories | MSHS Pathogen Surveillance Program | Adolfo García-Sastre; Adriana van de Guchte; Ajay Obla; Alberto Paniz-Mondolfi; Ana S. Gonzalez-Reiche; Angela Amoako; Ashley Salimbangon; Betsaida Salom Melo; Bremy Alburquerque; Brianne Ciferri; Charles Gleason; Daniel Floda; Deena R. Altman; Denise Jurczyszak; Emilia Mia Sordillo; Gintaras Deikus; Giulio Kleiner; Gopi Patel; Hala Alshammary; Harm van Bakel; Irina Oussenko; Jayeeta Dutta; Juan Soto; Julia Matthews; Katherine Beach; Kathryn Twyman; Kayla Russo; Komal Srivastava; Levy Sominsky; Mahmoud Awawda; Marta Luksza; Matthew M. Hernandez; Melissa Gitman; Michael D. Nowak; Mitchell J. Sullivan; Nancy Francoeur; Robert Sebra; Sarah Schaefer; Shelcie Fabre; Shwetha Hara Sridhar; Viviana Simon; Ying-Chih Wang; Zenab Khan |
| EPI_ISL_2489014 | Mako Medical | Centers for Disease Control and Prevention Division of Viral Diseases, Pathogen Discovery | Adrian Paskey; Benjamin Rambo-Martin; Christopher Gulvick; Clinton R. Paden; Dakota Howard; Darlene Wagner; Dhwani Batra; Duncan MacCannell; Jason Caravas; Kara Moser; Lauren Moon; Matthew Schmerer; Matthew Tugwell; Peter W. Cook; Scott Sammons; Shatavia Morrison; Yvette Unoarumhi |
| EPI_ISL_1017472, EPI_ISL_1017474, EPI_ISL_1017475, EPI_ISL_1017476, EPI_ISL_1017477, EPI_ISL_1017478, EPI_ISL_1017479, EPI_ISL_1017480, EPI_ISL_1017481, EPI_ISL_1017482, EPI_ISL_1017483, EPI_ISL_1017484, EPI_ISL_1017485, EPI_ISL_1017486, EPI_ISL_1017487, EPI_ISL_1017488, EPI_ISL_1017490, EPI_ISL_1017492, EPI_ISL_1017493, EPI_ISL_1017494, EPI_ISL_1017495, EPI_ISL_1017496, EPI_ISL_1017497, EPI_ISL_1017498, EPI_ISL_1017499, EPI_ISL_1038994, EPI_ISL_1038995, EPI_ISL_1067641, EPI_ISL_1067650, EPI_ISL_1067656, EPI_ISL_1067657, EPI_ISL_1067658, EPI_ISL_1067659, EPI_ISL_1067669, EPI_ISL_1067692, EPI_ISL_1067693, EPI_ISL_1067694, EPI_ISL_1091801, EPI_ISL_1091802, EPI_ISL_1091803, EPI_ISL_1091806, EPI_ISL_1091807, EPI_ISL_1091810, EPI_ISL_1091812, EPI_ISL_1091813, EPI_ISL_1091817, EPI_ISL_1091818, EPI_ISL_1091819, EPI_ISL_1091824, EPI_ISL_1091826, EPI_ISL_1091827, EPI_ISL_1091830, EPI_ISL_1091833, EPI_ISL_1091834, EPI_ISL_1091842, EPI_ISL_1091849, EPI_ISL_1091850, EPI_ISL_1091851, EPI_ISL_1091852, EPI_ISL_1091853, EPI_ISL_1091858, EPI_ISL_1091859, EPI_ISL_1091860, EPI_ISL_1091861, EPI_ISL_1091862, EPI_ISL_1091863, EPI_ISL_1091864, EPI_ISL_1091866, EPI_ISL_1091867, EPI_ISL_1091868, EPI_ISL_1112934, EPI_ISL_1112937, EPI_ISL_1112939 | | | |
| see above | Murphy Medical Associates | Grubaugh Lab - Yale School of Public Health | Anderson Brito; Anne Wyllie; Annie Watkins; Caleb Neal; Chaney Kalinich; Chantal Vogels; Isabel Ott; Joseph Fauver; Mallery Breban; Mary Petrone; Nathan Grubaugh; Steven Murphy; Tara Alpert |
| EPI_ISL_1182088, EPI_ISL_1182089, EPI_ISL_1182090, EPI_ISL_1182093, EPI_ISL_1182094, EPI_ISL_1225682, EPI_ISL_1225701, EPI_ISL_1225713, EPI_ISL_1293129, EPI_ISL_1293130, EPI_ISL_1293131, EPI_ISL_1293132, EPI_ISL_1293133, EPI_ISL_1293134, EPI_ISL_1293135, EPI_ISL_1293136, EPI_ISL_1293137, EPI_ISL_1293139, EPI_ISL_1378822, EPI_ISL_1378823, EPI_ISL_1378827, EPI_ISL_1378828, EPI_ISL_1378829, EPI_ISL_1509262, EPI_ISL_1509263, EPI_ISL_1509264, EPI_ISL_1509265, EPI_ISL_1509266, EPI_ISL_1509267, EPI_ISL_1509268, EPI_ISL_1509269, EPI_ISL_1674698, EPI_ISL_1674699, EPI_ISL_1674700, EPI_ISL_1674701, EPI_ISL_1674702, EPI_ISL_1674704, EPI_ISL_1674705, EPI_ISL_2159668, EPI_ISL_2159670, EPI_ISL_2159672 | | | |
| see above | Murphy Medical Association | Grubaugh Lab - Yale School of Public Health | Anderson Brito; Annie Watkins; Caleb Neal; Chaney Kalinich; Chantal Vogels; Eva Laszlo; Isabel Ott; Isabell Ott; Jessica Rothman; Joseph Fauver; Mallery Breban; Mary Petrone; Nathan Grubaugh; Steven Murphy; Tara Alpert |

| | | | |
|---|---|---|---|
| EPI_ISL_1789900, EPI_ISL_1789927, EPI_ISL_1790026, EPI_ISL_2663536 | NYU Langone Health | Departments of Pathology and Medicine, New York University School of Medicine | Adriana Heguy; Christian Marier; Dacia Dimartino; Emily Guzman; Gael Westby; Guiqing Wang; Paolo Cotzia; Paul Zappile; Peter Meyn; Sitharam Ramaswami; Yutong Zhang |

EPI_ISL_854842, EPI_ISL_854843, EPI_ISL_854844, EPI_ISL_854845, EPI_ISL_854846, EPI_ISL_854847, EPI_ISL_854848, EPI_ISL_854849, EPI_ISL_854850, EPI_ISL_854851, EPI_ISL_854852, EPI_ISL_854853, EPI_ISL_854854, EPI_ISL_854855, EPI_ISL_854856, EPI_ISL_854857, EPI_ISL_854858, EPI_ISL_854859, EPI_ISL_854860, EPI_ISL_854861, EPI_ISL_854862, EPI_ISL_854863, EPI_ISL_854864, EPI_ISL_854865,
EPI_ISL_854866, EPI_ISL_854867, EPI_ISL_854868, EPI_ISL_854869, EPI_ISL_854870, EPI_ISL_854871, EPI_ISL_854872, EPI_ISL_855081, EPI_ISL_855082, EPI_ISL_855083, EPI_ISL_855084, EPI_ISL_855085, EPI_ISL_855086, EPI_ISL_855087, EPI_ISL_855088, EPI_ISL_855089, EPI_ISL_855090, EPI_ISL_855091, EPI_ISL_855092, EPI_ISL_855093, EPI_ISL_855094, EPI_ISL_855095, EPI_ISL_877189, EPI_ISL_877198,
EPI_ISL_877199, EPI_ISL_877200, EPI_ISL_903526, EPI_ISL_903527, EPI_ISL_903528, EPI_ISL_903529, EPI_ISL_903530, EPI_ISL_904131, EPI_ISL_906424, EPI_ISL_906425, EPI_ISL_906426, EPI_ISL_906427, EPI_ISL_906428, EPI_ISL_906429

| | | | |
|---|---|---|---|
| see above | Quest Diagnostics | Quest Diagnostics | Anderson, B.; Bernstein; D.F.; Gerasimova, A.; Hua, M.; I.A.; K.E.; Kagan; L.E.; Lacbawan, F.; Liu Y.; Livingston; Owen, R.; Perez, A.; R.M.; Rosenthal; S.H.; Shalhout; Shlyakhter; Tanpaiboon, P. |

EPI_ISL_1159278, EPI_ISL_1159289, EPI_ISL_1159313, EPI_ISL_1193397, EPI_ISL_1193400, EPI_ISL_1193418, EPI_ISL_1193425, EPI_ISL_1193449, EPI_ISL_1193467, EPI_ISL_1193468, EPI_ISL_1193469, EPI_ISL_1193473, EPI_ISL_1193475, EPI_ISL_1193477, EPI_ISL_1193480, EPI_ISL_1193488, EPI_ISL_1193507, EPI_ISL_1193509, EPI_ISL_1193513, EPI_ISL_1193519, EPI_ISL_1193528, EPI_ISL_1193531,
EPI_ISL_1193550, EPI_ISL_1193551, EPI_ISL_1193574, EPI_ISL_1193597, EPI_ISL_1193598, EPI_ISL_1193603, EPI_ISL_1193604, EPI_ISL_1193623, EPI_ISL_1193645, EPI_ISL_1193655, EPI_ISL_1193657, EPI_ISL_1193779, EPI_ISL_1193782, EPI_ISL_1193800, EPI_ISL_1193801, EPI_ISL_1193868, EPI_ISL_1193895, EPI_ISL_1194056, EPI_ISL_1194059, EPI_ISL_1194071, EPI_ISL_1194083, EPI_ISL_1194103,
EPI_ISL_1194134, EPI_ISL_1194137, EPI_ISL_1194148, EPI_ISL_1194167, EPI_ISL_1194170, EPI_ISL_1194180, EPI_ISL_1194189, EPI_ISL_1194199, EPI_ISL_1194201, EPI_ISL_1194203, EPI_ISL_1194205, EPI_ISL_1194209, EPI_ISL_1194228, EPI_ISL_1194230, EPI_ISL_1194422, EPI_ISL_1194423, EPI_ISL_1194425, EPI_ISL_1194559, EPI_ISL_1194564, EPI_ISL_1194570, EPI_ISL_1220425, EPI_ISL_1220434,
EPI_ISL_1220518, EPI_ISL_1220534, EPI_ISL_1220569, EPI_ISL_1220630, EPI_ISL_1220638, EPI_ISL_1220641, EPI_ISL_1220642, EPI_ISL_1220682, EPI_ISL_1220703, EPI_ISL_1220704, EPI_ISL_1220713, EPI_ISL_1252921, EPI_ISL_1252954, EPI_ISL_1252968, EPI_ISL_1252983, EPI_ISL_1252994, EPI_ISL_1253000, EPI_ISL_1253009, EPI_ISL_1253012, EPI_ISL_1253015, EPI_ISL_1253199, EPI_ISL_1253211,
EPI_ISL_1253417, EPI_ISL_1253418, EPI_ISL_1253421, EPI_ISL_1253435, EPI_ISL_1253451, EPI_ISL_1253454, EPI_ISL_1253479, EPI_ISL_1253488, EPI_ISL_1253496, EPI_ISL_1253504, EPI_ISL_1253510, EPI_ISL_1253526, EPI_ISL_1253532, EPI_ISL_1254090, EPI_ISL_1254142, EPI_ISL_1254189, EPI_ISL_1254309, EPI_ISL_1254310, EPI_ISL_1254322, EPI_ISL_1254326, EPI_ISL_1254327, EPI_ISL_1254329,
EPI_ISL_1254335, EPI_ISL_1254336, EPI_ISL_1254342, EPI_ISL_1254343, EPI_ISL_1254347, EPI_ISL_1254354, EPI_ISL_1254355, EPI_ISL_1254368, EPI_ISL_1254369, EPI_ISL_1254370, EPI_ISL_1254372, EPI_ISL_1254375, EPI_ISL_1254383, EPI_ISL_1254395, EPI_ISL_1254401, EPI_ISL_1254402, EPI_ISL_1254404, EPI_ISL_1254412, EPI_ISL_1254419, EPI_ISL_1254420, EPI_ISL_1254523, EPI_ISL_1254530,
EPI_ISL_1265921, EPI_ISL_1265944, EPI_ISL_1265971, EPI_ISL_1265987, EPI_ISL_1266091, EPI_ISL_1266097, EPI_ISL_1266150, EPI_ISL_1266157, EPI_ISL_1266224, EPI_ISL_1266784, EPI_ISL_1266816, EPI_ISL_1266881, EPI_ISL_1267036, EPI_ISL_1267075, EPI_ISL_1267146, EPI_ISL_1267152, EPI_ISL_1267153, EPI_ISL_1267157, EPI_ISL_1267158, EPI_ISL_1267160, EPI_ISL_1267169, EPI_ISL_1267175,
EPI_ISL_1267176, EPI_ISL_1267187, EPI_ISL_1267209, EPI_ISL_1291175, EPI_ISL_1291185, EPI_ISL_1291388, EPI_ISL_1291391, EPI_ISL_1291402, EPI_ISL_1291406, EPI_ISL_1291407, EPI_ISL_1291408, EPI_ISL_1291483, EPI_ISL_1291489, EPI_ISL_1291492, EPI_ISL_1291493, EPI_ISL_1291494, EPI_ISL_1291498, EPI_ISL_1291508, EPI_ISL_1291509, EPI_ISL_1291522, EPI_ISL_1291531, EPI_ISL_1291534,
EPI_ISL_1291540, EPI_ISL_1291572, EPI_ISL_1291574, EPI_ISL_1291580, EPI_ISL_1297685, EPI_ISL_1297686, EPI_ISL_1297692, EPI_ISL_1297696, EPI_ISL_1297698, EPI_ISL_1297707, EPI_ISL_1297710, EPI_ISL_1297722, EPI_ISL_1297726, EPI_ISL_1297730, EPI_ISL_1297734, EPI_ISL_1297743, EPI_ISL_1297755, EPI_ISL_1298227, EPI_ISL_1298232, EPI_ISL_1298233, EPI_ISL_1298237, EPI_ISL_1314842,
EPI_ISL_1314900, EPI_ISL_1314936, EPI_ISL_1314947, EPI_ISL_1314964, EPI_ISL_1335910, EPI_ISL_1335922, EPI_ISL_1335943, EPI_ISL_1335944, EPI_ISL_1335947, EPI_ISL_1335965, EPI_ISL_1335998, EPI_ISL_1336038, EPI_ISL_1336043, EPI_ISL_1336045, EPI_ISL_1336074, EPI_ISL_1336075, EPI_ISL_1336097, EPI_ISL_1336111, EPI_ISL_1336113, EPI_ISL_1336115, EPI_ISL_1336116, EPI_ISL_1336117,
EPI_ISL_1336121, EPI_ISL_1336123, EPI_ISL_1336124, EPI_ISL_1336125, EPI_ISL_1336126, EPI_ISL_1336127, EPI_ISL_1336128, EPI_ISL_1339904, EPI_ISL_1339909, EPI_ISL_1339920, EPI_ISL_1339921, EPI_ISL_1339923, EPI_ISL_1339927, EPI_ISL_1339929, EPI_ISL_1339930, EPI_ISL_1339931, EPI_ISL_1339934, EPI_ISL_1339936, EPI_ISL_1339937, EPI_ISL_1339942, EPI_ISL_1339944, EPI_ISL_1339945,
EPI_ISL_1339946, EPI_ISL_1339948, EPI_ISL_1339950, EPI_ISL_1339952, EPI_ISL_1339954, EPI_ISL_1339958, EPI_ISL_1339961, EPI_ISL_1339963, EPI_ISL_1339965, EPI_ISL_1339967, EPI_ISL_1340497, EPI_ISL_1340523, EPI_ISL_1340532, EPI_ISL_1340540, EPI_ISL_1340553, EPI_ISL_1340561, EPI_ISL_1340582, EPI_ISL_1340592, EPI_ISL_1340596, EPI_ISL_1340601, EPI_ISL_1340602, EPI_ISL_1340628,
EPI_ISL_1340632, EPI_ISL_1340641, EPI_ISL_1340688, EPI_ISL_1340689, EPI_ISL_1340706, EPI_ISL_1340709, EPI_ISL_1340717, EPI_ISL_1340719, EPI_ISL_1340731, EPI_ISL_1367264, EPI_ISL_1367265, EPI_ISL_1367269, EPI_ISL_1367279, EPI_ISL_1367310, EPI_ISL_1367449, EPI_ISL_1367462, EPI_ISL_1391427, EPI_ISL_1391655, EPI_ISL_1391660, EPI_ISL_1391663, EPI_ISL_1391666, EPI_ISL_1391689,
EPI_ISL_1422338, EPI_ISL_1422357, EPI_ISL_1422400, EPI_ISL_1422419, EPI_ISL_1422422, EPI_ISL_1422429, EPI_ISL_1422452, EPI_ISL_1422462, EPI_ISL_1422470, EPI_ISL_1422493, EPI_ISL_1422506, EPI_ISL_1422508, EPI_ISL_1422521, EPI_ISL_1422532, EPI_ISL_1422534, EPI_ISL_1422546, EPI_ISL_1422548, EPI_ISL_1422553, EPI_ISL_1422554, EPI_ISL_1422648, EPI_ISL_1422689, EPI_ISL_1422691,
EPI_ISL_1422698, EPI_ISL_1422703, EPI_ISL_1422709, EPI_ISL_1422776, EPI_ISL_1422782, EPI_ISL_1422788, EPI_ISL_1422790, EPI_ISL_1422791, EPI_ISL_1422794, EPI_ISL_1422799, EPI_ISL_1422800, EPI_ISL_1422801, EPI_ISL_1422938, EPI_ISL_1422950, EPI_ISL_1422977, EPI_ISL_1444348, EPI_ISL_1444356, EPI_ISL_1444369, EPI_ISL_1444382, EPI_ISL_1444393, EPI_ISL_1444460, EPI_ISL_1444467,
EPI_ISL_1444502, EPI_ISL_1444528, EPI_ISL_1444531, EPI_ISL_1444532, EPI_ISL_1444534, EPI_ISL_1444535, EPI_ISL_1444544, EPI_ISL_1444546, EPI_ISL_1444556, EPI_ISL_1444561, EPI_ISL_1444562, EPI_ISL_1444689, EPI_ISL_1444711, EPI_ISL_1460738, EPI_ISL_1479760, EPI_ISL_1479796, EPI_ISL_1479915, EPI_ISL_1479918, EPI_ISL_1479924, EPI_ISL_1479930, EPI_ISL_1479932, EPI_ISL_1479938,
EPI_ISL_1479943, EPI_ISL_1479944, EPI_ISL_1479945, EPI_ISL_1479946, EPI_ISL_1479947, EPI_ISL_1479949, EPI_ISL_1479952, EPI_ISL_1479955, EPI_ISL_1479960, EPI_ISL_1480528, EPI_ISL_1480594, EPI_ISL_1480609, EPI_ISL_1480621, EPI_ISL_1480630, EPI_ISL_1480631, EPI_ISL_1480639, EPI_ISL_1480647, EPI_ISL_1480664, EPI_ISL_1480668, EPI_ISL_1480670, EPI_ISL_1480784, EPI_ISL_1480790,
EPI_ISL_1480810, EPI_ISL_1480822, EPI_ISL_1480823, EPI_ISL_1480837, EPI_ISL_1480844, EPI_ISL_1480855, EPI_ISL_1480871, EPI_ISL_1480925, EPI_ISL_1491952, EPI_ISL_1491953, EPI_ISL_1491957, EPI_ISL_1491959, EPI_ISL_1491960, EPI_ISL_1491962, EPI_ISL_1491963, EPI_ISL_1491972, EPI_ISL_1491975, EPI_ISL_1491982, EPI_ISL_1491991, EPI_ISL_1492006, EPI_ISL_1492007, EPI_ISL_1492009,
EPI_ISL_1492013, EPI_ISL_1492017, EPI_ISL_1492018, EPI_ISL_1492026, EPI_ISL_1493609, EPI_ISL_1493641, EPI_ISL_1493654, EPI_ISL_1493961, EPI_ISL_1493985, EPI_ISL_1493989, EPI_ISL_1493991, EPI_ISL_1493992, EPI_ISL_1493993, EPI_ISL_1493994, EPI_ISL_1493996, EPI_ISL_1493998, EPI_ISL_1494001, EPI_ISL_1494002, EPI_ISL_1494003, EPI_ISL_1494005, EPI_ISL_1494007, EPI_ISL_1494008,
EPI_ISL_1494012, EPI_ISL_1494017, EPI_ISL_1494022, EPI_ISL_1494023, EPI_ISL_1494024, EPI_ISL_1494025, EPI_ISL_1494026, EPI_ISL_1494027, EPI_ISL_1494029, EPI_ISL_1494030, EPI_ISL_1526066, EPI_ISL_1551754, EPI_ISL_1551758, EPI_ISL_1551760, EPI_ISL_1551823, EPI_ISL_1551828, EPI_ISL_1551839, EPI_ISL_1551854, EPI_ISL_1551860, EPI_ISL_1551868, EPI_ISL_1551871, EPI_ISL_1551879,
EPI_ISL_1551965, EPI_ISL_1551971, EPI_ISL_1551972, EPI_ISL_1551980, EPI_ISL_1551981, EPI_ISL_1552001, EPI_ISL_1552161, EPI_ISL_1552187, EPI_ISL_1552190, EPI_ISL_1552203, EPI_ISL_1552321, EPI_ISL_1552342, EPI_ISL_1552349, EPI_ISL_1552353, EPI_ISL_1552358, EPI_ISL_1552362, EPI_ISL_1552364, EPI_ISL_1552365, EPI_ISL_1552368, EPI_ISL_1552371, EPI_ISL_1552379, EPI_ISL_1552382,
EPI_ISL_1552385, EPI_ISL_1552390, EPI_ISL_1552394, EPI_ISL_1552395, EPI_ISL_1552400, EPI_ISL_1552402, EPI_ISL_1552403, EPI_ISL_1552408, EPI_ISL_1553130, EPI_ISL_1553149, EPI_ISL_1553157, EPI_ISL_1553166, EPI_ISL_1582042, EPI_ISL_1582049, EPI_ISL_1582072, EPI_ISL_1582074, EPI_ISL_1582085, EPI_ISL_1582153, EPI_ISL_1582169, EPI_ISL_1622020, EPI_ISL_1648359, EPI_ISL_1668788,
EPI_ISL_1702580, EPI_ISL_1702602, EPI_ISL_1702609, EPI_ISL_1702611, EPI_ISL_1702612, EPI_ISL_1702630, EPI_ISL_1702635, EPI_ISL_1702641, EPI_ISL_1702649, EPI_ISL_1702655, EPI_ISL_1702656, EPI_ISL_1702662, EPI_ISL_1702667, EPI_ISL_1702670, EPI_ISL_1702681, EPI_ISL_1702687, EPI_ISL_1702694, EPI_ISL_1702703, EPI_ISL_1702708, EPI_ISL_1702712, EPI_ISL_1737882, EPI_ISL_1737883,
EPI_ISL_1737884, EPI_ISL_1737887, EPI_ISL_1737888, EPI_ISL_1737889, EPI_ISL_1737892, EPI_ISL_1737894, EPI_ISL_1737896, EPI_ISL_1737898, EPI_ISL_1737900, EPI_ISL_1737902, EPI_ISL_1737903, EPI_ISL_1737909, EPI_ISL_1737910, EPI_ISL_1737912, EPI_ISL_1737914, EPI_ISL_1737918, EPI_ISL_1737920, EPI_ISL_1737921, EPI_ISL_1737923, EPI_ISL_1737938, EPI_ISL_1737940, EPI_ISL_1737944,
EPI_ISL_1737945, EPI_ISL_1737946, EPI_ISL_1737947, EPI_ISL_1737948, EPI_ISL_1737951, EPI_ISL_1737952, EPI_ISL_1737953, EPI_ISL_1737955, EPI_ISL_1737956, EPI_ISL_1737962, EPI_ISL_1737963, EPI_ISL_1737968, EPI_ISL_1737969, EPI_ISL_1737970, EPI_ISL_1737975, EPI_ISL_1737976, EPI_ISL_1737980, EPI_ISL_1737984, EPI_ISL_1737985, EPI_ISL_1737986, EPI_ISL_1737999, EPI_ISL_1738000,
EPI_ISL_1738001, EPI_ISL_1738002, EPI_ISL_1738004, EPI_ISL_1738005, EPI_ISL_1738007, EPI_ISL_1738009, EPI_ISL_1738012, EPI_ISL_1738014, EPI_ISL_1738017, EPI_ISL_1738019, EPI_ISL_1738021, EPI_ISL_1738027, EPI_ISL_1738029, EPI_ISL_1738032, EPI_ISL_1738047, EPI_ISL_1738053, EPI_ISL_1738055, EPI_ISL_1738058, EPI_ISL_1738059, EPI_ISL_1738072, EPI_ISL_1738085, EPI_ISL_1738099,
EPI_ISL_1738100, EPI_ISL_1738142, EPI_ISL_1738168, EPI_ISL_1738327, EPI_ISL_1738328, EPI_ISL_1738332, EPI_ISL_1738371, EPI_ISL_1738373, EPI_ISL_1738402, EPI_ISL_1738451, EPI_ISL_1738472, EPI_ISL_1738517, EPI_ISL_1738555, EPI_ISL_1738564, EPI_ISL_1738609, EPI_ISL_1738633, EPI_ISL_1738634, EPI_ISL_1738638, EPI_ISL_1738645, EPI_ISL_1738656, EPI_ISL_1738657, EPI_ISL_1738693,
EPI_ISL_1738702, EPI_ISL_1738715, EPI_ISL_1738735, EPI_ISL_1738737, EPI_ISL_1738746, EPI_ISL_1738749, EPI_ISL_1753065, EPI_ISL_1753067, EPI_ISL_1753072, EPI_ISL_1753115, EPI_ISL_1753119, EPI_ISL_1753120, EPI_ISL_1753129, EPI_ISL_1753142, EPI_ISL_1753192, EPI_ISL_1753209, EPI_ISL_1753285, EPI_ISL_1753291, EPI_ISL_1753297, EPI_ISL_1753298, EPI_ISL_1753303, EPI_ISL_1753305,
EPI_ISL_1753312, EPI_ISL_1753329, EPI_ISL_1753357, EPI_ISL_1753358, EPI_ISL_1753359, EPI_ISL_1753368, EPI_ISL_1753394, EPI_ISL_1753409, EPI_ISL_1753417, EPI_ISL_1753613, EPI_ISL_1753618, EPI_ISL_1753620, EPI_ISL_1753640, EPI_ISL_1753648, EPI_ISL_1753658, EPI_ISL_1761882, EPI_ISL_1761887, EPI_ISL_1761889, EPI_ISL_1761892, EPI_ISL_1797881, EPI_ISL_1797883, EPI_ISL_1797884,
EPI_ISL_1797890, EPI_ISL_1797892, EPI_ISL_1797895, EPI_ISL_1797900, EPI_ISL_1797902, EPI_ISL_1797906, EPI_ISL_1797909, EPI_ISL_1797914, EPI_ISL_1797917, EPI_ISL_1797920, EPI_ISL_1797924, EPI_ISL_1797928, EPI_ISL_1797936, EPI_ISL_1797938, EPI_ISL_1797943, EPI_ISL_1797946, EPI_ISL_1797947, EPI_ISL_1797953, EPI_ISL_1797958, EPI_ISL_1797959, EPI_ISL_1797960, EPI_ISL_1797962,
EPI_ISL_1797965, EPI_ISL_1797966, EPI_ISL_1797971, EPI_ISL_1797974, EPI_ISL_1797975, EPI_ISL_1797976, EPI_ISL_1797979, EPI_ISL_1797991, EPI_ISL_1797997, EPI_ISL_1798000, EPI_ISL_1798001, EPI_ISL_1798005, EPI_ISL_1798006, EPI_ISL_1798008, EPI_ISL_1798010, EPI_ISL_1798014, EPI_ISL_1798017, EPI_ISL_1798025, EPI_ISL_1798026, EPI_ISL_1798030, EPI_ISL_1798033, EPI_ISL_1818057,
EPI_ISL_1818140, EPI_ISL_1818150, EPI_ISL_1818151, EPI_ISL_1818160, EPI_ISL_1818162, EPI_ISL_1818169, EPI_ISL_1818172, EPI_ISL_1818173, EPI_ISL_1818175, EPI_ISL_1818178, EPI_ISL_1818179, EPI_ISL_1818181, EPI_ISL_1818182, EPI_ISL_1818186, EPI_ISL_1818187, EPI_ISL_1818188, EPI_ISL_1818194, EPI_ISL_1818197, EPI_ISL_1818205, EPI_ISL_1818208, EPI_ISL_1818210, EPI_ISL_1818216,
EPI_ISL_1818217, EPI_ISL_1818218, EPI_ISL_1818221, EPI_ISL_1818235, EPI_ISL_1818237, EPI_ISL_1818238, EPI_ISL_1818239, EPI_ISL_1818242, EPI_ISL_1818250, EPI_ISL_1818252, EPI_ISL_1818254, EPI_ISL_1818266, EPI_ISL_1818267, EPI_ISL_1818268, EPI_ISL_1818270, EPI_ISL_1818273, EPI_ISL_1818276, EPI_ISL_1818536, EPI_ISL_1834228, EPI_ISL_1834230, EPI_ISL_1834233, EPI_ISL_1834263,
EPI_ISL_1834273, EPI_ISL_1834276, EPI_ISL_1834280, EPI_ISL_1834315, EPI_ISL_1834323, EPI_ISL_1834360, EPI_ISL_1834365, EPI_ISL_1834372, EPI_ISL_1834377, EPI_ISL_1834379, EPI_ISL_1834391, EPI_ISL_1834428, EPI_ISL_1834441, EPI_ISL_1834442, EPI_ISL_1838963, EPI_ISL_1838982, EPI_ISL_1838992, EPI_ISL_1840270, EPI_ISL_1840470, EPI_ISL_1840483, EPI_ISL_1840491, EPI_ISL_1840554,
EPI_ISL_1840596, EPI_ISL_1906409, EPI_ISL_1990411, EPI_ISL_1990418, EPI_ISL_1990423, EPI_ISL_1990424, EPI_ISL_1990425, EPI_ISL_1990426, EPI_ISL_1990431, EPI_ISL_1990435, EPI_ISL_1990444, EPI_ISL_1990447, EPI_ISL_1990456, EPI_ISL_1990476, EPI_ISL_1990478, EPI_ISL_1990484, EPI_ISL_1990485, EPI_ISL_1990488, EPI_ISL_1990504, EPI_ISL_1990512, EPI_ISL_1990522, EPI_ISL_1990531,
EPI_ISL_1990535, EPI_ISL_1990537, EPI_ISL_1990538, EPI_ISL_1990540, EPI_ISL_2011027, EPI_ISL_2011029, EPI_ISL_2011031, EPI_ISL_2011032, EPI_ISL_2011039, EPI_ISL_2011044, EPI_ISL_2011048, EPI_ISL_2011049, EPI_ISL_2011051, EPI_ISL_2011052, EPI_ISL_2011054, EPI_ISL_2011056, EPI_ISL_2011057, EPI_ISL_2011058, EPI_ISL_2011063, EPI_ISL_2011068, EPI_ISL_2011069, EPI_ISL_2011077,
EPI_ISL_2011080, EPI_ISL_2011083, EPI_ISL_2011086, EPI_ISL_2011126, EPI_ISL_2011138, EPI_ISL_2011146, EPI_ISL_2011148, EPI_ISL_2011151, EPI_ISL_2011168, EPI_ISL_2011170, EPI_ISL_2011176, EPI_ISL_2011190, EPI_ISL_2011192, EPI_ISL_2011203, EPI_ISL_2011228, EPI_ISL_2011229, EPI_ISL_2035324, EPI_ISL_2035334, EPI_ISL_2035339, EPI_ISL_2035344, EPI_ISL_2046985, EPI_ISL_2047097,
EPI_ISL_2047125, EPI_ISL_2090473, EPI_ISL_2090483, EPI_ISL_2090490, EPI_ISL_2090503, EPI_ISL_2090509, EPI_ISL_2090518, EPI_ISL_2090541, EPI_ISL_2090555, EPI_ISL_2090559, EPI_ISL_2090568, EPI_ISL_2090585, EPI_ISL_2098289, EPI_ISL_2098300, EPI_ISL_2098314, EPI_ISL_2098321, EPI_ISL_2098364, EPI_ISL_2098380, EPI_ISL_2132580, EPI_ISL_2133097, EPI_ISL_2133100, EPI_ISL_2133101,
EPI_ISL_2133103, EPI_ISL_2133105, EPI_ISL_2133107, EPI_ISL_2133109, EPI_ISL_2133112, EPI_ISL_2133115, EPI_ISL_2133117, EPI_ISL_2133120, EPI_ISL_2133124, EPI_ISL_2133140, EPI_ISL_2133146, EPI_ISL_2133147, EPI_ISL_2133149, EPI_ISL_2133151, EPI_ISL_2133152, EPI_ISL_2133153, EPI_ISL_2133163, EPI_ISL_2133164, EPI_ISL_2133166, EPI_ISL_2133169, EPI_ISL_2133175, EPI_ISL_2133183,
EPI_ISL_2133189, EPI_ISL_2133193, EPI_ISL_2133195, EPI_ISL_2133196, EPI_ISL_2133197, EPI_ISL_2133198, EPI_ISL_2133200, EPI_ISL_2133201, EPI_ISL_2133212, EPI_ISL_2133213, EPI_ISL_2133219, EPI_ISL_2133220, EPI_ISL_2133231, EPI_ISL_2133332, EPI_ISL_2133395, EPI_ISL_2133396, EPI_ISL_2133409, EPI_ISL_2133411, EPI_ISL_2133422, EPI_ISL_2133439, EPI_ISL_2133452, EPI_ISL_2133455,
EPI_ISL_2133465, EPI_ISL_2133466, EPI_ISL_2133468, EPI_ISL_2133469, EPI_ISL_2133473, EPI_ISL_2133478, EPI_ISL_2133486, EPI_ISL_2133488, EPI_ISL_2133508, EPI_ISL_2133519, EPI_ISL_2133523, EPI_ISL_2133526, EPI_ISL_2133659, EPI_ISL_2133663, EPI_ISL_2140733, EPI_ISL_2140737, EPI_ISL_2140739, EPI_ISL_2143027, EPI_ISL_2143035, EPI_ISL_2143038, EPI_ISL_2143048, EPI_ISL_2143056,
EPI_ISL_2143058, EPI_ISL_2143072, EPI_ISL_2143074, EPI_ISL_2143088, EPI_ISL_2143093, EPI_ISL_2143103, EPI_ISL_2143113, EPI_ISL_2143114, EPI_ISL_2143135, EPI_ISL_2143140, EPI_ISL_2143146, EPI_ISL_2143157, EPI_ISL_2179971, EPI_ISL_2180464, EPI_ISL_2180476, EPI_ISL_2180514, EPI_ISL_2180515, EPI_ISL_2187245, EPI_ISL_2187246, EPI_ISL_2187251, EPI_ISL_2268408, EPI_ISL_2268417,
EPI_ISL_2268431, EPI_ISL_2268476, EPI_ISL_2268481, EPI_ISL_2268495, EPI_ISL_2268504, EPI_ISL_2268523, EPI_ISL_2268547, EPI_ISL_2268554, EPI_ISL_2268570, EPI_ISL_2268593, EPI_ISL_2268644, EPI_ISL_2268674, EPI_ISL_2268679, EPI_ISL_2268681, EPI_ISL_2268682, EPI_ISL_2268683, EPI_ISL_2268686, EPI_ISL_2268687, EPI_ISL_2268693, EPI_ISL_2268694, EPI_ISL_2268695, EPI_ISL_2268700,
EPI_ISL_2268708, EPI_ISL_2268715, EPI_ISL_2268716, EPI_ISL_2268718, EPI_ISL_2268719, EPI_ISL_2268720, EPI_ISL_2268724, EPI_ISL_2268733, EPI_ISL_2268734, EPI_ISL_2268736, EPI_ISL_2268740, EPI_ISL_2268747, EPI_ISL_2268756, EPI_ISL_2268759, EPI_ISL_2268760, EPI_ISL_2268768, EPI_ISL_2268770, EPI_ISL_2268776, EPI_ISL_2268778, EPI_ISL_2268779, EPI_ISL_2268780, EPI_ISL_2268781,
EPI_ISL_2268785, EPI_ISL_2268791, EPI_ISL_2268801, EPI_ISL_2268804, EPI_ISL_2268805, EPI_ISL_2268810, EPI_ISL_2268812, EPI_ISL_2268813, EPI_ISL_2268827, EPI_ISL_2268830, EPI_ISL_2268831, EPI_ISL_2268837, EPI_ISL_2268838, EPI_ISL_2268839, EPI_ISL_2268841, EPI_ISL_2268845, EPI_ISL_2268848, EPI_ISL_2268859, EPI_ISL_2268860, EPI_ISL_2268862, EPI_ISL_2268867, EPI_ISL_2268868,
EPI_ISL_2268869, EPI_ISL_2268875, EPI_ISL_2268876, EPI_ISL_2268879, EPI_ISL_2268893, EPI_ISL_2268900, EPI_ISL_2268909, EPI_ISL_2268914, EPI_ISL_2268919, EPI_ISL_2268922, EPI_ISL_2268923, EPI_ISL_2268925, EPI_ISL_2268926, EPI_ISL_2268930, EPI_ISL_2268935, EPI_ISL_2269079, EPI_ISL_2269095, EPI_ISL_2269096, EPI_ISL_2269098, EPI_ISL_2269103, EPI_ISL_2269110, EPI_ISL_2269113,
EPI_ISL_2269117, EPI_ISL_2269122, EPI_ISL_2269130, EPI_ISL_2269131, EPI_ISL_2269136, EPI_ISL_2269137, EPI_ISL_2269142, EPI_ISL_2269146, EPI_ISL_2269169, EPI_ISL_2269175, EPI_ISL_2269178, EPI_ISL_2269192, EPI_ISL_2269194, EPI_ISL_2269195, EPI_ISL_2269203, EPI_ISL_2269206, EPI_ISL_2269212, EPI_ISL_2269213, EPI_ISL_2280172, EPI_ISL_2367954, EPI_ISL_2367959, EPI_ISL_2367962,
EPI_ISL_2367967, EPI_ISL_2367968, EPI_ISL_2367975, EPI_ISL_2367981, EPI_ISL_2367988, EPI_ISL_2367993, EPI_ISL_2367998, EPI_ISL_2368000, EPI_ISL_2368008, EPI_ISL_2368017, EPI_ISL_2368027, EPI_ISL_2368028, EPI_ISL_2368029, EPI_ISL_2368030, EPI_ISL_2368047, EPI_ISL_2368063, EPI_ISL_2371725, EPI_ISL_2371742, EPI_ISL_2371762, EPI_ISL_2371763, EPI_ISL_2371764, EPI_ISL_2371765,
EPI_ISL_2371768, EPI_ISL_2371769, EPI_ISL_2371782, EPI_ISL_2371783, EPI_ISL_2371786, EPI_ISL_2371790, EPI_ISL_2371795, EPI_ISL_2371825, EPI_ISL_2371827, EPI_ISL_2371832, EPI_ISL_2371835, EPI_ISL_2371836, EPI_ISL_2371842, EPI_ISL_2371857, EPI_ISL_2371871, EPI_ISL_2371884, EPI_ISL_2371888, EPI_ISL_2371889, EPI_ISL_2371896, EPI_ISL_2371917, EPI_ISL_2371928, EPI_ISL_2371954,
EPI_ISL_2371961, EPI_ISL_2371976, EPI_ISL_2372005, EPI_ISL_2372008, EPI_ISL_2372010, EPI_ISL_2372024, EPI_ISL_2372031, EPI_ISL_2372046, EPI_ISL_2372070, EPI_ISL_2372081, EPI_ISL_2372085, EPI_ISL_2372104, EPI_ISL_2372111, EPI_ISL_2372113, EPI_ISL_2372116, EPI_ISL_2372126, EPI_ISL_2372127, EPI_ISL_2372129, EPI_ISL_2372139, EPI_ISL_2372144, EPI_ISL_2372148, EPI_ISL_2372152,
EPI_ISL_2372163, EPI_ISL_2372164, EPI_ISL_2372181, EPI_ISL_2372182, EPI_ISL_2372186, EPI_ISL_2372189, EPI_ISL_2372204, EPI_ISL_2372205, EPI_ISL_2372208, EPI_ISL_2372218, EPI_ISL_2372223, EPI_ISL_2372226, EPI_ISL_2372231, EPI_ISL_2372233, EPI_ISL_2372235, EPI_ISL_2372237, EPI_ISL_2372257, EPI_ISL_2372263, EPI_ISL_2372905, EPI_ISL_2372910, EPI_ISL_2372911, EPI_ISL_2372926,
EPI_ISL_2372934, EPI_ISL_2372935, EPI_ISL_2372937, EPI_ISL_2397329, EPI_ISL_2397330, EPI_ISL_2397334, EPI_ISL_2397353, EPI_ISL_2397364, EPI_ISL_2397399, EPI_ISL_2397428, EPI_ISL_2397468, EPI_ISL_2397503, EPI_ISL_2397521, EPI_ISL_2397547, EPI_ISL_2397551, EPI_ISL_2397557, EPI_ISL_2397561, EPI_ISL_2397578, EPI_ISL_2397582, EPI_ISL_2397597, EPI_ISL_2397598, EPI_ISL_2397599,
EPI_ISL_2397603, EPI_ISL_2397608, EPI_ISL_2397617, EPI_ISL_2397620, EPI_ISL_2397625, EPI_ISL_2397628, EPI_ISL_2397632, EPI_ISL_2397633, EPI_ISL_2397641, EPI_ISL_2397655, EPI_ISL_2397656, EPI_ISL_2397657, EPI_ISL_2397661, EPI_ISL_2397662, EPI_ISL_2397670, EPI_ISL_2397677, EPI_ISL_2397690, EPI_ISL_2397692, EPI_ISL_2397693, EPI_ISL_2397700, EPI_ISL_2397702, EPI_ISL_2397704,
EPI_ISL_2397706, EPI_ISL_2397708, EPI_ISL_2397724, EPI_ISL_2397730, EPI_ISL_2397733, EPI_ISL_2397740, EPI_ISL_2397742, EPI_ISL_2397746, EPI_ISL_2397751, EPI_ISL_2397754, EPI_ISL_2397766, EPI_ISL_2397776, EPI_ISL_2397778, EPI_ISL_2397779, EPI_ISL_2397797, EPI_ISL_2399472, EPI_ISL_2399495, EPI_ISL_2399501, EPI_ISL_2439296, EPI_ISL_2439566, EPI_ISL_2439573, EPI_ISL_2439575,
EPI_ISL_2439578, EPI_ISL_2439579, EPI_ISL_2439588, EPI_ISL_2439592, EPI_ISL_2439598, EPI_ISL_2439600, EPI_ISL_2439603, EPI_ISL_2439605, EPI_ISL_2439618, EPI_ISL_2439632, EPI_ISL_2439652, EPI_ISL_2439654, EPI_ISL_2439657, EPI_ISL_2439659, EPI_ISL_2439663, EPI_ISL_2439671, EPI_ISL_2439679, EPI_ISL_2439690, EPI_ISL_2439691, EPI_ISL_2439752, EPI_ISL_2439882, EPI_ISL_2439917,
EPI_ISL_2439938, EPI_ISL_2439939, EPI_ISL_2439951, EPI_ISL_2440549, EPI_ISL_2440552, EPI_ISL_2440557, EPI_ISL_2440565, EPI_ISL_2440566, EPI_ISL_2440581, EPI_ISL_2440589, EPI_ISL_2440616, EPI_ISL_2440618, EPI_ISL_2440631, EPI_ISL_2440642, EPI_ISL_2440649, EPI_ISL_2440658, EPI_ISL_2440659, EPI_ISL_2440663, EPI_ISL_2440672, EPI_ISL_2440673, EPI_ISL_2440679, EPI_ISL_2440680,
EPI_ISL_2440700, EPI_ISL_2440705, EPI_ISL_2440710, EPI_ISL_2440720, EPI_ISL_2440742, EPI_ISL_2440770, EPI_ISL_2440771, EPI_ISL_2440791, EPI_ISL_2440795, EPI_ISL_2440796, EPI_ISL_2440803, EPI_ISL_2440816, EPI_ISL_2440836, EPI_ISL_2440847, EPI_ISL_2440884, EPI_ISL_2440891, EPI_ISL_2440892, EPI_ISL_2440894, EPI_ISL_2451290, EPI_ISL_2451304, EPI_ISL_2451309, EPI_ISL_2451319,
EPI_ISL_2451322, EPI_ISL_2451334, EPI_ISL_2451349, EPI_ISL_2451355, EPI_ISL_2451361, EPI_ISL_2451367, EPI_ISL_2451368, EPI_ISL_2451378, EPI_ISL_2451382, EPI_ISL_2451394, EPI_ISL_2451480, EPI_ISL_2451531, EPI_ISL_2451533, EPI_ISL_2451537, EPI_ISL_2451539, EPI_ISL_2451543, EPI_ISL_2451556, EPI_ISL_2463019, EPI_ISL_2463034, EPI_ISL_2463040, EPI_ISL_2463042, EPI_ISL_2463064,
EPI_ISL_2463066, EPI_ISL_2463067, EPI_ISL_2463086, EPI_ISL_2463089, EPI_ISL_2463090, EPI_ISL_2463098, EPI_ISL_2463100, EPI_ISL_2463101, EPI_ISL_2463106, EPI_ISL_2463107, EPI_ISL_2463116, EPI_ISL_2463119, EPI_ISL_2463153, EPI_ISL_2463204, EPI_ISL_2463216, EPI_ISL_2463231, EPI_ISL_2463234, EPI_ISL_2463250, EPI_ISL_2488832, EPI_ISL_2488858, EPI_ISL_2526788, EPI_ISL_2526972,
EPI_ISL_2529102, EPI_ISL_2529166, EPI_ISL_2529190, EPI_ISL_2529212, EPI_ISL_2529236, EPI_ISL_2529256, EPI_ISL_2529318, EPI_ISL_2529338, EPI_ISL_2534555, EPI_ISL_2534563, EPI_ISL_2599655, EPI_ISL_2599680, EPI_ISL_2599687, EPI_ISL_2599722, EPI_ISL_2599870, EPI_ISL_2599893, EPI_ISL_2611833, EPI_ISL_2611838, EPI_ISL_2611862, EPI_ISL_2649948, EPI_ISL_2649950, EPI_ISL_2649954,
EPI_ISL_2649989, EPI_ISL_2652272, EPI_ISL_2652304, EPI_ISL_2652305, EPI_ISL_2652309, EPI_ISL_2652313, EPI_ISL_2652315, EPI_ISL_2652346, EPI_ISL_2652451, EPI_ISL_2686368, EPI_ISL_2686371, EPI_ISL_2686373, EPI_ISL_2868813, EPI_ISL_2868919, EPI_ISL_2869056, EPI_ISL_2869090, EPI_ISL_2869101, EPI_ISL_2870363, EPI_ISL_2870427, EPI_ISL_2870542, EPI_ISL_2870660, EPI_ISL_2871006,
EPI_ISL_2873905, EPI_ISL_2874058, EPI_ISL_2874113, EPI_ISL_2874156, EPI_ISL_2874516, EPI_ISL_2876139, EPI_ISL_2876740, EPI_ISL_2876753, EPI_ISL_2876762, EPI_ISL_2876764, EPI_ISL_2876851, EPI_ISL_2930003, EPI_ISL_2930104, EPI_ISL_2930186, EPI_ISL_2930238, EPI_ISL_2930270, EPI_ISL_2930273, EPI_ISL_2930289, EPI_ISL_2930312, EPI_ISL_2930321, EPI_ISL_3018282, EPI_ISL_3018391,
EPI_ISL_3018417, EPI_ISL_3018509, EPI_ISL_3085838, EPI_ISL_3113761, EPI_ISL_3113808, EPI_ISL_3113827, EPI_ISL_3113867, EPI_ISL_3113906, EPI_ISL_3113909, EPI_ISL_3113943, EPI_ISL_3114162, EPI_ISL_3114246, EPI_ISL_3114367, EPI_ISL_3114418, EPI_ISL_3114429, EPI_ISL_3114457, EPI_ISL_3114523, EPI_ISL_3114605, EPI_ISL_3114659, EPI_ISL_3114692, EPI_ISL_3305191

| | | | |
|---|---|---|---|
| see above | Quest Diagnostics Incorporated | Centers for Disease Control and Prevention Division of Viral Diseases, Pathogen Discovery | A. Gerasimova; A. Perez; Adrian Paskey; B. Anderson; Ben L. Rambo-Martin; Benjamin Rambo-Martin; Christopher Gulvick; Clinton R. Paden; Dakota Howard; Darlene Wagner; Dhwani Batra; Duncan MacCannell; F. Lacbawan; I. A. Shlyakhter; Jason Caravas; K.E. Livingston; Kara Moser; L.E. Bernstein; M. Hua; Matthew Schmerer; P. Tanpaiboon; Peter W. Cook; R. M. Kagan; R. Owen; R. V. Rolando; S. H. Rosenthal; Scott Sammons; Shatavia Morrison; Suxiang Tong; Y. Liu; Yvette Unoarumhi |

EPI_ISL_1086672, EPI_ISL_1086675, EPI_ISL_1086692, EPI_ISL_1086734, EPI_ISL_1086751, EPI_ISL_1086757, EPI_ISL_1086789, EPI_ISL_1086791, EPI_ISL_1086812, EPI_ISL_1086821, EPI_ISL_1086848, EPI_ISL_1086878, EPI_ISL_1087042, EPI_ISL_1087053, EPI_ISL_1087070, EPI_ISL_1087078, EPI_ISL_1087089, EPI_ISL_1087090, EPI_ISL_1087102, EPI_ISL_1087438, EPI_ISL_1087440, EPI_ISL_1087640,
EPI_ISL_1087791, EPI_ISL_1087792, EPI_ISL_1087798, EPI_ISL_1087815, EPI_ISL_1087821, EPI_ISL_1087823, EPI_ISL_1087841, EPI_ISL_1087848, EPI_ISL_1087849, EPI_ISL_1087861, EPI_ISL_1087862, EPI_ISL_1087865, EPI_ISL_1087869, EPI_ISL_1087876, EPI_ISL_1087880, EPI_ISL_1087889, EPI_ISL_1090542, EPI_ISL_1090546, EPI_ISL_1090563, EPI_ISL_1090567, EPI_ISL_1090573, EPI_ISL_1090574,
EPI_ISL_1090576, EPI_ISL_1090577, EPI_ISL_1090586, EPI_ISL_1090596, EPI_ISL_1090598, EPI_ISL_1090601, EPI_ISL_1090602, EPI_ISL_1090603, EPI_ISL_1090613, EPI_ISL_1090614, EPI_ISL_1090621, EPI_ISL_1090898, EPI_ISL_1090905, EPI_ISL_1090918, EPI_ISL_1113909, EPI_ISL_1113912, EPI_ISL_1113948, EPI_ISL_1113955, EPI_ISL_1113958, EPI_ISL_1113959, EPI_ISL_1113986, EPI_ISL_1113993,
EPI_ISL_1114019, EPI_ISL_1114022, EPI_ISL_1114025, EPI_ISL_1139233, EPI_ISL_1139261, EPI_ISL_1139281, EPI_ISL_1139298, EPI_ISL_1139310, EPI_ISL_1139313, EPI_ISL_1139318, EPI_ISL_1139342, EPI_ISL_1139366, EPI_ISL_1139526, EPI_ISL_1139534, EPI_ISL_1139538, EPI_ISL_1139552, EPI_ISL_1139566, EPI_ISL_1139567, EPI_ISL_1139569, EPI_ISL_1139573, EPI_ISL_1139575, EPI_ISL_1139577,
EPI_ISL_1139579, EPI_ISL_1139581, EPI_ISL_1139587, EPI_ISL_1139622, EPI_ISL_1139702, EPI_ISL_1139704, EPI_ISL_1158937, EPI_ISL_1158943, EPI_ISL_1158945, EPI_ISL_1158960, EPI_ISL_1158972, EPI_ISL_1158974, EPI_ISL_1158978, EPI_ISL_1158982, EPI_ISL_1159002, EPI_ISL_1159003, EPI_ISL_1159005, EPI_ISL_1159011, EPI_ISL_1159022, EPI_ISL_1159028, EPI_ISL_1159032, EPI_ISL_1159038,
EPI_ISL_1159039, EPI_ISL_1159044, EPI_ISL_1159048, EPI_ISL_1159050, EPI_ISL_1159051, EPI_ISL_1159058, EPI_ISL_1159060, EPI_ISL_1159586, EPI_ISL_1159652, EPI_ISL_1159663

| | | | |
|---|---|---|---|
| see above | Quest Diagnostics Incorporated | Respiratory Viruses Branch, Division of Viral Diseases, Centers for Disease Control and Prevention | A. Gerasimova; A. Perez; B. Anderson; Ben L. Rambo-Martin; Clinton R. Paden; Dakota Howard; Dhwani Batra; Duncan MacCannell; F. Lacbawan; I. A. Shlyakhter; K.E. Livingston; L.E. Bernstein; M. Hua; P. Tanpaiboon; Peter W. Cook; R. M. Kagan; R. Owen; R. V. Rolando; S. H. Rosenthal; Suxiang Tong; Y. Liu |

EPI_ISL_884697, EPI_ISL_2254280, EPI_ISL_2254447, EPI_ISL_3091584, EPI_ISL_3091815, EPI_ISL_3091943, EPI_ISL_3091989, EPI_ISL_3091998, EPI_ISL_3092047

| | | | |
|---|---|---|---|
| see above | Respiratory Viruses Branch, Centers for Disease Control and Prevention | Respiratory Viruses Branch, Centers for Disease Control and Prevention | Allen, E.; Anderson, B.; Antico, J.; Ascencio, A.; B.L.; Barret; Batra, D.; Becker, D.; Bernstein, L.; Bolze, A.; C.R.; Campbell, P.; Caravas, J.; Case, R.; Cassens, T.; Cho, R.; Cirulli, E.; Clark, C.; Cook; Febbo, P.; Galloway, S.; Gerasimova, A.; Ghorpade, V.; Gietzen, K.; Gulvick, C.; Hardison, M.; Houdeshell, H.; Howard, D.; Hua, M.; I.A.; Isaksson, M.; K.S.; Kagan; Kvalvaag, O.; Lacbawan, F.; Laurent, M.; Lee, W.; Levan, G.; Liu, J.; Liu, Y.; Livingston, K.; Lu, J.; MacCannell, D.; Morrison, S.; Moser, K.; N.L.; Nall, D.; Nguyen, J.; Owen, R.; P.W.; Paden; Paskey, A.; Perez, A.; R.M.; R.V.; Rambo-Martin; Rambo-Martin, B.; Ramirez, J.; Rivera-Garcia, C.; Rolando; Rosenthal; S.H.; Sammons, S.; Sanders, E.; Sandoval, E.; Schmerer, M.; Shlyakhter; Sickler, B.; Tanpaiboon, P.; Tolentino, M.; Tong, S.; Tran, C.; Unoarumhi, Y.; Vest, A.; Wagner, D.; Wang, S.; Washington; Westlund, S.; White, S.; Wickline, S.; de Feo, E. |

| | | | |
|---|---|---|---|
| EPI_ISL_1378768, EPI_ISL_1378773, EPI_ISL_1378774 | Tempus | Grubaugh Lab - Yale School of Public Health | Anderson Brito; Annie Watkins; Chaney Kalinich; Chantal Vogels; Christopher E. Mason; Gaurav Khullar; Isabell Ott; Jessica Metti; Jessica Rothman; Joel T. Dudley; Joseph Fauver; Mallery Breban; Mary Petrone; Matthew J. MacKay; Megan Nash; Nathan Grubaugh; Nike Beaubier; Tara Alpert |

EPI_ISL_891137, EPI_ISL_891138, EPI_ISL_891139, EPI_ISL_891140, EPI_ISL_979366, EPI_ISL_979367, EPI_ISL_979368, EPI_ISL_979369, EPI_ISL_979370, EPI_ISL_979371, EPI_ISL_979372, EPI_ISL_979373, EPI_ISL_979374, EPI_ISL_979375, EPI_ISL_979376, EPI_ISL_979377, EPI_ISL_979378, EPI_ISL_1203819, EPI_ISL_1203820, EPI_ISL_1203821, EPI_ISL_1203822, EPI_ISL_1203823, EPI_ISL_1203824,
EPI_ISL_1203826, EPI_ISL_1203827, EPI_ISL_1203828, EPI_ISL_1203829, EPI_ISL_1203830, EPI_ISL_1203831, EPI_ISL_1203832, EPI_ISL_1203833, EPI_ISL_1203834, EPI_ISL_1203835, EPI_ISL_1203836, EPI_ISL_1203837, EPI_ISL_1203838, EPI_ISL_1203839, EPI_ISL_1203840, EPI_ISL_1203842, EPI_ISL_1273306, EPI_ISL_1273308, EPI_ISL_1273310, EPI_ISL_1273311, EPI_ISL_1273312, EPI_ISL_1273315,
EPI_ISL_1273316, EPI_ISL_1273317, EPI_ISL_1273318, EPI_ISL_1273319, EPI_ISL_1273320, EPI_ISL_1273321, EPI_ISL_1273322, EPI_ISL_1273323, EPI_ISL_1273324, EPI_ISL_1273325, EPI_ISL_1273326, EPI_ISL_1273327, EPI_ISL_1273328, EPI_ISL_1273329, EPI_ISL_1273330, EPI_ISL_1273331, EPI_ISL_1273332, EPI_ISL_1273334, EPI_ISL_1273335, EPI_ISL_1273336, EPI_ISL_1273339, EPI_ISL_1273341,
EPI_ISL_1273342, EPI_ISL_1273343, EPI_ISL_1273344, EPI_ISL_1273346, EPI_ISL_1273348, EPI_ISL_1273349, EPI_ISL_1273350, EPI_ISL_1273351, EPI_ISL_1273353, EPI_ISL_1273354, EPI_ISL_1273355, EPI_ISL_1273356, EPI_ISL_1273357, EPI_ISL_1273358, EPI_ISL_1273359, EPI_ISL_1273360, EPI_ISL_1273361, EPI_ISL_1273362, EPI_ISL_1273363, EPI_ISL_1273364, EPI_ISL_1273365, EPI_ISL_1273366,
EPI_ISL_1273367, EPI_ISL_1273368, EPI_ISL_1273369, EPI_ISL_1273370, EPI_ISL_1273371, EPI_ISL_1273374, EPI_ISL_1273375, EPI_ISL_1273376, EPI_ISL_1273378, EPI_ISL_1289785, EPI_ISL_1289786, EPI_ISL_1289787, EPI_ISL_1289788, EPI_ISL_1315126, EPI_ISL_1315127, EPI_ISL_1315128, EPI_ISL_1315129, EPI_ISL_1315130, EPI_ISL_1315131, EPI_ISL_1315132, EPI_ISL_1315133, EPI_ISL_1315134,
EPI_ISL_1315135, EPI_ISL_1315136, EPI_ISL_1315137, EPI_ISL_1315138, EPI_ISL_1315139, EPI_ISL_1315140, EPI_ISL_1315141, EPI_ISL_1315142, EPI_ISL_1315143, EPI_ISL_1315146, EPI_ISL_1315148, EPI_ISL_1315150, EPI_ISL_1315151, EPI_ISL_1315152, EPI_ISL_1315153, EPI_ISL_1315154, EPI_ISL_1315155, EPI_ISL_1315156, EPI_ISL_1315157, EPI_ISL_1315158, EPI_ISL_1315159, EPI_ISL_1315160,
EPI_ISL_1315161, EPI_ISL_1315162, EPI_ISL_1315163, EPI_ISL_1315164, EPI_ISL_1315165, EPI_ISL_1315166, EPI_ISL_1315167, EPI_ISL_1315168, EPI_ISL_1315169, EPI_ISL_1315170, EPI_ISL_1315171, EPI_ISL_1315173, EPI_ISL_1315175, EPI_ISL_1315176, EPI_ISL_1315177, EPI_ISL_1315178, EPI_ISL_1315179, EPI_ISL_1315180, EPI_ISL_1315181, EPI_ISL_1315182, EPI_ISL_1315183, EPI_ISL_1315184,
EPI_ISL_1315185, EPI_ISL_1315186, EPI_ISL_1315187, EPI_ISL_1315188, EPI_ISL_1315191, EPI_ISL_1315192, EPI_ISL_1315193, EPI_ISL_1315194, EPI_ISL_1315195, EPI_ISL_1315196, EPI_ISL_1315198, EPI_ISL_1315200, EPI_ISL_1315201, EPI_ISL_1315203, EPI_ISL_1315204, EPI_ISL_1315205, EPI_ISL_1315206, EPI_ISL_1315207, EPI_ISL_1315208, EPI_ISL_1315209, EPI_ISL_1315210, EPI_ISL_1315211,
EPI_ISL_1315212, EPI_ISL_1315213, EPI_ISL_1315214, EPI_ISL_1315215, EPI_ISL_1315216, EPI_ISL_1315220, EPI_ISL_1315221, EPI_ISL_1315223, EPI_ISL_1401474, EPI_ISL_1401475, EPI_ISL_1401476, EPI_ISL_1401480, EPI_ISL_1401482, EPI_ISL_1401485, EPI_ISL_1401486, EPI_ISL_1401487, EPI_ISL_1401488, EPI_ISL_1401489, EPI_ISL_1401491, EPI_ISL_1401493, EPI_ISL_1401494, EPI_ISL_1401495,
EPI_ISL_1401496, EPI_ISL_1401497, EPI_ISL_1401498, EPI_ISL_1401499, EPI_ISL_1401500, EPI_ISL_1401501, EPI_ISL_1401502, EPI_ISL_1401503, EPI_ISL_1401505, EPI_ISL_1401506, EPI_ISL_1401509, EPI_ISL_1401510, EPI_ISL_1401511, EPI_ISL_1401512, EPI_ISL_1401513, EPI_ISL_1401514, EPI_ISL_1401516, EPI_ISL_1401518, EPI_ISL_1401519, EPI_ISL_1401520, EPI_ISL_1401522, EPI_ISL_1401524,

EPI_ISL_1401525, EPI_ISL_1401526, EPI_ISL_1401527, EPI_ISL_1401528, EPI_ISL_1401529, EPI_ISL_1401531, EPI_ISL_1401532, EPI_ISL_1401533, EPI_ISL_1401534, EPI_ISL_1401535, EPI_ISL_1401536, EPI_ISL_1401537, EPI_ISL_1401538, EPI_ISL_1401539, EPI_ISL_1401540, EPI_ISL_1401541, EPI_ISL_1401543, EPI_ISL_1401544, EPI_ISL_1401545, EPI_ISL_1401546, EPI_ISL_1401547, EPI_ISL_1401548, EPI_ISL_1401549, EPI_ISL_1401551, EPI_ISL_1401552, EPI_ISL_1401554, EPI_ISL_1401555, EPI_ISL_1401556, EPI_ISL_1401557, EPI_ISL_1401558, EPI_ISL_1401562, EPI_ISL_1401563, EPI_ISL_1401564, EPI_ISL_1401565, EPI_ISL_1470561, EPI_ISL_1470563, EPI_ISL_1470565, EPI_ISL_1470566, EPI_ISL_1470568, EPI_ISL_1470569, EPI_ISL_1470570, EPI_ISL_1470571, EPI_ISL_1470572, EPI_ISL_1470573, EPI_ISL_1470574, EPI_ISL_1470576, EPI_ISL_1470577, EPI_ISL_1470578, EPI_ISL_1470579, EPI_ISL_1470580, EPI_ISL_1470581, EPI_ISL_1470582, EPI_ISL_1470583, EPI_ISL_1470584, EPI_ISL_1470585, EPI_ISL_1470586, EPI_ISL_1470588, EPI_ISL_1470589, EPI_ISL_1470591, EPI_ISL_1470592, EPI_ISL_1470593, EPI_ISL_1470594, EPI_ISL_1470595, EPI_ISL_1470596, EPI_ISL_1470597, EPI_ISL_1470598, EPI_ISL_1470600, EPI_ISL_1470601, EPI_ISL_1470602, EPI_ISL_1470603, EPI_ISL_1470604, EPI_ISL_1470605, EPI_ISL_1470606, EPI_ISL_1470607, EPI_ISL_1470609, EPI_ISL_1470611, EPI_ISL_1470613, EPI_ISL_1470614, EPI_ISL_1470615, EPI_ISL_1470616, EPI_ISL_1470617, EPI_ISL_1470618, EPI_ISL_1470619, EPI_ISL_1470620, EPI_ISL_1470621, EPI_ISL_1470622, EPI_ISL_1470623, EPI_ISL_1470624, EPI_ISL_1470625, EPI_ISL_1470627, EPI_ISL_1470629, EPI_ISL_1470630, EPI_ISL_1470632, EPI_ISL_1470633, EPI_ISL_1470634, EPI_ISL_1470635, EPI_ISL_1470638, EPI_ISL_1470639, EPI_ISL_1470640, EPI_ISL_1470641, EPI_ISL_1470642, EPI_ISL_1470643, EPI_ISL_1470644, EPI_ISL_1470645, EPI_ISL_1470646, EPI_ISL_1470647, EPI_ISL_1470649, EPI_ISL_1470650, EPI_ISL_1470652, EPI_ISL_1470653, EPI_ISL_1470655, EPI_ISL_1470656, EPI_ISL_1470657, EPI_ISL_1470658, EPI_ISL_1470659, EPI_ISL_1470660, EPI_ISL_1470661, EPI_ISL_1470663, EPI_ISL_1470664, EPI_ISL_1470665, EPI_ISL_1470666, EPI_ISL_1470667, EPI_ISL_1470668, EPI_ISL_1470669, EPI_ISL_1470670, EPI_ISL_1470671, EPI_ISL_1470673, EPI_ISL_1470674, EPI_ISL_1470675, EPI_ISL_1470678, EPI_ISL_1470679, EPI_ISL_1470680, EPI_ISL_1470682, EPI_ISL_1470684, EPI_ISL_1470685, EPI_ISL_1470687, EPI_ISL_1470689, EPI_ISL_1470690, EPI_ISL_1470691, EPI_ISL_1470692, EPI_ISL_1470694, EPI_ISL_1470695, EPI_ISL_1470696, EPI_ISL_1470697, EPI_ISL_1470698, EPI_ISL_1470702, EPI_ISL_1470703, EPI_ISL_1470705, EPI_ISL_1470706, EPI_ISL_1470709, EPI_ISL_1470710, EPI_ISL_1470711, EPI_ISL_1470712, EPI_ISL_1470713, EPI_ISL_1470714, EPI_ISL_1470715, EPI_ISL_1470716, EPI_ISL_1470717, EPI_ISL_1470719, EPI_ISL_1470720, EPI_ISL_1470721, EPI_ISL_1470722, EPI_ISL_1470723, EPI_ISL_1470724, EPI_ISL_1470725, EPI_ISL_1470726, EPI_ISL_1470727, EPI_ISL_1470728, EPI_ISL_1470729, EPI_ISL_1470730, EPI_ISL_1470731, EPI_ISL_1470732, EPI_ISL_1470733, EPI_ISL_1470734, EPI_ISL_1470735, EPI_ISL_1470736, EPI_ISL_1470737, EPI_ISL_1470739, EPI_ISL_1470740, EPI_ISL_1540714, EPI_ISL_1540715, EPI_ISL_1540716, EPI_ISL_1540717, EPI_ISL_1540719, EPI_ISL_1540720, EPI_ISL_1540721, EPI_ISL_1540724, EPI_ISL_1540725, EPI_ISL_1540726, EPI_ISL_1540728, EPI_ISL_1540729, EPI_ISL_1540730, EPI_ISL_1540731, EPI_ISL_1540733, EPI_ISL_1540735, EPI_ISL_1540736, EPI_ISL_1540737, EPI_ISL_1540738, EPI_ISL_1540739, EPI_ISL_1540740, EPI_ISL_1540741, EPI_ISL_1540742, EPI_ISL_1540743, EPI_ISL_1540744, EPI_ISL_1540745, EPI_ISL_1540746, EPI_ISL_1540748, EPI_ISL_1540749, EPI_ISL_1540750, EPI_ISL_1540752, EPI_ISL_1540753, EPI_ISL_1540754, EPI_ISL_1540755, EPI_ISL_1540756, EPI_ISL_1540758, EPI_ISL_1540762, EPI_ISL_1540763, EPI_ISL_1540764, EPI_ISL_1540767, EPI_ISL_1540769, EPI_ISL_1540770, EPI_ISL_1540771, EPI_ISL_1540772, EPI_ISL_1540773, EPI_ISL_1540776, EPI_ISL_1540778, EPI_ISL_1540779, EPI_ISL_1540780, EPI_ISL_1540781, EPI_ISL_1540782, EPI_ISL_1540783, EPI_ISL_1540784, EPI_ISL_1540785, EPI_ISL_1540786, EPI_ISL_1540787, EPI_ISL_1540788, EPI_ISL_1540789, EPI_ISL_1540791, EPI_ISL_1540792, EPI_ISL_1540793, EPI_ISL_1540794, EPI_ISL_1540795, EPI_ISL_1540796, EPI_ISL_1540797, EPI_ISL_1540798, EPI_ISL_1540799, EPI_ISL_1540800, EPI_ISL_1540802, EPI_ISL_1540803, EPI_ISL_1540804, EPI_ISL_1540807, EPI_ISL_1540808, EPI_ISL_1540809, EPI_ISL_1540810, EPI_ISL_1540811, EPI_ISL_1540812, EPI_ISL_1540813, EPI_ISL_1540814, EPI_ISL_1540815, EPI_ISL_1540816, EPI_ISL_1540817, EPI_ISL_1540818, EPI_ISL_1540819, EPI_ISL_1540820, EPI_ISL_1540821, EPI_ISL_1540822, EPI_ISL_1540823, EPI_ISL_1540824, EPI_ISL_1540826, EPI_ISL_1540830, EPI_ISL_1540833, EPI_ISL_1540834, EPI_ISL_1540835, EPI_ISL_1540838, EPI_ISL_1540839, EPI_ISL_1540840, EPI_ISL_1540841, EPI_ISL_1540842, EPI_ISL_1540843, EPI_ISL_1540844, EPI_ISL_1540845, EPI_ISL_1540846, EPI_ISL_1540847, EPI_ISL_1540850, EPI_ISL_1540852, EPI_ISL_1625540, EPI_ISL_1625541, EPI_ISL_1625542, EPI_ISL_1625543, EPI_ISL_1625545, EPI_ISL_1625546, EPI_ISL_1625547, EPI_ISL_1625548, EPI_ISL_1625549, EPI_ISL_1625550, EPI_ISL_1625551, EPI_ISL_1625552, EPI_ISL_1625553, EPI_ISL_1625554, EPI_ISL_1625555, EPI_ISL_1625556, EPI_ISL_1625557, EPI_ISL_1625558, EPI_ISL_1625559, EPI_ISL_1625560, EPI_ISL_1625561, EPI_ISL_1625562, EPI_ISL_1625563, EPI_ISL_1625564, EPI_ISL_1625565, EPI_ISL_1625566, EPI_ISL_1625567, EPI_ISL_1625568, EPI_ISL_1625569, EPI_ISL_1625570, EPI_ISL_1625571, EPI_ISL_1625572, EPI_ISL_1625573, EPI_ISL_1625574, EPI_ISL_1625575, EPI_ISL_1625576, EPI_ISL_1625577, EPI_ISL_1625578, EPI_ISL_1625579, EPI_ISL_1625580, EPI_ISL_1625581, EPI_ISL_1625583, EPI_ISL_1625584, EPI_ISL_1625585, EPI_ISL_1625586, EPI_ISL_1625587, EPI_ISL_1625588, EPI_ISL_1625589, EPI_ISL_1625590, EPI_ISL_1625591, EPI_ISL_1625592, EPI_ISL_1625593, EPI_ISL_1625594, EPI_ISL_1625595, EPI_ISL_1625596, EPI_ISL_1625597, EPI_ISL_1625598, EPI_ISL_1625599, EPI_ISL_1625600, EPI_ISL_1625601, EPI_ISL_1625602, EPI_ISL_1625603, EPI_ISL_1625604, EPI_ISL_1625605, EPI_ISL_1625606, EPI_ISL_1625607, EPI_ISL_1625608, EPI_ISL_1625609, EPI_ISL_1625610, EPI_ISL_1625611, EPI_ISL_1625612, EPI_ISL_1625613, EPI_ISL_1625614, EPI_ISL_1625615, EPI_ISL_1625626, EPI_ISL_1625627, EPI_ISL_1625628, EPI_ISL_1625629, EPI_ISL_1625630, EPI_ISL_1625631, EPI_ISL_1625632, EPI_ISL_1625633, EPI_ISL_1625634, EPI_ISL_1625641, EPI_ISL_1625643, EPI_ISL_1625644, EPI_ISL_1625645, EPI_ISL_1625646, EPI_ISL_1625647, EPI_ISL_1625648, EPI_ISL_1625649, EPI_ISL_1625650, EPI_ISL_1625651, EPI_ISL_1625652, EPI_ISL_1625653, EPI_ISL_1625654, EPI_ISL_1625655, EPI_ISL_1625656, EPI_ISL_1625657, EPI_ISL_1625658, EPI_ISL_1625659, EPI_ISL_1625660, EPI_ISL_1625661, EPI_ISL_1625662, EPI_ISL_1625663, EPI_ISL_1625664, EPI_ISL_1625665, EPI_ISL_1625666, EPI_ISL_1625667, EPI_ISL_1625668, EPI_ISL_1625669, EPI_ISL_1739426, EPI_ISL_1739427, EPI_ISL_1739428, EPI_ISL_1739429, EPI_ISL_1739430, EPI_ISL_1739431, EPI_ISL_1739433, EPI_ISL_1739436, EPI_ISL_1739437, EPI_ISL_1739438, EPI_ISL_1739439, EPI_ISL_1739443, EPI_ISL_1739444, EPI_ISL_1739445, EPI_ISL_1739447, EPI_ISL_1739449, EPI_ISL_1739450, EPI_ISL_1739451, EPI_ISL_1739452, EPI_ISL_1739453, EPI_ISL_1739454, EPI_ISL_1739456, EPI_ISL_1739457, EPI_ISL_1739458, EPI_ISL_1739459, EPI_ISL_1739460, EPI_ISL_1739462, EPI_ISL_1739463, EPI_ISL_1739465, EPI_ISL_1739466, EPI_ISL_1739467, EPI_ISL_1739468, EPI_ISL_1739469, EPI_ISL_1739471, EPI_ISL_1739472, EPI_ISL_1739473, EPI_ISL_1739474, EPI_ISL_1739475, EPI_ISL_1739476, EPI_ISL_1739477, EPI_ISL_1739478, EPI_ISL_1739479, EPI_ISL_1739480, EPI_ISL_1739481, EPI_ISL_1739482, EPI_ISL_1739483, EPI_ISL_1739484, EPI_ISL_1739486, EPI_ISL_1739487, EPI_ISL_1739488, EPI_ISL_1739489, EPI_ISL_1739490, EPI_ISL_1739491, EPI_ISL_1739493, EPI_ISL_1739494, EPI_ISL_1739496, EPI_ISL_1739497, EPI_ISL_1739498, EPI_ISL_1739499, EPI_ISL_1739500, EPI_ISL_1739502, EPI_ISL_1739503, EPI_ISL_1739504, EPI_ISL_1739505, EPI_ISL_1739506, EPI_ISL_1739507, EPI_ISL_1739508, EPI_ISL_1739509, EPI_ISL_1739510, EPI_ISL_1739511, EPI_ISL_1739514, EPI_ISL_1739515, EPI_ISL_1739516, EPI_ISL_1739517, EPI_ISL_1739518, EPI_ISL_1739519, EPI_ISL_1739520, EPI_ISL_1739521, EPI_ISL_1739522, EPI_ISL_1739523, EPI_ISL_1739524, EPI_ISL_1739525, EPI_ISL_1739526, EPI_ISL_1739527, EPI_ISL_1739528, EPI_ISL_1739529, EPI_ISL_1739530, EPI_ISL_1739531, EPI_ISL_1739532, EPI_ISL_1739533, EPI_ISL_1739534, EPI_ISL_1739535, EPI_ISL_1739536, EPI_ISL_1739537, EPI_ISL_1739538, EPI_ISL_1739539, EPI_ISL_1739540, EPI_ISL_1739541, EPI_ISL_1739542, EPI_ISL_1739543, EPI_ISL_1826473, EPI_ISL_1826479, EPI_ISL_1826482, EPI_ISL_1826484, EPI_ISL_1826485, EPI_ISL_1826486, EPI_ISL_1826487, EPI_ISL_1826488, EPI_ISL_1826489, EPI_ISL_1826490, EPI_ISL_1826491, EPI_ISL_1826492, EPI_ISL_1826493, EPI_ISL_1826494, EPI_ISL_1826495, EPI_ISL_1826496, EPI_ISL_1826497, EPI_ISL_1826498, EPI_ISL_1826499, EPI_ISL_1826500, EPI_ISL_1826501, EPI_ISL_1826502, EPI_ISL_1826504, EPI_ISL_1826505, EPI_ISL_1826507, EPI_ISL_1826510, EPI_ISL_1826512, EPI_ISL_1826513, EPI_ISL_1826514, EPI_ISL_1826516, EPI_ISL_1826517, EPI_ISL_1826520, EPI_ISL_1826521, EPI_ISL_1826524, EPI_ISL_1826525, EPI_ISL_1826526, EPI_ISL_1826527, EPI_ISL_1826529, EPI_ISL_1826530, EPI_ISL_1826531, EPI_ISL_1826533, EPI_ISL_1826534, EPI_ISL_1826535, EPI_ISL_1826536, EPI_ISL_1826538, EPI_ISL_1826540, EPI_ISL_1826541, EPI_ISL_1826542, EPI_ISL_1826543, EPI_ISL_1826544, EPI_ISL_1826545, EPI_ISL_1826546, EPI_ISL_1826547, EPI_ISL_1826549, EPI_ISL_1826550, EPI_ISL_1826552, EPI_ISL_1826553, EPI_ISL_1826554, EPI_ISL_1970631, EPI_ISL_1970633, EPI_ISL_1970634, EPI_ISL_1970637, EPI_ISL_1970638, EPI_ISL_1970639, EPI_ISL_1970640, EPI_ISL_1970641, EPI_ISL_1970642, EPI_ISL_1970643, EPI_ISL_1970644, EPI_ISL_1970645, EPI_ISL_1970646, EPI_ISL_1970648, EPI_ISL_1970649, EPI_ISL_1970650, EPI_ISL_1970651, EPI_ISL_1970652, EPI_ISL_1970653, EPI_ISL_1970654, EPI_ISL_1970655, EPI_ISL_1970656, EPI_ISL_1970657, EPI_ISL_1970659, EPI_ISL_1970660, EPI_ISL_1970662, EPI_ISL_1970663, EPI_ISL_1970664, EPI_ISL_1970665, EPI_ISL_1970666, EPI_ISL_1970667, EPI_ISL_1970668, EPI_ISL_1970669, EPI_ISL_1970670, EPI_ISL_1970671, EPI_ISL_1970672, EPI_ISL_1970673, EPI_ISL_1970674, EPI_ISL_1970675, EPI_ISL_1970676, EPI_ISL_1970677, EPI_ISL_1970678, EPI_ISL_1970679, EPI_ISL_1970680, EPI_ISL_1970681, EPI_ISL_1970682, EPI_ISL_1970683, EPI_ISL_1970684, EPI_ISL_1970685, EPI_ISL_1970686, EPI_ISL_2133345, EPI_ISL_2133346, EPI_ISL_2133349, EPI_ISL_2133352, EPI_ISL_2133353, EPI_ISL_2133354, EPI_ISL_2133355, EPI_ISL_2133358, EPI_ISL_2133360, EPI_ISL_2133361, EPI_ISL_2133362, EPI_ISL_2133363, EPI_ISL_2133364, EPI_ISL_2133365, EPI_ISL_2133366, EPI_ISL_2133367, EPI_ISL_2133368, EPI_ISL_2133369, EPI_ISL_2133370, EPI_ISL_2133371, EPI_ISL_2133372, EPI_ISL_2133373, EPI_ISL_2133375, EPI_ISL_2133376, EPI_ISL_2133377, EPI_ISL_2133378, EPI_ISL_2133379, EPI_ISL_2133380, EPI_ISL_2230023, EPI_ISL_2230024, EPI_ISL_2230025, EPI_ISL_2230026, EPI_ISL_2230027, EPI_ISL_2230028, EPI_ISL_2230029, EPI_ISL_2230030, EPI_ISL_2230031, EPI_ISL_2230032, EPI_ISL_2230033, EPI_ISL_2230034, EPI_ISL_2230035, EPI_ISL_2230037, EPI_ISL_2230038, EPI_ISL_2230039, EPI_ISL_2346323, EPI_ISL_2346324, EPI_ISL_2346325, EPI_ISL_2346326, EPI_ISL_2346327, EPI_ISL_2346328, EPI_ISL_2346329, EPI_ISL_2346330, EPI_ISL_2346331, EPI_ISL_2346332, EPI_ISL_2346333, EPI_ISL_2346334, EPI_ISL_2346335, EPI_ISL_2346336, EPI_ISL_2346338, EPI_ISL_2466704, EPI_ISL_2466707, EPI_ISL_2466708, EPI_ISL_2466709, EPI_ISL_2562123, EPI_ISL_2562124, EPI_ISL_2562125, EPI_ISL_2716209, EPI_ISL_2716210, EPI_ISL_2716211, EPI_ISL_2716212, EPI_ISL_2716213, EPI_ISL_2716214, EPI_ISL_2716215, EPI_ISL_2716216, EPI_ISL_2716220, EPI_ISL_3066877, EPI_ISL_3066878, EPI_ISL_3066879, EPI_ISL_3066881, EPI_ISL_3216755, EPI_ISL_3216756, EPI_ISL_3216757, EPI_ISL_3216758, EPI_ISL_3216759, EPI_ISL_3216760, EPI_ISL_3216761, EPI_ISL_3216762, EPI_ISL_3216763, EPI_ISL_3216764, EPI_ISL_3216765, EPI_ISL_3216766, EPI_ISL_3216767, EPI_ISL_3319120, EPI_ISL_3319121, EPI_ISL_3319122, EPI_ISL_3319123, EPI_ISL_3319124, EPI_ISL_3319125, EPI_ISL_3319126, EPI_ISL_3319127, EPI_ISL_3319128, EPI_ISL_3319129, EPI_ISL_3373118

| | | | |
|---|---|---|---|
| see above | The Jackson Laboratory | The Jackson Laboratory | Adams M; Bergeron D; Kelly K; Li L; Lloyd M; Long J; Maurya R; Omerza G; Renzette N; Sanderson B; Srivastava A; Uvalic J; Wei C L |
| EPI_ISL_2383323 | US Air Force School of Aerospace Medicine | US Air Force School of Aerospace Medicine | Amanda Javorina; Anthony Fries; Carol Garrett; Clarise Starr; Elizabeth Macias; Jennifer Meyer; Sarah Purves; William Gruner |

EPI_ISL_2382295, EPI_ISL_2382296, EPI_ISL_2382297, EPI_ISL_2382299, EPI_ISL_2382300, EPI_ISL_2382301, EPI_ISL_2382302, EPI_ISL_2382304, EPI_ISL_2382305

| | | | |
|---|---|---|---|
| see above | Uconn Health Clinical Microbiology | Brenton Graveley Lab, UConn Health | Brenton Graveley; Christina Caron; Kimberly Hayes; Lori Avery; Michael Senh; Sara Olson; Stacey Ferraro; Xintao Wei |

EPI_ISL_2283903, EPI_ISL_2283906, EPI_ISL_2283909, EPI_ISL_2283911, EPI_ISL_2283912, EPI_ISL_2283913, EPI_ISL_2283918, EPI_ISL_2283919, EPI_ISL_2283928, EPI_ISL_2283930, EPI_ISL_2283933, EPI_ISL_2283934, EPI_ISL_2283935, EPI_ISL_2283937, EPI_ISL_2283938, EPI_ISL_2283939, EPI_ISL_2283942, EPI_ISL_2283943, EPI_ISL_2283945, EPI_ISL_2283951, EPI_ISL_2283952, EPI_ISL_2283953, EPI_ISL_2283954, EPI_ISL_2283955, EPI_ISL_2283956, EPI_ISL_2283961, EPI_ISL_2283962, EPI_ISL_2283963, EPI_ISL_2283966, EPI_ISL_2283967, EPI_ISL_2283968, EPI_ISL_2283969, EPI_ISL_2283971, EPI_ISL_2283973, EPI_ISL_2283976, EPI_ISL_2283982, EPI_ISL_2283984, EPI_ISL_2283986, EPI_ISL_2283987, EPI_ISL_2284178, EPI_ISL_2284180, EPI_ISL_2284181, EPI_ISL_2284187, EPI_ISL_2284188, EPI_ISL_2284189, EPI_ISL_2284192, EPI_ISL_2284196, EPI_ISL_2284198

| | | | |
|---|---|---|---|
| see above | VA Connecticut Healthcare System | Yale Center for Genomic Analysis | Brooke Sullivan; Curt Scharfe; Irina Tikhonova; Kaya Bilguvar; Shrikant Mane |

EPI_ISL_861750, EPI_ISL_861752, EPI_ISL_861753, EPI_ISL_861771, EPI_ISL_912160, EPI_ISL_1038982, EPI_ISL_1038983, EPI_ISL_1038984, EPI_ISL_1038985, EPI_ISL_1038986, EPI_ISL_1038987, EPI_ISL_1038988, EPI_ISL_1038989, EPI_ISL_1038990, EPI_ISL_1038991, EPI_ISL_1038992, EPI_ISL_1038993, EPI_ISL_1163667, EPI_ISL_1163668, EPI_ISL_1163669, EPI_ISL_1163670, EPI_ISL_1163671, EPI_ISL_1163672, EPI_ISL_1163673, EPI_ISL_1163674, EPI_ISL_1163675, EPI_ISL_1163676, EPI_ISL_1163677, EPI_ISL_1163678, EPI_ISL_1163679, EPI_ISL_1163680, EPI_ISL_1163681, EPI_ISL_1163682, EPI_ISL_1163683, EPI_ISL_1163684, EPI_ISL_1163685, EPI_ISL_1163686, EPI_ISL_1163687, EPI_ISL_1182015, EPI_ISL_1182016, EPI_ISL_1182017, EPI_ISL_1182018, EPI_ISL_1182019, EPI_ISL_1182020, EPI_ISL_1182021, EPI_ISL_1182022, EPI_ISL_1182023, EPI_ISL_1182024, EPI_ISL_1182025, EPI_ISL_1182026, EPI_ISL_1225584, EPI_ISL_1225585, EPI_ISL_1225586, EPI_ISL_1225587, EPI_ISL_1225588, EPI_ISL_1225589, EPI_ISL_1225590, EPI_ISL_1225591, EPI_ISL_1225592, EPI_ISL_1225594, EPI_ISL_1225596, EPI_ISL_1225597, EPI_ISL_1225598, EPI_ISL_1225601, EPI_ISL_1225602, EPI_ISL_1225603, EPI_ISL_1225607, EPI_ISL_1225608, EPI_ISL_1225609, EPI_ISL_1225610, EPI_ISL_1225615, EPI_ISL_1225616, EPI_ISL_1225617, EPI_ISL_1225620, EPI_ISL_1225621, EPI_ISL_1225623, EPI_ISL_1225624, EPI_ISL_1225648, EPI_ISL_1225649, EPI_ISL_1225650, EPI_ISL_1225651, EPI_ISL_1225652, EPI_ISL_1225653, EPI_ISL_1225658, EPI_ISL_1225659, EPI_ISL_1225660, EPI_ISL_1225663, EPI_ISL_1225665, EPI_ISL_1225668, EPI_ISL_1225670, EPI_ISL_1225672, EPI_ISL_1225674, EPI_ISL_1225679, EPI_ISL_1225680, EPI_ISL_1225681, EPI_ISL_1225683, EPI_ISL_1225684, EPI_ISL_1225685, EPI_ISL_1225686, EPI_ISL_1225687, EPI_ISL_1225689, EPI_ISL_1225690, EPI_ISL_1225691, EPI_ISL_1225692, EPI_ISL_1225693, EPI_ISL_1225694, EPI_ISL_1225696, EPI_ISL_1225697, EPI_ISL_1225698, EPI_ISL_1225700, EPI_ISL_1225702, EPI_ISL_1225703, EPI_ISL_1225704, EPI_ISL_1225705, EPI_ISL_1225708, EPI_ISL_1225709, EPI_ISL_1225710, EPI_ISL_1225712, EPI_ISL_1225714, EPI_ISL_1225716, EPI_ISL_1225721, EPI_ISL_1225724, EPI_ISL_1225725, EPI_ISL_1225726, EPI_ISL_1225729, EPI_ISL_1225730, EPI_ISL_1225731, EPI_ISL_1225732, EPI_ISL_1225733, EPI_ISL_1225734, EPI_ISL_1225735, EPI_ISL_1292979, EPI_ISL_1292980, EPI_ISL_1292981, EPI_ISL_1292982, EPI_ISL_1292983, EPI_ISL_1292984, EPI_ISL_1292986, EPI_ISL_1292987, EPI_ISL_1292988, EPI_ISL_1292989, EPI_ISL_1292990, EPI_ISL_1292991, EPI_ISL_1292993, EPI_ISL_1292994, EPI_ISL_1292997, EPI_ISL_1293149, EPI_ISL_1293150, EPI_ISL_1293151, EPI_ISL_1293152, EPI_ISL_1293153, EPI_ISL_1293154, EPI_ISL_1293155, EPI_ISL_1293156, EPI_ISL_1293157, EPI_ISL_1293158, EPI_ISL_1293159, EPI_ISL_1293160, EPI_ISL_1293161, EPI_ISL_1293162, EPI_ISL_1293163, EPI_ISL_1293164, EPI_ISL_1293165, EPI_ISL_1293166, EPI_ISL_1293167, EPI_ISL_1293168, EPI_ISL_1293169, EPI_ISL_1293170, EPI_ISL_1293171, EPI_ISL_1293172, EPI_ISL_1293173, EPI_ISL_1293174, EPI_ISL_1293175, EPI_ISL_1293176, EPI_ISL_1293177, EPI_ISL_1293179, EPI_ISL_1293180, EPI_ISL_1293181, EPI_ISL_1293182, EPI_ISL_1293183, EPI_ISL_1293184, EPI_ISL_1293185, EPI_ISL_1293186, EPI_ISL_1293187, EPI_ISL_1293188, EPI_ISL_1293189, EPI_ISL_1293190, EPI_ISL_1293191, EPI_ISL_1293192, EPI_ISL_1293193, EPI_ISL_1293194, EPI_ISL_1293195, EPI_ISL_1293196, EPI_ISL_1293197, EPI_ISL_1293198, EPI_ISL_1293199, EPI_ISL_1293200, EPI_ISL_1293201, EPI_ISL_1293202, EPI_ISL_1293203, EPI_ISL_1293204, EPI_ISL_1293205, EPI_ISL_1293206, EPI_ISL_1293207, EPI_ISL_1293208, EPI_ISL_1293209, EPI_ISL_1293210, EPI_ISL_1293211, EPI_ISL_1293212, EPI_ISL_1293213, EPI_ISL_1293214, EPI_ISL_1293215, EPI_ISL_1293216, EPI_ISL_1293219, EPI_ISL_1293220, EPI_ISL_1293221, EPI_ISL_1293222, EPI_ISL_1293223, EPI_ISL_1293224, EPI_ISL_1293225, EPI_ISL_1293226, EPI_ISL_1293227, EPI_ISL_1293228, EPI_ISL_1293229, EPI_ISL_1293230, EPI_ISL_1293231, EPI_ISL_1293232, EPI_ISL_1293233, EPI_ISL_1293234, EPI_ISL_1293235, EPI_ISL_1378673, EPI_ISL_1378674, EPI_ISL_1378675, EPI_ISL_1378676, EPI_ISL_1378677, EPI_ISL_1378678, EPI_ISL_1378679, EPI_ISL_1378680, EPI_ISL_1378681, EPI_ISL_1378682, EPI_ISL_1378683, EPI_ISL_1378684, EPI_ISL_1378685, EPI_ISL_1378686, EPI_ISL_1378688, EPI_ISL_1378689, EPI_ISL_1378690, EPI_ISL_1378691, EPI_ISL_1378692, EPI_ISL_1378693, EPI_ISL_1378697, EPI_ISL_1378698, EPI_ISL_1378699, EPI_ISL_1378700, EPI_ISL_1378701, EPI_ISL_1378702, EPI_ISL_1378703, EPI_ISL_1378704, EPI_ISL_1378705, EPI_ISL_1378706, EPI_ISL_1378707, EPI_ISL_1378708, EPI_ISL_1378709, EPI_ISL_1378710, EPI_ISL_1378711, EPI_ISL_1378712, EPI_ISL_1378713, EPI_ISL_1378714, EPI_ISL_1378715, EPI_ISL_1378716, EPI_ISL_1378717, EPI_ISL_1378718, EPI_ISL_1378719, EPI_ISL_1378721, EPI_ISL_1378722, EPI_ISL_1378723, EPI_ISL_1378725, EPI_ISL_1378727, EPI_ISL_1378729, EPI_ISL_1378730, EPI_ISL_1378731, EPI_ISL_1378732, EPI_ISL_1378733, EPI_ISL_1378734, EPI_ISL_1378735, EPI_ISL_1378736, EPI_ISL_1378737, EPI_ISL_1378739, EPI_ISL_1378740, EPI_ISL_1378742, EPI_ISL_1378743, EPI_ISL_1378744, EPI_ISL_1378745, EPI_ISL_1378746, EPI_ISL_1378747, EPI_ISL_1378748, EPI_ISL_1378749, EPI_ISL_1378750, EPI_ISL_1378751, EPI_ISL_1378754, EPI_ISL_1378755, EPI_ISL_1378756, EPI_ISL_1378757, EPI_ISL_1378758, EPI_ISL_1378759, EPI_ISL_1378760, EPI_ISL_1378761, EPI_ISL_1378762, EPI_ISL_1378763, EPI_ISL_1447510, EPI_ISL_1447511, EPI_ISL_1447512, EPI_ISL_1447513, EPI_ISL_1447515, EPI_ISL_1447516, EPI_ISL_1447517, EPI_ISL_1447518, EPI_ISL_1447519, EPI_ISL_1447521, EPI_ISL_1447522, EPI_ISL_1447523, EPI_ISL_1447524, EPI_ISL_1447525, EPI_ISL_1447526, EPI_ISL_1447527, EPI_ISL_1447528, EPI_ISL_1447529, EPI_ISL_1447530, EPI_ISL_1447532, EPI_ISL_1447533, EPI_ISL_1447534, EPI_ISL_1447535, EPI_ISL_1447536, EPI_ISL_1447537, EPI_ISL_1447538, EPI_ISL_1447539, EPI_ISL_1447540, EPI_ISL_1447541, EPI_ISL_1447542, EPI_ISL_1447543, EPI_ISL_1447544, EPI_ISL_1447545, EPI_ISL_1447546, EPI_ISL_1447547, EPI_ISL_1447548, EPI_ISL_1447549, EPI_ISL_1447550, EPI_ISL_1447552, EPI_ISL_1447553, EPI_ISL_1447557, EPI_ISL_1447558, EPI_ISL_1447559, EPI_ISL_1447560, EPI_ISL_1447561, EPI_ISL_1447562, EPI_ISL_1447564, EPI_ISL_1447565, EPI_ISL_1447566, EPI_ISL_1447567, EPI_ISL_1447568, EPI_ISL_1447569, EPI_ISL_1447570, EPI_ISL_1447571, EPI_ISL_1447573, EPI_ISL_1447576, EPI_ISL_1447578, EPI_ISL_1447580, EPI_ISL_1447581, EPI_ISL_1447582, EPI_ISL_1447583, EPI_ISL_1447584, EPI_ISL_1447585, EPI_ISL_1447586, EPI_ISL_1447587, EPI_ISL_1447589, EPI_ISL_1447590, EPI_ISL_1447591, EPI_ISL_1447592, EPI_ISL_1447593, EPI_ISL_1447594, EPI_ISL_1447595, EPI_ISL_1447596, EPI_ISL_1447597, EPI_ISL_1447598, EPI_ISL_1447599, EPI_ISL_1447600, EPI_ISL_1447602, EPI_ISL_1447603, EPI_ISL_1447604, EPI_ISL_1447605, EPI_ISL_1447606, EPI_ISL_1447607, EPI_ISL_1447608, EPI_ISL_1447611, EPI_ISL_1447612, EPI_ISL_1447613, EPI_ISL_1447614, EPI_ISL_1447616, EPI_ISL_1447617, EPI_ISL_1447618, EPI_ISL_1447619, EPI_ISL_1447620, EPI_ISL_1447621, EPI_ISL_1447623, EPI_ISL_1447624, EPI_ISL_1447625, EPI_ISL_1447627, EPI_ISL_1447628, EPI_ISL_1447629, EPI_ISL_1447630, EPI_ISL_1447631, EPI_ISL_1447634, EPI_ISL_1447635, EPI_ISL_1447636, EPI_ISL_1447637, EPI_ISL_1447638, EPI_ISL_1447639, EPI_ISL_1447640, EPI_ISL_1447641, EPI_ISL_1447642, EPI_ISL_1447643, EPI_ISL_1447644, EPI_ISL_1447645, EPI_ISL_1447646, EPI_ISL_1447647, EPI_ISL_1447648, EPI_ISL_1447649, EPI_ISL_1447650, EPI_ISL_1447651, EPI_ISL_1447652, EPI_ISL_1447653, EPI_ISL_1447656, EPI_ISL_1447657, EPI_ISL_1447659, EPI_ISL_1447660, EPI_ISL_1447661, EPI_ISL_1447662, EPI_ISL_1447663, EPI_ISL_1447664, EPI_ISL_1447665, EPI_ISL_1447666, EPI_ISL_1447667, EPI_ISL_1447668, EPI_ISL_1447669, EPI_ISL_1447670, EPI_ISL_1509103, EPI_ISL_1509104, EPI_ISL_1509107, EPI_ISL_1509108, EPI_ISL_1509109, EPI_ISL_1509111, EPI_ISL_1509112, EPI_ISL_1509113, EPI_ISL_1509115, EPI_ISL_1509116, EPI_ISL_1509117, EPI_ISL_1509118, EPI_ISL_1509119, EPI_ISL_1509120, EPI_ISL_1509121, EPI_ISL_1509123, EPI_ISL_1509124, EPI_ISL_1509125, EPI_ISL_1509126, EPI_ISL_1509127, EPI_ISL_1509128, EPI_ISL_1509129, EPI_ISL_1509130, EPI_ISL_1509131, EPI_ISL_1509132, EPI_ISL_1509133, EPI_ISL_1509134, EPI_ISL_1509135, EPI_ISL_1509136, EPI_ISL_1509137, EPI_ISL_1509138, EPI_ISL_1509139, EPI_ISL_1509141, EPI_ISL_1509142, EPI_ISL_1509143, EPI_ISL_1509144, EPI_ISL_1509145, EPI_ISL_1509146, EPI_ISL_1509149, EPI_ISL_1509151, EPI_ISL_1509152, EPI_ISL_1509153, EPI_ISL_1509154, EPI_ISL_1509155, EPI_ISL_1509156, EPI_ISL_1509157, EPI_ISL_1509158, EPI_ISL_1509159, EPI_ISL_1509160, EPI_ISL_1509161, EPI_ISL_1509163, EPI_ISL_1509164, EPI_ISL_1509165, EPI_ISL_1509166, EPI_ISL_1509167, EPI_ISL_1509168, EPI_ISL_1509169, EPI_ISL_1509170, EPI_ISL_1509171, EPI_ISL_1509172, EPI_ISL_1509173, EPI_ISL_1509174, EPI_ISL_1509175, EPI_ISL_1509176, EPI_ISL_1509177, EPI_ISL_1509178, EPI_ISL_1509179, EPI_ISL_1509181, EPI_ISL_1509182, EPI_ISL_1509183, EPI_ISL_1509184, EPI_ISL_1509185, EPI_ISL_1509186, EPI_ISL_1509187, EPI_ISL_1509188, EPI_ISL_1509190, EPI_ISL_1509191, EPI_ISL_1509192, EPI_ISL_1509193, EPI_ISL_1509194, EPI_ISL_1509195, EPI_ISL_1509196, EPI_ISL_1509197, EPI_ISL_1509201, EPI_ISL_1509202, EPI_ISL_1509204, EPI_ISL_1509205, EPI_ISL_1509206, EPI_ISL_1509207, EPI_ISL_1509208, EPI_ISL_1509209, EPI_ISL_1509210, EPI_ISL_1509211, EPI_ISL_1509212, EPI_ISL_1509213, EPI_ISL_1509214, EPI_ISL_1509215, EPI_ISL_1509216, EPI_ISL_1509217, EPI_ISL_1509218, EPI_ISL_1509219, EPI_ISL_1509220, EPI_ISL_1509221, EPI_ISL_1509222, EPI_ISL_1509223, EPI_ISL_1509224, EPI_ISL_1509225, EPI_ISL_1509226, EPI_ISL_1509227, EPI_ISL_1509228, EPI_ISL_1509229, EPI_ISL_1509230, EPI_ISL_1509231, EPI_ISL_1509233, EPI_ISL_1509234, EPI_ISL_1509235, EPI_ISL_1509236, EPI_ISL_1509237, EPI_ISL_1509239, EPI_ISL_1509240, EPI_ISL_1509241, EPI_ISL_1509242, EPI_ISL_1509243, EPI_ISL_1509244, EPI_ISL_1509245, EPI_ISL_1509246, EPI_ISL_1509247, EPI_ISL_1509248, EPI_ISL_1509249, EPI_ISL_1509250, EPI_ISL_1509251, EPI_ISL_1509252, EPI_ISL_1509253, EPI_ISL_1509255, EPI_ISL_1509256, EPI_ISL_1509257, EPI_ISL_1509258, EPI_ISL_1509259, EPI_ISL_1509260, EPI_ISL_1509261, EPI_ISL_1587258, EPI_ISL_1587259, EPI_ISL_1587260, EPI_ISL_1587261, EPI_ISL_1587262, EPI_ISL_1587263, EPI_ISL_1587264, EPI_ISL_1587265, EPI_ISL_1587266, EPI_ISL_1587268, EPI_ISL_1587269, EPI_ISL_1587271, EPI_ISL_1587272, EPI_ISL_1587273, EPI_ISL_1587275, EPI_ISL_1587276, EPI_ISL_1587277, EPI_ISL_1587279, EPI_ISL_1587280, EPI_ISL_1587281, EPI_ISL_1587283, EPI_ISL_1587284, EPI_ISL_1587286, EPI_ISL_1587287, EPI_ISL_1587288, EPI_ISL_1587289, EPI_ISL_1587290, EPI_ISL_1587291, EPI_ISL_1587292, EPI_ISL_1587293, EPI_ISL_1587294, EPI_ISL_1587295, EPI_ISL_1587296, EPI_ISL_1587297, EPI_ISL_1587298, EPI_ISL_1587299, EPI_ISL_1587300, EPI_ISL_1587301, EPI_ISL_1587302, EPI_ISL_1587303, EPI_ISL_1587305, EPI_ISL_1587306, EPI_ISL_1587307, EPI_ISL_1587310, EPI_ISL_1587311, EPI_ISL_1587312, EPI_ISL_1587313, EPI_ISL_1587314, EPI_ISL_1587315, EPI_ISL_1587316, EPI_ISL_1587317, EPI_ISL_1587318, EPI_ISL_1587319, EPI_ISL_1587320, EPI_ISL_1587321, EPI_ISL_1587322, EPI_ISL_1587323, EPI_ISL_1587324, EPI_ISL_1587325, EPI_ISL_1587326, EPI_ISL_1587327, EPI_ISL_1587328, EPI_ISL_1587329, EPI_ISL_1587330, EPI_ISL_1587331, EPI_ISL_1587332, EPI_ISL_1587333, EPI_ISL_1587335, EPI_ISL_1587336, EPI_ISL_1587337, EPI_ISL_1587338, EPI_ISL_1587340, EPI_ISL_1587341, EPI_ISL_1587344, EPI_ISL_1587345, EPI_ISL_1587346, EPI_ISL_1587348, EPI_ISL_1587349, EPI_ISL_1587350, EPI_ISL_1587351, EPI_ISL_1587352, EPI_ISL_1587353, EPI_ISL_1587355, EPI_ISL_1587356, EPI_ISL_1587359, EPI_ISL_1587360, EPI_ISL_1587361, EPI_ISL_1587362, EPI_ISL_1587363, EPI_ISL_1587364, EPI_ISL_1587365, EPI_ISL_1587366, EPI_ISL_1587367, EPI_ISL_1587368, EPI_ISL_1587369, EPI_ISL_1587370, EPI_ISL_1587371, EPI_ISL_1587372, EPI_ISL_1587373, EPI_ISL_1587374, EPI_ISL_1587375, EPI_ISL_1587376, EPI_ISL_1587377, EPI_ISL_1587378, EPI_ISL_1587379, EPI_ISL_1587380, EPI_ISL_1587381, EPI_ISL_1587384, EPI_ISL_1587385, EPI_ISL_1587386, EPI_ISL_1587387, EPI_ISL_1587388, EPI_ISL_1587389, EPI_ISL_1587391, EPI_ISL_1587392, EPI_ISL_1587393, EPI_ISL_1587394, EPI_ISL_1587396, EPI_ISL_1587397, EPI_ISL_1587399, EPI_ISL_1587400, EPI_ISL_1587401, EPI_ISL_1587402, EPI_ISL_1587403, EPI_ISL_1587404, EPI_ISL_1587405, EPI_ISL_1587406, EPI_ISL_1587407, EPI_ISL_1587408, EPI_ISL_1587410, EPI_ISL_1587412, EPI_ISL_1587413, EPI_ISL_1587414, EPI_ISL_1587416, EPI_ISL_1587417, EPI_ISL_1587418, EPI_ISL_1587419, EPI_ISL_1587420, EPI_ISL_1587422, EPI_ISL_1587425, EPI_ISL_1674535, EPI_ISL_1674536, EPI_ISL_1674537, EPI_ISL_1674538, EPI_ISL_1674539, EPI_ISL_1674540, EPI_ISL_1674541, EPI_ISL_1674542, EPI_ISL_1674543, EPI_ISL_1674544, EPI_ISL_1674545, EPI_ISL_1674546, EPI_ISL_1674547, EPI_ISL_1674548, EPI_ISL_1674549, EPI_ISL_1674550, EPI_ISL_1674551, EPI_ISL_1674552, EPI_ISL_1674553, EPI_ISL_1674554, EPI_ISL_1674555, EPI_ISL_1674556, EPI_ISL_1674557, EPI_ISL_1674558, EPI_ISL_1674559, EPI_ISL_1674560, EPI_ISL_1674561, EPI_ISL_1674562, EPI_ISL_1674563, EPI_ISL_1674564, EPI_ISL_1674565, EPI_ISL_1674566, EPI_ISL_1674567, EPI_ISL_1674568, EPI_ISL_1674569, EPI_ISL_1674570, EPI_ISL_1674571, EPI_ISL_1674572, EPI_ISL_1674573, EPI_ISL_1674574, EPI_ISL_1674575, EPI_ISL_1674576, EPI_ISL_1674577, EPI_ISL_1674578, EPI_ISL_1674579, EPI_ISL_1674580, EPI_ISL_1674581, EPI_ISL_1674582, EPI_ISL_1674583, EPI_ISL_1674584, EPI_ISL_1674586, EPI_ISL_1674587, EPI_ISL_1674588, EPI_ISL_1674589, EPI_ISL_1674590, EPI_ISL_1674591, EPI_ISL_1674592, EPI_ISL_1674593, EPI_ISL_1674594, EPI_ISL_1674595, EPI_ISL_1674596, EPI_ISL_1674597, EPI_ISL_1674598, EPI_ISL_1674599, EPI_ISL_1674600, EPI_ISL_1674601, EPI_ISL_1674604, EPI_ISL_1674605, EPI_ISL_1674606, EPI_ISL_1674607, EPI_ISL_1674608, EPI_ISL_1674609, EPI_ISL_1674611, EPI_ISL_1674612, EPI_ISL_1674613, EPI_ISL_1674614, EPI_ISL_1674615, EPI_ISL_1674616, EPI_ISL_1674617, EPI_ISL_1674618, EPI_ISL_1674619, EPI_ISL_1674620, EPI_ISL_1674621, EPI_ISL_1674622, EPI_ISL_1674623, EPI_ISL_1674624, EPI_ISL_1674625, EPI_ISL_1674626, EPI_ISL_1674627, EPI_ISL_1674628, EPI_ISL_1674629, EPI_ISL_1674630, EPI_ISL_1674631, EPI_ISL_1674632, EPI_ISL_1674633, EPI_ISL_1674638, EPI_ISL_1674639, EPI_ISL_1674640, EPI_ISL_1674641, EPI_ISL_1674642, EPI_ISL_1674643, EPI_ISL_1674644, EPI_ISL_1674645, EPI_ISL_1674646, EPI_ISL_1674647, EPI_ISL_1674648, EPI_ISL_1674649, EPI_ISL_1674650, EPI_ISL_1674651, EPI_ISL_1674652, EPI_ISL_1674655, EPI_ISL_1674656, EPI_ISL_1674658, EPI_ISL_1674659, EPI_ISL_1674660, EPI_ISL_1674662, EPI_ISL_1674663, EPI_ISL_1674664, EPI_ISL_1674665, EPI_ISL_1674666, EPI_ISL_1674667, EPI_ISL_1674668, EPI_ISL_1674669, EPI_ISL_1674670, EPI_ISL_1674671, EPI_ISL_1674672, EPI_ISL_1674673, EPI_ISL_1674674, EPI_ISL_1674675, EPI_ISL_1674676, EPI_ISL_1674678, EPI_ISL_1674679, EPI_ISL_1674680, EPI_ISL_1674681, EPI_ISL_1674682, EPI_ISL_1674683, EPI_ISL_1674684, EPI_ISL_1674685, EPI_ISL_1674686, EPI_ISL_1674687, EPI_ISL_1674688, EPI_ISL_1674689, EPI_ISL_1674690, EPI_ISL_1674691, EPI_ISL_1674692, EPI_ISL_1674693, EPI_ISL_1674694, EPI_ISL_1674695, EPI_ISL_1674697, EPI_ISL_1792929, EPI_ISL_1792930, EPI_ISL_1792931, EPI_ISL_1792932, EPI_ISL_1792933, EPI_ISL_1792934, EPI_ISL_1792935, EPI_ISL_1792936, EPI_ISL_1792937, EPI_ISL_1792938, EPI_ISL_1792939, EPI_ISL_1792940, EPI_ISL_1792941, EPI_ISL_1792942, EPI_ISL_1792943, EPI_ISL_1792946, EPI_ISL_1792947, EPI_ISL_1792949, EPI_ISL_1792951, EPI_ISL_1792952, EPI_ISL_1792953, EPI_ISL_1792954, EPI_ISL_1792955, EPI_ISL_1792956, EPI_ISL_1792957, EPI_ISL_1792958, EPI_ISL_1792959, EPI_ISL_1792960, EPI_ISL_1792961, EPI_ISL_1792962, EPI_ISL_1792964, EPI_ISL_1792965, EPI_ISL_1792966, EPI_ISL_1792967, EPI_ISL_1792968, EPI_ISL_1792969, EPI_ISL_1792970, EPI_ISL_1792971, EPI_ISL_1792972, EPI_ISL_1792973, EPI_ISL_1792974, EPI_ISL_1792976, EPI_ISL_1792977, EPI_ISL_1792978, EPI_ISL_1792979, EPI_ISL_1792980, EPI_ISL_1792981, EPI_ISL_1792982, EPI_ISL_1792983, EPI_ISL_1792984, EPI_ISL_1792985, EPI_ISL_1792986, EPI_ISL_1792988, EPI_ISL_1792991, EPI_ISL_1792992, EPI_ISL_1792994, EPI_ISL_1792996, EPI_ISL_1792997, EPI_ISL_1792998, EPI_ISL_1792999, EPI_ISL_1793000, EPI_ISL_1793001, EPI_ISL_1793003, EPI_ISL_1793004, EPI_ISL_1793005, EPI_ISL_1793006, EPI_ISL_1793007, EPI_ISL_1793008, EPI_ISL_1793009, EPI_ISL_1793010, EPI_ISL_1793011, EPI_ISL_1793013, EPI_ISL_1793015, EPI_ISL_1793016, EPI_ISL_1793017, EPI_ISL_1793018, EPI_ISL_1793019, EPI_ISL_1793020, EPI_ISL_1793021, EPI_ISL_1793022, EPI_ISL_1793023, EPI_ISL_1793024, EPI_ISL_1793025, EPI_ISL_1793026, EPI_ISL_1793027, EPI_ISL_1793028, EPI_ISL_1793031, EPI_ISL_1793033, EPI_ISL_1793035, EPI_ISL_1793037, EPI_ISL_1793038, EPI_ISL_1793039, EPI_ISL_1793041, EPI_ISL_1793042, EPI_ISL_1793044, EPI_ISL_1793045, EPI_ISL_1793046, EPI_ISL_1793047, EPI_ISL_1793048, EPI_ISL_1793049, EPI_ISL_1793051, EPI_ISL_1793052, EPI_ISL_1793054, EPI_ISL_1793056, EPI_ISL_1793057, EPI_ISL_1793058, EPI_ISL_1793059, EPI_ISL_1793060, EPI_ISL_1793061, EPI_ISL_1793062, EPI_ISL_1793063, EPI_ISL_1793065, EPI_ISL_1793066, EPI_ISL_1793067, EPI_ISL_1793068, EPI_ISL_1793070, EPI_ISL_1793071, EPI_ISL_1793072, EPI_ISL_1793073, EPI_ISL_1793075, EPI_ISL_1793076, EPI_ISL_1793077, EPI_ISL_1793079, EPI_ISL_1793080, EPI_ISL_1793082, EPI_ISL_1793083, EPI_ISL_1793084, EPI_ISL_1793085, EPI_ISL_1793087, EPI_ISL_1793088, EPI_ISL_1793089, EPI_ISL_1793090, EPI_ISL_1793091, EPI_ISL_1793092, EPI_ISL_1793093, EPI_ISL_1793094, EPI_ISL_1793095, EPI_ISL_1963645, EPI_ISL_1963646, EPI_ISL_1963647, EPI_ISL_1963648, EPI_ISL_1963649, EPI_ISL_1963650, EPI_ISL_1963651, EPI_ISL_1963652, EPI_ISL_1963653, EPI_ISL_1963654, EPI_ISL_1963655, EPI_ISL_1963656, EPI_ISL_1963657, EPI_ISL_1963658, EPI_ISL_1963659, EPI_ISL_1963660, EPI_ISL_1963662, EPI_ISL_1963663, EPI_ISL_1963664, EPI_ISL_1963665, EPI_ISL_1963666, EPI_ISL_1963668, EPI_ISL_1963669, EPI_ISL_1963670, EPI_ISL_1963671, EPI_ISL_1963672, EPI_ISL_1963674, EPI_ISL_1963677, EPI_ISL_1963678, EPI_ISL_1963679, EPI_ISL_1963680, EPI_ISL_1963681, EPI_ISL_1963682, EPI_ISL_1963683, EPI_ISL_1963684, EPI_ISL_1963685, EPI_ISL_1963687, EPI_ISL_1963688, EPI_ISL_1963689, EPI_ISL_1963690, EPI_ISL_1963691, EPI_ISL_1963692, EPI_ISL_1963693, EPI_ISL_1963694, EPI_ISL_1963695, EPI_ISL_1963697, EPI_ISL_1963699, EPI_ISL_1963700, EPI_ISL_1963701, EPI_ISL_1963702, EPI_ISL_1963704, EPI_ISL_1963705, EPI_ISL_1963706, EPI_ISL_1963707, EPI_ISL_1963708, EPI_ISL_1963709, EPI_ISL_1963711, EPI_ISL_1963712, EPI_ISL_1963713, EPI_ISL_1963714, EPI_ISL_1963716, EPI_ISL_1963717, EPI_ISL_1963720, EPI_ISL_1963721, EPI_ISL_1963724, EPI_ISL_1963725, EPI_ISL_1963726, EPI_ISL_1963729, EPI_ISL_1963730, EPI_ISL_1963731, EPI_ISL_1963732, EPI_ISL_1963733, EPI_ISL_1963734, EPI_ISL_1963736, EPI_ISL_1963738, EPI_ISL_1963740, EPI_ISL_1963741, EPI_ISL_1963742, EPI_ISL_1963743, EPI_ISL_1963744, EPI_ISL_1963745, EPI_ISL_1963746, EPI_ISL_1963747, EPI_ISL_1963748, EPI_ISL_1963749, EPI_ISL_1963750, EPI_ISL_1963751, EPI_ISL_1963752, EPI_ISL_1963753, EPI_ISL_1963754, EPI_ISL_1963755, EPI_ISL_1963756, EPI_ISL_1963757, EPI_ISL_1963758, EPI_ISL_1963759, EPI_ISL_1963760, EPI_ISL_1963761, EPI_ISL_1963762, EPI_ISL_1963763, EPI_ISL_1963764, EPI_ISL_1963765, EPI_ISL_1963766, EPI_ISL_1963768, EPI_ISL_1963769, EPI_ISL_1963770, EPI_ISL_1963771, EPI_ISL_1963772, EPI_ISL_1963773, EPI_ISL_1963774, EPI_ISL_1963775, EPI_ISL_1963776, EPI_ISL_1963777, EPI_ISL_1963778, EPI_ISL_1963779, EPI_ISL_1963780, EPI_ISL_1963781, EPI_ISL_1963782, EPI_ISL_1963783, EPI_ISL_1963784, EPI_ISL_1963785, EPI_ISL_1963786, EPI_ISL_1963787, EPI_ISL_1963788, EPI_ISL_1963789, EPI_ISL_1963790, EPI_ISL_1963791, EPI_ISL_1963792, EPI_ISL_1963793, EPI_ISL_1963794, EPI_ISL_1963795, EPI_ISL_1963796, EPI_ISL_1963797, EPI_ISL_1963799, EPI_ISL_1963800, EPI_ISL_1963801, EPI_ISL_1963802, EPI_ISL_1963803, EPI_ISL_1963804, EPI_ISL_1963805, EPI_ISL_1963806, EPI_ISL_1963807, EPI_ISL_1963808, EPI_ISL_1963810, EPI_ISL_1963811, EPI_ISL_1963812, EPI_ISL_1963813, EPI_ISL_1963814, EPI_ISL_1963815, EPI_ISL_1963816, EPI_ISL_1963817, EPI_ISL_1963818, EPI_ISL_1963819, EPI_ISL_1963820, EPI_ISL_1963821, EPI_ISL_1963822, EPI_ISL_1963823, EPI_ISL_1963824, EPI_ISL_1963825, EPI_ISL_1963826, EPI_ISL_2023050, EPI_ISL_2023051, EPI_ISL_2023052, EPI_ISL_2023053, EPI_ISL_2023054, EPI_ISL_2023055, EPI_ISL_2023056, EPI_ISL_2023057, EPI_ISL_2023058, EPI_ISL_2023061, EPI_ISL_2023062, EPI_ISL_2023063, EPI_ISL_2023064, EPI_ISL_2023065, EPI_ISL_2023066, EPI_ISL_2023067, EPI_ISL_2023068, EPI_ISL_2023069, EPI_ISL_2023070, EPI_ISL_2023072, EPI_ISL_2023073, EPI_ISL_2023074, EPI_ISL_2023075, EPI_ISL_2023076, EPI_ISL_2023077, EPI_ISL_2023078, EPI_ISL_2023079, EPI_ISL_2023080, EPI_ISL_2023081, EPI_ISL_2023082, EPI_ISL_2023084, EPI_ISL_2023085, EPI_ISL_2023086, EPI_ISL_2023087, EPI_ISL_2023088, EPI_ISL_2023089, EPI_ISL_2023090, EPI_ISL_2023091, EPI_ISL_2023092, EPI_ISL_2023093, EPI_ISL_2023094, EPI_ISL_2023095, EPI_ISL_2023096, EPI_ISL_2023097, EPI_ISL_2023098, EPI_ISL_2023099, EPI_ISL_2023100, EPI_ISL_2023101, EPI_ISL_2023102, EPI_ISL_2023103, EPI_ISL_2023104, EPI_ISL_2023105, EPI_ISL_2023106, EPI_ISL_2023108, EPI_ISL_2023109, EPI_ISL_2023110, EPI_ISL_2023111, EPI_ISL_2023112, EPI_ISL_2023114, EPI_ISL_2023115, EPI_ISL_2023116, EPI_ISL_2023117, EPI_ISL_2023118, EPI_ISL_2023119, EPI_ISL_2023120, EPI_ISL_2023121, EPI_ISL_2023122, EPI_ISL_2023123, EPI_ISL_2023124, EPI_ISL_2023125, EPI_ISL_2023126, EPI_ISL_2023127, EPI_ISL_2023128, EPI_ISL_2023129, EPI_ISL_2023130, EPI_ISL_2023131, EPI_ISL_2023132, EPI_ISL_2023133,

EPI_ISL_2023134, EPI_ISL_2023135, EPI_ISL_2023136, EPI_ISL_2023137, EPI_ISL_2023138, EPI_ISL_2023139, EPI_ISL_2023140, EPI_ISL_2023141, EPI_ISL_2023142, EPI_ISL_2023143, EPI_ISL_2023144, EPI_ISL_2023145, EPI_ISL_2023146, EPI_ISL_2023147, EPI_ISL_2023149, EPI_ISL_2023150, EPI_ISL_2023151, EPI_ISL_2023152, EPI_ISL_2023153, EPI_ISL_2023154, EPI_ISL_2023155, EPI_ISL_2023156, EPI_ISL_2023157, EPI_ISL_2023159, EPI_ISL_2023160, EPI_ISL_2023161, EPI_ISL_2023163, EPI_ISL_2023164, EPI_ISL_2023165, EPI_ISL_2023166, EPI_ISL_2023167, EPI_ISL_2023168, EPI_ISL_2023169, EPI_ISL_2023170, EPI_ISL_2023171, EPI_ISL_2023172, EPI_ISL_2023173, EPI_ISL_2023174, EPI_ISL_2023176, EPI_ISL_2023177, EPI_ISL_2023178, EPI_ISL_2023179, EPI_ISL_2023180, EPI_ISL_2023181, EPI_ISL_2023182, EPI_ISL_2023183, EPI_ISL_2023184, EPI_ISL_2023186, EPI_ISL_2023187, EPI_ISL_2023188, EPI_ISL_2023189, EPI_ISL_2023190, EPI_ISL_2023191, EPI_ISL_2023192, EPI_ISL_2023193, EPI_ISL_2023194, EPI_ISL_2023195, EPI_ISL_2023196, EPI_ISL_2023198, EPI_ISL_2023199, EPI_ISL_2023200, EPI_ISL_2023201, EPI_ISL_2023202, EPI_ISL_2023203, EPI_ISL_2023204, EPI_ISL_2023205, EPI_ISL_2023207, EPI_ISL_2023208, EPI_ISL_2023209, EPI_ISL_2023210, EPI_ISL_2023211, EPI_ISL_2023212, EPI_ISL_2023213, EPI_ISL_2159576, EPI_ISL_2159578, EPI_ISL_2159580, EPI_ISL_2159581, EPI_ISL_2159583, EPI_ISL_2159585, EPI_ISL_2159586, EPI_ISL_2159588, EPI_ISL_2159589, EPI_ISL_2159591, EPI_ISL_2159593, EPI_ISL_2159594, EPI_ISL_2159596, EPI_ISL_2159597, EPI_ISL_2159599, EPI_ISL_2159600, EPI_ISL_2159602, EPI_ISL_2159603, EPI_ISL_2159605, EPI_ISL_2159606, EPI_ISL_2159608, EPI_ISL_2159609, EPI_ISL_2159611, EPI_ISL_2159613, EPI_ISL_2159615, EPI_ISL_2159618, EPI_ISL_2159621, EPI_ISL_2159623, EPI_ISL_2159625, EPI_ISL_2159626, EPI_ISL_2159629, EPI_ISL_2159631, EPI_ISL_2159632, EPI_ISL_2159634, EPI_ISL_2159636, EPI_ISL_2159637, EPI_ISL_2159639, EPI_ISL_2159640, EPI_ISL_2159642, EPI_ISL_2159643, EPI_ISL_2159645, EPI_ISL_2159646, EPI_ISL_2159648, EPI_ISL_2159649, EPI_ISL_2159651, EPI_ISL_2159653, EPI_ISL_2159656, EPI_ISL_2159657, EPI_ISL_2159659, EPI_ISL_2159661, EPI_ISL_2159662, EPI_ISL_2159664, EPI_ISL_2159665, EPI_ISL_2159667, EPI_ISL_2296381, EPI_ISL_2296382, EPI_ISL_2296383, EPI_ISL_2296384, EPI_ISL_2296385, EPI_ISL_2296386, EPI_ISL_2296387, EPI_ISL_2296388, EPI_ISL_2296389, EPI_ISL_2296390, EPI_ISL_2296391, EPI_ISL_2296392, EPI_ISL_2296393, EPI_ISL_2296394, EPI_ISL_2296395, EPI_ISL_2296396, EPI_ISL_2296398, EPI_ISL_2296399, EPI_ISL_2296400, EPI_ISL_2296401, EPI_ISL_2296402, EPI_ISL_2296403, EPI_ISL_2296404, EPI_ISL_2296405, EPI_ISL_2296408, EPI_ISL_2296409, EPI_ISL_2296410, EPI_ISL_2296412, EPI_ISL_2296413, EPI_ISL_2296414, EPI_ISL_2296415, EPI_ISL_2296416, EPI_ISL_2296417, EPI_ISL_2296418, EPI_ISL_2296419, EPI_ISL_2296420, EPI_ISL_2296421, EPI_ISL_2296422, EPI_ISL_2296423, EPI_ISL_2296425, EPI_ISL_2296426, EPI_ISL_2296427, EPI_ISL_2296428, EPI_ISL_2296429, EPI_ISL_2296430, EPI_ISL_2296431, EPI_ISL_2296432, EPI_ISL_2296433, EPI_ISL_2296434, EPI_ISL_2296435, EPI_ISL_2296436, EPI_ISL_2296437, EPI_ISL_2296438, EPI_ISL_2296439, EPI_ISL_2296440, EPI_ISL_2296441, EPI_ISL_2296442, EPI_ISL_2296443, EPI_ISL_2296444, EPI_ISL_2296446, EPI_ISL_2296447, EPI_ISL_2296448, EPI_ISL_2296449, EPI_ISL_2296450, EPI_ISL_2296451, EPI_ISL_2296452, EPI_ISL_2296453, EPI_ISL_2296454, EPI_ISL_2296455, EPI_ISL_2296456, EPI_ISL_2296458, EPI_ISL_2296459, EPI_ISL_2296460, EPI_ISL_2296461, EPI_ISL_2296463, EPI_ISL_2296464, EPI_ISL_2296465, EPI_ISL_2296466, EPI_ISL_2426099, EPI_ISL_2426100, EPI_ISL_2426101, EPI_ISL_2426102, EPI_ISL_2426103, EPI_ISL_2426104, EPI_ISL_2426105, EPI_ISL_2426106, EPI_ISL_2426107, EPI_ISL_2426108, EPI_ISL_2426110, EPI_ISL_2426111, EPI_ISL_2426113, EPI_ISL_2426114, EPI_ISL_2426115, EPI_ISL_2426117, EPI_ISL_2426118, EPI_ISL_2426119, EPI_ISL_2426121, EPI_ISL_2426122, EPI_ISL_2426124, EPI_ISL_2426125, EPI_ISL_2426126, EPI_ISL_2426127, EPI_ISL_2426128, EPI_ISL_2426129, EPI_ISL_2426130, EPI_ISL_2426131, EPI_ISL_2426132, EPI_ISL_2426133, EPI_ISL_2426134, EPI_ISL_2426135, EPI_ISL_2426136, EPI_ISL_2426137, EPI_ISL_2426138, EPI_ISL_2426139, EPI_ISL_2426140, EPI_ISL_2501663, EPI_ISL_2501666, EPI_ISL_2501667, EPI_ISL_2501668, EPI_ISL_2501669, EPI_ISL_2501672, EPI_ISL_2501673, EPI_ISL_2501674, EPI_ISL_2501675, EPI_ISL_2501676, EPI_ISL_2501677, EPI_ISL_2501678, EPI_ISL_2501679, EPI_ISL_2501680, EPI_ISL_2501681, EPI_ISL_2501682, EPI_ISL_2501683, EPI_ISL_2501684, EPI_ISL_2501686, EPI_ISL_2501687, EPI_ISL_2501688, EPI_ISL_2501689, EPI_ISL_2501690, EPI_ISL_2501691, EPI_ISL_2501692, EPI_ISL_2501693, EPI_ISL_2501694, EPI_ISL_2501695, EPI_ISL_2501696, EPI_ISL_2501697, EPI_ISL_2501699, EPI_ISL_2501703, EPI_ISL_2501704, EPI_ISL_2501705, EPI_ISL_2501706, EPI_ISL_2501719, EPI_ISL_2501721, EPI_ISL_2501722, EPI_ISL_2501724, EPI_ISL_2501725, EPI_ISL_2501726, EPI_ISL_2501727, EPI_ISL_2598670, EPI_ISL_2598671, EPI_ISL_2598672, EPI_ISL_2598673, EPI_ISL_2598674, EPI_ISL_2598675, EPI_ISL_2598677, EPI_ISL_2598678, EPI_ISL_2598679, EPI_ISL_2598680, EPI_ISL_2598681, EPI_ISL_2598682, EPI_ISL_2598683, EPI_ISL_2598684, EPI_ISL_2598685, EPI_ISL_2598686, EPI_ISL_2598687, EPI_ISL_2598688, EPI_ISL_2598690, EPI_ISL_2598691, EPI_ISL_2598693, EPI_ISL_2598694, EPI_ISL_2598695, EPI_ISL_2598696, EPI_ISL_2716222, EPI_ISL_2716223, EPI_ISL_2716224, EPI_ISL_2716225, EPI_ISL_2716226, EPI_ISL_2716227, EPI_ISL_2716228, EPI_ISL_2716229, EPI_ISL_2716230, EPI_ISL_2716231, EPI_ISL_2716232, EPI_ISL_2716233, EPI_ISL_2716234, EPI_ISL_2716235, EPI_ISL_2716236, EPI_ISL_2716238, EPI_ISL_2716239, EPI_ISL_2716241, EPI_ISL_2716242, EPI_ISL_2716243, EPI_ISL_2716244, EPI_ISL_2716246, EPI_ISL_2716247, EPI_ISL_2716248, EPI_ISL_2716249, EPI_ISL_2716250, EPI_ISL_2716251, EPI_ISL_2716252, EPI_ISL_2716253, EPI_ISL_2716254, EPI_ISL_2776208, EPI_ISL_2776209, EPI_ISL_2776210, EPI_ISL_2776212, EPI_ISL_2776213, EPI_ISL_2860304, EPI_ISL_2860305, EPI_ISL_2860306, EPI_ISL_2860307, EPI_ISL_2860308, EPI_ISL_2860309, EPI_ISL_2860311, EPI_ISL_2860312, EPI_ISL_2860313, EPI_ISL_2860314, EPI_ISL_2860315, EPI_ISL_2860316, EPI_ISL_2860317, EPI_ISL_2960035, EPI_ISL_2960036, EPI_ISL_2960037, EPI_ISL_2960038, EPI_ISL_2960039, EPI_ISL_2960040, EPI_ISL_2960041, EPI_ISL_2960042, EPI_ISL_2960044, EPI_ISL_2960045, EPI_ISL_2960046, EPI_ISL_2960047, EPI_ISL_2960048, EPI_ISL_2960049, EPI_ISL_2960050, EPI_ISL_3025414, EPI_ISL_3025415, EPI_ISL_3025416, EPI_ISL_3025417, EPI_ISL_3025418, EPI_ISL_3025419, EPI_ISL_3025420, EPI_ISL_3025421, EPI_ISL_3025422, EPI_ISL_3025423, EPI_ISL_3025424, EPI_ISL_3025425, EPI_ISL_3025426, EPI_ISL_3025427, EPI_ISL_3025428, EPI_ISL_3025429, EPI_ISL_3025430, EPI_ISL_3025431, EPI_ISL_3025432, EPI_ISL_3025433, EPI_ISL_3025434, EPI_ISL_3025435, EPI_ISL_3025436, EPI_ISL_3025437, EPI_ISL_3025438, EPI_ISL_3025439, EPI_ISL_3025440, EPI_ISL_3025441, EPI_ISL_3025442, EPI_ISL_3025443, EPI_ISL_3104658, EPI_ISL_3104660, EPI_ISL_3104661, EPI_ISL_3104676, EPI_ISL_3104681, EPI_ISL_3104682, EPI_ISL_3104686, EPI_ISL_3104687, EPI_ISL_3104688, EPI_ISL_3104689, EPI_ISL_3104692, EPI_ISL_3104693, EPI_ISL_3104694, EPI_ISL_3104711, EPI_ISL_3104712, EPI_ISL_3104713, EPI_ISL_3104720, EPI_ISL_3104721, EPI_ISL_3104726, EPI_ISL_3104727, EPI_ISL_3104728, EPI_ISL_3104729, EPI_ISL_3104730, EPI_ISL_3104731, EPI_ISL_3104732, EPI_ISL_3104733, EPI_ISL_3104734, EPI_ISL_3104735, EPI_ISL_3104736, EPI_ISL_3104737, EPI_ISL_3104738, EPI_ISL_3104739, EPI_ISL_3104740, EPI_ISL_3104741, EPI_ISL_3104742, EPI_ISL_3104743, EPI_ISL_3104744, EPI_ISL_3104745, EPI_ISL_3104747, EPI_ISL_3104748, EPI_ISL_3104749, EPI_ISL_3104750, EPI_ISL_3104751, EPI_ISL_3105419, EPI_ISL_3105421, EPI_ISL_3105422, EPI_ISL_3105433, EPI_ISL_3105442, EPI_ISL_3105453, EPI_ISL_3105454, EPI_ISL_3105456, EPI_ISL_3105457, EPI_ISL_3105458, EPI_ISL_3105459, EPI_ISL_3105460, EPI_ISL_3105462, EPI_ISL_3105464, EPI_ISL_3105470, EPI_ISL_3105472, EPI_ISL_3105473, EPI_ISL_3105474, EPI_ISL_3105475, EPI_ISL_3105476, EPI_ISL_3105477, EPI_ISL_3105478, EPI_ISL_3105479, EPI_ISL_3105481, EPI_ISL_3105482, EPI_ISL_3105483, EPI_ISL_3105484, EPI_ISL_3105485, EPI_ISL_3105487, EPI_ISL_3105489, EPI_ISL_3105492, EPI_ISL_3105493, EPI_ISL_3105495, EPI_ISL_3105499, EPI_ISL_3105500, EPI_ISL_3105505, EPI_ISL_3105509, EPI_ISL_3105514, EPI_ISL_3105515, EPI_ISL_3105516, EPI_ISL_3105517, EPI_ISL_3105518, EPI_ISL_3105519, EPI_ISL_3105522, EPI_ISL_3105523, EPI_ISL_3105525, EPI_ISL_3105540, EPI_ISL_3105542, EPI_ISL_3105543, EPI_ISL_3105545, EPI_ISL_3133022, EPI_ISL_3133023, EPI_ISL_3188563, EPI_ISL_3188566, EPI_ISL_3188573, EPI_ISL_3188575, EPI_ISL_3188592, EPI_ISL_3188593, EPI_ISL_3188594, EPI_ISL_3188595, EPI_ISL_3188596, EPI_ISL_3188616, EPI_ISL_3188621, EPI_ISL_3236362, EPI_ISL_3236363, EPI_ISL_3236364, EPI_ISL_3236365, EPI_ISL_3236366, EPI_ISL_3236367, EPI_ISL_3236368, EPI_ISL_3236369, EPI_ISL_3236370, EPI_ISL_3236371, EPI_ISL_3236372, EPI_ISL_3236373, EPI_ISL_3236374, EPI_ISL_3236375, EPI_ISL_3236376, EPI_ISL_3236377, EPI_ISL_3236378, EPI_ISL_3236379, EPI_ISL_3236380, EPI_ISL_3236381, EPI_ISL_3236382, EPI_ISL_3236383, EPI_ISL_3236384, EPI_ISL_3236385, EPI_ISL_3236386, EPI_ISL_3236387, EPI_ISL_3236388, EPI_ISL_3236389, EPI_ISL_3236390, EPI_ISL_3236391, EPI_ISL_3236392, EPI_ISL_3236393, EPI_ISL_3236394, EPI_ISL_3236395, EPI_ISL_3236396, EPI_ISL_3236397, EPI_ISL_3236398, EPI_ISL_3236399, EPI_ISL_3236400, EPI_ISL_3236402, EPI_ISL_3236403, EPI_ISL_3236404, EPI_ISL_3236405, EPI_ISL_3236406, EPI_ISL_3236407, EPI_ISL_3236408, EPI_ISL_3236409, EPI_ISL_3236410, EPI_ISL_3236411, EPI_ISL_3236412, EPI_ISL_3236413, EPI_ISL_3236414, EPI_ISL_3236415, EPI_ISL_3236416, EPI_ISL_3236417, EPI_ISL_3236418, EPI_ISL_3236419, EPI_ISL_3236420, EPI_ISL_3236421, EPI_ISL_3236422, EPI_ISL_3236423, EPI_ISL_3236424, EPI_ISL_3236425, EPI_ISL_3236426, EPI_ISL_3236427, EPI_ISL_3236428, EPI_ISL_3236429, EPI_ISL_3236430, EPI_ISL_3236431, EPI_ISL_3236432, EPI_ISL_3236453, EPI_ISL_3236454, EPI_ISL_3236455, EPI_ISL_3236456, EPI_ISL_3236457, EPI_ISL_3236458, EPI_ISL_3236459, EPI_ISL_3236460, EPI_ISL_3236461, EPI_ISL_3236462, EPI_ISL_3236463, EPI_ISL_3236464, EPI_ISL_3236465, EPI_ISL_3236466, EPI_ISL_3236467, EPI_ISL_3236468, EPI_ISL_3236469, EPI_ISL_3236470, EPI_ISL_3236471, EPI_ISL_3236472, EPI_ISL_3236473, EPI_ISL_3236474, EPI_ISL_3236475, EPI_ISL_3236476, EPI_ISL_3236477, EPI_ISL_3236478, EPI_ISL_3236479, EPI_ISL_3236480, EPI_ISL_3236481, EPI_ISL_3236482, EPI_ISL_3236483, EPI_ISL_3236484, EPI_ISL_3236485, EPI_ISL_3236486, EPI_ISL_3236487, EPI_ISL_3236488, EPI_ISL_3236489, EPI_ISL_3236490, EPI_ISL_3236491, EPI_ISL_3236492, EPI_ISL_3236493, EPI_ISL_3236494, EPI_ISL_3347233, EPI_ISL_3347234, EPI_ISL_3347235, EPI_ISL_3347236, EPI_ISL_3347237, EPI_ISL_3347238, EPI_ISL_3347239, EPI_ISL_3347240, EPI_ISL_3347241, EPI_ISL_3347242, EPI_ISL_3347243, EPI_ISL_3347244, EPI_ISL_3347245, EPI_ISL_3347246, EPI_ISL_3347247, EPI_ISL_3347248, EPI_ISL_3347249, EPI_ISL_3347250, EPI_ISL_3347251, EPI_ISL_3347252, EPI_ISL_3347253, EPI_ISL_3347254, EPI_ISL_3347255, EPI_ISL_3347256, EPI_ISL_3347257, EPI_ISL_3347258, EPI_ISL_3347259, EPI_ISL_3347260, EPI_ISL_3347261, EPI_ISL_3347262, EPI_ISL_3347263, EPI_ISL_3347264, EPI_ISL_3347265, EPI_ISL_3347266, EPI_ISL_3347267, EPI_ISL_3347268, EPI_ISL_3347269, EPI_ISL_3347270, EPI_ISL_3347271, EPI_ISL_3347272, EPI_ISL_3347273, EPI_ISL_3347274, EPI_ISL_3347275, EPI_ISL_3347276, EPI_ISL_3369781, EPI_ISL_3369782, EPI_ISL_3370105, EPI_ISL_3370106, EPI_ISL_3370107, EPI_ISL_3370108, EPI_ISL_3370109, EPI_ISL_3370110, EPI_ISL_3370111, EPI_ISL_3370112, EPI_ISL_3370113, EPI_ISL_3370114, EPI_ISL_3370115, EPI_ISL_3370116, EPI_ISL_3370117, EPI_ISL_3370118, EPI_ISL_3370119, EPI_ISL_3370120, EPI_ISL_3370122, EPI_ISL_3370124, EPI_ISL_3370125, EPI_ISL_3370126, EPI_ISL_3370127, EPI_ISL_3370128, EPI_ISL_3370129, EPI_ISL_3370130, EPI_ISL_3370131, EPI_ISL_3370132, EPI_ISL_3370133, EPI_ISL_3370134, EPI_ISL_3370135, EPI_ISL_3370136, EPI_ISL_3370137, EPI_ISL_3370138, EPI_ISL_3370139, EPI_ISL_3370140, EPI_ISL_3370142, EPI_ISL_3370143, EPI_ISL_3370144, EPI_ISL_3370145, EPI_ISL_3370146, EPI_ISL_3370147, EPI_ISL_3370148, EPI_ISL_3370149, EPI_ISL_3370150, EPI_ISL_3370151, EPI_ISL_3370152, EPI_ISL_3370153, EPI_ISL_3370154, EPI_ISL_3370155, EPI_ISL_3370156, EPI_ISL_3370157, EPI_ISL_3370158, EPI_ISL_3370159, EPI_ISL_3370160, EPI_ISL_3370161, EPI_ISL_3370162, EPI_ISL_3370163, EPI_ISL_3370164, EPI_ISL_3370165, EPI_ISL_3370166, EPI_ISL_3370167, EPI_ISL_3370168, EPI_ISL_3370169, EPI_ISL_3370170, EPI_ISL_3370171, EPI_ISL_3370172, EPI_ISL_3370173, EPI_ISL_3370174, EPI_ISL_3370175, EPI_ISL_3370176, EPI_ISL_3370177, EPI_ISL_3370178, EPI_ISL_3370179, EPI_ISL_3370180, EPI_ISL_3370181, EPI_ISL_3370182, EPI_ISL_3370183, EPI_ISL_3370184, EPI_ISL_3370185, EPI_ISL_3370186, EPI_ISL_3370188, EPI_ISL_3370189, EPI_ISL_3370190, EPI_ISL_3370191, EPI_ISL_3370192, EPI_ISL_3370193, EPI_ISL_3370194, EPI_ISL_3370196, EPI_ISL_3370197, EPI_ISL_3370198, EPI_ISL_3370199, EPI_ISL_3370200, EPI_ISL_3370201, EPI_ISL_3370202, EPI_ISL_3370203, EPI_ISL_3370204, EPI_ISL_3370205, EPI_ISL_3370206, EPI_ISL_3370207, EPI_ISL_3370208, EPI_ISL_3370209, EPI_ISL_3370210, EPI_ISL_3370211, EPI_ISL_3370212, EPI_ISL_3370213, EPI_ISL_3370214, EPI_ISL_3370215, EPI_ISL_3370216, EPI_ISL_3370217, EPI_ISL_3370218, EPI_ISL_3370219, EPI_ISL_3370220, EPI_ISL_3370221, EPI_ISL_3370222, EPI_ISL_3370223, EPI_ISL_3370224, EPI_ISL_3370225, EPI_ISL_3370226, EPI_ISL_3370227, EPI_ISL_3370228, EPI_ISL_3370229, EPI_ISL_3370231, EPI_ISL_3370232, EPI_ISL_3370233, EPI_ISL_3370234, EPI_ISL_3370235, EPI_ISL_3370236, EPI_ISL_3370237, EPI_ISL_3370238, EPI_ISL_3370240, EPI_ISL_3370241, EPI_ISL_3370242, EPI_ISL_3370243, EPI_ISL_3370244, EPI_ISL_3370245, EPI_ISL_3370246, EPI_ISL_3370248, EPI_ISL_3370249, EPI_ISL_3370250, EPI_ISL_3370251, EPI_ISL_3370252, EPI_ISL_3370253, EPI_ISL_3370254, EPI_ISL_3370255, EPI_ISL_3370256, EPI_ISL_3370257, EPI_ISL_3370258, EPI_ISL_3370259, EPI_ISL_3370260, EPI_ISL_3370261, EPI_ISL_3370262, EPI_ISL_3370263, EPI_ISL_3370264, EPI_ISL_3370265, EPI_ISL_3370266, EPI_ISL_3370267

| | | | |
|---|---|---|---|
| see above | Yale Clinical Virology Lab | Grubaugh Lab - Yale School of Public Health | Anderson Brito; Anne Wyllie; Annie Watkins; Chaney Kalinich; Chantal Vogels; Isabel Ott; Isabell Ott; Jessica Rothman; Joseph Fauver; Kendall Billig; Mallery Breban; Marie L. Landry; Mary Petrone; Nathan Grubaugh; Tara Alpert; Tobias Koch |

EPI_ISL_1793385, EPI_ISL_1793386, EPI_ISL_1793387, EPI_ISL_1793388, EPI_ISL_1793389, EPI_ISL_1793390, EPI_ISL_1793391, EPI_ISL_1793392, EPI_ISL_1793394, EPI_ISL_1793395, EPI_ISL_1793396, EPI_ISL_1793398, EPI_ISL_1793399, EPI_ISL_1793400, EPI_ISL_1793402, EPI_ISL_1793403, EPI_ISL_1793404, EPI_ISL_1793405, EPI_ISL_1793406, EPI_ISL_1793407, EPI_ISL_1793408, EPI_ISL_1793409, EPI_ISL_1793410, EPI_ISL_1793411, EPI_ISL_1793412, EPI_ISL_1793415, EPI_ISL_1793416, EPI_ISL_1793417, EPI_ISL_1793419, EPI_ISL_1793420, EPI_ISL_1793421, EPI_ISL_1793422, EPI_ISL_1793423, EPI_ISL_1793424, EPI_ISL_1793425, EPI_ISL_1793426, EPI_ISL_1793427, EPI_ISL_1793428, EPI_ISL_1793430, EPI_ISL_1793431, EPI_ISL_1793432, EPI_ISL_1793433, EPI_ISL_1793434, EPI_ISL_1793435, EPI_ISL_1793436, EPI_ISL_1793437, EPI_ISL_1793438, EPI_ISL_1793439, EPI_ISL_1793440, EPI_ISL_1793441, EPI_ISL_1793444, EPI_ISL_1793445, EPI_ISL_1793446, EPI_ISL_1793447, EPI_ISL_1793448, EPI_ISL_1793451, EPI_ISL_1793452, EPI_ISL_1793453, EPI_ISL_1793454, EPI_ISL_1793455, EPI_ISL_1793456, EPI_ISL_1793457, EPI_ISL_1793458, EPI_ISL_1793460, EPI_ISL_1793461, EPI_ISL_1793462, EPI_ISL_1793464, EPI_ISL_1793465, EPI_ISL_1793466, EPI_ISL_1793467, EPI_ISL_1793468, EPI_ISL_1793469, EPI_ISL_1793470, EPI_ISL_1793471, EPI_ISL_1793472, EPI_ISL_1793473, EPI_ISL_1793474, EPI_ISL_1793475, EPI_ISL_1793476, EPI_ISL_1793477, EPI_ISL_1793478, EPI_ISL_1793479, EPI_ISL_1793480, EPI_ISL_1793481, EPI_ISL_1793482, EPI_ISL_1793483, EPI_ISL_1793484, EPI_ISL_1793485, EPI_ISL_1793486, EPI_ISL_1793487, EPI_ISL_1793488, EPI_ISL_1793489, EPI_ISL_1793491, EPI_ISL_1793492, EPI_ISL_1793494, EPI_ISL_1793495, EPI_ISL_1793496, EPI_ISL_1793497, EPI_ISL_1793498, EPI_ISL_1793499, EPI_ISL_1793500, EPI_ISL_1793501, EPI_ISL_1793503, EPI_ISL_1793504, EPI_ISL_1793506, EPI_ISL_1793508, EPI_ISL_1793509, EPI_ISL_1793511, EPI_ISL_1793512, EPI_ISL_1793513, EPI_ISL_1793515, EPI_ISL_1793517, EPI_ISL_1793518, EPI_ISL_1793519, EPI_ISL_1793521, EPI_ISL_1793522, EPI_ISL_1793523, EPI_ISL_1793524, EPI_ISL_1793525, EPI_ISL_1793526, EPI_ISL_1793527, EPI_ISL_1793529, EPI_ISL_1793530, EPI_ISL_1793531, EPI_ISL_1793532, EPI_ISL_1793533, EPI_ISL_1793535, EPI_ISL_1793536, EPI_ISL_1793537, EPI_ISL_1793538, EPI_ISL_1793539, EPI_ISL_1793540, EPI_ISL_1793541, EPI_ISL_1793543, EPI_ISL_1793544, EPI_ISL_1793545, EPI_ISL_1793546, EPI_ISL_1793547, EPI_ISL_1793549, EPI_ISL_1793550, EPI_ISL_1793551, EPI_ISL_1793552, EPI_ISL_1793553, EPI_ISL_1793555, EPI_ISL_1793556, EPI_ISL_1793557, EPI_ISL_1793559, EPI_ISL_1793560, EPI_ISL_1793561, EPI_ISL_1793562, EPI_ISL_1793563, EPI_ISL_1793565, EPI_ISL_1793566, EPI_ISL_1793567, EPI_ISL_1793568, EPI_ISL_1793569, EPI_ISL_1793570, EPI_ISL_1793571, EPI_ISL_1793572, EPI_ISL_1793573, EPI_ISL_1793574, EPI_ISL_1793575, EPI_ISL_1793576, EPI_ISL_1793577, EPI_ISL_1793579, EPI_ISL_1793580, EPI_ISL_1793581, EPI_ISL_1793582, EPI_ISL_1793583, EPI_ISL_1793584, EPI_ISL_1793585, EPI_ISL_1793586, EPI_ISL_1793587, EPI_ISL_1793588, EPI_ISL_1793589, EPI_ISL_1793590, EPI_ISL_1793592, EPI_ISL_1793595, EPI_ISL_1793596, EPI_ISL_1793597, EPI_ISL_1793598, EPI_ISL_1793599, EPI_ISL_1793600, EPI_ISL_1793602, EPI_ISL_1793603, EPI_ISL_1793604, EPI_ISL_1793606, EPI_ISL_1793607, EPI_ISL_1793608, EPI_ISL_1793610, EPI_ISL_1793611, EPI_ISL_1793612, EPI_ISL_1793613, EPI_ISL_1793614, EPI_ISL_1793616, EPI_ISL_1793618, EPI_ISL_1793619, EPI_ISL_1793620, EPI_ISL_1793621, EPI_ISL_1793622, EPI_ISL_1793623, EPI_ISL_1793624, EPI_ISL_1793625, EPI_ISL_1793626, EPI_ISL_1793627, EPI_ISL_1793628, EPI_ISL_1793629, EPI_ISL_1793630, EPI_ISL_1793631, EPI_ISL_1793632, EPI_ISL_1793633, EPI_ISL_1793634, EPI_ISL_1793635, EPI_ISL_1793636, EPI_ISL_1793637, EPI_ISL_1793638, EPI_ISL_1793639, EPI_ISL_1793640, EPI_ISL_1793641, EPI_ISL_1793642, EPI_ISL_1793643, EPI_ISL_1793644, EPI_ISL_1793645, EPI_ISL_2020688, EPI_ISL_2020690, EPI_ISL_2020691, EPI_ISL_2020692, EPI_ISL_2020694, EPI_ISL_2020695, EPI_ISL_2020696, EPI_ISL_2020697, EPI_ISL_2020698, EPI_ISL_2020699, EPI_ISL_2020700, EPI_ISL_2020701, EPI_ISL_2020702, EPI_ISL_2020703, EPI_ISL_2020704, EPI_ISL_2020706, EPI_ISL_2020707, EPI_ISL_2020708, EPI_ISL_2020709, EPI_ISL_2020710, EPI_ISL_2020711, EPI_ISL_2020712, EPI_ISL_2020713, EPI_ISL_2020714, EPI_ISL_2020715, EPI_ISL_2020716, EPI_ISL_2020717, EPI_ISL_2020718, EPI_ISL_2020719, EPI_ISL_2020720, EPI_ISL_2020721, EPI_ISL_2020722, EPI_ISL_2020723, EPI_ISL_2020724, EPI_ISL_2020725, EPI_ISL_2020726, EPI_ISL_2020728, EPI_ISL_2020729, EPI_ISL_2020730, EPI_ISL_2020731, EPI_ISL_2020732, EPI_ISL_2020733, EPI_ISL_2020734, EPI_ISL_2020735, EPI_ISL_2020736, EPI_ISL_2020737, EPI_ISL_2020738, EPI_ISL_2020739, EPI_ISL_2020740, EPI_ISL_2020741, EPI_ISL_2020742, EPI_ISL_2020743, EPI_ISL_2020744, EPI_ISL_2020745, EPI_ISL_2020746, EPI_ISL_2020747, EPI_ISL_2020748, EPI_ISL_2020749, EPI_ISL_2020751, EPI_ISL_2020752, EPI_ISL_2020753, EPI_ISL_2020755, EPI_ISL_2020756, EPI_ISL_2020757, EPI_ISL_2020758, EPI_ISL_2020759, EPI_ISL_2020760, EPI_ISL_2020761, EPI_ISL_2020762, EPI_ISL_2020763, EPI_ISL_2020764, EPI_ISL_2020765, EPI_ISL_2020766, EPI_ISL_2020768, EPI_ISL_2020769, EPI_ISL_2020770, EPI_ISL_2020771, EPI_ISL_2020772, EPI_ISL_2020773, EPI_ISL_2020774, EPI_ISL_2020775, EPI_ISL_2020776, EPI_ISL_2020778, EPI_ISL_2020779, EPI_ISL_2020780, EPI_ISL_2020782, EPI_ISL_2020783, EPI_ISL_2020784, EPI_ISL_2020785, EPI_ISL_2020786, EPI_ISL_2020787, EPI_ISL_2020789, EPI_ISL_2020790, EPI_ISL_2020791, EPI_ISL_2020792, EPI_ISL_2020793, EPI_ISL_2020794, EPI_ISL_2020796, EPI_ISL_2020797, EPI_ISL_2020798, EPI_ISL_2020799, EPI_ISL_2020800, EPI_ISL_2020801, EPI_ISL_2020802, EPI_ISL_2020803, EPI_ISL_2020804, EPI_ISL_2020805, EPI_ISL_2020806, EPI_ISL_2020808, EPI_ISL_2020809, EPI_ISL_2020810, EPI_ISL_2020811, EPI_ISL_2020812, EPI_ISL_2020813, EPI_ISL_2020814, EPI_ISL_2020815, EPI_ISL_2020816, EPI_ISL_2020817, EPI_ISL_2020818, EPI_ISL_2020819, EPI_ISL_2020820, EPI_ISL_2020821, EPI_ISL_2020822, EPI_ISL_2020823, EPI_ISL_2020824, EPI_ISL_2020825, EPI_ISL_2020826, EPI_ISL_2020827, EPI_ISL_2020828, EPI_ISL_2020829, EPI_ISL_2020830, EPI_ISL_2020831, EPI_ISL_2020832, EPI_ISL_2020833, EPI_ISL_2020834, EPI_ISL_2020835, EPI_ISL_2020836, EPI_ISL_2020837, EPI_ISL_2020838, EPI_ISL_2020839, EPI_ISL_2020841, EPI_ISL_2020842, EPI_ISL_2020844, EPI_ISL_2020845, EPI_ISL_2020846, EPI_ISL_2020847, EPI_ISL_2020848, EPI_ISL_2020849, EPI_ISL_2020850, EPI_ISL_2020852, EPI_ISL_2020853, EPI_ISL_2020854, EPI_ISL_2020855, EPI_ISL_2020856, EPI_ISL_2020857, EPI_ISL_2020858, EPI_ISL_2020859, EPI_ISL_2020860, EPI_ISL_2020861, EPI_ISL_2034754, EPI_ISL_2034756, EPI_ISL_2034757, EPI_ISL_2034759, EPI_ISL_2034760, EPI_ISL_2034761, EPI_ISL_2034763, EPI_ISL_2034764, EPI_ISL_2034765, EPI_ISL_2034767, EPI_ISL_2034768, EPI_ISL_2034770, EPI_ISL_2034771, EPI_ISL_2034772, EPI_ISL_2034774, EPI_ISL_2034775, EPI_ISL_2034777, EPI_ISL_2034778, EPI_ISL_2034780, EPI_ISL_2034783, EPI_ISL_2034786, EPI_ISL_2034787, EPI_ISL_2034788, EPI_ISL_2034790, EPI_ISL_2034792, EPI_ISL_2034794, EPI_ISL_2034795, EPI_ISL_2034797, EPI_ISL_2034798, EPI_ISL_2034800, EPI_ISL_2034801, EPI_ISL_2034802, EPI_ISL_2034803, EPI_ISL_2034805, EPI_ISL_2034806, EPI_ISL_2034807, EPI_ISL_2034809, EPI_ISL_2034810, EPI_ISL_2034812, EPI_ISL_2034814, EPI_ISL_2034816, EPI_ISL_2034819, EPI_ISL_2034820, EPI_ISL_2034821, EPI_ISL_2034823, EPI_ISL_2034824, EPI_ISL_2034826, EPI_ISL_2034827, EPI_ISL_2034828, EPI_ISL_2034830, EPI_ISL_2034831, EPI_ISL_2034832, EPI_ISL_2034834, EPI_ISL_2034835, EPI_ISL_2034837, EPI_ISL_2034838, EPI_ISL_2034839, EPI_ISL_2034841, EPI_ISL_2034842, EPI_ISL_2034843, EPI_ISL_2034845, EPI_ISL_2034846, EPI_ISL_2034847, EPI_ISL_2034848, EPI_ISL_2034850, EPI_ISL_2034851, EPI_ISL_2034852, EPI_ISL_2034853, EPI_ISL_2034854, EPI_ISL_2034855, EPI_ISL_2034856, EPI_ISL_2034857, EPI_ISL_2034858, EPI_ISL_2034859, EPI_ISL_2034861, EPI_ISL_2034862, EPI_ISL_2034863, EPI_ISL_2034864, EPI_ISL_2034865, EPI_ISL_2034866, EPI_ISL_2034867, EPI_ISL_2034868, EPI_ISL_2034869, EPI_ISL_2034870, EPI_ISL_2034873, EPI_ISL_2034875, EPI_ISL_2034876, EPI_ISL_2034877, EPI_ISL_2034879, EPI_ISL_2034880, EPI_ISL_2034882, EPI_ISL_2034883, EPI_ISL_2034885, EPI_ISL_2034887, EPI_ISL_2034889, EPI_ISL_2034891, EPI_ISL_2034892, EPI_ISL_2034893, EPI_ISL_2034895, EPI_ISL_2034896, EPI_ISL_2034897, EPI_ISL_2034899, EPI_ISL_2034900, EPI_ISL_2034902, EPI_ISL_2034903, EPI_ISL_2034904, EPI_ISL_2034906, EPI_ISL_2034907, EPI_ISL_2034908, EPI_ISL_2034910, EPI_ISL_2034911, EPI_ISL_2034914, EPI_ISL_2034916, EPI_ISL_2034917, EPI_ISL_2034918, EPI_ISL_2034920, EPI_ISL_2034924, EPI_ISL_2034925, EPI_ISL_2034927, EPI_ISL_2034928, EPI_ISL_2034929, EPI_ISL_2034931, EPI_ISL_2034932, EPI_ISL_2034935, EPI_ISL_2034936, EPI_ISL_2034938, EPI_ISL_2034941, EPI_ISL_2034944, EPI_ISL_2034945, EPI_ISL_2034946, EPI_ISL_2034951, EPI_ISL_2034952, EPI_ISL_2034957, EPI_ISL_2034958, EPI_ISL_2034959, EPI_ISL_2034961, EPI_ISL_2034965, EPI_ISL_2034966, EPI_ISL_2034968, EPI_ISL_2034969, EPI_ISL_2034970, EPI_ISL_2034972, EPI_ISL_2034973, EPI_ISL_2034974, EPI_ISL_2034976, EPI_ISL_2034977, EPI_ISL_2034978, EPI_ISL_2034979, EPI_ISL_2034981, EPI_ISL_2034983, EPI_ISL_2034984, EPI_ISL_2034985, EPI_ISL_2034986, EPI_ISL_2034988, EPI_ISL_2034989, EPI_ISL_2034990, EPI_ISL_2034991, EPI_ISL_2034992, EPI_ISL_2034993, EPI_ISL_2034994, EPI_ISL_2034995, EPI_ISL_2034998, EPI_ISL_2034999, EPI_ISL_2035001, EPI_ISL_2035002, EPI_ISL_2035003, EPI_ISL_2035005, EPI_ISL_2035006, EPI_ISL_2035008, EPI_ISL_2035009, EPI_ISL_2035010, EPI_ISL_2035011, EPI_ISL_2035012, EPI_ISL_2035013, EPI_ISL_2035014, EPI_ISL_2035015, EPI_ISL_2035016, EPI_ISL_2035017, EPI_ISL_2035018, EPI_ISL_2035019, EPI_ISL_2035020, EPI_ISL_2035022, EPI_ISL_2035023, EPI_ISL_2035025, EPI_ISL_2035027, EPI_ISL_2035028, EPI_ISL_2035032, EPI_ISL_2035033, EPI_ISL_2035035, EPI_ISL_2035037, EPI_ISL_2035038, EPI_ISL_2035040, EPI_ISL_2035042, EPI_ISL_2035044, EPI_ISL_2035045, EPI_ISL_2035047, EPI_ISL_2035048, EPI_ISL_2035050, EPI_ISL_2035051, EPI_ISL_2035054, EPI_ISL_2035055, EPI_ISL_2035057, EPI_ISL_2035058, EPI_ISL_2035060, EPI_ISL_2035061, EPI_ISL_2035062, EPI_ISL_2035064, EPI_ISL_2035066, EPI_ISL_2035068, EPI_ISL_2035070, EPI_ISL_2035072, EPI_ISL_2035074, EPI_ISL_2035076, EPI_ISL_2035077, EPI_ISL_2035079, EPI_ISL_2035080, EPI_ISL_2035082, EPI_ISL_2035083, EPI_ISL_2035085, EPI_ISL_2035087, EPI_ISL_2035088, EPI_ISL_2035090, EPI_ISL_2035091, EPI_ISL_2035093, EPI_ISL_2035095, EPI_ISL_2035097, EPI_ISL_2035098, EPI_ISL_2035100, EPI_ISL_2035101, EPI_ISL_2158975, EPI_ISL_2158977, EPI_ISL_2158978, EPI_ISL_2158980, EPI_ISL_2158981, EPI_ISL_2158983, EPI_ISL_2158986, EPI_ISL_2158988, EPI_ISL_2158989, EPI_ISL_2158991, EPI_ISL_2158992, EPI_ISL_2158994, EPI_ISL_2158995, EPI_ISL_2158997, EPI_ISL_2158998, EPI_ISL_2159000, EPI_ISL_2159002, EPI_ISL_2159003, EPI_ISL_2159005, EPI_ISL_2159006, EPI_ISL_2159008, EPI_ISL_2159010, EPI_ISL_2159011, EPI_ISL_2159014, EPI_ISL_2159016, EPI_ISL_2159017, EPI_ISL_2159019, EPI_ISL_2159020, EPI_ISL_2159022, EPI_ISL_2159023, EPI_ISL_2159025, EPI_ISL_2159027, EPI_ISL_2159028, EPI_ISL_2159030, EPI_ISL_2159031, EPI_ISL_2159033, EPI_ISL_2159035, EPI_ISL_2159036, EPI_ISL_2159038, EPI_ISL_2159039, EPI_ISL_2159041, EPI_ISL_2159042, EPI_ISL_2159044, EPI_ISL_2159047, EPI_ISL_2159050, EPI_ISL_2159051, EPI_ISL_2159055, EPI_ISL_2159056, EPI_ISL_2159058, EPI_ISL_2159059, EPI_ISL_2159061, EPI_ISL_2159063, EPI_ISL_2159064, EPI_ISL_2159067, EPI_ISL_2159069, EPI_ISL_2159070, EPI_ISL_2159072, EPI_ISL_2159073, EPI_ISL_2159075, EPI_ISL_2159076, EPI_ISL_2159078, EPI_ISL_2159080, EPI_ISL_2159081, EPI_ISL_2159083, EPI_ISL_2159084, EPI_ISL_2159086, EPI_ISL_2159088, EPI_ISL_2159090, EPI_ISL_2159091, EPI_ISL_2159093, EPI_ISL_2159095, EPI_ISL_2159096, EPI_ISL_2159098, EPI_ISL_2159099, EPI_ISL_2159101, EPI_ISL_2159102, EPI_ISL_2159104, EPI_ISL_2159105, EPI_ISL_2159107, EPI_ISL_2159108, EPI_ISL_2159110, EPI_ISL_2159111, EPI_ISL_2159113, EPI_ISL_2159114, EPI_ISL_3105418, EPI_ISL_3105423, EPI_ISL_3105424, EPI_ISL_3105425, EPI_ISL_3105430, EPI_ISL_3105443, EPI_ISL_3105448, EPI_ISL_3105451, EPI_ISL_3105455, EPI_ISL_3105461, EPI_ISL_3105465, EPI_ISL_3105480, EPI_ISL_3105486, EPI_ISL_3105494, EPI_ISL_3105504, EPI_ISL_3105506, EPI_ISL_3105512, EPI_ISL_3105526, EPI_ISL_3105527

| | | | |
|---|---|---|---|
| see above | Yale Clinical Virology Lab | Yale Center for Genomic Analysis | Brooke Sullivan; Christopher Castaldi; Curt Scharfe; Irina Tikhonova; Kaya Bilguvar; Shrikant Mane |

EPI_ISL_779151, EPI_ISL_1091800, EPI_ISL_1091804, EPI_ISL_1091814, EPI_ISL_1091815, EPI_ISL_1091816, EPI_ISL_1091825, EPI_ISL_1091832

| | | | |
|---|---|---|---|
| see above | Yale Clinical Virology Laboratory | Grubaugh Lab - Yale School of Public Health | Adam Moore; Akiko Iwasaki; Albert Ko; Alice Lu; Allison Nelson; Anderson Brito; Anne Wyllie; Annie Watkins; Arnau Casanovas; Catherine Muenker; Chaney Kalinich; Chantal Vogels; Charlese Dela Cruz; Isabel Ott; Joseph Fauver; Mallery Breban; Maria Tokuyama; Mary Petrone; Nathan Grubaugh; Patrick Wong; Peiwen Lu; Richard Martinello; Saad Omer; Shelli Farhadian; Tara Alpert |
| EPI_ISL_847832, EPI_ISL_847833, EPI_ISL_847837 | Yale New Haven Hospital | Grubaugh Lab - Yale School of Public Health | Anderson Brito; Anne Wyllie; Chaney Kalinich; Chantal Vogels; Isabel Ott; Joseph Fauver; Mallery Breban; Mary Petrone; Nathan Grubaugh; Tara Alpert |

EPI_ISL_779153, EPI_ISL_779154, EPI_ISL_861734, EPI_ISL_861736, EPI_ISL_861745, EPI_ISL_861764, EPI_ISL_861767, EPI_ISL_861768, EPI_ISL_861770, EPI_ISL_912167, EPI_ISL_912168, EPI_ISL_912179, EPI_ISL_912180, EPI_ISL_912181, EPI_ISL_912182, EPI_ISL_912183, EPI_ISL_912184, EPI_ISL_1009125, EPI_ISL_1009126, EPI_ISL_1038998, EPI_ISL_1038999, EPI_ISL_1039000, EPI_ISL_1168762, EPI_ISL_1168763, EPI_ISL_1168764, EPI_ISL_1168765, EPI_ISL_1225613, EPI_ISL_1225662, EPI_ISL_1293089, EPI_ISL_1293090, EPI_ISL_1293091, EPI_ISL_1293092, EPI_ISL_1293144, EPI_ISL_1293145, EPI_ISL_1293146, EPI_ISL_1293147, EPI_ISL_1293148, EPI_ISL_1378814, EPI_ISL_1378815, EPI_ISL_1378816, EPI_ISL_1378817, EPI_ISL_1378818, EPI_ISL_1378819, EPI_ISL_1378820, EPI_ISL_1378821, EPI_ISL_1447671, EPI_ISL_2023214, EPI_ISL_2023215, EPI_ISL_2023216, EPI_ISL_2023217, EPI_ISL_2159673, EPI_ISL_2159675, EPI_ISL_2296467, EPI_ISL_2296468, EPI_ISL_2296469, EPI_ISL_2296470, EPI_ISL_2426142, EPI_ISL_2776219, EPI_ISL_2960104, EPI_ISL_2960105, EPI_ISL_2960106, EPI_ISL_2960107, EPI_ISL_3025444, EPI_ISL_3025445, EPI_ISL_3104793, EPI_ISL_3105513, EPI_ISL_3105524, EPI_ISL_3236573, EPI_ISL_3236574, EPI_ISL_3236575, EPI_ISL_3236576, EPI_ISL_3236577, EPI_ISL_3236578, EPI_ISL_3236579, EPI_ISL_3236580, EPI_ISL_3236581, EPI_ISL_3236582, EPI_ISL_3236583, EPI_ISL_3236584, EPI_ISL_3347354, EPI_ISL_3347355, EPI_ISL_3347356, EPI_ISL_3347357, EPI_ISL_3347358, EPI_ISL_3347359, EPI_ISL_3347360, EPI_ISL_3347361, EPI_ISL_3347362, EPI_ISL_3347363, EPI_ISL_3347364, EPI_ISL_3347365, EPI_ISL_3347366

| | | | |
|---|---|---|---|
| see above | Yale Pathology Lab | Grubaugh Lab - Yale School of Public Health | Adam Moore; Akiko Iwasaki; Albert Ko; Alice Lu; Allison Nelson; Anderson Brito; Anne Wyllie; Annie Watkins; Arnau Casanovas; Catherine Muenker; Chaney Kalinich; Chantal Vogels; Charlese Dela Cruz; Chen Liu; Isabel Ott; Isabell Ott; Jessica Rothman; Jianhui Wang; Joseph Fauver; Kendall Billig; Mallery Breban; Maria Tokuyama; Mary Petrone; Nathan Grubaugh; Patrick Wong; Pei Hui; Peiwen Lu; Richard Martinello; Saad Omer; Shelli Farhadian; Susan Bell and Han Zhou; Tara Alpert; Tobias Koch |
