## Supplementary material for "Comparative transmissibility of SARS-CoV-2 variants Delta and Alpha in New England, USA": DataS1-7: DataS3_table_Massachusetts-1.pdf

We gratefully acknowledge the following Authors from the Originating laboratories responsible for obtaining the specimens, as well as the Submitting laboratories where the genome data were generated and shared via GISAID, on which this research is based.

All Submitters of data may be contacted directly via www.gisaid.org

Authors are sorted alphabetically.

| Accession ID | Originating Laboratory | Submitting Laboratory | Authors |
|---|---|---|---|
| EPI_ISL_1445391, EPI_ISL_1445393, EPI_ISL_1445437, EPI_ISL_1445476, EPI_ISL_1445477, EPI_ISL_1445479, EPI_ISL_1445492, EPI_ISL_1445513, EPI_ISL_1445536, EPI_ISL_1445606, EPI_ISL_1445645, EPI_ISL_1445648, EPI_ISL_1445708, EPI_ISL_1445711, EPI_ISL_1445718, EPI_ISL_1445770, EPI_ISL_1445854, EPI_ISL_1445864, EPI_ISL_1445911, EPI_ISL_1445925, EPI_ISL_1445936, EPI_ISL_1446022, EPI_ISL_1446109, EPI_ISL_1446146, EPI_ISL_1446169, EPI_ISL_1446178, EPI_ISL_1446180, EPI_ISL_1479510, EPI_ISL_1479540, EPI_ISL_1479559, EPI_ISL_1479572, EPI_ISL_1479580, EPI_ISL_1479621, EPI_ISL_1479622, EPI_ISL_1479623, EPI_ISL_1479625, EPI_ISL_1479803, EPI_ISL_1479804, EPI_ISL_1479809, EPI_ISL_1479826, EPI_ISL_1479857, EPI_ISL_1491749, EPI_ISL_1491753, EPI_ISL_1491763, EPI_ISL_1491832, EPI_ISL_1491844, EPI_ISL_1491857, EPI_ISL_1491858, EPI_ISL_1491864, EPI_ISL_1491919, EPI_ISL_1493546, EPI_ISL_1493549, EPI_ISL_1513157, EPI_ISL_1513196, EPI_ISL_1513211, EPI_ISL_1513232, EPI_ISL_1513237, EPI_ISL_1513250, EPI_ISL_1513367, EPI_ISL_1513369, EPI_ISL_1513382, EPI_ISL_1513394, EPI_ISL_1513399, EPI_ISL_1513425, EPI_ISL_1513493, EPI_ISL_1513516, EPI_ISL_1513531, EPI_ISL_1513559, EPI_ISL_1513628, EPI_ISL_1513629, EPI_ISL_1525052, EPI_ISL_1525071, EPI_ISL_1525103, EPI_ISL_1525138, EPI_ISL_1525173, EPI_ISL_1525212, EPI_ISL_1525214, EPI_ISL_1525227, EPI_ISL_1525240, EPI_ISL_1550393, EPI_ISL_1550429, EPI_ISL_1550493, EPI_ISL_1550648, EPI_ISL_1550665, EPI_ISL_1550667, EPI_ISL_1550698, EPI_ISL_1550702, EPI_ISL_1550706, EPI_ISL_1550708, EPI_ISL_1550730, EPI_ISL_1550758, EPI_ISL_1550762, EPI_ISL_1550830, EPI_ISL_1550862, EPI_ISL_1550991, EPI_ISL_1550994, EPI_ISL_1550995, EPI_ISL_1550996, EPI_ISL_1551011, EPI_ISL_1551040, EPI_ISL_1551041, EPI_ISL_1551086, EPI_ISL_1551088, EPI_ISL_1551089, EPI_ISL_1551114, EPI_ISL_1551198, EPI_ISL_1551319, EPI_ISL_1551363, EPI_ISL_1551374, EPI_ISL_1551375, EPI_ISL_1551415, EPI_ISL_1551470, EPI_ISL_1551509, EPI_ISL_1551555, EPI_ISL_1551567, EPI_ISL_1551568, EPI_ISL_1551569, EPI_ISL_1551570, EPI_ISL_1551579, EPI_ISL_1551584, EPI_ISL_1551635, EPI_ISL_1551638, EPI_ISL_1559763, EPI_ISL_1559779, EPI_ISL_1559780, EPI_ISL_1559781, EPI_ISL_1559782, EPI_ISL_1559783, EPI_ISL_1559785, EPI_ISL_1559786, EPI_ISL_1559836, EPI_ISL_1559851, EPI_ISL_1559928, EPI_ISL_1559998, EPI_ISL_1560112, EPI_ISL_1560114, EPI_ISL_1560260, EPI_ISL_1560276, EPI_ISL_1560294, EPI_ISL_1560295, EPI_ISL_1560296, EPI_ISL_1560375, EPI_ISL_1560485, EPI_ISL_1560486, EPI_ISL_1560488, EPI_ISL_1560531, EPI_ISL_1560532, EPI_ISL_1560598, EPI_ISL_1560599, EPI_ISL_1560640, EPI_ISL_1560642, EPI_ISL_1560686, EPI_ISL_1560694, EPI_ISL_1560723, EPI_ISL_1560724, EPI_ISL_1560725, EPI_ISL_1560742, EPI_ISL_1560754, EPI_ISL_1560894, EPI_ISL_1561035, EPI_ISL_1561036, EPI_ISL_1561143, EPI_ISL_1561300, EPI_ISL_1561307, EPI_ISL_1561308, EPI_ISL_1561323, EPI_ISL_1561351, EPI_ISL_1561360, EPI_ISL_1561399, EPI_ISL_1561446, EPI_ISL_1561450, EPI_ISL_1561451, EPI_ISL_1561505, EPI_ISL_1561510, EPI_ISL_1561511, EPI_ISL_1561516, EPI_ISL_1561718, EPI_ISL_1561827, EPI_ISL_1561850, EPI_ISL_1561881, EPI_ISL_1561915, EPI_ISL_1561937, EPI_ISL_1562048, EPI_ISL_1562052, EPI_ISL_1562095, EPI_ISL_1562100, EPI_ISL_1562138, EPI_ISL_1562140, EPI_ISL_1562163, EPI_ISL_1562164, EPI_ISL_1562178, EPI_ISL_1562200, EPI_ISL_1562204, EPI_ISL_1562241, EPI_ISL_1562249, EPI_ISL_1562253, EPI_ISL_1562256, EPI_ISL_1562290, EPI_ISL_1562307, EPI_ISL_1562361, EPI_ISL_1562374, EPI_ISL_1562376, EPI_ISL_1562382, EPI_ISL_1562482, EPI_ISL_1562501, EPI_ISL_1562598, EPI_ISL_1562610, EPI_ISL_1562611, EPI_ISL_1562613, EPI_ISL_1562635, EPI_ISL_1562636, EPI_ISL_1562637, EPI_ISL_1562645, EPI_ISL_1562646, EPI_ISL_1562651, EPI_ISL_1562654, EPI_ISL_1562679, EPI_ISL_1562703, EPI_ISL_1562706, EPI_ISL_1562707, EPI_ISL_1562708, EPI_ISL_1562709, EPI_ISL_1562711, EPI_ISL_1562720, EPI_ISL_1562739, EPI_ISL_1562793, EPI_ISL_1562795, EPI_ISL_1562826, EPI_ISL_1562938, EPI_ISL_1562963, EPI_ISL_1562965, EPI_ISL_1562966, EPI_ISL_1562972, EPI_ISL_1562973, EPI_ISL_1562974, EPI_ISL_1562985, EPI_ISL_1563016, EPI_ISL_1563017, EPI_ISL_1563074, EPI_ISL_1563078, EPI_ISL_1563108, EPI_ISL_1563123, EPI_ISL_1563126, EPI_ISL_1563127, EPI_ISL_1563143, EPI_ISL_1563165, EPI_ISL_1563166, EPI_ISL_1563228, EPI_ISL_1563369, EPI_ISL_1563412, EPI_ISL_1563565, EPI_ISL_1563601, EPI_ISL_1563604, EPI_ISL_1648978, EPI_ISL_1648990, EPI_ISL_1649127, EPI_ISL_1649160, EPI_ISL_1649161, EPI_ISL_1649166, EPI_ISL_1649177, EPI_ISL_1649212, EPI_ISL_1649242, EPI_ISL_1649255, EPI_ISL_1649302, EPI_ISL_1649308, EPI_ISL_1649310, EPI_ISL_1649317, EPI_ISL_1649356, EPI_ISL_1649472, EPI_ISL_1649526, EPI_ISL_1649528, EPI_ISL_1649557, EPI_ISL_1649567, EPI_ISL_1649655, EPI_ISL_1649667, EPI_ISL_1649670, EPI_ISL_1649671, EPI_ISL_1649672, EPI_ISL_1649678, EPI_ISL_1649695, EPI_ISL_1649740, EPI_ISL_1649741, EPI_ISL_1649746, EPI_ISL_1649748, EPI_ISL_1649751, EPI_ISL_1649752, EPI_ISL_1649754, EPI_ISL_1649790, EPI_ISL_1649819, EPI_ISL_1649843, EPI_ISL_1649847, EPI_ISL_1649880, EPI_ISL_1649936, EPI_ISL_1649939, EPI_ISL_1649942, EPI_ISL_1649965, EPI_ISL_1649966, EPI_ISL_1649967, EPI_ISL_1649983, EPI_ISL_1650017, EPI_ISL_1650019, EPI_ISL_1650048, EPI_ISL_1650070, EPI_ISL_1650115, EPI_ISL_1650154, EPI_ISL_1650239, EPI_ISL_1650262, EPI_ISL_1650302, EPI_ISL_1650348, EPI_ISL_1650381, EPI_ISL_1650382, EPI_ISL_1650386, EPI_ISL_1650398, EPI_ISL_1650399, EPI_ISL_1650450, EPI_ISL_1650474, EPI_ISL_1650488, EPI_ISL_1650489, EPI_ISL_1650533, EPI_ISL_1650549, EPI_ISL_1650613, EPI_ISL_1650619, EPI_ISL_1650621, EPI_ISL_1650628, EPI_ISL_1650646, EPI_ISL_1650649, EPI_ISL_1650650, EPI_ISL_1650652, EPI_ISL_1650653, EPI_ISL_1650690, EPI_ISL_1650694, EPI_ISL_1650739, EPI_ISL_1650741, EPI_ISL_1650742, EPI_ISL_1650804, EPI_ISL_1650828, EPI_ISL_1650901, EPI_ISL_1650955, EPI_ISL_1651044, EPI_ISL_1651062, EPI_ISL_1651063, EPI_ISL_1651064, EPI_ISL_1651140, EPI_ISL_1667456, EPI_ISL_1667467, EPI_ISL_1667468, EPI_ISL_1667470, EPI_ISL_1667477, EPI_ISL_1667499, EPI_ISL_1667511, EPI_ISL_1667532, EPI_ISL_1667533, EPI_ISL_1667545, EPI_ISL_1667550, EPI_ISL_1667551, EPI_ISL_1667575, EPI_ISL_1667626, EPI_ISL_1667641, EPI_ISL_1667649, EPI_ISL_1667669, EPI_ISL_1667670, EPI_ISL_1667681, EPI_ISL_1667718, EPI_ISL_1667806, EPI_ISL_1667921, EPI_ISL_1668018, EPI_ISL_1668162, EPI_ISL_1668166, EPI_ISL_1668168, EPI_ISL_1668187, EPI_ISL_1668196, EPI_ISL_1668315, EPI_ISL_1668346, EPI_ISL_1668443, EPI_ISL_1685486, EPI_ISL_1685489, EPI_ISL_1685522, EPI_ISL_1685576, EPI_ISL_1685577, EPI_ISL_1685730, EPI_ISL_1685817, EPI_ISL_1685818, EPI_ISL_1685836, EPI_ISL_1685837, EPI_ISL_1685861, EPI_ISL_1685893, EPI_ISL_1685897, EPI_ISL_1685899, EPI_ISL_1685900, EPI_ISL_1685905, EPI_ISL_1685932, EPI_ISL_1685986, EPI_ISL_1685987, EPI_ISL_1685988, EPI_ISL_1685996, EPI_ISL_1686116, EPI_ISL_1686172, EPI_ISL_1686180, EPI_ISL_1686230, EPI_ISL_1686375, EPI_ISL_1686380, EPI_ISL_1686574, EPI_ISL_1686861, EPI_ISL_1686864, EPI_ISL_1686893, EPI_ISL_1686902, EPI_ISL_1687005, EPI_ISL_1687226, EPI_ISL_1687238, EPI_ISL_1687270, EPI_ISL_1687288, EPI_ISL_1687296, EPI_ISL_1687298, EPI_ISL_1687317, EPI_ISL_1687320, EPI_ISL_1687324, EPI_ISL_1687344, EPI_ISL_1687395, EPI_ISL_1687396, EPI_ISL_1687412, EPI_ISL_1687425, EPI_ISL_1687439, EPI_ISL_1687458, EPI_ISL_1687459, EPI_ISL_1687462, EPI_ISL_1687463, EPI_ISL_1687584, EPI_ISL_1687585, EPI_ISL_1687586, EPI_ISL_1687694, EPI_ISL_1687721, EPI_ISL_1687784, EPI_ISL_1687805, EPI_ISL_1687939, EPI_ISL_1687962, EPI_ISL_1687966, EPI_ISL_1687973, EPI_ISL_1687978, EPI_ISL_1687985, EPI_ISL_1687996, EPI_ISL_1688000, EPI_ISL_1688008, EPI_ISL_1688013, EPI_ISL_1688015, EPI_ISL_1688016, EPI_ISL_1688091, EPI_ISL_1688099, EPI_ISL_1688107, EPI_ISL_1688143, EPI_ISL_1688182, EPI_ISL_1688183, EPI_ISL_1688228, EPI_ISL_1688230, EPI_ISL_1688258, EPI_ISL_1688259, EPI_ISL_1688281, EPI_ISL_1688297, EPI_ISL_1688333, EPI_ISL_1688416, EPI_ISL_1688425, EPI_ISL_1688435, EPI_ISL_1688455, EPI_ISL_1688465, EPI_ISL_1688467, EPI_ISL_1688468, EPI_ISL_1688471, EPI_ISL_1688472, EPI_ISL_1688570, EPI_ISL_1688632, EPI_ISL_1688940, EPI_ISL_1688944, EPI_ISL_1688949, EPI_ISL_1688970, EPI_ISL_1688973, EPI_ISL_1688974, EPI_ISL_1688977, EPI_ISL_1688979, EPI_ISL_1689127, EPI_ISL_1689133, EPI_ISL_1689144, EPI_ISL_1689146, EPI_ISL_1689150, EPI_ISL_1689245, EPI_ISL_1689305, EPI_ISL_1689718, EPI_ISL_1689786, EPI_ISL_1689826, EPI_ISL_1689828, EPI_ISL_1689830, EPI_ISL_1689831, EPI_ISL_1689834, EPI_ISL_1689835, EPI_ISL_1689836, EPI_ISL_1689838, EPI_ISL_1689849, EPI_ISL_1689850, EPI_ISL_1689872, EPI_ISL_1689876, EPI_ISL_1689940, EPI_ISL_1689943, EPI_ISL_1689970, EPI_ISL_1690160, EPI_ISL_1690169, EPI_ISL_1695111, EPI_ISL_1695131, EPI_ISL_1695139, EPI_ISL_1695141, EPI_ISL_1695164, EPI_ISL_1695167, EPI_ISL_1695299, EPI_ISL_1695425, EPI_ISL_1701934, EPI_ISL_1701945, EPI_ISL_1701961, EPI_ISL_1701998, EPI_ISL_1702015, EPI_ISL_1702019, EPI_ISL_1702060, EPI_ISL_1702074, EPI_ISL_1702091, EPI_ISL_1702103, EPI_ISL_1702125, EPI_ISL_1702131, EPI_ISL_1702752, EPI_ISL_1702785, EPI_ISL_1702850, EPI_ISL_1702879, EPI_ISL_1735746, EPI_ISL_1735764, EPI_ISL_1735768, EPI_ISL_1735769, EPI_ISL_1735780, EPI_ISL_1735861, EPI_ISL_1735863, EPI_ISL_1735864, EPI_ISL_1735878, EPI_ISL_1735882, EPI_ISL_1735901, EPI_ISL_1735942, EPI_ISL_1735951, EPI_ISL_1735953, EPI_ISL_1735954, EPI_ISL_1736158, EPI_ISL_1736179, EPI_ISL_1736181, EPI_ISL_1736204, EPI_ISL_1736210, EPI_ISL_1736211, EPI_ISL_1736213, EPI_ISL_1736214, EPI_ISL_1736222, EPI_ISL_1736223, EPI_ISL_1736224, EPI_ISL_1736226, EPI_ISL_1736227, EPI_ISL_1736229, EPI_ISL_1736230, EPI_ISL_1736233, EPI_ISL_1736242, EPI_ISL_1736244, EPI_ISL_1736245, EPI_ISL_1736247, EPI_ISL_1736406, EPI_ISL_1736442, EPI_ISL_1736454, EPI_ISL_1736467, EPI_ISL_1736469, EPI_ISL_1736475, EPI_ISL_1736542, EPI_ISL_1736547, EPI_ISL_1736703, EPI_ISL_1736712, EPI_ISL_1736715, EPI_ISL_1736717, EPI_ISL_1736801, EPI_ISL_1736808, EPI_ISL_1736814, EPI_ISL_1736846, EPI_ISL_1736859, EPI_ISL_1736932, EPI_ISL_1736933, EPI_ISL_1736937, EPI_ISL_1737029, EPI_ISL_1737037, EPI_ISL_1737068, EPI_ISL_1737069, EPI_ISL_1737090, EPI_ISL_1737184, EPI_ISL_1737242, EPI_ISL_1737243, EPI_ISL_1737244, EPI_ISL_1737250, EPI_ISL_1737288, EPI_ISL_1737339, EPI_ISL_1737358, EPI_ISL_1737376, EPI_ISL_1737380, EPI_ISL_1737390, EPI_ISL_1737393, EPI_ISL_1737415, EPI_ISL_1737457, EPI_ISL_1737519, EPI_ISL_1737594, EPI_ISL_1737595, EPI_ISL_1737623, EPI_ISL_1737624, EPI_ISL_1737625, EPI_ISL_1737627, EPI_ISL_1737629, EPI_ISL_1737630, EPI_ISL_1737754, EPI_ISL_1737757, EPI_ISL_1737803, EPI_ISL_1738779, EPI_ISL_1753545, EPI_ISL_1753599, EPI_ISL_1834445, EPI_ISL_1834447, EPI_ISL_1834452, EPI_ISL_1834464, EPI_ISL_1834476, EPI_ISL_1834497, EPI_ISL_1834498, EPI_ISL_1834508, EPI_ISL_1834578, EPI_ISL_1834649, EPI_ISL_1834775, EPI_ISL_1834789, EPI_ISL_1834859, EPI_ISL_1834860, EPI_ISL_1834885, EPI_ISL_1834897, EPI_ISL_1834956, EPI_ISL_1835148, EPI_ISL_1835179, EPI_ISL_1835180, EPI_ISL_1835208, EPI_ISL_1835281, EPI_ISL_1835282, EPI_ISL_1835313, EPI_ISL_1835314, EPI_ISL_1835320, EPI_ISL_1835345, EPI_ISL_1835349, EPI_ISL_1835365, EPI_ISL_1835366, EPI_ISL_1835400, EPI_ISL_1835402, EPI_ISL_1835403, EPI_ISL_1835409, EPI_ISL_1835413, EPI_ISL_1835416, EPI_ISL_1835424, EPI_ISL_1835426, EPI_ISL_1835427, EPI_ISL_1835430, EPI_ISL_1835469, EPI_ISL_1835474, EPI_ISL_1835505, EPI_ISL_1835579, EPI_ISL_1835600, EPI_ISL_1835664, EPI_ISL_1835692, EPI_ISL_1835738, EPI_ISL_1835784, EPI_ISL_1835913, EPI_ISL_1835915, EPI_ISL_1835917, EPI_ISL_1835954, EPI_ISL_1835980, EPI_ISL_1835983, EPI_ISL_1835985, EPI_ISL_1836006, EPI_ISL_1836007, EPI_ISL_1836008, EPI_ISL_1836011, EPI_ISL_1836036, EPI_ISL_1836063, EPI_ISL_1836114, EPI_ISL_1836131, EPI_ISL_1836141, EPI_ISL_1836147, EPI_ISL_1836176, EPI_ISL_1836177, EPI_ISL_1836185, EPI_ISL_1836207, EPI_ISL_1836208, EPI_ISL_1836211, EPI_ISL_1836271, EPI_ISL_1836302, EPI_ISL_1836357, EPI_ISL_1836358, EPI_ISL_1836360, EPI_ISL_1836361, EPI_ISL_1836407, EPI_ISL_1836408, EPI_ISL_1836410, EPI_ISL_1836415, EPI_ISL_1836435, EPI_ISL_1836436, EPI_ISL_1836446, EPI_ISL_1836531, EPI_ISL_1836670, EPI_ISL_1836705, EPI_ISL_1836708, EPI_ISL_1836770, EPI_ISL_1836804, EPI_ISL_1836943, EPI_ISL_1836962, EPI_ISL_1836991, EPI_ISL_1837054, EPI_ISL_1837069, EPI_ISL_1837136, EPI_ISL_1837309, EPI_ISL_1837398, EPI_ISL_1837421, EPI_ISL_1837424, EPI_ISL_1837426, EPI_ISL_1837511, EPI_ISL_1837547, EPI_ISL_1837586, EPI_ISL_1837590, EPI_ISL_1837592, EPI_ISL_1837608, EPI_ISL_1837612, EPI_ISL_1837678, EPI_ISL_1837695, EPI_ISL_1837713, EPI_ISL_1837783, EPI_ISL_1837893, EPI_ISL_1837897, EPI_ISL_1837899, EPI_ISL_1837930, EPI_ISL_1837960, EPI_ISL_1837976, EPI_ISL_1838025, EPI_ISL_1838030, EPI_ISL_1838041, EPI_ISL_1838053, EPI_ISL_1838058, EPI_ISL_1838135, EPI_ISL_1838137, EPI_ISL_1838138, EPI_ISL_1838146, EPI_ISL_1838258, EPI_ISL_1838283, EPI_ISL_1838447, EPI_ISL_1900181, EPI_ISL_1900246, EPI_ISL_1900295, EPI_ISL_1900330, EPI_ISL_1900345, EPI_ISL_1900393, EPI_ISL_1900397, EPI_ISL_1922914, EPI_ISL_1922963, EPI_ISL_1923105, EPI_ISL_1923120, EPI_ISL_1925095, EPI_ISL_1925137, EPI_ISL_1925141, EPI_ISL_1925143, EPI_ISL_1925197, EPI_ISL_1925201, EPI_ISL_1925209, EPI_ISL_1925256, EPI_ISL_1925271, EPI_ISL_1925272, EPI_ISL_1925274, EPI_ISL_1925276, EPI_ISL_1925277, EPI_ISL_1925280, EPI_ISL_1925313, EPI_ISL_1925400, EPI_ISL_1925405, EPI_ISL_1925446, EPI_ISL_1925457, EPI_ISL_1925497, EPI_ISL_1925516, EPI_ISL_1925517, EPI_ISL_1925520, EPI_ISL_1942223, EPI_ISL_1942228, EPI_ISL_1995031, EPI_ISL_1995087, EPI_ISL_1995106, EPI_ISL_1995138, EPI_ISL_1995171, EPI_ISL_1995211, EPI_ISL_1995254, EPI_ISL_1995263, EPI_ISL_1995270, EPI_ISL_1995273, EPI_ISL_1995274, EPI_ISL_1995275, EPI_ISL_1995276, EPI_ISL_1995293, EPI_ISL_1995294, EPI_ISL_1995295, EPI_ISL_1995308, EPI_ISL_1995313, EPI_ISL_1995317, EPI_ISL_1995318, EPI_ISL_1995319, EPI_ISL_1995320, EPI_ISL_1995321, EPI_ISL_1995327, EPI_ISL_1995359, EPI_ISL_1995360, EPI_ISL_1995361, EPI_ISL_1995362, EPI_ISL_1995365, EPI_ISL_1995535, EPI_ISL_1995573, EPI_ISL_1995587, EPI_ISL_1995765, EPI_ISL_1995824, EPI_ISL_1995979, EPI_ISL_1996121, EPI_ISL_1996123, EPI_ISL_1996125, EPI_ISL_1996128, EPI_ISL_1996129, EPI_ISL_1996130, EPI_ISL_1996134, EPI_ISL_1996151, EPI_ISL_1996218, EPI_ISL_1996269, EPI_ISL_1996270, EPI_ISL_1996276, EPI_ISL_1996290, EPI_ISL_1996439, EPI_ISL_1996624, EPI_ISL_1996627, EPI_ISL_1996628, EPI_ISL_1996629, EPI_ISL_1996630, EPI_ISL_1996631, EPI_ISL_1996632, EPI_ISL_1996633, EPI_ISL_1996635, EPI_ISL_1996636, EPI_ISL_1996637, EPI_ISL_1996638, EPI_ISL_1996640, EPI_ISL_1996934, EPI_ISL_1996943, EPI_ISL_1997311, EPI_ISL_1997336, EPI_ISL_1997337, EPI_ISL_1997338, EPI_ISL_1997341, EPI_ISL_1997342, EPI_ISL_1997343, EPI_ISL_1997344, EPI_ISL_1997345, EPI_ISL_1997347, EPI_ISL_1997349, EPI_ISL_1997350, EPI_ISL_1997351, EPI_ISL_1997355, EPI_ISL_1997356, EPI_ISL_1997357, EPI_ISL_1997422, EPI_ISL_1998021, EPI_ISL_1998023, EPI_ISL_1998027, EPI_ISL_1998056, EPI_ISL_1998058, EPI_ISL_1998059, EPI_ISL_1998062, EPI_ISL_1998091, EPI_ISL_1998106, EPI_ISL_1998107, EPI_ISL_1998223, EPI_ISL_1998241, EPI_ISL_1998346, EPI_ISL_1998448, EPI_ISL_1998450, EPI_ISL_1998462, EPI_ISL_1998487, EPI_ISL_1998500, EPI_ISL_1998669, EPI_ISL_1998694, EPI_ISL_1998713, EPI_ISL_1998723, EPI_ISL_1998754, EPI_ISL_1998798, EPI_ISL_1998866, EPI_ISL_1998996, EPI_ISL_1998997, EPI_ISL_1999005, EPI_ISL_1999136, EPI_ISL_1999137, EPI_ISL_1999158, EPI_ISL_1999166, EPI_ISL_1999180, EPI_ISL_1999202, EPI_ISL_1999205, EPI_ISL_1999208, EPI_ISL_1999242, EPI_ISL_1999252, EPI_ISL_1999313, EPI_ISL_1999348, EPI_ISL_1999349, EPI_ISL_1999352, EPI_ISL_1999465, EPI_ISL_1999566, EPI_ISL_1999571, EPI_ISL_1999631, EPI_ISL_1999722, EPI_ISL_1999752, EPI_ISL_1999906, EPI_ISL_1999911, EPI_ISL_1999932, EPI_ISL_1999935, EPI_ISL_1999977, EPI_ISL_1999985, EPI_ISL_1999986, EPI_ISL_1999987, EPI_ISL_1999988, EPI_ISL_1999991, EPI_ISL_1999992, EPI_ISL_1999993, EPI_ISL_1999995, EPI_ISL_2000052, EPI_ISL_2000056, EPI_ISL_2000059, EPI_ISL_2000133, EPI_ISL_2000147, EPI_ISL_2000165, EPI_ISL_2000176, EPI_ISL_2000189, EPI_ISL_2000200, EPI_ISL_2000240, EPI_ISL_2000264, EPI_ISL_2000265, EPI_ISL_2000269, EPI_ISL_2000276, EPI_ISL_2000307, EPI_ISL_2000324, EPI_ISL_2000336, EPI_ISL_2000373, EPI_ISL_2000427, EPI_ISL_2000474, EPI_ISL_2000479, EPI_ISL_2000480, EPI_ISL_2000499 | | | |
| see above | Aegis Sciences Corporation | Centers for Disease Control and Prevention Division of Viral Diseases, Pathogen Discovery | Adrian Paskey; Alec Vest; Benjamin Rambo-Martin; Christopher Gulvick; Clinton R. Paden; Cyndi Clark; Dakota Howard; Darlene Wagner; Dhwani Batra; Dillon Nall; Duncan MacCannell; Ethan Sanders; Holly Houdeshell; Jason Caravas; Kara Moser; Matthew Hardison; Matthew Schmerer; Ola Kvalvaag; Patrick Campbell; Peter W. Cook; Rob Case; Scott Sammons; Shatavia Morrison; Shaun Westlund; Vikramsinha Ghorpade; Yvette Unoarumhi |
| EPI_ISL_976941, EPI_ISL_976942, EPI_ISL_976943, EPI_ISL_976944, EPI_ISL_976945, EPI_ISL_976946, EPI_ISL_976947, EPI_ISL_976948, EPI_ISL_976949, EPI_ISL_976950, EPI_ISL_976951, EPI_ISL_976952, EPI_ISL_976953, EPI_ISL_976954, EPI_ISL_976955, EPI_ISL_976956, EPI_ISL_976957, EPI_ISL_976958, EPI_ISL_976959, EPI_ISL_976960, EPI_ISL_976961, EPI_ISL_976962, EPI_ISL_976963, EPI_ISL_976964, EPI_ISL_976965, EPI_ISL_976966, EPI_ISL_976967, EPI_ISL_976968, EPI_ISL_976969, EPI_ISL_976970, EPI_ISL_976971, EPI_ISL_976972, EPI_ISL_976973, EPI_ISL_976974, EPI_ISL_976975, EPI_ISL_976976, EPI_ISL_976977, EPI_ISL_976978, EPI_ISL_976979, EPI_ISL_976980, EPI_ISL_976982, EPI_ISL_976983, EPI_ISL_976984, EPI_ISL_976985, EPI_ISL_976986, EPI_ISL_976987, EPI_ISL_976988, EPI_ISL_976989, EPI_ISL_976990, EPI_ISL_976991, EPI_ISL_976992, EPI_ISL_976993, EPI_ISL_976994, EPI_ISL_976995, EPI_ISL_976996, EPI_ISL_976997, EPI_ISL_976998, EPI_ISL_976999, EPI_ISL_977000, EPI_ISL_977001, EPI_ISL_977002, EPI_ISL_977003, EPI_ISL_977004, EPI_ISL_977005, EPI_ISL_977006, EPI_ISL_977007, EPI_ISL_977008, EPI_ISL_977009, EPI_ISL_977010, EPI_ISL_977011, EPI_ISL_977012, EPI_ISL_977013, EPI_ISL_977014, EPI_ISL_977015, EPI_ISL_977016, EPI_ISL_977017, EPI_ISL_977018, EPI_ISL_977019, EPI_ISL_977020, EPI_ISL_977021, EPI_ISL_977022, EPI_ISL_977023, EPI_ISL_1011678, EPI_ISL_1011679, EPI_ISL_1011680, EPI_ISL_1011681, EPI_ISL_1011682, EPI_ISL_1011683, EPI_ISL_1011684, EPI_ISL_1011685, EPI_ISL_1011686, EPI_ISL_1011687, EPI_ISL_1011688, EPI_ISL_1011689, EPI_ISL_1011690, EPI_ISL_1011691, EPI_ISL_1011692, EPI_ISL_1011693, EPI_ISL_1011694, EPI_ISL_1011695, EPI_ISL_1011696, EPI_ISL_1011697, EPI_ISL_1011698, EPI_ISL_1011699, EPI_ISL_1011700, EPI_ISL_1011701, EPI_ISL_1011702, EPI_ISL_1011703, EPI_ISL_1011704, EPI_ISL_1011705, EPI_ISL_1067739, EPI_ISL_1067740, EPI_ISL_1067741, EPI_ISL_1067742, EPI_ISL_1067743, EPI_ISL_1067744, EPI_ISL_1067745, EPI_ISL_1067746, EPI_ISL_1067747, EPI_ISL_1067748, EPI_ISL_1067749, EPI_ISL_1067750, EPI_ISL_1067751, EPI_ISL_1067752, EPI_ISL_1067753, EPI_ISL_1067754, EPI_ISL_1067755, EPI_ISL_1067756, EPI_ISL_1067757, EPI_ISL_1067758, EPI_ISL_1067759, EPI_ISL_1067760, EPI_ISL_1067761, EPI_ISL_1067762, EPI_ISL_1067763, EPI_ISL_1067764, EPI_ISL_1067765, EPI_ISL_1067766, EPI_ISL_1067767, EPI_ISL_1067768, EPI_ISL_1067769, EPI_ISL_1067770, EPI_ISL_1067771, EPI_ISL_1067772, EPI_ISL_1067773, EPI_ISL_1067774, EPI_ISL_1067775, EPI_ISL_1067776, EPI_ISL_1067777, EPI_ISL_1067778, EPI_ISL_1067779, EPI_ISL_1067780, EPI_ISL_1067781, EPI_ISL_1067782, EPI_ISL_1067783, EPI_ISL_1067784, EPI_ISL_1067785, EPI_ISL_1067786, EPI_ISL_1067787, EPI_ISL_1067788, EPI_ISL_1067789, EPI_ISL_1067790, EPI_ISL_1067791, EPI_ISL_1067792, EPI_ISL_1067793, EPI_ISL_1067794, EPI_ISL_1067795, EPI_ISL_1067796, EPI_ISL_1067797, EPI_ISL_1067798, EPI_ISL_1067799, EPI_ISL_1067800, EPI_ISL_1067801, EPI_ISL_1067802, EPI_ISL_1067803, EPI_ISL_1067804, EPI_ISL_1067805, EPI_ISL_1067806, EPI_ISL_1067807, EPI_ISL_1067808, EPI_ISL_1067809, EPI_ISL_1067810, EPI_ISL_1067811, EPI_ISL_1067812, EPI_ISL_1067813, EPI_ISL_1067814, EPI_ISL_1067815, EPI_ISL_1067816, EPI_ISL_1067817, EPI_ISL_1121633, EPI_ISL_1121634, EPI_ISL_1121635, EPI_ISL_1121636, EPI_ISL_1121637, EPI_ISL_1121638, EPI_ISL_1121639, EPI_ISL_1121640, EPI_ISL_1121641, EPI_ISL_1121642, EPI_ISL_1121643, EPI_ISL_1121644, EPI_ISL_1121645, EPI_ISL_1121646, EPI_ISL_1121647, EPI_ISL_1121648, EPI_ISL_1121649, EPI_ISL_1121650, EPI_ISL_1121651, EPI_ISL_1121652, EPI_ISL_1121653, EPI_ISL_1121654, EPI_ISL_1121655, EPI_ISL_1121656, EPI_ISL_1121657, EPI_ISL_1121658, EPI_ISL_1121659, EPI_ISL_1121660, EPI_ISL_1121661, EPI_ISL_1121662, EPI_ISL_1121663, EPI_ISL_1121664, EPI_ISL_1121665, EPI_ISL_1121666, EPI_ISL_1121667, EPI_ISL_1121668, EPI_ISL_1121669, EPI_ISL_1121670, EPI_ISL_1121671, EPI_ISL_1121672, EPI_ISL_1121673, EPI_ISL_1121674, EPI_ISL_1121675, EPI_ISL_1121676, EPI_ISL_1121677, EPI_ISL_1121678, EPI_ISL_1121679, EPI_ISL_1121680, EPI_ISL_1121681, EPI_ISL_1121682, EPI_ISL_1121683, EPI_ISL_1121684, EPI_ISL_1121685, EPI_ISL_1121686, EPI_ISL_1121687, EPI_ISL_1121688, EPI_ISL_1121689, EPI_ISL_1121690, EPI_ISL_1121691, EPI_ISL_1121692, EPI_ISL_1121693, EPI_ISL_1121694, EPI_ISL_1121695, EPI_ISL_1121696, EPI_ISL_1121697, EPI_ISL_1121698, EPI_ISL_1121699, EPI_ISL_1121700, EPI_ISL_1121701, EPI_ISL_1121702, EPI_ISL_1121879, EPI_ISL_1121880, EPI_ISL_1121881, EPI_ISL_1121882, EPI_ISL_1121883, EPI_ISL_1121884, EPI_ISL_1121885, EPI_ISL_1121886, EPI_ISL_1121963, EPI_ISL_1121964, EPI_ISL_1121965, EPI_ISL_1121966, EPI_ISL_1121967, EPI_ISL_1121968, EPI_ISL_1121973, EPI_ISL_1253719, EPI_ISL_1253720, EPI_ISL_1253722, EPI_ISL_1253723, EPI_ISL_1253724, EPI_ISL_1253725, EPI_ISL_1253727, EPI_ISL_1253728, EPI_ISL_1253729, EPI_ISL_1253730, EPI_ISL_1253731, EPI_ISL_1253732, EPI_ISL_1253733, EPI_ISL_1253734, EPI_ISL_1253735, EPI_ISL_1253736, EPI_ISL_1253739, EPI_ISL_1253740, EPI_ISL_1253741, EPI_ISL_1253742, EPI_ISL_1253743, EPI_ISL_1253744, EPI_ISL_1253745, EPI_ISL_1253747, EPI_ISL_1253748, EPI_ISL_1253749, EPI_ISL_1253750, EPI_ISL_1253752, EPI_ISL_1253753, EPI_ISL_1253755, EPI_ISL_1253756, EPI_ISL_1253757, EPI_ISL_1253758, EPI_ISL_1253759, EPI_ISL_1253760, EPI_ISL_1253761, EPI_ISL_1253762, EPI_ISL_1253763, EPI_ISL_1253765, EPI_ISL_1253767, EPI_ISL_1253768, EPI_ISL_1253769, EPI_ISL_1253770, EPI_ISL_1253771, EPI_ISL_1253772, EPI_ISL_1253773, EPI_ISL_1253774, EPI_ISL_1253775, EPI_ISL_1253776, EPI_ISL_1253778, EPI_ISL_1253780, EPI_ISL_1253781, EPI_ISL_1253782, EPI_ISL_1253784, EPI_ISL_1253785, EPI_ISL_1253787, EPI_ISL_1253788, EPI_ISL_1253789, EPI_ISL_1253790, EPI_ISL_1253791, EPI_ISL_1253792, EPI_ISL_1253793, EPI_ISL_1253794, EPI_ISL_1253795, EPI_ISL_1253796, EPI_ISL_1253797, EPI_ISL_1253798, EPI_ISL_1253799, EPI_ISL_1253800, EPI_ISL_1253801, EPI_ISL_1253802, EPI_ISL_1253803, EPI_ISL_1253804, EPI_ISL_1253805, EPI_ISL_1253806, EPI_ISL_1253807, EPI_ISL_1253808, EPI_ISL_1253809, EPI_ISL_1253810, EPI_ISL_1253811, EPI_ISL_1253812, EPI_ISL_1253813, EPI_ISL_1253814, EPI_ISL_1253815, EPI_ISL_1253816, EPI_ISL_1253817, EPI_ISL_1253818, EPI_ISL_1253819, EPI_ISL_1253820, EPI_ISL_1253821, EPI_ISL_1253822, EPI_ISL_1253823, EPI_ISL_1253824, EPI_ISL_1253825, EPI_ISL_1253826, EPI_ISL_1253827, EPI_ISL_1253828, EPI_ISL_1253829, EPI_ISL_1253830, EPI_ISL_1253831, EPI_ISL_1253832, EPI_ISL_1253833, EPI_ISL_1253834, EPI_ISL_1253835, EPI_ISL_1253836, EPI_ISL_1253837, EPI_ISL_1253838, EPI_ISL_1253839, EPI_ISL_1253840, EPI_ISL_1253841, EPI_ISL_1253842, EPI_ISL_1253843, EPI_ISL_1253844, EPI_ISL_1253845, EPI_ISL_1253846, EPI_ISL_1253847, EPI_ISL_1253848, EPI_ISL_1253849, EPI_ISL_1253850, EPI_ISL_1253851, EPI_ISL_1253852, EPI_ISL_1253853, EPI_ISL_1253854, EPI_ISL_1253855, EPI_ISL_1253856, EPI_ISL_1253857, EPI_ISL_1253858, EPI_ISL_1253859, EPI_ISL_1253860, EPI_ISL_1253861, EPI_ISL_1253862, EPI_ISL_1253863, EPI_ISL_1253864, EPI_ISL_1253865, EPI_ISL_1253866, EPI_ISL_1253867, EPI_ISL_1253868, EPI_ISL_1253869, EPI_ISL_1253870, EPI_ISL_1253871, EPI_ISL_1253872, EPI_ISL_1253873, EPI_ISL_1253874, EPI_ISL_1253875, EPI_ISL_1253876, EPI_ISL_1253877, EPI_ISL_1253878, EPI_ISL_1253879, EPI_ISL_1253880, EPI_ISL_1253881, EPI_ISL_1253882, EPI_ISL_1253883, EPI_ISL_1253884, EPI_ISL_1407254, EPI_ISL_1407255, EPI_ISL_1407256, EPI_ISL_1407258, EPI_ISL_1407259, EPI_ISL_1407260, EPI_ISL_1407261, EPI_ISL_1407262, EPI_ISL_1407263, EPI_ISL_1407264, EPI_ISL_1407266, EPI_ISL_1407267, EPI_ISL_1407268, EPI_ISL_1407269, EPI_ISL_1407271, EPI_ISL_1407273, EPI_ISL_1407274, EPI_ISL_1407275, EPI_ISL_1407276, EPI_ISL_1407277, EPI_ISL_1407278, EPI_ISL_1407279, EPI_ISL_1407280, EPI_ISL_1407281, EPI_ISL_1407282, EPI_ISL_1407283, EPI_ISL_1407285, EPI_ISL_1407286, EPI_ISL_1407287, EPI_ISL_1407288, EPI_ISL_1407291, EPI_ISL_1407292, EPI_ISL_1407293, EPI_ISL_1407294, EPI_ISL_1407295, EPI_ISL_1407296, EPI_ISL_1407297, EPI_ISL_1407298, EPI_ISL_1407299, EPI_ISL_1407300, EPI_ISL_1407301, EPI_ISL_1407302, EPI_ISL_1407303, EPI_ISL_1407304, EPI_ISL_1407305, EPI_ISL_1407306, EPI_ISL_1407307, EPI_ISL_1407308, EPI_ISL_1407309, EPI_ISL_1407312, EPI_ISL_1407314, EPI_ISL_1407315, EPI_ISL_1407316, EPI_ISL_1407318, EPI_ISL_1407319, EPI_ISL_1407320, EPI_ISL_1407321, EPI_ISL_1407322, EPI_ISL_1407323, EPI_ISL_1407325, EPI_ISL_1407327, EPI_ISL_1407328, EPI_ISL_1407329, EPI_ISL_1407330, EPI_ISL_1413366, EPI_ISL_1413369, EPI_ISL_1413380, EPI_ISL_1413385, EPI_ISL_1413388, EPI_ISL_1413390, EPI_ISL_1413393, EPI_ISL_1413399, EPI_ISL_1413401, EPI_ISL_1413404, EPI_ISL_1413407, EPI_ISL_1413410, EPI_ISL_1413413, EPI_ISL_1413416, EPI_ISL_1413418, EPI_ISL_1413421, EPI_ISL_1413424, EPI_ISL_1413426, EPI_ISL_1413429, EPI_ISL_1413431, EPI_ISL_1413434, EPI_ISL_1413437, EPI_ISL_1413439, EPI_ISL_1413442, EPI_ISL_1413445, EPI_ISL_1413450, EPI_ISL_1413452, EPI_ISL_1413455, EPI_ISL_1413458, EPI_ISL_1413460, EPI_ISL_1413463, EPI_ISL_1413465, EPI_ISL_1413468, EPI_ISL_1413471, EPI_ISL_1413479, EPI_ISL_1413482, EPI_ISL_1413490, EPI_ISL_1413493, EPI_ISL_1413501, EPI_ISL_1413504, EPI_ISL_1413506, EPI_ISL_1413509, EPI_ISL_1413511, EPI_ISL_1413789, EPI_ISL_1413794, EPI_ISL_1413797, EPI_ISL_1413799, EPI_ISL_1413803, EPI_ISL_1413805, EPI_ISL_1413807, EPI_ISL_1413810, EPI_ISL_1413813, EPI_ISL_1413815, EPI_ISL_1413818, EPI_ISL_1413821, EPI_ISL_1413823, EPI_ISL_1413826, EPI_ISL_1413829, EPI_ISL_1413831, EPI_ISL_1413835, EPI_ISL_1413837, EPI_ISL_1413840, EPI_ISL_1413842, EPI_ISL_1413845, EPI_ISL_1413848, EPI_ISL_1413851, EPI_ISL_1413853, EPI_ISL_1413856, EPI_ISL_1413859, EPI_ISL_1413861, EPI_ISL_1413864, EPI_ISL_1516476, EPI_ISL_1516477, EPI_ISL_1516478, EPI_ISL_1516479, EPI_ISL_1516480, EPI_ISL_1516482, EPI_ISL_1516485, EPI_ISL_1516488, EPI_ISL_1516489, EPI_ISL_1516491, EPI_ISL_1516492, EPI_ISL_1516493, EPI_ISL_1516494, EPI_ISL_1516495, EPI_ISL_1516496, EPI_ISL_1516498, EPI_ISL_1516502, EPI_ISL_1516503, EPI_ISL_1516504, EPI_ISL_1516505, EPI_ISL_1516506, EPI_ISL_1516507, EPI_ISL_1516508, EPI_ISL_1516509, EPI_ISL_1516510, EPI_ISL_1516511, EPI_ISL_1516512, EPI_ISL_1516513, EPI_ISL_1516514, EPI_ISL_1516515, EPI_ISL_1516516, EPI_ISL_1516517, EPI_ISL_1516518, EPI_ISL_1516520, EPI_ISL_1516521, EPI_ISL_1516522, EPI_ISL_1516596, EPI_ISL_1516597, EPI_ISL_1516665, EPI_ISL_1516666, EPI_ISL_1516668, EPI_ISL_1516669, EPI_ISL_1516670, EPI_ISL_1516671, EPI_ISL_1516672, EPI_ISL_1516673, EPI_ISL_1516674, EPI_ISL_1516675, EPI_ISL_1516676, EPI_ISL_1516677, EPI_ISL_1516678, EPI_ISL_1516680, EPI_ISL_1516681, EPI_ISL_1516683, EPI_ISL_1516684, EPI_ISL_1516685, EPI_ISL_1516686, EPI_ISL_1516687, EPI_ISL_1516688, EPI_ISL_1516690, EPI_ISL_1516691, EPI_ISL_1516692, EPI_ISL_1516693, EPI_ISL_1516694, EPI_ISL_1516695, EPI_ISL_1516697, EPI_ISL_1516699, EPI_ISL_1516700, EPI_ISL_1516701, EPI_ISL_1516702, EPI_ISL_1516703, EPI_ISL_1516704, EPI_ISL_1516705, EPI_ISL_1516706, EPI_ISL_1516707, EPI_ISL_1516708, EPI_ISL_1516709, EPI_ISL_1516711, EPI_ISL_1516712, EPI_ISL_1516713, EPI_ISL_1516714, EPI_ISL_1516715, EPI_ISL_1516716, EPI_ISL_1516717, EPI_ISL_1516718, EPI_ISL_1516719, EPI_ISL_1516720, EPI_ISL_1516721, EPI_ISL_1516722, EPI_ISL_1516723, EPI_ISL_1516724, EPI_ISL_1516725, EPI_ISL_1516726, EPI_ISL_1516727, EPI_ISL_1516728, EPI_ISL_1516729, EPI_ISL_1516730, EPI_ISL_1516731, EPI_ISL_1516732, EPI_ISL_1516733, EPI_ISL_1516734, EPI_ISL_1516735, EPI_ISL_1516736, EPI_ISL_1516737, EPI_ISL_1516738, EPI_ISL_1516739, EPI_ISL_1516740, EPI_ISL_1516741, EPI_ISL_1516742, EPI_ISL_1516743, EPI_ISL_1516745, EPI_ISL_1578140, EPI_ISL_1578141, EPI_ISL_1578142, EPI_ISL_1578143, EPI_ISL_1578149, EPI_ISL_1578161, EPI_ISL_1578164, EPI_ISL_1578166, EPI_ISL_1578167, EPI_ISL_1578168, EPI_ISL_1578169, EPI_ISL_1578172, EPI_ISL_1578173, EPI_ISL_1578174, EPI_ISL_1578178, EPI_ISL_1578179, EPI_ISL_1578184, EPI_ISL_1578187, EPI_ISL_1578189, EPI_ISL_1578190, EPI_ISL_1578193, EPI_ISL_1578198, EPI_ISL_1578199, EPI_ISL_1578200, EPI_ISL_1578204, EPI_ISL_1578205, EPI_ISL_1578206, EPI_ISL_1578207, EPI_ISL_1578209, EPI_ISL_1578212, EPI_ISL_1578216, EPI_ISL_1578218, EPI_ISL_1578220, EPI_ISL_1578222, EPI_ISL_1578224, EPI_ISL_1578229, EPI_ISL_1578230, EPI_ISL_1578232, EPI_ISL_1578233, EPI_ISL_1578234, EPI_ISL_1578236, EPI_ISL_1578237, EPI_ISL_1578238, EPI_ISL_1578239, EPI_ISL_1578240, EPI_ISL_1578242, EPI_ISL_1578243, EPI_ISL_1578244, EPI_ISL_1578245, EPI_ISL_1578246, EPI_ISL_1578247, EPI_ISL_1578248, EPI_ISL_1578249, EPI_ISL_1578250, EPI_ISL_1578251, EPI_ISL_1578252, EPI_ISL_1578253, EPI_ISL_1578254, EPI_ISL_1578255, EPI_ISL_1578256, EPI_ISL_1578257, EPI_ISL_1578258, EPI_ISL_1578259, EPI_ISL_1578260, EPI_ISL_1578261, EPI_ISL_1578262, EPI_ISL_1578263, EPI_ISL_1578264, EPI_ISL_1578265, EPI_ISL_1578266, EPI_ISL_1578267, EPI_ISL_1578300, EPI_ISL_1578301, EPI_ISL_1578302, EPI_ISL_1578304, EPI_ISL_1578305, EPI_ISL_1578306, EPI_ISL_1578307, EPI_ISL_1578308, EPI_ISL_1578309, EPI_ISL_1578310, EPI_ISL_1578311, EPI_ISL_1578312, EPI_ISL_1578313, EPI_ISL_1578314, EPI_ISL_1578340, EPI_ISL_1578342, EPI_ISL_1578343, EPI_ISL_1578344, EPI_ISL_1578345, EPI_ISL_1578346, EPI_ISL_1578347, EPI_ISL_1578350, EPI_ISL_1578351, EPI_ISL_1578352, EPI_ISL_1578378, EPI_ISL_1578379, EPI_ISL_1578391, EPI_ISL_1578392, EPI_ISL_1578394, EPI_ISL_1578395, EPI_ISL_1578411, EPI_ISL_1578412, EPI_ISL_1578413, EPI_ISL_1578415, EPI_ISL_1578416, EPI_ISL_1578417, EPI_ISL_1578418, EPI_ISL_1578419, EPI_ISL_1578420, EPI_ISL_1578421, EPI_ISL_1578422, EPI_ISL_1578423, EPI_ISL_1578424, EPI_ISL_1578425, EPI_ISL_1578426, EPI_ISL_1578427, EPI_ISL_1578428, EPI_ISL_1578429, EPI_ISL_1578430, EPI_ISL_1578431, EPI_ISL_1710150, EPI_ISL_1710151, EPI_ISL_1710152, EPI_ISL_1710153, EPI_ISL_1710154, EPI_ISL_1710156, EPI_ISL_1710158, EPI_ISL_1710159, EPI_ISL_1710160, EPI_ISL_1710161, EPI_ISL_1710162, EPI_ISL_1710163, EPI_ISL_1710164, EPI_ISL_1710165, EPI_ISL_1710166, EPI_ISL_1710168, EPI_ISL_1710170, EPI_ISL_1710171, EPI_ISL_1710172, EPI_ISL_1710173, EPI_ISL_1710174, EPI_ISL_1710175, EPI_ISL_1710179, EPI_ISL_1710180, EPI_ISL_1710245, EPI_ISL_1710246, EPI_ISL_1710247, EPI_ISL_1710248, EPI_ISL_1710249, EPI_ISL_1710250, EPI_ISL_1710251, EPI_ISL_1710253, EPI_ISL_1710254, EPI_ISL_1710255, EPI_ISL_1710256, EPI_ISL_1710257, EPI_ISL_1710258, EPI_ISL_1710260, EPI_ISL_1710261, EPI_ISL_1710262, EPI_ISL_1710264, EPI_ISL_1710265, EPI_ISL_1710266, EPI_ISL_1710267, EPI_ISL_1710268, EPI_ISL_1710269, EPI_ISL_1710270, EPI_ISL_1710271, EPI_ISL_1710272, EPI_ISL_1710273, EPI_ISL_1710274, EPI_ISL_1710275, EPI_ISL_1710276, EPI_ISL_1710277, EPI_ISL_1710278, EPI_ISL_1710279, EPI_ISL_1710281, EPI_ISL_1710282, EPI_ISL_1710283, EPI_ISL_1710284, EPI_ISL_1710285, EPI_ISL_1710286, EPI_ISL_1710287, EPI_ISL_1710288, EPI_ISL_1710289, EPI_ISL_1710290, EPI_ISL_1710292, EPI_ISL_1710293, EPI_ISL_1710294, EPI_ISL_1710295, EPI_ISL_1710296, EPI_ISL_1710297, EPI_ISL_1710301, EPI_ISL_1710302, EPI_ISL_1710303, EPI_ISL_1710304, EPI_ISL_1710306, EPI_ISL_1710307, EPI_ISL_1710308, EPI_ISL_1710309, EPI_ISL_1710310, EPI_ISL_1710311, EPI_ISL_1710313, EPI_ISL_1710314, EPI_ISL_1710315, EPI_ISL_1710316, EPI_ISL_1710317, EPI_ISL_1710318, EPI_ISL_1710319, EPI_ISL_1710321, EPI_ISL_1710324, EPI_ISL_1710325, EPI_ISL_1710326, EPI_ISL_1710327, EPI_ISL_1710328, EPI_ISL_1710329, EPI_ISL_1710330, EPI_ISL_1710331, EPI_ISL_1710332, EPI_ISL_1710333, EPI_ISL_1710334, EPI_ISL_1710335, EPI_ISL_1710336, EPI_ISL_1710337, EPI_ISL_1710338, EPI_ISL_1710339, EPI_ISL_1710340, EPI_ISL_1710341, EPI_ISL_1710342, EPI_ISL_1710343, EPI_ISL_1710344, EPI_ISL_1710345, EPI_ISL_1710346, EPI_ISL_1710348, EPI_ISL_1710349, EPI_ISL_1710350, EPI_ISL_1710351, EPI_ISL_1710352, EPI_ISL_1710354, EPI_ISL_1710355, EPI_ISL_1710356, EPI_ISL_1710357, EPI_ISL_1710358, EPI_ISL_1710359, EPI_ISL_1710360, EPI_ISL_1710362, EPI_ISL_1710363, EPI_ISL_1710364, EPI_ISL_1710365, EPI_ISL_1710368, EPI_ISL_1710369, EPI_ISL_1710370, EPI_ISL_1710373, EPI_ISL_1710374, EPI_ISL_1710375, EPI_ISL_1710376, EPI_ISL_1710377, EPI_ISL_1710378, EPI_ISL_1710379, EPI_ISL_1710380, EPI_ISL_1710381, EPI_ISL_1710382, EPI_ISL_1710384, EPI_ISL_1710385, EPI_ISL_1710386, EPI_ISL_1710387, EPI_ISL_1710388, EPI_ISL_1710389, EPI_ISL_1710390, EPI_ISL_1710391, EPI_ISL_1710392, EPI_ISL_1710393, EPI_ISL_1710394, EPI_ISL_1710395, EPI_ISL_1710396, EPI_ISL_1710397, EPI_ISL_1710398, EPI_ISL_1710400, EPI_ISL_1710401, EPI_ISL_1710402, EPI_ISL_1710403, EPI_ISL_1710404, EPI_ISL_1710405, EPI_ISL_1710406, EPI_ISL_1710407, EPI_ISL_1710410, EPI_ISL_1710411, EPI_ISL_1710412, EPI_ISL_1710413, EPI_ISL_1710414, EPI_ISL_1710415, EPI_ISL_1710416, EPI_ISL_1710417, EPI_ISL_1710418, EPI_ISL_1710419, EPI_ISL_1710420, EPI_ISL_1710421, EPI_ISL_1710422, EPI_ISL_1710423, EPI_ISL_1710424, EPI_ISL_1710425, EPI_ISL_1710426, EPI_ISL_1710427, EPI_ISL_1710428, EPI_ISL_1710429, EPI_ISL_1710430, EPI_ISL_1710431, EPI_ISL_1710432, EPI_ISL_1710433, EPI_ISL_1710434, EPI_ISL_1710436, EPI_ISL_1710437, EPI_ISL_1710438, EPI_ISL_1710439, EPI_ISL_1710440, EPI_ISL_1710442, EPI_ISL_1710444, EPI_ISL_1710446, EPI_ISL_1710447, EPI_ISL_1710448, EPI_ISL_1710449, EPI_ISL_1710450, EPI_ISL_1710451, EPI_ISL_1710452, EPI_ISL_1710453, EPI_ISL_1710454, EPI_ISL_1710455, EPI_ISL_1710456, EPI_ISL_1710457, EPI_ISL_1710458, EPI_ISL_1710459, EPI_ISL_1710460, EPI_ISL_1710461, EPI_ISL_1710462, EPI_ISL_1710463, EPI_ISL_1710464, EPI_ISL_1710465, EPI_ISL_1710466, EPI_ISL_1710467, EPI_ISL_1710468, EPI_ISL_1710469, EPI_ISL_1710470, EPI_ISL_1710471, EPI_ISL_1710472, EPI_ISL_1710474, EPI_ISL_1710475, EPI_ISL_1710476, EPI_ISL_1710478, EPI_ISL_1710479, EPI_ISL_1710480, EPI_ISL_1710481, EPI_ISL_1710482, EPI_ISL_1710483, EPI_ISL_1710484, EPI_ISL_1710485, EPI_ISL_1710486, EPI_ISL_1710487, EPI_ISL_1710488, EPI_ISL_1710489, EPI_ISL_1710490, EPI_ISL_1710491, EPI_ISL_1710492, EPI_ISL_1710493, EPI_ISL_1710494, EPI_ISL_1710495, EPI_ISL_1710496, EPI_ISL_1710497, EPI_ISL_1710498, EPI_ISL_1710499, EPI_ISL_1710500, EPI_ISL_1710501, EPI_ISL_1710502, EPI_ISL_1710503, EPI_ISL_1710504, EPI_ISL_1710505, EPI_ISL_1743719, EPI_ISL_1743721, EPI_ISL_1743722, EPI_ISL_1743723, EPI_ISL_1743726, EPI_ISL_1743727, EPI_ISL_1743728, EPI_ISL_1743730, EPI_ISL_1743732, EPI_ISL_1743733, EPI_ISL_1743734, EPI_ISL_1743736, EPI_ISL_1743737, EPI_ISL_1743738, EPI_ISL_1743739, EPI_ISL_1743740, EPI_ISL_1743741, EPI_ISL_1743742, EPI_ISL_1743744, EPI_ISL_1743746, EPI_ISL_1743747, EPI_ISL_1743749, EPI_ISL_1743751, EPI_ISL_1743752, EPI_ISL_1743753, EPI_ISL_1743754, EPI_ISL_1743755, EPI_ISL_1743757, EPI_ISL_1743758, EPI_ISL_1743759, EPI_ISL_1743761, EPI_ISL_1743762, EPI_ISL_1743763, EPI_ISL_1743764, EPI_ISL_1743766, EPI_ISL_1743767, EPI_ISL_1743768, EPI_ISL_1743769, EPI_ISL_1743770, EPI_ISL_1743771, EPI_ISL_1743772, EPI_ISL_1743773, EPI_ISL_1743774, EPI_ISL_1743775, EPI_ISL_1743776, EPI_ISL_1743777, EPI_ISL_1743778, EPI_ISL_1743779, EPI_ISL_1743780, EPI_ISL_1743781, EPI_ISL_1743782, EPI_ISL_1743783, EPI_ISL_1743784, EPI_ISL_1743786, EPI_ISL_1743787, EPI_ISL_1743788, EPI_ISL_1743789, EPI_ISL_1743790, | | | |

EPI_ISL_1743791, EPI_ISL_1743792, EPI_ISL_1743793, EPI_ISL_1743794, EPI_ISL_1743795, EPI_ISL_1743796, EPI_ISL_1743797, EPI_ISL_1743799, EPI_ISL_1743800, EPI_ISL_1743801, EPI_ISL_1743802, EPI_ISL_1743803, EPI_ISL_1743804, EPI_ISL_1743805, EPI_ISL_1743806, EPI_ISL_1743807, EPI_ISL_1743808, EPI_ISL_1743809, EPI_ISL_1743810, EPI_ISL_1743811, EPI_ISL_1743812, EPI_ISL_1743813,
EPI_ISL_1743814, EPI_ISL_1743815, EPI_ISL_1743816, EPI_ISL_1743817, EPI_ISL_1743818, EPI_ISL_1743819, EPI_ISL_1743820, EPI_ISL_1743821, EPI_ISL_1743822, EPI_ISL_1743823, EPI_ISL_1743824, EPI_ISL_1743826, EPI_ISL_1743827, EPI_ISL_1743830, EPI_ISL_1743831, EPI_ISL_1743832, EPI_ISL_1743835, EPI_ISL_1743836, EPI_ISL_1743837, EPI_ISL_1743838, EPI_ISL_1743839, EPI_ISL_1743840,
EPI_ISL_1743841, EPI_ISL_1743842, EPI_ISL_1743843, EPI_ISL_1743844, EPI_ISL_1743845, EPI_ISL_1743846, EPI_ISL_1743847, EPI_ISL_1743848, EPI_ISL_1743849, EPI_ISL_1743850, EPI_ISL_1743852, EPI_ISL_1743853, EPI_ISL_1743855, EPI_ISL_1743856, EPI_ISL_1743857, EPI_ISL_1743858, EPI_ISL_1743859, EPI_ISL_1743860, EPI_ISL_1743861, EPI_ISL_1743862, EPI_ISL_1743863, EPI_ISL_1743864,
EPI_ISL_1743865, EPI_ISL_1743867, EPI_ISL_1743868, EPI_ISL_1743869, EPI_ISL_1743870, EPI_ISL_1743871, EPI_ISL_1743872, EPI_ISL_1743873, EPI_ISL_1743875, EPI_ISL_1743876, EPI_ISL_1743877, EPI_ISL_1743878, EPI_ISL_1743879, EPI_ISL_1743880, EPI_ISL_1743881, EPI_ISL_1743883, EPI_ISL_1743884, EPI_ISL_1743885, EPI_ISL_1743887, EPI_ISL_1743888, EPI_ISL_1743889, EPI_ISL_1743890,
EPI_ISL_1743892, EPI_ISL_1743894, EPI_ISL_1743895, EPI_ISL_1743896, EPI_ISL_1743897, EPI_ISL_1743898, EPI_ISL_1743899, EPI_ISL_1743900, EPI_ISL_1743901, EPI_ISL_1743902, EPI_ISL_1743903, EPI_ISL_1743904, EPI_ISL_1743905, EPI_ISL_1743906, EPI_ISL_1743907, EPI_ISL_1743908, EPI_ISL_1743909, EPI_ISL_1743910, EPI_ISL_1743911, EPI_ISL_1743912, EPI_ISL_1743913, EPI_ISL_1743914,
EPI_ISL_1743915, EPI_ISL_1743916, EPI_ISL_1743917, EPI_ISL_1743918, EPI_ISL_1743919, EPI_ISL_1743920, EPI_ISL_1743922, EPI_ISL_1743923, EPI_ISL_1743924, EPI_ISL_1743925, EPI_ISL_1743926, EPI_ISL_1743927, EPI_ISL_1743928, EPI_ISL_1743929, EPI_ISL_1743930, EPI_ISL_1743931, EPI_ISL_1743932, EPI_ISL_1743933, EPI_ISL_1743934, EPI_ISL_1743935, EPI_ISL_1743936, EPI_ISL_1743939,
EPI_ISL_1743941, EPI_ISL_1743942, EPI_ISL_1743943, EPI_ISL_1743944, EPI_ISL_1743945, EPI_ISL_1743946, EPI_ISL_1743947, EPI_ISL_1743948, EPI_ISL_1743949, EPI_ISL_1743950, EPI_ISL_1743951, EPI_ISL_1743952, EPI_ISL_1743953, EPI_ISL_1743954, EPI_ISL_1743956, EPI_ISL_1743958, EPI_ISL_1743959, EPI_ISL_1743960, EPI_ISL_1743961, EPI_ISL_1743962, EPI_ISL_1743963, EPI_ISL_1743964,
EPI_ISL_1743965, EPI_ISL_1743966, EPI_ISL_1743967, EPI_ISL_1743968, EPI_ISL_1743969, EPI_ISL_1743970, EPI_ISL_1743971, EPI_ISL_1743972, EPI_ISL_1743973, EPI_ISL_1743974, EPI_ISL_1743975, EPI_ISL_1743976, EPI_ISL_1743977, EPI_ISL_1743979, EPI_ISL_1743980, EPI_ISL_1743981, EPI_ISL_1743982, EPI_ISL_1743983, EPI_ISL_1743984, EPI_ISL_1743985, EPI_ISL_1743987, EPI_ISL_1743988,
EPI_ISL_1743989, EPI_ISL_1743990, EPI_ISL_1743991, EPI_ISL_1743993, EPI_ISL_1743994, EPI_ISL_1743995, EPI_ISL_1743996, EPI_ISL_1743997, EPI_ISL_1744000, EPI_ISL_1744001, EPI_ISL_1744002, EPI_ISL_1744003, EPI_ISL_1744004, EPI_ISL_1744005, EPI_ISL_1744006, EPI_ISL_1744007, EPI_ISL_1744008, EPI_ISL_1744009, EPI_ISL_1744010, EPI_ISL_1744012, EPI_ISL_1744013, EPI_ISL_1744014,
EPI_ISL_1744015, EPI_ISL_1744016, EPI_ISL_1744017, EPI_ISL_1744018, EPI_ISL_1744019, EPI_ISL_1744020, EPI_ISL_1744021, EPI_ISL_1744022, EPI_ISL_1744024, EPI_ISL_1744025, EPI_ISL_1744027, EPI_ISL_1744028, EPI_ISL_1744029, EPI_ISL_1744030, EPI_ISL_1744031, EPI_ISL_1744033, EPI_ISL_1744034, EPI_ISL_1744035, EPI_ISL_1744036, EPI_ISL_1744037, EPI_ISL_1744038, EPI_ISL_1744039,
EPI_ISL_1744040, EPI_ISL_1744041, EPI_ISL_1744043, EPI_ISL_1744044, EPI_ISL_1744045, EPI_ISL_1744047, EPI_ISL_1744048, EPI_ISL_1744049, EPI_ISL_1744050, EPI_ISL_1744051, EPI_ISL_1744052, EPI_ISL_1744054, EPI_ISL_1744055, EPI_ISL_1744056, EPI_ISL_1744057, EPI_ISL_1744058, EPI_ISL_1744059, EPI_ISL_1744060, EPI_ISL_1744061, EPI_ISL_1744062, EPI_ISL_1744063, EPI_ISL_1744064,
EPI_ISL_1744065, EPI_ISL_1744066, EPI_ISL_1744068, EPI_ISL_1744069, EPI_ISL_1744070, EPI_ISL_1744071, EPI_ISL_1744072, EPI_ISL_1744073, EPI_ISL_1744074, EPI_ISL_1744075, EPI_ISL_1744076, EPI_ISL_1744078, EPI_ISL_1744079, EPI_ISL_1744080, EPI_ISL_1744081, EPI_ISL_1744082, EPI_ISL_1744084, EPI_ISL_1744085, EPI_ISL_1744086, EPI_ISL_1744088, EPI_ISL_1744089, EPI_ISL_1744090,
EPI_ISL_1744091, EPI_ISL_1744092, EPI_ISL_1744093, EPI_ISL_1744094, EPI_ISL_1744095, EPI_ISL_1744096, EPI_ISL_1744097, EPI_ISL_1744098, EPI_ISL_1744099, EPI_ISL_1744100, EPI_ISL_1744101, EPI_ISL_1744102, EPI_ISL_1744103, EPI_ISL_1744104, EPI_ISL_1744105, EPI_ISL_1744106, EPI_ISL_1744107, EPI_ISL_1744108, EPI_ISL_1744110, EPI_ISL_1744111, EPI_ISL_1744112, EPI_ISL_1744113,
EPI_ISL_1744115, EPI_ISL_1744116, EPI_ISL_1744117, EPI_ISL_1744118, EPI_ISL_1744119, EPI_ISL_1744120, EPI_ISL_1744121, EPI_ISL_1744122, EPI_ISL_1744123, EPI_ISL_1744124, EPI_ISL_1744125, EPI_ISL_1744127, EPI_ISL_1744128, EPI_ISL_1744129, EPI_ISL_1744130, EPI_ISL_1744131, EPI_ISL_1744132, EPI_ISL_1744133, EPI_ISL_1744134, EPI_ISL_1744135, EPI_ISL_1744137, EPI_ISL_1744138,
EPI_ISL_1744139, EPI_ISL_1744140, EPI_ISL_1744141, EPI_ISL_1744142, EPI_ISL_1744143, EPI_ISL_1744144, EPI_ISL_1744145, EPI_ISL_1744146, EPI_ISL_1744147, EPI_ISL_1744148, EPI_ISL_1744149, EPI_ISL_1744150, EPI_ISL_1744151, EPI_ISL_1744153, EPI_ISL_1744154, EPI_ISL_1744155, EPI_ISL_1744156, EPI_ISL_1744157, EPI_ISL_1744158, EPI_ISL_1744159, EPI_ISL_1744160, EPI_ISL_1744161,
EPI_ISL_1744162, EPI_ISL_1744163, EPI_ISL_1744164, EPI_ISL_1744165, EPI_ISL_1744166, EPI_ISL_1744168, EPI_ISL_1744169, EPI_ISL_1744170, EPI_ISL_1744172, EPI_ISL_1744173, EPI_ISL_1744174, EPI_ISL_1744175, EPI_ISL_1744177, EPI_ISL_1744178, EPI_ISL_1744179, EPI_ISL_1744180, EPI_ISL_1744181, EPI_ISL_1744182, EPI_ISL_1744183, EPI_ISL_1744184, EPI_ISL_1744186, EPI_ISL_1744187,
EPI_ISL_1744188, EPI_ISL_1744189, EPI_ISL_1744190, EPI_ISL_1744192, EPI_ISL_1744193, EPI_ISL_1744194, EPI_ISL_1744195, EPI_ISL_1744196, EPI_ISL_1744197, EPI_ISL_1744198, EPI_ISL_1744199, EPI_ISL_1744200, EPI_ISL_1744202, EPI_ISL_1744203, EPI_ISL_1744204, EPI_ISL_1744205, EPI_ISL_1744206, EPI_ISL_1744207, EPI_ISL_1744208, EPI_ISL_1744209, EPI_ISL_1744210, EPI_ISL_1744212,
EPI_ISL_1744213, EPI_ISL_1744214, EPI_ISL_1744215, EPI_ISL_1744216, EPI_ISL_1744217, EPI_ISL_1744218, EPI_ISL_1744219, EPI_ISL_1744220, EPI_ISL_1744221, EPI_ISL_1744222, EPI_ISL_1744223, EPI_ISL_1744224, EPI_ISL_1744225, EPI_ISL_1744226, EPI_ISL_1744227, EPI_ISL_1744228, EPI_ISL_1744229, EPI_ISL_1744230, EPI_ISL_1744232, EPI_ISL_1744234, EPI_ISL_1744235, EPI_ISL_1744236,
EPI_ISL_1744237, EPI_ISL_1744238, EPI_ISL_1744239, EPI_ISL_1744240, EPI_ISL_1744241, EPI_ISL_1744242, EPI_ISL_1744243, EPI_ISL_1744244, EPI_ISL_1744245, EPI_ISL_1744246, EPI_ISL_1744247, EPI_ISL_1744248, EPI_ISL_1744249, EPI_ISL_1744250, EPI_ISL_1744251, EPI_ISL_1744252, EPI_ISL_1744253, EPI_ISL_1744254, EPI_ISL_1744255, EPI_ISL_1744256, EPI_ISL_1744257, EPI_ISL_1744258,
EPI_ISL_1744259, EPI_ISL_1744260, EPI_ISL_1744261, EPI_ISL_1744262, EPI_ISL_1744263, EPI_ISL_1744264, EPI_ISL_1744265, EPI_ISL_1744266, EPI_ISL_1744267, EPI_ISL_1744268, EPI_ISL_1744269, EPI_ISL_1744270, EPI_ISL_1744271, EPI_ISL_1744272, EPI_ISL_1744273, EPI_ISL_1744276, EPI_ISL_1744277, EPI_ISL_1744278, EPI_ISL_1744279, EPI_ISL_1744280, EPI_ISL_1744281, EPI_ISL_1744282,
EPI_ISL_1744283, EPI_ISL_1744284, EPI_ISL_1744285, EPI_ISL_1744286, EPI_ISL_1744287, EPI_ISL_1744288, EPI_ISL_1744289, EPI_ISL_1744290, EPI_ISL_1744292, EPI_ISL_1744293, EPI_ISL_1744294, EPI_ISL_1744295, EPI_ISL_1744296, EPI_ISL_1744297, EPI_ISL_1744298, EPI_ISL_1744299, EPI_ISL_1744301, EPI_ISL_1744302, EPI_ISL_1744303, EPI_ISL_1756987, EPI_ISL_1756988, EPI_ISL_1756989,
EPI_ISL_1756990, EPI_ISL_1756991, EPI_ISL_1756993, EPI_ISL_1756994, EPI_ISL_1756996, EPI_ISL_1756997, EPI_ISL_1756998, EPI_ISL_1756999, EPI_ISL_1757000, EPI_ISL_1757001, EPI_ISL_1757002, EPI_ISL_1757005, EPI_ISL_1757006, EPI_ISL_1757007, EPI_ISL_1757009, EPI_ISL_1757010, EPI_ISL_1757011, EPI_ISL_1757012, EPI_ISL_1757013, EPI_ISL_1757014, EPI_ISL_1757015, EPI_ISL_1757016,
EPI_ISL_1757017, EPI_ISL_1757018, EPI_ISL_1757019, EPI_ISL_1757021, EPI_ISL_1757022, EPI_ISL_1757024, EPI_ISL_1757025, EPI_ISL_1757026, EPI_ISL_1757027, EPI_ISL_1757028, EPI_ISL_1757029, EPI_ISL_1757030, EPI_ISL_1757031, EPI_ISL_1757032, EPI_ISL_1757033, EPI_ISL_1757035, EPI_ISL_1757037, EPI_ISL_1757038, EPI_ISL_1757039, EPI_ISL_1757040, EPI_ISL_1757043, EPI_ISL_1757045,
EPI_ISL_1757046, EPI_ISL_1757078, EPI_ISL_1757079, EPI_ISL_1757080, EPI_ISL_1757093, EPI_ISL_1757094, EPI_ISL_1757095, EPI_ISL_1757097, EPI_ISL_1757098, EPI_ISL_1757099, EPI_ISL_1757100, EPI_ISL_1757101, EPI_ISL_1757102, EPI_ISL_1757103, EPI_ISL_1757104, EPI_ISL_1757105, EPI_ISL_1757106, EPI_ISL_1757107, EPI_ISL_1757108, EPI_ISL_1757109, EPI_ISL_1757111, EPI_ISL_1757112,
EPI_ISL_1757113, EPI_ISL_1757114, EPI_ISL_1757115, EPI_ISL_1757117, EPI_ISL_1757118, EPI_ISL_1757119, EPI_ISL_1757120, EPI_ISL_1757121, EPI_ISL_1757181, EPI_ISL_1757184, EPI_ISL_1757185, EPI_ISL_1757186, EPI_ISL_1757187, EPI_ISL_1757188, EPI_ISL_1757189, EPI_ISL_1757190, EPI_ISL_1757191, EPI_ISL_1757192, EPI_ISL_1757194, EPI_ISL_1757195, EPI_ISL_1757196, EPI_ISL_1757197,
EPI_ISL_1757198, EPI_ISL_1757199, EPI_ISL_1757200, EPI_ISL_1757201, EPI_ISL_1757202, EPI_ISL_1757203, EPI_ISL_1757204, EPI_ISL_1757205, EPI_ISL_1757206, EPI_ISL_1757207, EPI_ISL_1757210, EPI_ISL_1757211, EPI_ISL_1757212, EPI_ISL_1757213, EPI_ISL_1757214, EPI_ISL_1757215, EPI_ISL_1757216, EPI_ISL_1757217, EPI_ISL_1757218, EPI_ISL_1757219, EPI_ISL_1757220, EPI_ISL_1757222,
EPI_ISL_1757223, EPI_ISL_1757224, EPI_ISL_1757225, EPI_ISL_1757226, EPI_ISL_1757227, EPI_ISL_1757229, EPI_ISL_1757230, EPI_ISL_1757231, EPI_ISL_1757232, EPI_ISL_1757233, EPI_ISL_1757234, EPI_ISL_1757235, EPI_ISL_1757236, EPI_ISL_1757239, EPI_ISL_1757240, EPI_ISL_1757241, EPI_ISL_1757242, EPI_ISL_1757243, EPI_ISL_1757244, EPI_ISL_1757245, EPI_ISL_1757246, EPI_ISL_1757247,
EPI_ISL_1757248, EPI_ISL_1757249, EPI_ISL_1757250, EPI_ISL_1757251, EPI_ISL_1757252, EPI_ISL_1757294, EPI_ISL_1757298, EPI_ISL_1757299, EPI_ISL_1757300, EPI_ISL_1757301, EPI_ISL_1757302, EPI_ISL_1757303, EPI_ISL_1757304, EPI_ISL_1757305, EPI_ISL_1757307, EPI_ISL_1757308, EPI_ISL_1757309, EPI_ISL_1757310, EPI_ISL_1757311, EPI_ISL_1757312, EPI_ISL_1757313, EPI_ISL_1757314,
EPI_ISL_1757315, EPI_ISL_1757317, EPI_ISL_1757319, EPI_ISL_1757321, EPI_ISL_1757322, EPI_ISL_1757323, EPI_ISL_1757324, EPI_ISL_1757325, EPI_ISL_1786733, EPI_ISL_1786734, EPI_ISL_1786735, EPI_ISL_1786736, EPI_ISL_1786737, EPI_ISL_1786738, EPI_ISL_1786739, EPI_ISL_1786740, EPI_ISL_1786741, EPI_ISL_1786743, EPI_ISL_1786745, EPI_ISL_1786746, EPI_ISL_1786747, EPI_ISL_1786748,
EPI_ISL_1786749, EPI_ISL_1786750, EPI_ISL_1786751, EPI_ISL_1786752, EPI_ISL_1786753, EPI_ISL_1786754, EPI_ISL_1786756, EPI_ISL_1786757, EPI_ISL_1786758, EPI_ISL_1786759, EPI_ISL_1786760, EPI_ISL_1786761, EPI_ISL_1786762, EPI_ISL_1786763, EPI_ISL_1786764, EPI_ISL_1786765, EPI_ISL_1786766, EPI_ISL_1786767, EPI_ISL_1786768, EPI_ISL_1786769, EPI_ISL_1786770, EPI_ISL_1786771,
EPI_ISL_1786772, EPI_ISL_1786773, EPI_ISL_1786774, EPI_ISL_1786776, EPI_ISL_1786777, EPI_ISL_1786778, EPI_ISL_1786779, EPI_ISL_1786780, EPI_ISL_1786781, EPI_ISL_1786782, EPI_ISL_1786783, EPI_ISL_1786784, EPI_ISL_1786785, EPI_ISL_1786786, EPI_ISL_1786787, EPI_ISL_1786788, EPI_ISL_1786790, EPI_ISL_1786791, EPI_ISL_1786792, EPI_ISL_1786793, EPI_ISL_1786795, EPI_ISL_1786796,
EPI_ISL_1786798, EPI_ISL_1786799, EPI_ISL_1786800, EPI_ISL_1786801, EPI_ISL_1786802, EPI_ISL_1786803, EPI_ISL_1786804, EPI_ISL_1786805, EPI_ISL_1786806, EPI_ISL_1786807, EPI_ISL_1786808, EPI_ISL_1786809, EPI_ISL_1786810, EPI_ISL_1786811, EPI_ISL_1786813, EPI_ISL_1786814, EPI_ISL_1786815, EPI_ISL_1786816, EPI_ISL_1786818, EPI_ISL_1786819, EPI_ISL_1786820, EPI_ISL_1786821,
EPI_ISL_1786822, EPI_ISL_1786823, EPI_ISL_1786824, EPI_ISL_1786826, EPI_ISL_1786827, EPI_ISL_1786828, EPI_ISL_1786829, EPI_ISL_1786830, EPI_ISL_1786831, EPI_ISL_1786832, EPI_ISL_1786833, EPI_ISL_1786834, EPI_ISL_1786836, EPI_ISL_1786837, EPI_ISL_1786839, EPI_ISL_1786840, EPI_ISL_1786841, EPI_ISL_1786842, EPI_ISL_1786843, EPI_ISL_1786844, EPI_ISL_1786845, EPI_ISL_1786847,
EPI_ISL_1786848, EPI_ISL_1786849, EPI_ISL_1786850, EPI_ISL_1786851, EPI_ISL_1786852, EPI_ISL_1786853, EPI_ISL_1786854, EPI_ISL_1786855, EPI_ISL_1786856, EPI_ISL_1786857, EPI_ISL_1786858, EPI_ISL_1786859, EPI_ISL_1786860, EPI_ISL_1786861, EPI_ISL_1786862, EPI_ISL_1786863, EPI_ISL_1786864, EPI_ISL_1786865, EPI_ISL_1786866, EPI_ISL_1786869, EPI_ISL_1786870, EPI_ISL_1786871,
EPI_ISL_1786873, EPI_ISL_1786874, EPI_ISL_1786876, EPI_ISL_1786877, EPI_ISL_1786878, EPI_ISL_1786879, EPI_ISL_1786880, EPI_ISL_1786881, EPI_ISL_1786882, EPI_ISL_1786884, EPI_ISL_1786885, EPI_ISL_1786886, EPI_ISL_1786887, EPI_ISL_1786888, EPI_ISL_1786889, EPI_ISL_1786890, EPI_ISL_1786891, EPI_ISL_1786892, EPI_ISL_1786894, EPI_ISL_1786895, EPI_ISL_1786896, EPI_ISL_1786897,
EPI_ISL_1786898, EPI_ISL_1786899, EPI_ISL_1786900, EPI_ISL_1786901, EPI_ISL_1786902, EPI_ISL_1786903, EPI_ISL_1786904, EPI_ISL_1786905, EPI_ISL_1786909, EPI_ISL_1786910, EPI_ISL_1786911, EPI_ISL_1786912, EPI_ISL_1786913, EPI_ISL_1786914, EPI_ISL_1786915, EPI_ISL_1786916, EPI_ISL_1786917, EPI_ISL_1786918, EPI_ISL_1786920, EPI_ISL_1786923, EPI_ISL_1786924, EPI_ISL_1786925,
EPI_ISL_1786926, EPI_ISL_1786927, EPI_ISL_1786928, EPI_ISL_1786929, EPI_ISL_1786930, EPI_ISL_1786931, EPI_ISL_1786932, EPI_ISL_1786933, EPI_ISL_1786934, EPI_ISL_1786935, EPI_ISL_1786936, EPI_ISL_1786937, EPI_ISL_1786938, EPI_ISL_1786939, EPI_ISL_1786940, EPI_ISL_1786941, EPI_ISL_1786942, EPI_ISL_1786943, EPI_ISL_1786945, EPI_ISL_1786946, EPI_ISL_1786947, EPI_ISL_1786948,
EPI_ISL_1786949, EPI_ISL_1786950, EPI_ISL_1786951, EPI_ISL_1786952, EPI_ISL_1786953, EPI_ISL_1786954, EPI_ISL_1786955, EPI_ISL_1786956, EPI_ISL_1786957, EPI_ISL_1786958, EPI_ISL_1786960, EPI_ISL_1786961, EPI_ISL_1786962, EPI_ISL_1786963, EPI_ISL_1786964, EPI_ISL_1786966, EPI_ISL_1786967, EPI_ISL_1786968, EPI_ISL_1786970, EPI_ISL_1786971, EPI_ISL_1786972, EPI_ISL_1786974,
EPI_ISL_1786975, EPI_ISL_1786976, EPI_ISL_1786977, EPI_ISL_1786978, EPI_ISL_1786979, EPI_ISL_1786980, EPI_ISL_1786981, EPI_ISL_1786982, EPI_ISL_1786983, EPI_ISL_1786984, EPI_ISL_1786985, EPI_ISL_1786986, EPI_ISL_1786987, EPI_ISL_1786988, EPI_ISL_1786989, EPI_ISL_1786990, EPI_ISL_1786991, EPI_ISL_1786992, EPI_ISL_1786993, EPI_ISL_1786995, EPI_ISL_1786996, EPI_ISL_1786997,
EPI_ISL_1786998, EPI_ISL_1786999, EPI_ISL_1787001, EPI_ISL_1787002, EPI_ISL_1787003, EPI_ISL_1787004, EPI_ISL_1787005, EPI_ISL_1787006, EPI_ISL_1787007, EPI_ISL_1787008, EPI_ISL_1787009, EPI_ISL_1787010, EPI_ISL_1787012, EPI_ISL_1787013, EPI_ISL_1787014, EPI_ISL_1787015, EPI_ISL_1787017, EPI_ISL_1787018, EPI_ISL_1787019, EPI_ISL_1787020, EPI_ISL_1787022, EPI_ISL_1787023,
EPI_ISL_1787024, EPI_ISL_1787025, EPI_ISL_1787026, EPI_ISL_1787027, EPI_ISL_1787028, EPI_ISL_1787030, EPI_ISL_1787031, EPI_ISL_1787032, EPI_ISL_1787033, EPI_ISL_1787034, EPI_ISL_1787035, EPI_ISL_1787036, EPI_ISL_1787037, EPI_ISL_1787038, EPI_ISL_1787039, EPI_ISL_1787040, EPI_ISL_1787041, EPI_ISL_1787042, EPI_ISL_1787043, EPI_ISL_1787046, EPI_ISL_1787047, EPI_ISL_1787048,
EPI_ISL_1787049, EPI_ISL_1787050, EPI_ISL_1787051, EPI_ISL_1787052, EPI_ISL_1787053, EPI_ISL_1787054, EPI_ISL_1787055, EPI_ISL_1787056, EPI_ISL_1787057, EPI_ISL_1787058, EPI_ISL_1787060, EPI_ISL_1787061, EPI_ISL_1787062, EPI_ISL_1787063, EPI_ISL_1787064, EPI_ISL_1787065, EPI_ISL_1787066, EPI_ISL_1787067, EPI_ISL_1787068, EPI_ISL_1787069, EPI_ISL_1787072, EPI_ISL_1787073,
EPI_ISL_1787074, EPI_ISL_1787075, EPI_ISL_1787076, EPI_ISL_1787077, EPI_ISL_1787078, EPI_ISL_1787079, EPI_ISL_1787080, EPI_ISL_1787081, EPI_ISL_1787082, EPI_ISL_1787083, EPI_ISL_1787084, EPI_ISL_1787086, EPI_ISL_1787087, EPI_ISL_1787088, EPI_ISL_1787089, EPI_ISL_1787091, EPI_ISL_1787093, EPI_ISL_1787094, EPI_ISL_1787095, EPI_ISL_1787096, EPI_ISL_1787097, EPI_ISL_1787098,
EPI_ISL_1787099, EPI_ISL_1787100, EPI_ISL_1787101, EPI_ISL_1787102, EPI_ISL_1787103, EPI_ISL_1787104, EPI_ISL_1787106, EPI_ISL_1787107, EPI_ISL_1787108, EPI_ISL_1787110, EPI_ISL_1787111, EPI_ISL_1787112, EPI_ISL_1787114, EPI_ISL_1787115, EPI_ISL_1787116, EPI_ISL_1787117, EPI_ISL_1787118, EPI_ISL_1787119, EPI_ISL_1787120, EPI_ISL_1787121, EPI_ISL_1787122, EPI_ISL_1787123,
EPI_ISL_1787124, EPI_ISL_1787125, EPI_ISL_1787126, EPI_ISL_1787127, EPI_ISL_1787128, EPI_ISL_1787129, EPI_ISL_1787130, EPI_ISL_1787131, EPI_ISL_1787132, EPI_ISL_1787133, EPI_ISL_1787134, EPI_ISL_1787135, EPI_ISL_1787136, EPI_ISL_1787137, EPI_ISL_1787138, EPI_ISL_1787139, EPI_ISL_1787140, EPI_ISL_1787141, EPI_ISL_1787142, EPI_ISL_1787143, EPI_ISL_1787144, EPI_ISL_1787145,
EPI_ISL_1787146, EPI_ISL_1787148, EPI_ISL_1787149, EPI_ISL_1787150, EPI_ISL_1787151, EPI_ISL_1787152, EPI_ISL_1787153, EPI_ISL_1787154, EPI_ISL_1787156, EPI_ISL_1787157, EPI_ISL_1787158, EPI_ISL_1787159, EPI_ISL_1787161, EPI_ISL_1787162, EPI_ISL_1787163, EPI_ISL_1787164, EPI_ISL_1787165, EPI_ISL_1787166, EPI_ISL_1787168, EPI_ISL_1787169, EPI_ISL_1787170, EPI_ISL_1787172,
EPI_ISL_1787174, EPI_ISL_1787175, EPI_ISL_1787176, EPI_ISL_1787177, EPI_ISL_1787178, EPI_ISL_1787179, EPI_ISL_1787180, EPI_ISL_1787181, EPI_ISL_1787182, EPI_ISL_1787183, EPI_ISL_1787184, EPI_ISL_1787185, EPI_ISL_1787186, EPI_ISL_1787187, EPI_ISL_1787188, EPI_ISL_1787189, EPI_ISL_1787190, EPI_ISL_1787191, EPI_ISL_1787192, EPI_ISL_1787193, EPI_ISL_1787194, EPI_ISL_1787195,
EPI_ISL_1787196, EPI_ISL_1787198, EPI_ISL_1787199, EPI_ISL_1787200, EPI_ISL_1787202, EPI_ISL_1787203, EPI_ISL_1787204, EPI_ISL_1787205, EPI_ISL_1787206, EPI_ISL_1787207, EPI_ISL_1787208, EPI_ISL_1787209, EPI_ISL_1787210, EPI_ISL_1787212, EPI_ISL_1787213, EPI_ISL_1787214, EPI_ISL_1787215, EPI_ISL_1787216, EPI_ISL_1787217, EPI_ISL_1787219, EPI_ISL_1787221, EPI_ISL_1787222,
EPI_ISL_1787223, EPI_ISL_1787224, EPI_ISL_1787225, EPI_ISL_1787226, EPI_ISL_1787227, EPI_ISL_1787229, EPI_ISL_1787230, EPI_ISL_1825144, EPI_ISL_1825145, EPI_ISL_1825146, EPI_ISL_1825147, EPI_ISL_1825149, EPI_ISL_1825150, EPI_ISL_1825151, EPI_ISL_1825152, EPI_ISL_1825153, EPI_ISL_1825154, EPI_ISL_1825155, EPI_ISL_1825156, EPI_ISL_1825157, EPI_ISL_1825158, EPI_ISL_1825159,
EPI_ISL_1825160, EPI_ISL_1825161, EPI_ISL_1825162, EPI_ISL_1825163, EPI_ISL_1825165, EPI_ISL_1825166, EPI_ISL_1825167, EPI_ISL_1825168, EPI_ISL_1825169, EPI_ISL_1825170, EPI_ISL_1825171, EPI_ISL_1825172, EPI_ISL_1825173, EPI_ISL_1825174, EPI_ISL_1825175, EPI_ISL_1825177, EPI_ISL_1825178, EPI_ISL_1825179, EPI_ISL_1825180, EPI_ISL_1825182, EPI_ISL_1825184, EPI_ISL_1825186,
EPI_ISL_1825187, EPI_ISL_1825188, EPI_ISL_1825190, EPI_ISL_1825191, EPI_ISL_1825192, EPI_ISL_1825193, EPI_ISL_1825194, EPI_ISL_1825195, EPI_ISL_1825196, EPI_ISL_1825198, EPI_ISL_1825199, EPI_ISL_1825200, EPI_ISL_1825201, EPI_ISL_1825202, EPI_ISL_1825203, EPI_ISL_1825204, EPI_ISL_1825205, EPI_ISL_1825206, EPI_ISL_1825207, EPI_ISL_1825208, EPI_ISL_1825210, EPI_ISL_1825212,
EPI_ISL_1825221, EPI_ISL_1825224, EPI_ISL_1825255, EPI_ISL_1825256, EPI_ISL_1825257, EPI_ISL_1825260, EPI_ISL_1825261, EPI_ISL_1825262, EPI_ISL_1825264, EPI_ISL_1825265, EPI_ISL_1825266, EPI_ISL_1825273, EPI_ISL_1825278, EPI_ISL_1825279, EPI_ISL_1825280, EPI_ISL_1825281, EPI_ISL_1825288, EPI_ISL_1825289, EPI_ISL_1825290, EPI_ISL_1825291, EPI_ISL_1825293, EPI_ISL_1825296,
EPI_ISL_1825297, EPI_ISL_1825298, EPI_ISL_1825300, EPI_ISL_1825301, EPI_ISL_1825302, EPI_ISL_1825303, EPI_ISL_1825305, EPI_ISL_1825306, EPI_ISL_1825307, EPI_ISL_1825308, EPI_ISL_1825309, EPI_ISL_1825310, EPI_ISL_1825311, EPI_ISL_1825312, EPI_ISL_1825313, EPI_ISL_1825315, EPI_ISL_1825316, EPI_ISL_1825317, EPI_ISL_1825318, EPI_ISL_1825319, EPI_ISL_1825320, EPI_ISL_1825321,
EPI_ISL_1825322, EPI_ISL_1825323, EPI_ISL_1825324, EPI_ISL_1825325, EPI_ISL_1825326, EPI_ISL_1825327, EPI_ISL_1825328, EPI_ISL_1825329, EPI_ISL_1825330, EPI_ISL_1825331, EPI_ISL_1825332, EPI_ISL_1825333, EPI_ISL_1825337, EPI_ISL_1825338, EPI_ISL_1825339, EPI_ISL_1825340, EPI_ISL_1825341, EPI_ISL_1825342, EPI_ISL_1825343, EPI_ISL_1825344, EPI_ISL_1825345, EPI_ISL_1825346,
EPI_ISL_1825348, EPI_ISL_1825349, EPI_ISL_1825350, EPI_ISL_1825351, EPI_ISL_1825352, EPI_ISL_1825353, EPI_ISL_1825354, EPI_ISL_1825355, EPI_ISL_1825356, EPI_ISL_1825357, EPI_ISL_1825358, EPI_ISL_1825359, EPI_ISL_1825360, EPI_ISL_1825361, EPI_ISL_1825362, EPI_ISL_1825363, EPI_ISL_1825364, EPI_ISL_1825365, EPI_ISL_1825366, EPI_ISL_1825367, EPI_ISL_1825369, EPI_ISL_1825370,
EPI_ISL_1825371, EPI_ISL_1825375, EPI_ISL_1825376, EPI_ISL_1825377, EPI_ISL_1825378, EPI_ISL_1825380, EPI_ISL_1825382, EPI_ISL_1825383, EPI_ISL_1825384, EPI_ISL_1825386, EPI_ISL_1825387, EPI_ISL_1825388, EPI_ISL_1825391, EPI_ISL_1825392, EPI_ISL_1825393, EPI_ISL_1825394, EPI_ISL_1825395, EPI_ISL_1825396, EPI_ISL_1825397, EPI_ISL_1825398, EPI_ISL_1825400, EPI_ISL_1825401,
EPI_ISL_1825405, EPI_ISL_1825406, EPI_ISL_1825407, EPI_ISL_1825408, EPI_ISL_1825409, EPI_ISL_1825410, EPI_ISL_1825411, EPI_ISL_1825412, EPI_ISL_1825414, EPI_ISL_1825415, EPI_ISL_1825416, EPI_ISL_1825417, EPI_ISL_1825418, EPI_ISL_1825419, EPI_ISL_1825420, EPI_ISL_1825421, EPI_ISL_1825422, EPI_ISL_1825423, EPI_ISL_1825424, EPI_ISL_1825425, EPI_ISL_1825426, EPI_ISL_1825427,
EPI_ISL_1825428, EPI_ISL_1825429, EPI_ISL_1825430, EPI_ISL_1825431, EPI_ISL_1825432, EPI_ISL_1825433, EPI_ISL_1825434, EPI_ISL_1825435, EPI_ISL_1825436, EPI_ISL_1825437, EPI_ISL_1825438, EPI_ISL_1825439, EPI_ISL_1825440, EPI_ISL_1825441, EPI_ISL_1825442, EPI_ISL_1825443, EPI_ISL_1825444, EPI_ISL_1825445, EPI_ISL_1825446, EPI_ISL_1825447, EPI_ISL_1825448, EPI_ISL_1825449,
EPI_ISL_1825450, EPI_ISL_1825451, EPI_ISL_1825452, EPI_ISL_1825457, EPI_ISL_1825459, EPI_ISL_1825460, EPI_ISL_1825461, EPI_ISL_1825462, EPI_ISL_1825463, EPI_ISL_1825466, EPI_ISL_1825468, EPI_ISL_1825470, EPI_ISL_1825471, EPI_ISL_1825472, EPI_ISL_1825477, EPI_ISL_1825490, EPI_ISL_1825493, EPI_ISL_1825499, EPI_ISL_1825502, EPI_ISL_1825503, EPI_ISL_1825507, EPI_ISL_1825508,
EPI_ISL_1825510, EPI_ISL_1825513, EPI_ISL_1825520, EPI_ISL_1825522, EPI_ISL_1825523, EPI_ISL_1825525, EPI_ISL_1825526, EPI_ISL_1825530, EPI_ISL_1825533, EPI_ISL_1825536, EPI_ISL_1825537, EPI_ISL_1825539, EPI_ISL_1825541, EPI_ISL_1825542, EPI_ISL_1825543, EPI_ISL_1825544, EPI_ISL_1825545, EPI_ISL_1825546, EPI_ISL_1825548, EPI_ISL_1825549, EPI_ISL_1825550, EPI_ISL_1825551,
EPI_ISL_1825552, EPI_ISL_1825553, EPI_ISL_1825554, EPI_ISL_1825555, EPI_ISL_1825557, EPI_ISL_1825560, EPI_ISL_1825562, EPI_ISL_1825566, EPI_ISL_1825567, EPI_ISL_1825568, EPI_ISL_1825569, EPI_ISL_1825570, EPI_ISL_1825575, EPI_ISL_1825576, EPI_ISL_1825577, EPI_ISL_1825581, EPI_ISL_1825582, EPI_ISL_1825583, EPI_ISL_1825585, EPI_ISL_1825586, EPI_ISL_1825587, EPI_ISL_1825591,
EPI_ISL_1825593, EPI_ISL_1825594, EPI_ISL_1825595, EPI_ISL_1825596, EPI_ISL_1825597, EPI_ISL_1825598, EPI_ISL_1825599, EPI_ISL_1825600, EPI_ISL_1825602, EPI_ISL_1825603, EPI_ISL_1825604, EPI_ISL_1825605, EPI_ISL_1825606, EPI_ISL_1825607, EPI_ISL_1825608, EPI_ISL_1825609, EPI_ISL_1825610, EPI_ISL_1825611, EPI_ISL_1825612, EPI_ISL_1825613, EPI_ISL_1825614, EPI_ISL_1825615,
EPI_ISL_1825616, EPI_ISL_1825617, EPI_ISL_1825618, EPI_ISL_1825619, EPI_ISL_1825620, EPI_ISL_1825622, EPI_ISL_1825623, EPI_ISL_1825625, EPI_ISL_1825626, EPI_ISL_1825627, EPI_ISL_1825628, EPI_ISL_1825629, EPI_ISL_1825630, EPI_ISL_1825631, EPI_ISL_1825632, EPI_ISL_1825633, EPI_ISL_1825634, EPI_ISL_1825635, EPI_ISL_1825636, EPI_ISL_1825637, EPI_ISL_1825638, EPI_ISL_1825640,
EPI_ISL_1825641, EPI_ISL_1825642, EPI_ISL_1825643, EPI_ISL_1825644, EPI_ISL_1825645, EPI_ISL_1825646, EPI_ISL_1825647, EPI_ISL_1825648, EPI_ISL_1825649, EPI_ISL_1825650, EPI_ISL_1825651, EPI_ISL_1825652, EPI_ISL_1825654, EPI_ISL_1825656, EPI_ISL_1825657, EPI_ISL_1825658, EPI_ISL_1825659, EPI_ISL_1825661, EPI_ISL_1825662, EPI_ISL_1825663, EPI_ISL_1825664, EPI_ISL_1825665,
EPI_ISL_1825666, EPI_ISL_1825667, EPI_ISL_1825668, EPI_ISL_1825669, EPI_ISL_1825670, EPI_ISL_1825671, EPI_ISL_1825672, EPI_ISL_1825673, EPI_ISL_1825674, EPI_ISL_1825675, EPI_ISL_1918411, EPI_ISL_1918412, EPI_ISL_1918413, EPI_ISL_1918414, EPI_ISL_1918416, EPI_ISL_1918417, EPI_ISL_1918418, EPI_ISL_1918419, EPI_ISL_1918421, EPI_ISL_1918422, EPI_ISL_1918423, EPI_ISL_1918424,
EPI_ISL_1918425, EPI_ISL_1918426, EPI_ISL_1918427, EPI_ISL_1918429, EPI_ISL_1918430, EPI_ISL_1918431, EPI_ISL_1918432, EPI_ISL_1918433, EPI_ISL_1918434, EPI_ISL_1918435, EPI_ISL_1918436, EPI_ISL_1918437, EPI_ISL_1918438, EPI_ISL_1918439, EPI_ISL_1918440, EPI_ISL_1918441, EPI_ISL_1918442, EPI_ISL_1918443, EPI_ISL_1918444, EPI_ISL_1918445, EPI_ISL_1918446, EPI_ISL_1918447,
EPI_ISL_1918449, EPI_ISL_1918451, EPI_ISL_1918452, EPI_ISL_1918453, EPI_ISL_1918454, EPI_ISL_1918455, EPI_ISL_1918457, EPI_ISL_1918458, EPI_ISL_1918460, EPI_ISL_1918461, EPI_ISL_1918462, EPI_ISL_1918463, EPI_ISL_1918464, EPI_ISL_1918466, EPI_ISL_1918467, EPI_ISL_1918468, EPI_ISL_1918469, EPI_ISL_1918470, EPI_ISL_1918472, EPI_ISL_1918473, EPI_ISL_1918474, EPI_ISL_1918475,
EPI_ISL_1918476, EPI_ISL_1918477, EPI_ISL_1918478, EPI_ISL_1918479, EPI_ISL_1918480, EPI_ISL_1918482, EPI_ISL_1918483, EPI_ISL_1918485, EPI_ISL_1918486, EPI_ISL_1918487, EPI_ISL_1918488, EPI_ISL_1918489, EPI_ISL_1918491, EPI_ISL_1918492, EPI_ISL_1918493, EPI_ISL_1918494, EPI_ISL_1918495, EPI_ISL_1918496, EPI_ISL_1918497, EPI_ISL_1918499, EPI_ISL_1918500, EPI_ISL_1918501,
EPI_ISL_1918502, EPI_ISL_1918503, EPI_ISL_1918504, EPI_ISL_1918506, EPI_ISL_1918507, EPI_ISL_1918508, EPI_ISL_1918509, EPI_ISL_1918510, EPI_ISL_1918511, EPI_ISL_1918512, EPI_ISL_1918513, EPI_ISL_1918514, EPI_ISL_1918515, EPI_ISL_1918516, EPI_ISL_1918517, EPI_ISL_1918518, EPI_ISL_1918520, EPI_ISL_1918521, EPI_ISL_1918522, EPI_ISL_1918523, EPI_ISL_1918525, EPI_ISL_1918526,
EPI_ISL_1918527, EPI_ISL_1918528, EPI_ISL_1918529, EPI_ISL_1918530, EPI_ISL_1918531, EPI_ISL_1918532, EPI_ISL_1918533, EPI_ISL_1918535, EPI_ISL_1918536, EPI_ISL_1918537, EPI_ISL_1918538, EPI_ISL_1918539, EPI_ISL_1918540, EPI_ISL_1918541, EPI_ISL_1918542, EPI_ISL_1918543, EPI_ISL_1918544, EPI_ISL_1918545, EPI_ISL_1918546, EPI_ISL_1918547, EPI_ISL_1918549, EPI_ISL_1918550,
EPI_ISL_1918551, EPI_ISL_1918552, EPI_ISL_1918553, EPI_ISL_1918554, EPI_ISL_1918555, EPI_ISL_1918556, EPI_ISL_1918557, EPI_ISL_1918559, EPI_ISL_1918560, EPI_ISL_1918561, EPI_ISL_1918562, EPI_ISL_1918563, EPI_ISL_1918564, EPI_ISL_1918566, EPI_ISL_1918567, EPI_ISL_1918568, EPI_ISL_1918569, EPI_ISL_1918570, EPI_ISL_1918571, EPI_ISL_1918572, EPI_ISL_1918573, EPI_ISL_1918574,
EPI_ISL_1918575, EPI_ISL_1918576, EPI_ISL_1918578, EPI_ISL_1918579, EPI_ISL_1918580, EPI_ISL_1918581, EPI_ISL_1918582, EPI_ISL_1918583, EPI_ISL_1918584, EPI_ISL_1918585, EPI_ISL_1918586, EPI_ISL_1918587, EPI_ISL_1918588, EPI_ISL_1918589, EPI_ISL_1918590, EPI_ISL_1918591, EPI_ISL_1918592, EPI_ISL_1918593, EPI_ISL_1918594, EPI_ISL_1918595, EPI_ISL_1918596, EPI_ISL_1918599,
EPI_ISL_1918600, EPI_ISL_1918602, EPI_ISL_1918603, EPI_ISL_1918604, EPI_ISL_1918606, EPI_ISL_1918607, EPI_ISL_1918608, EPI_ISL_1918609, EPI_ISL_1918610, EPI_ISL_1918611, EPI_ISL_1918612, EPI_ISL_1918614, EPI_ISL_1918615, EPI_ISL_1918616, EPI_ISL_1918617, EPI_ISL_1918618, EPI_ISL_1918619, EPI_ISL_1918620, EPI_ISL_1918621, EPI_ISL_1918622, EPI_ISL_1918623, EPI_ISL_1918624,
EPI_ISL_1918625, EPI_ISL_1918626, EPI_ISL_1918627, EPI_ISL_1918628, EPI_ISL_1918629, EPI_ISL_1918630, EPI_ISL_1918631, EPI_ISL_1918632, EPI_ISL_1918633, EPI_ISL_1918634, EPI_ISL_1918635, EPI_ISL_1918637, EPI_ISL_1918638, EPI_ISL_1918640, EPI_ISL_1918641, EPI_ISL_1918642, EPI_ISL_1918643, EPI_ISL_1918644, EPI_ISL_1918646, EPI_ISL_1918647, EPI_ISL_1918648, EPI_ISL_1918649,
EPI_ISL_1918651, EPI_ISL_1918652, EPI_ISL_1918653, EPI_ISL_1918654, EPI_ISL_1918655, EPI_ISL_1918656, EPI_ISL_1918657, EPI_ISL_1918658, EPI_ISL_1918659, EPI_ISL_1918661, EPI_ISL_1918662, EPI_ISL_1918664, EPI_ISL_1918666, EPI_ISL_1918667, EPI_ISL_1918669, EPI_ISL_1918670, EPI_ISL_1918671, EPI_ISL_1918672, EPI_ISL_1918673, EPI_ISL_1918674, EPI_ISL_1918675, EPI_ISL_1918676,
EPI_ISL_1918677, EPI_ISL_1918678, EPI_ISL_1918679, EPI_ISL_1918680, EPI_ISL_1918681, EPI_ISL_1918682, EPI_ISL_1918683, EPI_ISL_1918684, EPI_ISL_1918685, EPI_ISL_1918686, EPI_ISL_1918688, EPI_ISL_1918689, EPI_ISL_1918690, EPI_ISL_1918691, EPI_ISL_1918692, EPI_ISL_1918694, EPI_ISL_1918695, EPI_ISL_1918696, EPI_ISL_1918697, EPI_ISL_1918698, EPI_ISL_1918699, EPI_ISL_1918700,
EPI_ISL_1918701, EPI_ISL_1918702, EPI_ISL_1918703, EPI_ISL_1918704, EPI_ISL_1918705, EPI_ISL_1918706, EPI_ISL_1918707, EPI_ISL_1918708, EPI_ISL_1918709, EPI_ISL_1918710, EPI_ISL_1918711, EPI_ISL_1918712, EPI_ISL_1918713, EPI_ISL_1918714, EPI_ISL_1918715, EPI_ISL_1918716, EPI_ISL_1918717, EPI_ISL_1918718, EPI_ISL_1918719, EPI_ISL_1918720, EPI_ISL_1918721, EPI_ISL_1918722,
EPI_ISL_1918723, EPI_ISL_1918724, EPI_ISL_1918725, EPI_ISL_1918726, EPI_ISL_1918727, EPI_ISL_1918728, EPI_ISL_1918729, EPI_ISL_1918730, EPI_ISL_1918731, EPI_ISL_1918732, EPI_ISL_1918733, EPI_ISL_1918734, EPI_ISL_1918735, EPI_ISL_1918736, EPI_ISL_1918737, EPI_ISL_1918738, EPI_ISL_1918739, EPI_ISL_1918740, EPI_ISL_1918742, EPI_ISL_1918743, EPI_ISL_1918744, EPI_ISL_1918745,
EPI_ISL_1918746, EPI_ISL_1918747, EPI_ISL_1918749, EPI_ISL_1918750, EPI_ISL_1918751, EPI_ISL_1918752, EPI_ISL_1918754, EPI_ISL_1918755, EPI_ISL_1918756, EPI_ISL_1918757, EPI_ISL_1918758, EPI_ISL_1918759, EPI_ISL_1918762, EPI_ISL_1918763, EPI_ISL_1918764, EPI_ISL_1918765, EPI_ISL_1918766, EPI_ISL_1918767, EPI_ISL_1918769, EPI_ISL_1918771, EPI_ISL_1918773, EPI_ISL_1918774,
EPI_ISL_1918776, EPI_ISL_1918777, EPI_ISL_1918778, EPI_ISL_1918780, EPI_ISL_1918781, EPI_ISL_1918782, EPI_ISL_1918783, EPI_ISL_1918784, EPI_ISL_1918785, EPI_ISL_1918786, EPI_ISL_1918787, EPI_ISL_1918788, EPI_ISL_1918789, EPI_ISL_1918790, EPI_ISL_1918791, EPI_ISL_1918792, EPI_ISL_1918793, EPI_ISL_1918794, EPI_ISL_1918796, EPI_ISL_1918797, EPI_ISL_1918798, EPI_ISL_1918799,
EPI_ISL_1918800, EPI_ISL_1918801, EPI_ISL_1918802, EPI_ISL_1918803, EPI_ISL_1918804, EPI_ISL_1918806, EPI_ISL_1918807, EPI_ISL_1918808, EPI_ISL_1918809, EPI_ISL_1918810, EPI_ISL_1918811, EPI_ISL_1918813, EPI_ISL_1918814, EPI_ISL_1918815, EPI_ISL_1918816, EPI_ISL_1918817, EPI_ISL_1918818, EPI_ISL_1918819, EPI_ISL_1918820, EPI_ISL_1918821, EPI_ISL_1918822, EPI_ISL_1918823,
EPI_ISL_1918824, EPI_ISL_1918826, EPI_ISL_1918827, EPI_ISL_1918829, EPI_ISL_1918830, EPI_ISL_1918832, EPI_ISL_1918833, EPI_ISL_1918835, EPI_ISL_1918836, EPI_ISL_1918837, EPI_ISL_1918838, EPI_ISL_1918840, EPI_ISL_1918841, EPI_ISL_1918843, EPI_ISL_1918844, EPI_ISL_1918845, EPI_ISL_1918846, EPI_ISL_1918847, EPI_ISL_1918848, EPI_ISL_1918849, EPI_ISL_1918850, EPI_ISL_1918851,
EPI_ISL_1918853, EPI_ISL_1918854, EPI_ISL_1918855, EPI_ISL_1918856, EPI_ISL_1918857, EPI_ISL_1918858, EPI_ISL_1918859, EPI_ISL_1918860, EPI_ISL_1918861, EPI_ISL_1918862, EPI_ISL_1918864, EPI_ISL_1918865, EPI_ISL_1918867, EPI_ISL_1918868, EPI_ISL_1918869, EPI_ISL_1918870, EPI_ISL_1918871, EPI_ISL_1918872, EPI_ISL_1918873, EPI_ISL_1918874, EPI_ISL_1918875, EPI_ISL_1918876,
EPI_ISL_1918877, EPI_ISL_1918878, EPI_ISL_1918879, EPI_ISL_1918880, EPI_ISL_1918881, EPI_ISL_1918882, EPI_ISL_1918883, EPI_ISL_1918884, EPI_ISL_1918885, EPI_ISL_1918886, EPI_ISL_1918887, EPI_ISL_1918888, EPI_ISL_1918889, EPI_ISL_1918891, EPI_ISL_1918892, EPI_ISL_1918893, EPI_ISL_1918894, EPI_ISL_1918895, EPI_ISL_1918896, EPI_ISL_1918897, EPI_ISL_1918898, EPI_ISL_1918899,
EPI_ISL_1918900, EPI_ISL_1918901, EPI_ISL_1918902, EPI_ISL_1918904, EPI_ISL_1918905, EPI_ISL_1918906, EPI_ISL_1918907, EPI_ISL_1918908, EPI_ISL_1918909, EPI_ISL_1918910, EPI_ISL_1918911, EPI_ISL_1918912, EPI_ISL_1918913, EPI_ISL_1918915, EPI_ISL_1918917, EPI_ISL_1918919, EPI_ISL_1918920, EPI_ISL_1918921, EPI_ISL_1918922, EPI_ISL_1918923, EPI_ISL_1918924, EPI_ISL_1918925,
EPI_ISL_1918926, EPI_ISL_1918927, EPI_ISL_1918928, EPI_ISL_1918929, EPI_ISL_1918930, EPI_ISL_1918931, EPI_ISL_1918932, EPI_ISL_1918934, EPI_ISL_1918935, EPI_ISL_1918936, EPI_ISL_1918937, EPI_ISL_1918938, EPI_ISL_1918939, EPI_ISL_1918940, EPI_ISL_1918941, EPI_ISL_1918942, EPI_ISL_1918943, EPI_ISL_1918944, EPI_ISL_1918945, EPI_ISL_1918946, EPI_ISL_1918947, EPI_ISL_1919014,
EPI_ISL_1919016, EPI_ISL_1919017, EPI_ISL_1919018, EPI_ISL_1919019, EPI_ISL_1919020, EPI_ISL_1919021, EPI_ISL_1919022, EPI_ISL_1919023, EPI_ISL_1919024, EPI_ISL_1919025, EPI_ISL_1919026, EPI_ISL_1919027, EPI_ISL_1919028, EPI_ISL_1919029, EPI_ISL_1919030, EPI_ISL_1919031, EPI_ISL_1919032, EPI_ISL_1919034, EPI_ISL_1919035, EPI_ISL_1919036, EPI_ISL_1919037, EPI_ISL_1919038,
EPI_ISL_1919039, EPI_ISL_1919040, EPI_ISL_1919041, EPI_ISL_1919042, EPI_ISL_1919043, EPI_ISL_1919044, EPI_ISL_1919045, EPI_ISL_1919046, EPI_ISL_1919047, EPI_ISL_1967957, EPI_ISL_1967958, EPI_ISL_1967959, EPI_ISL_1967962, EPI_ISL_1967963, EPI_ISL_1967964, EPI_ISL_1967965, EPI_ISL_1967966, EPI_ISL_1967967, EPI_ISL_1967969, EPI_ISL_1967970, EPI_ISL_1967971, EPI_ISL_1967972,
EPI_ISL_1967973, EPI_ISL_1967974, EPI_ISL_1967975, EPI_ISL_1967976, EPI_ISL_1967977, EPI_ISL_1967978, EPI_ISL_1967979, EPI_ISL_1967981, EPI_ISL_1967982, EPI_ISL_1967983, EPI_ISL_1967984, EPI_ISL_1967985, EPI_ISL_1967986, EPI_ISL_1967987, EPI_ISL_1967988, EPI_ISL_1967989, EPI_ISL_1967990, EPI_ISL_1967993, EPI_ISL_1967994, EPI_ISL_1967996, EPI_ISL_1967997, EPI_ISL_1967998,
EPI_ISL_1967999, EPI_ISL_1968000, EPI_ISL_1968002, EPI_ISL_1968003, EPI_ISL_1968004, EPI_ISL_1968005, EPI_ISL_1968006, EPI_ISL_1968007, EPI_ISL_1968009, EPI_ISL_1968011, EPI_ISL_1968012, EPI_ISL_1968013, EPI_ISL_1968014, EPI_ISL_1968015, EPI_ISL_1968016, EPI_ISL_1968017, EPI_ISL_1968018, EPI_ISL_1968020, EPI_ISL_1968021, EPI_ISL_1968023, EPI_ISL_1968024, EPI_ISL_1968025,
EPI_ISL_1968026, EPI_ISL_1968027, EPI_ISL_1968028, EPI_ISL_1968029, EPI_ISL_1968030, EPI_ISL_1968031, EPI_ISL_1968032, EPI_ISL_1968033, EPI_ISL_1968034, EPI_ISL_1968035, EPI_ISL_1968036, EPI_ISL_1968037, EPI_ISL_1968038, EPI_ISL_1968039, EPI_ISL_1968040, EPI_ISL_1968041, EPI_ISL_1968042, EPI_ISL_1968043, EPI_ISL_1968044, EPI_ISL_1968045, EPI_ISL_1968046, EPI_ISL_1968047,
EPI_ISL_1968048, EPI_ISL_1968049, EPI_ISL_1968050, EPI_ISL_1968052, EPI_ISL_1968053, EPI_ISL_1968054, EPI_ISL_1968055, EPI_ISL_1968056, EPI_ISL_1968058, EPI_ISL_1968060, EPI_ISL_1968062, EPI_ISL_1968063, EPI_ISL_1968064, EPI_ISL_1968066, EPI_ISL_1968067, EPI_ISL_1968068, EPI_ISL_1968069, EPI_ISL_1968070, EPI_ISL_1968071, EPI_ISL_1968072, EPI_ISL_1968073, EPI_ISL_1968074,
EPI_ISL_1968075, EPI_ISL_1968076, EPI_ISL_1968078, EPI_ISL_1968079, EPI_ISL_1968080, EPI_ISL_1968081, EPI_ISL_1968082, EPI_ISL_1968083, EPI_ISL_1968084, EPI_ISL_1968085, EPI_ISL_1968086, EPI_ISL_1968087, EPI_ISL_1968088, EPI_ISL_1968089, EPI_ISL_1968090, EPI_ISL_1968091, EPI_ISL_1968093, EPI_ISL_1968094, EPI_ISL_1968095, EPI_ISL_1968096, EPI_ISL_1968097, EPI_ISL_1968100,
EPI_ISL_1968101, EPI_ISL_1968102, EPI_ISL_1968103, EPI_ISL_1968104, EPI_ISL_1968105, EPI_ISL_1968106, EPI_ISL_1968107, EPI_ISL_1968109, EPI_ISL_1968110, EPI_ISL_1968112, EPI_ISL_1968114, EPI_ISL_1968115, EPI_ISL_1968116, EPI_ISL_1968117, EPI_ISL_1968118, EPI_ISL_1968119, EPI_ISL_1968120, EPI_ISL_1968121, EPI_ISL_1968122, EPI_ISL_1968123, EPI_ISL_1968124, EPI_ISL_1968125,
EPI_ISL_1968126, EPI_ISL_1968127, EPI_ISL_1968128, EPI_ISL_1968129, EPI_ISL_1968130, EPI_ISL_1968131, EPI_ISL_1968132, EPI_ISL_1968134, EPI_ISL_1968135, EPI_ISL_1968136, EPI_ISL_1968137, EPI_ISL_1968138, EPI_ISL_1968139, EPI_ISL_1968140, EPI_ISL_1968142, EPI_ISL_1968144, EPI_ISL_1968145, EPI_ISL_1968146, EPI_ISL_1968147, EPI_ISL_1968148, EPI_ISL_1968150, EPI_ISL_1968151,
EPI_ISL_1968153, EPI_ISL_1968154, EPI_ISL_1968155, EPI_ISL_1968156, EPI_ISL_1968157, EPI_ISL_1968158, EPI_ISL_1968159, EPI_ISL_1968161, EPI_ISL_1968162, EPI_ISL_1968163, EPI_ISL_1968164, EPI_ISL_1968166, EPI_ISL_1968169, EPI_ISL_1968170, EPI_ISL_1968171, EPI_ISL_1968172, EPI_ISL_1968173, EPI_ISL_1968174, EPI_ISL_1968175, EPI_ISL_1968176, EPI_ISL_1968178, EPI_ISL_1968179,
EPI_ISL_1968180, EPI_ISL_1968181, EPI_ISL_1968182, EPI_ISL_1968183, EPI_ISL_1968185, EPI_ISL_1968186, EPI_ISL_1968187, EPI_ISL_1968188, EPI_ISL_1968189, EPI_ISL_1968190, EPI_ISL_1968191, EPI_ISL_1968192, EPI_ISL_1968195, EPI_ISL_1968196, EPI_ISL_1968197, EPI_ISL_1968198, EPI_ISL_1968199, EPI_ISL_1968200, EPI_ISL_1968201, EPI_ISL_1968202, EPI_ISL_1968203, EPI_ISL_1968204,
EPI_ISL_1968205, EPI_ISL_1968206, EPI_ISL_1968207, EPI_ISL_1968208, EPI_ISL_1968209, EPI_ISL_1968210, EPI_ISL_1968211, EPI_ISL_1968212, EPI_ISL_1968213, EPI_ISL_1968214, EPI_ISL_1968217, EPI_ISL_1968218, EPI_ISL_1968219, EPI_ISL_1968220, EPI_ISL_1968221, EPI_ISL_1968222, EPI_ISL_1968223, EPI_ISL_1968224, EPI_ISL_1968225, EPI_ISL_1968226, EPI_ISL_1968227, EPI_ISL_1968228,
EPI_ISL_1968229, EPI_ISL_1968230, EPI_ISL_1968231, EPI_ISL_1968232, EPI_ISL_1968233, EPI_ISL_1968234, EPI_ISL_1968235, EPI_ISL_1968236, EPI_ISL_1968237, EPI_ISL_1968238, EPI_ISL_1968240, EPI_ISL_1968241, EPI_ISL_1968242, EPI_ISL_1968243, EPI_ISL_1968245, EPI_ISL_1968246, EPI_ISL_1968247, EPI_ISL_1968248, EPI_ISL_1968249, EPI_ISL_1968250, EPI_ISL_1968251, EPI_ISL_1968255,
EPI_ISL_1968256, EPI_ISL_1968257, EPI_ISL_1968258, EPI_ISL_1968259, EPI_ISL_1968260, EPI_ISL_1968261, EPI_ISL_1968262, EPI_ISL_1968263, EPI_ISL_1968264, EPI_ISL_1968265, EPI_ISL_1968266, EPI_ISL_1968267, EPI_ISL_1968268, EPI_ISL_1968269, EPI_ISL_1968270, EPI_ISL_1968271, EPI_ISL_1968272, EPI_ISL_1968273, EPI_ISL_1968274, EPI_ISL_1968275, EPI_ISL_1968276, EPI_ISL_1968277,
EPI_ISL_1968279, EPI_ISL_1968280, EPI_ISL_1968282, EPI_ISL_1968283, EPI_ISL_1968284, EPI_ISL_1968286, EPI_ISL_1968287, EPI_ISL_1968288, EPI_ISL_1968289, EPI_ISL_1968290, EPI_ISL_1968291, EPI_ISL_1968292, EPI_ISL_1968293, EPI_ISL_1968294, EPI_ISL_1968295, EPI_ISL_1968296, EPI_ISL_1968297, EPI_ISL_1968298, EPI_ISL_1968300, EPI_ISL_1968302, EPI_ISL_1968303, EPI_ISL_1968304,
EPI_ISL_1968306, EPI_ISL_1968309, EPI_ISL_1968310, EPI_ISL_1968311, EPI_ISL_1968313, EPI_ISL_1968314, EPI_ISL_1968315, EPI_ISL_1968316, EPI_ISL_1968317, EPI_ISL_1968318, EPI_ISL_1968319, EPI_ISL_1968320, EPI_ISL_1968322, EPI_ISL_1968323, EPI_ISL_1968324, EPI_ISL_1968325, EPI_ISL_1968327, EPI_ISL_1968328, EPI_ISL_1968329, EPI_ISL_1968330, EPI_ISL_1968331, EPI_ISL_1968332,
EPI_ISL_1968333, EPI_ISL_1968334, EPI_ISL_1968335, EPI_ISL_1968336, EPI_ISL_1968337, EPI_ISL_1968338, EPI_ISL_1968339, EPI_ISL_1968340, EPI_ISL_1968341, EPI_ISL_1968342, EPI_ISL_1968343, EPI_ISL_1968344, EPI_ISL_1968345, EPI_ISL_1968347, EPI_ISL_1968348, EPI_ISL_1968350, EPI_ISL_1968351, EPI_ISL_1968352, EPI_ISL_1968353, EPI_ISL_1968354, EPI_ISL_1968355, EPI_ISL_1968356,
EPI_ISL_1968357, EPI_ISL_1968358, EPI_ISL_1968359, EPI_ISL_1968360, EPI_ISL_1968361, EPI_ISL_1968362, EPI_ISL_1968363, EPI_ISL_1968364, EPI_ISL_1968365, EPI_ISL_1968366, EPI_ISL_1968367, EPI_ISL_1968368, EPI_ISL_1968369, EPI_ISL_1968370, EPI_ISL_1968371, EPI_ISL_1968372, EPI_ISL_1968373, EPI_ISL_1968374, EPI_ISL_1968375, EPI_ISL_1968376, EPI_ISL_1968378, EPI_ISL_1968379,
EPI_ISL_1968380, EPI_ISL_1968381, EPI_ISL_1968383, EPI_ISL_1968384, EPI_ISL_1968385, EPI_ISL_1968386, EPI_ISL_1968387, EPI_ISL_1968388, EPI_ISL_1968389, EPI_ISL_1968390, EPI_ISL_1968392, EPI_ISL_1968393, EPI_ISL_1968394, EPI_ISL_1968395, EPI_ISL_1968396, EPI_ISL_1968397, EPI_ISL_1968398, EPI_ISL_1968400, EPI_ISL_1968401, EPI_ISL_1968402, EPI_ISL_1968403, EPI_ISL_1968404,
EPI_ISL_1968405, EPI_ISL_1968406, EPI_ISL_1968407, EPI_ISL_1968408, EPI_ISL_1968409, EPI_ISL_1968411, EPI_ISL_1968412, EPI_ISL_1968413, EPI_ISL_1968414, EPI_ISL_1968416, EPI_ISL_1968417, EPI_ISL_1968418, EPI_ISL_1968420, EPI_ISL_1968422, EPI_ISL_1968423, EPI_ISL_1968424, EPI_ISL_1968425, EPI_ISL_1968426, EPI_ISL_1968427, EPI_ISL_1968428, EPI_ISL_1968430, EPI_ISL_1968431,
EPI_ISL_1968432, EPI_ISL_1968433, EPI_ISL_1968435, EPI_ISL_1968436, EPI_ISL_1968437, EPI_ISL_1968438, EPI_ISL_1968439, EPI_ISL_1968440, EPI_ISL_1968441, EPI_ISL_1968442, EPI_ISL_1968443, EPI_ISL_1968445, EPI_ISL_1968446, EPI_ISL_1968448, EPI_ISL_1968450, EPI_ISL_1968451, EPI_ISL_1968452, EPI_ISL_1968453, EPI_ISL_1968454, EPI_ISL_1968455, EPI_ISL_1968456, EPI_ISL_1968457,
EPI_ISL_1968458, EPI_ISL_1968459, EPI_ISL_1968460, EPI_ISL_1968461, EPI_ISL_1968462, EPI_ISL_1968463, EPI_ISL_1968465, EPI_ISL_1968466, EPI_ISL_1968467, EPI_ISL_1968469, EPI_ISL_1968470, EPI_ISL_1968471, EPI_ISL_1968472, EPI_ISL_1968473, EPI_ISL_1968474, EPI_ISL_1968476, EPI_ISL_1968477, EPI_ISL_1968478, EPI_ISL_1968479, EPI_ISL_1968480, EPI_ISL_1971537, EPI_ISL_1971538,
EPI_ISL_1971539, EPI_ISL_1971540, EPI_ISL_1971542, EPI_ISL_1971543, EPI_ISL_1971544, EPI_ISL_1971545, EPI_ISL_1971546, EPI_ISL_1971547, EPI_ISL_1971548, EPI_ISL_1971549, EPI_ISL_1971550, EPI_ISL_1971551, EPI_ISL_1971552, EPI_ISL_1971553, EPI_ISL_1971554, EPI_ISL_1971555, EPI_ISL_1971556, EPI_ISL_1971558, EPI_ISL_1971559, EPI_ISL_1971560, EPI_ISL_1971561, EPI_ISL_1971563,
EPI_ISL_1971564, EPI_ISL_1971566, EPI_ISL_1971567, EPI_ISL_1971568, EPI_ISL_1971569, EPI_ISL_1971570, EPI_ISL_1971571, EPI_ISL_1971572, EPI_ISL_1971573, EPI_ISL_1971574, EPI_ISL_1971575, EPI_ISL_1971576, EPI_ISL_1971577, EPI_ISL_1971578, EPI_ISL_1971579, EPI_ISL_1971580, EPI_ISL_1971581, EPI_ISL_1971582, EPI_ISL_1971583, EPI_ISL_1971584, EPI_ISL_1971585, EPI_ISL_1971588,

EPI_ISL_1971590, EPI_ISL_1971591, EPI_ISL_1971594, EPI_ISL_1971595, EPI_ISL_1971596, EPI_ISL_1971597, EPI_ISL_1971598, EPI_ISL_1971599, EPI_ISL_1971600, EPI_ISL_1971601, EPI_ISL_1971602, EPI_ISL_1971603, EPI_ISL_1971604, EPI_ISL_1971605, EPI_ISL_1971606, EPI_ISL_1971607, EPI_ISL_1971608, EPI_ISL_1971609, EPI_ISL_1971610, EPI_ISL_1971611, EPI_ISL_1971612, EPI_ISL_1971613, EPI_ISL_1971614, EPI_ISL_1971615, EPI_ISL_1971616, EPI_ISL_1971619, EPI_ISL_1971620, EPI_ISL_1971621, EPI_ISL_1971622, EPI_ISL_1971623, EPI_ISL_1971624, EPI_ISL_1971627, EPI_ISL_1971628, EPI_ISL_1971629, EPI_ISL_1971630, EPI_ISL_1971631, EPI_ISL_1971632, EPI_ISL_1971633, EPI_ISL_1971634, EPI_ISL_1971635, EPI_ISL_1971636, EPI_ISL_1971637, EPI_ISL_1971638, EPI_ISL_1971639, EPI_ISL_1971641, EPI_ISL_1971642, EPI_ISL_1971643, EPI_ISL_1971645, EPI_ISL_1971646, EPI_ISL_1971647, EPI_ISL_1971648, EPI_ISL_1971649, EPI_ISL_1971650, EPI_ISL_1971651, EPI_ISL_1971653, EPI_ISL_1971654, EPI_ISL_1971655, EPI_ISL_1971656, EPI_ISL_1971657, EPI_ISL_1971658, EPI_ISL_1971659, EPI_ISL_1971660, EPI_ISL_1971661, EPI_ISL_1971662, EPI_ISL_1971664, EPI_ISL_1971665, EPI_ISL_1971666, EPI_ISL_1971667, EPI_ISL_1971668, EPI_ISL_1971669, EPI_ISL_1971670, EPI_ISL_1971671, EPI_ISL_1971672, EPI_ISL_1971673, EPI_ISL_1971674, EPI_ISL_1971675, EPI_ISL_1971676, EPI_ISL_1971677, EPI_ISL_1971678, EPI_ISL_1971679, EPI_ISL_1971680, EPI_ISL_1971682, EPI_ISL_1971683, EPI_ISL_1971684, EPI_ISL_1971686, EPI_ISL_1971687, EPI_ISL_1971688, EPI_ISL_1971689, EPI_ISL_1971692, EPI_ISL_1971693, EPI_ISL_1971694, EPI_ISL_1971695, EPI_ISL_1971696, EPI_ISL_1971699, EPI_ISL_1971700, EPI_ISL_1971701, EPI_ISL_1971702, EPI_ISL_1971704, EPI_ISL_1971705, EPI_ISL_1971706, EPI_ISL_1971707, EPI_ISL_1971708, EPI_ISL_1971709, EPI_ISL_1971710, EPI_ISL_1971711, EPI_ISL_1971712, EPI_ISL_1971713, EPI_ISL_1971714, EPI_ISL_1971715, EPI_ISL_1971716, EPI_ISL_1971717, EPI_ISL_1971718, EPI_ISL_1971719, EPI_ISL_1971720, EPI_ISL_1971721, EPI_ISL_1971722, EPI_ISL_1971724, EPI_ISL_1971725, EPI_ISL_1971726, EPI_ISL_1971727, EPI_ISL_1971728, EPI_ISL_1971729, EPI_ISL_1971730, EPI_ISL_1971731, EPI_ISL_1971732, EPI_ISL_1971733, EPI_ISL_1971734, EPI_ISL_1971735, EPI_ISL_1971736, EPI_ISL_1971737, EPI_ISL_1971738, EPI_ISL_1971739, EPI_ISL_1971740, EPI_ISL_1971741, EPI_ISL_1971742, EPI_ISL_1971743, EPI_ISL_1971744, EPI_ISL_1971745, EPI_ISL_1971746, EPI_ISL_1971747, EPI_ISL_1971748, EPI_ISL_1971749, EPI_ISL_1971750, EPI_ISL_1971751, EPI_ISL_1971752, EPI_ISL_1971753, EPI_ISL_1971754, EPI_ISL_1971755, EPI_ISL_1971756, EPI_ISL_1971757, EPI_ISL_1971758, EPI_ISL_1971759, EPI_ISL_1971760, EPI_ISL_1971761, EPI_ISL_1971762, EPI_ISL_1971763, EPI_ISL_1971764, EPI_ISL_1971765, EPI_ISL_1971766, EPI_ISL_1971767, EPI_ISL_1971768, EPI_ISL_1971769, EPI_ISL_1971770, EPI_ISL_1971771, EPI_ISL_1971772, EPI_ISL_1971775, EPI_ISL_1971776, EPI_ISL_1971778, EPI_ISL_1971779, EPI_ISL_1971780, EPI_ISL_1971781, EPI_ISL_1971782, EPI_ISL_1971783, EPI_ISL_1971784, EPI_ISL_1971786, EPI_ISL_1971787, EPI_ISL_1971788, EPI_ISL_1971789, EPI_ISL_1971790, EPI_ISL_1971791, EPI_ISL_1971792, EPI_ISL_1971794, EPI_ISL_1971795, EPI_ISL_1971796, EPI_ISL_1971797, EPI_ISL_1971798, EPI_ISL_1971799, EPI_ISL_1971800, EPI_ISL_1971801, EPI_ISL_1971802, EPI_ISL_1971803, EPI_ISL_1971804, EPI_ISL_1971805, EPI_ISL_1971806, EPI_ISL_1971807, EPI_ISL_1971808, EPI_ISL_1971809, EPI_ISL_1971811, EPI_ISL_1971812, EPI_ISL_1971813, EPI_ISL_1971814, EPI_ISL_1971815, EPI_ISL_1971816, EPI_ISL_1971817, EPI_ISL_1971818, EPI_ISL_1971819, EPI_ISL_1971820, EPI_ISL_1971821, EPI_ISL_1971822, EPI_ISL_1971823, EPI_ISL_1971824, EPI_ISL_1971825, EPI_ISL_1971827, EPI_ISL_1971828, EPI_ISL_1971829, EPI_ISL_1971830, EPI_ISL_1971831, EPI_ISL_1971832, EPI_ISL_1971833, EPI_ISL_1971834, EPI_ISL_1971835, EPI_ISL_1971836, EPI_ISL_1971837, EPI_ISL_1971838, EPI_ISL_1971839, EPI_ISL_1971841, EPI_ISL_1971842, EPI_ISL_1971843, EPI_ISL_1971844, EPI_ISL_1971845, EPI_ISL_1971846, EPI_ISL_1971847, EPI_ISL_1971848, EPI_ISL_1971849, EPI_ISL_1971850, EPI_ISL_1971851, EPI_ISL_1971852, EPI_ISL_1971853, EPI_ISL_1971854, EPI_ISL_1971855, EPI_ISL_1971857, EPI_ISL_1971858, EPI_ISL_1971859, EPI_ISL_1971860, EPI_ISL_1971861, EPI_ISL_1971862, EPI_ISL_1971864, EPI_ISL_1971865, EPI_ISL_1971866, EPI_ISL_1971867, EPI_ISL_1971869, EPI_ISL_1971870, EPI_ISL_1971872, EPI_ISL_1971873, EPI_ISL_1971874, EPI_ISL_1971875, EPI_ISL_1971876, EPI_ISL_1971877, EPI_ISL_1971878, EPI_ISL_1971879, EPI_ISL_1971880, EPI_ISL_1971882, EPI_ISL_1971883, EPI_ISL_1971884, EPI_ISL_1971885, EPI_ISL_1971886, EPI_ISL_1971887, EPI_ISL_1971888, EPI_ISL_1971889, EPI_ISL_1971890, EPI_ISL_1971891, EPI_ISL_1971892, EPI_ISL_1971893, EPI_ISL_1971894, EPI_ISL_1971895, EPI_ISL_1971896, EPI_ISL_1971897, EPI_ISL_1971898, EPI_ISL_1971899, EPI_ISL_1971900, EPI_ISL_1971901, EPI_ISL_1971903, EPI_ISL_1971904, EPI_ISL_1971905, EPI_ISL_1971906, EPI_ISL_1971907, EPI_ISL_1971908, EPI_ISL_1971909, EPI_ISL_1971910, EPI_ISL_1971911, EPI_ISL_1971913, EPI_ISL_1971914, EPI_ISL_1971915, EPI_ISL_1971916, EPI_ISL_1971917, EPI_ISL_1971918, EPI_ISL_1971919, EPI_ISL_1971920, EPI_ISL_1971921, EPI_ISL_1971922, EPI_ISL_1971923, EPI_ISL_1971924, EPI_ISL_1971925, EPI_ISL_1971926, EPI_ISL_1971927, EPI_ISL_1971928, EPI_ISL_1971929, EPI_ISL_1971930, EPI_ISL_1971931, EPI_ISL_1971933, EPI_ISL_1971934, EPI_ISL_1971935, EPI_ISL_1971936, EPI_ISL_1971937, EPI_ISL_1971938, EPI_ISL_1971939, EPI_ISL_1971940, EPI_ISL_1971941, EPI_ISL_1971942, EPI_ISL_1971943, EPI_ISL_1971944, EPI_ISL_1971946, EPI_ISL_1971948, EPI_ISL_1971949, EPI_ISL_1971950, EPI_ISL_1971951, EPI_ISL_1971953, EPI_ISL_1971954, EPI_ISL_1971956, EPI_ISL_1971957, EPI_ISL_1971958, EPI_ISL_1971960, EPI_ISL_1971962, EPI_ISL_1971963, EPI_ISL_1971965, EPI_ISL_1971966, EPI_ISL_1971967, EPI_ISL_1971969, EPI_ISL_1971970, EPI_ISL_1971971, EPI_ISL_1971972, EPI_ISL_1971973, EPI_ISL_1971974, EPI_ISL_1971976, EPI_ISL_1971977, EPI_ISL_1971978, EPI_ISL_1971979

| | | | |
|---|---|---|---|
| see above | Broad Institute Clinical Research Sequencing Platform | Infectious Disease Program, Broad Institute of Harvard and MIT | Adams, G.; B.L.; B.W.; Bauer, M.; Birren; Blumenstiel, B.; Brown, C.; Carter, A.; Chaluvadi, S.; D.J.; DeFelice, M.; DeRuff, K.; Dodge, S.; Gabriel, S.; Gallagher, G.; Gladden-Young, A.; Granger, B.; J.E.; K.J.; Lagerborg, K.; Larkin, K.; Lee, M.; Lemieux; Lennon, N.; Loreth, C.; MacInnis; Madoff, L.; McGovern, S.; Meldrim, J.; Normandin, E.; P.C.; Park; Pearlman, L.; Reilly, S.; Rudy, M.; Sabeti; Siddle; Smole, S.; Tomkins-Tinch, C.; Vicente, G.; and MacInnis; and Sabeti |

EPI_ISL_936464, EPI_ISL_936465, EPI_ISL_936466, EPI_ISL_936468, EPI_ISL_936471, EPI_ISL_936472, EPI_ISL_936473, EPI_ISL_936474, EPI_ISL_936477, EPI_ISL_936478, EPI_ISL_936481, EPI_ISL_936483, EPI_ISL_936485, EPI_ISL_1018064, EPI_ISL_1018065, EPI_ISL_1018066, EPI_ISL_1018067, EPI_ISL_1018068, EPI_ISL_1018069

| | | | |
|---|---|---|---|
| see above | DPH, Massachusetts State Public Health Lab | DPH, Massachusetts State Public Health Lab | A.S.; Fink, T.; G.R.; Gallagher; Lang; S.C.; Smole |

EPI_ISL_1460272, EPI_ISL_1555578, EPI_ISL_1556698, EPI_ISL_1556975, EPI_ISL_1613176, EPI_ISL_1614356, EPI_ISL_1667081, EPI_ISL_1733828, EPI_ISL_1734518

| | | | |
|---|---|---|---|
| see above | Fulgent Genetics | Centers for Disease Control and Prevention Division of Viral Diseases, Pathogen Discovery | Adrian Paskey; Becky Tsai; Benafsh Sapra; Benjamin Rambo-Martin; Christopher Gulvick; Clinton R. Paden; Dakota Howard; Darlene Wagner; Dhwani Batra; Doreen Ng; Duncan MacCannell; Harry Gao; James Xie; Jason Caravas; John Gao; Joseph Fierro; Kara Moser; Matthew Schmerer; Mickey Li; Peter W. Cook; Scott Sammons; Shatavia Morrison; Yan Meng; Yvette Unoarumhi |

EPI_ISL_850713, EPI_ISL_850716, EPI_ISL_850717, EPI_ISL_850725, EPI_ISL_850726, EPI_ISL_850727, EPI_ISL_850747, EPI_ISL_850840, EPI_ISL_850843, EPI_ISL_850848, EPI_ISL_850849, EPI_ISL_850850, EPI_ISL_850878, EPI_ISL_850880, EPI_ISL_850891, EPI_ISL_850892, EPI_ISL_850912

| | | | |
|---|---|---|---|
| see above | Helix / Illumina | Genomics and Discovery, Respiratory Viruses Branch, Division of Viral Diseases, Centers for Disease Control and Prevention | Alexandre Bolze; Ary Ascencio; Ben L. Rambo-Martin Eileen de Feo; Brad Sickler; Charlotte Rivera-Garcia; Christine Tran; Clinton R. Paden; David Becker; Dhwani Batra; Duncan MacCannell; Efren Sandoval; Elizabeth Cirulli; Eric Allen; Geraint Levan; James Lu; Jan Antico; Jason Nguyen; Jimmy Ramirez; Jingtao Liu; Kelly Schiabor Barrett; Kim Gietzen; Magnus Isaksson; Marc Laurent; Matthew Tolentino; Nicole L. Washington; Peter W. Cook; Phil Febbo; Ryan Cho; Shannon Wickline; Sherry Wang; Simon White; Summer Galloway; Suxiang Tong; Tyler Cassens; William Lee |

EPI_ISL_1088653, EPI_ISL_1088668, EPI_ISL_1088708, EPI_ISL_1088761, EPI_ISL_1088808, EPI_ISL_1088818, EPI_ISL_1088894, EPI_ISL_1088920, EPI_ISL_1088934, EPI_ISL_1088949, EPI_ISL_1088971, EPI_ISL_1089022, EPI_ISL_1089031, EPI_ISL_1089047, EPI_ISL_1089051, EPI_ISL_1089078, EPI_ISL_1089097, EPI_ISL_1089182, EPI_ISL_1089196, EPI_ISL_1089199, EPI_ISL_1089200, EPI_ISL_1089206, EPI_ISL_1089224, EPI_ISL_1089227, EPI_ISL_1089236, EPI_ISL_1089241, EPI_ISL_1255409, EPI_ISL_1255413

| | | | |
|---|---|---|---|
| see above | Helix / Illumina | Respiratory Viruses Branch, Division of Viral Diseases, Centers for Disease Control and Prevention | Alexandre Bolze; Ary Ascencio; Ben L. Rambo-Martin; Brad Sickler; Charlotte Rivera-Garcia; Christine Tran; Clinton R. Paden; Dakota Howard; David Becker; Dhwani Batra; Duncan MacCannell; Efren Sandoval; Eileen de Feo; Elizabeth Cirulli; Eric Allen; Geraint Levan; James Lu; Jan Antico; Jason Nguyen; Jimmy Ramirez; Jingtao Liu; Kelly Schiabor Barrett; Kim Gietzen; Magnus Isaksson; Marc Laurent; Matthew Tolentino; Nicole L. Washington; Peter W. Cook; Phil Febbo; Ryan Cho; Shannon Wickline; Sherry Wang; Simon White; Summer Galloway; Suxiang Tong; Tyler Cassens; William Lee |

EPI_ISL_1194739, EPI_ISL_1194767, EPI_ISL_1194815, EPI_ISL_1194821, EPI_ISL_1194861, EPI_ISL_1261898, EPI_ISL_1261902, EPI_ISL_1261909, EPI_ISL_1261913, EPI_ISL_1261916, EPI_ISL_1261924, EPI_ISL_1261926, EPI_ISL_1261927, EPI_ISL_1262038, EPI_ISL_1262119, EPI_ISL_1262131, EPI_ISL_1262132, EPI_ISL_1262136, EPI_ISL_1262148, EPI_ISL_1262156, EPI_ISL_1262181, EPI_ISL_1262182, EPI_ISL_1262189, EPI_ISL_1262209, EPI_ISL_1262216, EPI_ISL_1262221, EPI_ISL_1262223, EPI_ISL_1262237, EPI_ISL_1262241, EPI_ISL_1262254, EPI_ISL_1262259, EPI_ISL_1262264, EPI_ISL_1262323, EPI_ISL_1262335, EPI_ISL_1262356, EPI_ISL_1262411, EPI_ISL_1262418, EPI_ISL_1262420, EPI_ISL_1262430, EPI_ISL_1262447, EPI_ISL_1267514, EPI_ISL_1267517, EPI_ISL_1267518, EPI_ISL_1267528, EPI_ISL_1267617, EPI_ISL_1267641, EPI_ISL_1267647, EPI_ISL_1267651, EPI_ISL_1267654, EPI_ISL_1267673, EPI_ISL_1267679, EPI_ISL_1267685, EPI_ISL_1267687, EPI_ISL_1267688, EPI_ISL_1267698, EPI_ISL_1267716, EPI_ISL_1267731, EPI_ISL_1267732, EPI_ISL_1267757, EPI_ISL_1267769, EPI_ISL_1267773, EPI_ISL_1267778, EPI_ISL_1267807, EPI_ISL_1267860, EPI_ISL_1267969, EPI_ISL_1267977, EPI_ISL_1267985, EPI_ISL_1267986, EPI_ISL_1267994, EPI_ISL_1268010, EPI_ISL_1268011, EPI_ISL_1268070, EPI_ISL_1268074, EPI_ISL_1268079, EPI_ISL_1268084, EPI_ISL_1268097, EPI_ISL_1268104, EPI_ISL_1268108, EPI_ISL_1268120, EPI_ISL_1268179, EPI_ISL_1268184, EPI_ISL_1268192, EPI_ISL_1268206, EPI_ISL_1268208, EPI_ISL_1268256, EPI_ISL_1268258, EPI_ISL_1268268, EPI_ISL_1268274, EPI_ISL_1268281, EPI_ISL_1268302, EPI_ISL_1268307, EPI_ISL_1268316, EPI_ISL_1268347, EPI_ISL_1268394, EPI_ISL_1268396, EPI_ISL_1268408, EPI_ISL_1268409, EPI_ISL_1268412, EPI_ISL_1268420, EPI_ISL_1268431, EPI_ISL_1268448, EPI_ISL_1268499, EPI_ISL_1268506, EPI_ISL_1268532, EPI_ISL_1268550, EPI_ISL_1268586, EPI_ISL_1268617, EPI_ISL_1268618, EPI_ISL_1268693, EPI_ISL_1268699, EPI_ISL_1268827, EPI_ISL_1268938, EPI_ISL_1268942, EPI_ISL_1269004, EPI_ISL_1269024, EPI_ISL_1269096, EPI_ISL_1269247, EPI_ISL_1269281, EPI_ISL_1269282, EPI_ISL_1269288, EPI_ISL_1269293, EPI_ISL_1269329, EPI_ISL_1269704, EPI_ISL_1269749, EPI_ISL_1269817, EPI_ISL_1270052, EPI_ISL_1270061, EPI_ISL_1270062, EPI_ISL_1270066, EPI_ISL_1270067, EPI_ISL_1270070, EPI_ISL_1270080, EPI_ISL_1270081, EPI_ISL_1270088, EPI_ISL_1270089, EPI_ISL_1270103, EPI_ISL_1270105, EPI_ISL_1270117, EPI_ISL_1270127, EPI_ISL_1270188, EPI_ISL_1270221, EPI_ISL_1270240, EPI_ISL_1270257, EPI_ISL_1270298, EPI_ISL_1270305, EPI_ISL_1270313, EPI_ISL_1270316, EPI_ISL_1270319, EPI_ISL_1270321, EPI_ISL_1270343, EPI_ISL_1270365, EPI_ISL_1270368, EPI_ISL_1270381, EPI_ISL_1270410, EPI_ISL_1270415, EPI_ISL_1270423, EPI_ISL_1270427, EPI_ISL_1270435, EPI_ISL_1270528, EPI_ISL_1270583, EPI_ISL_1270595, EPI_ISL_1270613, EPI_ISL_1270614, EPI_ISL_1270635, EPI_ISL_1270642, EPI_ISL_1270643, EPI_ISL_1270721, EPI_ISL_1270734, EPI_ISL_1270739, EPI_ISL_1270745, EPI_ISL_1270753, EPI_ISL_1270758, EPI_ISL_1270760, EPI_ISL_1270765, EPI_ISL_1270766, EPI_ISL_1270774, EPI_ISL_1270775, EPI_ISL_1270782, EPI_ISL_1270785, EPI_ISL_1283389, EPI_ISL_1283425, EPI_ISL_1283433, EPI_ISL_1283440, EPI_ISL_1283468, EPI_ISL_1283587, EPI_ISL_1283595, EPI_ISL_1283617, EPI_ISL_1283620, EPI_ISL_1283634, EPI_ISL_1283636, EPI_ISL_1283647, EPI_ISL_1283650, EPI_ISL_1283652, EPI_ISL_1283655, EPI_ISL_1283754, EPI_ISL_1283767, EPI_ISL_1283783, EPI_ISL_1283808, EPI_ISL_1284023, EPI_ISL_1284060, EPI_ISL_1284278, EPI_ISL_1284326, EPI_ISL_1284389, EPI_ISL_1284409, EPI_ISL_1284456, EPI_ISL_1297857, EPI_ISL_1297872, EPI_ISL_1297881, EPI_ISL_1297885, EPI_ISL_1297890, EPI_ISL_1297897, EPI_ISL_1297902, EPI_ISL_1297903, EPI_ISL_1297908, EPI_ISL_1297949, EPI_ISL_1298018, EPI_ISL_1298104, EPI_ISL_1298109, EPI_ISL_1298121, EPI_ISL_1298124, EPI_ISL_1298127, EPI_ISL_1298141, EPI_ISL_1298142, EPI_ISL_1298153, EPI_ISL_1298158, EPI_ISL_1336968, EPI_ISL_1336970, EPI_ISL_1336983, EPI_ISL_1336991, EPI_ISL_1336997, EPI_ISL_1337028, EPI_ISL_1337034, EPI_ISL_1337063, EPI_ISL_1337075, EPI_ISL_1337078, EPI_ISL_1337093, EPI_ISL_1337094, EPI_ISL_1337102, EPI_ISL_1337113, EPI_ISL_1337121, EPI_ISL_1337184, EPI_ISL_1337293, EPI_ISL_1337333, EPI_ISL_1337388, EPI_ISL_1337400, EPI_ISL_1338019, EPI_ISL_1338030, EPI_ISL_1338038, EPI_ISL_1338046, EPI_ISL_1338049, EPI_ISL_1338060, EPI_ISL_1338062, EPI_ISL_1338063, EPI_ISL_1338065, EPI_ISL_1338069, EPI_ISL_1338075, EPI_ISL_1338081, EPI_ISL_1338102, EPI_ISL_1338150, EPI_ISL_1338158, EPI_ISL_1338200, EPI_ISL_1338219, EPI_ISL_1338227, EPI_ISL_1338252, EPI_ISL_1338267, EPI_ISL_1338268, EPI_ISL_1338310, EPI_ISL_1338374, EPI_ISL_1338376, EPI_ISL_1338387, EPI_ISL_1340079, EPI_ISL_1340114, EPI_ISL_1340165, EPI_ISL_1340170, EPI_ISL_1340197, EPI_ISL_1340255, EPI_ISL_1340266, EPI_ISL_1340338, EPI_ISL_1340340, EPI_ISL_1340342, EPI_ISL_1340344, EPI_ISL_1340346, EPI_ISL_1340350, EPI_ISL_1340352, EPI_ISL_1340357, EPI_ISL_1340358, EPI_ISL_1340360, EPI_ISL_1340399, EPI_ISL_1340401, EPI_ISL_1340405, EPI_ISL_1340413, EPI_ISL_1340414, EPI_ISL_1340422, EPI_ISL_1340430, EPI_ISL_1340433, EPI_ISL_1340434, EPI_ISL_1340435, EPI_ISL_1340438, EPI_ISL_1340439, EPI_ISL_1340446, EPI_ISL_1340449, EPI_ISL_1391709, EPI_ISL_1391710, EPI_ISL_1391713, EPI_ISL_1391718, EPI_ISL_1391719, EPI_ISL_1391793, EPI_ISL_1391802, EPI_ISL_1391826, EPI_ISL_1391833, EPI_ISL_1391837, EPI_ISL_1391838, EPI_ISL_1391850, EPI_ISL_1391855, EPI_ISL_1391857, EPI_ISL_1391913, EPI_ISL_1391928, EPI_ISL_1391938, EPI_ISL_1391944, EPI_ISL_1391948, EPI_ISL_1391953, EPI_ISL_1391963, EPI_ISL_1420879, EPI_ISL_1420891, EPI_ISL_1420893, EPI_ISL_1420913, EPI_ISL_1420963, EPI_ISL_1420965, EPI_ISL_1420967, EPI_ISL_1421017, EPI_ISL_1421054, EPI_ISL_1421161, EPI_ISL_1421163, EPI_ISL_1421172, EPI_ISL_1421182, EPI_ISL_1421309, EPI_ISL_1421339, EPI_ISL_1421356, EPI_ISL_1421358, EPI_ISL_1421364, EPI_ISL_1421367, EPI_ISL_1421449, EPI_ISL_1421521, EPI_ISL_1421551, EPI_ISL_1421623, EPI_ISL_1421691, EPI_ISL_1421728, EPI_ISL_1421740, EPI_ISL_1443822, EPI_ISL_1443840, EPI_ISL_1443844, EPI_ISL_1443864, EPI_ISL_1443920, EPI_ISL_1443923, EPI_ISL_1443967, EPI_ISL_1443986, EPI_ISL_1444032, EPI_ISL_1444037, EPI_ISL_1444039, EPI_ISL_1444108, EPI_ISL_1444145, EPI_ISL_1444167, EPI_ISL_1444171, EPI_ISL_1444174, EPI_ISL_1444177, EPI_ISL_1444186, EPI_ISL_1444187, EPI_ISL_1444191, EPI_ISL_1444195, EPI_ISL_1444198, EPI_ISL_1444207, EPI_ISL_1444208, EPI_ISL_1444215, EPI_ISL_1444220, EPI_ISL_1444221, EPI_ISL_1444222, EPI_ISL_1444226, EPI_ISL_1444228, EPI_ISL_1444230, EPI_ISL_1444236, EPI_ISL_1444254, EPI_ISL_1444257, EPI_ISL_1460834, EPI_ISL_1460837, EPI_ISL_1460839, EPI_ISL_1460841, EPI_ISL_1460848, EPI_ISL_1460859, EPI_ISL_1460861, EPI_ISL_1460869, EPI_ISL_1460872, EPI_ISL_1460898, EPI_ISL_1460957, EPI_ISL_1479245, EPI_ISL_1479261, EPI_ISL_1479262, EPI_ISL_1479263, EPI_ISL_1479281, EPI_ISL_1479287, EPI_ISL_1479288, EPI_ISL_1479297, EPI_ISL_1479306, EPI_ISL_1479307, EPI_ISL_1479312, EPI_ISL_1479318, EPI_ISL_1479356, EPI_ISL_1479357, EPI_ISL_1479360, EPI_ISL_1479363, EPI_ISL_1479364, EPI_ISL_1479373, EPI_ISL_1479376, EPI_ISL_1479385, EPI_ISL_1479388, EPI_ISL_1479392, EPI_ISL_1479394, EPI_ISL_1479397, EPI_ISL_1479398, EPI_ISL_1479407, EPI_ISL_1479408, EPI_ISL_1479409, EPI_ISL_1480020, EPI_ISL_1480139, EPI_ISL_1480143, EPI_ISL_1480147, EPI_ISL_1480185, EPI_ISL_1480193, EPI_ISL_1480195, EPI_ISL_1480198, EPI_ISL_1480204, EPI_ISL_1480211, EPI_ISL_1480224, EPI_ISL_1480227, EPI_ISL_1480280, EPI_ISL_1480284, EPI_ISL_1480285, EPI_ISL_1480331, EPI_ISL_1480358, EPI_ISL_1480378, EPI_ISL_1480395, EPI_ISL_1480407, EPI_ISL_1480410, EPI_ISL_1480420, EPI_ISL_1480437, EPI_ISL_1480477, EPI_ISL_1480478, EPI_ISL_1480491, EPI_ISL_1480515, EPI_ISL_1480520, EPI_ISL_1493725, EPI_ISL_1493727, EPI_ISL_1493795, EPI_ISL_1493801, EPI_ISL_1493808, EPI_ISL_1493836, EPI_ISL_1493922, EPI_ISL_1493924, EPI_ISL_1493929, EPI_ISL_1511893, EPI_ISL_1511898, EPI_ISL_1511911, EPI_ISL_1511927, EPI_ISL_1511934, EPI_ISL_1511956, EPI_ISL_1511960, EPI_ISL_1511966, EPI_ISL_1512015, EPI_ISL_1512030, EPI_ISL_1512033, EPI_ISL_1512039, EPI_ISL_1512043, EPI_ISL_1512114, EPI_ISL_1512128, EPI_ISL_1512136, EPI_ISL_1512138, EPI_ISL_1512141, EPI_ISL_1512142, EPI_ISL_1512145, EPI_ISL_1512153, EPI_ISL_1512158, EPI_ISL_1512159, EPI_ISL_1512185, EPI_ISL_1512192, EPI_ISL_1512206, EPI_ISL_1512210, EPI_ISL_1512217, EPI_ISL_1512221, EPI_ISL_1512237, EPI_ISL_1512244, EPI_ISL_1512304, EPI_ISL_1512309, EPI_ISL_1512332, EPI_ISL_1512334, EPI_ISL_1512336, EPI_ISL_1512382, EPI_ISL_1512447, EPI_ISL_1512486, EPI_ISL_1512534, EPI_ISL_1512543, EPI_ISL_1512549, EPI_ISL_1512551, EPI_ISL_1512553, EPI_ISL_1512560, EPI_ISL_1512577, EPI_ISL_1512595, EPI_ISL_1512668, EPI_ISL_1512676, EPI_ISL_1512677, EPI_ISL_1512684, EPI_ISL_1512693, EPI_ISL_1512696, EPI_ISL_1512697, EPI_ISL_1512724, EPI_ISL_1553289, EPI_ISL_1553314, EPI_ISL_1553315, EPI_ISL_1553317, EPI_ISL_1553319, EPI_ISL_1553321, EPI_ISL_1553326, EPI_ISL_1553329, EPI_ISL_1553333, EPI_ISL_1553347, EPI_ISL_1553353, EPI_ISL_1553357, EPI_ISL_1553399, EPI_ISL_1553419, EPI_ISL_1553423, EPI_ISL_1553430, EPI_ISL_1553438, EPI_ISL_1553483, EPI_ISL_1553547, EPI_ISL_1553552, EPI_ISL_1553556, EPI_ISL_1553576, EPI_ISL_1553577, EPI_ISL_1553582, EPI_ISL_1553606, EPI_ISL_1553612, EPI_ISL_1553627, EPI_ISL_1553628, EPI_ISL_1553637, EPI_ISL_1553640, EPI_ISL_1553641, EPI_ISL_1553643, EPI_ISL_1553686, EPI_ISL_1553699, EPI_ISL_1553711, EPI_ISL_1553727, EPI_ISL_1553734, EPI_ISL_1553746, EPI_ISL_1553748, EPI_ISL_1553853, EPI_ISL_1553898, EPI_ISL_1553899, EPI_ISL_1553916, EPI_ISL_1553933, EPI_ISL_1553948, EPI_ISL_1553954, EPI_ISL_1553955, EPI_ISL_1553959, EPI_ISL_1553963, EPI_ISL_1553972, EPI_ISL_1553978, EPI_ISL_1554010, EPI_ISL_1554059, EPI_ISL_1554062, EPI_ISL_1554063, EPI_ISL_1554069, EPI_ISL_1554082, EPI_ISL_1554095, EPI_ISL_1554098, EPI_ISL_1554108, EPI_ISL_1554112, EPI_ISL_1554113, EPI_ISL_1554115, EPI_ISL_1554120, EPI_ISL_1554127, EPI_ISL_1554129, EPI_ISL_1554134, EPI_ISL_1554145, EPI_ISL_1554148, EPI_ISL_1554156, EPI_ISL_1554157, EPI_ISL_1554162, EPI_ISL_1554165, EPI_ISL_1554167, EPI_ISL_1554168, EPI_ISL_1554231, EPI_ISL_1554237, EPI_ISL_1554244, EPI_ISL_1554265, EPI_ISL_1554275, EPI_ISL_1554290, EPI_ISL_1554321, EPI_ISL_1554322, EPI_ISL_1554323, EPI_ISL_1554332, EPI_ISL_1554340, EPI_ISL_1554341, EPI_ISL_1554356, EPI_ISL_1554378, EPI_ISL_1554394, EPI_ISL_1554396, EPI_ISL_1554399, EPI_ISL_1554411, EPI_ISL_1554432, EPI_ISL_1554438, EPI_ISL_1554451, EPI_ISL_1554457, EPI_ISL_1554484, EPI_ISL_1554518, EPI_ISL_1554521, EPI_ISL_1554522, EPI_ISL_1554531, EPI_ISL_1554539, EPI_ISL_1554540, EPI_ISL_1554551, EPI_ISL_1554552, EPI_ISL_1554569, EPI_ISL_1554596, EPI_ISL_1554601, EPI_ISL_1554602, EPI_ISL_1554611, EPI_ISL_1554625, EPI_ISL_1554711, EPI_ISL_1554737, EPI_ISL_1554740, EPI_ISL_1554751, EPI_ISL_1554753, EPI_ISL_1554757, EPI_ISL_1554758, EPI_ISL_1554779, EPI_ISL_1554785, EPI_ISL_1554811, EPI_ISL_1554821, EPI_ISL_1554825, EPI_ISL_1554881, EPI_ISL_1575485, EPI_ISL_1575495, EPI_ISL_1575505, EPI_ISL_1575519, EPI_ISL_1575555, EPI_ISL_1575626, EPI_ISL_1575659, EPI_ISL_1575682, EPI_ISL_1575685, EPI_ISL_1575688, EPI_ISL_1575706, EPI_ISL_1575729, EPI_ISL_1575735, EPI_ISL_1575738, EPI_ISL_1575745, EPI_ISL_1575751, EPI_ISL_1575764, EPI_ISL_1575765, EPI_ISL_1575784, EPI_ISL_1575785, EPI_ISL_1575789, EPI_ISL_1575797, EPI_ISL_1575798, EPI_ISL_1575801, EPI_ISL_1575860, EPI_ISL_1575914, EPI_ISL_1575921, EPI_ISL_1575929, EPI_ISL_1575934, EPI_ISL_1575939, EPI_ISL_1575940, EPI_ISL_1575946, EPI_ISL_1575958, EPI_ISL_1575989, EPI_ISL_1575991, EPI_ISL_1576005, EPI_ISL_1576009, EPI_ISL_1576011, EPI_ISL_1576024, EPI_ISL_1576054, EPI_ISL_1576075, EPI_ISL_1576079, EPI_ISL_1576097, EPI_ISL_1576108, EPI_ISL_1576128, EPI_ISL_1576139, EPI_ISL_1576150, EPI_ISL_1576152, EPI_ISL_1576157, EPI_ISL_1576162, EPI_ISL_1576165, EPI_ISL_1576166, EPI_ISL_1576167, EPI_ISL_1576176, EPI_ISL_1576178, EPI_ISL_1576179, EPI_ISL_1576184, EPI_ISL_1576190, EPI_ISL_1576195, EPI_ISL_1576202, EPI_ISL_1576211, EPI_ISL_1576216, EPI_ISL_1576239, EPI_ISL_1576253, EPI_ISL_1576405, EPI_ISL_1576412, EPI_ISL_1576416, EPI_ISL_1576436, EPI_ISL_1576444, EPI_ISL_1576450, EPI_ISL_1576455, EPI_ISL_1576462, EPI_ISL_1576463, EPI_ISL_1576473, EPI_ISL_1576491, EPI_ISL_1576496, EPI_ISL_1576503, EPI_ISL_1576510, EPI_ISL_1576512, EPI_ISL_1576519, EPI_ISL_1576524, EPI_ISL_1576527, EPI_ISL_1576530, EPI_ISL_1576531, EPI_ISL_1576533, EPI_ISL_1576536, EPI_ISL_1576539, EPI_ISL_1576542, EPI_ISL_1576551, EPI_ISL_1576552, EPI_ISL_1576557, EPI_ISL_1576560, EPI_ISL_1576563, EPI_ISL_1576575, EPI_ISL_1576604, EPI_ISL_1576618, EPI_ISL_1576626, EPI_ISL_1576628, EPI_ISL_1576731, EPI_ISL_1580754, EPI_ISL_1580791, EPI_ISL_1580802, EPI_ISL_1580803, EPI_ISL_1580805, EPI_ISL_1580806, EPI_ISL_1580819, EPI_ISL_1580823, EPI_ISL_1580825, EPI_ISL_1580837, EPI_ISL_1580847, EPI_ISL_1580850, EPI_ISL_1580851, EPI_ISL_1580859, EPI_ISL_1580874, EPI_ISL_1580940, EPI_ISL_1580952, EPI_ISL_1580966, EPI_ISL_1580968, EPI_ISL_1580978, EPI_ISL_1581002, EPI_ISL_1581018, EPI_ISL_1581023, EPI_ISL_1581039, EPI_ISL_1581061, EPI_ISL_1581103, EPI_ISL_1581133, EPI_ISL_1581237, EPI_ISL_1581249, EPI_ISL_1581273, EPI_ISL_1581285, EPI_ISL_1581287, EPI_ISL_1581304, EPI_ISL_1581305, EPI_ISL_1581317, EPI_ISL_1581320, EPI_ISL_1581323, EPI_ISL_1581360, EPI_ISL_1581370, EPI_ISL_1581384, EPI_ISL_1581430, EPI_ISL_1581435, EPI_ISL_1581537, EPI_ISL_1581567, EPI_ISL_1581586, EPI_ISL_1581605, EPI_ISL_1581638, EPI_ISL_1581641, EPI_ISL_1581652, EPI_ISL_1581760, EPI_ISL_1581776, EPI_ISL_1581778, EPI_ISL_1581791, EPI_ISL_1581799, EPI_ISL_1581803, EPI_ISL_1581805, EPI_ISL_1581824, EPI_ISL_1581827, EPI_ISL_1581842, EPI_ISL_1592410, EPI_ISL_1592421, EPI_ISL_1592442, EPI_ISL_1592453, EPI_ISL_1592454, EPI_ISL_1592482, EPI_ISL_1592485, EPI_ISL_1592486, EPI_ISL_1592503, EPI_ISL_1592509, EPI_ISL_1592513, EPI_ISL_1592515, EPI_ISL_1592532, EPI_ISL_1592539, EPI_ISL_1592540, EPI_ISL_1592556, EPI_ISL_1592568, EPI_ISL_1592586, EPI_ISL_1592587, EPI_ISL_1592702, EPI_ISL_1592703, EPI_ISL_1592715, EPI_ISL_1592725, EPI_ISL_1592737, EPI_ISL_1592771, EPI_ISL_1592776, EPI_ISL_1592777, EPI_ISL_1592803, EPI_ISL_1592836, EPI_ISL_1592850, EPI_ISL_1592867, EPI_ISL_1592894, EPI_ISL_1592905, EPI_ISL_1592918, EPI_ISL_1592919, EPI_ISL_1592920, EPI_ISL_1592927, EPI_ISL_1592936, EPI_ISL_1592939, EPI_ISL_1592951, EPI_ISL_1593082, EPI_ISL_1593128, EPI_ISL_1593165, EPI_ISL_1593168, EPI_ISL_1593172, EPI_ISL_1593176, EPI_ISL_1593184, EPI_ISL_1593192, EPI_ISL_1593195, EPI_ISL_1593200, EPI_ISL_1615163, EPI_ISL_1615165, EPI_ISL_1615186, EPI_ISL_1615308, EPI_ISL_1615316, EPI_ISL_1615318, EPI_ISL_1615324, EPI_ISL_1615327, EPI_ISL_1615328, EPI_ISL_1615338, EPI_ISL_1615339, EPI_ISL_1615345, EPI_ISL_1615350, EPI_ISL_1615359, EPI_ISL_1615360, EPI_ISL_1615365, EPI_ISL_1615371, EPI_ISL_1615394, EPI_ISL_1615469, EPI_ISL_1615484, EPI_ISL_1615489, EPI_ISL_1615494, EPI_ISL_1615508, EPI_ISL_1615511, EPI_ISL_1615523, EPI_ISL_1615524, EPI_ISL_1615532, EPI_ISL_1615539, EPI_ISL_1615548, EPI_ISL_1678891, EPI_ISL_1678956, EPI_ISL_1678957, EPI_ISL_1678960, EPI_ISL_1678967, EPI_ISL_1678971, EPI_ISL_1678979, EPI_ISL_1678988, EPI_ISL_1679007, EPI_ISL_1679010, EPI_ISL_1679011, EPI_ISL_1679017, EPI_ISL_1679026, EPI_ISL_1679028, EPI_ISL_1679089, EPI_ISL_1679104, EPI_ISL_1679113, EPI_ISL_1679171, EPI_ISL_1679184, EPI_ISL_1679381, EPI_ISL_1679518, EPI_ISL_1679528, EPI_ISL_1679538, EPI_ISL_1679540, EPI_ISL_1679544, EPI_ISL_1679571, EPI_ISL_1679579, EPI_ISL_1679600, EPI_ISL_1679611, EPI_ISL_1679615, EPI_ISL_1679652, EPI_ISL_1679657, EPI_ISL_1679661, EPI_ISL_1679664, EPI_ISL_1679690, EPI_ISL_1679692, EPI_ISL_1679695, EPI_ISL_1679696, EPI_ISL_1679704, EPI_ISL_1679742, EPI_ISL_1679768, EPI_ISL_1679772, EPI_ISL_1679789, EPI_ISL_1679807, EPI_ISL_1679865, EPI_ISL_1679894, EPI_ISL_1679919, EPI_ISL_1679942, EPI_ISL_1680005, EPI_ISL_1680105, EPI_ISL_1680121, EPI_ISL_1680139, EPI_ISL_1680153, EPI_ISL_1690281, EPI_ISL_1690317, EPI_ISL_1690322, EPI_ISL_1690332, EPI_ISL_1690345, EPI_ISL_1690357, EPI_ISL_1690359, EPI_ISL_1690657, EPI_ISL_1690748, EPI_ISL_1690815, EPI_ISL_1690945, EPI_ISL_1690963, EPI_ISL_1690992, EPI_ISL_1691000, EPI_ISL_1691002, EPI_ISL_1691011, EPI_ISL_1691012, EPI_ISL_1691030, EPI_ISL_1691037, EPI_ISL_1691059, EPI_ISL_1691091, EPI_ISL_1691125, EPI_ISL_1691204, EPI_ISL_1691233, EPI_ISL_1691238, EPI_ISL_1691239, EPI_ISL_1691265, EPI_ISL_1691276, EPI_ISL_1691277, EPI_ISL_1691281, EPI_ISL_1691290, EPI_ISL_1691353, EPI_ISL_1691367, EPI_ISL_1691375, EPI_ISL_1691389, EPI_ISL_1694649, EPI_ISL_1694658, EPI_ISL_1694660, EPI_ISL_1694681, EPI_ISL_1701837, EPI_ISL_1702145, EPI_ISL_1702161, EPI_ISL_1734579, EPI_ISL_1734603, EPI_ISL_1734630, EPI_ISL_1734632, EPI_ISL_1734656, EPI_ISL_1734676, EPI_ISL_1734697, EPI_ISL_1734717, EPI_ISL_1734718, EPI_ISL_1734824, EPI_ISL_1734828, EPI_ISL_1734856, EPI_ISL_1734932, EPI_ISL_1734947, EPI_ISL_1734971, EPI_ISL_1734983, EPI_ISL_1734990, EPI_ISL_1734993, EPI_ISL_1734994, EPI_ISL_1735049, EPI_ISL_1735076, EPI_ISL_1735085, EPI_ISL_1735128, EPI_ISL_1735154, EPI_ISL_1735168, EPI_ISL_1735176, EPI_ISL_1735182, EPI_ISL_1735185, EPI_ISL_1735193, EPI_ISL_1735194, EPI_ISL_1735210, EPI_ISL_1735216, EPI_ISL_1735219, EPI_ISL_1735231, EPI_ISL_1735236, EPI_ISL_1735237, EPI_ISL_1735256, EPI_ISL_1735257, EPI_ISL_1735260, EPI_ISL_1735263, EPI_ISL_1735267, EPI_ISL_1735278, EPI_ISL_1735294, EPI_ISL_1735309, EPI_ISL_1735439, EPI_ISL_1735448, EPI_ISL_1735449, EPI_ISL_1735450, EPI_ISL_1735471, EPI_ISL_1735492, EPI_ISL_1735493, EPI_ISL_1735504, EPI_ISL_1735559, EPI_ISL_1735574, EPI_ISL_1735583, EPI_ISL_1735595, EPI_ISL_1735599, EPI_ISL_1735606, EPI_ISL_1735624, EPI_ISL_1735626, EPI_ISL_1735634, EPI_ISL_1735636, EPI_ISL_1735639, EPI_ISL_1735642, EPI_ISL_1735658, EPI_ISL_1735676, EPI_ISL_1753511, EPI_ISL_1796142, EPI_ISL_1796153, EPI_ISL_1796158, EPI_ISL_1796195, EPI_ISL_1796206, EPI_ISL_1796207, EPI_ISL_1796227, EPI_ISL_1796246, EPI_ISL_1796247, EPI_ISL_1796265, EPI_ISL_1796320, EPI_ISL_1796340, EPI_ISL_1796360, EPI_ISL_1796368, EPI_ISL_1796377, EPI_ISL_1796384, EPI_ISL_1796402, EPI_ISL_1796422, EPI_ISL_1796427, EPI_ISL_1796434, EPI_ISL_1796445, EPI_ISL_1796451, EPI_ISL_1796452, EPI_ISL_1796456, EPI_ISL_1796512, EPI_ISL_1796533, EPI_ISL_1796536, EPI_ISL_1796545, EPI_ISL_1796547, EPI_ISL_1796579, EPI_ISL_1796593, EPI_ISL_1796606, EPI_ISL_1796616, EPI_ISL_1796633, EPI_ISL_1796662, EPI_ISL_1796663, EPI_ISL_1796670, EPI_ISL_1796671, EPI_ISL_1796713, EPI_ISL_1796714, EPI_ISL_1796724, EPI_ISL_1796734, EPI_ISL_1796735, EPI_ISL_1796756, EPI_ISL_1796771, EPI_ISL_1796786, EPI_ISL_1796787, EPI_ISL_1796791, EPI_ISL_1796799, EPI_ISL_1796805, EPI_ISL_1796819, EPI_ISL_1796820, EPI_ISL_1796821, EPI_ISL_1796822, EPI_ISL_1796831, EPI_ISL_1796851, EPI_ISL_1796901, EPI_ISL_1796906, EPI_ISL_1796931, EPI_ISL_1796942, EPI_ISL_1796944, EPI_ISL_1796957, EPI_ISL_1796961, EPI_ISL_1796971, EPI_ISL_1796989, EPI_ISL_1797092, EPI_ISL_1797103, EPI_ISL_1797110, EPI_ISL_1797127, EPI_ISL_1797131, EPI_ISL_1797287, EPI_ISL_1797298, EPI_ISL_1797376, EPI_ISL_1797381, EPI_ISL_1797399, EPI_ISL_1797406, EPI_ISL_1797409, EPI_ISL_1797410, EPI_ISL_1797411, EPI_ISL_1797420, EPI_ISL_1797433, EPI_ISL_1803505, EPI_ISL_1803537, EPI_ISL_1803539, EPI_ISL_1803558, EPI_ISL_1803570, EPI_ISL_1803578, EPI_ISL_1803581, EPI_ISL_1803603, EPI_ISL_1803612, EPI_ISL_1803624, EPI_ISL_1803665, EPI_ISL_1803706, EPI_ISL_1803707, EPI_ISL_1803712, EPI_ISL_1803713, EPI_ISL_1803720, EPI_ISL_1803732, EPI_ISL_1803761, EPI_ISL_1803771, EPI_ISL_1803774, EPI_ISL_1803776, EPI_ISL_1803777, EPI_ISL_1803781, EPI_ISL_1803784, EPI_ISL_1803803, EPI_ISL_1803856, EPI_ISL_1804032, EPI_ISL_1804050, EPI_ISL_1804052, EPI_ISL_1804067, EPI_ISL_1804084, EPI_ISL_1804234, EPI_ISL_1804299, EPI_ISL_1804358, EPI_ISL_1804378, EPI_ISL_1804412, EPI_ISL_1804413, EPI_ISL_1804414, EPI_ISL_1804435, EPI_ISL_1804454, EPI_ISL_1804464, EPI_ISL_1804465, EPI_ISL_1804467, EPI_ISL_1804472, EPI_ISL_1804477, EPI_ISL_1804478, EPI_ISL_1804487, EPI_ISL_1804488, EPI_ISL_1804490, EPI_ISL_1804511, EPI_ISL_1804546, EPI_ISL_1804580, EPI_ISL_1804587, EPI_ISL_1804594, EPI_ISL_1804607, EPI_ISL_1804615, EPI_ISL_1804617, EPI_ISL_1804620, EPI_ISL_1804636, EPI_ISL_1804660, EPI_ISL_1818007, EPI_ISL_1818014, EPI_ISL_1818024, EPI_ISL_1818040, EPI_ISL_1818041, EPI_ISL_1818044, EPI_ISL_1906895, EPI_ISL_1907002, EPI_ISL_1907036, EPI_ISL_1907064, EPI_ISL_1907119, EPI_ISL_1907134, EPI_ISL_1907299, EPI_ISL_1907311, EPI_ISL_1907330, EPI_ISL_1907331, EPI_ISL_1907342, EPI_ISL_1907364, EPI_ISL_1907373, EPI_ISL_1907386, EPI_ISL_1907406, EPI_ISL_1907415, EPI_ISL_1907486, EPI_ISL_1907487, EPI_ISL_1907533, EPI_ISL_1907540, EPI_ISL_1907543, EPI_ISL_1907558, EPI_ISL_1907570, EPI_ISL_1907581, EPI_ISL_1907582, EPI_ISL_1907583, EPI_ISL_1907591, EPI_ISL_1907593, EPI_ISL_1907625, EPI_ISL_1907627, EPI_ISL_1907638, EPI_ISL_1907670, EPI_ISL_1907682, EPI_ISL_1907721, EPI_ISL_1907764, EPI_ISL_1907775, EPI_ISL_1907829, EPI_ISL_1907839, EPI_ISL_1907973, EPI_ISL_1908043, EPI_ISL_1908053, EPI_ISL_1908064, EPI_ISL_1908067, EPI_ISL_1908074, EPI_ISL_1908092, EPI_ISL_1908104, EPI_ISL_1908115, EPI_ISL_1908137, EPI_ISL_1908139, EPI_ISL_1923253, EPI_ISL_1923331, EPI_ISL_1923354, EPI_ISL_1923372, EPI_ISL_1923373, EPI_ISL_1923375, EPI_ISL_1923382, EPI_ISL_1923384, EPI_ISL_1923390, EPI_ISL_1923397, EPI_ISL_1923401, EPI_ISL_1923405, EPI_ISL_1923432, EPI_ISL_1923501, EPI_ISL_1923502, EPI_ISL_1923577, EPI_ISL_1923635, EPI_ISL_1923648, EPI_ISL_1923666, EPI_ISL_1923676, EPI_ISL_1923685, EPI_ISL_1923706, EPI_ISL_1923713, EPI_ISL_1923714, EPI_ISL_1923718, EPI_ISL_1923726, EPI_ISL_1923738, EPI_ISL_1923744, EPI_ISL_1923747, EPI_ISL_1923759, EPI_ISL_1923762, EPI_ISL_1923777, EPI_ISL_1923788, EPI_ISL_1923811, EPI_ISL_1923835, EPI_ISL_1923840, EPI_ISL_1923862, EPI_ISL_1923877, EPI_ISL_1923885, EPI_ISL_1923886, EPI_ISL_1923895, EPI_ISL_1923898, EPI_ISL_1924002, EPI_ISL_1924041, EPI_ISL_1924077, EPI_ISL_1924100, EPI_ISL_1924115, EPI_ISL_1924125, EPI_ISL_1924144, EPI_ISL_1924145, EPI_ISL_1924155, EPI_ISL_1924156, EPI_ISL_1924161, EPI_ISL_1924167, EPI_ISL_1924174, EPI_ISL_1924175, EPI_ISL_1924193, EPI_ISL_1924271, EPI_ISL_1935584, EPI_ISL_1935587, EPI_ISL_1942155

| | | | |
|---|---|---|---|
| see above | Helix/Illumina | Centers for Disease Control and Prevention Division of Viral Diseases, Pathogen Discovery | Adrian Paskey; Alexandre Bolze; Ary Ascencio; Ben L. Rambo-Martin; Benjamin Rambo-Martin; Brad Sickler; Charlotte Rivera-Garcia; Christine Tran; Christopher Gulvick; Clinton R. Paden; Dakota Howard; Darlene Wagner; David Becker; Dhwani Batra; Duncan MacCannell; Efren Sandoval; Eileen de Feo; Elizabeth Cirulli; Eric Allen; Geraint Levan; James Lu; Jan Antico; Jason Caravas; Jason Nguyen; Jimmy Ramirez; Jingtao Liu; Kara Moser; Kelly Schiabor Barrett; Kim Gietzen; Magnus Isaksson; Marc Laurent; Matthew Schmerer; Matthew Tolentino; Nicole L. Washington; Peter W. Cook; Phil Febbo; Ryan Cho; Scott Sammons; Shannon Wickline; Shatavia Morrison; Sherry Wang; Simon White; Summer Galloway; Suxiang Tong; Tyler Cassens; William Lee; Yvette Unoarumhi |

EPI_ISL_850560, EPI_ISL_850566, EPI_ISL_850986, EPI_ISL_850989, EPI_ISL_850997, EPI_ISL_876654, EPI_ISL_876655, EPI_ISL_876700, EPI_ISL_876701, EPI_ISL_876702

| | | | |
|---|---|---|---|
| see above | Helix/Illumina | Genomics and Discovery, Respiratory Viruses Branch, Division of Viral Diseases, Centers | ; Alexandre Bolze; Ary Ascencio; Ben L. Rambo-Martin; Ben L. Rambo-Martin Eileen de Feo; Brad Sickler; Charlotte Rivera-Garcia; Christine Tran; Clinton R. Paden; David Becker; Dhwani Batra; Duncan MacCannell; Efren Sandoval; Eileen de Feo; Elizabeth Cirulli; Eric Allen; Geraint Levan; James Lu; Jan Antico; Jason Nguyen; Jimmy Ramirez; Jingtao Liu; Kelly Schiabor Barrett; Kim Gietzen; Magnus Isaksson; Marc Laurent; Matthew Tolentino; Nicole L. Washington; Peter W. Cook; Phil Febbo; Ryan Cho; Shannon Wickline; Sherry Wang; Simon White; Summer Galloway; Suxiang Tong; Tyler Cassens; William Lee |

| | | | |
|---|---|---|---|
| | | for Disease Control and Prevention | |
| EPI_ISL_966527, EPI_ISL_966616, EPI_ISL_966617, EPI_ISL_966642, EPI_ISL_966647, EPI_ISL_966688, EPI_ISL_966731, EPI_ISL_966872, EPI_ISL_966911, EPI_ISL_966914, EPI_ISL_966932, EPI_ISL_966941, EPI_ISL_966953, EPI_ISL_967032, EPI_ISL_967074, EPI_ISL_967093, EPI_ISL_967100, EPI_ISL_967133, EPI_ISL_967136, EPI_ISL_967142, EPI_ISL_967160, EPI_ISL_967186, EPI_ISL_967187, EPI_ISL_967188, EPI_ISL_967189, EPI_ISL_967190, EPI_ISL_967191, EPI_ISL_967200, EPI_ISL_967205, EPI_ISL_967255, EPI_ISL_967266, EPI_ISL_967271, EPI_ISL_967285, EPI_ISL_967309, EPI_ISL_967377, EPI_ISL_967386, EPI_ISL_967401, EPI_ISL_967408, EPI_ISL_967414, EPI_ISL_967459, EPI_ISL_967466, EPI_ISL_967547, EPI_ISL_967655, EPI_ISL_967826, EPI_ISL_967833, EPI_ISL_967852, EPI_ISL_978614, EPI_ISL_978650, EPI_ISL_978656, EPI_ISL_978664, EPI_ISL_978673, EPI_ISL_978679, EPI_ISL_978701, EPI_ISL_978712, EPI_ISL_978716, EPI_ISL_978724, EPI_ISL_978742, EPI_ISL_978769, EPI_ISL_1016519, EPI_ISL_1016561, EPI_ISL_1016637, EPI_ISL_1016685, EPI_ISL_1016800, EPI_ISL_1016811, EPI_ISL_1081050, EPI_ISL_1110650, EPI_ISL_1110653, EPI_ISL_1110659, EPI_ISL_1133800, EPI_ISL_1133833, EPI_ISL_1133893, EPI_ISL_1133943, EPI_ISL_1133951, EPI_ISL_1133991, EPI_ISL_1133997, EPI_ISL_1133999, EPI_ISL_1134013, EPI_ISL_1134033, EPI_ISL_1134188, EPI_ISL_1134190, EPI_ISL_1134198, EPI_ISL_1134211, EPI_ISL_1134212, EPI_ISL_1134215, EPI_ISL_1134226, EPI_ISL_1134236, EPI_ISL_1134244, EPI_ISL_1134253, EPI_ISL_1134293, EPI_ISL_1134294, EPI_ISL_1134317, EPI_ISL_1134341, EPI_ISL_1134357, EPI_ISL_1134390, EPI_ISL_1134391, EPI_ISL_1134393, EPI_ISL_1134405, EPI_ISL_1134420, EPI_ISL_1134422, EPI_ISL_1134427, EPI_ISL_1134428, EPI_ISL_1134451, EPI_ISL_1134456, EPI_ISL_1134588, EPI_ISL_1134610, EPI_ISL_1134618, EPI_ISL_1134619, EPI_ISL_1134636, EPI_ISL_1134637, EPI_ISL_1134647, EPI_ISL_1134735, EPI_ISL_1134741, EPI_ISL_1134748, EPI_ISL_1134749, EPI_ISL_1134775 | | | |
| see above | Helix/Illumina | Respiratory Viruses Branch, Division of Viral Diseases, Centers for Disease Control and Prevention | ; Alexandre Bolze; Ary Ascencio; Ben L. Rambo-Martin; Brad Sickler; Charlotte Rivera-Garcia; Christine Tran; Clinton R. Paden; Dakota Howard; David Becker; Dhwani Batra; Duncan MacCannell; Efren Sandoval; Eileen de Feo; Elizabeth Cirulli; Eric Allen; Geraint Levan; James Lu; Jan Antico; Jason Nguyen; Jimmy Ramirez; Jingtao Liu; Kelly Schiabor Barrett; Kim Gietzen; Magnus Isaksson; Marc Laurent; Matthew Tolentino; Nicole L. Washington; Peter W. Cook; Phil Febbo; Ryan Cho; Shannon Wickline; Sherry Wang; Simon White; Summer Galloway; Suxiang Tong; Tyler Cassens; William Lee |
| EPI_ISL_907068 | Infectious Diseases, Quest Diagnostics | Infectious Diseases, Quest Diagnostics | Anderson, B.; Bernstein; D.F.; Gerasimova, A.; Hua, M.; K.E.; Kagan; L.E.; Lacbawan, F.; Liu, Y.; Livingston; Owen, R.; R.M.; Rosenthal; S.H.; Shalhout |
| EPI_ISL_1512965, EPI_ISL_1588429, EPI_ISL_1691967, EPI_ISL_1692006, EPI_ISL_1692094, EPI_ISL_1692142, EPI_ISL_1692364, EPI_ISL_1692698, EPI_ISL_1693114, EPI_ISL_1694000, EPI_ISL_1694247, EPI_ISL_1694369, EPI_ISL_1694466, EPI_ISL_1694531, EPI_ISL_1702235, EPI_ISL_1795450, EPI_ISL_1795455, EPI_ISL_1795539, EPI_ISL_1795557, EPI_ISL_1795631, EPI_ISL_1803208 | | | |
| see above | Infinity Biologix | Centers for Disease Control and Prevention Division of Viral Diseases, Pathogen Discovery | Adrian Paskey; Benjamin Rambo-Martin; Chirayu Goswami; Christian Bixby; Christopher Gulvick; Clinton R. Paden; Dakota Howard; Darlene Wagner; Dhwani Batra; Duncan MacCannell; Jason Caravas; Jonathan Schultz; Kara Moser; Matthew Schmerer; Peter W. Cook; Robin Grimwood; Russ Hager; Scott Sammons; Shatavia Morrison; Yihe Wang; Yvette Unoarumhi |
| EPI_ISL_886311, EPI_ISL_886535, EPI_ISL_886897, EPI_ISL_886990, EPI_ISL_887773, EPI_ISL_888127 | Labcorp | Genomics and Discovery, Respiratory Viruses Branch, Division of Viral Diseases, Centers for Disease Control and Prevention | ; Amanda Douglas; Amanda Suchanek; Andrea Throop; Ayla Burns; Ben L. Rambo-Martin; Bobbi Croy; Brian Krueger; Brian Norvell; Christos Petropoulos; Clinton R. Paden; Craig Lukasik; Debbie Boles; Dhwani Batra; Duncan MacCannell; Eyad Almasri; Goran Stevovic; Howard Engler; Hrushikesh Deshmukh; Jake Humphrey; Jana Schroth; Joe Voshell; John Pruitt; Jonathan Meltzer; Jonathan Williams; Kimberly Wagner; Lax Iyer; Lyndon Tilson; Manoj Jain; Marcia Eisenberg; Mary Ann Cristobal; Mary Williamson; Michael Levandoski; Mike Sapeta; Mindy Nye; Minoo Agarwal; Mohan Kolli; Nuthawin Charoensri; Oren Cohen; Peter W. Cook; Prashant Gupta; Qian Zeng; Rama Ghatti; Scott Parker; Scott Ryan; Stanley Letovsky; Steven Ragan; Summer Galloway; Suresh Babu Selvaraju; Susan Countryman; Susan Hicks; Suxiang Tong; Suzanne Dale; Thomas Urban; Tim Kuphal; Tricia Zwiefelhofer; Vincent Drouillon |
| EPI_ISL_1193304, EPI_ISL_1193309, EPI_ISL_1193325, EPI_ISL_1193328, EPI_ISL_1193329, EPI_ISL_1193330, EPI_ISL_1193358, EPI_ISL_1220968, EPI_ISL_1220969, EPI_ISL_1221018, EPI_ISL_1221079, EPI_ISL_1221088, EPI_ISL_1221303, EPI_ISL_1221305, EPI_ISL_1221319, EPI_ISL_1221320, EPI_ISL_1221321, EPI_ISL_1221322, EPI_ISL_1221323, EPI_ISL_1221333, EPI_ISL_1221425, EPI_ISL_1221426, EPI_ISL_1221763, EPI_ISL_1221766, EPI_ISL_1221772, EPI_ISL_1221773, EPI_ISL_1221786, EPI_ISL_1221878, EPI_ISL_1221881, EPI_ISL_1221882, EPI_ISL_1221887, EPI_ISL_1222043, EPI_ISL_1222044, EPI_ISL_1222047, EPI_ISL_1222066, EPI_ISL_1222075, EPI_ISL_1222076, EPI_ISL_1222077, EPI_ISL_1222078, EPI_ISL_1222086, EPI_ISL_1222113, EPI_ISL_1222117, EPI_ISL_1222119, EPI_ISL_1222121, EPI_ISL_1255092, EPI_ISL_1255094, EPI_ISL_1255096, EPI_ISL_1255097, EPI_ISL_1291245, EPI_ISL_1291246, EPI_ISL_1291255, EPI_ISL_1291260, EPI_ISL_1291302, EPI_ISL_1291303, EPI_ISL_1298254, EPI_ISL_1298261, EPI_ISL_1298353, EPI_ISL_1298355, EPI_ISL_1298357, EPI_ISL_1319167, EPI_ISL_1319506, EPI_ISL_1319510, EPI_ISL_1319516, EPI_ISL_1319517, EPI_ISL_1319518, EPI_ISL_1319532, EPI_ISL_1319583, EPI_ISL_1319615, EPI_ISL_1319760, EPI_ISL_1319843, EPI_ISL_1319853, EPI_ISL_1320032, EPI_ISL_1320360, EPI_ISL_1320361, EPI_ISL_1320362, EPI_ISL_1320368, EPI_ISL_1320371, EPI_ISL_1320498, EPI_ISL_1320538, EPI_ISL_1320539, EPI_ISL_1320542, EPI_ISL_1320693, EPI_ISL_1320694, EPI_ISL_1320695, EPI_ISL_1320726, EPI_ISL_1320729, EPI_ISL_1320748, EPI_ISL_1320771, EPI_ISL_1320772, EPI_ISL_1320780, EPI_ISL_1320781, EPI_ISL_1320804, EPI_ISL_1320805, EPI_ISL_1320806, EPI_ISL_1320808, EPI_ISL_1320840, EPI_ISL_1320841, EPI_ISL_1320842, EPI_ISL_1320843, EPI_ISL_1320846, EPI_ISL_1320873, EPI_ISL_1320912, EPI_ISL_1320917, EPI_ISL_1320918, EPI_ISL_1320951, EPI_ISL_1320967, EPI_ISL_1320968, EPI_ISL_1320969, EPI_ISL_1320970, EPI_ISL_1320979, EPI_ISL_1320980, EPI_ISL_1320982, EPI_ISL_1320983, EPI_ISL_1320986, EPI_ISL_1320989, EPI_ISL_1320993, EPI_ISL_1320994, EPI_ISL_1320995, EPI_ISL_1320997, EPI_ISL_1320998, EPI_ISL_1321006, EPI_ISL_1321007, EPI_ISL_1338410, EPI_ISL_1338411, EPI_ISL_1338412, EPI_ISL_1338415, EPI_ISL_1338416, EPI_ISL_1338417, EPI_ISL_1338436, EPI_ISL_1338437, EPI_ISL_1338438, EPI_ISL_1338439, EPI_ISL_1338440, EPI_ISL_1338441, EPI_ISL_1338442, EPI_ISL_1338450, EPI_ISL_1338451, EPI_ISL_1338452, EPI_ISL_1338454, EPI_ISL_1338456, EPI_ISL_1338457, EPI_ISL_1338461, EPI_ISL_1338462, EPI_ISL_1338471, EPI_ISL_1338472, EPI_ISL_1338473, EPI_ISL_1338516, EPI_ISL_1338517, EPI_ISL_1338520, EPI_ISL_1338549, EPI_ISL_1338550, EPI_ISL_1338551, EPI_ISL_1338553, EPI_ISL_1338556, EPI_ISL_1338557, EPI_ISL_1338566, EPI_ISL_1338568, EPI_ISL_1338573, EPI_ISL_1338574, EPI_ISL_1338586, EPI_ISL_1338588, EPI_ISL_1338589, EPI_ISL_1338590, EPI_ISL_1338591, EPI_ISL_1338592, EPI_ISL_1338597, EPI_ISL_1338656, EPI_ISL_1421880, EPI_ISL_1421881, EPI_ISL_1421998, EPI_ISL_1422026, EPI_ISL_1422071, EPI_ISL_1422072, EPI_ISL_1422080, EPI_ISL_1422089, EPI_ISL_1422096, EPI_ISL_1422118, EPI_ISL_1422139, EPI_ISL_1422140, EPI_ISL_1422141, EPI_ISL_1422143, EPI_ISL_1422144, EPI_ISL_1422151, EPI_ISL_1422152, EPI_ISL_1422154, EPI_ISL_1422155, EPI_ISL_1422156, EPI_ISL_1422158, EPI_ISL_1422160, EPI_ISL_1462017, EPI_ISL_1462336, EPI_ISL_1462345, EPI_ISL_1462349, EPI_ISL_1462350, EPI_ISL_1462353, EPI_ISL_1462354, EPI_ISL_1462356, EPI_ISL_1462373, EPI_ISL_1462374, EPI_ISL_1462388, EPI_ISL_1462392, EPI_ISL_1462393, EPI_ISL_1462397, EPI_ISL_1462405, EPI_ISL_1462406, EPI_ISL_1462407, EPI_ISL_1462408, EPI_ISL_1462410, EPI_ISL_1462424, EPI_ISL_1462436, EPI_ISL_1462437, EPI_ISL_1462439, EPI_ISL_1462443, EPI_ISL_1462444, EPI_ISL_1462445, EPI_ISL_1462446, EPI_ISL_1462448, EPI_ISL_1462449, EPI_ISL_1462455, EPI_ISL_1462459, EPI_ISL_1462462, EPI_ISL_1462466, EPI_ISL_1462467, EPI_ISL_1462468, EPI_ISL_1462481, EPI_ISL_1462482, EPI_ISL_1462491, EPI_ISL_1462529, EPI_ISL_1462532, EPI_ISL_1462544, EPI_ISL_1462547, EPI_ISL_1462565, EPI_ISL_1462808, EPI_ISL_1462815, EPI_ISL_1462816, EPI_ISL_1462817, EPI_ISL_1462818, EPI_ISL_1462829, EPI_ISL_1462832, EPI_ISL_1462833, EPI_ISL_1462834, EPI_ISL_1462841, EPI_ISL_1462843, EPI_ISL_1462845, EPI_ISL_1462846, EPI_ISL_1462851, EPI_ISL_1462853, EPI_ISL_1463175, EPI_ISL_1463198, EPI_ISL_1463202, EPI_ISL_1463219, EPI_ISL_1463220, EPI_ISL_1463221, EPI_ISL_1463234, EPI_ISL_1463235, EPI_ISL_1463240, EPI_ISL_1463241, EPI_ISL_1463245, EPI_ISL_1463246, EPI_ISL_1463247, EPI_ISL_1463251, EPI_ISL_1463255, EPI_ISL_1463258, EPI_ISL_1463259, EPI_ISL_1463273, EPI_ISL_1463274, EPI_ISL_1463275, EPI_ISL_1463276, EPI_ISL_1463279, EPI_ISL_1463283, EPI_ISL_1463288, EPI_ISL_1463289, EPI_ISL_1463292, EPI_ISL_1463293, EPI_ISL_1463304, EPI_ISL_1463305, EPI_ISL_1463322, EPI_ISL_1463323, EPI_ISL_1463334, EPI_ISL_1463336, EPI_ISL_1463340, EPI_ISL_1463341, EPI_ISL_1463342, EPI_ISL_1463344, EPI_ISL_1463345, EPI_ISL_1463347, EPI_ISL_1463361, EPI_ISL_1463365, EPI_ISL_1463366, EPI_ISL_1463369, EPI_ISL_1463372, EPI_ISL_1463375, EPI_ISL_1463377, EPI_ISL_1463380, EPI_ISL_1463381, EPI_ISL_1463383, EPI_ISL_1463384, EPI_ISL_1463385, EPI_ISL_1463396, EPI_ISL_1463397, EPI_ISL_1463401, EPI_ISL_1463405, EPI_ISL_1463406, EPI_ISL_1463412, EPI_ISL_1463413, EPI_ISL_1463414, EPI_ISL_1463419, EPI_ISL_1463421, EPI_ISL_1463422, EPI_ISL_1463425, EPI_ISL_1463429, EPI_ISL_1463431, EPI_ISL_1463438, EPI_ISL_1463439, EPI_ISL_1463440, EPI_ISL_1463441, EPI_ISL_1480985, EPI_ISL_1481026, EPI_ISL_1481027, EPI_ISL_1481030, EPI_ISL_1481059, EPI_ISL_1481062, EPI_ISL_1481064, EPI_ISL_1481065, EPI_ISL_1481066, EPI_ISL_1481091, EPI_ISL_1481093, EPI_ISL_1513956, EPI_ISL_1513980, EPI_ISL_1513981, EPI_ISL_1513982, EPI_ISL_1513983, EPI_ISL_1513995, EPI_ISL_1513998, EPI_ISL_1513999, EPI_ISL_1514004, EPI_ISL_1514005, EPI_ISL_1514008, EPI_ISL_1514009, EPI_ISL_1514654, EPI_ISL_1514712, EPI_ISL_1515152, EPI_ISL_1548253, EPI_ISL_1548435, EPI_ISL_1548437, EPI_ISL_1548452, EPI_ISL_1548453, EPI_ISL_1548454, EPI_ISL_1548455, EPI_ISL_1548456, EPI_ISL_1548457, EPI_ISL_1548458, EPI_ISL_1548459, EPI_ISL_1548481, EPI_ISL_1548488, EPI_ISL_1548489, EPI_ISL_1548490, EPI_ISL_1548492, EPI_ISL_1548493, EPI_ISL_1548495, EPI_ISL_1548496, EPI_ISL_1548497, EPI_ISL_1548498, EPI_ISL_1548500, EPI_ISL_1548510, EPI_ISL_1548512, EPI_ISL_1548527, EPI_ISL_1548528, EPI_ISL_1548712, EPI_ISL_1548713, EPI_ISL_1548731, EPI_ISL_1548732, EPI_ISL_1548733, EPI_ISL_1548734, EPI_ISL_1548735, EPI_ISL_1548736, EPI_ISL_1548737, EPI_ISL_1548738, EPI_ISL_1549031, EPI_ISL_1549036, EPI_ISL_1549037, EPI_ISL_1549039, EPI_ISL_1549043, EPI_ISL_1549048, EPI_ISL_1549049, EPI_ISL_1549050, EPI_ISL_1549051, EPI_ISL_1549052, EPI_ISL_1549053, EPI_ISL_1549055, EPI_ISL_1549056, EPI_ISL_1549057, EPI_ISL_1549059, EPI_ISL_1549060, EPI_ISL_1549061, EPI_ISL_1549062, EPI_ISL_1549069, EPI_ISL_1549070, EPI_ISL_1549071, EPI_ISL_1549072, EPI_ISL_1549073, EPI_ISL_1549074, EPI_ISL_1549076, EPI_ISL_1549078, EPI_ISL_1549083, EPI_ISL_1549084, EPI_ISL_1549085, EPI_ISL_1549087, EPI_ISL_1549091, EPI_ISL_1549092, EPI_ISL_1549093, EPI_ISL_1549094, EPI_ISL_1549108, EPI_ISL_1549109, EPI_ISL_1549110, EPI_ISL_1549112, EPI_ISL_1549114, EPI_ISL_1549117, EPI_ISL_1549121, EPI_ISL_1549125, EPI_ISL_1549127, EPI_ISL_1549128, EPI_ISL_1549129, EPI_ISL_1549130, EPI_ISL_1549345, EPI_ISL_1549359, EPI_ISL_1549360, EPI_ISL_1549363, EPI_ISL_1549364, EPI_ISL_1549375, EPI_ISL_1549381, EPI_ISL_1549382, EPI_ISL_1549385, EPI_ISL_1549386, EPI_ISL_1549387, EPI_ISL_1549536, EPI_ISL_1549792, EPI_ISL_1609361, EPI_ISL_1609362, EPI_ISL_1609373, EPI_ISL_1609374, EPI_ISL_1609773, EPI_ISL_1609959, EPI_ISL_1610228, EPI_ISL_1610231, EPI_ISL_1610232, EPI_ISL_1610233, EPI_ISL_1610234, EPI_ISL_1610235, EPI_ISL_1610244, EPI_ISL_1610250, EPI_ISL_1610251, EPI_ISL_1610252, EPI_ISL_1610253, EPI_ISL_1610264, EPI_ISL_1610268, EPI_ISL_1610270, EPI_ISL_1610272, EPI_ISL_1610273, EPI_ISL_1610274, EPI_ISL_1610275, EPI_ISL_1610277, EPI_ISL_1610279, EPI_ISL_1610280, EPI_ISL_1610283, EPI_ISL_1610286, EPI_ISL_1610301, EPI_ISL_1610304, EPI_ISL_1610312, EPI_ISL_1610313, EPI_ISL_1610315, EPI_ISL_1610316, EPI_ISL_1610317, EPI_ISL_1610325, EPI_ISL_1610329, EPI_ISL_1610330, EPI_ISL_1610331, EPI_ISL_1610333, EPI_ISL_1610334, EPI_ISL_1610356, EPI_ISL_1610360, EPI_ISL_1610362, EPI_ISL_1610363, EPI_ISL_1610399, EPI_ISL_1610400, EPI_ISL_1610402, EPI_ISL_1610403, EPI_ISL_1610405, EPI_ISL_1610407, EPI_ISL_1610409, EPI_ISL_1610433, EPI_ISL_1610435, EPI_ISL_1610445, EPI_ISL_1610449, EPI_ISL_1610451, EPI_ISL_1610453, EPI_ISL_1610454, EPI_ISL_1610456, EPI_ISL_1610459, EPI_ISL_1610460, EPI_ISL_1610462, EPI_ISL_1610463, EPI_ISL_1610474, EPI_ISL_1610487, EPI_ISL_1610489, EPI_ISL_1610492, EPI_ISL_1610566, EPI_ISL_1610574, EPI_ISL_1610578, EPI_ISL_1610581, EPI_ISL_1610586, EPI_ISL_1610591, EPI_ISL_1610592, EPI_ISL_1610606, EPI_ISL_1610610, EPI_ISL_1610612, EPI_ISL_1610615, EPI_ISL_1610620, EPI_ISL_1610622, EPI_ISL_1610630, EPI_ISL_1610634, EPI_ISL_1610636, EPI_ISL_1610638, EPI_ISL_1610639, EPI_ISL_1610644, EPI_ISL_1610648, EPI_ISL_1610658, EPI_ISL_1610666, EPI_ISL_1610667, EPI_ISL_1610668, EPI_ISL_1610669, EPI_ISL_1610891, EPI_ISL_1610892, EPI_ISL_1610896, EPI_ISL_1610897, EPI_ISL_1610898, EPI_ISL_1610909, EPI_ISL_1610911, EPI_ISL_1610912, EPI_ISL_1610913, EPI_ISL_1610921, EPI_ISL_1610922, EPI_ISL_1610923, EPI_ISL_1610926, EPI_ISL_1610927, EPI_ISL_1610928, EPI_ISL_1610930, EPI_ISL_1610931, EPI_ISL_1610935, EPI_ISL_1610936, EPI_ISL_1610937, EPI_ISL_1610938, EPI_ISL_1610950, EPI_ISL_1610952, EPI_ISL_1610966, EPI_ISL_1610967, EPI_ISL_1610968, EPI_ISL_1610969, EPI_ISL_1610975, EPI_ISL_1610976, EPI_ISL_1610978, EPI_ISL_1610979, EPI_ISL_1610981, EPI_ISL_1610997, EPI_ISL_1611032, EPI_ISL_1611034, EPI_ISL_1611106, EPI_ISL_1611108, EPI_ISL_1611210, EPI_ISL_1611284, EPI_ISL_1611349, EPI_ISL_1611466, EPI_ISL_1611949, EPI_ISL_1680471, EPI_ISL_1680472, EPI_ISL_1680476, EPI_ISL_1680478, EPI_ISL_1680497, EPI_ISL_1680498, EPI_ISL_1680499, EPI_ISL_1680501, EPI_ISL_1680502, EPI_ISL_1680503, EPI_ISL_1680504, EPI_ISL_1680505, EPI_ISL_1680506, EPI_ISL_1680507, EPI_ISL_1680508, EPI_ISL_1680509, EPI_ISL_1680510, EPI_ISL_1680518, EPI_ISL_1680520, EPI_ISL_1680521, EPI_ISL_1680522, EPI_ISL_1680525, EPI_ISL_1680526, EPI_ISL_1680527, EPI_ISL_1680528, EPI_ISL_1680529, EPI_ISL_1680531, EPI_ISL_1680532, EPI_ISL_1680537, EPI_ISL_1680546, EPI_ISL_1680547, EPI_ISL_1680560, EPI_ISL_1680562, EPI_ISL_1680564, EPI_ISL_1680565, EPI_ISL_1680567, EPI_ISL_1680568, EPI_ISL_1680572, EPI_ISL_1680573, EPI_ISL_1680574, EPI_ISL_1680578, EPI_ISL_1680579, EPI_ISL_1680582, EPI_ISL_1680587, EPI_ISL_1680588, EPI_ISL_1680601, EPI_ISL_1680602, EPI_ISL_1680604, EPI_ISL_1680606, EPI_ISL_1680607, EPI_ISL_1680608, EPI_ISL_1680609, EPI_ISL_1680610, EPI_ISL_1680611, EPI_ISL_1680613, EPI_ISL_1680653, EPI_ISL_1680654, EPI_ISL_1680656, EPI_ISL_1680661, EPI_ISL_1680673, EPI_ISL_1680675, EPI_ISL_1680678, EPI_ISL_1680679, EPI_ISL_1680680, EPI_ISL_1680681, EPI_ISL_1680682, EPI_ISL_1680704, EPI_ISL_1680707, EPI_ISL_1680708, EPI_ISL_1680709, EPI_ISL_1680710, EPI_ISL_1680712, EPI_ISL_1680713, EPI_ISL_1680716, EPI_ISL_1680717, EPI_ISL_1680764, EPI_ISL_1680766, EPI_ISL_1680801, EPI_ISL_1680815, EPI_ISL_1680822, EPI_ISL_1680827, EPI_ISL_1680828, EPI_ISL_1680829, EPI_ISL_1680832, EPI_ISL_1680846, EPI_ISL_1680870, EPI_ISL_1680871, EPI_ISL_1680872, EPI_ISL_1680873, EPI_ISL_1680874, EPI_ISL_1680875, EPI_ISL_1681167, EPI_ISL_1681177, EPI_ISL_1681178, EPI_ISL_1681180, EPI_ISL_1681189, EPI_ISL_1681191, EPI_ISL_1681192, EPI_ISL_1681194, EPI_ISL_1681196, EPI_ISL_1681210, EPI_ISL_1681211, EPI_ISL_1681217, EPI_ISL_1681218, EPI_ISL_1681219, EPI_ISL_1681220, EPI_ISL_1681222, EPI_ISL_1681227, EPI_ISL_1681229, EPI_ISL_1681232, EPI_ISL_1681233, EPI_ISL_1681234, EPI_ISL_1681235, EPI_ISL_1681236, EPI_ISL_1681237, EPI_ISL_1681239, EPI_ISL_1681240, EPI_ISL_1681243, EPI_ISL_1681244, EPI_ISL_1681262, EPI_ISL_1681268, EPI_ISL_1681272, EPI_ISL_1681275, EPI_ISL_1681277, EPI_ISL_1681284, EPI_ISL_1681293, EPI_ISL_1681294, EPI_ISL_1681297, EPI_ISL_1681299, EPI_ISL_1681303, EPI_ISL_1681304, EPI_ISL_1681319, EPI_ISL_1681320, EPI_ISL_1681329, EPI_ISL_1681352, EPI_ISL_1681353, EPI_ISL_1681354, EPI_ISL_1681355, EPI_ISL_1681356, EPI_ISL_1681357, EPI_ISL_1681360, EPI_ISL_1681361, EPI_ISL_1681363, EPI_ISL_1681369, EPI_ISL_1681408, EPI_ISL_1681435, EPI_ISL_1681436, EPI_ISL_1681437, EPI_ISL_1681438, EPI_ISL_1681439, EPI_ISL_1681449, EPI_ISL_1681450, EPI_ISL_1681462, EPI_ISL_1681465, EPI_ISL_1681466, EPI_ISL_1681469, EPI_ISL_1681470, EPI_ISL_1681478, EPI_ISL_1681484, EPI_ISL_1681487, EPI_ISL_1681501, EPI_ISL_1681507, EPI_ISL_1681509, EPI_ISL_1681514, EPI_ISL_1681522, EPI_ISL_1681535, EPI_ISL_1681539, EPI_ISL_1681540, EPI_ISL_1681556, EPI_ISL_1681557, EPI_ISL_1681558, EPI_ISL_1681559, EPI_ISL_1681561, EPI_ISL_1681563, EPI_ISL_1681603, EPI_ISL_1681604, EPI_ISL_1681605, EPI_ISL_1681657, EPI_ISL_1683109, EPI_ISL_1683115, EPI_ISL_1683208, EPI_ISL_1683234, EPI_ISL_1683235, EPI_ISL_1683268, EPI_ISL_1683562, EPI_ISL_1683821, EPI_ISL_1683827, EPI_ISL_1683829, EPI_ISL_1683845, EPI_ISL_1684304, EPI_ISL_1701867, EPI_ISL_1701868, EPI_ISL_1701869, EPI_ISL_1798838, EPI_ISL_1798878, EPI_ISL_1798895, EPI_ISL_1800530, EPI_ISL_1800696, EPI_ISL_1800791, EPI_ISL_1801294, EPI_ISL_1801303, EPI_ISL_1801306, EPI_ISL_1801307, EPI_ISL_1801308, EPI_ISL_1801309, EPI_ISL_1801310, EPI_ISL_1801313, EPI_ISL_1801315, EPI_ISL_1801316, EPI_ISL_1801318, EPI_ISL_1801322, EPI_ISL_1801326, EPI_ISL_1801348, EPI_ISL_1801354, EPI_ISL_1801356, EPI_ISL_1801360, EPI_ISL_1801361, EPI_ISL_1801362, EPI_ISL_1801363, EPI_ISL_1801364, EPI_ISL_1801365, EPI_ISL_1801402, EPI_ISL_1801422, EPI_ISL_1801428, EPI_ISL_1801432, EPI_ISL_1801440, EPI_ISL_1801446, EPI_ISL_1801447, EPI_ISL_1801505, EPI_ISL_1801506, EPI_ISL_1801511, EPI_ISL_1801512, EPI_ISL_1801515, EPI_ISL_1801535, EPI_ISL_1801536, EPI_ISL_1801539, EPI_ISL_1801552, EPI_ISL_1801554, EPI_ISL_1801555, EPI_ISL_1801558, EPI_ISL_1801560, EPI_ISL_1801561, EPI_ISL_1801578, EPI_ISL_1801581, EPI_ISL_1801596, EPI_ISL_1801601, EPI_ISL_1801625, EPI_ISL_1801629, EPI_ISL_1801635, EPI_ISL_1801636, EPI_ISL_1801715, EPI_ISL_1801721, EPI_ISL_1801729, EPI_ISL_1801730, EPI_ISL_1801736, EPI_ISL_1801739, EPI_ISL_1801750, EPI_ISL_1801786, EPI_ISL_1801788, EPI_ISL_1801850, EPI_ISL_1801892, EPI_ISL_1801893, EPI_ISL_1801894, EPI_ISL_1801933, EPI_ISL_1801934, EPI_ISL_1801942, EPI_ISL_1801944, EPI_ISL_1801945, EPI_ISL_1801947, EPI_ISL_1801948, EPI_ISL_1801959, EPI_ISL_1801960, EPI_ISL_1801961, EPI_ISL_1802008, EPI_ISL_1818077, EPI_ISL_1926080, EPI_ISL_1926308, EPI_ISL_1926371, EPI_ISL_1926373, EPI_ISL_1926374, EPI_ISL_1926375, EPI_ISL_1926410, EPI_ISL_1926411, EPI_ISL_1926471, EPI_ISL_1926472, EPI_ISL_1926481, EPI_ISL_1926492, EPI_ISL_1926493, EPI_ISL_1926494, EPI_ISL_1926495, EPI_ISL_1926496, EPI_ISL_1926545, EPI_ISL_1926546, EPI_ISL_1926594, EPI_ISL_1926595, EPI_ISL_1926604, EPI_ISL_1926626, EPI_ISL_1929409, EPI_ISL_1929412, EPI_ISL_1929415, EPI_ISL_1929434, EPI_ISL_1929437, EPI_ISL_1930074, EPI_ISL_1930077, EPI_ISL_1930081, EPI_ISL_1930087, EPI_ISL_1930100, EPI_ISL_1930126, EPI_ISL_1930129, EPI_ISL_1930143, EPI_ISL_1930153, EPI_ISL_1930156, EPI_ISL_1930173, EPI_ISL_1930176, EPI_ISL_1930179, EPI_ISL_1930190, EPI_ISL_1930213, EPI_ISL_1930216, EPI_ISL_1930219, EPI_ISL_1930222, EPI_ISL_1930225, EPI_ISL_1930228, EPI_ISL_1930253, EPI_ISL_1930255, EPI_ISL_1930258, EPI_ISL_1930273, EPI_ISL_1930313, EPI_ISL_1930315, EPI_ISL_1930333, EPI_ISL_1930339, EPI_ISL_1930343, EPI_ISL_1930394, EPI_ISL_1930455, EPI_ISL_1930461, EPI_ISL_1930464, EPI_ISL_1990051 | | | |
| see above | Laboratory Corporation of America | Centers for Disease Control and Prevention Division of Viral Diseases, Pathogen Discovery | Adrian Paskey; Amanda Douglas; Amanda Suchanek; Andrea Throop; Ayla Burns; Ben L. Rambo-Martin; Benjamin Rambo-Martin; Bobbi Croy; Brian Krueger; Brian Norvell; Christopher Gulvick; Christos Petropoulos; Clinton R. Paden; Craig Lukasik; Dakota Howard; Darlene Wagner; Debbie Boles; Dhwani Batra; Duncan MacCannell; Eyad Almasri; Eyad Almasri Debbie Boles; Goran Stevovic; Howard Engler; Hrushikesh Deshmukh; Jake Humphrey; Jana Schroth; Jason Caravas; Joe Voshell; John Pruitt; Jonathan Meltzer; Jonathan Williams; Kara Moser; Kimberly Wagner; Lax Iyer; Lyndon Tilson; Manoj Jain; Marcia Eisenberg; Mary Ann Cristobal; Mary Williamson; Matthew Schmerer; Michael Levandoski; Mike Sapeta; Mindy Nye; Minoo Agarwal; Mohan Kolli; Nuthawin Charoensri; Oren Cohen; Peter W. Cook; Prashant Gupta; Qian Zeng; Rama Ghatti; Scott Parker; Scott Ryan; Scott Sammons; Shatavia Morrison; Stanley Letovsky; Steven Ragan; Suresh Babu Selvaraju; Susan Countryman; Susan Hicks; Suxiang Tong; Suzanne Dale; Thomas Urban; Tim Kuphal; Tricia Zwiefelhofer; Vincent Drouillon; Yvette Unoarumhi |
| EPI_ISL_1020808, EPI_ISL_1020879, EPI_ISL_1021357, EPI_ISL_1021405, EPI_ISL_1021407, EPI_ISL_1021423, EPI_ISL_1021527, EPI_ISL_1021565, EPI_ISL_1021583, EPI_ISL_1021594, EPI_ISL_1021599, EPI_ISL_1021633, EPI_ISL_1021666, EPI_ISL_1021699, EPI_ISL_1021700, EPI_ISL_1021752, EPI_ISL_1021808, EPI_ISL_1021816, EPI_ISL_1021817, EPI_ISL_1021863, EPI_ISL_1021869, EPI_ISL_1021875, EPI_ISL_1021876, EPI_ISL_1021883, EPI_ISL_1021894, EPI_ISL_1021903, EPI_ISL_1021925, EPI_ISL_1021927, EPI_ISL_1021929, EPI_ISL_1021930, EPI_ISL_1021947, EPI_ISL_1021950, EPI_ISL_1021951, EPI_ISL_1021952, EPI_ISL_1021953, EPI_ISL_1021966, EPI_ISL_1021971, EPI_ISL_1021977, EPI_ISL_1021980, EPI_ISL_1021983, EPI_ISL_1021985, EPI_ISL_1021996, EPI_ISL_1021997, EPI_ISL_1021999, EPI_ISL_1022000, EPI_ISL_1022054, EPI_ISL_1022059, EPI_ISL_1022061, EPI_ISL_1022071, EPI_ISL_1022078, EPI_ISL_1022112, EPI_ISL_1022114, EPI_ISL_1022118, EPI_ISL_1022119, EPI_ISL_1022142, EPI_ISL_1022147, EPI_ISL_1022150, EPI_ISL_1022151, EPI_ISL_1022152, EPI_ISL_1022153, EPI_ISL_1022179, EPI_ISL_1022180, EPI_ISL_1022181, EPI_ISL_1022198, EPI_ISL_1022274, EPI_ISL_1022289, EPI_ISL_1022302, EPI_ISL_1022303, EPI_ISL_1022312, EPI_ISL_1022313, EPI_ISL_1022314, EPI_ISL_1022323, EPI_ISL_1022324, EPI_ISL_1022363, EPI_ISL_1022365, EPI_ISL_1022367, EPI_ISL_1022368, EPI_ISL_1022391, EPI_ISL_1025979, EPI_ISL_1026084, EPI_ISL_1026558, EPI_ISL_1026650, EPI_ISL_1027718, EPI_ISL_1028012, EPI_ISL_1029898, EPI_ISL_1030605, EPI_ISL_1030606, EPI_ISL_1030621, EPI_ISL_1030705, EPI_ISL_1030786, EPI_ISL_1031287, EPI_ISL_1031293, EPI_ISL_1031497, EPI_ISL_1031509, EPI_ISL_1031513, EPI_ISL_1031566, EPI_ISL_1031866, EPI_ISL_1032164, EPI_ISL_1032400, EPI_ISL_1032403, EPI_ISL_1036996, EPI_ISL_1037024, EPI_ISL_1037060, EPI_ISL_1037061, EPI_ISL_1037071, EPI_ISL_1037174, EPI_ISL_1037183, EPI_ISL_1037260, EPI_ISL_1037641, EPI_ISL_1037681, EPI_ISL_1037700, EPI_ISL_1037713, EPI_ISL_1037723, EPI_ISL_1037731, EPI_ISL_1037834, EPI_ISL_1037837, EPI_ISL_1037860, EPI_ISL_1038107, EPI_ISL_1038126, EPI_ISL_1038130, EPI_ISL_1038193, EPI_ISL_1038294, EPI_ISL_1038369, EPI_ISL_1038370, EPI_ISL_1038532, EPI_ISL_1038657, EPI_ISL_1038743, EPI_ISL_1038775, EPI_ISL_1038786, EPI_ISL_1159935, EPI_ISL_1159974, EPI_ISL_1159985, EPI_ISL_1159986, EPI_ISL_1160037, EPI_ISL_1160051, EPI_ISL_1160110, EPI_ISL_1160151, EPI_ISL_1160259, EPI_ISL_1160268, EPI_ISL_1160401, EPI_ISL_1160413, EPI_ISL_1160433, EPI_ISL_1160440, EPI_ISL_1160451, EPI_ISL_1160522, EPI_ISL_1160548, EPI_ISL_1160553, EPI_ISL_1160598, EPI_ISL_1160609, EPI_ISL_1160642, EPI_ISL_1160685, EPI_ISL_1160770, EPI_ISL_1160784, EPI_ISL_1160850, EPI_ISL_1160851, EPI_ISL_1160853, EPI_ISL_1160855, EPI_ISL_1160922, EPI_ISL_1160945, EPI_ISL_1160948, EPI_ISL_1160994, EPI_ISL_1161133, EPI_ISL_1161136, EPI_ISL_1161137, EPI_ISL_1161142, EPI_ISL_1161153, EPI_ISL_1161154, EPI_ISL_1161365, EPI_ISL_1161366, EPI_ISL_1161371, EPI_ISL_1161376, EPI_ISL_1161377, EPI_ISL_1161378, EPI_ISL_1161379, EPI_ISL_1161390, EPI_ISL_1161392, EPI_ISL_1161393, EPI_ISL_1161452, EPI_ISL_1161469, EPI_ISL_1161476, EPI_ISL_1161495, EPI_ISL_1161499, EPI_ISL_1161539, EPI_ISL_1161576, EPI_ISL_1161594, EPI_ISL_1161607, EPI_ISL_1161616, EPI_ISL_1161617, EPI_ISL_1161618, EPI_ISL_1161658, EPI_ISL_1161659, EPI_ISL_1161679, EPI_ISL_1161682, EPI_ISL_1161697, EPI_ISL_1161727, EPI_ISL_1161739, EPI_ISL_1161762, EPI_ISL_1161763, EPI_ISL_1161775, EPI_ISL_1161791, EPI_ISL_1161857, EPI_ISL_1161866, EPI_ISL_1161879, EPI_ISL_1162264, EPI_ISL_1162284, EPI_ISL_1162329, EPI_ISL_1162347, EPI_ISL_1162364, EPI_ISL_1162431, EPI_ISL_1162446, EPI_ISL_1162450, EPI_ISL_1162457, EPI_ISL_1162473, EPI_ISL_1162501, EPI_ISL_1162506, EPI_ISL_1162533, EPI_ISL_1162538, EPI_ISL_1162548, EPI_ISL_1162572, EPI_ISL_1162577, EPI_ISL_1162586, EPI_ISL_1162592, EPI_ISL_1162595, EPI_ISL_1162607, EPI_ISL_1162633, EPI_ISL_1162660, EPI_ISL_1162716, EPI_ISL_1162755, EPI_ISL_1162779, EPI_ISL_1162782, EPI_ISL_1162793, EPI_ISL_1162821, EPI_ISL_1162824, EPI_ISL_1162828, EPI_ISL_1162873, EPI_ISL_1162894, EPI_ISL_1162898, EPI_ISL_1162906, EPI_ISL_1162913, EPI_ISL_1162918, EPI_ISL_1162921, EPI_ISL_1162957, EPI_ISL_1162988, EPI_ISL_1162999, EPI_ISL_1163011, EPI_ISL_1163017, EPI_ISL_1163101, EPI_ISL_1163145, EPI_ISL_1163146, EPI_ISL_1163159, EPI_ISL_1163160, EPI_ISL_1163164, EPI_ISL_1163165, EPI_ISL_1163166, EPI_ISL_1163167, EPI_ISL_1163168, EPI_ISL_1163170, EPI_ISL_1163177, EPI_ISL_1163200, EPI_ISL_1163201, EPI_ISL_1163202, EPI_ISL_1163225, EPI_ISL_1163287, EPI_ISL_1163289, EPI_ISL_1163290, EPI_ISL_1163291, EPI_ISL_1163299, EPI_ISL_1163300, EPI_ISL_1163309, EPI_ISL_1163310, EPI_ISL_1163396, EPI_ISL_1163412, EPI_ISL_1163456, EPI_ISL_1163457, EPI_ISL_1163512, EPI_ISL_1163527, EPI_ISL_1255299, EPI_ISL_1255392, EPI_ISL_1255393 | | | |
| see above | Laboratory Corporation of America | Respiratory Viruses Branch, Division of Viral Diseases, Centers for Disease Control and Prevention | Amanda Douglas; Amanda Suchanek; Andrea Throop; Ayla Burns; Ben L. Rambo-Martin; Bobbi Croy; Brian Krueger; Brian Norvell; Christos Petropoulos; Clinton R. Paden; Craig Lukasik; Dakota Howard; Dhwani Batra; Duncan MacCannell; Eyad Almasri Debbie Boles; Goran Stevovic; Howard Engler; Hrushikesh Deshmukh; Jake Humphrey; Jana Schroth; Joe Voshell; John Pruitt; Jonathan Meltzer; Jonathan Williams; Kimberly Wagner; Lax Iyer; Lyndon Tilson; Manoj Jain; Marcia Eisenberg; Mary Ann Cristobal; Mary Williamson; Michael Levandoski; Mike Sapeta; Mindy Nye; Minoo Agarwal; Mohan Kolli; Nuthawin Charoensri; Oren Cohen; Peter W. Cook; Prashant Gupta; Qian Zeng; Rama Ghatti; Scott Parker; Scott Ryan; Stanley Letovsky; Steven Ragan; Suresh Babu Selvaraju; Susan Countryman; Susan Hicks; Suxiang Tong; Suzanne Dale; Thomas Urban; Tim Kuphal; Tricia Zwiefelhofer; Vincent Drouillon |
| EPI_ISL_1318505, EPI_ISL_1318506, EPI_ISL_1318507, EPI_ISL_1318508, EPI_ISL_1318509, EPI_ISL_1446992, EPI_ISL_1446993, EPI_ISL_1446994, EPI_ISL_1446995, EPI_ISL_1446996, EPI_ISL_1446997, EPI_ISL_1446998 | | | |
| see above | MA State Public Health Laboratory | Centers for Disease Control and Prevention Division of Viral Diseases, Pathogen Discovery | Alison Laufer Halpin; Anna Montmayeur; Anna Uehara; Ben L. Rambo-Martin; Clinton R. Paden; Dakota Howard; Darlene Wagner; Dave Wentworth; Dhwani Batra; Haibin Wang; Jasmine Padilla; Jing Zhang; Justin Lee; Katie Dillon; Krista Queen; Kristen Knipe; Kristine Lacek; Lori Rowe; Mark Burroughs; Matthew Schmerer; Mili Sheth; Peter Cook; Peter W. Cook; Rachel Marine; Sam Shepard; Sarah Nobles; Shoshona Le; Suxiang Tong; Vivien Dugan; Yan Li; Ying Tao; Yvette Unoarumhi |
| EPI_ISL_977083, EPI_ISL_977101, EPI_ISL_977102, EPI_ISL_977103, EPI_ISL_977104, EPI_ISL_977105, EPI_ISL_977106, EPI_ISL_977107, EPI_ISL_977108, EPI_ISL_977109, EPI_ISL_977110, EPI_ISL_977111, EPI_ISL_977112, EPI_ISL_977113, EPI_ISL_977114, EPI_ISL_977115, EPI_ISL_977116, EPI_ISL_977117, EPI_ISL_977118, EPI_ISL_977119, EPI_ISL_1011535, EPI_ISL_1011536, EPI_ISL_1011538, EPI_ISL_1011539, EPI_ISL_1011540, EPI_ISL_1011541, EPI_ISL_1011542, EPI_ISL_1011543, EPI_ISL_1011544, EPI_ISL_1011545, EPI_ISL_1011546, EPI_ISL_1011547, EPI_ISL_1011548, EPI_ISL_1011549, EPI_ISL_1011550, EPI_ISL_1011551, EPI_ISL_1011552, EPI_ISL_1011553, EPI_ISL_1011554, EPI_ISL_1011555, EPI_ISL_1011556, EPI_ISL_1011557, EPI_ISL_1011558, EPI_ISL_1011559, EPI_ISL_1011560, EPI_ISL_1011561, EPI_ISL_1011562, EPI_ISL_1011563, EPI_ISL_1011564, EPI_ISL_1011565, EPI_ISL_1011566, EPI_ISL_1011567, EPI_ISL_1011568, EPI_ISL_1011569, EPI_ISL_1011570, EPI_ISL_1011571, EPI_ISL_1011572, EPI_ISL_1011573, EPI_ISL_1011574, EPI_ISL_1011575, EPI_ISL_1011576, EPI_ISL_1011577, EPI_ISL_1011578, EPI_ISL_1011579, EPI_ISL_1011580, EPI_ISL_1011581, EPI_ISL_1011582, EPI_ISL_1011584, EPI_ISL_1011586, EPI_ISL_1011587, EPI_ISL_1011590, EPI_ISL_1011591, EPI_ISL_1011592, EPI_ISL_1011593, EPI_ISL_1011594, EPI_ISL_1011595, EPI_ISL_1011596, EPI_ISL_1011597, EPI_ISL_1121760, EPI_ISL_1121761, EPI_ISL_1121762, EPI_ISL_1121763, EPI_ISL_1121764, EPI_ISL_1121765, EPI_ISL_1121766, EPI_ISL_1121767, EPI_ISL_1121768, EPI_ISL_1121769, EPI_ISL_1121770, EPI_ISL_1121771, EPI_ISL_1121772, EPI_ISL_1121773, EPI_ISL_1121774, EPI_ISL_1121775, EPI_ISL_1121776, EPI_ISL_1121777, EPI_ISL_1121778, EPI_ISL_1121779, EPI_ISL_1121780, EPI_ISL_1121781, EPI_ISL_1121782, EPI_ISL_1121783, EPI_ISL_1121784, EPI_ISL_1121785, EPI_ISL_1121786, EPI_ISL_1121787, EPI_ISL_1121788, EPI_ISL_1121789, EPI_ISL_1121790, EPI_ISL_1121791, EPI_ISL_1121792, EPI_ISL_1121793, EPI_ISL_1121794, EPI_ISL_1121795, EPI_ISL_1121796, EPI_ISL_1121797, EPI_ISL_1121798, EPI_ISL_1121799, EPI_ISL_1121800, EPI_ISL_1121801, EPI_ISL_1121802, EPI_ISL_1121803, EPI_ISL_1121804, EPI_ISL_1121805, EPI_ISL_1121806, EPI_ISL_1121807, EPI_ISL_1121808, EPI_ISL_1121809, EPI_ISL_1121810, EPI_ISL_1121811, EPI_ISL_1121812, EPI_ISL_1121971, EPI_ISL_1413136 | | | |
| see above | Massachusetts General Hospital | Infectious Disease Program, Broad Institute of Harvard and MIT | A.E.; Adams, G.; Anahtar, M.; B.L.; B.W.; Bauer, M.; Birren; Branda, J.; Carter, A.; Cerrato, F.; Chaluvadi, S.; Chapman; Cusick, C.; D.J.; DeRuff, K.; E. and Sabeti; Flowers, K.; Gallagher, G.; Gladden-Young, A.; Gnirke, A.; Harris, J.; J.E.; K.J.; LaRocque, R.; Lagerborg, K.; Lemieux; Lin; Loreth, C.; MacInnis; Neumann, A.; Normandin, E.; P.C.; Park; Pierce, V.; Reilly, S.; Rosenberg; Rudy, M.; Ryan, E.; S.B.; Shaw, B.; Siddle; Slater, D.; Smole, S.; Tomkins-Tinch, C.; Turbett, S. |
| EPI_ISL_876046, EPI_ISL_876050, EPI_ISL_876052, EPI_ISL_876053, EPI_ISL_876056, EPI_ISL_876059, EPI_ISL_876060, EPI_ISL_876061, EPI_ISL_876062, EPI_ISL_876063, EPI_ISL_876065, EPI_ISL_876070, EPI_ISL_876077, EPI_ISL_876080, EPI_ISL_876081, EPI_ISL_876087, EPI_ISL_876095, EPI_ISL_876096, EPI_ISL_876099, EPI_ISL_876101, EPI_ISL_876111, EPI_ISL_876114, EPI_ISL_876118, EPI_ISL_876124, EPI_ISL_876127, EPI_ISL_876128, EPI_ISL_876131, EPI_ISL_876136, EPI_ISL_876137, EPI_ISL_876143, EPI_ISL_876144, EPI_ISL_876146, EPI_ISL_876147, EPI_ISL_876149, EPI_ISL_876153, EPI_ISL_876155, EPI_ISL_876161, EPI_ISL_876162, EPI_ISL_876163, EPI_ISL_876172, EPI_ISL_876173, EPI_ISL_876176, EPI_ISL_876178, EPI_ISL_876180, EPI_ISL_876184, EPI_ISL_876186, EPI_ISL_876190, EPI_ISL_876193, EPI_ISL_876196, EPI_ISL_876197, EPI_ISL_876198, EPI_ISL_876199, EPI_ISL_876200, EPI_ISL_876202, EPI_ISL_876203, EPI_ISL_876208, EPI_ISL_876215, EPI_ISL_876218, EPI_ISL_876219, EPI_ISL_876220, EPI_ISL_876221, EPI_ISL_876222, EPI_ISL_876223, EPI_ISL_876224, EPI_ISL_876225, EPI_ISL_876226, EPI_ISL_876227, EPI_ISL_876228, EPI_ISL_876229, EPI_ISL_876230, EPI_ISL_876231, EPI_ISL_876232, EPI_ISL_876233, EPI_ISL_876234, EPI_ISL_876235, EPI_ISL_876236, EPI_ISL_876237, EPI_ISL_876238, EPI_ISL_876239, EPI_ISL_876240, EPI_ISL_876241, EPI_ISL_876242, EPI_ISL_876243, EPI_ISL_876244, EPI_ISL_876245, EPI_ISL_876246, EPI_ISL_876247, EPI_ISL_876248, EPI_ISL_876249, EPI_ISL_876250, EPI_ISL_876251, EPI_ISL_913393, EPI_ISL_913394, EPI_ISL_913395, EPI_ISL_913396, EPI_ISL_913397, EPI_ISL_913398, EPI_ISL_913399, EPI_ISL_913400, EPI_ISL_913401, EPI_ISL_913402, EPI_ISL_913403, EPI_ISL_913404, EPI_ISL_913405, EPI_ISL_913406, EPI_ISL_913407, EPI_ISL_913408, EPI_ISL_913409, EPI_ISL_913410, EPI_ISL_913411, EPI_ISL_913412, EPI_ISL_913413, EPI_ISL_913414, EPI_ISL_913415, EPI_ISL_913416, EPI_ISL_913417, EPI_ISL_913418, EPI_ISL_913419, EPI_ISL_913420, EPI_ISL_913421, EPI_ISL_913422, EPI_ISL_913423, EPI_ISL_913424, EPI_ISL_913425, EPI_ISL_913426, EPI_ISL_913427, EPI_ISL_913428, EPI_ISL_913429, EPI_ISL_913430, EPI_ISL_913431, EPI_ISL_913432, EPI_ISL_913433, EPI_ISL_913434, EPI_ISL_913435, EPI_ISL_965865, EPI_ISL_965866, EPI_ISL_965867, EPI_ISL_965868, EPI_ISL_965869, EPI_ISL_965870, EPI_ISL_965871, EPI_ISL_965872, EPI_ISL_965873, EPI_ISL_965874, EPI_ISL_965875, EPI_ISL_965876, EPI_ISL_965877, EPI_ISL_965878, EPI_ISL_965879, EPI_ISL_965880, EPI_ISL_965881, EPI_ISL_965882, EPI_ISL_965883, EPI_ISL_965884, EPI_ISL_965885, EPI_ISL_965886, EPI_ISL_965887, EPI_ISL_965888, EPI_ISL_965889, EPI_ISL_965890, EPI_ISL_965891, EPI_ISL_965892, EPI_ISL_965893, EPI_ISL_965894, EPI_ISL_965895, EPI_ISL_965896, EPI_ISL_965897, EPI_ISL_965898, EPI_ISL_965899, EPI_ISL_965900, EPI_ISL_965901, EPI_ISL_965902, EPI_ISL_965903, EPI_ISL_965904, EPI_ISL_965905, EPI_ISL_1009713, EPI_ISL_1009715, EPI_ISL_1009716, EPI_ISL_1009717, EPI_ISL_1009718, EPI_ISL_1009719, EPI_ISL_1009720, EPI_ISL_1009721, EPI_ISL_1009722, EPI_ISL_1009723, EPI_ISL_1009724, EPI_ISL_1009725, EPI_ISL_1009726, EPI_ISL_1009727, EPI_ISL_1009728, | | | |

EPI_ISL_1009729, EPI_ISL_1009730, EPI_ISL_1009731, EPI_ISL_1009732, EPI_ISL_1009733, EPI_ISL_1009734, EPI_ISL_1009735, EPI_ISL_1009736, EPI_ISL_1009737, EPI_ISL_1009738, EPI_ISL_1009739, EPI_ISL_1009740, EPI_ISL_1009741, EPI_ISL_1009742, EPI_ISL_1009743, EPI_ISL_1009744, EPI_ISL_1009745, EPI_ISL_1009746, EPI_ISL_1009747, EPI_ISL_1009748, EPI_ISL_1009749, EPI_ISL_1009750, EPI_ISL_1009751, EPI_ISL_1009752, EPI_ISL_1009753, EPI_ISL_1009754, EPI_ISL_1009755, EPI_ISL_1009756, EPI_ISL_1009757, EPI_ISL_1009758, EPI_ISL_1009759, EPI_ISL_1009760, EPI_ISL_1009761, EPI_ISL_1009762, EPI_ISL_1009763, EPI_ISL_1009764, EPI_ISL_1009765, EPI_ISL_1009766, EPI_ISL_1009767, EPI_ISL_1009768, EPI_ISL_1009769, EPI_ISL_1009770, EPI_ISL_1009771, EPI_ISL_1009772, EPI_ISL_1009773, EPI_ISL_1009774, EPI_ISL_1009775, EPI_ISL_1009776, EPI_ISL_1009777, EPI_ISL_1009778, EPI_ISL_1009779, EPI_ISL_1009780, EPI_ISL_1009781, EPI_ISL_1009782, EPI_ISL_1009783, EPI_ISL_1009784, EPI_ISL_1009785, EPI_ISL_1009786, EPI_ISL_1009787, EPI_ISL_1009788, EPI_ISL_1009789, EPI_ISL_1009790, EPI_ISL_1009791, EPI_ISL_1009792, EPI_ISL_1233254, EPI_ISL_1233255, EPI_ISL_1233256, EPI_ISL_1233257, EPI_ISL_1233258, EPI_ISL_1233259, EPI_ISL_1233260, EPI_ISL_1233261, EPI_ISL_1233262, EPI_ISL_1233271, EPI_ISL_1233272, EPI_ISL_1233273, EPI_ISL_1233274, EPI_ISL_1233275, EPI_ISL_1233276, EPI_ISL_1233277, EPI_ISL_1233278, EPI_ISL_1233279, EPI_ISL_1233280, EPI_ISL_1233281, EPI_ISL_1233282, EPI_ISL_1233283, EPI_ISL_1233284, EPI_ISL_1233285, EPI_ISL_1233286, EPI_ISL_1233287, EPI_ISL_1233288, EPI_ISL_1233289, EPI_ISL_1233290, EPI_ISL_1233291, EPI_ISL_1233292, EPI_ISL_1233293, EPI_ISL_1233303, EPI_ISL_1233304, EPI_ISL_1233305, EPI_ISL_1233306, EPI_ISL_1233307, EPI_ISL_1233308, EPI_ISL_1233309, EPI_ISL_1233310, EPI_ISL_1233311, EPI_ISL_1233312, EPI_ISL_1233313, EPI_ISL_1233314, EPI_ISL_1233315, EPI_ISL_1233316, EPI_ISL_1233317, EPI_ISL_1233318, EPI_ISL_1233319, EPI_ISL_1233320, EPI_ISL_1233321, EPI_ISL_1233322, EPI_ISL_1233323, EPI_ISL_1233324, EPI_ISL_1233325, EPI_ISL_1233326, EPI_ISL_1233327, EPI_ISL_1233328, EPI_ISL_1233329, EPI_ISL_1233330, EPI_ISL_1233331, EPI_ISL_1233332, EPI_ISL_1233333, EPI_ISL_1233334, EPI_ISL_1233335, EPI_ISL_1233336, EPI_ISL_1233337, EPI_ISL_1233338, EPI_ISL_1233339, EPI_ISL_1233340, EPI_ISL_1233341, EPI_ISL_1233342, EPI_ISL_1233343, EPI_ISL_1233344, EPI_ISL_1233345, EPI_ISL_1233346, EPI_ISL_1233347, EPI_ISL_1233358, EPI_ISL_1233359, EPI_ISL_1233360, EPI_ISL_1233361, EPI_ISL_1233362, EPI_ISL_1233363, EPI_ISL_1233364, EPI_ISL_1233365, EPI_ISL_1233366, EPI_ISL_1233367, EPI_ISL_1233368, EPI_ISL_1233369, EPI_ISL_1233370, EPI_ISL_1233371, EPI_ISL_1233372, EPI_ISL_1233373, EPI_ISL_1233374, EPI_ISL_1233375, EPI_ISL_1233376, EPI_ISL_1404617, EPI_ISL_1404618, EPI_ISL_1404619, EPI_ISL_1404620, EPI_ISL_1404621, EPI_ISL_1404622, EPI_ISL_1404623, EPI_ISL_1406697, EPI_ISL_1406698, EPI_ISL_1406699, EPI_ISL_1406700, EPI_ISL_1406701, EPI_ISL_1406702, EPI_ISL_1406703, EPI_ISL_1406704, EPI_ISL_1503460, EPI_ISL_1503462, EPI_ISL_1503463, EPI_ISL_1503464, EPI_ISL_1503465, EPI_ISL_1503466, EPI_ISL_1503468, EPI_ISL_1503469, EPI_ISL_1503470, EPI_ISL_1503471, EPI_ISL_1503472, EPI_ISL_1503473, EPI_ISL_1503474, EPI_ISL_1503475, EPI_ISL_1503476, EPI_ISL_1503477, EPI_ISL_1503478, EPI_ISL_1503479, EPI_ISL_1503480, EPI_ISL_1503498, EPI_ISL_1503499, EPI_ISL_1503500, EPI_ISL_1503501, EPI_ISL_1503502, EPI_ISL_1503503, EPI_ISL_1503504, EPI_ISL_1503505, EPI_ISL_1503506, EPI_ISL_1503514, EPI_ISL_1503515, EPI_ISL_1503516, EPI_ISL_1503517, EPI_ISL_1503518, EPI_ISL_1503519, EPI_ISL_1503520, EPI_ISL_1503521, EPI_ISL_1503522, EPI_ISL_1503523, EPI_ISL_1503524, EPI_ISL_1503525, EPI_ISL_1503526, EPI_ISL_1503527, EPI_ISL_1503528, EPI_ISL_1503529, EPI_ISL_1503530, EPI_ISL_1503531, EPI_ISL_1503532, EPI_ISL_1503533, EPI_ISL_1503534, EPI_ISL_1503535, EPI_ISL_1503536, EPI_ISL_1503537, EPI_ISL_1503538, EPI_ISL_1503539, EPI_ISL_1503540, EPI_ISL_1503541, EPI_ISL_1503542, EPI_ISL_1503543, EPI_ISL_1503544, EPI_ISL_1503545, EPI_ISL_1503546, EPI_ISL_1503547, EPI_ISL_1503548, EPI_ISL_1503549, EPI_ISL_1503561, EPI_ISL_1503562, EPI_ISL_1503563, EPI_ISL_1503564, EPI_ISL_1503565, EPI_ISL_1503566, EPI_ISL_1503567, EPI_ISL_1503568, EPI_ISL_1503569, EPI_ISL_1503570, EPI_ISL_1503571, EPI_ISL_1503572, EPI_ISL_1503573, EPI_ISL_1503574, EPI_ISL_1503575, EPI_ISL_1503576, EPI_ISL_1503578, EPI_ISL_1503579, EPI_ISL_1503580, EPI_ISL_1503581, EPI_ISL_1503582, EPI_ISL_1503584, EPI_ISL_1503585, EPI_ISL_1503586, EPI_ISL_1503587, EPI_ISL_1503588, EPI_ISL_1503589, EPI_ISL_1503590, EPI_ISL_1503591, EPI_ISL_1503592, EPI_ISL_1503593, EPI_ISL_1503595, EPI_ISL_1503596, EPI_ISL_1503597, EPI_ISL_1503598, EPI_ISL_1503599, EPI_ISL_1503600, EPI_ISL_1503601, EPI_ISL_1503602, EPI_ISL_1503603, EPI_ISL_1503604, EPI_ISL_1503605, EPI_ISL_1503606, EPI_ISL_1503607, EPI_ISL_1503608, EPI_ISL_1503609, EPI_ISL_1503610, EPI_ISL_1503611, EPI_ISL_1503612, EPI_ISL_1503613, EPI_ISL_1503614, EPI_ISL_1503615, EPI_ISL_1503616, EPI_ISL_1503617, EPI_ISL_1503618, EPI_ISL_1503619, EPI_ISL_1503620, EPI_ISL_1503621, EPI_ISL_1503622, EPI_ISL_1503623, EPI_ISL_1503624, EPI_ISL_1503625, EPI_ISL_1503626, EPI_ISL_1503627, EPI_ISL_1503628, EPI_ISL_1503629, EPI_ISL_1503630, EPI_ISL_1503631, EPI_ISL_1503632, EPI_ISL_1503633, EPI_ISL_1503634, EPI_ISL_1503635, EPI_ISL_1503636, EPI_ISL_1503637, EPI_ISL_1503639, EPI_ISL_1503640, EPI_ISL_1503641, EPI_ISL_1503642, EPI_ISL_1503643, EPI_ISL_1503644, EPI_ISL_1503645, EPI_ISL_1503646, EPI_ISL_1503647, EPI_ISL_1503648, EPI_ISL_1503649, EPI_ISL_1503650, EPI_ISL_1503651, EPI_ISL_1503656, EPI_ISL_1503657, EPI_ISL_1503658, EPI_ISL_1503659, EPI_ISL_1503660, EPI_ISL_1503661, EPI_ISL_1503662, EPI_ISL_1503663, EPI_ISL_1503664, EPI_ISL_1503665, EPI_ISL_1503666, EPI_ISL_1503667, EPI_ISL_1503668, EPI_ISL_1503669, EPI_ISL_1503670, EPI_ISL_1503671, EPI_ISL_1503672, EPI_ISL_1503673, EPI_ISL_1503674, EPI_ISL_1503675, EPI_ISL_1503676, EPI_ISL_1503677, EPI_ISL_1503678, EPI_ISL_1503679, EPI_ISL_1503680, EPI_ISL_1503681, EPI_ISL_1503682, EPI_ISL_1503683, EPI_ISL_1503684, EPI_ISL_1503685, EPI_ISL_1503686, EPI_ISL_1503687, EPI_ISL_1503688, EPI_ISL_1503690, EPI_ISL_1503691, EPI_ISL_1503692, EPI_ISL_1503693, EPI_ISL_1503694, EPI_ISL_1503695, EPI_ISL_1503696, EPI_ISL_1503697, EPI_ISL_1503698, EPI_ISL_1503699, EPI_ISL_1503700, EPI_ISL_1503701, EPI_ISL_1503702, EPI_ISL_1503703, EPI_ISL_1503704, EPI_ISL_1503705, EPI_ISL_1503706, EPI_ISL_1503707, EPI_ISL_1503708, EPI_ISL_1503709, EPI_ISL_1503710, EPI_ISL_1503711, EPI_ISL_1503712, EPI_ISL_1503713, EPI_ISL_1503714, EPI_ISL_1503715, EPI_ISL_1503717, EPI_ISL_1503718, EPI_ISL_1503719, EPI_ISL_1503720, EPI_ISL_1503721, EPI_ISL_1503722, EPI_ISL_1503723, EPI_ISL_1503724, EPI_ISL_1503725, EPI_ISL_1503726, EPI_ISL_1503727, EPI_ISL_1503728, EPI_ISL_1503729, EPI_ISL_1503730, EPI_ISL_1503731, EPI_ISL_1503732, EPI_ISL_1503733, EPI_ISL_1503734, EPI_ISL_1503735, EPI_ISL_1503736, EPI_ISL_1503737, EPI_ISL_1503738, EPI_ISL_1503739, EPI_ISL_1503740, EPI_ISL_1503741, EPI_ISL_1503742, EPI_ISL_1503743, EPI_ISL_1503744, EPI_ISL_1503745, EPI_ISL_1503746, EPI_ISL_1503747, EPI_ISL_1503748, EPI_ISL_1503756, EPI_ISL_1503757, EPI_ISL_1503758, EPI_ISL_1503759, EPI_ISL_1503760, EPI_ISL_1503761, EPI_ISL_1503762, EPI_ISL_1503763, EPI_ISL_1503764, EPI_ISL_1503765, EPI_ISL_1503766, EPI_ISL_1503767, EPI_ISL_1503768, EPI_ISL_1503769, EPI_ISL_1503770, EPI_ISL_1503771, EPI_ISL_1527091, EPI_ISL_1527093, EPI_ISL_1527094, EPI_ISL_1527097, EPI_ISL_1527098, EPI_ISL_1527099, EPI_ISL_1527100, EPI_ISL_1527106, EPI_ISL_1527107, EPI_ISL_1527108, EPI_ISL_1527109, EPI_ISL_1527110, EPI_ISL_1527112, EPI_ISL_1527113, EPI_ISL_1527114, EPI_ISL_1527115, EPI_ISL_1527116, EPI_ISL_1527118, EPI_ISL_1527119, EPI_ISL_1527120, EPI_ISL_1527121, EPI_ISL_1527122, EPI_ISL_1527123, EPI_ISL_1527124, EPI_ISL_1527125, EPI_ISL_1527126, EPI_ISL_1527127, EPI_ISL_1527128, EPI_ISL_1527129, EPI_ISL_1527130, EPI_ISL_1527131, EPI_ISL_1527132, EPI_ISL_1527133, EPI_ISL_1527134, EPI_ISL_1527135, EPI_ISL_1527136, EPI_ISL_1527137, EPI_ISL_1527138, EPI_ISL_1527139, EPI_ISL_1527140, EPI_ISL_1527141, EPI_ISL_1527142, EPI_ISL_1527143, EPI_ISL_1527144, EPI_ISL_1527145, EPI_ISL_1527146, EPI_ISL_1527147, EPI_ISL_1527148, EPI_ISL_1527149, EPI_ISL_1527150, EPI_ISL_1527153, EPI_ISL_1527154, EPI_ISL_1527155, EPI_ISL_1527158, EPI_ISL_1527159, EPI_ISL_1527160, EPI_ISL_1527162, EPI_ISL_1527163, EPI_ISL_1527164, EPI_ISL_1527165, EPI_ISL_1527166, EPI_ISL_1527167, EPI_ISL_1527168, EPI_ISL_1527169, EPI_ISL_1527170, EPI_ISL_1527171, EPI_ISL_1527172, EPI_ISL_1527173, EPI_ISL_1527174, EPI_ISL_1527176, EPI_ISL_1527177, EPI_ISL_1527178, EPI_ISL_1527179, EPI_ISL_1527180, EPI_ISL_1527181, EPI_ISL_1527182, EPI_ISL_1527183, EPI_ISL_1527184, EPI_ISL_1527185, EPI_ISL_1527186, EPI_ISL_1527187, EPI_ISL_1527188, EPI_ISL_1527189, EPI_ISL_1527190, EPI_ISL_1527191, EPI_ISL_1527192, EPI_ISL_1527201, EPI_ISL_1527203, EPI_ISL_1527204, EPI_ISL_1527205, EPI_ISL_1527209, EPI_ISL_1527210, EPI_ISL_1527211, EPI_ISL_1527213, EPI_ISL_1527214, EPI_ISL_1527216, EPI_ISL_1527217, EPI_ISL_1527218, EPI_ISL_1527219, EPI_ISL_1527220, EPI_ISL_1527221, EPI_ISL_1527222, EPI_ISL_1527225, EPI_ISL_1527226, EPI_ISL_1527227, EPI_ISL_1527228, EPI_ISL_1528093, EPI_ISL_1528094, EPI_ISL_1528095, EPI_ISL_1528096, EPI_ISL_1528097, EPI_ISL_1528098, EPI_ISL_1528099, EPI_ISL_1528100, EPI_ISL_1528101, EPI_ISL_1528102, EPI_ISL_1528103, EPI_ISL_1528104, EPI_ISL_1528105, EPI_ISL_1528106, EPI_ISL_1528107, EPI_ISL_1528108, EPI_ISL_1528109, EPI_ISL_1528110, EPI_ISL_1528111, EPI_ISL_1528112, EPI_ISL_1528113, EPI_ISL_1528114, EPI_ISL_1528115, EPI_ISL_1528116, EPI_ISL_1528117, EPI_ISL_1528118, EPI_ISL_1528119, EPI_ISL_1528120, EPI_ISL_1528121, EPI_ISL_1528122, EPI_ISL_1528123, EPI_ISL_1528124, EPI_ISL_1528125, EPI_ISL_1528126, EPI_ISL_1528127, EPI_ISL_1528128, EPI_ISL_1528129, EPI_ISL_1528130, EPI_ISL_1528131, EPI_ISL_1528132, EPI_ISL_1528133, EPI_ISL_1528134, EPI_ISL_1528135, EPI_ISL_1528136, EPI_ISL_1528137, EPI_ISL_1528138, EPI_ISL_1528139, EPI_ISL_1528140, EPI_ISL_1528141, EPI_ISL_1528142, EPI_ISL_1528143, EPI_ISL_1528144, EPI_ISL_1528145, EPI_ISL_1528146, EPI_ISL_1528147, EPI_ISL_1528148, EPI_ISL_1528149, EPI_ISL_1528150, EPI_ISL_1528153, EPI_ISL_1528156, EPI_ISL_1528157, EPI_ISL_1528158, EPI_ISL_1528159, EPI_ISL_1624489, EPI_ISL_1624490, EPI_ISL_1624491, EPI_ISL_1624492, EPI_ISL_1624493, EPI_ISL_1624494, EPI_ISL_1624495, EPI_ISL_1624497, EPI_ISL_1624498, EPI_ISL_1624499, EPI_ISL_1624500, EPI_ISL_1624501, EPI_ISL_1624502, EPI_ISL_1624503, EPI_ISL_1624504, EPI_ISL_1624505, EPI_ISL_1624506, EPI_ISL_1624507, EPI_ISL_1624508, EPI_ISL_1624509, EPI_ISL_1624510, EPI_ISL_1624511, EPI_ISL_1624512, EPI_ISL_1624513, EPI_ISL_1624515, EPI_ISL_1624516, EPI_ISL_1624517, EPI_ISL_1624518, EPI_ISL_1624519, EPI_ISL_1624520, EPI_ISL_1624521, EPI_ISL_1624522, EPI_ISL_1624524, EPI_ISL_1624525, EPI_ISL_1624526, EPI_ISL_1624527, EPI_ISL_1624528, EPI_ISL_1624529, EPI_ISL_1624530, EPI_ISL_1624531, EPI_ISL_1624532, EPI_ISL_1624533, EPI_ISL_1624534, EPI_ISL_1624535, EPI_ISL_1624537, EPI_ISL_1624538, EPI_ISL_1624539, EPI_ISL_1624540, EPI_ISL_1624541, EPI_ISL_1624542, EPI_ISL_1624543, EPI_ISL_1624545, EPI_ISL_1624547, EPI_ISL_1624548, EPI_ISL_1624549, EPI_ISL_1624550, EPI_ISL_1624551, EPI_ISL_1624552, EPI_ISL_1624553, EPI_ISL_1624554, EPI_ISL_1624555, EPI_ISL_1624556, EPI_ISL_1624558, EPI_ISL_1624559, EPI_ISL_1624560, EPI_ISL_1624561, EPI_ISL_1624562, EPI_ISL_1624563, EPI_ISL_1624564, EPI_ISL_1624565, EPI_ISL_1624566, EPI_ISL_1624567, EPI_ISL_1624568, EPI_ISL_1624569, EPI_ISL_1624570, EPI_ISL_1624571, EPI_ISL_1624573, EPI_ISL_1624574, EPI_ISL_1624575, EPI_ISL_1624576, EPI_ISL_1624577, EPI_ISL_1624578, EPI_ISL_1624579, EPI_ISL_1624580, EPI_ISL_1624581, EPI_ISL_1624582, EPI_ISL_1624583, EPI_ISL_1624584, EPI_ISL_1624585, EPI_ISL_1624586, EPI_ISL_1624587, EPI_ISL_1624588, EPI_ISL_1624589, EPI_ISL_1624590, EPI_ISL_1624591, EPI_ISL_1624592, EPI_ISL_1624593, EPI_ISL_1624594, EPI_ISL_1624595, EPI_ISL_1624596, EPI_ISL_1624597, EPI_ISL_1624598, EPI_ISL_1624599, EPI_ISL_1624600, EPI_ISL_1624601, EPI_ISL_1624602, EPI_ISL_1624603, EPI_ISL_1624604, EPI_ISL_1624605, EPI_ISL_1624606, EPI_ISL_1624607, EPI_ISL_1624608, EPI_ISL_1624609, EPI_ISL_1624610, EPI_ISL_1624611, EPI_ISL_1624612, EPI_ISL_1624613, EPI_ISL_1624614, EPI_ISL_1624615, EPI_ISL_1624616, EPI_ISL_1624617, EPI_ISL_1624618, EPI_ISL_1624619, EPI_ISL_1624620, EPI_ISL_1624621, EPI_ISL_1624622, EPI_ISL_1624623, EPI_ISL_1624624, EPI_ISL_1624625, EPI_ISL_1624626, EPI_ISL_1624627, EPI_ISL_1624628, EPI_ISL_1624629, EPI_ISL_1624630, EPI_ISL_1624631, EPI_ISL_1624632, EPI_ISL_1624633, EPI_ISL_1624634, EPI_ISL_1624637, EPI_ISL_1624638, EPI_ISL_1624639, EPI_ISL_1624640, EPI_ISL_1624641, EPI_ISL_1624642, EPI_ISL_1624643, EPI_ISL_1624644, EPI_ISL_1624645, EPI_ISL_1624646, EPI_ISL_1624647, EPI_ISL_1624648, EPI_ISL_1624649, EPI_ISL_1624650, EPI_ISL_1624651, EPI_ISL_1624652, EPI_ISL_1624653, EPI_ISL_1624654, EPI_ISL_1624655, EPI_ISL_1624656, EPI_ISL_1624657, EPI_ISL_1624661, EPI_ISL_1624662, EPI_ISL_1624663, EPI_ISL_1624664, EPI_ISL_1624665, EPI_ISL_1624666, EPI_ISL_1624667, EPI_ISL_1624668, EPI_ISL_1624669, EPI_ISL_1624670, EPI_ISL_1624671, EPI_ISL_1624672, EPI_ISL_1624673, EPI_ISL_1624674, EPI_ISL_1624677, EPI_ISL_1624679, EPI_ISL_1624680, EPI_ISL_1624681, EPI_ISL_1624683, EPI_ISL_1624684, EPI_ISL_1624685, EPI_ISL_1624686, EPI_ISL_1624687, EPI_ISL_1624688, EPI_ISL_1624689, EPI_ISL_1624690, EPI_ISL_1624692, EPI_ISL_1624694, EPI_ISL_1624697, EPI_ISL_1624698, EPI_ISL_1624699, EPI_ISL_1624700, EPI_ISL_1624703, EPI_ISL_1624706, EPI_ISL_1624707, EPI_ISL_1624708, EPI_ISL_1624709, EPI_ISL_1624710, EPI_ISL_1624711, EPI_ISL_1624712, EPI_ISL_1624713, EPI_ISL_1624714, EPI_ISL_1624715, EPI_ISL_1624716, EPI_ISL_1710599, EPI_ISL_1710600, EPI_ISL_1710601, EPI_ISL_1710602, EPI_ISL_1710603, EPI_ISL_1710605, EPI_ISL_1710607, EPI_ISL_1710608, EPI_ISL_1710609, EPI_ISL_1710610, EPI_ISL_1710611, EPI_ISL_1710612, EPI_ISL_1710614, EPI_ISL_1710616, EPI_ISL_1710618, EPI_ISL_1710622, EPI_ISL_1710624, EPI_ISL_1710627, EPI_ISL_1710628, EPI_ISL_1710629, EPI_ISL_1710630, EPI_ISL_1710632, EPI_ISL_1710637, EPI_ISL_1710638, EPI_ISL_1710639, EPI_ISL_1710640, EPI_ISL_1710641, EPI_ISL_1710642, EPI_ISL_1710643, EPI_ISL_1710644, EPI_ISL_1710645, EPI_ISL_1710646, EPI_ISL_1710648, EPI_ISL_1710649, EPI_ISL_1710650, EPI_ISL_1710652, EPI_ISL_1710653, EPI_ISL_1710654, EPI_ISL_1710655, EPI_ISL_1710658, EPI_ISL_1710659, EPI_ISL_1710660

| | | | |
|---|---|---|---|
| see above | Massachusetts State Public Health Laboratory | Massachusetts State Public Health Laboratory | Andrew Lang; Glen Gallagher; Sandra Smole; Timelia Fink |

EPI_ISL_854874, EPI_ISL_854875, EPI_ISL_854876, EPI_ISL_854877, EPI_ISL_854878, EPI_ISL_854879, EPI_ISL_854880, EPI_ISL_854881, EPI_ISL_854882, EPI_ISL_854883, EPI_ISL_854884, EPI_ISL_854885, EPI_ISL_854886, EPI_ISL_854887, EPI_ISL_854888, EPI_ISL_854889, EPI_ISL_854890, EPI_ISL_854891, EPI_ISL_854892, EPI_ISL_854893, EPI_ISL_854894, EPI_ISL_854895, EPI_ISL_854896, EPI_ISL_854897, EPI_ISL_854898, EPI_ISL_854899, EPI_ISL_854900, EPI_ISL_854901, EPI_ISL_854902, EPI_ISL_854903, EPI_ISL_854904, EPI_ISL_854905, EPI_ISL_854906, EPI_ISL_854907, EPI_ISL_854908, EPI_ISL_854909, EPI_ISL_854910, EPI_ISL_854911, EPI_ISL_854912, EPI_ISL_854913, EPI_ISL_854914, EPI_ISL_854915, EPI_ISL_854916, EPI_ISL_854917, EPI_ISL_854918, EPI_ISL_854919, EPI_ISL_854920, EPI_ISL_854921, EPI_ISL_854922, EPI_ISL_854923, EPI_ISL_854924, EPI_ISL_854925, EPI_ISL_854926, EPI_ISL_854927, EPI_ISL_854928, EPI_ISL_854929, EPI_ISL_854930, EPI_ISL_854931, EPI_ISL_854932, EPI_ISL_854933, EPI_ISL_854934, EPI_ISL_854935, EPI_ISL_854936, EPI_ISL_854937, EPI_ISL_854938, EPI_ISL_854939, EPI_ISL_854940, EPI_ISL_854941, EPI_ISL_854942, EPI_ISL_854943, EPI_ISL_854944, EPI_ISL_854945, EPI_ISL_854946, EPI_ISL_854947, EPI_ISL_854948, EPI_ISL_854949, EPI_ISL_854950, EPI_ISL_854951, EPI_ISL_854952, EPI_ISL_854953, EPI_ISL_854954, EPI_ISL_854955, EPI_ISL_854956, EPI_ISL_854957, EPI_ISL_854958, EPI_ISL_854959, EPI_ISL_854960, EPI_ISL_854961, EPI_ISL_854962, EPI_ISL_855096, EPI_ISL_855097, EPI_ISL_855098, EPI_ISL_855099, EPI_ISL_855100, EPI_ISL_855101, EPI_ISL_855102, EPI_ISL_855103, EPI_ISL_855104, EPI_ISL_855105, EPI_ISL_855106, EPI_ISL_855107, EPI_ISL_855108, EPI_ISL_855109, EPI_ISL_855110, EPI_ISL_855111, EPI_ISL_855112, EPI_ISL_855113, EPI_ISL_855114, EPI_ISL_855115, EPI_ISL_855116, EPI_ISL_855117, EPI_ISL_855118, EPI_ISL_855119, EPI_ISL_855120, EPI_ISL_855121, EPI_ISL_855122, EPI_ISL_855123, EPI_ISL_855124, EPI_ISL_855125, EPI_ISL_855126, EPI_ISL_855127, EPI_ISL_855128, EPI_ISL_855129, EPI_ISL_855130, EPI_ISL_855131, EPI_ISL_855132, EPI_ISL_855133, EPI_ISL_855134, EPI_ISL_855135, EPI_ISL_855136, EPI_ISL_855137, EPI_ISL_855138, EPI_ISL_855139, EPI_ISL_855140, EPI_ISL_855141, EPI_ISL_855142, EPI_ISL_855143, EPI_ISL_855144, EPI_ISL_855145, EPI_ISL_855146, EPI_ISL_855147, EPI_ISL_855148, EPI_ISL_855149, EPI_ISL_855150, EPI_ISL_855151, EPI_ISL_855152, EPI_ISL_855153, EPI_ISL_855154, EPI_ISL_877190, EPI_ISL_877191, EPI_ISL_877192, EPI_ISL_877193, EPI_ISL_877194, EPI_ISL_877195, EPI_ISL_877201, EPI_ISL_877202, EPI_ISL_877203, EPI_ISL_903531, EPI_ISL_903532, EPI_ISL_903533, EPI_ISL_903534, EPI_ISL_903535, EPI_ISL_903536, EPI_ISL_903537, EPI_ISL_903538, EPI_ISL_903539, EPI_ISL_903540, EPI_ISL_903541, EPI_ISL_903542, EPI_ISL_903543, EPI_ISL_903544, EPI_ISL_903545, EPI_ISL_903546, EPI_ISL_903547, EPI_ISL_903548, EPI_ISL_903549, EPI_ISL_903550, EPI_ISL_904132, EPI_ISL_904133, EPI_ISL_906432, EPI_ISL_906433, EPI_ISL_906434, EPI_ISL_906435, EPI_ISL_906436, EPI_ISL_906437, EPI_ISL_906438, EPI_ISL_906439, EPI_ISL_906440, EPI_ISL_906441, EPI_ISL_906442, EPI_ISL_906443, EPI_ISL_915323, EPI_ISL_915324, EPI_ISL_915325, EPI_ISL_915326

| | | | |
|---|---|---|---|
| see above | Quest Diagnostics | Quest Diagnostics | Anderson, B.; Bernstein; D.F.; Gerasimova, A.; Hua, M.; I.A.; K.E.; Kagan; L.E.; Lacbawan, F.; Liu Y.; Livingston; Owen, R.; Perez, A.; R.M.; Rosenthal; S.H.; Shalhout; Shlyakhter; Tanpaiboon, P. |

EPI_ISL_1159242, EPI_ISL_1159243, EPI_ISL_1159245, EPI_ISL_1159252, EPI_ISL_1159270, EPI_ISL_1159280, EPI_ISL_1159282, EPI_ISL_1159285, EPI_ISL_1159286, EPI_ISL_1159296, EPI_ISL_1159318, EPI_ISL_1159345, EPI_ISL_1193376, EPI_ISL_1193391, EPI_ISL_1193396, EPI_ISL_1193403, EPI_ISL_1193408, EPI_ISL_1193420, EPI_ISL_1193424, EPI_ISL_1193427, EPI_ISL_1193428, EPI_ISL_1193429, EPI_ISL_1193440, EPI_ISL_1193441, EPI_ISL_1193446, EPI_ISL_1193452, EPI_ISL_1193454, EPI_ISL_1193463, EPI_ISL_1193465, EPI_ISL_1193470, EPI_ISL_1193474, EPI_ISL_1193478, EPI_ISL_1193485, EPI_ISL_1193489, EPI_ISL_1193496, EPI_ISL_1193500, EPI_ISL_1193502, EPI_ISL_1193510, EPI_ISL_1193529, EPI_ISL_1193541, EPI_ISL_1193569, EPI_ISL_1193579, EPI_ISL_1193583, EPI_ISL_1193591, EPI_ISL_1193596, EPI_ISL_1193599, EPI_ISL_1193610, EPI_ISL_1193612, EPI_ISL_1193619, EPI_ISL_1193640, EPI_ISL_1193756, EPI_ISL_1193792, EPI_ISL_1193813, EPI_ISL_1193825, EPI_ISL_1193848, EPI_ISL_1193870, EPI_ISL_1193878, EPI_ISL_1193893, EPI_ISL_1193905, EPI_ISL_1193938, EPI_ISL_1193940, EPI_ISL_1193965, EPI_ISL_1193974, EPI_ISL_1194011, EPI_ISL_1194410, EPI_ISL_1194430, EPI_ISL_1194439, EPI_ISL_1194441, EPI_ISL_1194470, EPI_ISL_1194482, EPI_ISL_1194483, EPI_ISL_1194575, EPI_ISL_1220363, EPI_ISL_1220365, EPI_ISL_1220378, EPI_ISL_1220385, EPI_ISL_1220397, EPI_ISL_1220409, EPI_ISL_1220443, EPI_ISL_1220456, EPI_ISL_1220462, EPI_ISL_1220498, EPI_ISL_1220504, EPI_ISL_1220505, EPI_ISL_1220510, EPI_ISL_1220525, EPI_ISL_1220550, EPI_ISL_1220556, EPI_ISL_1220557, EPI_ISL_1220561, EPI_ISL_1220571, EPI_ISL_1220585, EPI_ISL_1220588, EPI_ISL_1220591, EPI_ISL_1220595, EPI_ISL_1220599, EPI_ISL_1220602, EPI_ISL_1220610, EPI_ISL_1220614, EPI_ISL_1220616, EPI_ISL_1220625, EPI_ISL_1220628, EPI_ISL_1220637, EPI_ISL_1220648, EPI_ISL_1220651, EPI_ISL_1220654, EPI_ISL_1220658, EPI_ISL_1220666, EPI_ISL_1220673, EPI_ISL_1220674, EPI_ISL_1220680, EPI_ISL_1220683, EPI_ISL_1220684, EPI_ISL_1220688, EPI_ISL_1220689, EPI_ISL_1220691, EPI_ISL_1220692, EPI_ISL_1220699, EPI_ISL_1220700, EPI_ISL_1252917, EPI_ISL_1252939, EPI_ISL_1252949, EPI_ISL_1252959, EPI_ISL_1252964, EPI_ISL_1252966, EPI_ISL_1252981, EPI_ISL_1252984, EPI_ISL_1252997, EPI_ISL_1253002, EPI_ISL_1253003, EPI_ISL_1253016, EPI_ISL_1253027, EPI_ISL_1253043, EPI_ISL_1253051, EPI_ISL_1253066, EPI_ISL_1253070, EPI_ISL_1253073, EPI_ISL_1253079, EPI_ISL_1253103, EPI_ISL_1253107, EPI_ISL_1253110, EPI_ISL_1253122, EPI_ISL_1253125, EPI_ISL_1253137, EPI_ISL_1253148, EPI_ISL_1253149, EPI_ISL_1253150, EPI_ISL_1253153, EPI_ISL_1253157, EPI_ISL_1253168, EPI_ISL_1253177, EPI_ISL_1253179, EPI_ISL_1253181, EPI_ISL_1253206, EPI_ISL_1253217, EPI_ISL_1253227, EPI_ISL_1253237, EPI_ISL_1253242, EPI_ISL_1253248, EPI_ISL_1253249, EPI_ISL_1253251, EPI_ISL_1253252, EPI_ISL_1253255, EPI_ISL_1253256, EPI_ISL_1253260, EPI_ISL_1253272, EPI_ISL_1254321, EPI_ISL_1254323, EPI_ISL_1254331, EPI_ISL_1254333, EPI_ISL_1254346, EPI_ISL_1254349, EPI_ISL_1254361, EPI_ISL_1254365, EPI_ISL_1254371, EPI_ISL_1254376, EPI_ISL_1254380, EPI_ISL_1254384, EPI_ISL_1254387, EPI_ISL_1254388, EPI_ISL_1254390, EPI_ISL_1254392, EPI_ISL_1254393, EPI_ISL_1254400, EPI_ISL_1254406, EPI_ISL_1254407, EPI_ISL_1254408, EPI_ISL_1254409, EPI_ISL_1265917, EPI_ISL_1265919, EPI_ISL_1265927, EPI_ISL_1265934, EPI_ISL_1265939, EPI_ISL_1265943, EPI_ISL_1265945, EPI_ISL_1265955, EPI_ISL_1265959, EPI_ISL_1265975, EPI_ISL_1265997, EPI_ISL_1266000, EPI_ISL_1266005, EPI_ISL_1266015, EPI_ISL_1266042, EPI_ISL_1266053, EPI_ISL_1266059, EPI_ISL_1266067, EPI_ISL_1266084, EPI_ISL_1266108, EPI_ISL_1266130, EPI_ISL_1266155, EPI_ISL_1266156, EPI_ISL_1266171, EPI_ISL_1266172, EPI_ISL_1266179, EPI_ISL_1266181, EPI_ISL_1266191, EPI_ISL_1266211, EPI_ISL_1266222, EPI_ISL_1266832, EPI_ISL_1266851, EPI_ISL_1266878, EPI_ISL_1266899, EPI_ISL_1266947, EPI_ISL_1266969, EPI_ISL_1266981, EPI_ISL_1267039, EPI_ISL_1267070, EPI_ISL_1267099, EPI_ISL_1267102, EPI_ISL_1267108, EPI_ISL_1267114, EPI_ISL_1267117, EPI_ISL_1267119, EPI_ISL_1267121, EPI_ISL_1267123, EPI_ISL_1267126, EPI_ISL_1267130, EPI_ISL_1267133, EPI_ISL_1267137, EPI_ISL_1267139, EPI_ISL_1267140, EPI_ISL_1267141, EPI_ISL_1267149, EPI_ISL_1267150, EPI_ISL_1267155, EPI_ISL_1267164, EPI_ISL_1267165, EPI_ISL_1267167, EPI_ISL_1267171, EPI_ISL_1267184, EPI_ISL_1267186, EPI_ISL_1267190, EPI_ISL_1267191, EPI_ISL_1267194, EPI_ISL_1267196, EPI_ISL_1267203, EPI_ISL_1267204, EPI_ISL_1267205, EPI_ISL_1267208, EPI_ISL_1267213, EPI_ISL_1267216, EPI_ISL_1267238, EPI_ISL_1267244, EPI_ISL_1267247, EPI_ISL_1267250, EPI_ISL_1267253, EPI_ISL_1267277, EPI_ISL_1267289, EPI_ISL_1267298, EPI_ISL_1267338, EPI_ISL_1267340, EPI_ISL_1267341, EPI_ISL_1267343, EPI_ISL_1267345, EPI_ISL_1267347, EPI_ISL_1267364, EPI_ISL_1267369, EPI_ISL_1267372, EPI_ISL_1267378, EPI_ISL_1267380, EPI_ISL_1267381, EPI_ISL_1267384, EPI_ISL_1267401, EPI_ISL_1267413, EPI_ISL_1267420, EPI_ISL_1267425, EPI_ISL_1267427, EPI_ISL_1267428, EPI_ISL_1267435, EPI_ISL_1267438, EPI_ISL_1267440, EPI_ISL_1267444, EPI_ISL_1267450, EPI_ISL_1267452, EPI_ISL_1267459, EPI_ISL_1267460, EPI_ISL_1267469, EPI_ISL_1267475, EPI_ISL_1267482, EPI_ISL_1267484, EPI_ISL_1291107, EPI_ISL_1291108, EPI_ISL_1291122, EPI_ISL_1291136, EPI_ISL_1291139, EPI_ISL_1291174, EPI_ISL_1291181, EPI_ISL_1291184, EPI_ISL_1291392, EPI_ISL_1291393, EPI_ISL_1291401, EPI_ISL_1291403, EPI_ISL_1291405, EPI_ISL_1291409, EPI_ISL_1291462, EPI_ISL_1291475, EPI_ISL_1291481, EPI_ISL_1291533, EPI_ISL_1291536, EPI_ISL_1291571, EPI_ISL_1291578, EPI_ISL_1297590, EPI_ISL_1297594, EPI_ISL_1297596, EPI_ISL_1297597, EPI_ISL_1297607, EPI_ISL_1297608, EPI_ISL_1297609, EPI_ISL_1297611, EPI_ISL_1297612, EPI_ISL_1297615, EPI_ISL_1297617, EPI_ISL_1297618, EPI_ISL_1297620, EPI_ISL_1297625, EPI_ISL_1297628, EPI_ISL_1297629, EPI_ISL_1297630, EPI_ISL_1297633, EPI_ISL_1297637, EPI_ISL_1297638, EPI_ISL_1297641, EPI_ISL_1297646, EPI_ISL_1297650, EPI_ISL_1297651, EPI_ISL_1297652, EPI_ISL_1297659, EPI_ISL_1297664, EPI_ISL_1297668, EPI_ISL_1297675, EPI_ISL_1297676, EPI_ISL_1297704, EPI_ISL_1297706, EPI_ISL_1297711, EPI_ISL_1297714, EPI_ISL_1297715, EPI_ISL_1297717, EPI_ISL_1297721, EPI_ISL_1297723, EPI_ISL_1297725, EPI_ISL_1297728, EPI_ISL_1297729, EPI_ISL_1297731, EPI_ISL_1297736, EPI_ISL_1297739, EPI_ISL_1297740, EPI_ISL_1297742, EPI_ISL_1297746, EPI_ISL_1297750, EPI_ISL_1297753, EPI_ISL_1297762, EPI_ISL_1297763, EPI_ISL_1314545, EPI_ISL_1314547, EPI_ISL_1314549, EPI_ISL_1314554, EPI_ISL_1314563, EPI_ISL_1314571, EPI_ISL_1314574, EPI_ISL_1314576, EPI_ISL_1314577, EPI_ISL_1314583, EPI_ISL_1314587, EPI_ISL_1314592, EPI_ISL_1314593, EPI_ISL_1314598, EPI_ISL_1314602, EPI_ISL_1314607, EPI_ISL_1314612, EPI_ISL_1314627, EPI_ISL_1314631, EPI_ISL_1314632, EPI_ISL_1314636, EPI_ISL_1314639, EPI_ISL_1314640, EPI_ISL_1314690, EPI_ISL_1314748, EPI_ISL_1314755, EPI_ISL_1314776, EPI_ISL_1314839, EPI_ISL_1314894, EPI_ISL_1314912, EPI_ISL_1314935, EPI_ISL_1335908, EPI_ISL_1335928, EPI_ISL_1335937, EPI_ISL_1335957, EPI_ISL_1335959, EPI_ISL_1335961, EPI_ISL_1335964, EPI_ISL_1335966, EPI_ISL_1335972, EPI_ISL_1335991, EPI_ISL_1336036, EPI_ISL_1336037, EPI_ISL_1336042, EPI_ISL_1336112, EPI_ISL_1339823, EPI_ISL_1339826, EPI_ISL_1339903, EPI_ISL_1339906, EPI_ISL_1339907, EPI_ISL_1339919, EPI_ISL_1339925, EPI_ISL_1339926, EPI_ISL_1339939, EPI_ISL_1339943, EPI_ISL_1339951, EPI_ISL_1339953, EPI_ISL_1339957, EPI_ISL_1340538, EPI_ISL_1340544, EPI_ISL_1340556, EPI_ISL_1340558, EPI_ISL_1340560, EPI_ISL_1340567, EPI_ISL_1340568, EPI_ISL_1340577, EPI_ISL_1340578, EPI_ISL_1340583, EPI_ISL_1340584, EPI_ISL_1340585, EPI_ISL_1340638, EPI_ISL_1340662, EPI_ISL_1340663, EPI_ISL_1340664, EPI_ISL_1340668, EPI_ISL_1340676, EPI_ISL_1340704, EPI_ISL_1340707, EPI_ISL_1340713, EPI_ISL_1340742, EPI_ISL_1367164, EPI_ISL_1367193, EPI_ISL_1367205, EPI_ISL_1367208, EPI_ISL_1367216, EPI_ISL_1367235, EPI_ISL_1367240, EPI_ISL_1367245, EPI_ISL_1367246, EPI_ISL_1367255, EPI_ISL_1367258, EPI_ISL_1367260, EPI_ISL_1367270, EPI_ISL_1367272, EPI_ISL_1367275, EPI_ISL_1367276, EPI_ISL_1367280, EPI_ISL_1367287, EPI_ISL_1367289, EPI_ISL_1367291, EPI_ISL_1367292, EPI_ISL_1367295, EPI_ISL_1367297, EPI_ISL_1367299, EPI_ISL_1367304, EPI_ISL_1367308, EPI_ISL_1367317, EPI_ISL_1367320, EPI_ISL_1367323, EPI_ISL_1367327, EPI_ISL_1367416, EPI_ISL_1367427, EPI_ISL_1367433, EPI_ISL_1367454, EPI_ISL_1367459, EPI_ISL_1367473, EPI_ISL_1391420, EPI_ISL_1391426, EPI_ISL_1391428, EPI_ISL_1391434, EPI_ISL_1391438, EPI_ISL_1391445, EPI_ISL_1391448, EPI_ISL_1391461, EPI_ISL_1391468, EPI_ISL_1391472, EPI_ISL_1391648, EPI_ISL_1391662, EPI_ISL_1422330, EPI_ISL_1422344, EPI_ISL_1422353, EPI_ISL_1422376, EPI_ISL_1422395, EPI_ISL_1422404, EPI_ISL_1422407, EPI_ISL_1422482, EPI_ISL_1422489, EPI_ISL_1422499, EPI_ISL_1422502, EPI_ISL_1422529, EPI_ISL_1422531, EPI_ISL_1422537, EPI_ISL_1422538, EPI_ISL_1422539, EPI_ISL_1422542, EPI_ISL_1422559, EPI_ISL_1422560, EPI_ISL_1422591, EPI_ISL_1422593, EPI_ISL_1422601, EPI_ISL_1422603, EPI_ISL_1422619, EPI_ISL_1422625, EPI_ISL_1422628, EPI_ISL_1422631, EPI_ISL_1422632, EPI_ISL_1422670, EPI_ISL_1422676, EPI_ISL_1422678, EPI_ISL_1422695, EPI_ISL_1422704, EPI_ISL_1422754, EPI_ISL_1422760, EPI_ISL_1422762, EPI_ISL_1422763, EPI_ISL_1422764, EPI_ISL_1422765, EPI_ISL_1422767, EPI_ISL_1422768, EPI_ISL_1422771, EPI_ISL_1422773, EPI_ISL_1422774, EPI_ISL_1422775, EPI_ISL_1422780, EPI_ISL_1422784, EPI_ISL_1422785, EPI_ISL_1422786, EPI_ISL_1422789, EPI_ISL_1422792, EPI_ISL_1422793, EPI_ISL_1422796, EPI_ISL_1422797, EPI_ISL_1422855, EPI_ISL_1422856, EPI_ISL_1422857, EPI_ISL_1422861, EPI_ISL_1422862, EPI_ISL_1422867, EPI_ISL_1422872, EPI_ISL_1422874, EPI_ISL_1422882, EPI_ISL_1422884, EPI_ISL_1422887, EPI_ISL_1422929, EPI_ISL_1422963, EPI_ISL_1422979, EPI_ISL_1422987, EPI_ISL_1444500, EPI_ISL_1444506, EPI_ISL_1444510, EPI_ISL_1444511, EPI_ISL_1444512, EPI_ISL_1444514, EPI_ISL_1444521, EPI_ISL_1444522, EPI_ISL_1444526, EPI_ISL_1444533, EPI_ISL_1444541, EPI_ISL_1444547, EPI_ISL_1444550, EPI_ISL_1444553, EPI_ISL_1444554, EPI_ISL_1444555, EPI_ISL_1444557, EPI_ISL_1444707, EPI_ISL_1444710, EPI_ISL_1444722, EPI_ISL_1444725, EPI_ISL_1444727, EPI_ISL_1444729, EPI_ISL_1479755, EPI_ISL_1479925, EPI_ISL_1479927, EPI_ISL_1479931, EPI_ISL_1479937, EPI_ISL_1479940, EPI_ISL_1479957, EPI_ISL_1479959, EPI_ISL_1480534, EPI_ISL_1480547, EPI_ISL_1480568, EPI_ISL_1480618, EPI_ISL_1480624, EPI_ISL_1480625, EPI_ISL_1480633, EPI_ISL_1480652, EPI_ISL_1480657, EPI_ISL_1480659, EPI_ISL_1480676, EPI_ISL_1480685, EPI_ISL_1480690, EPI_ISL_1480697, EPI_ISL_1480698, EPI_ISL_1480702, EPI_ISL_1480709, EPI_ISL_1480714, EPI_ISL_1480726, EPI_ISL_1480730, EPI_ISL_1480731, EPI_ISL_1480733, EPI_ISL_1480785, EPI_ISL_1480796, EPI_ISL_1480800, EPI_ISL_1480802, EPI_ISL_1480804, EPI_ISL_1480809, EPI_ISL_1480827, EPI_ISL_1480828, EPI_ISL_1480833, EPI_ISL_1480843, EPI_ISL_1480847, EPI_ISL_1480850, EPI_ISL_1480851, EPI_ISL_1480854, EPI_ISL_1480857, EPI_ISL_1480864, EPI_ISL_1480865, EPI_ISL_1480866, EPI_ISL_1480867, EPI_ISL_1480868, EPI_ISL_1480873, EPI_ISL_1480957, EPI_ISL_1480960, EPI_ISL_1491966, EPI_ISL_1491973, EPI_ISL_1491977, EPI_ISL_1491990, EPI_ISL_1491999, EPI_ISL_1492016, EPI_ISL_1492024, EPI_ISL_1493603, EPI_ISL_1493604, EPI_ISL_1493611, EPI_ISL_1493613, EPI_ISL_1493634, EPI_ISL_1493635, EPI_ISL_1493637, EPI_ISL_1493644, EPI_ISL_1493651, EPI_ISL_1493661, EPI_ISL_1493987, EPI_ISL_1493990, EPI_ISL_1493997, EPI_ISL_1493999, EPI_ISL_1494000, EPI_ISL_1494013, EPI_ISL_1494014, EPI_ISL_1494015, EPI_ISL_1494018, EPI_ISL_1494019, EPI_ISL_1494020, EPI_ISL_1494028, EPI_ISL_1511673, EPI_ISL_1525947, EPI_ISL_1551735, EPI_ISL_1551759, EPI_ISL_1551768, EPI_ISL_1551796, EPI_ISL_1551811, EPI_ISL_1551812, EPI_ISL_1551815, EPI_ISL_1551821, EPI_ISL_1551824, EPI_ISL_1551826, EPI_ISL_1551827, EPI_ISL_1551832, EPI_ISL_1551836, EPI_ISL_1551837, EPI_ISL_1551838, EPI_ISL_1551840, EPI_ISL_1551843, EPI_ISL_1551848, EPI_ISL_1551849, EPI_ISL_1551852, EPI_ISL_1551853, EPI_ISL_1551856, EPI_ISL_1551858, EPI_ISL_1551859, EPI_ISL_1551862, EPI_ISL_1551863, EPI_ISL_1551870, EPI_ISL_1551874, EPI_ISL_1551876, EPI_ISL_1551877, EPI_ISL_1551878, EPI_ISL_1551925, EPI_ISL_1551928, EPI_ISL_1551933, EPI_ISL_1551935, EPI_ISL_1551943, EPI_ISL_1551948, EPI_ISL_1551953, EPI_ISL_1551959, EPI_ISL_1551962, EPI_ISL_1551978, EPI_ISL_1551988, EPI_ISL_1552003, EPI_ISL_1552165, EPI_ISL_1552218, EPI_ISL_1552220, EPI_ISL_1552327, EPI_ISL_1552333, EPI_ISL_1552336, EPI_ISL_1552337, EPI_ISL_1552339, EPI_ISL_1552355, EPI_ISL_1552356, EPI_ISL_1552361, EPI_ISL_1552378, EPI_ISL_1552388, EPI_ISL_1552405, EPI_ISL_1552409, EPI_ISL_1552976, EPI_ISL_1553133, EPI_ISL_1553161, EPI_ISL_1615034, EPI_ISL_1622051, EPI_ISL_1702700, EPI_ISL_1711970, EPI_ISL_1738149, EPI_ISL_1753157, EPI_ISL_1753227, EPI_ISL_1797910, EPI_ISL_1797964, EPI_ISL_1797982, EPI_ISL_1818060, EPI_ISL_1818061, EPI_ISL_1818141, EPI_ISL_1818142, EPI_ISL_1818148, EPI_ISL_1818149, EPI_ISL_1818152, EPI_ISL_1818153, EPI_ISL_1818155, EPI_ISL_1818159, EPI_ISL_1818164, EPI_ISL_1818170, EPI_ISL_1818180, EPI_ISL_1818183, EPI_ISL_1818196, EPI_ISL_1818202, EPI_ISL_1818203, EPI_ISL_1818204, EPI_ISL_1818206, EPI_ISL_1818209, EPI_ISL_1818215, EPI_ISL_1818224, EPI_ISL_1818227, EPI_ISL_1818247, EPI_ISL_1818249, EPI_ISL_1818256, EPI_ISL_1818258, EPI_ISL_1818259, EPI_ISL_1818260, EPI_ISL_1818262, EPI_ISL_1818264, EPI_ISL_1818269, EPI_ISL_1818274, EPI_ISL_1818278, EPI_ISL_1818402, EPI_ISL_1818414, EPI_ISL_1834209, EPI_ISL_1834214, EPI_ISL_1834220, EPI_ISL_1834226, EPI_ISL_1834254, EPI_ISL_1834256, EPI_ISL_1834257, EPI_ISL_1834270, EPI_ISL_1834295, EPI_ISL_1834298, EPI_ISL_1834316, EPI_ISL_1834352, EPI_ISL_1834373, EPI_ISL_1834378, EPI_ISL_1834390, EPI_ISL_1834401, EPI_ISL_1834404, EPI_ISL_1834410, EPI_ISL_1838922, EPI_ISL_1838927, EPI_ISL_1838938, EPI_ISL_1838941, EPI_ISL_1838945, EPI_ISL_1838947, EPI_ISL_1838966, EPI_ISL_1838968, EPI_ISL_1838970, EPI_ISL_1838979, EPI_ISL_1838984, EPI_ISL_1838989, EPI_ISL_1838997, EPI_ISL_1839018, EPI_ISL_1839026, EPI_ISL_1840421, EPI_ISL_1840437, EPI_ISL_1840444, EPI_ISL_1840477, EPI_ISL_1840482, EPI_ISL_1840493, EPI_ISL_1840502, EPI_ISL_1840581, EPI_ISL_1840609, EPI_ISL_1840614, EPI_ISL_1900040, EPI_ISL_1900046, EPI_ISL_1900048, EPI_ISL_1900051, EPI_ISL_1900057, EPI_ISL_1900124, EPI_ISL_1900134, EPI_ISL_1900137, EPI_ISL_1900428, EPI_ISL_1900429, EPI_ISL_1900431, EPI_ISL_1900432, EPI_ISL_1900435, EPI_ISL_1900464, EPI_ISL_1900465, EPI_ISL_1935570

| | | | |
|---|---|---|---|
| see above | Quest Diagnostics Incorporated | Centers for Disease Control and Prevention Division of Viral Diseases, Pathogen Discovery | A. Gerasimova; A. Perez; Adrian Paskey; B. Anderson; Ben L. Rambo-Martin; Benjamin Rambo-Martin; Christopher Gulvick; Clinton R. Paden; Dakota Howard; Darlene Wagner; Dhwani Batra; Duncan MacCannell; F. Lacbawan; I. A. Shlyakhter; Jason Caravas; K.E. Livingston; Kara Moser; L.E. Bernstein; M. Hua; Matthew Schmerer; P. Tanpaiboon; Peter W. Cook; R. M. Kagan; R. Owen; R. V. Rolando; S. H. Rosenthal; Scott Sammons; Shatavia Morrison; Suxiang Tong; Y. Liu; Yvette Unoarumhi |

EPI_ISL_1086676, EPI_ISL_1086696, EPI_ISL_1086702, EPI_ISL_1086705, EPI_ISL_1086708, EPI_ISL_1086724, EPI_ISL_1086727, EPI_ISL_1086763, EPI_ISL_1086766, EPI_ISL_1086786, EPI_ISL_1086788, EPI_ISL_1086800, EPI_ISL_1086856, EPI_ISL_1086987, EPI_ISL_1087020, EPI_ISL_1087041, EPI_ISL_1087058, EPI_ISL_1087076, EPI_ISL_1087561, EPI_ISL_1087622, EPI_ISL_1087624, EPI_ISL_1087629, EPI_ISL_1087637, EPI_ISL_1087653, EPI_ISL_1087654, EPI_ISL_1087658, EPI_ISL_1087664, EPI_ISL_1087678, EPI_ISL_1087680, EPI_ISL_1087687, EPI_ISL_1087688, EPI_ISL_1087699, EPI_ISL_1087700, EPI_ISL_1087706, EPI_ISL_1087712, EPI_ISL_1087717, EPI_ISL_1087722, EPI_ISL_1087833, EPI_ISL_1087977, EPI_ISL_1090417, EPI_ISL_1090534, EPI_ISL_1090538, EPI_ISL_1090543, EPI_ISL_1090547, EPI_ISL_1090555, EPI_ISL_1090556, EPI_ISL_1090558, EPI_ISL_1090570, EPI_ISL_1090571, EPI_ISL_1090584, EPI_ISL_1090591, EPI_ISL_1090592, EPI_ISL_1090594, EPI_ISL_1090624, EPI_ISL_1090848, EPI_ISL_1090852, EPI_ISL_1090886, EPI_ISL_1090894, EPI_ISL_1090895, EPI_ISL_1090900, EPI_ISL_1090917, EPI_ISL_1090944, EPI_ISL_1091007, EPI_ISL_1113764, EPI_ISL_1113766, EPI_ISL_1113767, EPI_ISL_1113768, EPI_ISL_1113770, EPI_ISL_1113777, EPI_ISL_1113782, EPI_ISL_1113785, EPI_ISL_1113787, EPI_ISL_1113793, EPI_ISL_1113806, EPI_ISL_1113807, EPI_ISL_1113811, EPI_ISL_1113812, EPI_ISL_1113815, EPI_ISL_1113818, EPI_ISL_1113819, EPI_ISL_1113820, EPI_ISL_1113822, EPI_ISL_1113824, EPI_ISL_1113833, EPI_ISL_1113835, EPI_ISL_1113836, EPI_ISL_1113839, EPI_ISL_1113841, EPI_ISL_1113847, EPI_ISL_1113848, EPI_ISL_1113850, EPI_ISL_1113852, EPI_ISL_1113856, EPI_ISL_1113858, EPI_ISL_1113860, EPI_ISL_1113862, EPI_ISL_1113868, EPI_ISL_1113870, EPI_ISL_1113872, EPI_ISL_1113876, EPI_ISL_1113879, EPI_ISL_1113880, EPI_ISL_1113885, EPI_ISL_1113887, EPI_ISL_1113889, EPI_ISL_1113890, EPI_ISL_1113894, EPI_ISL_1113907, EPI_ISL_1113910, EPI_ISL_1113911, EPI_ISL_1113914, EPI_ISL_1113921, EPI_ISL_1113923, EPI_ISL_1113930, EPI_ISL_1113931, EPI_ISL_1113946, EPI_ISL_1113947, EPI_ISL_1113956, EPI_ISL_1113965, EPI_ISL_1114101, EPI_ISL_1139219, EPI_ISL_1139221, EPI_ISL_1139222, EPI_ISL_1139226, EPI_ISL_1139266, EPI_ISL_1139271, EPI_ISL_1139278, EPI_ISL_1139279, EPI_ISL_1139280, EPI_ISL_1139283, EPI_ISL_1139288, EPI_ISL_1139311, EPI_ISL_1139316, EPI_ISL_1139329, EPI_ISL_1139364, EPI_ISL_1139375, EPI_ISL_1139402, EPI_ISL_1139403, EPI_ISL_1139423, EPI_ISL_1139486, EPI_ISL_1139487, EPI_ISL_1139488, EPI_ISL_1139512, EPI_ISL_1139520, EPI_ISL_1139532, EPI_ISL_1139533, EPI_ISL_1139536, EPI_ISL_1139537, EPI_ISL_1139539, EPI_ISL_1139542, EPI_ISL_1139545, EPI_ISL_1139546, EPI_ISL_1139553, EPI_ISL_1139555, EPI_ISL_1139556, EPI_ISL_1139558, EPI_ISL_1139560, EPI_ISL_1139563, EPI_ISL_1139565, EPI_ISL_1139570, EPI_ISL_1139572, EPI_ISL_1139574, EPI_ISL_1139580, EPI_ISL_1139582, EPI_ISL_1139583, EPI_ISL_1139584, EPI_ISL_1139585, EPI_ISL_1139586, EPI_ISL_1139592, EPI_ISL_1139644, EPI_ISL_1139647, EPI_ISL_1139685, EPI_ISL_1139701, EPI_ISL_1139854, EPI_ISL_1158946, EPI_ISL_1158949, EPI_ISL_1158950, EPI_ISL_1158951, EPI_ISL_1158953, EPI_ISL_1158964, EPI_ISL_1158966, EPI_ISL_1158971, EPI_ISL_1158973, EPI_ISL_1158976, EPI_ISL_1158985, EPI_ISL_1158987, EPI_ISL_1159007, EPI_ISL_1159027, EPI_ISL_1159029, EPI_ISL_1159033, EPI_ISL_1159034, EPI_ISL_1159049, EPI_ISL_1159052, EPI_ISL_1159053, EPI_ISL_1159054, EPI_ISL_1159061, EPI_ISL_1159065, EPI_ISL_1159559, EPI_ISL_1159563, EPI_ISL_1159564, EPI_ISL_1159570, EPI_ISL_1159573, EPI_ISL_1159580, EPI_ISL_1159581, EPI_ISL_1159583, EPI_ISL_1159585, EPI_ISL_1159606, EPI_ISL_1159622, EPI_ISL_1159627, EPI_ISL_1159628, EPI_ISL_1159632, EPI_ISL_1159633, EPI_ISL_1159635, EPI_ISL_1159638, EPI_ISL_1159641, EPI_ISL_1159645, EPI_ISL_1159651, EPI_ISL_1159657, EPI_ISL_1159658, EPI_ISL_1159665, EPI_ISL_1159666, EPI_ISL_1159667, EPI_ISL_1159672, EPI_ISL_1159677, EPI_ISL_1290857, EPI_ISL_1293333

| | | | |
|---|---|---|---|
| see above | Quest Diagnostics Incorporated | Respiratory Viruses Branch, Division of Viral Diseases, Centers | A. Gerasimova; A. Perez; B. Anderson; Ben L. Rambo-Martin; Clinton R. Paden; Dakota Howard; Dhwani Batra; Duncan MacCannell; F. Lacbawan; I. A. Shlyakhter; K.E. Livingston; L.E. Bernstein; M. Hua; P. Tanpaiboon; Peter W. Cook; R. M. Kagan; R. Owen; R. V. Rolando; S. H. Rosenthal; Suxiang Tong; Y. Liu |

| | | | |
|---|---|---|---|
| | | for Disease Control and Prevention | |
| EPI_ISL_884618, EPI_ISL_884632, EPI_ISL_884670, EPI_ISL_884675, EPI_ISL_884683, EPI_ISL_884688, EPI_ISL_884785, EPI_ISL_884789, EPI_ISL_884792, EPI_ISL_884808, EPI_ISL_884814, EPI_ISL_1233886 | | | |
| see above | Respiratory Viruses Branch, Centers for Disease Control and Prevention | Respiratory Viruses Branch, Centers for Disease Control and Prevention | Allen, E.; Antico, J.; Ascencio, A.; B.L.; Barret; Barrett; Batra, D.; Becker, D.; Bolze, A.; C.R.; Cassens, T.; Cho, R.; Cirulli, E.; Cook; D.T.; Febbo, P.; Galloway, S.; Gietzen, K.; Howard; Isaksson, M.; K.S.; Laurent, M.; Lee, W.; Levan, G.; Liu, J.; Lu, J.; MacCannell, D.; N.L.; Nguyen, J.; P.W.; Paden; Rambo-Martin; Ramirez, J.; Rivera-Garcia, C.; Sandoval, E.; Sickler, B.; Tolentino, M.; Tong, S.; Tran, C.; Wang, S.; Washington; White, S.; Wickline, S.; de Feo, E. |
| EPI_ISL_861747 | Tempus | Grubaugh Lab - Yale School of Public Health | Anderson Brito; Anne Wyllie; Annie Watkins; Chaney Kalinich; Chantal Vogels; Isabel Ott; Joseph Fauver; Mallery Breban; Mary Petrone; Nathan Grubaugh; Tara Alpert |
| EPI_ISL_1315149, EPI_ISL_1540732, EPI_ISL_1540831 | The Jackson Laboratory | The Jackson Laboratory | Adams M; Bergeron D; Kelly K; Li L; Long J; Omerza G; Renzette N |
| EPI_ISL_1447575, EPI_ISL_1447579, EPI_ISL_1447588, EPI_ISL_1447626 | Yale Clinical Virology Lab | Grubaugh Lab - Yale School of Public Health | Anderson Brito; Annie Watkins; Chaney Kalinich; Chantal Vogels; Isabel Ott; Jessica Rothman; Joseph Fauver; Mallery Breban; Marie L. Landry; Mary Petrone; Nathan Grubaugh; Tara Alpert |
