## Supplementary material for "Comparative transmissibility of SARS-CoV-2 variants Delta and Alpha in New England, USA": DataS1-7: DataS4_Massachusetts-2.pdf

We gratefully acknowledge the following Authors from the Originating laboratories responsible for obtaining the specimens, as well as the Submitting laboratories where the genome data were generated and shared via GISAID, on which this research is based.

All Submitters of data may be contacted directly via www.gisaid.org

Authors are sorted alphabetically.

| Accession ID | Originating Laboratory | Submitting Laboratory | Authors |
|---|---|---|---|
| EPI_ISL_1997051, EPI_ISL_1997066, EPI_ISL_1997099, EPI_ISL_1997178, EPI_ISL_1997255, EPI_ISL_1997340, EPI_ISL_1997562, EPI_ISL_1997563, EPI_ISL_1997564, EPI_ISL_1997607, EPI_ISL_1997631, EPI_ISL_1997632, EPI_ISL_1997633, EPI_ISL_1997634, EPI_ISL_1997656, EPI_ISL_1997657, EPI_ISL_1997658, EPI_ISL_1997659, EPI_ISL_1997660, EPI_ISL_1997661, EPI_ISL_1997662, EPI_ISL_1997663, EPI_ISL_1997664, EPI_ISL_1997670, EPI_ISL_1997733, EPI_ISL_1997792, EPI_ISL_1997795, EPI_ISL_1997818, EPI_ISL_1997825, EPI_ISL_1997826, EPI_ISL_1997841, EPI_ISL_2035149, EPI_ISL_2035168, EPI_ISL_2035205, EPI_ISL_2035218, EPI_ISL_2035224, EPI_ISL_2035249, EPI_ISL_2035250, EPI_ISL_2035259, EPI_ISL_2035265, EPI_ISL_2038996, EPI_ISL_2039121, EPI_ISL_2039164, EPI_ISL_2039277, EPI_ISL_2039287, EPI_ISL_2039289, EPI_ISL_2039290, EPI_ISL_2039292, EPI_ISL_2039293, EPI_ISL_2039294, EPI_ISL_2039295, EPI_ISL_2039308, EPI_ISL_2039452, EPI_ISL_2039587, EPI_ISL_2039590, EPI_ISL_2039591, EPI_ISL_2039617, EPI_ISL_2039655, EPI_ISL_2039709, EPI_ISL_2039718, EPI_ISL_2039789, EPI_ISL_2039839, EPI_ISL_2039843, EPI_ISL_2039882, EPI_ISL_2040036, EPI_ISL_2040048, EPI_ISL_2040050, EPI_ISL_2040084, EPI_ISL_2040109, EPI_ISL_2040149, EPI_ISL_2040174, EPI_ISL_2040200, EPI_ISL_2040267, EPI_ISL_2040294, EPI_ISL_2040354, EPI_ISL_2040355, EPI_ISL_2040371, EPI_ISL_2040454, EPI_ISL_2040491, EPI_ISL_2040492, EPI_ISL_2040503, EPI_ISL_2040512, EPI_ISL_2040532, EPI_ISL_2040541, EPI_ISL_2040566, EPI_ISL_2040669, EPI_ISL_2040670, EPI_ISL_2040674, EPI_ISL_2040745, EPI_ISL_2040785, EPI_ISL_2040907, EPI_ISL_2040945, EPI_ISL_2040947, EPI_ISL_2041005, EPI_ISL_2041055, EPI_ISL_2041081, EPI_ISL_2041083, EPI_ISL_2041084, EPI_ISL_2041085, EPI_ISL_2041086, EPI_ISL_2041088, EPI_ISL_2041151, EPI_ISL_2041164, EPI_ISL_2041165, EPI_ISL_2041166, EPI_ISL_2041185, EPI_ISL_2041192, EPI_ISL_2041194, EPI_ISL_2041195, EPI_ISL_2041196, EPI_ISL_2041197, EPI_ISL_2041198, EPI_ISL_2041200, EPI_ISL_2041207, EPI_ISL_2041208, EPI_ISL_2041210, EPI_ISL_2041211, EPI_ISL_2041212, EPI_ISL_2041213, EPI_ISL_2041214, EPI_ISL_2041219, EPI_ISL_2041220, EPI_ISL_2041221, EPI_ISL_2041222, EPI_ISL_2041223, EPI_ISL_2041225, EPI_ISL_2041256, EPI_ISL_2041303, EPI_ISL_2041392, EPI_ISL_2041408, EPI_ISL_2041410, EPI_ISL_2041411, EPI_ISL_2041413, EPI_ISL_2041417, EPI_ISL_2041421, EPI_ISL_2041425, EPI_ISL_2041485, EPI_ISL_2041505, EPI_ISL_2041664, EPI_ISL_2041724, EPI_ISL_2041769, EPI_ISL_2041892, EPI_ISL_2041893, EPI_ISL_2041920, EPI_ISL_2041923, EPI_ISL_2041924, EPI_ISL_2041929, EPI_ISL_2041932, EPI_ISL_2041945, EPI_ISL_2041970, EPI_ISL_2042121, EPI_ISL_2042124, EPI_ISL_2042158, EPI_ISL_2042159, EPI_ISL_2042169, EPI_ISL_2042170, EPI_ISL_2042171, EPI_ISL_2042172, EPI_ISL_2042174, EPI_ISL_2042252, EPI_ISL_2042265, EPI_ISL_2042282, EPI_ISL_2042284, EPI_ISL_2042288, EPI_ISL_2042290, EPI_ISL_2042291, EPI_ISL_2042306, EPI_ISL_2042311, EPI_ISL_2042350, EPI_ISL_2042391, EPI_ISL_2042460, EPI_ISL_2042474, EPI_ISL_2042477, EPI_ISL_2042569, EPI_ISL_2042597, EPI_ISL_2042670, EPI_ISL_2042684, EPI_ISL_2042707, EPI_ISL_2042711, EPI_ISL_2042725, EPI_ISL_2042735, EPI_ISL_2042741, EPI_ISL_2042760, EPI_ISL_2042793, EPI_ISL_2042865, EPI_ISL_2042867, EPI_ISL_2043000, EPI_ISL_2043178, EPI_ISL_2043182, EPI_ISL_2043186, EPI_ISL_2043194, EPI_ISL_2043220, EPI_ISL_2043221, EPI_ISL_2043251, EPI_ISL_2043268, EPI_ISL_2043311, EPI_ISL_2043349, EPI_ISL_2043456, EPI_ISL_2043469, EPI_ISL_2043490, EPI_ISL_2043535, EPI_ISL_2089472, EPI_ISL_2089489, EPI_ISL_2089506, EPI_ISL_2089515, EPI_ISL_2089572, EPI_ISL_2089676, EPI_ISL_2089782, EPI_ISL_2089786, EPI_ISL_2089852, EPI_ISL_2089937, EPI_ISL_2089944, EPI_ISL_2089946, EPI_ISL_2089949, EPI_ISL_2089998, EPI_ISL_2090009, EPI_ISL_2090113, EPI_ISL_2090118, EPI_ISL_2090145, EPI_ISL_2090146, EPI_ISL_2090147, EPI_ISL_2090148, EPI_ISL_2090149, EPI_ISL_2090150, EPI_ISL_2090152, EPI_ISL_2090168, EPI_ISL_2090212, EPI_ISL_2090213, EPI_ISL_2090214, EPI_ISL_2090215, EPI_ISL_2090252, EPI_ISL_2090423, EPI_ISL_2090424, EPI_ISL_2090425, EPI_ISL_2090440, EPI_ISL_2090447, EPI_ISL_2090457, EPI_ISL_2090462, EPI_ISL_2090463, EPI_ISL_2090467, EPI_ISL_2132539, EPI_ISL_2132555, EPI_ISL_2132571, EPI_ISL_2145813, EPI_ISL_2145876, EPI_ISL_2145884, EPI_ISL_2145965, EPI_ISL_2145971, EPI_ISL_2146044, EPI_ISL_2146054, EPI_ISL_2146076, EPI_ISL_2146184, EPI_ISL_2146260, EPI_ISL_2146261, EPI_ISL_2146290, EPI_ISL_2146301, EPI_ISL_2146315, EPI_ISL_2146320, EPI_ISL_2146382, EPI_ISL_2146413, EPI_ISL_2146445, EPI_ISL_2146490, EPI_ISL_2146499, EPI_ISL_2146520, EPI_ISL_2146644, EPI_ISL_2146684, EPI_ISL_2146709, EPI_ISL_2146710, EPI_ISL_2146776, EPI_ISL_2146784, EPI_ISL_2146786, EPI_ISL_2146812, EPI_ISL_2146827, EPI_ISL_2146860, EPI_ISL_2146864, EPI_ISL_2146876, EPI_ISL_2146911, EPI_ISL_2147014, EPI_ISL_2147164, EPI_ISL_2147424, EPI_ISL_2147425, EPI_ISL_2147444, EPI_ISL_2147627, EPI_ISL_2147628, EPI_ISL_2147648, EPI_ISL_2147667, EPI_ISL_2147675, EPI_ISL_2147678, EPI_ISL_2147941, EPI_ISL_2147970, EPI_ISL_2147972, EPI_ISL_2147979, EPI_ISL_2147984, EPI_ISL_2147993, EPI_ISL_2148068, EPI_ISL_2148074, EPI_ISL_2148122, EPI_ISL_2148123, EPI_ISL_2148124, EPI_ISL_2148125, EPI_ISL_2148196, EPI_ISL_2148245, EPI_ISL_2148246, EPI_ISL_2148250, EPI_ISL_2148278, EPI_ISL_2148281, EPI_ISL_2148335, EPI_ISL_2148351, EPI_ISL_2148354, EPI_ISL_2148355, EPI_ISL_2148358, EPI_ISL_2148359, EPI_ISL_2148363, EPI_ISL_2148364, EPI_ISL_2148365, EPI_ISL_2148366, EPI_ISL_2148367, EPI_ISL_2148368, EPI_ISL_2148369, EPI_ISL_2148370, EPI_ISL_2148371, EPI_ISL_2148372, EPI_ISL_2148373, EPI_ISL_2148374, EPI_ISL_2148435, EPI_ISL_2148616, EPI_ISL_2148618, EPI_ISL_2148631, EPI_ISL_2148715, EPI_ISL_2148724, EPI_ISL_2148725, EPI_ISL_2148726, EPI_ISL_2148919, EPI_ISL_2148920, EPI_ISL_2149220, EPI_ISL_2149304, EPI_ISL_2149336, EPI_ISL_2149337, EPI_ISL_2149338, EPI_ISL_2149339, EPI_ISL_2149382, EPI_ISL_2149390, EPI_ISL_2149442, EPI_ISL_2149456, EPI_ISL_2149472, EPI_ISL_2149588, EPI_ISL_2149589, EPI_ISL_2149595, EPI_ISL_2149599, EPI_ISL_2149603, EPI_ISL_2149608, EPI_ISL_2149624, EPI_ISL_2149662, EPI_ISL_2149688, EPI_ISL_2149765, EPI_ISL_2149817, EPI_ISL_2149969, EPI_ISL_2149975, EPI_ISL_2149977, EPI_ISL_2150159, EPI_ISL_2150213, EPI_ISL_2150218, EPI_ISL_2150254, EPI_ISL_2150324, EPI_ISL_2150333, EPI_ISL_2150363, EPI_ISL_2150366, EPI_ISL_2150376, EPI_ISL_2150379, EPI_ISL_2150411, EPI_ISL_2150417, EPI_ISL_2150449, EPI_ISL_2150457, EPI_ISL_2150470, EPI_ISL_2150501, EPI_ISL_2150509, EPI_ISL_2150533, EPI_ISL_2150561, EPI_ISL_2150677, EPI_ISL_2150679, EPI_ISL_2150683, EPI_ISL_2150716, EPI_ISL_2150745, EPI_ISL_2150746, EPI_ISL_2150747, EPI_ISL_2150815, EPI_ISL_2150870, EPI_ISL_2150881, EPI_ISL_2150898, EPI_ISL_2150902, EPI_ISL_2150911, EPI_ISL_2150967, EPI_ISL_2158957, EPI_ISL_2159052, EPI_ISL_2159135, EPI_ISL_2159152, EPI_ISL_2159174, EPI_ISL_2159265, EPI_ISL_2159436, EPI_ISL_2159456, EPI_ISL_2180540, EPI_ISL_2180577, EPI_ISL_2180578, EPI_ISL_2180580, EPI_ISL_2180719, EPI_ISL_2180731, EPI_ISL_2180734, EPI_ISL_2180794, EPI_ISL_2180876, EPI_ISL_2181007, EPI_ISL_2181045, EPI_ISL_2181052, EPI_ISL_2181087, EPI_ISL_2181119, EPI_ISL_2181185, EPI_ISL_2181200, EPI_ISL_2181204, EPI_ISL_2181224, EPI_ISL_2181361, EPI_ISL_2181373, EPI_ISL_2181381, EPI_ISL_2181423, EPI_ISL_2181433, EPI_ISL_2181434, EPI_ISL_2181446, EPI_ISL_2181482, EPI_ISL_2181532, EPI_ISL_2185635, EPI_ISL_2185636, EPI_ISL_2185639, EPI_ISL_2185650, EPI_ISL_2185672, EPI_ISL_2185691, EPI_ISL_2185694, EPI_ISL_2185833, EPI_ISL_2185836, EPI_ISL_2185856, EPI_ISL_2185860, EPI_ISL_2185861, EPI_ISL_2185864, EPI_ISL_2185964, EPI_ISL_2186015, EPI_ISL_2186103, EPI_ISL_2186108, EPI_ISL_2186109, EPI_ISL_2186112, EPI_ISL_2186114, EPI_ISL_2186118, EPI_ISL_2186128, EPI_ISL_2186134, EPI_ISL_2186146, EPI_ISL_2186156, EPI_ISL_2186160, EPI_ISL_2186161, EPI_ISL_2186178, EPI_ISL_2186216, EPI_ISL_2186242, EPI_ISL_2187374, EPI_ISL_2187377, EPI_ISL_2187381, EPI_ISL_2187382, EPI_ISL_2187440, EPI_ISL_2187472, EPI_ISL_2187496, EPI_ISL_2187542, EPI_ISL_2187556, EPI_ISL_2201237, EPI_ISL_2201289, EPI_ISL_2201486, EPI_ISL_2201566, EPI_ISL_2202112, EPI_ISL_2202114, EPI_ISL_2202115, EPI_ISL_2202304, EPI_ISL_2202367, EPI_ISL_2202375, EPI_ISL_2202376, EPI_ISL_2202377, EPI_ISL_2202403, EPI_ISL_2202615, EPI_ISL_2202624, EPI_ISL_2202699, EPI_ISL_2202711, EPI_ISL_2202712, EPI_ISL_2202840, EPI_ISL_2203090, EPI_ISL_2203140, EPI_ISL_2204549, EPI_ISL_2204595, EPI_ISL_2204604, EPI_ISL_2204854, EPI_ISL_2204968, EPI_ISL_2241664, EPI_ISL_2241670, EPI_ISL_2241720, EPI_ISL_2241783, EPI_ISL_2241813, EPI_ISL_2241869, EPI_ISL_2241940, EPI_ISL_2241941, EPI_ISL_2241942, EPI_ISL_2241959, EPI_ISL_2241978, EPI_ISL_2242022, EPI_ISL_2242024, EPI_ISL_2242025, EPI_ISL_2242057, EPI_ISL_2242079, EPI_ISL_2242162, EPI_ISL_2242165, EPI_ISL_2242179, EPI_ISL_2242194, EPI_ISL_2242281, EPI_ISL_2242282, EPI_ISL_2242290, EPI_ISL_2242305, EPI_ISL_2242341, EPI_ISL_2242349, EPI_ISL_2242350, EPI_ISL_2242351, EPI_ISL_2242353, EPI_ISL_2242379, EPI_ISL_2242403, EPI_ISL_2242430, EPI_ISL_2242434, EPI_ISL_2242438, EPI_ISL_2242462, EPI_ISL_2242463, EPI_ISL_2242465, EPI_ISL_2242483, EPI_ISL_2242522, EPI_ISL_2242524, EPI_ISL_2242613, EPI_ISL_2242627, EPI_ISL_2242718, EPI_ISL_2242721, EPI_ISL_2242722, EPI_ISL_2242728, EPI_ISL_2242735, EPI_ISL_2242763, EPI_ISL_2242841, EPI_ISL_2242864, EPI_ISL_2242884, EPI_ISL_2242903, EPI_ISL_2242915, EPI_ISL_2242949, EPI_ISL_2242973, EPI_ISL_2242991, EPI_ISL_2243045, EPI_ISL_2243116, EPI_ISL_2243126, EPI_ISL_2243130, EPI_ISL_2243153, EPI_ISL_2243175, EPI_ISL_2243176, EPI_ISL_2243248, EPI_ISL_2243266, EPI_ISL_2243324, EPI_ISL_2243333, EPI_ISL_2243342, EPI_ISL_2243369, EPI_ISL_2243419, EPI_ISL_2243420, EPI_ISL_2243425, EPI_ISL_2243437, EPI_ISL_2243453, EPI_ISL_2243456, EPI_ISL_2243470, EPI_ISL_2243477, EPI_ISL_2243621, EPI_ISL_2243623, EPI_ISL_2243663, EPI_ISL_2243664, EPI_ISL_2243684, EPI_ISL_2243743, EPI_ISL_2243785, EPI_ISL_2243802, EPI_ISL_2243816, EPI_ISL_2243941, EPI_ISL_2243951, EPI_ISL_2243973, EPI_ISL_2243982, EPI_ISL_2244022, EPI_ISL_2244024, EPI_ISL_2244045, EPI_ISL_2244073, EPI_ISL_2244084, EPI_ISL_2244101, EPI_ISL_2244154, EPI_ISL_2244164, EPI_ISL_2244256, EPI_ISL_2244260, EPI_ISL_2244267, EPI_ISL_2244283, EPI_ISL_2244287, EPI_ISL_2244311, EPI_ISL_2244371, EPI_ISL_2244402, EPI_ISL_2244403, EPI_ISL_2244409, EPI_ISL_2244417, EPI_ISL_2244448, EPI_ISL_2244463, EPI_ISL_2244464, EPI_ISL_2244499, EPI_ISL_2244569, EPI_ISL_2244586, EPI_ISL_2244649, EPI_ISL_2244673, EPI_ISL_2244721, EPI_ISL_2244799, EPI_ISL_2244824, EPI_ISL_2244825, EPI_ISL_2244826, EPI_ISL_2244893, EPI_ISL_2268035, EPI_ISL_2268065, EPI_ISL_2268256, EPI_ISL_2280402, EPI_ISL_2280403, EPI_ISL_2280404, EPI_ISL_2280405, EPI_ISL_2280553, EPI_ISL_2280640, EPI_ISL_2280722, EPI_ISL_2280734, EPI_ISL_2280735, EPI_ISL_2280762, EPI_ISL_2280793, EPI_ISL_2280815, EPI_ISL_2280880, EPI_ISL_2280902, EPI_ISL_2280949, EPI_ISL_2281000, EPI_ISL_2281029, EPI_ISL_2281046, EPI_ISL_2283842, EPI_ISL_2283843, EPI_ISL_2283865, EPI_ISL_2283884, EPI_ISL_2321257, EPI_ISL_2321262, EPI_ISL_2321368, EPI_ISL_2368289, EPI_ISL_2368290, EPI_ISL_2368334, EPI_ISL_2368337, EPI_ISL_2368350, EPI_ISL_2368359, EPI_ISL_2368362, EPI_ISL_2368455, EPI_ISL_2368806, EPI_ISL_2368826, EPI_ISL_2368827, EPI_ISL_2368909, EPI_ISL_2369947, EPI_ISL_2370020, EPI_ISL_2370042, EPI_ISL_2370172, EPI_ISL_2370219, EPI_ISL_2370221, EPI_ISL_2370235, EPI_ISL_2370437, EPI_ISL_2370538, EPI_ISL_2370562, EPI_ISL_2370564, EPI_ISL_2370566, EPI_ISL_2370571, EPI_ISL_2370589, EPI_ISL_2370595, EPI_ISL_2421385, EPI_ISL_2421387, EPI_ISL_2421435, EPI_ISL_2421602, EPI_ISL_2421630, EPI_ISL_2421678, EPI_ISL_2421681, EPI_ISL_2421778, EPI_ISL_2421779, EPI_ISL_2421803, EPI_ISL_2421814, EPI_ISL_2422005, EPI_ISL_2482339, EPI_ISL_2482343, EPI_ISL_2482398, EPI_ISL_2482399, EPI_ISL_2489118, EPI_ISL_2489190, EPI_ISL_2489191, EPI_ISL_2489209, EPI_ISL_2489212, EPI_ISL_2489213, EPI_ISL_2489233, EPI_ISL_2489312, EPI_ISL_2489340, EPI_ISL_2489361, EPI_ISL_2489395, EPI_ISL_2489456, EPI_ISL_2489480, EPI_ISL_2489494, EPI_ISL_2489519, EPI_ISL_2489569, EPI_ISL_2489571, EPI_ISL_2489576, EPI_ISL_2489587, EPI_ISL_2489644, EPI_ISL_2489713, EPI_ISL_2489765, EPI_ISL_2489846, EPI_ISL_2489884, EPI_ISL_2527893, EPI_ISL_2527895, EPI_ISL_2527950, EPI_ISL_2528214, EPI_ISL_2528360, EPI_ISL_2528504, EPI_ISL_2528515, EPI_ISL_2528550, EPI_ISL_2528879, EPI_ISL_2528949, EPI_ISL_2528955, EPI_ISL_2529005, EPI_ISL_2531023, EPI_ISL_2531089, EPI_ISL_2534535, EPI_ISL_2599943, EPI_ISL_2599969, EPI_ISL_2599988, EPI_ISL_2599990, EPI_ISL_2600012, EPI_ISL_2600035, EPI_ISL_2600053, EPI_ISL_2600094, EPI_ISL_2600117, EPI_ISL_2600137, EPI_ISL_2600236, EPI_ISL_2600286, EPI_ISL_2686916, EPI_ISL_2686941, EPI_ISL_2687016, EPI_ISL_2687044, EPI_ISL_2687087, EPI_ISL_2687251, EPI_ISL_2687252, EPI_ISL_2687475, EPI_ISL_2687484, EPI_ISL_2709513, EPI_ISL_2784860, EPI_ISL_2785307, EPI_ISL_2785330, EPI_ISL_2785331, EPI_ISL_2785514, EPI_ISL_2875851, EPI_ISL_2875901, EPI_ISL_2875902, EPI_ISL_2875903, EPI_ISL_2875924, EPI_ISL_3217278, EPI_ISL_3217379, EPI_ISL_3217478, EPI_ISL_3217507, EPI_ISL_3217863, EPI_ISL_3218073, EPI_ISL_3218116, EPI_ISL_3218126, EPI_ISL_3218145, EPI_ISL_3218417, EPI_ISL_3218494, EPI_ISL_3218553, EPI_ISL_3218768, EPI_ISL_3218791, EPI_ISL_3218792, EPI_ISL_3219953, EPI_ISL_3220003, EPI_ISL_3220024, EPI_ISL_3220215, EPI_ISL_3220216, EPI_ISL_3220375, EPI_ISL_3220378, EPI_ISL_3220422, EPI_ISL_3220541, EPI_ISL_3220640, EPI_ISL_3220803, EPI_ISL_3220969, EPI_ISL_3305294, EPI_ISL_3305374, EPI_ISL_3305397, EPI_ISL_3305488, EPI_ISL_3321501, EPI_ISL_3321544, EPI_ISL_3321563, EPI_ISL_3321752, EPI_ISL_3321847, EPI_ISL_3321871, EPI_ISL_3321873, EPI_ISL_3321972, EPI_ISL_3321973, EPI_ISL_3322001, EPI_ISL_3322002, EPI_ISL_3322033, EPI_ISL_3322050, EPI_ISL_3322092, EPI_ISL_3322140, EPI_ISL_3322381, EPI_ISL_3322391, EPI_ISL_3322412, EPI_ISL_3322444, EPI_ISL_3322548, EPI_ISL_3322550, EPI_ISL_3322665, EPI_ISL_3322744, EPI_ISL_3323028, EPI_ISL_3323036, EPI_ISL_3323094, EPI_ISL_3323159, EPI_ISL_3323472, EPI_ISL_3323473, EPI_ISL_3323671, EPI_ISL_3323682, EPI_ISL_3323683, EPI_ISL_3323705, EPI_ISL_3323821, EPI_ISL_3324108, EPI_ISL_3324289, EPI_ISL_3324351, EPI_ISL_3324355, EPI_ISL_3324412, EPI_ISL_3324416, EPI_ISL_3324475, EPI_ISL_3324500, EPI_ISL_3324681, EPI_ISL_3325031, EPI_ISL_3325130, EPI_ISL_3325198 | | | |
| see above | Aegis Sciences Corporation | Centers for Disease Control and Prevention Division of Viral Diseases, Pathogen Discovery | Adrian Paskey; Alec Vest; Benjamin Rambo-Martin; Christopher Gulvick; Clinton R. Paden; Cyndi Clark; Dakota Howard; Darlene Wagner; Dhwani Batra; Dillon Nall; Duncan MacCannell; Ethan Sanders; Holly Houdeshell; Jason Caravas; Kara Moser; Matthew Hardison; Matthew Schmerer; Ola Kvalvaag; Patrick Campbell; Peter W. Cook; Rob Case; Scott Sammons; Shatavia Morrison; Shaun Westlund; Vikramsinha Ghorpade; Yvette Unoarumhi |
| EPI_ISL_3016318, EPI_ISL_3016319 | Boston University CTL | Boston Universit/National Emerging Infectious Disease Laboratories | Catherine Klapperich; Jacquelyn Turcinovic; John H. Connor; Lena Landeverde; Lynn Doucette-Stamm |
| EPI_ISL_2991502, EPI_ISL_2991503, EPI_ISL_2991504, EPI_ISL_2991505, EPI_ISL_2991506, EPI_ISL_2991507, EPI_ISL_2991509, EPI_ISL_2991510, EPI_ISL_2991511, EPI_ISL_2991512, EPI_ISL_2991513, EPI_ISL_2991515, EPI_ISL_2991516, EPI_ISL_2991517, EPI_ISL_2991519, EPI_ISL_2991520, EPI_ISL_2991521, EPI_ISL_2991522, EPI_ISL_2991523, EPI_ISL_2991524, EPI_ISL_2991526, EPI_ISL_2991527, EPI_ISL_2991528, EPI_ISL_2991529, EPI_ISL_2991530, EPI_ISL_2991531, EPI_ISL_2991532, EPI_ISL_2991533, EPI_ISL_2991534, EPI_ISL_2991535, EPI_ISL_2991536, EPI_ISL_2991537, EPI_ISL_2991538, EPI_ISL_2991539, EPI_ISL_2991540, EPI_ISL_2991541, EPI_ISL_2991542, EPI_ISL_2991543, EPI_ISL_2991544, EPI_ISL_2991545, EPI_ISL_2991546, EPI_ISL_2991547, EPI_ISL_2991548, EPI_ISL_2991549, EPI_ISL_2991550, EPI_ISL_2991551, EPI_ISL_2991552, EPI_ISL_2991553, EPI_ISL_2991555, EPI_ISL_2991556, EPI_ISL_2991557, EPI_ISL_2991558, EPI_ISL_2991561, EPI_ISL_2991562, EPI_ISL_2991563, EPI_ISL_2991564, EPI_ISL_2991565, EPI_ISL_2991566, EPI_ISL_2991568, EPI_ISL_2991569, EPI_ISL_2991570, EPI_ISL_2991571, EPI_ISL_2991572, EPI_ISL_2991573, EPI_ISL_2991574, EPI_ISL_2991575, EPI_ISL_2991576, EPI_ISL_2991577, EPI_ISL_2991578, EPI_ISL_2991579, EPI_ISL_2991580, EPI_ISL_2991581, EPI_ISL_2991582, EPI_ISL_2991583, EPI_ISL_2991584, EPI_ISL_2991586, EPI_ISL_2991587, EPI_ISL_2991588, EPI_ISL_2991589, EPI_ISL_2991590, EPI_ISL_2991591, EPI_ISL_2991592, EPI_ISL_2991593, EPI_ISL_2991594, EPI_ISL_2991595, EPI_ISL_2991596, EPI_ISL_2991597, EPI_ISL_2991599, EPI_ISL_2991600, EPI_ISL_2991602, EPI_ISL_2991603, EPI_ISL_2991604, EPI_ISL_2991605, EPI_ISL_2991606, EPI_ISL_2991607, EPI_ISL_2991608, EPI_ISL_2991609, EPI_ISL_2991611, EPI_ISL_2991612, EPI_ISL_2991613, EPI_ISL_2991614, EPI_ISL_2991615, EPI_ISL_2991616, EPI_ISL_2991617, EPI_ISL_2991618, EPI_ISL_2991619, EPI_ISL_2991621, EPI_ISL_2991622, EPI_ISL_2991623, EPI_ISL_2991624, EPI_ISL_2991625, EPI_ISL_2991626, EPI_ISL_2991628, EPI_ISL_2991629, EPI_ISL_2991630, EPI_ISL_2991631, EPI_ISL_2991632, EPI_ISL_2991633, EPI_ISL_2991634, EPI_ISL_2991635, EPI_ISL_2991636, EPI_ISL_2991637, EPI_ISL_2991639, EPI_ISL_2991641, EPI_ISL_2991642, EPI_ISL_2991643, EPI_ISL_2991644, EPI_ISL_2991645, EPI_ISL_2991646, EPI_ISL_2991647, EPI_ISL_2991648, EPI_ISL_2991649, EPI_ISL_2991651, EPI_ISL_2991652, EPI_ISL_2991653, EPI_ISL_2991655, EPI_ISL_2991656, EPI_ISL_2991657, EPI_ISL_2991658, EPI_ISL_2991659, EPI_ISL_2991660, EPI_ISL_2991661, EPI_ISL_2991662, EPI_ISL_2991663, EPI_ISL_2991664, EPI_ISL_2991665, EPI_ISL_2991666, EPI_ISL_2991667, EPI_ISL_2991668, EPI_ISL_2991669, EPI_ISL_2991670, EPI_ISL_2991673, EPI_ISL_2991674, EPI_ISL_2991675, EPI_ISL_2991676, EPI_ISL_2991677, EPI_ISL_2991678, EPI_ISL_2991679, EPI_ISL_2991680, EPI_ISL_2991681, EPI_ISL_2991682, EPI_ISL_2991683, EPI_ISL_2991684, EPI_ISL_2991685, EPI_ISL_2991686, EPI_ISL_2991687, EPI_ISL_2991689, EPI_ISL_2991690, EPI_ISL_2991691, EPI_ISL_2991692, EPI_ISL_2991693, EPI_ISL_2991694, EPI_ISL_2991695, EPI_ISL_2991696, EPI_ISL_2991697, EPI_ISL_2991699, EPI_ISL_2991700, EPI_ISL_2991701, EPI_ISL_2991702, EPI_ISL_2991703, EPI_ISL_2991705, EPI_ISL_2991706, EPI_ISL_2991707, EPI_ISL_2991708, EPI_ISL_2991709, EPI_ISL_2991710, EPI_ISL_2991711, EPI_ISL_2991712, EPI_ISL_2991713, EPI_ISL_2991714, EPI_ISL_2991715, EPI_ISL_2991716, EPI_ISL_2991717, EPI_ISL_2991718, EPI_ISL_2991719, EPI_ISL_2991720, EPI_ISL_2991721, EPI_ISL_2991722, EPI_ISL_2991723, EPI_ISL_2991724, EPI_ISL_2991725, EPI_ISL_2991726, EPI_ISL_2991727, EPI_ISL_2991728, EPI_ISL_2991729, EPI_ISL_2991730, EPI_ISL_2991731, EPI_ISL_2991732, EPI_ISL_2991733, EPI_ISL_2991736, EPI_ISL_2991737, EPI_ISL_2991738, EPI_ISL_2991739, EPI_ISL_2991740, EPI_ISL_2991741, EPI_ISL_2991742, EPI_ISL_2991743, EPI_ISL_2991744, EPI_ISL_2991745, EPI_ISL_2991746, EPI_ISL_2991747, EPI_ISL_2991748, EPI_ISL_2991750, EPI_ISL_2991752, EPI_ISL_2991753, EPI_ISL_2991754, EPI_ISL_2991755, EPI_ISL_2991756, EPI_ISL_2991757, EPI_ISL_2991758, EPI_ISL_2991759, EPI_ISL_2991761, EPI_ISL_2991763, EPI_ISL_2991764, EPI_ISL_2991765, EPI_ISL_2991767, EPI_ISL_2991768, EPI_ISL_2991769, EPI_ISL_2991770, EPI_ISL_2991771, EPI_ISL_2991772, EPI_ISL_2991773, EPI_ISL_2991774, EPI_ISL_2991775, EPI_ISL_2991777, EPI_ISL_2991778, EPI_ISL_2991779, EPI_ISL_2991780, EPI_ISL_2991781, EPI_ISL_2991783, EPI_ISL_2991784, EPI_ISL_2991785, EPI_ISL_2991786, EPI_ISL_2991787, EPI_ISL_2991788, EPI_ISL_2991789, EPI_ISL_2991790, EPI_ISL_2991791, EPI_ISL_2991792, EPI_ISL_2991793, EPI_ISL_2991794, EPI_ISL_2991795, EPI_ISL_2991796, EPI_ISL_2991797, EPI_ISL_2991799, EPI_ISL_2991800, EPI_ISL_2991801, EPI_ISL_2991802, EPI_ISL_2991803, EPI_ISL_2991804, EPI_ISL_2991805, EPI_ISL_2991806, EPI_ISL_2991807, EPI_ISL_2991808, EPI_ISL_2991810, EPI_ISL_2991811, EPI_ISL_2991812, EPI_ISL_2991813, EPI_ISL_2991814, EPI_ISL_2991815, EPI_ISL_2991816, EPI_ISL_2991817, EPI_ISL_2991818, EPI_ISL_2991819, EPI_ISL_2991820, EPI_ISL_2991821, EPI_ISL_2991822, EPI_ISL_2991823, EPI_ISL_2991824, EPI_ISL_2991825, EPI_ISL_2991826, EPI_ISL_2991827, EPI_ISL_2991828, EPI_ISL_2991829, EPI_ISL_2991830, EPI_ISL_2991831, EPI_ISL_2991833, EPI_ISL_2991834, EPI_ISL_2991835, EPI_ISL_2991836, EPI_ISL_2991837, EPI_ISL_2991838, EPI_ISL_2991839, EPI_ISL_2991840, EPI_ISL_2991841, EPI_ISL_2991842, EPI_ISL_2991843, EPI_ISL_2991844, EPI_ISL_2991845, EPI_ISL_2991846, EPI_ISL_2991847, EPI_ISL_2991849, EPI_ISL_2991850, EPI_ISL_2991852, EPI_ISL_2991853, EPI_ISL_2991854, EPI_ISL_2991855, EPI_ISL_2991856, EPI_ISL_2991857, EPI_ISL_2991858, EPI_ISL_2991859, EPI_ISL_2991860, EPI_ISL_2991862, EPI_ISL_2991863, EPI_ISL_2991864, EPI_ISL_2991865, EPI_ISL_2991866, EPI_ISL_2991867, EPI_ISL_2991868, EPI_ISL_2991869, EPI_ISL_2991871, EPI_ISL_2991872, EPI_ISL_2991873, EPI_ISL_2991874, EPI_ISL_2991875, EPI_ISL_2991876, EPI_ISL_2991877, EPI_ISL_2991878, EPI_ISL_2991879, EPI_ISL_2991880, EPI_ISL_2991881, EPI_ISL_2991882, EPI_ISL_2991883, EPI_ISL_2991884, EPI_ISL_2991885, EPI_ISL_2991886, EPI_ISL_2991887, EPI_ISL_2991888, EPI_ISL_2991889, EPI_ISL_2991890, EPI_ISL_2991892, EPI_ISL_2991893, EPI_ISL_2991894, EPI_ISL_2991895, EPI_ISL_2991896, EPI_ISL_2991897, EPI_ISL_2991898, EPI_ISL_2991899, EPI_ISL_2991900, EPI_ISL_2991901, EPI_ISL_2991902, EPI_ISL_2991903, EPI_ISL_2991904, EPI_ISL_2991905, EPI_ISL_2991906, EPI_ISL_2991907, EPI_ISL_2991908, EPI_ISL_2991909, EPI_ISL_2991911, EPI_ISL_2991913, EPI_ISL_2991914, EPI_ISL_2991915, EPI_ISL_2991916, EPI_ISL_2991917, EPI_ISL_2991918, EPI_ISL_2991919, EPI_ISL_2991920, EPI_ISL_2991921, EPI_ISL_2991922, EPI_ISL_2991923, EPI_ISL_2991924, EPI_ISL_2991925, EPI_ISL_2991926, EPI_ISL_2991927, EPI_ISL_2991928, EPI_ISL_2991929, EPI_ISL_2991930, EPI_ISL_2991931, EPI_ISL_2991932, EPI_ISL_2991933, EPI_ISL_2991934, EPI_ISL_2991935, EPI_ISL_2991937, EPI_ISL_2991938, EPI_ISL_2991939, EPI_ISL_2991940, EPI_ISL_2991941, EPI_ISL_2991942, EPI_ISL_2991943, EPI_ISL_2991944 | | | |
| see above | Broad Institute Clinical Research Sequencing Platform | Infectious Disease Program, Broad Institute of Harvard and MIT | A.E.; Adams, G.; Anahtar, M.; B.L.; B.W.; Bauer, M.; Birren; Branda, J.; Carter, A.; Cerrato, F.; Chaluvadi, S.; Chapman; Cusick, C.; D.J.; DeRuff, K.; Flowers, K.; Gallagher, G.; Gladden-Young, A.; Gnirke, A.; Harris, J.; J.E.; K.J.; LaRocque, R.; Lagerborg, K.; Lemieux; Lin; Loreth, C.; MacInnis; Neumann, A.; Normandin, E.; P.C.; Park; Pierce, V.; Reilly, S.; Rosenberg, E.; Rudy, M.; Ryan, E.; S.B.; Sabeti; Shaw, B.; Siddle; Slater, D.; Smole, S.; Tomkins-Tinch, C.; Turbett, S. |
| EPI_ISL_2016384, EPI_ISL_2016385, EPI_ISL_2016386, EPI_ISL_2016387, EPI_ISL_2016388, EPI_ISL_2016389, EPI_ISL_2016390, EPI_ISL_2016391, EPI_ISL_2016392, EPI_ISL_2016393, EPI_ISL_2016394, EPI_ISL_2016395, EPI_ISL_2016396, EPI_ISL_2016397, EPI_ISL_2016398, EPI_ISL_2016399, EPI_ISL_2016400, EPI_ISL_2016402, EPI_ISL_2016403, EPI_ISL_2016404, EPI_ISL_2016406, EPI_ISL_2016407, EPI_ISL_2016408, EPI_ISL_2016409, EPI_ISL_2016410, EPI_ISL_2016411, EPI_ISL_2016412, EPI_ISL_2016413, EPI_ISL_2016414, EPI_ISL_2016415, EPI_ISL_2016416, EPI_ISL_2016417, EPI_ISL_2016418, EPI_ISL_2016419, EPI_ISL_2016420, EPI_ISL_2016421, EPI_ISL_2016422, EPI_ISL_2016423, EPI_ISL_2016424, EPI_ISL_2016425, EPI_ISL_2016426, EPI_ISL_2016427, EPI_ISL_2016429, EPI_ISL_2016430, EPI_ISL_2016431, EPI_ISL_2016432, EPI_ISL_2016433, EPI_ISL_2016434, EPI_ISL_2016435, EPI_ISL_2016436, EPI_ISL_2016437, EPI_ISL_2016438, EPI_ISL_2096228, EPI_ISL_2096229, EPI_ISL_2096230, EPI_ISL_2096231, EPI_ISL_2096232, EPI_ISL_2096234, EPI_ISL_2096235, EPI_ISL_2096236, EPI_ISL_2096237, EPI_ISL_2096238, EPI_ISL_2096239, EPI_ISL_2096240, EPI_ISL_2096241, EPI_ISL_2096242, EPI_ISL_2096243, EPI_ISL_2096244, EPI_ISL_2096245, EPI_ISL_2096246, EPI_ISL_2096248, EPI_ISL_2096250, EPI_ISL_2096251, EPI_ISL_2096252, EPI_ISL_2096253, EPI_ISL_2096254, EPI_ISL_2096255, EPI_ISL_2096256, EPI_ISL_2096258, EPI_ISL_2096259, EPI_ISL_2096260, EPI_ISL_2096262, EPI_ISL_2096263, EPI_ISL_2096264, EPI_ISL_2096265, EPI_ISL_2096266, EPI_ISL_2096267, EPI_ISL_2096268, EPI_ISL_2096269, EPI_ISL_2096270, EPI_ISL_2096272, EPI_ISL_2096273, EPI_ISL_2096274, EPI_ISL_2096275, EPI_ISL_2096276, EPI_ISL_2096277, EPI_ISL_2096278, EPI_ISL_2096279, EPI_ISL_2096282, EPI_ISL_2096283, EPI_ISL_2096284, EPI_ISL_2096285, EPI_ISL_2096286, EPI_ISL_2096287, EPI_ISL_2096288, EPI_ISL_2096289, EPI_ISL_2096291, EPI_ISL_2096292, EPI_ISL_2096293, EPI_ISL_2096294, EPI_ISL_2096295, EPI_ISL_2096296, EPI_ISL_2096297, EPI_ISL_2096298, EPI_ISL_2096299, EPI_ISL_2096300, EPI_ISL_2096301, EPI_ISL_2096303, EPI_ISL_2096304, EPI_ISL_2096305, EPI_ISL_2096306, EPI_ISL_2096307, EPI_ISL_2096308, EPI_ISL_2096309, EPI_ISL_2096310, EPI_ISL_2096313, EPI_ISL_2096314, EPI_ISL_2096315, EPI_ISL_2096317, EPI_ISL_2096319, EPI_ISL_2096320, EPI_ISL_2096321, EPI_ISL_2096322, EPI_ISL_2096323, EPI_ISL_2096325, EPI_ISL_2096327, EPI_ISL_2096328, EPI_ISL_2096329, EPI_ISL_2096330, EPI_ISL_2096332, EPI_ISL_2096333, EPI_ISL_2096334, EPI_ISL_2096335, EPI_ISL_2096336, EPI_ISL_2096337, EPI_ISL_2096338, EPI_ISL_2096339, EPI_ISL_2096340, EPI_ISL_2096341, EPI_ISL_2096342, EPI_ISL_2096343, EPI_ISL_2096344, EPI_ISL_2096346, EPI_ISL_2096347, EPI_ISL_2096348, EPI_ISL_2096349, EPI_ISL_2096350, EPI_ISL_2096351, EPI_ISL_2096352, EPI_ISL_2096353, EPI_ISL_2096354, EPI_ISL_2096356, EPI_ISL_2096358, EPI_ISL_2096359, EPI_ISL_2096360, EPI_ISL_2096362, EPI_ISL_2096363, EPI_ISL_2096364, EPI_ISL_2096366, EPI_ISL_2096367, EPI_ISL_2096368, EPI_ISL_2096369, EPI_ISL_2096370, EPI_ISL_2096371, EPI_ISL_2096373, EPI_ISL_2096374, EPI_ISL_2096375, EPI_ISL_2096376, EPI_ISL_2096377, EPI_ISL_2096378, EPI_ISL_2096379, EPI_ISL_2096381, EPI_ISL_2096383, EPI_ISL_2096384, EPI_ISL_2096385, EPI_ISL_2096388, EPI_ISL_2096389, EPI_ISL_2096390, EPI_ISL_2096391, EPI_ISL_2096393, EPI_ISL_2096394, EPI_ISL_2096395, EPI_ISL_2096396, EPI_ISL_2096397, EPI_ISL_2096398, EPI_ISL_2096399, EPI_ISL_2096400, EPI_ISL_2096401, EPI_ISL_2096402, EPI_ISL_2096403, EPI_ISL_2096404, EPI_ISL_2096405, EPI_ISL_2096406, EPI_ISL_2096408, EPI_ISL_2096409, EPI_ISL_2096411, EPI_ISL_2096412, EPI_ISL_2096413, EPI_ISL_2096414, EPI_ISL_2096416, EPI_ISL_2096417, EPI_ISL_2096418, EPI_ISL_2096419, EPI_ISL_2096420, EPI_ISL_2096421, EPI_ISL_2096423, EPI_ISL_2096424, EPI_ISL_2096425, EPI_ISL_2096426, EPI_ISL_2096427, EPI_ISL_2096428, EPI_ISL_2096429, EPI_ISL_2096430, EPI_ISL_2096431, EPI_ISL_2096432, EPI_ISL_2096433, EPI_ISL_2096434, EPI_ISL_2096435, EPI_ISL_2096436, EPI_ISL_2096437, EPI_ISL_2096438, EPI_ISL_2096440, EPI_ISL_2096441, EPI_ISL_2096442, EPI_ISL_2096443, EPI_ISL_2096444, EPI_ISL_2096445, EPI_ISL_2096446, EPI_ISL_2096447, EPI_ISL_2096448, EPI_ISL_2096449, EPI_ISL_2096450, EPI_ISL_2096452, EPI_ISL_2096453, EPI_ISL_2096454, EPI_ISL_2096455, EPI_ISL_2096457, EPI_ISL_2096458, EPI_ISL_2096459, EPI_ISL_2096461, EPI_ISL_2096462, EPI_ISL_2096463, EPI_ISL_2096464, EPI_ISL_2096465, EPI_ISL_2096466, EPI_ISL_2096468, EPI_ISL_2096469, EPI_ISL_2096470, EPI_ISL_2096471, EPI_ISL_2096473, EPI_ISL_2096474, EPI_ISL_2096475, EPI_ISL_2096476, EPI_ISL_2096478, EPI_ISL_2096479, EPI_ISL_2096480, EPI_ISL_2096481, EPI_ISL_2096482, EPI_ISL_2096483, EPI_ISL_2096484, EPI_ISL_2096485, EPI_ISL_2096487, EPI_ISL_2096488, EPI_ISL_2096489, EPI_ISL_2096490, EPI_ISL_2096491, EPI_ISL_2096492, EPI_ISL_2096493, EPI_ISL_2096494, EPI_ISL_2096495, EPI_ISL_2096497, EPI_ISL_2096498, EPI_ISL_2096500, EPI_ISL_2096501, EPI_ISL_2096502, EPI_ISL_2096503, EPI_ISL_2096504, EPI_ISL_2096505, EPI_ISL_2096506, EPI_ISL_2096507, EPI_ISL_2096509, EPI_ISL_2096510, EPI_ISL_2096511, EPI_ISL_2096512, EPI_ISL_2096513, EPI_ISL_2096514, EPI_ISL_2096515, EPI_ISL_2096517, EPI_ISL_2096518, EPI_ISL_2096520, EPI_ISL_2096521, EPI_ISL_2096522, EPI_ISL_2096523, EPI_ISL_2096524, EPI_ISL_2096525, EPI_ISL_2096526, EPI_ISL_2096528, EPI_ISL_2096529, EPI_ISL_2096530, EPI_ISL_2096531, EPI_ISL_2096532, EPI_ISL_2096533, EPI_ISL_2096535, EPI_ISL_2096536, EPI_ISL_2096537, EPI_ISL_2096538, EPI_ISL_2096539, EPI_ISL_2096540, EPI_ISL_2096541, EPI_ISL_2096542, EPI_ISL_2096543, EPI_ISL_2096544, EPI_ISL_2096545, EPI_ISL_2096546, EPI_ISL_2096547, EPI_ISL_2096548, EPI_ISL_2096549, EPI_ISL_2096550, EPI_ISL_2096551, EPI_ISL_2096552, EPI_ISL_2096553, EPI_ISL_2096554, EPI_ISL_2096555, EPI_ISL_2096556, EPI_ISL_2096557, EPI_ISL_2096558, EPI_ISL_2096560, EPI_ISL_2096562, EPI_ISL_2096563, EPI_ISL_2096564, EPI_ISL_2096565, EPI_ISL_2096567, EPI_ISL_2096569, EPI_ISL_2096571, EPI_ISL_2096572, EPI_ISL_2096574, EPI_ISL_2096575, EPI_ISL_2096576, EPI_ISL_2096577, EPI_ISL_2096578, EPI_ISL_2096579, EPI_ISL_2096580, EPI_ISL_2096581, EPI_ISL_2096582, EPI_ISL_2096583, EPI_ISL_2096584, EPI_ISL_2096586, EPI_ISL_2096587, EPI_ISL_2096588, EPI_ISL_2096590, EPI_ISL_2096591, EPI_ISL_2096592, EPI_ISL_2096593, EPI_ISL_2096594, EPI_ISL_2096595, EPI_ISL_2096596, EPI_ISL_2096597, EPI_ISL_2096598, EPI_ISL_2096599, EPI_ISL_2096600, EPI_ISL_2096601, EPI_ISL_2096602, EPI_ISL_2096603, EPI_ISL_2096604, EPI_ISL_2096605, EPI_ISL_2096606, EPI_ISL_2096607, EPI_ISL_2096608, EPI_ISL_2096609, EPI_ISL_2096610, EPI_ISL_2096612, EPI_ISL_2096613, EPI_ISL_2096614, EPI_ISL_2096616, EPI_ISL_2096617, EPI_ISL_2096618, EPI_ISL_2096619, EPI_ISL_2096620, EPI_ISL_2096621, EPI_ISL_2096622, EPI_ISL_2096623, EPI_ISL_2096624, EPI_ISL_2096625, EPI_ISL_2096626, EPI_ISL_2096627, EPI_ISL_2096628, EPI_ISL_2096629, EPI_ISL_2096630, EPI_ISL_2096631, EPI_ISL_2096632, EPI_ISL_2096633, EPI_ISL_2096634, EPI_ISL_2096635, EPI_ISL_2096637, EPI_ISL_2096638, EPI_ISL_2096639, EPI_ISL_2096640, EPI_ISL_2096641, EPI_ISL_2096642, EPI_ISL_2096643, EPI_ISL_2096644, EPI_ISL_2096645, EPI_ISL_2096646, EPI_ISL_2096647, EPI_ISL_2096648, EPI_ISL_2096649, EPI_ISL_2096651, EPI_ISL_2096652, EPI_ISL_2096653, EPI_ISL_2096654, EPI_ISL_2096655, EPI_ISL_2096656, EPI_ISL_2096657, EPI_ISL_2096658, EPI_ISL_2096659, EPI_ISL_2096661, EPI_ISL_2096662, EPI_ISL_2096663, EPI_ISL_2096664, EPI_ISL_2096665, EPI_ISL_2096666, EPI_ISL_2096667, EPI_ISL_2096668, EPI_ISL_2096669, EPI_ISL_2096670, EPI_ISL_2096671, EPI_ISL_2096672, EPI_ISL_2096673, EPI_ISL_2103835, EPI_ISL_2103836, EPI_ISL_2103837, EPI_ISL_2103838, EPI_ISL_2103839, EPI_ISL_2103840, EPI_ISL_2103841, EPI_ISL_2103842, EPI_ISL_2103843, EPI_ISL_2103844, EPI_ISL_2103845, EPI_ISL_2103846, EPI_ISL_2103848, EPI_ISL_2103849, EPI_ISL_2103850, EPI_ISL_2103851, EPI_ISL_2103852, EPI_ISL_2103854, EPI_ISL_2103855, EPI_ISL_2103856, EPI_ISL_2103857, EPI_ISL_2103858, EPI_ISL_2103859, EPI_ISL_2103860, EPI_ISL_2103861, EPI_ISL_2103862, EPI_ISL_2103863, EPI_ISL_2103864, EPI_ISL_2103866, EPI_ISL_2103867, EPI_ISL_2103868, EPI_ISL_2103869, EPI_ISL_2103870, EPI_ISL_2103871, EPI_ISL_2103872, EPI_ISL_2103873, EPI_ISL_2103874, EPI_ISL_2103875, EPI_ISL_2103876, EPI_ISL_2103878, EPI_ISL_2103879, EPI_ISL_2103880, EPI_ISL_2103882, EPI_ISL_2103883, EPI_ISL_2103884, EPI_ISL_2103885, EPI_ISL_2103886, EPI_ISL_2103887, EPI_ISL_2103888, EPI_ISL_2103889, EPI_ISL_2103890, EPI_ISL_2103891, EPI_ISL_2103892, EPI_ISL_2103893, EPI_ISL_2103894, EPI_ISL_2103895, EPI_ISL_2103896, EPI_ISL_2103897, EPI_ISL_2103898, EPI_ISL_2103899, EPI_ISL_2103900, EPI_ISL_2103901, EPI_ISL_2103902, EPI_ISL_2103903, EPI_ISL_2103904, EPI_ISL_2103905, EPI_ISL_2103906, EPI_ISL_2103907, EPI_ISL_2103908, EPI_ISL_2103909, EPI_ISL_2103910, EPI_ISL_2103911, EPI_ISL_2103912, EPI_ISL_2103913, EPI_ISL_2103914, EPI_ISL_2103915, EPI_ISL_2103917, EPI_ISL_2103918, EPI_ISL_2103919, EPI_ISL_2103920, EPI_ISL_2103921, EPI_ISL_2103922, EPI_ISL_2103923, EPI_ISL_2103925, EPI_ISL_2103926, EPI_ISL_2103927, EPI_ISL_2103928, EPI_ISL_2103931, EPI_ISL_2103932, EPI_ISL_2103933, EPI_ISL_2103934, EPI_ISL_2103935, EPI_ISL_2103936, EPI_ISL_2103937, EPI_ISL_2103938, EPI_ISL_2103939, EPI_ISL_2103940, EPI_ISL_2103941, EPI_ISL_2103942, EPI_ISL_2103943, EPI_ISL_2103944, EPI_ISL_2103945, EPI_ISL_2103946, EPI_ISL_2103947, EPI_ISL_2103948, EPI_ISL_2103949, EPI_ISL_2103950, EPI_ISL_2103951, EPI_ISL_2103952, EPI_ISL_2103953, EPI_ISL_2103954, EPI_ISL_2103955, EPI_ISL_2103956, EPI_ISL_2103957, EPI_ISL_2103959, EPI_ISL_2103960, EPI_ISL_2103961, EPI_ISL_2103962, EPI_ISL_2103963, EPI_ISL_2103964, EPI_ISL_2103965, EPI_ISL_2103967, EPI_ISL_2103968, EPI_ISL_2103969, EPI_ISL_2103970, EPI_ISL_2103971, EPI_ISL_2103972, EPI_ISL_2103973, EPI_ISL_2103974, EPI_ISL_2103975, EPI_ISL_2103976, EPI_ISL_2103977, EPI_ISL_2103978, EPI_ISL_2103979, EPI_ISL_2103980, EPI_ISL_2103981, EPI_ISL_2103983, EPI_ISL_2103984, EPI_ISL_2103985, EPI_ISL_2103987, EPI_ISL_2103988, EPI_ISL_2103989, EPI_ISL_2103990, EPI_ISL_2103991, EPI_ISL_2103992, EPI_ISL_2103993, EPI_ISL_2103994, EPI_ISL_2103995, EPI_ISL_2103996, EPI_ISL_2103997, EPI_ISL_2103998, EPI_ISL_2103999, EPI_ISL_2104000, EPI_ISL_2104002, EPI_ISL_2104003, EPI_ISL_2104004, EPI_ISL_2104005, EPI_ISL_2104008, EPI_ISL_2104009, EPI_ISL_2104010, EPI_ISL_2104011, EPI_ISL_2104012, EPI_ISL_2104013, EPI_ISL_2104014, EPI_ISL_2104015, EPI_ISL_2104016, EPI_ISL_2104017, EPI_ISL_2104019, EPI_ISL_2104020, EPI_ISL_2104021, EPI_ISL_2104022, EPI_ISL_2104023, EPI_ISL_2104024, EPI_ISL_2104025, EPI_ISL_2104026, EPI_ISL_2104027, EPI_ISL_2104028, EPI_ISL_2104029, EPI_ISL_2104030, EPI_ISL_2104031, EPI_ISL_2104032, EPI_ISL_2104033, EPI_ISL_2104034, EPI_ISL_2104036, EPI_ISL_2104037, EPI_ISL_2104038, EPI_ISL_2104039, EPI_ISL_2104040, EPI_ISL_2104043, EPI_ISL_2104045, EPI_ISL_2104046, EPI_ISL_2104047, EPI_ISL_2104048, EPI_ISL_2104049, EPI_ISL_2104050, EPI_ISL_2104051, EPI_ISL_2104052, EPI_ISL_2104053, EPI_ISL_2104054, EPI_ISL_2104055, | | | |

EPI_ISL_2104057, EPI_ISL_2104058, EPI_ISL_2104059, EPI_ISL_2104060, EPI_ISL_2104061, EPI_ISL_2104062, EPI_ISL_2104063, EPI_ISL_2104065, EPI_ISL_2104066, EPI_ISL_2104067, EPI_ISL_2104068, EPI_ISL_2104069, EPI_ISL_2104071, EPI_ISL_2104072, EPI_ISL_2104073, EPI_ISL_2104074, EPI_ISL_2104075, EPI_ISL_2104076, EPI_ISL_2104077, EPI_ISL_2104078, EPI_ISL_2104080, EPI_ISL_2104081,
EPI_ISL_2104082, EPI_ISL_2104083, EPI_ISL_2104084, EPI_ISL_2104085, EPI_ISL_2104086, EPI_ISL_2104087, EPI_ISL_2104088, EPI_ISL_2104089, EPI_ISL_2104091, EPI_ISL_2104092, EPI_ISL_2104093, EPI_ISL_2104095, EPI_ISL_2104096, EPI_ISL_2104097, EPI_ISL_2104099, EPI_ISL_2104100, EPI_ISL_2104101, EPI_ISL_2104102, EPI_ISL_2104103, EPI_ISL_2104104, EPI_ISL_2104105, EPI_ISL_2104107,
EPI_ISL_2104108, EPI_ISL_2104109, EPI_ISL_2104110, EPI_ISL_2104111, EPI_ISL_2104112, EPI_ISL_2104113, EPI_ISL_2104114, EPI_ISL_2104115, EPI_ISL_2104116, EPI_ISL_2104117, EPI_ISL_2104118, EPI_ISL_2104119, EPI_ISL_2104120, EPI_ISL_2104121, EPI_ISL_2104122, EPI_ISL_2104123, EPI_ISL_2104124, EPI_ISL_2104125, EPI_ISL_2104127, EPI_ISL_2104128, EPI_ISL_2104129, EPI_ISL_2104130,
EPI_ISL_2104131, EPI_ISL_2104133, EPI_ISL_2104134, EPI_ISL_2104135, EPI_ISL_2104136, EPI_ISL_2104137, EPI_ISL_2104138, EPI_ISL_2104139, EPI_ISL_2104140, EPI_ISL_2104142, EPI_ISL_2104143, EPI_ISL_2104144, EPI_ISL_2104145, EPI_ISL_2104146, EPI_ISL_2104147, EPI_ISL_2104149, EPI_ISL_2104150, EPI_ISL_2104151, EPI_ISL_2104152, EPI_ISL_2104153, EPI_ISL_2104154, EPI_ISL_2104155,
EPI_ISL_2104157, EPI_ISL_2104158, EPI_ISL_2104159, EPI_ISL_2104160, EPI_ISL_2104161, EPI_ISL_2104162, EPI_ISL_2104163, EPI_ISL_2104164, EPI_ISL_2104165, EPI_ISL_2104166, EPI_ISL_2104167, EPI_ISL_2104168, EPI_ISL_2104169, EPI_ISL_2104170, EPI_ISL_2104171, EPI_ISL_2104172, EPI_ISL_2104173, EPI_ISL_2104174, EPI_ISL_2104175, EPI_ISL_2104177, EPI_ISL_2104178, EPI_ISL_2104180,
EPI_ISL_2104181, EPI_ISL_2104183, EPI_ISL_2104184, EPI_ISL_2104185, EPI_ISL_2104186, EPI_ISL_2104187, EPI_ISL_2104188, EPI_ISL_2104189, EPI_ISL_2104190, EPI_ISL_2104192, EPI_ISL_2104193, EPI_ISL_2104194, EPI_ISL_2104195, EPI_ISL_2104196, EPI_ISL_2104197, EPI_ISL_2104198, EPI_ISL_2104199, EPI_ISL_2104200, EPI_ISL_2104202, EPI_ISL_2104203, EPI_ISL_2104205, EPI_ISL_2104206,
EPI_ISL_2104207, EPI_ISL_2104208, EPI_ISL_2104209, EPI_ISL_2104210, EPI_ISL_2104211, EPI_ISL_2104212, EPI_ISL_2104213, EPI_ISL_2104214, EPI_ISL_2104215, EPI_ISL_2104216, EPI_ISL_2104217, EPI_ISL_2104218, EPI_ISL_2104219, EPI_ISL_2104220, EPI_ISL_2104221, EPI_ISL_2104222, EPI_ISL_2104224, EPI_ISL_2104225, EPI_ISL_2104226, EPI_ISL_2104227, EPI_ISL_2104228, EPI_ISL_2104230,
EPI_ISL_2104231, EPI_ISL_2104232, EPI_ISL_2104233, EPI_ISL_2104234, EPI_ISL_2104235, EPI_ISL_2104236, EPI_ISL_2104237, EPI_ISL_2104238, EPI_ISL_2104240, EPI_ISL_2104241, EPI_ISL_2104242, EPI_ISL_2104243, EPI_ISL_2104244, EPI_ISL_2104245, EPI_ISL_2104246, EPI_ISL_2104247, EPI_ISL_2104249, EPI_ISL_2104250, EPI_ISL_2104251, EPI_ISL_2104252, EPI_ISL_2104253, EPI_ISL_2104254,
EPI_ISL_2104255, EPI_ISL_2104256, EPI_ISL_2104257, EPI_ISL_2104258, EPI_ISL_2104259, EPI_ISL_2104260, EPI_ISL_2104262, EPI_ISL_2104263, EPI_ISL_2104264, EPI_ISL_2104266, EPI_ISL_2104267, EPI_ISL_2104268, EPI_ISL_2104269, EPI_ISL_2104271, EPI_ISL_2104272, EPI_ISL_2104273, EPI_ISL_2104274, EPI_ISL_2104275, EPI_ISL_2104276, EPI_ISL_2104277, EPI_ISL_2104278, EPI_ISL_2104280,
EPI_ISL_2104281, EPI_ISL_2104282, EPI_ISL_2104283, EPI_ISL_2104284, EPI_ISL_2104285, EPI_ISL_2104286, EPI_ISL_2104287, EPI_ISL_2104288, EPI_ISL_2104289, EPI_ISL_2104290, EPI_ISL_2104292, EPI_ISL_2104294, EPI_ISL_2104295, EPI_ISL_2192595, EPI_ISL_2192596, EPI_ISL_2192597, EPI_ISL_2192598, EPI_ISL_2192599, EPI_ISL_2192600, EPI_ISL_2192601, EPI_ISL_2192602, EPI_ISL_2192603,
EPI_ISL_2192604, EPI_ISL_2192605, EPI_ISL_2192606, EPI_ISL_2192607, EPI_ISL_2192608, EPI_ISL_2192609, EPI_ISL_2192610, EPI_ISL_2192611, EPI_ISL_2192613, EPI_ISL_2192614, EPI_ISL_2192615, EPI_ISL_2192616, EPI_ISL_2192617, EPI_ISL_2192618, EPI_ISL_2192619, EPI_ISL_2192620, EPI_ISL_2192622, EPI_ISL_2192623, EPI_ISL_2192624, EPI_ISL_2192626, EPI_ISL_2192627, EPI_ISL_2192628,
EPI_ISL_2192629, EPI_ISL_2192630, EPI_ISL_2192632, EPI_ISL_2192634, EPI_ISL_2192635, EPI_ISL_2192636, EPI_ISL_2192637, EPI_ISL_2192639, EPI_ISL_2192641, EPI_ISL_2192643, EPI_ISL_2192644, EPI_ISL_2192645, EPI_ISL_2192646, EPI_ISL_2192647, EPI_ISL_2192649, EPI_ISL_2192651, EPI_ISL_2192652, EPI_ISL_2192655, EPI_ISL_2192656, EPI_ISL_2192657, EPI_ISL_2192658, EPI_ISL_2192659,
EPI_ISL_2192661, EPI_ISL_2192662, EPI_ISL_2192663, EPI_ISL_2192664, EPI_ISL_2192665, EPI_ISL_2192666, EPI_ISL_2192667, EPI_ISL_2192668, EPI_ISL_2192669, EPI_ISL_2192670, EPI_ISL_2192671, EPI_ISL_2192672, EPI_ISL_2192673, EPI_ISL_2192675, EPI_ISL_2192676, EPI_ISL_2192677, EPI_ISL_2192678, EPI_ISL_2192680, EPI_ISL_2192682, EPI_ISL_2192683, EPI_ISL_2192684, EPI_ISL_2192685,
EPI_ISL_2192686, EPI_ISL_2192687, EPI_ISL_2192688, EPI_ISL_2192689, EPI_ISL_2192690, EPI_ISL_2192691, EPI_ISL_2192692, EPI_ISL_2192693, EPI_ISL_2192694, EPI_ISL_2192695, EPI_ISL_2192697, EPI_ISL_2192698, EPI_ISL_2192699, EPI_ISL_2192700, EPI_ISL_2192701, EPI_ISL_2192702, EPI_ISL_2192703, EPI_ISL_2192704, EPI_ISL_2192705, EPI_ISL_2192706, EPI_ISL_2192707, EPI_ISL_2192708,
EPI_ISL_2192710, EPI_ISL_2192712, EPI_ISL_2192713, EPI_ISL_2192714, EPI_ISL_2192715, EPI_ISL_2192716, EPI_ISL_2192717, EPI_ISL_2192718, EPI_ISL_2192719, EPI_ISL_2192720, EPI_ISL_2192721, EPI_ISL_2192722, EPI_ISL_2192723, EPI_ISL_2192725, EPI_ISL_2192726, EPI_ISL_2192727, EPI_ISL_2192728, EPI_ISL_2192729, EPI_ISL_2192731, EPI_ISL_2192732, EPI_ISL_2192733, EPI_ISL_2192735,
EPI_ISL_2192736, EPI_ISL_2192737, EPI_ISL_2192738, EPI_ISL_2192739, EPI_ISL_2192740, EPI_ISL_2192741, EPI_ISL_2192742, EPI_ISL_2192744, EPI_ISL_2192745, EPI_ISL_2192746, EPI_ISL_2192747, EPI_ISL_2192748, EPI_ISL_2192750, EPI_ISL_2192751, EPI_ISL_2192752, EPI_ISL_2192753, EPI_ISL_2192754, EPI_ISL_2192755, EPI_ISL_2192756, EPI_ISL_2192757, EPI_ISL_2192758, EPI_ISL_2192759,
EPI_ISL_2192760, EPI_ISL_2192761, EPI_ISL_2192762, EPI_ISL_2192763, EPI_ISL_2192765, EPI_ISL_2192766, EPI_ISL_2192767, EPI_ISL_2192768, EPI_ISL_2192769, EPI_ISL_2192770, EPI_ISL_2192771, EPI_ISL_2192772, EPI_ISL_2192773, EPI_ISL_2192775, EPI_ISL_2192776, EPI_ISL_2192777, EPI_ISL_2192778, EPI_ISL_2192780, EPI_ISL_2192781, EPI_ISL_2192782, EPI_ISL_2192783, EPI_ISL_2192784,
EPI_ISL_2192785, EPI_ISL_2192786, EPI_ISL_2192787, EPI_ISL_2192788, EPI_ISL_2192789, EPI_ISL_2192790, EPI_ISL_2192791, EPI_ISL_2192793, EPI_ISL_2192794, EPI_ISL_2192796, EPI_ISL_2192797, EPI_ISL_2192798, EPI_ISL_2192799, EPI_ISL_2192800, EPI_ISL_2192801, EPI_ISL_2192802, EPI_ISL_2192803, EPI_ISL_2192804, EPI_ISL_2192805, EPI_ISL_2192807, EPI_ISL_2192808, EPI_ISL_2192811,
EPI_ISL_2192812, EPI_ISL_2192813, EPI_ISL_2192814, EPI_ISL_2192816, EPI_ISL_2192817, EPI_ISL_2192818, EPI_ISL_2192819, EPI_ISL_2192820, EPI_ISL_2192821, EPI_ISL_2192822, EPI_ISL_2192823, EPI_ISL_2192826, EPI_ISL_2192827, EPI_ISL_2192828, EPI_ISL_2192829, EPI_ISL_2192830, EPI_ISL_2192831, EPI_ISL_2192832, EPI_ISL_2192833, EPI_ISL_2192834, EPI_ISL_2192835, EPI_ISL_2192836,
EPI_ISL_2192837, EPI_ISL_2192838, EPI_ISL_2192839, EPI_ISL_2192841, EPI_ISL_2192842, EPI_ISL_2192843, EPI_ISL_2192845, EPI_ISL_2192846, EPI_ISL_2192847, EPI_ISL_2192848, EPI_ISL_2192849, EPI_ISL_2192850, EPI_ISL_2192851, EPI_ISL_2192852, EPI_ISL_2192853, EPI_ISL_2192854, EPI_ISL_2192855, EPI_ISL_2192856, EPI_ISL_2192857, EPI_ISL_2192858, EPI_ISL_2192860, EPI_ISL_2192861,
EPI_ISL_2192862, EPI_ISL_2192863, EPI_ISL_2192865, EPI_ISL_2192866, EPI_ISL_2192867, EPI_ISL_2192868, EPI_ISL_2192869, EPI_ISL_2192870, EPI_ISL_2192871, EPI_ISL_2192872, EPI_ISL_2192873, EPI_ISL_2192874, EPI_ISL_2192875, EPI_ISL_2192876, EPI_ISL_2192879, EPI_ISL_2192880, EPI_ISL_2192881, EPI_ISL_2192882, EPI_ISL_2192883, EPI_ISL_2192884, EPI_ISL_2192885, EPI_ISL_2192886,
EPI_ISL_2192887, EPI_ISL_2192888, EPI_ISL_2192889, EPI_ISL_2192890, EPI_ISL_2192891, EPI_ISL_2192892, EPI_ISL_2192893, EPI_ISL_2192894, EPI_ISL_2192896, EPI_ISL_2192897, EPI_ISL_2192898, EPI_ISL_2192899, EPI_ISL_2192900, EPI_ISL_2192901, EPI_ISL_2192902, EPI_ISL_2192903, EPI_ISL_2192904, EPI_ISL_2192906, EPI_ISL_2192907, EPI_ISL_2192908, EPI_ISL_2192909, EPI_ISL_2192910,
EPI_ISL_2192911, EPI_ISL_2192912, EPI_ISL_2192913, EPI_ISL_2192914, EPI_ISL_2192916, EPI_ISL_2192917, EPI_ISL_2192918, EPI_ISL_2192919, EPI_ISL_2192920, EPI_ISL_2192921, EPI_ISL_2192922, EPI_ISL_2192923, EPI_ISL_2192925, EPI_ISL_2192926, EPI_ISL_2192927, EPI_ISL_2192928, EPI_ISL_2192929, EPI_ISL_2192930, EPI_ISL_2192931, EPI_ISL_2192932, EPI_ISL_2192933, EPI_ISL_2192934,
EPI_ISL_2192935, EPI_ISL_2192936, EPI_ISL_2192937, EPI_ISL_2192938, EPI_ISL_2192940, EPI_ISL_2192942, EPI_ISL_2192943, EPI_ISL_2192945, EPI_ISL_2192946, EPI_ISL_2192947, EPI_ISL_2192948, EPI_ISL_2192949, EPI_ISL_2192950, EPI_ISL_2192952, EPI_ISL_2192953, EPI_ISL_2192954, EPI_ISL_2192955, EPI_ISL_2192956, EPI_ISL_2192957, EPI_ISL_2192958, EPI_ISL_2192960, EPI_ISL_2192961,
EPI_ISL_2192962, EPI_ISL_2192963, EPI_ISL_2192964, EPI_ISL_2192965, EPI_ISL_2192966, EPI_ISL_2192967, EPI_ISL_2192968, EPI_ISL_2192969, EPI_ISL_2192971, EPI_ISL_2192972, EPI_ISL_2192973, EPI_ISL_2192974, EPI_ISL_2192975, EPI_ISL_2192976, EPI_ISL_2192978, EPI_ISL_2192979, EPI_ISL_2192980, EPI_ISL_2192981, EPI_ISL_2192982, EPI_ISL_2192983, EPI_ISL_2192984, EPI_ISL_2192985,
EPI_ISL_2192986, EPI_ISL_2192988, EPI_ISL_2192989, EPI_ISL_2192990, EPI_ISL_2192991, EPI_ISL_2192992, EPI_ISL_2192993, EPI_ISL_2192994, EPI_ISL_2192996, EPI_ISL_2192997, EPI_ISL_2192998, EPI_ISL_2192999, EPI_ISL_2193000, EPI_ISL_2193001, EPI_ISL_2193002, EPI_ISL_2193003, EPI_ISL_2193004, EPI_ISL_2193005, EPI_ISL_2193006, EPI_ISL_2193007, EPI_ISL_2193008, EPI_ISL_2193009,
EPI_ISL_2193010, EPI_ISL_2193011, EPI_ISL_2193013, EPI_ISL_2193014, EPI_ISL_2193015, EPI_ISL_2193017, EPI_ISL_2193018, EPI_ISL_2193019, EPI_ISL_2193020, EPI_ISL_2193021, EPI_ISL_2193023, EPI_ISL_2193024, EPI_ISL_2193025, EPI_ISL_2193026, EPI_ISL_2193027, EPI_ISL_2193028, EPI_ISL_2193029, EPI_ISL_2193030, EPI_ISL_2193031, EPI_ISL_2193032, EPI_ISL_2193033, EPI_ISL_2193034,
EPI_ISL_2193035, EPI_ISL_2193036, EPI_ISL_2193037, EPI_ISL_2193038, EPI_ISL_2193039, EPI_ISL_2193040, EPI_ISL_2193043, EPI_ISL_2193044, EPI_ISL_2193046, EPI_ISL_2193047, EPI_ISL_2193048, EPI_ISL_2193049, EPI_ISL_2193050, EPI_ISL_2193051, EPI_ISL_2193052, EPI_ISL_2193053, EPI_ISL_2193054, EPI_ISL_2193055, EPI_ISL_2193056, EPI_ISL_2193057, EPI_ISL_2193058, EPI_ISL_2193059,
EPI_ISL_2193060, EPI_ISL_2193061, EPI_ISL_2193062, EPI_ISL_2193063, EPI_ISL_2193064, EPI_ISL_2193065, EPI_ISL_2193066, EPI_ISL_2193067, EPI_ISL_2193068, EPI_ISL_2193069, EPI_ISL_2193070, EPI_ISL_2193071, EPI_ISL_2193072, EPI_ISL_2193074, EPI_ISL_2193075, EPI_ISL_2193076, EPI_ISL_2193077, EPI_ISL_2193079, EPI_ISL_2193080, EPI_ISL_2193081, EPI_ISL_2193083, EPI_ISL_2193085,
EPI_ISL_2193087, EPI_ISL_2193088, EPI_ISL_2193089, EPI_ISL_2193090, EPI_ISL_2193091, EPI_ISL_2193092, EPI_ISL_2193093, EPI_ISL_2193094, EPI_ISL_2193095, EPI_ISL_2193096, EPI_ISL_2193097, EPI_ISL_2193098, EPI_ISL_2193099, EPI_ISL_2193100, EPI_ISL_2193101, EPI_ISL_2193102, EPI_ISL_2193103, EPI_ISL_2193104, EPI_ISL_2193105, EPI_ISL_2193106, EPI_ISL_2193107, EPI_ISL_2193108,
EPI_ISL_2193109, EPI_ISL_2193110, EPI_ISL_2311868, EPI_ISL_2311869, EPI_ISL_2311870, EPI_ISL_2311871, EPI_ISL_2311872, EPI_ISL_2311873, EPI_ISL_2311874, EPI_ISL_2311875, EPI_ISL_2311876, EPI_ISL_2311877, EPI_ISL_2311878, EPI_ISL_2311879, EPI_ISL_2311880, EPI_ISL_2311881, EPI_ISL_2311882, EPI_ISL_2311883, EPI_ISL_2311884, EPI_ISL_2311885, EPI_ISL_2311886, EPI_ISL_2311887,
EPI_ISL_2311888, EPI_ISL_2311889, EPI_ISL_2311890, EPI_ISL_2311892, EPI_ISL_2311893, EPI_ISL_2311895, EPI_ISL_2311896, EPI_ISL_2311897, EPI_ISL_2311898, EPI_ISL_2311899, EPI_ISL_2311900, EPI_ISL_2311901, EPI_ISL_2311902, EPI_ISL_2311905, EPI_ISL_2311906, EPI_ISL_2311907, EPI_ISL_2311908, EPI_ISL_2311911, EPI_ISL_2311912, EPI_ISL_2311913, EPI_ISL_2311914, EPI_ISL_2311915,
EPI_ISL_2311916, EPI_ISL_2311917, EPI_ISL_2311918, EPI_ISL_2311919, EPI_ISL_2311920, EPI_ISL_2311921, EPI_ISL_2311922, EPI_ISL_2311923, EPI_ISL_2311924, EPI_ISL_2311925, EPI_ISL_2311926, EPI_ISL_2311927, EPI_ISL_2311928, EPI_ISL_2311929, EPI_ISL_2311930, EPI_ISL_2311931, EPI_ISL_2311932, EPI_ISL_2311933, EPI_ISL_2311934, EPI_ISL_2311935, EPI_ISL_2311936, EPI_ISL_2311937,
EPI_ISL_2311938, EPI_ISL_2311940, EPI_ISL_2311941, EPI_ISL_2311942, EPI_ISL_2311943, EPI_ISL_2311945, EPI_ISL_2311946, EPI_ISL_2311947, EPI_ISL_2311948, EPI_ISL_2311949, EPI_ISL_2311950, EPI_ISL_2311951, EPI_ISL_2311952, EPI_ISL_2311953, EPI_ISL_2311954, EPI_ISL_2311955, EPI_ISL_2311956, EPI_ISL_2311957, EPI_ISL_2311958, EPI_ISL_2311959, EPI_ISL_2311960, EPI_ISL_2311961,
EPI_ISL_2311962, EPI_ISL_2311963, EPI_ISL_2311964, EPI_ISL_2311965, EPI_ISL_2311966, EPI_ISL_2311967, EPI_ISL_2311968, EPI_ISL_2311969, EPI_ISL_2311970, EPI_ISL_2311971, EPI_ISL_2311972, EPI_ISL_2311973, EPI_ISL_2311974, EPI_ISL_2311975, EPI_ISL_2311976, EPI_ISL_2311977, EPI_ISL_2311978, EPI_ISL_2311979, EPI_ISL_2311980, EPI_ISL_2311982, EPI_ISL_2311983, EPI_ISL_2311984,
EPI_ISL_2311985, EPI_ISL_2311986, EPI_ISL_2311987, EPI_ISL_2311988, EPI_ISL_2311989, EPI_ISL_2311991, EPI_ISL_2311992, EPI_ISL_2311993, EPI_ISL_2311994, EPI_ISL_2311995, EPI_ISL_2311996, EPI_ISL_2311997, EPI_ISL_2311998, EPI_ISL_2311999, EPI_ISL_2312000, EPI_ISL_2312001, EPI_ISL_2312003, EPI_ISL_2312004, EPI_ISL_2312005, EPI_ISL_2312006, EPI_ISL_2312007, EPI_ISL_2312008,
EPI_ISL_2312009, EPI_ISL_2312010, EPI_ISL_2312011, EPI_ISL_2312014, EPI_ISL_2312015, EPI_ISL_2312016, EPI_ISL_2312017, EPI_ISL_2312018, EPI_ISL_2312020, EPI_ISL_2312021, EPI_ISL_2312022, EPI_ISL_2312023, EPI_ISL_2312024, EPI_ISL_2312025, EPI_ISL_2312026, EPI_ISL_2312027, EPI_ISL_2312028, EPI_ISL_2312029, EPI_ISL_2312030, EPI_ISL_2312031, EPI_ISL_2312032, EPI_ISL_2312033,
EPI_ISL_2312034, EPI_ISL_2312035, EPI_ISL_2312037, EPI_ISL_2312038, EPI_ISL_2312039, EPI_ISL_2312040, EPI_ISL_2312041, EPI_ISL_2312042, EPI_ISL_2312045, EPI_ISL_2312047, EPI_ISL_2312048, EPI_ISL_2312049, EPI_ISL_2312050, EPI_ISL_2312051, EPI_ISL_2312052, EPI_ISL_2312053, EPI_ISL_2312054, EPI_ISL_2312055, EPI_ISL_2312057, EPI_ISL_2312058, EPI_ISL_2312059, EPI_ISL_2312060,
EPI_ISL_2312062, EPI_ISL_2312063, EPI_ISL_2312064, EPI_ISL_2312065, EPI_ISL_2312066, EPI_ISL_2312067, EPI_ISL_2312068, EPI_ISL_2312069, EPI_ISL_2312070, EPI_ISL_2312071, EPI_ISL_2312072, EPI_ISL_2312073, EPI_ISL_2312075, EPI_ISL_2312076, EPI_ISL_2312077, EPI_ISL_2312078, EPI_ISL_2312080, EPI_ISL_2312081, EPI_ISL_2312082, EPI_ISL_2312083, EPI_ISL_2312084, EPI_ISL_2312085,
EPI_ISL_2312087, EPI_ISL_2312088, EPI_ISL_2312091, EPI_ISL_2312092, EPI_ISL_2312093, EPI_ISL_2312094, EPI_ISL_2312095, EPI_ISL_2312096, EPI_ISL_2312097, EPI_ISL_2312098, EPI_ISL_2312099, EPI_ISL_2312100, EPI_ISL_2312101, EPI_ISL_2312102, EPI_ISL_2312103, EPI_ISL_2312104, EPI_ISL_2312105, EPI_ISL_2312106, EPI_ISL_2312107, EPI_ISL_2312108, EPI_ISL_2312109, EPI_ISL_2312110,
EPI_ISL_2312111, EPI_ISL_2312113, EPI_ISL_2312114, EPI_ISL_2312115, EPI_ISL_2312116, EPI_ISL_2312117, EPI_ISL_2312118, EPI_ISL_2312119, EPI_ISL_2312120, EPI_ISL_2312121, EPI_ISL_2312122, EPI_ISL_2312123, EPI_ISL_2312124, EPI_ISL_2312125, EPI_ISL_2312126, EPI_ISL_2312128, EPI_ISL_2312129, EPI_ISL_2312130, EPI_ISL_2312131, EPI_ISL_2312132, EPI_ISL_2312133, EPI_ISL_2312134,
EPI_ISL_2312135, EPI_ISL_2312136, EPI_ISL_2312137, EPI_ISL_2312139, EPI_ISL_2312140, EPI_ISL_2312141, EPI_ISL_2312142, EPI_ISL_2312143, EPI_ISL_2312144, EPI_ISL_2312146, EPI_ISL_2312147, EPI_ISL_2312148, EPI_ISL_2312149, EPI_ISL_2312151, EPI_ISL_2312152, EPI_ISL_2312153, EPI_ISL_2312154, EPI_ISL_2312155, EPI_ISL_2312156, EPI_ISL_2312157, EPI_ISL_2312158, EPI_ISL_2312159,
EPI_ISL_2312160, EPI_ISL_2312161, EPI_ISL_2312162, EPI_ISL_2312163, EPI_ISL_2312165, EPI_ISL_2312166, EPI_ISL_2312167, EPI_ISL_2312168, EPI_ISL_2312169, EPI_ISL_2312170, EPI_ISL_2312171, EPI_ISL_2312173, EPI_ISL_2312174, EPI_ISL_2312175, EPI_ISL_2312176, EPI_ISL_2312177, EPI_ISL_2312179, EPI_ISL_2312180, EPI_ISL_2312181, EPI_ISL_2312182, EPI_ISL_2312184, EPI_ISL_2312185,
EPI_ISL_2312186, EPI_ISL_2312187, EPI_ISL_2312188, EPI_ISL_2312189, EPI_ISL_2312190, EPI_ISL_2312191, EPI_ISL_2312193, EPI_ISL_2312194, EPI_ISL_2312195, EPI_ISL_2312196, EPI_ISL_2312197, EPI_ISL_2312198, EPI_ISL_2312199, EPI_ISL_2312200, EPI_ISL_2312201, EPI_ISL_2312202, EPI_ISL_2312203, EPI_ISL_2312204, EPI_ISL_2312205, EPI_ISL_2312206, EPI_ISL_2312208, EPI_ISL_2312209,
EPI_ISL_2312211, EPI_ISL_2312212, EPI_ISL_2312213, EPI_ISL_2312214, EPI_ISL_2312215, EPI_ISL_2312216, EPI_ISL_2312217, EPI_ISL_2312219, EPI_ISL_2312220, EPI_ISL_2312221, EPI_ISL_2312222, EPI_ISL_2312223, EPI_ISL_2312224, EPI_ISL_2312225, EPI_ISL_2312227, EPI_ISL_2312228, EPI_ISL_2312229, EPI_ISL_2312231, EPI_ISL_2312232, EPI_ISL_2312234, EPI_ISL_2312235, EPI_ISL_2312236,
EPI_ISL_2312237, EPI_ISL_2312238, EPI_ISL_2312239, EPI_ISL_2312240, EPI_ISL_2312241, EPI_ISL_2312242, EPI_ISL_2312243, EPI_ISL_2312244, EPI_ISL_2312245, EPI_ISL_2312247, EPI_ISL_2312248, EPI_ISL_2312249, EPI_ISL_2312250, EPI_ISL_2312251, EPI_ISL_2312252, EPI_ISL_2312254, EPI_ISL_2312255, EPI_ISL_2312256, EPI_ISL_2312257, EPI_ISL_2312258, EPI_ISL_2312259, EPI_ISL_2312260,
EPI_ISL_2312261, EPI_ISL_2312262, EPI_ISL_2312263, EPI_ISL_2312264, EPI_ISL_2312265, EPI_ISL_2312266, EPI_ISL_2312267, EPI_ISL_2312268, EPI_ISL_2312270, EPI_ISL_2312271, EPI_ISL_2312272, EPI_ISL_2312273, EPI_ISL_2312275, EPI_ISL_2312278, EPI_ISL_2312279, EPI_ISL_2312280, EPI_ISL_2312281, EPI_ISL_2312282, EPI_ISL_2312283, EPI_ISL_2312284, EPI_ISL_2312285, EPI_ISL_2312286,
EPI_ISL_2312287, EPI_ISL_2312288, EPI_ISL_2312289, EPI_ISL_2312290, EPI_ISL_2312291, EPI_ISL_2312292, EPI_ISL_2312293, EPI_ISL_2312294, EPI_ISL_2312295, EPI_ISL_2312296, EPI_ISL_2312297, EPI_ISL_2312298, EPI_ISL_2312299, EPI_ISL_2312300, EPI_ISL_2312301, EPI_ISL_2312302, EPI_ISL_2312303, EPI_ISL_2312304, EPI_ISL_2312305, EPI_ISL_2312306, EPI_ISL_2312308, EPI_ISL_2312309,
EPI_ISL_2312310, EPI_ISL_2312311, EPI_ISL_2312312, EPI_ISL_2312313, EPI_ISL_2312314, EPI_ISL_2312315, EPI_ISL_2312316, EPI_ISL_2312317, EPI_ISL_2312318, EPI_ISL_2312319, EPI_ISL_2312320, EPI_ISL_2312321, EPI_ISL_2312322, EPI_ISL_2312323, EPI_ISL_2312324, EPI_ISL_2312325, EPI_ISL_2312326, EPI_ISL_2312327, EPI_ISL_2312328, EPI_ISL_2312329, EPI_ISL_2312330, EPI_ISL_2312331,
EPI_ISL_2312332, EPI_ISL_2312333, EPI_ISL_2312334, EPI_ISL_2312336, EPI_ISL_2312338, EPI_ISL_2312339, EPI_ISL_2312340, EPI_ISL_2312341, EPI_ISL_2312342, EPI_ISL_2312343, EPI_ISL_2312345, EPI_ISL_2312346, EPI_ISL_2312348, EPI_ISL_2312350, EPI_ISL_2312351, EPI_ISL_2312352, EPI_ISL_2312353, EPI_ISL_2312354, EPI_ISL_2312355, EPI_ISL_2312356, EPI_ISL_2312357, EPI_ISL_2312358,
EPI_ISL_2312359, EPI_ISL_2312360, EPI_ISL_2312362, EPI_ISL_2312363, EPI_ISL_2312364, EPI_ISL_2312366, EPI_ISL_2312368, EPI_ISL_2312369, EPI_ISL_2312370, EPI_ISL_2312371, EPI_ISL_2312372, EPI_ISL_2312374, EPI_ISL_2312377, EPI_ISL_2312378, EPI_ISL_2312379, EPI_ISL_2312380, EPI_ISL_2312382, EPI_ISL_2312383, EPI_ISL_2312384, EPI_ISL_2312385, EPI_ISL_2312386, EPI_ISL_2312387,
EPI_ISL_2312388, EPI_ISL_2312389, EPI_ISL_2312390, EPI_ISL_2312391, EPI_ISL_2312392, EPI_ISL_2312393, EPI_ISL_2312395, EPI_ISL_2312396, EPI_ISL_2312397, EPI_ISL_2312398, EPI_ISL_2312399, EPI_ISL_2312400, EPI_ISL_2312402, EPI_ISL_2312403, EPI_ISL_2312404, EPI_ISL_2312405, EPI_ISL_2312406, EPI_ISL_2312407, EPI_ISL_2312408, EPI_ISL_2312409, EPI_ISL_2312410, EPI_ISL_2312411,
EPI_ISL_2312412, EPI_ISL_2312414, EPI_ISL_2312415, EPI_ISL_2312416, EPI_ISL_2312419, EPI_ISL_2312420, EPI_ISL_2312421, EPI_ISL_3007308, EPI_ISL_3007309, EPI_ISL_3007310, EPI_ISL_3007311, EPI_ISL_3007312, EPI_ISL_3007313, EPI_ISL_3007314, EPI_ISL_3007315, EPI_ISL_3007316, EPI_ISL_3007317, EPI_ISL_3007318, EPI_ISL_3007320, EPI_ISL_3007322, EPI_ISL_3007323, EPI_ISL_3007324,
EPI_ISL_3007325, EPI_ISL_3007326, EPI_ISL_3007327, EPI_ISL_3007328, EPI_ISL_3007329, EPI_ISL_3007330, EPI_ISL_3007331, EPI_ISL_3007332, EPI_ISL_3007333, EPI_ISL_3007334, EPI_ISL_3007335, EPI_ISL_3007336, EPI_ISL_3007337, EPI_ISL_3007338, EPI_ISL_3007339, EPI_ISL_3007340, EPI_ISL_3007342, EPI_ISL_3007343, EPI_ISL_3007344, EPI_ISL_3007345, EPI_ISL_3007346, EPI_ISL_3007347,
EPI_ISL_3007348, EPI_ISL_3007349, EPI_ISL_3007350, EPI_ISL_3007351, EPI_ISL_3007352, EPI_ISL_3007353, EPI_ISL_3007354, EPI_ISL_3007355, EPI_ISL_3007356, EPI_ISL_3007357, EPI_ISL_3007358, EPI_ISL_3007359, EPI_ISL_3007361, EPI_ISL_3007362, EPI_ISL_3007363, EPI_ISL_3007364, EPI_ISL_3007365, EPI_ISL_3007366, EPI_ISL_3007367, EPI_ISL_3007368, EPI_ISL_3007369, EPI_ISL_3007370,
EPI_ISL_3007371, EPI_ISL_3007372, EPI_ISL_3007373, EPI_ISL_3007374, EPI_ISL_3007375, EPI_ISL_3007377, EPI_ISL_3007378, EPI_ISL_3007379, EPI_ISL_3007380, EPI_ISL_3007381, EPI_ISL_3007382, EPI_ISL_3007383, EPI_ISL_3007384, EPI_ISL_3007385, EPI_ISL_3007386, EPI_ISL_3007387, EPI_ISL_3007388, EPI_ISL_3007390, EPI_ISL_3007391, EPI_ISL_3007392, EPI_ISL_3007393, EPI_ISL_3007394,
EPI_ISL_3007395, EPI_ISL_3007396, EPI_ISL_3007397, EPI_ISL_3007398, EPI_ISL_3007399, EPI_ISL_3007400, EPI_ISL_3007401, EPI_ISL_3007402, EPI_ISL_3007403, EPI_ISL_3007404, EPI_ISL_3007405, EPI_ISL_3007406, EPI_ISL_3007407, EPI_ISL_3007408, EPI_ISL_3007409, EPI_ISL_3007410, EPI_ISL_3007411, EPI_ISL_3007412, EPI_ISL_3007413, EPI_ISL_3007414, EPI_ISL_3007415, EPI_ISL_3007416,
EPI_ISL_3007417, EPI_ISL_3007418, EPI_ISL_3007419, EPI_ISL_3007421, EPI_ISL_3007422, EPI_ISL_3007423, EPI_ISL_3007424, EPI_ISL_3007425, EPI_ISL_3007426, EPI_ISL_3007427, EPI_ISL_3007428, EPI_ISL_3007429, EPI_ISL_3007430, EPI_ISL_3007431, EPI_ISL_3007432, EPI_ISL_3007433, EPI_ISL_3007434, EPI_ISL_3007435, EPI_ISL_3007436, EPI_ISL_3007437, EPI_ISL_3007438, EPI_ISL_3007439,
EPI_ISL_3007440, EPI_ISL_3007441, EPI_ISL_3007442, EPI_ISL_3007443, EPI_ISL_3007444, EPI_ISL_3007445, EPI_ISL_3007446, EPI_ISL_3007447, EPI_ISL_3007448, EPI_ISL_3007449, EPI_ISL_3007450, EPI_ISL_3007451, EPI_ISL_3007452, EPI_ISL_3007453, EPI_ISL_3007454, EPI_ISL_3007455, EPI_ISL_3007456, EPI_ISL_3007457, EPI_ISL_3007458, EPI_ISL_3084586, EPI_ISL_3084587, EPI_ISL_3084588,
EPI_ISL_3084589, EPI_ISL_3084590, EPI_ISL_3084591, EPI_ISL_3084592, EPI_ISL_3084593, EPI_ISL_3084594, EPI_ISL_3084595, EPI_ISL_3084596, EPI_ISL_3084598, EPI_ISL_3084599, EPI_ISL_3084600, EPI_ISL_3084601, EPI_ISL_3084602, EPI_ISL_3084603, EPI_ISL_3084604, EPI_ISL_3084605, EPI_ISL_3084606, EPI_ISL_3084607, EPI_ISL_3084608, EPI_ISL_3084609, EPI_ISL_3084610, EPI_ISL_3084611,
EPI_ISL_3084612, EPI_ISL_3084613, EPI_ISL_3084614, EPI_ISL_3084615, EPI_ISL_3084616, EPI_ISL_3084617, EPI_ISL_3084618, EPI_ISL_3084619, EPI_ISL_3084620, EPI_ISL_3084621, EPI_ISL_3084622, EPI_ISL_3084623, EPI_ISL_3084624, EPI_ISL_3084625, EPI_ISL_3084626, EPI_ISL_3084627, EPI_ISL_3084628, EPI_ISL_3084629, EPI_ISL_3084630, EPI_ISL_3084631, EPI_ISL_3084632, EPI_ISL_3084633,
EPI_ISL_3084634, EPI_ISL_3084635, EPI_ISL_3084636, EPI_ISL_3084637, EPI_ISL_3084638, EPI_ISL_3084639, EPI_ISL_3084640, EPI_ISL_3084641, EPI_ISL_3084642, EPI_ISL_3084643, EPI_ISL_3084644, EPI_ISL_3084645, EPI_ISL_3084646, EPI_ISL_3084647, EPI_ISL_3084648, EPI_ISL_3084649, EPI_ISL_3084650, EPI_ISL_3084651, EPI_ISL_3084652, EPI_ISL_3084653, EPI_ISL_3084654, EPI_ISL_3084655,
EPI_ISL_3084656, EPI_ISL_3084657, EPI_ISL_3084658, EPI_ISL_3084659, EPI_ISL_3084660, EPI_ISL_3084661, EPI_ISL_3084662, EPI_ISL_3084663, EPI_ISL_3084664, EPI_ISL_3084665, EPI_ISL_3084666, EPI_ISL_3084667, EPI_ISL_3084668, EPI_ISL_3084669, EPI_ISL_3084670, EPI_ISL_3084671, EPI_ISL_3084672, EPI_ISL_3084673, EPI_ISL_3084674, EPI_ISL_3084675, EPI_ISL_3084676, EPI_ISL_3084677,
EPI_ISL_3084678, EPI_ISL_3084679, EPI_ISL_3084680, EPI_ISL_3084681, EPI_ISL_3084682, EPI_ISL_3084683, EPI_ISL_3084684, EPI_ISL_3084685, EPI_ISL_3084686, EPI_ISL_3084687, EPI_ISL_3084688, EPI_ISL_3084689, EPI_ISL_3084690, EPI_ISL_3084691, EPI_ISL_3084692, EPI_ISL_3084693, EPI_ISL_3084694, EPI_ISL_3084695, EPI_ISL_3084696, EPI_ISL_3084697, EPI_ISL_3084698, EPI_ISL_3084699,
EPI_ISL_3084700, EPI_ISL_3084701, EPI_ISL_3084702, EPI_ISL_3084703, EPI_ISL_3084704, EPI_ISL_3084705, EPI_ISL_3084706, EPI_ISL_3084707, EPI_ISL_3084708, EPI_ISL_3084709, EPI_ISL_3084710, EPI_ISL_3084711, EPI_ISL_3084712, EPI_ISL_3084713, EPI_ISL_3084714, EPI_ISL_3084715, EPI_ISL_3084716, EPI_ISL_3084717, EPI_ISL_3084718, EPI_ISL_3084719, EPI_ISL_3084720, EPI_ISL_3084721,
EPI_ISL_3084722, EPI_ISL_3084723, EPI_ISL_3084724, EPI_ISL_3084725, EPI_ISL_3084726, EPI_ISL_3084727, EPI_ISL_3084728, EPI_ISL_3084729, EPI_ISL_3084730, EPI_ISL_3084731, EPI_ISL_3084732, EPI_ISL_3084733, EPI_ISL_3084734, EPI_ISL_3084735, EPI_ISL_3084736, EPI_ISL_3084737, EPI_ISL_3084738, EPI_ISL_3084739, EPI_ISL_3084740, EPI_ISL_3084741, EPI_ISL_3084742, EPI_ISL_3084743,
EPI_ISL_3084744, EPI_ISL_3084745, EPI_ISL_3084746, EPI_ISL_3084747, EPI_ISL_3084748, EPI_ISL_3084749, EPI_ISL_3084750, EPI_ISL_3084751, EPI_ISL_3084752, EPI_ISL_3084753, EPI_ISL_3084754, EPI_ISL_3084755, EPI_ISL_3084756, EPI_ISL_3084757, EPI_ISL_3084758, EPI_ISL_3084759, EPI_ISL_3084760, EPI_ISL_3084761, EPI_ISL_3084762, EPI_ISL_3084763, EPI_ISL_3084764, EPI_ISL_3084765,
EPI_ISL_3084766, EPI_ISL_3084767, EPI_ISL_3084768, EPI_ISL_3084769, EPI_ISL_3084770, EPI_ISL_3084771, EPI_ISL_3084772, EPI_ISL_3084773, EPI_ISL_3084774, EPI_ISL_3084775, EPI_ISL_3084776, EPI_ISL_3084777, EPI_ISL_3084778, EPI_ISL_3084779, EPI_ISL_3084780, EPI_ISL_3084781, EPI_ISL_3084782, EPI_ISL_3084783, EPI_ISL_3084784, EPI_ISL_3084785, EPI_ISL_3084786, EPI_ISL_3084787,
EPI_ISL_3084788, EPI_ISL_3084789, EPI_ISL_3084790, EPI_ISL_3084791, EPI_ISL_3084792, EPI_ISL_3084793, EPI_ISL_3084794, EPI_ISL_3084795, EPI_ISL_3084796, EPI_ISL_3084797, EPI_ISL_3084798, EPI_ISL_3084799, EPI_ISL_3084800, EPI_ISL_3084801, EPI_ISL_3084802, EPI_ISL_3084803, EPI_ISL_3084804, EPI_ISL_3084805, EPI_ISL_3084806, EPI_ISL_3084807, EPI_ISL_3084808, EPI_ISL_3084809,
EPI_ISL_3084810, EPI_ISL_3084811, EPI_ISL_3084812, EPI_ISL_3084813, EPI_ISL_3084814, EPI_ISL_3084815, EPI_ISL_3084816, EPI_ISL_3084817, EPI_ISL_3084818, EPI_ISL_3084819, EPI_ISL_3084820, EPI_ISL_3084821, EPI_ISL_3084822, EPI_ISL_3084823, EPI_ISL_3084824, EPI_ISL_3084825, EPI_ISL_3084826, EPI_ISL_3084827, EPI_ISL_3084828, EPI_ISL_3084829, EPI_ISL_3084830, EPI_ISL_3084831,
EPI_ISL_3084832, EPI_ISL_3084833, EPI_ISL_3084834, EPI_ISL_3084835, EPI_ISL_3084836, EPI_ISL_3084837, EPI_ISL_3084838, EPI_ISL_3084839, EPI_ISL_3084840, EPI_ISL_3084841, EPI_ISL_3084842, EPI_ISL_3084843, EPI_ISL_3084844, EPI_ISL_3084845, EPI_ISL_3084846, EPI_ISL_3084847, EPI_ISL_3084848, EPI_ISL_3084849, EPI_ISL_3084850, EPI_ISL_3084851, EPI_ISL_3084852, EPI_ISL_3084853,
EPI_ISL_3084855, EPI_ISL_3084856, EPI_ISL_3084857, EPI_ISL_3084858, EPI_ISL_3084859, EPI_ISL_3084860, EPI_ISL_3084861, EPI_ISL_3084862, EPI_ISL_3084863, EPI_ISL_3084864, EPI_ISL_3084865, EPI_ISL_3084866, EPI_ISL_3084867, EPI_ISL_3084868, EPI_ISL_3084869, EPI_ISL_3084870, EPI_ISL_3084871, EPI_ISL_3084872, EPI_ISL_3084873, EPI_ISL_3084874, EPI_ISL_3084875, EPI_ISL_3084876,
EPI_ISL_3084877, EPI_ISL_3084878, EPI_ISL_3084879, EPI_ISL_3084880, EPI_ISL_3084881, EPI_ISL_3084882, EPI_ISL_3084883, EPI_ISL_3084884, EPI_ISL_3084885, EPI_ISL_3084886, EPI_ISL_3084887, EPI_ISL_3084888, EPI_ISL_3084889, EPI_ISL_3084890, EPI_ISL_3084891, EPI_ISL_3084892, EPI_ISL_3084893, EPI_ISL_3084894, EPI_ISL_3084895, EPI_ISL_3084896, EPI_ISL_3084897, EPI_ISL_3084898,
EPI_ISL_3084899, EPI_ISL_3084900, EPI_ISL_3084901, EPI_ISL_3084902, EPI_ISL_3084903, EPI_ISL_3084904, EPI_ISL_3084905, EPI_ISL_3084906, EPI_ISL_3084907, EPI_ISL_3084908, EPI_ISL_3084909, EPI_ISL_3084910, EPI_ISL_3084911, EPI_ISL_3084912, EPI_ISL_3084913, EPI_ISL_3084914, EPI_ISL_3084915, EPI_ISL_3084916, EPI_ISL_3084917, EPI_ISL_3084918, EPI_ISL_3084919, EPI_ISL_3084920,
EPI_ISL_3084921, EPI_ISL_3084922, EPI_ISL_3084923, EPI_ISL_3084924, EPI_ISL_3084925, EPI_ISL_3084926, EPI_ISL_3084927, EPI_ISL_3084928, EPI_ISL_3084929, EPI_ISL_3084930, EPI_ISL_3084931, EPI_ISL_3084932, EPI_ISL_3084933, EPI_ISL_3084934, EPI_ISL_3084935, EPI_ISL_3084936, EPI_ISL_3084937, EPI_ISL_3084938, EPI_ISL_3084939, EPI_ISL_3084940, EPI_ISL_3084941, EPI_ISL_3084942,
EPI_ISL_3084943, EPI_ISL_3084944, EPI_ISL_3084945, EPI_ISL_3084946, EPI_ISL_3084947, EPI_ISL_3084948, EPI_ISL_3084949, EPI_ISL_3084950, EPI_ISL_3084951, EPI_ISL_3084952, EPI_ISL_3084953, EPI_ISL_3084954, EPI_ISL_3084955, EPI_ISL_3084956, EPI_ISL_3084957, EPI_ISL_3084958, EPI_ISL_3084959, EPI_ISL_3084960, EPI_ISL_3084961, EPI_ISL_3084962, EPI_ISL_3084963, EPI_ISL_3084964,
EPI_ISL_3084965, EPI_ISL_3084966, EPI_ISL_3084967, EPI_ISL_3084968, EPI_ISL_3084969, EPI_ISL_3084970, EPI_ISL_3084971, EPI_ISL_3084972, EPI_ISL_3084973, EPI_ISL_3084974, EPI_ISL_3084975, EPI_ISL_3084976, EPI_ISL_3084977, EPI_ISL_3084978, EPI_ISL_3084979, EPI_ISL_3084980, EPI_ISL_3084981, EPI_ISL_3084982, EPI_ISL_3084983, EPI_ISL_3084984, EPI_ISL_3084985, EPI_ISL_3084986,
EPI_ISL_3084987, EPI_ISL_3084988, EPI_ISL_3084989, EPI_ISL_3084990, EPI_ISL_3084991, EPI_ISL_3084992, EPI_ISL_3084993, EPI_ISL_3084994, EPI_ISL_3084995, EPI_ISL_3084996, EPI_ISL_3084997, EPI_ISL_3084998, EPI_ISL_3084999, EPI_ISL_3085000, EPI_ISL_3085001, EPI_ISL_3085002, EPI_ISL_3085003, EPI_ISL_3085004, EPI_ISL_3085005, EPI_ISL_3085006, EPI_ISL_3085007, EPI_ISL_3085008,
EPI_ISL_3085009, EPI_ISL_3085010, EPI_ISL_3085011, EPI_ISL_3085012, EPI_ISL_3085013, EPI_ISL_3085014, EPI_ISL_3085015, EPI_ISL_3085016, EPI_ISL_3085017, EPI_ISL_3085018, EPI_ISL_3085019, EPI_ISL_3085020, EPI_ISL_3085021, EPI_ISL_3085022, EPI_ISL_3085023, EPI_ISL_3085024, EPI_ISL_3085025, EPI_ISL_3085026, EPI_ISL_3085027, EPI_ISL_3085028, EPI_ISL_3085029, EPI_ISL_3085030,
EPI_ISL_3085031, EPI_ISL_3085032, EPI_ISL_3085033, EPI_ISL_3085034, EPI_ISL_3085035, EPI_ISL_3085036, EPI_ISL_3085037, EPI_ISL_3085038, EPI_ISL_3085039, EPI_ISL_3085040, EPI_ISL_3085041, EPI_ISL_3085042, EPI_ISL_3085043, EPI_ISL_3085044, EPI_ISL_3085045, EPI_ISL_3085046, EPI_ISL_3085047, EPI_ISL_3085048, EPI_ISL_3085050, EPI_ISL_3085052, EPI_ISL_3085054, EPI_ISL_3085057,
EPI_ISL_3085058, EPI_ISL_3085060, EPI_ISL_3085062, EPI_ISL_3085064, EPI_ISL_3085066, EPI_ISL_3085068, EPI_ISL_3085070, EPI_ISL_3085072, EPI_ISL_3085074, EPI_ISL_3085076, EPI_ISL_3085078, EPI_ISL_3085080, EPI_ISL_3085082, EPI_ISL_3085084, EPI_ISL_3085086, EPI_ISL_3085088, EPI_ISL_3085090, EPI_ISL_3085092, EPI_ISL_3085094, EPI_ISL_3085096, EPI_ISL_3085098, EPI_ISL_3085100,
EPI_ISL_3085102, EPI_ISL_3085103, EPI_ISL_3085104, EPI_ISL_3127202, EPI_ISL_3127203, EPI_ISL_3127204, EPI_ISL_3127205, EPI_ISL_3127206, EPI_ISL_3127207, EPI_ISL_3127208, EPI_ISL_3127209, EPI_ISL_3127210, EPI_ISL_3127211, EPI_ISL_3127212, EPI_ISL_3127213, EPI_ISL_3127214, EPI_ISL_3127215, EPI_ISL_3127216, EPI_ISL_3127217, EPI_ISL_3127218, EPI_ISL_3127219, EPI_ISL_3127220,
EPI_ISL_3127221, EPI_ISL_3127222, EPI_ISL_3127223, EPI_ISL_3127224, EPI_ISL_3127225, EPI_ISL_3127226, EPI_ISL_3127227, EPI_ISL_3127228, EPI_ISL_3127229, EPI_ISL_3127230, EPI_ISL_3127231, EPI_ISL_3127232, EPI_ISL_3127233, EPI_ISL_3127234, EPI_ISL_3127235, EPI_ISL_3127236, EPI_ISL_3127237, EPI_ISL_3127238, EPI_ISL_3127239, EPI_ISL_3127240, EPI_ISL_3127241, EPI_ISL_3127242,
EPI_ISL_3127243, EPI_ISL_3127244, EPI_ISL_3127245, EPI_ISL_3127246, EPI_ISL_3127247, EPI_ISL_3127248, EPI_ISL_3127249, EPI_ISL_3127250, EPI_ISL_3127251, EPI_ISL_3127252, EPI_ISL_3127253, EPI_ISL_3127254, EPI_ISL_3127255, EPI_ISL_3127256, EPI_ISL_3127257, EPI_ISL_3127258, EPI_ISL_3127259, EPI_ISL_3127260, EPI_ISL_3127261, EPI_ISL_3127262, EPI_ISL_3127263, EPI_ISL_3127264,
EPI_ISL_3127265, EPI_ISL_3127266, EPI_ISL_3127267, EPI_ISL_3127268, EPI_ISL_3127269, EPI_ISL_3127270, EPI_ISL_3127271, EPI_ISL_3127272, EPI_ISL_3127273, EPI_ISL_3127274, EPI_ISL_3127275, EPI_ISL_3127276, EPI_ISL_3127277, EPI_ISL_3127278, EPI_ISL_3127279, EPI_ISL_3127280, EPI_ISL_3127281, EPI_ISL_3127282, EPI_ISL_3127283, EPI_ISL_3127284, EPI_ISL_3127285, EPI_ISL_3127286,
EPI_ISL_3127287, EPI_ISL_3127288, EPI_ISL_3127289, EPI_ISL_3127290, EPI_ISL_3127291, EPI_ISL_3127292, EPI_ISL_3127293, EPI_ISL_3127294, EPI_ISL_3127295, EPI_ISL_3127296, EPI_ISL_3127297, EPI_ISL_3127298, EPI_ISL_3127299, EPI_ISL_3127300, EPI_ISL_3127301, EPI_ISL_3127302, EPI_ISL_3127303, EPI_ISL_3127304, EPI_ISL_3127305, EPI_ISL_3127306, EPI_ISL_3127307, EPI_ISL_3127308,
EPI_ISL_3127309, EPI_ISL_3127310, EPI_ISL_3127311, EPI_ISL_3127312, EPI_ISL_3127313, EPI_ISL_3127314, EPI_ISL_3127315, EPI_ISL_3127316, EPI_ISL_3127317, EPI_ISL_3127318, EPI_ISL_3127319, EPI_ISL_3127320, EPI_ISL_3127321, EPI_ISL_3127322, EPI_ISL_3127323, EPI_ISL_3127324, EPI_ISL_3127325, EPI_ISL_3127326, EPI_ISL_3127327, EPI_ISL_3127328, EPI_ISL_3127329, EPI_ISL_3127330,
EPI_ISL_3127331, EPI_ISL_3127332, EPI_ISL_3127333, EPI_ISL_3127334, EPI_ISL_3127335, EPI_ISL_3127336, EPI_ISL_3127337, EPI_ISL_3127338, EPI_ISL_3127339, EPI_ISL_3127340, EPI_ISL_3127341, EPI_ISL_3127342, EPI_ISL_3127343, EPI_ISL_3127344, EPI_ISL_3127345, EPI_ISL_3127346, EPI_ISL_3127347, EPI_ISL_3127348, EPI_ISL_3127349, EPI_ISL_3127350, EPI_ISL_3127351, EPI_ISL_3127352,
EPI_ISL_3127353, EPI_ISL_3127354, EPI_ISL_3127355, EPI_ISL_3127356, EPI_ISL_3127357, EPI_ISL_3127358, EPI_ISL_3127359, EPI_ISL_3127360, EPI_ISL_3127361, EPI_ISL_3127362, EPI_ISL_3127363, EPI_ISL_3127364, EPI_ISL_3127365, EPI_ISL_3127366, EPI_ISL_3127367, EPI_ISL_3127368, EPI_ISL_3127369, EPI_ISL_3127370, EPI_ISL_3127371, EPI_ISL_3127372, EPI_ISL_3127373, EPI_ISL_3127374,
EPI_ISL_3127375, EPI_ISL_3127376, EPI_ISL_3127377, EPI_ISL_3127378, EPI_ISL_3127379, EPI_ISL_3127380, EPI_ISL_3127381, EPI_ISL_3127382, EPI_ISL_3127383, EPI_ISL_3127384, EPI_ISL_3127385, EPI_ISL_3127386, EPI_ISL_3127387, EPI_ISL_3127388, EPI_ISL_3134285, EPI_ISL_3134286, EPI_ISL_3134287, EPI_ISL_3134288, EPI_ISL_3134289, EPI_ISL_3134290, EPI_ISL_3134291, EPI_ISL_3134292,
EPI_ISL_3134293, EPI_ISL_3134294, EPI_ISL_3134295, EPI_ISL_3134296, EPI_ISL_3134297, EPI_ISL_3134298, EPI_ISL_3134299, EPI_ISL_3134300, EPI_ISL_3134301, EPI_ISL_3134302, EPI_ISL_3134303, EPI_ISL_3134304, EPI_ISL_3134305, EPI_ISL_3134306, EPI_ISL_3134307, EPI_ISL_3134308, EPI_ISL_3134309, EPI_ISL_3134310, EPI_ISL_3134311, EPI_ISL_3134312, EPI_ISL_3134313, EPI_ISL_3134314,
EPI_ISL_3134315, EPI_ISL_3134316, EPI_ISL_3134317, EPI_ISL_3134318, EPI_ISL_3134319, EPI_ISL_3134320, EPI_ISL_3134321, EPI_ISL_3134322, EPI_ISL_3134323, EPI_ISL_3134324, EPI_ISL_3134325, EPI_ISL_3134326, EPI_ISL_3134327, EPI_ISL_3134328, EPI_ISL_3134329, EPI_ISL_3134330, EPI_ISL_3134331, EPI_ISL_3134332, EPI_ISL_3134333, EPI_ISL_3134334, EPI_ISL_3134335, EPI_ISL_3134336,
EPI_ISL_3134337, EPI_ISL_3134338, EPI_ISL_3134339, EPI_ISL_3134340, EPI_ISL_3134341, EPI_ISL_3134342, EPI_ISL_3134343, EPI_ISL_3134344, EPI_ISL_3134345, EPI_ISL_3134346, EPI_ISL_3134347, EPI_ISL_3134348, EPI_ISL_3134349, EPI_ISL_3134350, EPI_ISL_3134351, EPI_ISL_3134352, EPI_ISL_3134353, EPI_ISL_3134354, EPI_ISL_3134355, EPI_ISL_3134356, EPI_ISL_3134357, EPI_ISL_3134358,
EPI_ISL_3134359, EPI_ISL_3134360, EPI_ISL_3134361, EPI_ISL_3134362, EPI_ISL_3134363, EPI_ISL_3134364, EPI_ISL_3134365, EPI_ISL_3134366, EPI_ISL_3134367, EPI_ISL_3134368, EPI_ISL_3134369, EPI_ISL_3134370, EPI_ISL_3134371, EPI_ISL_3134372, EPI_ISL_3134373, EPI_ISL_3134374, EPI_ISL_3134375, EPI_ISL_3134376, EPI_ISL_3134377, EPI_ISL_3134378, EPI_ISL_3134379, EPI_ISL_3134380,
EPI_ISL_3134381, EPI_ISL_3134382, EPI_ISL_3134383, EPI_ISL_3134384, EPI_ISL_3134385, EPI_ISL_3134386, EPI_ISL_3134387, EPI_ISL_3134388, EPI_ISL_3134389, EPI_ISL_3134390, EPI_ISL_3134391, EPI_ISL_3134392, EPI_ISL_3134393, EPI_ISL_3134394, EPI_ISL_3134395, EPI_ISL_3134396, EPI_ISL_3134397, EPI_ISL_3134398, EPI_ISL_3134399, EPI_ISL_3134400, EPI_ISL_3134401, EPI_ISL_3134402,
EPI_ISL_3134403, EPI_ISL_3134404, EPI_ISL_3134405, EPI_ISL_3134406, EPI_ISL_3134407, EPI_ISL_3134408, EPI_ISL_3134409, EPI_ISL_3134410, EPI_ISL_3134411, EPI_ISL_3134412, EPI_ISL_3134413, EPI_ISL_3134414, EPI_ISL_3134415, EPI_ISL_3134416, EPI_ISL_3134417, EPI_ISL_3134418, EPI_ISL_3134419, EPI_ISL_3134420, EPI_ISL_3134421, EPI_ISL_3134422, EPI_ISL_3134423, EPI_ISL_3134424,
EPI_ISL_3134425, EPI_ISL_3134426, EPI_ISL_3134427, EPI_ISL_3134428, EPI_ISL_3134429, EPI_ISL_3134430, EPI_ISL_3134431, EPI_ISL_3134432, EPI_ISL_3134433, EPI_ISL_3134434, EPI_ISL_3134435, EPI_ISL_3134436, EPI_ISL_3134437, EPI_ISL_3134438, EPI_ISL_3134439, EPI_ISL_3134440, EPI_ISL_3134441, EPI_ISL_3134442, EPI_ISL_3134443, EPI_ISL_3134444, EPI_ISL_3134445, EPI_ISL_3134446,
EPI_ISL_3134447, EPI_ISL_3134448, EPI_ISL_3134449, EPI_ISL_3134450, EPI_ISL_3134451, EPI_ISL_3134452, EPI_ISL_3134453, EPI_ISL_3134454, EPI_ISL_3134455, EPI_ISL_3134456, EPI_ISL_3134457, EPI_ISL_3134458, EPI_ISL_3134459, EPI_ISL_3134460, EPI_ISL_3134461, EPI_ISL_3134462, EPI_ISL_3134463, EPI_ISL_3134464, EPI_ISL_3134465, EPI_ISL_3134466, EPI_ISL_3134467, EPI_ISL_3134468,
EPI_ISL_3134469, EPI_ISL_3134470, EPI_ISL_3134471, EPI_ISL_3134472, EPI_ISL_3134473, EPI_ISL_3134474, EPI_ISL_3134475, EPI_ISL_3134476, EPI_ISL_3134477, EPI_ISL_3134478, EPI_ISL_3134479, EPI_ISL_3134480, EPI_ISL_3134481, EPI_ISL_3134482, EPI_ISL_3134483, EPI_ISL_3134484, EPI_ISL_3134485, EPI_ISL_3134486, EPI_ISL_3134487, EPI_ISL_3134488, EPI_ISL_3134489, EPI_ISL_3134490,
EPI_ISL_3134491, EPI_ISL_3134492, EPI_ISL_3134493, EPI_ISL_3134494, EPI_ISL_3134495, EPI_ISL_3134496, EPI_ISL_3134497, EPI_ISL_3134498, EPI_ISL_3134499, EPI_ISL_3134500, EPI_ISL_3134501, EPI_ISL_3134502, EPI_ISL_3134503, EPI_ISL_3134504, EPI_ISL_3134505, EPI_ISL_3134506, EPI_ISL_3134507, EPI_ISL_3134508, EPI_ISL_3134509, EPI_ISL_3134510, EPI_ISL_3134511, EPI_ISL_3134512,
EPI_ISL_3134513, EPI_ISL_3134514, EPI_ISL_3134515, EPI_ISL_3134516, EPI_ISL_3134517, EPI_ISL_3134518, EPI_ISL_3134519, EPI_ISL_3134520, EPI_ISL_3134521, EPI_ISL_3134522, EPI_ISL_3134523, EPI_ISL_3134524, EPI_ISL_3134525, EPI_ISL_3134526, EPI_ISL_3134527, EPI_ISL_3134528, EPI_ISL_3134529, EPI_ISL_3134530, EPI_ISL_3134531, EPI_ISL_3134532, EPI_ISL_3134533, EPI_ISL_3134534,
EPI_ISL_3134535, EPI_ISL_3152941, EPI_ISL_3152942, EPI_ISL_3152943, EPI_ISL_3152944, EPI_ISL_3152945, EPI_ISL_3152946, EPI_ISL_3152947, EPI_ISL_3152948, EPI_ISL_3152949, EPI_ISL_3152950, EPI_ISL_3152951, EPI_ISL_3152952, EPI_ISL_3152953, EPI_ISL_3152954, EPI_ISL_3152955, EPI_ISL_3152956, EPI_ISL_3152957, EPI_ISL_3152958, EPI_ISL_3152959, EPI_ISL_3152960, EPI_ISL_3152961,
EPI_ISL_3152962, EPI_ISL_3152963, EPI_ISL_3152964, EPI_ISL_3152965, EPI_ISL_3152966, EPI_ISL_3152967, EPI_ISL_3152968, EPI_ISL_3152969, EPI_ISL_3152970, EPI_ISL_3152971, EPI_ISL_3152972, EPI_ISL_3152973, EPI_ISL_3152974, EPI_ISL_3152975, EPI_ISL_3152976, EPI_ISL_3152977, EPI_ISL_3152978, EPI_ISL_3152979, EPI_ISL_3152980, EPI_ISL_3152981, EPI_ISL_3152982, EPI_ISL_3152983,
EPI_ISL_3152984, EPI_ISL_3152985, EPI_ISL_3152986, EPI_ISL_3152987, EPI_ISL_3152988, EPI_ISL_3152989, EPI_ISL_3152990, EPI_ISL_3152991, EPI_ISL_3152992, EPI_ISL_3152993, EPI_ISL_3152994, EPI_ISL_3152995, EPI_ISL_3152996, EPI_ISL_3152997, EPI_ISL_3152998, EPI_ISL_3152999, EPI_ISL_3153000, EPI_ISL_3153001, EPI_ISL_3153002, EPI_ISL_3153003, EPI_ISL_3153004, EPI_ISL_3153005,
EPI_ISL_3153006, EPI_ISL_3153007, EPI_ISL_3153008, EPI_ISL_3153009, EPI_ISL_3153010, EPI_ISL_3153011, EPI_ISL_3153012, EPI_ISL_3153013, EPI_ISL_3153014, EPI_ISL_3153015, EPI_ISL_3153016, EPI_ISL_3153017, EPI_ISL_3153018, EPI_ISL_3153019, EPI_ISL_3153020, EPI_ISL_3153021, EPI_ISL_3153022, EPI_ISL_3153023, EPI_ISL_3153024, EPI_ISL_3153025, EPI_ISL_3153026, EPI_ISL_3153027,
EPI_ISL_3153028, EPI_ISL_3153029, EPI_ISL_3153030, EPI_ISL_3153031, EPI_ISL_3153032, EPI_ISL_3153033, EPI_ISL_3153034, EPI_ISL_3153035, EPI_ISL_3153036, EPI_ISL_3153037, EPI_ISL_3153038, EPI_ISL_3153039, EPI_ISL_3153040, EPI_ISL_3153041, EPI_ISL_3153042, EPI_ISL_3153043, EPI_ISL_3153044, EPI_ISL_3153045, EPI_ISL_3153046, EPI_ISL_3153047, EPI_ISL_3153048, EPI_ISL_3153049,
EPI_ISL_3153050, EPI_ISL_3153051, EPI_ISL_3153052, EPI_ISL_3153053, EPI_ISL_3153054, EPI_ISL_3153055, EPI_ISL_3153056, EPI_ISL_3153057, EPI_ISL_3153058, EPI_ISL_3153059, EPI_ISL_3153060, EPI_ISL_3153061, EPI_ISL_3153062, EPI_ISL_3153063, EPI_ISL_3153064, EPI_ISL_3153065, EPI_ISL_3153066, EPI_ISL_3153067, EPI_ISL_3153068, EPI_ISL_3153069, EPI_ISL_3153070, EPI_ISL_3153071,
EPI_ISL_3153072, EPI_ISL_3153073, EPI_ISL_3153074, EPI_ISL_3153075, EPI_ISL_3153076, EPI_ISL_3153077, EPI_ISL_3153078, EPI_ISL_3153079, EPI_ISL_3153080, EPI_ISL_3153081, EPI_ISL_3153082, EPI_ISL_3153083, EPI_ISL_3153084, EPI_ISL_3153085, EPI_ISL_3153086, EPI_ISL_3153087, EPI_ISL_3153088, EPI_ISL_3153089, EPI_ISL_3153090, EPI_ISL_3153091, EPI_ISL_3153092, EPI_ISL_3153093,
EPI_ISL_3153094, EPI_ISL_3153095, EPI_ISL_3153096, EPI_ISL_3153097, EPI_ISL_3153098, EPI_ISL_3153099, EPI_ISL_3153100, EPI_ISL_3153101, EPI_ISL_3153102, EPI_ISL_3153103, EPI_ISL_3153104, EPI_ISL_3153105, EPI_ISL_3153106, EPI_ISL_3153107, EPI_ISL_3153108, EPI_ISL_3153109, EPI_ISL_3153110, EPI_ISL_3153111, EPI_ISL_3153112, EPI_ISL_3153113, EPI_ISL_3153114, EPI_ISL_3153115,
EPI_ISL_3153116, EPI_ISL_3153117, EPI_ISL_3153118, EPI_ISL_3153119, EPI_ISL_3153120, EPI_ISL_3153121, EPI_ISL_3153122, EPI_ISL_3153123, EPI_ISL_3153124, EPI_ISL_3153125, EPI_ISL_3153126, EPI_ISL_3153127, EPI_ISL_3153128, EPI_ISL_3153129, EPI_ISL_3153130, EPI_ISL_3153131, EPI_ISL_3153132, EPI_ISL_3153133, EPI_ISL_3153134, EPI_ISL_3153135, EPI_ISL_3153136, EPI_ISL_3153137,

| | | | |
|---|---|---|---|
| EPI_ISL_3153138, EPI_ISL_3153139, EPI_ISL_3153140, EPI_ISL_3153141, EPI_ISL_3153142, EPI_ISL_3153143, EPI_ISL_3153144, EPI_ISL_3153145, EPI_ISL_3153146, EPI_ISL_3153147, EPI_ISL_3153148, EPI_ISL_3153149, EPI_ISL_3153150, EPI_ISL_3153151, EPI_ISL_3153152, EPI_ISL_3153153, EPI_ISL_3153154, EPI_ISL_3153155, EPI_ISL_3153156, EPI_ISL_3153157, EPI_ISL_3153158, EPI_ISL_3153159, EPI_ISL_3153160, EPI_ISL_3153161, EPI_ISL_3153162, EPI_ISL_3153163, EPI_ISL_3153164, EPI_ISL_3153165, EPI_ISL_3153166, EPI_ISL_3153167, EPI_ISL_3153168, EPI_ISL_3153169, EPI_ISL_3153170, EPI_ISL_3153171, EPI_ISL_3153172, EPI_ISL_3153173, EPI_ISL_3153174, EPI_ISL_3153175, EPI_ISL_3155732, EPI_ISL_3155733, EPI_ISL_3155734, EPI_ISL_3155735, EPI_ISL_3155736, EPI_ISL_3155737, EPI_ISL_3155738, EPI_ISL_3155739, EPI_ISL_3155740, EPI_ISL_3155741, EPI_ISL_3155742, EPI_ISL_3155743, EPI_ISL_3155744, EPI_ISL_3155745, EPI_ISL_3155746, EPI_ISL_3155747, EPI_ISL_3155748, EPI_ISL_3155749, EPI_ISL_3155750, EPI_ISL_3155751, EPI_ISL_3155752, EPI_ISL_3155753, EPI_ISL_3155754, EPI_ISL_3155755, EPI_ISL_3155756, EPI_ISL_3155757, EPI_ISL_3155758, EPI_ISL_3155759, EPI_ISL_3155760, EPI_ISL_3155761, EPI_ISL_3155762, EPI_ISL_3155763, EPI_ISL_3155764, EPI_ISL_3155765, EPI_ISL_3155766, EPI_ISL_3155767, EPI_ISL_3155768, EPI_ISL_3155769, EPI_ISL_3155770, EPI_ISL_3155771, EPI_ISL_3155772, EPI_ISL_3155773, EPI_ISL_3155774, EPI_ISL_3155775, EPI_ISL_3155776, EPI_ISL_3155777, EPI_ISL_3155778, EPI_ISL_3155779, EPI_ISL_3155780, EPI_ISL_3155781, EPI_ISL_3155782, EPI_ISL_3155783, EPI_ISL_3155784, EPI_ISL_3155785, EPI_ISL_3155786, EPI_ISL_3155787, EPI_ISL_3155788, EPI_ISL_3155789, EPI_ISL_3155790, EPI_ISL_3155791, EPI_ISL_3155792, EPI_ISL_3155793, EPI_ISL_3155794, EPI_ISL_3155795, EPI_ISL_3155796, EPI_ISL_3155797, EPI_ISL_3155798, EPI_ISL_3155799, EPI_ISL_3155800, EPI_ISL_3155801, EPI_ISL_3155802, EPI_ISL_3155803, EPI_ISL_3155804, EPI_ISL_3155805, EPI_ISL_3155806, EPI_ISL_3155807, EPI_ISL_3155808, EPI_ISL_3155809, EPI_ISL_3155810, EPI_ISL_3155811, EPI_ISL_3155812, EPI_ISL_3155813, EPI_ISL_3155814, EPI_ISL_3155815, EPI_ISL_3155816, EPI_ISL_3155817, EPI_ISL_3155818, EPI_ISL_3155819, EPI_ISL_3155820, EPI_ISL_3155821, EPI_ISL_3155822, EPI_ISL_3155823, EPI_ISL_3155824, EPI_ISL_3155825, EPI_ISL_3155826, EPI_ISL_3155827, EPI_ISL_3155828, EPI_ISL_3155829, EPI_ISL_3155830, EPI_ISL_3155831, EPI_ISL_3155832, EPI_ISL_3155833, EPI_ISL_3155834, EPI_ISL_3155835, EPI_ISL_3155836, EPI_ISL_3155837, EPI_ISL_3155838, EPI_ISL_3155839, EPI_ISL_3155840, EPI_ISL_3155841, EPI_ISL_3155842, EPI_ISL_3155843, EPI_ISL_3155844, EPI_ISL_3155845, EPI_ISL_3155846, EPI_ISL_3155847, EPI_ISL_3155848, EPI_ISL_3155849, EPI_ISL_3155850, EPI_ISL_3155851, EPI_ISL_3155852, EPI_ISL_3155853, EPI_ISL_3155854, EPI_ISL_3155855, EPI_ISL_3155856, EPI_ISL_3155857, EPI_ISL_3155858, EPI_ISL_3155859, EPI_ISL_3155860, EPI_ISL_3155861, EPI_ISL_3155862, EPI_ISL_3155863, EPI_ISL_3155864, EPI_ISL_3155865, EPI_ISL_3155866, EPI_ISL_3155867, EPI_ISL_3155868, EPI_ISL_3155869, EPI_ISL_3155870, EPI_ISL_3155871, EPI_ISL_3155872, EPI_ISL_3155873, EPI_ISL_3155874, EPI_ISL_3155875, EPI_ISL_3155876, EPI_ISL_3155877, EPI_ISL_3155878, EPI_ISL_3155879, EPI_ISL_3155880, EPI_ISL_3155881, EPI_ISL_3155882, EPI_ISL_3155883, EPI_ISL_3155884, EPI_ISL_3155885, EPI_ISL_3155886, EPI_ISL_3155887, EPI_ISL_3155888, EPI_ISL_3155889, EPI_ISL_3155890, EPI_ISL_3155891, EPI_ISL_3155892, EPI_ISL_3155893, EPI_ISL_3155894, EPI_ISL_3155895, EPI_ISL_3155896, EPI_ISL_3155897, EPI_ISL_3155898, EPI_ISL_3155899, EPI_ISL_3155900, EPI_ISL_3155901, EPI_ISL_3155902, EPI_ISL_3155903, EPI_ISL_3155904, EPI_ISL_3155905, EPI_ISL_3155906, EPI_ISL_3155907, EPI_ISL_3155908, EPI_ISL_3155909, EPI_ISL_3155910, EPI_ISL_3155911, EPI_ISL_3155912, EPI_ISL_3155913, EPI_ISL_3155914, EPI_ISL_3155915, EPI_ISL_3155916, EPI_ISL_3155917, EPI_ISL_3155918, EPI_ISL_3155919, EPI_ISL_3155920, EPI_ISL_3155921, EPI_ISL_3155922, EPI_ISL_3155923, EPI_ISL_3155924, EPI_ISL_3155925, EPI_ISL_3155926, EPI_ISL_3155927, EPI_ISL_3155928, EPI_ISL_3155929, EPI_ISL_3155930, EPI_ISL_3155931, EPI_ISL_3155932, EPI_ISL_3155933, EPI_ISL_3155934, EPI_ISL_3155935, EPI_ISL_3155936, EPI_ISL_3155937, EPI_ISL_3155938, EPI_ISL_3155939, EPI_ISL_3155940, EPI_ISL_3155941, EPI_ISL_3155942, EPI_ISL_3155943, EPI_ISL_3155944, EPI_ISL_3155945, EPI_ISL_3155946, EPI_ISL_3155948, EPI_ISL_3155950, EPI_ISL_3155952, EPI_ISL_3155955, EPI_ISL_3155958, EPI_ISL_3155960, EPI_ISL_3155963, EPI_ISL_3155965, EPI_ISL_3155968, EPI_ISL_3155971, EPI_ISL_3155973, EPI_ISL_3155976, EPI_ISL_3155978, EPI_ISL_3155981, EPI_ISL_3155983, EPI_ISL_3155986, EPI_ISL_3155988, EPI_ISL_3155991, EPI_ISL_3155993, EPI_ISL_3155995, EPI_ISL_3155996, EPI_ISL_3155997, EPI_ISL_3155998, EPI_ISL_3155999, EPI_ISL_3156000, EPI_ISL_3156001, EPI_ISL_3156002, EPI_ISL_3156003, EPI_ISL_3156004, EPI_ISL_3156005, EPI_ISL_3156006, EPI_ISL_3156007, EPI_ISL_3156008, EPI_ISL_3156009, EPI_ISL_3156010, EPI_ISL_3156011, EPI_ISL_3156012, EPI_ISL_3156013, EPI_ISL_3156014, EPI_ISL_3156015, EPI_ISL_3156016 | | | |
| see above | Broad Institute Clinical Research Sequencing Platform | Infectious Disease Program, Broad Institute of Harvard and MIT | Adams, G.; B.L.; B.W.; Bauer, M.; Birren; Blumenstiel, B.; Brown, C.; Carter, A.; Chaluvadi, S.; D.J.; DeFelice, M.; DeRuff, K.; Dodge, S.; Gabriel, S.; Gallagher, G.; Gladden-Young, A.; Granger, B.; J.E.; K.J.; Lagerborg, K.; Larkin, K.; Lee, M.; Lemieux; Lennon, N.; Loreth, C.; Madoff, L.; McGovern, S.; Meldrim, J.; Normandin, E.; P.C.; Park; Pearlman, L.; Reilly, S.; Rudy, M.; Sabeti; Siddle; Smole, S.; Tomkins-Tinch, C.; Vicente, G.; and MacInnis |
| EPI_ISL_2106202, EPI_ISL_2106205, EPI_ISL_2106206, EPI_ISL_2349549, EPI_ISL_2349550, EPI_ISL_2349551 | DPH, Massachusetts State Public Health Lab | DPH, Massachusetts State Public Health Lab | A.S.; Fink, T.; G.R.; Gallagher; Lang; S.C.; Smole |
| EPI_ISL_2502578, EPI_ISL_2811829 | DPH, Massachusetts State Public Health Lab | DPH, Massachusetts State Public Health Lab | A.S.; Fink, T.; G.R. and Smole; Gallagher; Lang; S.C. |
| EPI_ISL_2037927, EPI_ISL_2038185, EPI_ISL_2134551, EPI_ISL_2280252, EPI_ISL_3064798, EPI_ISL_3212215, EPI_ISL_3299970, EPI_ISL_3302565, EPI_ISL_3344245 | | | |
| see above | Fulgent Genetics | Centers for Disease Control and Prevention Division of Viral Diseases, Pathogen Discovery | Adrian Paskey; Becky Tsai; Benafsh Sapra; Benjamin Rambo-Martin; Christopher Gulvick; Clinton R. Paden; Dakota Howard; Darlene Wagner; Dhwani Batra; Doreen Ng; Duncan MacCannell; Harry Gao; James Xie; Jason Caravas; John Gao; Joseph Fierro; Kara Moser; Matthew Schmerer; Mickey Li; Peter W. Cook; Scott Sammons; Shatavia Morrison; Yan Meng; Yvette Unoarumhi |
| EPI_ISL_1990576, EPI_ISL_1990615, EPI_ISL_1990721, EPI_ISL_1990839, EPI_ISL_1990851, EPI_ISL_1990854, EPI_ISL_1990870, EPI_ISL_1990912, EPI_ISL_1990919, EPI_ISL_1990921, EPI_ISL_1990927, EPI_ISL_1990930, EPI_ISL_1990953, EPI_ISL_1990961, EPI_ISL_1990990, EPI_ISL_1991017, EPI_ISL_1991027, EPI_ISL_1991028, EPI_ISL_1991029, EPI_ISL_1991039, EPI_ISL_1991114, EPI_ISL_1991171, EPI_ISL_1991175, EPI_ISL_1991214, EPI_ISL_1991226, EPI_ISL_1991227, EPI_ISL_1991237, EPI_ISL_1991238, EPI_ISL_1991372, EPI_ISL_1991409, EPI_ISL_1991427, EPI_ISL_1991436, EPI_ISL_1991450, EPI_ISL_1991460, EPI_ISL_1991469, EPI_ISL_1991473, EPI_ISL_1991565, EPI_ISL_1991567, EPI_ISL_1991575, EPI_ISL_1991579, EPI_ISL_1991581, EPI_ISL_1991586, EPI_ISL_1991591, EPI_ISL_1991592, EPI_ISL_1991598, EPI_ISL_1991642, EPI_ISL_1991663, EPI_ISL_1991676, EPI_ISL_1991677, EPI_ISL_1991701, EPI_ISL_1991704, EPI_ISL_1991705, EPI_ISL_1991712, EPI_ISL_1991726, EPI_ISL_1991765, EPI_ISL_2009846, EPI_ISL_2009853, EPI_ISL_2009854, EPI_ISL_2009875, EPI_ISL_2009883, EPI_ISL_2009885, EPI_ISL_2009895, EPI_ISL_2009906, EPI_ISL_2009914, EPI_ISL_2010001, EPI_ISL_2010048, EPI_ISL_2010061, EPI_ISL_2010071, EPI_ISL_2010121, EPI_ISL_2010137, EPI_ISL_2010139, EPI_ISL_2010202, EPI_ISL_2010218, EPI_ISL_2010239, EPI_ISL_2010242, EPI_ISL_2010245, EPI_ISL_2010268, EPI_ISL_2010279, EPI_ISL_2010310, EPI_ISL_2010332, EPI_ISL_2010351, EPI_ISL_2010404, EPI_ISL_2010425, EPI_ISL_2010428, EPI_ISL_2010431, EPI_ISL_2010529, EPI_ISL_2010574, EPI_ISL_2010576, EPI_ISL_2010580, EPI_ISL_2010584, EPI_ISL_2010626, EPI_ISL_2010630, EPI_ISL_2010631, EPI_ISL_2010637, EPI_ISL_2010731, EPI_ISL_2010732, EPI_ISL_2010754, EPI_ISL_2010755, EPI_ISL_2010779, EPI_ISL_2010812, EPI_ISL_2010824, EPI_ISL_2010832, EPI_ISL_2097358, EPI_ISL_2097368, EPI_ISL_2097392, EPI_ISL_2097418, EPI_ISL_2097493, EPI_ISL_2097513, EPI_ISL_2097538, EPI_ISL_2097550, EPI_ISL_2097572, EPI_ISL_2097592, EPI_ISL_2097604, EPI_ISL_2097610, EPI_ISL_2097614, EPI_ISL_2097631, EPI_ISL_2097635, EPI_ISL_2097648, EPI_ISL_2097649, EPI_ISL_2097654, EPI_ISL_2097659, EPI_ISL_2097681, EPI_ISL_2097754, EPI_ISL_2097786, EPI_ISL_2097801, EPI_ISL_2097832, EPI_ISL_2097857, EPI_ISL_2097881, EPI_ISL_2098015, EPI_ISL_2098036, EPI_ISL_2098037, EPI_ISL_2098047, EPI_ISL_2098061, EPI_ISL_2098075, EPI_ISL_2098163, EPI_ISL_2098188, EPI_ISL_2098190, EPI_ISL_2144077, EPI_ISL_2144104, EPI_ISL_2144116, EPI_ISL_2144126, EPI_ISL_2144137, EPI_ISL_2144138, EPI_ISL_2144139, EPI_ISL_2144149, EPI_ISL_2144245, EPI_ISL_2144270, EPI_ISL_2144321, EPI_ISL_2144360, EPI_ISL_2144370, EPI_ISL_2144390, EPI_ISL_2144397, EPI_ISL_2144399, EPI_ISL_2144400, EPI_ISL_2144539, EPI_ISL_2144540, EPI_ISL_2144626, EPI_ISL_2144670, EPI_ISL_2144716, EPI_ISL_2144717, EPI_ISL_2144847, EPI_ISL_2144956, EPI_ISL_2144957, EPI_ISL_2144959, EPI_ISL_2144961, EPI_ISL_2144977, EPI_ISL_2144982, EPI_ISL_2145029, EPI_ISL_2145032, EPI_ISL_2145037, EPI_ISL_2145052, EPI_ISL_2145054, EPI_ISL_2145060, EPI_ISL_2145061, EPI_ISL_2145068, EPI_ISL_2145070, EPI_ISL_2145090, EPI_ISL_2145132, EPI_ISL_2145201, EPI_ISL_2145306, EPI_ISL_2145325, EPI_ISL_2145384, EPI_ISL_2145433, EPI_ISL_2145467, EPI_ISL_2145492, EPI_ISL_2145512, EPI_ISL_2145546, EPI_ISL_2145556, EPI_ISL_2145600, EPI_ISL_2145603, EPI_ISL_2145663, EPI_ISL_2159477, EPI_ISL_2159485, EPI_ISL_2159497, EPI_ISL_2159508, EPI_ISL_2159524, EPI_ISL_2159526, EPI_ISL_2159529, EPI_ISL_2159535, EPI_ISL_2159538, EPI_ISL_2159547, EPI_ISL_2159557, EPI_ISL_2159702, EPI_ISL_2159716, EPI_ISL_2159734, EPI_ISL_2159736, EPI_ISL_2159744, EPI_ISL_2159764, EPI_ISL_2159767, EPI_ISL_2159775, EPI_ISL_2159776, EPI_ISL_2159778, EPI_ISL_2159784, EPI_ISL_2159786, EPI_ISL_2159814, EPI_ISL_2159819, EPI_ISL_2159824, EPI_ISL_2159826, EPI_ISL_2159836, EPI_ISL_2159889, EPI_ISL_2160021, EPI_ISL_2160114, EPI_ISL_2160121, EPI_ISL_2160140, EPI_ISL_2160169, EPI_ISL_2160175, EPI_ISL_2160182, EPI_ISL_2160184, EPI_ISL_2160186, EPI_ISL_2160194, EPI_ISL_2160198, EPI_ISL_2160199, EPI_ISL_2160205, EPI_ISL_2160206, EPI_ISL_2160212, EPI_ISL_2160214, EPI_ISL_2160221, EPI_ISL_2160222, EPI_ISL_2160223, EPI_ISL_2160247, EPI_ISL_2160255, EPI_ISL_2160269, EPI_ISL_2160271, EPI_ISL_2160276, EPI_ISL_2160278, EPI_ISL_2160287, EPI_ISL_2160312, EPI_ISL_2160432, EPI_ISL_2160471, EPI_ISL_2160488, EPI_ISL_2160494, EPI_ISL_2160498, EPI_ISL_2160544, EPI_ISL_2160549, EPI_ISL_2160610, EPI_ISL_2247355, EPI_ISL_2247364, EPI_ISL_2247366, EPI_ISL_2247369, EPI_ISL_2247375, EPI_ISL_2247390, EPI_ISL_2247487, EPI_ISL_2247497, EPI_ISL_2247501, EPI_ISL_2247588, EPI_ISL_2247589, EPI_ISL_2247595, EPI_ISL_2247612, EPI_ISL_2247614, EPI_ISL_2247630, EPI_ISL_2247632, EPI_ISL_2247636, EPI_ISL_2247646, EPI_ISL_2247650, EPI_ISL_2247682, EPI_ISL_2247714, EPI_ISL_2247819, EPI_ISL_2247820, EPI_ISL_2247821, EPI_ISL_2247822, EPI_ISL_2247832, EPI_ISL_2247843, EPI_ISL_2247848, EPI_ISL_2247861, EPI_ISL_2247864, EPI_ISL_2247873, EPI_ISL_2247874, EPI_ISL_2247885, EPI_ISL_2248134, EPI_ISL_2248151, EPI_ISL_2248155, EPI_ISL_2248298, EPI_ISL_2248369, EPI_ISL_2269616, EPI_ISL_2269621, EPI_ISL_2269627, EPI_ISL_2269688, EPI_ISL_2269699, EPI_ISL_2269709, EPI_ISL_2269758, EPI_ISL_2269770, EPI_ISL_2269797, EPI_ISL_2269801, EPI_ISL_2269802, EPI_ISL_2269827, EPI_ISL_2269831, EPI_ISL_2269839, EPI_ISL_2269842, EPI_ISL_2269870, EPI_ISL_2269897, EPI_ISL_2269908, EPI_ISL_2269945, EPI_ISL_2269963, EPI_ISL_2269976, EPI_ISL_2269981, EPI_ISL_2269986, EPI_ISL_2270000, EPI_ISL_2270024, EPI_ISL_2270119, EPI_ISL_2270188, EPI_ISL_2270198, EPI_ISL_2270199, EPI_ISL_2270212, EPI_ISL_2270231, EPI_ISL_2270382, EPI_ISL_2270411, EPI_ISL_2270456, EPI_ISL_2270466, EPI_ISL_2270472, EPI_ISL_2270477, EPI_ISL_2270494, EPI_ISL_2270495, EPI_ISL_2270501, EPI_ISL_2270505, EPI_ISL_2270508, EPI_ISL_2270514, EPI_ISL_2270519, EPI_ISL_2270529, EPI_ISL_2270531, EPI_ISL_2270559, EPI_ISL_2270562, EPI_ISL_2270564, EPI_ISL_2270584, EPI_ISL_2270612, EPI_ISL_2270665, EPI_ISL_2270714, EPI_ISL_2270734, EPI_ISL_2270737, EPI_ISL_2280212, EPI_ISL_2280219, EPI_ISL_2320037, EPI_ISL_2320047, EPI_ISL_2320067, EPI_ISL_2320068, EPI_ISL_2320079, EPI_ISL_2320117, EPI_ISL_2320137, EPI_ISL_2320140, EPI_ISL_2320194, EPI_ISL_2320235, EPI_ISL_2320248, EPI_ISL_2320327, EPI_ISL_2320388, EPI_ISL_2320403, EPI_ISL_2320409, EPI_ISL_2320417, EPI_ISL_2320418, EPI_ISL_2320419, EPI_ISL_2320427, EPI_ISL_2320434, EPI_ISL_2320440, EPI_ISL_2320441, EPI_ISL_2320482, EPI_ISL_2320499, EPI_ISL_2320502, EPI_ISL_2320574, EPI_ISL_2320595, EPI_ISL_2320596, EPI_ISL_2320609, EPI_ISL_2320620, EPI_ISL_2320623, EPI_ISL_2320632, EPI_ISL_2320638, EPI_ISL_2320645, EPI_ISL_2320668, EPI_ISL_2320849, EPI_ISL_2320857, EPI_ISL_2320859, EPI_ISL_2320869, EPI_ISL_2320879, EPI_ISL_2320883, EPI_ISL_2320885, EPI_ISL_2320898, EPI_ISL_2320899, EPI_ISL_2320905, EPI_ISL_2320913, EPI_ISL_2321050, EPI_ISL_2321061, EPI_ISL_2321065, EPI_ISL_2321113, EPI_ISL_2367640, EPI_ISL_2440063, EPI_ISL_2440065, EPI_ISL_2440072, EPI_ISL_2440086, EPI_ISL_2440144, EPI_ISL_2440145, EPI_ISL_2440167, EPI_ISL_2440193, EPI_ISL_2440198, EPI_ISL_2440287, EPI_ISL_2440338, EPI_ISL_2440466, EPI_ISL_2440475, EPI_ISL_2440478, EPI_ISL_2440519, EPI_ISL_2440523, EPI_ISL_2451495, EPI_ISL_2598860, EPI_ISL_2598864, EPI_ISL_2598871, EPI_ISL_2598878, EPI_ISL_2598886, EPI_ISL_2598904, EPI_ISL_2598912, EPI_ISL_2598950, EPI_ISL_2598955, EPI_ISL_2598967, EPI_ISL_2598968, EPI_ISL_2598988, EPI_ISL_2598992, EPI_ISL_2599003, EPI_ISL_2599022, EPI_ISL_2599042, EPI_ISL_2599231, EPI_ISL_2599256, EPI_ISL_2599364, EPI_ISL_2599383, EPI_ISL_2599399, EPI_ISL_2599410, EPI_ISL_2599449, EPI_ISL_2599456, EPI_ISL_2599459, EPI_ISL_2599477, EPI_ISL_2599478, EPI_ISL_2599479, EPI_ISL_2599485, EPI_ISL_2599488, EPI_ISL_2599496, EPI_ISL_2599499, EPI_ISL_2599510, EPI_ISL_2599512, EPI_ISL_2599513, EPI_ISL_2599522, EPI_ISL_2599531, EPI_ISL_2599538, EPI_ISL_2599552, EPI_ISL_2599560, EPI_ISL_2599563, EPI_ISL_2599570, EPI_ISL_2599591, EPI_ISL_2599595, EPI_ISL_2599611, EPI_ISL_2599622, EPI_ISL_2599625, EPI_ISL_2599629 | | | |
| see above | Helix/Illumina | Centers for Disease Control and Prevention Division of Viral Diseases, Pathogen Discovery | Adrian Paskey; Alexandre Bolze; Ary Ascencio; Benjamin Rambo-Martin; Brad Sickler; Charlotte Rivera-Garcia; Christine Tran; Christopher Gulvick; Clinton R. Paden; Dakota Howard; Darlene Wagner; David Becker; Dhwani Batra; Duncan MacCannell; Efren Sandoval; Eileen de Feo; Elizabeth Cirulli; Eric Allen; Geraint Levan; James Lu; Jan Antico; Jason Caravas; Jason Nguyen; Jimmy Ramirez; Jingtao Liu; Kara Moser; Kelly Schiabor Barrett; Kim Gietzen; Magnus Isaksson; Marc Laurent; Matthew Schmerer; Matthew Tolentino; Nicole L. Washington; Peter W. Cook; Phil Febbo; Ryan Cho; Scott Sammons; Shannon Wickline; Shatavia Morrison; Sherry Wang; Simon White; Tyler Cassens; William Lee; Yvette Unoarumhi |
| EPI_ISL_2335095, EPI_ISL_2335096, EPI_ISL_2335097, EPI_ISL_2335098, EPI_ISL_2335099, EPI_ISL_2427269, EPI_ISL_2427270, EPI_ISL_2427284, EPI_ISL_2427304, EPI_ISL_2427309, EPI_ISL_2500375, EPI_ISL_2500376, EPI_ISL_2500377, EPI_ISL_2500378, EPI_ISL_2500379, EPI_ISL_2500380, EPI_ISL_2500381, EPI_ISL_2500382, EPI_ISL_2534329, EPI_ISL_2598269, EPI_ISL_2598270, EPI_ISL_2598271, EPI_ISL_2598272, EPI_ISL_2789945, EPI_ISL_2789946, EPI_ISL_2789948, EPI_ISL_2789949, EPI_ISL_2789950, EPI_ISL_2801444, EPI_ISL_2801455, EPI_ISL_2801456, EPI_ISL_2801458, EPI_ISL_2801462, EPI_ISL_3218885, EPI_ISL_3218886, EPI_ISL_3281551 | | | |
| see above | IVY3 Central Lab, Vanderbilt University Medical Center | Lauring Lab, University of Michigan, Department of Microbiology and Immunology | Gilbert; Valesano |
| EPI_ISL_2106171, EPI_ISL_2991508, EPI_ISL_2991560, EPI_ISL_2991654 | Infectious Disease Program, Broad Institute of Harvard and MIT | Infectious Disease Program, Broad Institute of Harvard and MIT | A.E.; Adams, G.; Anahtar, M.; B.L.; B.W.; Bauer, M.; Birren; Branda, J.; Carter, A.; Cerrato, F.; Chaluvadi, S.; Chapman; Cusick, C.; D.J.; DeRuff, K.; Flowers, K.; Gallagher, G.; Gladden-Young, A.; Gnirke, A.; Harris, J.; J.E.; K.J.; LaRocque, R.; Lagerborg, K.; Lemieux; Lin; Loreth, C.; MacInnis; Neumann, A.; Normandin, E.; P.C.; Park; Pierce, V.; Reilly, S.; Rosenberg, E.; Rudy, M.; Ryan, E.; S.B.; Sabeti; Shaw, B.; Siddle; Slater, D.; Smole, S.; Tomkins-Tinch, C.; Turbett, S. |
| EPI_ISL_2088860, EPI_ISL_2089148, EPI_ISL_2089259, EPI_ISL_2089403, EPI_ISL_2089433, EPI_ISL_2186329, EPI_ISL_2186940, EPI_ISL_2282496, EPI_ISL_2283376, EPI_ISL_2283575, EPI_ISL_2283593, EPI_ISL_2323997, EPI_ISL_3299728 | | | |
| see above | Infinity Biologix | Centers for Disease Control and Prevention Division of Viral Diseases, Pathogen Discovery | Adrian Paskey; Benjamin Rambo-Martin; Chirayu Goswami; Christian Bixby; Christopher Gulvick; Clinton R. Paden; Dakota Howard; Darlene Wagner; Dhwani Batra; Duncan MacCannell; Jason Caravas; Jonathan Schultz; Kara Moser; Matthew Schmerer; Peter W. Cook; Robin Grimwood; Russ Hager; Scott Sammons; Shatavia Morrison; Yihe Wang; Yvette Unoarumhi |
| EPI_ISL_2044030, EPI_ISL_2045449, EPI_ISL_2045450, EPI_ISL_2045458, EPI_ISL_2045468, EPI_ISL_2045501, EPI_ISL_2045504, EPI_ISL_2045505, EPI_ISL_2045506, EPI_ISL_2045513, EPI_ISL_2045516, EPI_ISL_2045570, EPI_ISL_2045581, EPI_ISL_2045584, EPI_ISL_2045735, EPI_ISL_2045736, EPI_ISL_2045737, EPI_ISL_2045739, EPI_ISL_2045742, EPI_ISL_2045745, EPI_ISL_2045746, EPI_ISL_2045752, EPI_ISL_2045755, EPI_ISL_2045757, EPI_ISL_2045758, EPI_ISL_2045759, EPI_ISL_2045766, EPI_ISL_2045771, EPI_ISL_2045774, EPI_ISL_2045776, EPI_ISL_2045784, EPI_ISL_2045791, EPI_ISL_2045792, EPI_ISL_2045808, EPI_ISL_2045809, EPI_ISL_2045826, EPI_ISL_2045827, EPI_ISL_2045829, EPI_ISL_2045838, EPI_ISL_2045839, EPI_ISL_2045861, EPI_ISL_2045865, EPI_ISL_2045867, EPI_ISL_2045874, EPI_ISL_2045875, EPI_ISL_2045877, EPI_ISL_2045878, EPI_ISL_2045879, EPI_ISL_2045880, EPI_ISL_2045881, EPI_ISL_2045922, EPI_ISL_2045925, EPI_ISL_2045927, EPI_ISL_2181660, EPI_ISL_2181661, EPI_ISL_2181664, EPI_ISL_2181683, EPI_ISL_2181684, EPI_ISL_2181700, EPI_ISL_2181701, EPI_ISL_2181704, EPI_ISL_2181705, EPI_ISL_2181745, EPI_ISL_2181816, EPI_ISL_2181941, EPI_ISL_2182248, EPI_ISL_2182254, EPI_ISL_2182256, EPI_ISL_2182259, EPI_ISL_2182270, EPI_ISL_2182299, EPI_ISL_2182305, EPI_ISL_2182432, EPI_ISL_2182449, EPI_ISL_2182452, EPI_ISL_2182465, EPI_ISL_2182468, EPI_ISL_2182471, EPI_ISL_2182474, EPI_ISL_2182477, EPI_ISL_2182485, EPI_ISL_2182487, EPI_ISL_2182532, EPI_ISL_2182538, EPI_ISL_2182634, EPI_ISL_2182636, EPI_ISL_2182639, EPI_ISL_2182655, EPI_ISL_2182658, EPI_ISL_2182660, EPI_ISL_2182717, EPI_ISL_2182803, EPI_ISL_2182810, EPI_ISL_2182815, EPI_ISL_2182817, EPI_ISL_2182825, EPI_ISL_2182834, EPI_ISL_2182837, EPI_ISL_2182840, EPI_ISL_2182845, EPI_ISL_2182853, EPI_ISL_2182861, EPI_ISL_2183036, EPI_ISL_2183047, EPI_ISL_2183066, EPI_ISL_2183068, EPI_ISL_2183083, EPI_ISL_2183430, EPI_ISL_2184304, EPI_ISL_2305999, EPI_ISL_2306656, EPI_ISL_2306657, EPI_ISL_2306664, EPI_ISL_2306665, EPI_ISL_2306667, EPI_ISL_2306668, EPI_ISL_2306670, EPI_ISL_2306671, EPI_ISL_2306672, EPI_ISL_2306675, EPI_ISL_2306684, EPI_ISL_2306725, EPI_ISL_2306759, EPI_ISL_2306763, EPI_ISL_2306768, EPI_ISL_2306769, EPI_ISL_2306770, EPI_ISL_2306773, EPI_ISL_2306833, EPI_ISL_2306835, EPI_ISL_2306837, EPI_ISL_2306863, EPI_ISL_2306866, EPI_ISL_2306867, EPI_ISL_2306908, EPI_ISL_2306919, EPI_ISL_2306921, EPI_ISL_2306954, EPI_ISL_2306968, EPI_ISL_2306975, EPI_ISL_2306976, EPI_ISL_2306977, EPI_ISL_2306978, EPI_ISL_2306979, EPI_ISL_2306980, EPI_ISL_2306981, EPI_ISL_2306983, EPI_ISL_2306996, EPI_ISL_2398987, EPI_ISL_2398990, EPI_ISL_2398999, EPI_ISL_2399000, EPI_ISL_2399015, EPI_ISL_2399049, EPI_ISL_2399057, EPI_ISL_2399058, EPI_ISL_2399060, EPI_ISL_2399063, EPI_ISL_2399064, EPI_ISL_2399086, EPI_ISL_2399111, EPI_ISL_2399112, EPI_ISL_2399113, EPI_ISL_2399160, EPI_ISL_2399161, EPI_ISL_2399162, EPI_ISL_2399252, EPI_ISL_2399256, EPI_ISL_2399272, EPI_ISL_2399281, EPI_ISL_2399294, EPI_ISL_2399347, EPI_ISL_2399363, EPI_ISL_2399364, EPI_ISL_2399365, EPI_ISL_2399366, EPI_ISL_2399367, EPI_ISL_2399369, EPI_ISL_2399370, EPI_ISL_2399378, EPI_ISL_2399406, EPI_ISL_2480221, EPI_ISL_2480470, EPI_ISL_2480476, EPI_ISL_2480490, EPI_ISL_2480494, EPI_ISL_2480502, EPI_ISL_2480504, EPI_ISL_2480505, EPI_ISL_2480517, EPI_ISL_2480519, EPI_ISL_2480522, EPI_ISL_2480540, EPI_ISL_2480541, EPI_ISL_2481168, EPI_ISL_2611125, EPI_ISL_2611126, EPI_ISL_2611152, EPI_ISL_2611307, EPI_ISL_2611453, EPI_ISL_2872147, EPI_ISL_2872150, EPI_ISL_2872170, EPI_ISL_2872173, EPI_ISL_2872370, EPI_ISL_2926228, EPI_ISL_2926487, EPI_ISL_3016813, EPI_ISL_3111131, EPI_ISL_3111847, EPI_ISL_3111849, EPI_ISL_3111860, EPI_ISL_3111866, EPI_ISL_3111890, EPI_ISL_3111942, EPI_ISL_3112331, EPI_ISL_3112698, EPI_ISL_3112876, EPI_ISL_3112942, EPI_ISL_3327866, EPI_ISL_3328387, EPI_ISL_3328857, EPI_ISL_3330502, EPI_ISL_3330504, EPI_ISL_3330505, EPI_ISL_3330516, EPI_ISL_3330517, EPI_ISL_3330523, EPI_ISL_3330527, EPI_ISL_3330528, EPI_ISL_3330539, EPI_ISL_3330542, EPI_ISL_3330550, EPI_ISL_3330553, EPI_ISL_3330557, EPI_ISL_3330560, EPI_ISL_3330581, EPI_ISL_3330604 | | | |
| see above | Laboratory Corporation of America | Centers for Disease Control and Prevention Division of Viral Diseases, Pathogen Discovery | Adrian Paskey; Amanda Douglas; Amanda Suchanek; Andrea Throop; Ayla Burns; Benjamin Rambo-Martin; Bobbi Croy; Brian Krueger; Brian Norvell; Christopher Gulvick; Christos Petropoulos; Clinton R. Paden; Craig Lukasik; Dakota Howard; Darlene Wagner; Debbie Boles; Dhwani Batra; Duncan MacCannell; Eyad Almasri; Goran Stevovic; Howard Engler; Hrushikesh Deshmukh; Jake Humphrey; Jana Schroth; Jason Caravas; Joe Voshell; John Pruitt; Jonathan Meltzer; Jonathan Williams; Kara Moser; Kimberly Wagner; Lax Iyer; Lisa Pfefferle; Lyndon Tilson; Manoj Jain; Marcia Eisenberg; Mary Ann Cristobal; Mary Williamson; Matthew Robinson; Matthew Schmerer; Michael Levandoski; Mike Sapeta; Mindy Nye; Minoo Agarwal; Mohan Kolli; Nuthawin Charoensri; Oren Cohen; Peter W. Cook; Prashant Gupta; Qian Zeng; Rama Ghatti; Scott Parker; Scott Ryan; Scott Sammons; Shatavia Morrison; Stanley Letovsky; Steven Ragan; Suresh Babu Selvaraju; Susan Countryman; Susan Hicks; Suzanne Dale; Thomas Urban; Tim Kuphal; Tricia Zwiefelhofer; Vincent Drouillon; Yvette Unoarumhi |
| EPI_ISL_2018220, EPI_ISL_2018221, EPI_ISL_2018222, EPI_ISL_2018223, EPI_ISL_2018226, EPI_ISL_2018227, EPI_ISL_2451813, EPI_ISL_2451815, EPI_ISL_2451816, EPI_ISL_2451818, EPI_ISL_2451820, EPI_ISL_2451821, EPI_ISL_2451827, EPI_ISL_2451829, EPI_ISL_3354254 | | | |
| see above | MA State Public Health Laboratory | Centers for Disease Control and Prevention Division of Viral Diseases, Pathogen Discovery | Alex Burgin; Alison Laufer Halpin; Ben L. Rambo-Martin; Ben Rambo-Martin; Clinton Paden; Clinton R. Paden; Dakota Howard; Darlene Wagner; Dave Wentworth; Dhwani Batra; Jasmine Padilla; Justin Lee; Katie Dillon; Krista Queen; Kristen Knipe; Kristine Lacek; Mark Burroughs; Matthew Schmerer; Meghan Bentz; Mili Sheth; Peter Cook; Sam Shepard; Sarah Nobles; Shoshona Le; Suxiang Tong; Vivien Dugan; Yvette Unoarumhi |
| EPI_ISL_2016440, EPI_ISL_2016441, EPI_ISL_2016442, EPI_ISL_2016443, EPI_ISL_2016444, EPI_ISL_2016445, EPI_ISL_2016446, EPI_ISL_2016447, EPI_ISL_2016448, EPI_ISL_2016449, EPI_ISL_2016450, EPI_ISL_2016451, EPI_ISL_2016452, EPI_ISL_2016453, EPI_ISL_2016454, EPI_ISL_2016455, EPI_ISL_2016458, EPI_ISL_2016459, EPI_ISL_2016460, EPI_ISL_2016461, EPI_ISL_2016462, EPI_ISL_2016464, EPI_ISL_2016465, EPI_ISL_2016466, EPI_ISL_2016467, EPI_ISL_2016468, EPI_ISL_2016469, EPI_ISL_2016470, EPI_ISL_2016471, EPI_ISL_2016472, EPI_ISL_2016473, EPI_ISL_2016474, EPI_ISL_2016475, EPI_ISL_2016476, EPI_ISL_2016477, EPI_ISL_2016478, EPI_ISL_2016479, EPI_ISL_2016480, EPI_ISL_2016481, EPI_ISL_2016482, EPI_ISL_2016483, EPI_ISL_2016484, EPI_ISL_2016485, EPI_ISL_2016486, EPI_ISL_2362355, EPI_ISL_2362356, EPI_ISL_2362357, EPI_ISL_2362358, EPI_ISL_2362359, EPI_ISL_2362360, EPI_ISL_2362361, EPI_ISL_2362362, EPI_ISL_2362364, EPI_ISL_2362365, EPI_ISL_2362366, EPI_ISL_2362367, EPI_ISL_2362368, EPI_ISL_2362369, EPI_ISL_2362370, EPI_ISL_2362371, EPI_ISL_2362372, EPI_ISL_2362373, EPI_ISL_2362375 | | | |
| see above | Massachusetts General Hospital | Infectious Disease Program, Broad Institute of Harvard and MIT | A.E.; Adams, G.; Anahtar, M.; B.L.; B.W.; Bauer, M.; Birren; Branda, J.; Carter, A.; Cerrato, F.; Chaluvadi, S.; Chapman; Cusick, C.; D.J.; DeRuff, K.; E. and Sabeti; Flowers, K.; Gallagher, G.; Gladden-Young, A.; Gnirke, A.; Harris, J.; J.E.; K.J.; LaRocque, R.; Lagerborg, K.; Lemieux; Lin; Loreth, C.; MacInnis; Neumann, A.; Normandin, E.; P.C.; Park; Pierce, V.; Reilly, S.; Rosenberg; Rudy, M.; Ryan, E.; S.B.; Shaw, B.; Siddle; Slater, D.; Smole, S.; Tomkins-Tinch, C.; Turbett, S. |
| EPI_ISL_2008117, EPI_ISL_2008118, EPI_ISL_2008119, EPI_ISL_2008121, EPI_ISL_2008122, EPI_ISL_2008124, EPI_ISL_2008125, EPI_ISL_2008127, EPI_ISL_2008128, EPI_ISL_2008129, EPI_ISL_2008130, EPI_ISL_2008131, EPI_ISL_2008132, EPI_ISL_2008133, EPI_ISL_2008134, EPI_ISL_2008135, EPI_ISL_2008136, EPI_ISL_2008137, EPI_ISL_2008138, EPI_ISL_2008139, EPI_ISL_2008140, EPI_ISL_2008141, EPI_ISL_2008142, EPI_ISL_2008144, EPI_ISL_2008146, EPI_ISL_2008147, EPI_ISL_2008148, EPI_ISL_2008149, EPI_ISL_2008150, EPI_ISL_2008151, EPI_ISL_2008152, EPI_ISL_2008153, EPI_ISL_2008155, EPI_ISL_2008157, EPI_ISL_2008158, EPI_ISL_2008159, EPI_ISL_2008160, EPI_ISL_2008161, EPI_ISL_2008162, EPI_ISL_2008163, EPI_ISL_2008164, EPI_ISL_2008166, EPI_ISL_2008167, EPI_ISL_2008168, EPI_ISL_2008170, EPI_ISL_2008172, EPI_ISL_2008173, EPI_ISL_2008174, EPI_ISL_2008175, EPI_ISL_2008177, EPI_ISL_2008178, EPI_ISL_2008182, EPI_ISL_2008183, EPI_ISL_2008184, EPI_ISL_2008185, EPI_ISL_2008186, EPI_ISL_2008187, EPI_ISL_2008188, EPI_ISL_2008189, EPI_ISL_2008190, EPI_ISL_2008191, EPI_ISL_2008193, EPI_ISL_2008194, EPI_ISL_2008195, EPI_ISL_2008196, EPI_ISL_2008197, | | | |

| | | | |
|---|---|---|---|
| EPI_ISL_2008198, EPI_ISL_2008199, EPI_ISL_2008202, EPI_ISL_2008203, EPI_ISL_2008204, EPI_ISL_2008205, EPI_ISL_2008206, EPI_ISL_2008207, EPI_ISL_2008208, EPI_ISL_2008209, EPI_ISL_2008210, EPI_ISL_2008211, EPI_ISL_2008212, EPI_ISL_2008213, EPI_ISL_2008214, EPI_ISL_2008215, EPI_ISL_2008216, EPI_ISL_2008217, EPI_ISL_2008218, EPI_ISL_2008219, EPI_ISL_2008220, EPI_ISL_2008221, EPI_ISL_2008222, EPI_ISL_2008223, EPI_ISL_2008224, EPI_ISL_2008225, EPI_ISL_2008226, EPI_ISL_2008227, EPI_ISL_2008228, EPI_ISL_2008229, EPI_ISL_2008230, EPI_ISL_2008231, EPI_ISL_2008232, EPI_ISL_2008233, EPI_ISL_2008234, EPI_ISL_2008235, EPI_ISL_2008236, EPI_ISL_2008237, EPI_ISL_2008238, EPI_ISL_2008239, EPI_ISL_2008240, EPI_ISL_2008241, EPI_ISL_2008242, EPI_ISL_2008243, EPI_ISL_2008244, EPI_ISL_2008245, EPI_ISL_2008246, EPI_ISL_2008247, EPI_ISL_2008248, EPI_ISL_2008249, EPI_ISL_2008250, EPI_ISL_2008251, EPI_ISL_2008252, EPI_ISL_2008253, EPI_ISL_2008254, EPI_ISL_2008256, EPI_ISL_2008257, EPI_ISL_2008258, EPI_ISL_2008259, EPI_ISL_2008260, EPI_ISL_2008261, EPI_ISL_2008262, EPI_ISL_2008263, EPI_ISL_2008264, EPI_ISL_2008265, EPI_ISL_2008266, EPI_ISL_2008267, EPI_ISL_2008268, EPI_ISL_2008269, EPI_ISL_2008270, EPI_ISL_2008271, EPI_ISL_2008272, EPI_ISL_2008273, EPI_ISL_2008274, EPI_ISL_2008275, EPI_ISL_2008276, EPI_ISL_2008277, EPI_ISL_2008278, EPI_ISL_2008279, EPI_ISL_2008280, EPI_ISL_2008281, EPI_ISL_2008282, EPI_ISL_2008283, EPI_ISL_2008284, EPI_ISL_2008285, EPI_ISL_2008286, EPI_ISL_2008287, EPI_ISL_2008293, EPI_ISL_2008294, EPI_ISL_2008295, EPI_ISL_2008296, EPI_ISL_2008297, EPI_ISL_2008298, EPI_ISL_2008299, EPI_ISL_2008300, EPI_ISL_2008301, EPI_ISL_2008302, EPI_ISL_2008303, EPI_ISL_2008304, EPI_ISL_2008305, EPI_ISL_2008306, EPI_ISL_2008307, EPI_ISL_2008308, EPI_ISL_2008309, EPI_ISL_2008310, EPI_ISL_2008312, EPI_ISL_2008314, EPI_ISL_2008315, EPI_ISL_2008316, EPI_ISL_2008317, EPI_ISL_2008318, EPI_ISL_2008319, EPI_ISL_2008321, EPI_ISL_2008322, EPI_ISL_2008323, EPI_ISL_2008325, EPI_ISL_2008326, EPI_ISL_2008327, EPI_ISL_2008329, EPI_ISL_2008330, EPI_ISL_2008331, EPI_ISL_2008332, EPI_ISL_2008333, EPI_ISL_2008335, EPI_ISL_2008336, EPI_ISL_2008337, EPI_ISL_2008338, EPI_ISL_2008339, EPI_ISL_2008340, EPI_ISL_2008341, EPI_ISL_2008342, EPI_ISL_2008343, EPI_ISL_2008344, EPI_ISL_2008345, EPI_ISL_2008346, EPI_ISL_2008347, EPI_ISL_2008348, EPI_ISL_2008349, EPI_ISL_2008350, EPI_ISL_2008367, EPI_ISL_2008368, EPI_ISL_2008369, EPI_ISL_2008378, EPI_ISL_2008379, EPI_ISL_2008380, EPI_ISL_2008381, EPI_ISL_2008382, EPI_ISL_2008383, EPI_ISL_2008384, EPI_ISL_2008385, EPI_ISL_2008386, EPI_ISL_2008388, EPI_ISL_2008389, EPI_ISL_2008390, EPI_ISL_2008391, EPI_ISL_2008392, EPI_ISL_2008393, EPI_ISL_2008394, EPI_ISL_2008395, EPI_ISL_2008396, EPI_ISL_2008399, EPI_ISL_2229481, EPI_ISL_2229482, EPI_ISL_2229483, EPI_ISL_2229484, EPI_ISL_2229485, EPI_ISL_2229486, EPI_ISL_2229487, EPI_ISL_2229488, EPI_ISL_2229489, EPI_ISL_2229490, EPI_ISL_2229492, EPI_ISL_2229493, EPI_ISL_2229494, EPI_ISL_2229495, EPI_ISL_2229496, EPI_ISL_2229497, EPI_ISL_2229498, EPI_ISL_2229499, EPI_ISL_2229500, EPI_ISL_2229505, EPI_ISL_2229506, EPI_ISL_2229507, EPI_ISL_2229508, EPI_ISL_2229509, EPI_ISL_2229510, EPI_ISL_2229511, EPI_ISL_2229512, EPI_ISL_2229513, EPI_ISL_2229514, EPI_ISL_2229515, EPI_ISL_2229516, EPI_ISL_2229517, EPI_ISL_2229519, EPI_ISL_2229521, EPI_ISL_2229523, EPI_ISL_2229524, EPI_ISL_2229525, EPI_ISL_2229526, EPI_ISL_2229527, EPI_ISL_2229528, EPI_ISL_2229529, EPI_ISL_2229530, EPI_ISL_2229531, EPI_ISL_2229532, EPI_ISL_2229534, EPI_ISL_2229535, EPI_ISL_2229536, EPI_ISL_2229537, EPI_ISL_2229765, EPI_ISL_2229766, EPI_ISL_2229767, EPI_ISL_2229768, EPI_ISL_2229769, EPI_ISL_2229772, EPI_ISL_2229773, EPI_ISL_2229774, EPI_ISL_2229775, EPI_ISL_2229776, EPI_ISL_2229777, EPI_ISL_2229778, EPI_ISL_2229779, EPI_ISL_2229780, EPI_ISL_2229781, EPI_ISL_2229782, EPI_ISL_2229783, EPI_ISL_2229792, EPI_ISL_2229793, EPI_ISL_2229796, EPI_ISL_2229797, EPI_ISL_2229798, EPI_ISL_2229799, EPI_ISL_2229800, EPI_ISL_2229803, EPI_ISL_2229804, EPI_ISL_2229805, EPI_ISL_2229806, EPI_ISL_2229807, EPI_ISL_2229808, EPI_ISL_2229809, EPI_ISL_2229810, EPI_ISL_2229811, EPI_ISL_2229812, EPI_ISL_2229813, EPI_ISL_2400245, EPI_ISL_2400246, EPI_ISL_2400247, EPI_ISL_2400248, EPI_ISL_2400249, EPI_ISL_2400250, EPI_ISL_2400251, EPI_ISL_2400252, EPI_ISL_2483773, EPI_ISL_2483777, EPI_ISL_2483778, EPI_ISL_2483779, EPI_ISL_2483780, EPI_ISL_2483781, EPI_ISL_2483783, EPI_ISL_2483784, EPI_ISL_2483785, EPI_ISL_2483786, EPI_ISL_2483787, EPI_ISL_2483788, EPI_ISL_2483789, EPI_ISL_2483790, EPI_ISL_2483791, EPI_ISL_2483792, EPI_ISL_2483793, EPI_ISL_2483794, EPI_ISL_2483795, EPI_ISL_2483797, EPI_ISL_2483798, EPI_ISL_2483799, EPI_ISL_2483800, EPI_ISL_2483801, EPI_ISL_2483802, EPI_ISL_2483803, EPI_ISL_2483804, EPI_ISL_2483805, EPI_ISL_2483806, EPI_ISL_2483807, EPI_ISL_2483808, EPI_ISL_2483809, EPI_ISL_2483810, EPI_ISL_2484933, EPI_ISL_2484934, EPI_ISL_2484935, EPI_ISL_2484936, EPI_ISL_2484937, EPI_ISL_2484938, EPI_ISL_2484939, EPI_ISL_2484940, EPI_ISL_2484941, EPI_ISL_2484942, EPI_ISL_2484943, EPI_ISL_2484944, EPI_ISL_2484945, EPI_ISL_2484946, EPI_ISL_2484947, EPI_ISL_2484948, EPI_ISL_2484949, EPI_ISL_2484950, EPI_ISL_2484951, EPI_ISL_2484952, EPI_ISL_2484953, EPI_ISL_2484954, EPI_ISL_2484955, EPI_ISL_2484956, EPI_ISL_2484957, EPI_ISL_2484958, EPI_ISL_2484959, EPI_ISL_2688804, EPI_ISL_2688806, EPI_ISL_2688807, EPI_ISL_2688808, EPI_ISL_2688809, EPI_ISL_2688810, EPI_ISL_2688811, EPI_ISL_2688812, EPI_ISL_2688813, EPI_ISL_2688814, EPI_ISL_2688815, EPI_ISL_2688816, EPI_ISL_2688817, EPI_ISL_2688818, EPI_ISL_2688819, EPI_ISL_2688820, EPI_ISL_2920250, EPI_ISL_2920251, EPI_ISL_2920252, EPI_ISL_2920253, EPI_ISL_2920254, EPI_ISL_2920255, EPI_ISL_2920256, EPI_ISL_2920257, EPI_ISL_2920258, EPI_ISL_2920259, EPI_ISL_2920260, EPI_ISL_2920261, EPI_ISL_2920262, EPI_ISL_2920263, EPI_ISL_2920264, EPI_ISL_2920265, EPI_ISL_2920266, EPI_ISL_2920267, EPI_ISL_2920268, EPI_ISL_2920269, EPI_ISL_2920271 | | | |
| see above | Massachusetts State Public Health Laboratory | Massachusetts State Public Health Laboratory | Andrew Lang; Glen Gallagher; Sandra Smole; Timelia Fink |
| EPI_ISL_3236361, EPI_ISL_3236585 | New England Biolabs | New England Biolabs | Bradley William Langhorst |
| EPI_ISL_1990410, EPI_ISL_1990412, EPI_ISL_1990413, EPI_ISL_1990416, EPI_ISL_1990429, EPI_ISL_1990430, EPI_ISL_1990432, EPI_ISL_1990433, EPI_ISL_1990436, EPI_ISL_1990437, EPI_ISL_1990439, EPI_ISL_1990441, EPI_ISL_1990445, EPI_ISL_1990446, EPI_ISL_1990448, EPI_ISL_1990449, EPI_ISL_1990450, EPI_ISL_1990451, EPI_ISL_1990452, EPI_ISL_1990454, EPI_ISL_1990457, EPI_ISL_1990458, EPI_ISL_1990459, EPI_ISL_1990463, EPI_ISL_1990465, EPI_ISL_1990467, EPI_ISL_1990468, EPI_ISL_1990470, EPI_ISL_1990475, EPI_ISL_1990477, EPI_ISL_1990481, EPI_ISL_1990490, EPI_ISL_1990491, EPI_ISL_1990492, EPI_ISL_1990493, EPI_ISL_1990494, EPI_ISL_1990498, EPI_ISL_1990499, EPI_ISL_1990501, EPI_ISL_1990505, EPI_ISL_1990506, EPI_ISL_1990508, EPI_ISL_1990510, EPI_ISL_1990514, EPI_ISL_1990517, EPI_ISL_1990519, EPI_ISL_1990521, EPI_ISL_1990526, EPI_ISL_1990527, EPI_ISL_1990532, EPI_ISL_1990533, EPI_ISL_1990539, EPI_ISL_1990542, EPI_ISL_1990543, EPI_ISL_2011067, EPI_ISL_2011099, EPI_ISL_2011104, EPI_ISL_2011114, EPI_ISL_2011121, EPI_ISL_2011128, EPI_ISL_2011130, EPI_ISL_2011131, EPI_ISL_2011132, EPI_ISL_2011135, EPI_ISL_2011157, EPI_ISL_2011165, EPI_ISL_2011167, EPI_ISL_2011179, EPI_ISL_2011186, EPI_ISL_2011187, EPI_ISL_2011189, EPI_ISL_2011199, EPI_ISL_2011205, EPI_ISL_2011207, EPI_ISL_2011233, EPI_ISL_2011235, EPI_ISL_2011262, EPI_ISL_2011274, EPI_ISL_2011292, EPI_ISL_2011312, EPI_ISL_2011458, EPI_ISL_2011476, EPI_ISL_2011650, EPI_ISL_2011654, EPI_ISL_2011661, EPI_ISL_2011680, EPI_ISL_2034789, EPI_ISL_2034793, EPI_ISL_2034796, EPI_ISL_2034799, EPI_ISL_2034804, EPI_ISL_2034808, EPI_ISL_2034811, EPI_ISL_2034817, EPI_ISL_2034822, EPI_ISL_2034825, EPI_ISL_2034833, EPI_ISL_2034836, EPI_ISL_2034840, EPI_ISL_2034844, EPI_ISL_2035328, EPI_ISL_2035332, EPI_ISL_2035333, EPI_ISL_2035361, EPI_ISL_2046978, EPI_ISL_2047008, EPI_ISL_2047009, EPI_ISL_2047083, EPI_ISL_2047112, EPI_ISL_2047118, EPI_ISL_2090476, EPI_ISL_2090486, EPI_ISL_2090487, EPI_ISL_2090491, EPI_ISL_2090493, EPI_ISL_2090494, EPI_ISL_2090496, EPI_ISL_2090499, EPI_ISL_2090502, EPI_ISL_2090513, EPI_ISL_2090525, EPI_ISL_2090526, EPI_ISL_2090533, EPI_ISL_2090535, EPI_ISL_2090536, EPI_ISL_2090546, EPI_ISL_2090557, EPI_ISL_2090558, EPI_ISL_2090569, EPI_ISL_2090570, EPI_ISL_2090581, EPI_ISL_2090583, EPI_ISL_2090586, EPI_ISL_2090602, EPI_ISL_2098280, EPI_ISL_2098284, EPI_ISL_2098290, EPI_ISL_2098298, EPI_ISL_2098302, EPI_ISL_2098303, EPI_ISL_2098305, EPI_ISL_2098308, EPI_ISL_2098310, EPI_ISL_2098326, EPI_ISL_2098332, EPI_ISL_2098339, EPI_ISL_2098365, EPI_ISL_2098392, EPI_ISL_2098411, EPI_ISL_2098412, EPI_ISL_2132574, EPI_ISL_2132579, EPI_ISL_2132586, EPI_ISL_2132622, EPI_ISL_2132629, EPI_ISL_2142888, EPI_ISL_2142900, EPI_ISL_2142905, EPI_ISL_2142910, EPI_ISL_2142918, EPI_ISL_2142923, EPI_ISL_2142924, EPI_ISL_2142925, EPI_ISL_2142927, EPI_ISL_2142930, EPI_ISL_2142938, EPI_ISL_2142939, EPI_ISL_2142961, EPI_ISL_2142965, EPI_ISL_2142972, EPI_ISL_2142979, EPI_ISL_2142990, EPI_ISL_2142992, EPI_ISL_2142997, EPI_ISL_2142999, EPI_ISL_2143014, EPI_ISL_2143015, EPI_ISL_2143017, EPI_ISL_2143020, EPI_ISL_2143024, EPI_ISL_2143031, EPI_ISL_2143037, EPI_ISL_2143063, EPI_ISL_2143071, EPI_ISL_2143083, EPI_ISL_2143089, EPI_ISL_2143096, EPI_ISL_2143106, EPI_ISL_2143110, EPI_ISL_2143126, EPI_ISL_2143130, EPI_ISL_2143131, EPI_ISL_2143144, EPI_ISL_2143168, EPI_ISL_2143171, EPI_ISL_2143180, EPI_ISL_2143185, EPI_ISL_2143196, EPI_ISL_2143213, EPI_ISL_2143225, EPI_ISL_2143230, EPI_ISL_2143237, EPI_ISL_2143247, EPI_ISL_2143277, EPI_ISL_2143280, EPI_ISL_2143297, EPI_ISL_2143302, EPI_ISL_2143303, EPI_ISL_2143304, EPI_ISL_2143305, EPI_ISL_2143307, EPI_ISL_2143308, EPI_ISL_2143309, EPI_ISL_2143310, EPI_ISL_2143313, EPI_ISL_2143317, EPI_ISL_2143324, EPI_ISL_2143328, EPI_ISL_2143329, EPI_ISL_2143332, EPI_ISL_2143337, EPI_ISL_2143346, EPI_ISL_2143358, EPI_ISL_2143363, EPI_ISL_2143372, EPI_ISL_2143383, EPI_ISL_2143420, EPI_ISL_2143473, EPI_ISL_2143549, EPI_ISL_2143562, EPI_ISL_2143667, EPI_ISL_2143741, EPI_ISL_2143745, EPI_ISL_2143786, EPI_ISL_2143866, EPI_ISL_2143880, EPI_ISL_2143887, EPI_ISL_2143976, EPI_ISL_2143983, EPI_ISL_2143995, EPI_ISL_2179926, EPI_ISL_2179935, EPI_ISL_2179944, EPI_ISL_2179951, EPI_ISL_2179966, EPI_ISL_2179982, EPI_ISL_2180462, EPI_ISL_2180463, EPI_ISL_2180469, EPI_ISL_2180471, EPI_ISL_2180472, EPI_ISL_2180478, EPI_ISL_2180484, EPI_ISL_2180502, EPI_ISL_2180506, EPI_ISL_2180525, EPI_ISL_2187260, EPI_ISL_2187265, EPI_ISL_2187277, EPI_ISL_2187291, EPI_ISL_2187293, EPI_ISL_2187298, EPI_ISL_2187309, EPI_ISL_2187310, EPI_ISL_2187312, EPI_ISL_2187313, EPI_ISL_2187315, EPI_ISL_2187317, EPI_ISL_2241631, EPI_ISL_2241632, EPI_ISL_2241636, EPI_ISL_2241640, EPI_ISL_2241642, EPI_ISL_2268401, EPI_ISL_2268402, EPI_ISL_2268407, EPI_ISL_2268410, EPI_ISL_2268420, EPI_ISL_2268423, EPI_ISL_2268429, EPI_ISL_2268433, EPI_ISL_2268454, EPI_ISL_2268474, EPI_ISL_2268479, EPI_ISL_2268501, EPI_ISL_2268507, EPI_ISL_2268516, EPI_ISL_2268522, EPI_ISL_2268529, EPI_ISL_2268535, EPI_ISL_2268591, EPI_ISL_2268692, EPI_ISL_2268707, EPI_ISL_2268710, EPI_ISL_2268722, EPI_ISL_2268752, EPI_ISL_2268754, EPI_ISL_2268766, EPI_ISL_2268775, EPI_ISL_2268793, EPI_ISL_2268794, EPI_ISL_2268797, EPI_ISL_2268806, EPI_ISL_2268814, EPI_ISL_2268816, EPI_ISL_2268840, EPI_ISL_2268843, EPI_ISL_2268844, EPI_ISL_2268847, EPI_ISL_2268850, EPI_ISL_2268857, EPI_ISL_2268866, EPI_ISL_2268873, EPI_ISL_2268877, EPI_ISL_2268885, EPI_ISL_2268888, EPI_ISL_2268901, EPI_ISL_2268902, EPI_ISL_2268903, EPI_ISL_2268912, EPI_ISL_2268921, EPI_ISL_2268927, EPI_ISL_2269074, EPI_ISL_2269082, EPI_ISL_2269083, EPI_ISL_2269084, EPI_ISL_2269085, EPI_ISL_2269099, EPI_ISL_2269111, EPI_ISL_2269116, EPI_ISL_2269128, EPI_ISL_2269135, EPI_ISL_2269147, EPI_ISL_2269150, EPI_ISL_2269161, EPI_ISL_2269167, EPI_ISL_2269180, EPI_ISL_2269182, EPI_ISL_2269183, EPI_ISL_2269193, EPI_ISL_2269196, EPI_ISL_2269208, EPI_ISL_2269211, EPI_ISL_2280080, EPI_ISL_2280089, EPI_ISL_2280092, EPI_ISL_2280102, EPI_ISL_2280105, EPI_ISL_2280110, EPI_ISL_2280120, EPI_ISL_2280121, EPI_ISL_2280127, EPI_ISL_2280129, EPI_ISL_2280134, EPI_ISL_2280164, EPI_ISL_2280165, EPI_ISL_2280166, EPI_ISL_2280168, EPI_ISL_2280169, EPI_ISL_2280174, EPI_ISL_2280175, EPI_ISL_2304645, EPI_ISL_2304646, EPI_ISL_2304654, EPI_ISL_2304705, EPI_ISL_2307159, EPI_ISL_2367803, EPI_ISL_2367835, EPI_ISL_2367955, EPI_ISL_2367970, EPI_ISL_2367972, EPI_ISL_2368004, EPI_ISL_2368018, EPI_ISL_2368036, EPI_ISL_2368061, EPI_ISL_2369130, EPI_ISL_2369152, EPI_ISL_2369167, EPI_ISL_2369215, EPI_ISL_2369279, EPI_ISL_2369290, EPI_ISL_2369317, EPI_ISL_2369320, EPI_ISL_2369375, EPI_ISL_2369378, EPI_ISL_2369388, EPI_ISL_2371723, EPI_ISL_2371734, EPI_ISL_2371740, EPI_ISL_2371746, EPI_ISL_2371749, EPI_ISL_2371753, EPI_ISL_2371755, EPI_ISL_2371758, EPI_ISL_2371772, EPI_ISL_2371775, EPI_ISL_2371776, EPI_ISL_2371779, EPI_ISL_2371785, EPI_ISL_2371794, EPI_ISL_2371801, EPI_ISL_2371804, EPI_ISL_2371814, EPI_ISL_2371817, EPI_ISL_2371821, EPI_ISL_2371831, EPI_ISL_2371839, EPI_ISL_2371847, EPI_ISL_2371863, EPI_ISL_2371882, EPI_ISL_2371890, EPI_ISL_2371918, EPI_ISL_2371922, EPI_ISL_2371924, EPI_ISL_2371933, EPI_ISL_2371939, EPI_ISL_2371947, EPI_ISL_2371968, EPI_ISL_2371977, EPI_ISL_2371979, EPI_ISL_2372154, EPI_ISL_2372155, EPI_ISL_2372156, EPI_ISL_2372166, EPI_ISL_2372169, EPI_ISL_2372171, EPI_ISL_2372173, EPI_ISL_2372176, EPI_ISL_2372177, EPI_ISL_2372192, EPI_ISL_2372198, EPI_ISL_2372199, EPI_ISL_2372212, EPI_ISL_2372217, EPI_ISL_2372222, EPI_ISL_2372225, EPI_ISL_2372230, EPI_ISL_2372239, EPI_ISL_2372245, EPI_ISL_2372254, EPI_ISL_2372259, EPI_ISL_2372261, EPI_ISL_2372264, EPI_ISL_2372410, EPI_ISL_2372421, EPI_ISL_2372423, EPI_ISL_2372900, EPI_ISL_2372902, EPI_ISL_2372903, EPI_ISL_2372906, EPI_ISL_2372914, EPI_ISL_2372917, EPI_ISL_2372918, EPI_ISL_2372921, EPI_ISL_2372923, EPI_ISL_2372924, EPI_ISL_2372928, EPI_ISL_2372930, EPI_ISL_2372931, EPI_ISL_2372941, EPI_ISL_2372943, EPI_ISL_2372945, EPI_ISL_2372946, EPI_ISL_2372949, EPI_ISL_2372950, EPI_ISL_2372952, EPI_ISL_2397333, EPI_ISL_2397341, EPI_ISL_2397374, EPI_ISL_2397391, EPI_ISL_2397397, EPI_ISL_2397446, EPI_ISL_2397451, EPI_ISL_2397452, EPI_ISL_2397458, EPI_ISL_2397472, EPI_ISL_2397476, EPI_ISL_2397500, EPI_ISL_2397501, EPI_ISL_2397517, EPI_ISL_2397562, EPI_ISL_2397564, EPI_ISL_2397565, EPI_ISL_2397570, EPI_ISL_2397573, EPI_ISL_2397575, EPI_ISL_2397580, EPI_ISL_2397584, EPI_ISL_2397586, EPI_ISL_2397588, EPI_ISL_2397590, EPI_ISL_2397591, EPI_ISL_2397592, EPI_ISL_2397593, EPI_ISL_2397594, EPI_ISL_2397596, EPI_ISL_2397602, EPI_ISL_2397609, EPI_ISL_2397612, EPI_ISL_2397613, EPI_ISL_2397614, EPI_ISL_2397621, EPI_ISL_2397627, EPI_ISL_2397631, EPI_ISL_2397634, EPI_ISL_2397636, EPI_ISL_2397639, EPI_ISL_2397646, EPI_ISL_2397647, EPI_ISL_2397650, EPI_ISL_2397658, EPI_ISL_2397659, EPI_ISL_2397660, EPI_ISL_2397665, EPI_ISL_2397671, EPI_ISL_2397672, EPI_ISL_2397673, EPI_ISL_2397676, EPI_ISL_2397679, EPI_ISL_2397684, EPI_ISL_2397687, EPI_ISL_2397694, EPI_ISL_2397697, EPI_ISL_2397709, EPI_ISL_2397717, EPI_ISL_2397718, EPI_ISL_2397726, EPI_ISL_2397732, EPI_ISL_2397736, EPI_ISL_2397744, EPI_ISL_2397747, EPI_ISL_2397760, EPI_ISL_2397769, EPI_ISL_2397773, EPI_ISL_2397782, EPI_ISL_2397783, EPI_ISL_2397790, EPI_ISL_2397795, EPI_ISL_2399463, EPI_ISL_2399464, EPI_ISL_2399473, EPI_ISL_2399489, EPI_ISL_2399490, EPI_ISL_2399491, EPI_ISL_2399493, EPI_ISL_2399496, EPI_ISL_2399497, EPI_ISL_2399498, EPI_ISL_2399499, EPI_ISL_2399500, EPI_ISL_2399502, EPI_ISL_2399503, EPI_ISL_2399506, EPI_ISL_2399509, EPI_ISL_2399510, EPI_ISL_2399512, EPI_ISL_2439583, EPI_ISL_2439609, EPI_ISL_2439611, EPI_ISL_2439615, EPI_ISL_2439621, EPI_ISL_2439622, EPI_ISL_2439626, EPI_ISL_2439627, EPI_ISL_2439630, EPI_ISL_2439638, EPI_ISL_2439647, EPI_ISL_2439648, EPI_ISL_2439660, EPI_ISL_2439661, EPI_ISL_2439664, EPI_ISL_2439665, EPI_ISL_2439669, EPI_ISL_2439675, EPI_ISL_2439678, EPI_ISL_2439684, EPI_ISL_2439874, EPI_ISL_2439895, EPI_ISL_2439936, EPI_ISL_2439937, EPI_ISL_2439946, EPI_ISL_2439970, EPI_ISL_2440529, EPI_ISL_2440536, EPI_ISL_2440538, EPI_ISL_2440540, EPI_ISL_2440545, EPI_ISL_2440547, EPI_ISL_2440558, EPI_ISL_2440562, EPI_ISL_2440568, EPI_ISL_2440580, EPI_ISL_2440590, EPI_ISL_2440592, EPI_ISL_2440600, EPI_ISL_2440608, EPI_ISL_2440624, EPI_ISL_2440625, EPI_ISL_2440643, EPI_ISL_2440656, EPI_ISL_2440660, EPI_ISL_2440664, EPI_ISL_2440665, EPI_ISL_2440666, EPI_ISL_2440669, EPI_ISL_2440678, EPI_ISL_2440696, EPI_ISL_2440702, EPI_ISL_2440704, EPI_ISL_2440707, EPI_ISL_2440708, EPI_ISL_2440709, EPI_ISL_2440713, EPI_ISL_2440717, EPI_ISL_2440721, EPI_ISL_2440725, EPI_ISL_2440735, EPI_ISL_2440738, EPI_ISL_2440739, EPI_ISL_2440763, EPI_ISL_2440764, EPI_ISL_2440768, EPI_ISL_2440772, EPI_ISL_2440775, EPI_ISL_2440778, EPI_ISL_2440782, EPI_ISL_2440790, EPI_ISL_2440798, EPI_ISL_2440799, EPI_ISL_2440807, EPI_ISL_2440817, EPI_ISL_2440824, EPI_ISL_2440825, EPI_ISL_2440838, EPI_ISL_2440842, EPI_ISL_2440851, EPI_ISL_2440855, EPI_ISL_2440857, EPI_ISL_2440861, EPI_ISL_2440863, EPI_ISL_2440865, EPI_ISL_2440866, EPI_ISL_2440886, EPI_ISL_2440888, EPI_ISL_2451299, EPI_ISL_2451307, EPI_ISL_2451318, EPI_ISL_2451320, EPI_ISL_2451326, EPI_ISL_2451329, EPI_ISL_2451333, EPI_ISL_2451335, EPI_ISL_2451351, EPI_ISL_2451357, EPI_ISL_2451383, EPI_ISL_2451389, EPI_ISL_2451440, EPI_ISL_2451442, EPI_ISL_2451443, EPI_ISL_2451445, EPI_ISL_2451446, EPI_ISL_2451447, EPI_ISL_2451451, EPI_ISL_2451454, EPI_ISL_2451456, EPI_ISL_2451457, EPI_ISL_2451482, EPI_ISL_2451490, EPI_ISL_2451507, EPI_ISL_2451508, EPI_ISL_2451511, EPI_ISL_2451517, EPI_ISL_2451519, EPI_ISL_2451520, EPI_ISL_2451526, EPI_ISL_2451530, EPI_ISL_2451538, EPI_ISL_2451540, EPI_ISL_2451546, EPI_ISL_2451550, EPI_ISL_2451553, EPI_ISL_2451554, EPI_ISL_2451557, EPI_ISL_2451561, EPI_ISL_2451562, EPI_ISL_2463009, EPI_ISL_2463010, EPI_ISL_2463016, EPI_ISL_2463021, EPI_ISL_2463027, EPI_ISL_2463036, EPI_ISL_2463039, EPI_ISL_2463046, EPI_ISL_2463048, EPI_ISL_2463054, EPI_ISL_2463058, EPI_ISL_2463060, EPI_ISL_2463071, EPI_ISL_2463075, EPI_ISL_2463082, EPI_ISL_2463083, EPI_ISL_2463087, EPI_ISL_2463093, EPI_ISL_2463099, EPI_ISL_2463117, EPI_ISL_2463118, EPI_ISL_2463139, EPI_ISL_2463143, EPI_ISL_2463206, EPI_ISL_2463207, EPI_ISL_2488821, EPI_ISL_2488822, EPI_ISL_2488827, EPI_ISL_2488830, EPI_ISL_2488834, EPI_ISL_2488836, EPI_ISL_2488837, EPI_ISL_2488838, EPI_ISL_2488842, EPI_ISL_2488848, EPI_ISL_2526805, EPI_ISL_2526956, EPI_ISL_2527026, EPI_ISL_2527043, EPI_ISL_2529125, EPI_ISL_2529131, EPI_ISL_2529254, EPI_ISL_2529258, EPI_ISL_2529276, EPI_ISL_2534520, EPI_ISL_2534553, EPI_ISL_2534556, EPI_ISL_2599654, EPI_ISL_2599662, EPI_ISL_2599665, EPI_ISL_2599669, EPI_ISL_2599670, EPI_ISL_2599684, EPI_ISL_2599688, EPI_ISL_2599691, EPI_ISL_2599701, EPI_ISL_2599727, EPI_ISL_2599738, EPI_ISL_2611845, EPI_ISL_2611868, EPI_ISL_2649953, EPI_ISL_2649957, EPI_ISL_2649960, EPI_ISL_2652287, EPI_ISL_2652291, EPI_ISL_2652297, EPI_ISL_2652308, EPI_ISL_2652317, EPI_ISL_2652351, EPI_ISL_2652352, EPI_ISL_2686365, EPI_ISL_2868848, EPI_ISL_2869047, EPI_ISL_2870657, EPI_ISL_2871053, EPI_ISL_2871116, EPI_ISL_2871904, EPI_ISL_2872009, EPI_ISL_2874603, EPI_ISL_2874750, EPI_ISL_2874782, EPI_ISL_2876111, EPI_ISL_2876123, EPI_ISL_2876145, EPI_ISL_2876865, EPI_ISL_2930064, EPI_ISL_2930127, EPI_ISL_2930133, EPI_ISL_2930200, EPI_ISL_2930202, EPI_ISL_2930261, EPI_ISL_3018334, EPI_ISL_3018366, EPI_ISL_3018441, EPI_ISL_3113800, EPI_ISL_3113914, EPI_ISL_3114094, EPI_ISL_3114120, EPI_ISL_3114136, EPI_ISL_3114242, EPI_ISL_3114244, EPI_ISL_3114413, EPI_ISL_3114454, EPI_ISL_3114492, EPI_ISL_3114522, EPI_ISL_3114528, EPI_ISL_3114554, EPI_ISL_3114565, EPI_ISL_3114566, EPI_ISL_3114628, EPI_ISL_3212704, EPI_ISL_3212986, EPI_ISL_3213079, EPI_ISL_3303880, EPI_ISL_3305165, EPI_ISL_3305175, EPI_ISL_3305176, EPI_ISL_3330754, EPI_ISL_3330824, EPI_ISL_3330928, EPI_ISL_3330952, EPI_ISL_3331068, EPI_ISL_3331102, EPI_ISL_3331232 | | | |
| see above | Quest Diagnostics Incorporated | Centers for Disease Control and Prevention Division of Viral Diseases, Pathogen Discovery | A. Gerasimova; A. Perez; Adrian Paskey; B. Anderson; Benjamin Rambo-Martin; Christopher Gulvick; Clinton R. Paden; Dakota Howard; Darlene Wagner; Dhwani Batra; Duncan MacCannell; F. Lacbawan; I. A. Shlyakhter; Jason Caravas; K.E. Livingston; Kara Moser; L.E. Bernstein; M. Hua; Matthew Schmerer; P. Tanpaiboon; Peter W. Cook; R. M. Kagan; R. Owen; R. V. Rolando; S. H. Rosenthal; Scott Sammons; Shatavia Morrison; Y. Liu; Yvette Unoarumhi |
| EPI_ISL_2253143, EPI_ISL_2253150, EPI_ISL_2253157, EPI_ISL_2254303, EPI_ISL_2254373, EPI_ISL_2254379, EPI_ISL_2254397, EPI_ISL_2254403, EPI_ISL_2254411, EPI_ISL_2254438, EPI_ISL_2254462, EPI_ISL_2254492, EPI_ISL_3091705, EPI_ISL_3091735, EPI_ISL_3091765, EPI_ISL_3091911, EPI_ISL_3091920, EPI_ISL_3091940, EPI_ISL_3091983 | | | |
| see above | Respiratory Viruses Branch, Centers for Disease Control and Prevention | Respiratory Viruses Branch, Centers for Disease Control and Prevention | Anderson, B.; Batra, D.; Bernstein, L.; C.R.; Campbell, P.; Caravas, J.; Case, R.; Clark, C.; Cook; Gerasimova, A.; Ghorpade, V.; Gulvick, C.; Hardison, M.; Houdeshell, H.; Howard, D.; Hua, M.; I.A.; Kagan; Kvalvaag, O.; Lacbawan, F.; Liu, Y.; Livingston, K.; MacCannell, D.; Morrison, S.; Moser, K.; Nall, D.; Owen, R.; P.W.; Paden; Paskey, A.; Perez, A.; R.M.; R.V.; Rambo-Martin, B.; Rolando; Rosenthal; S.H.; Sammons, S.; Sanders, E.; Schmerer, M.; Shlyakhter; Tanpaiboon, P.; Unoarumhi, Y.; Vest, A.; Wagner, D.; Westlund, S. |
| EPI_ISL_2283890, EPI_ISL_2283891, EPI_ISL_2283892, EPI_ISL_2283893, EPI_ISL_2283895, EPI_ISL_2283896, EPI_ISL_2283897, EPI_ISL_2283898, EPI_ISL_2283899, EPI_ISL_2283900, EPI_ISL_2283902, EPI_ISL_2283907, EPI_ISL_2283908, EPI_ISL_2283910, EPI_ISL_2283914, EPI_ISL_2283915, EPI_ISL_2283916, EPI_ISL_2283920, EPI_ISL_2283922, EPI_ISL_2283923, EPI_ISL_2283924, EPI_ISL_2283925, EPI_ISL_2283927, EPI_ISL_2283932, EPI_ISL_2283936, EPI_ISL_2283940, EPI_ISL_2283944, EPI_ISL_2283946, EPI_ISL_2283948, EPI_ISL_2283949, EPI_ISL_2283957, EPI_ISL_2283958, EPI_ISL_2283960, EPI_ISL_2283964, EPI_ISL_2283970, EPI_ISL_2283972, EPI_ISL_2283974, EPI_ISL_2283975, EPI_ISL_2283977, EPI_ISL_2283978, EPI_ISL_2283979, EPI_ISL_2283980, EPI_ISL_2283981, EPI_ISL_2283983, EPI_ISL_2283985, EPI_ISL_2283988, EPI_ISL_2283989, EPI_ISL_2283990, EPI_ISL_2283991, EPI_ISL_2283992, EPI_ISL_2283993, EPI_ISL_2283994, EPI_ISL_2283995, EPI_ISL_2283996, EPI_ISL_2283999, EPI_ISL_2284001, EPI_ISL_2284003, EPI_ISL_2284004, EPI_ISL_2284005, EPI_ISL_2284006, EPI_ISL_2284007, EPI_ISL_2284008, EPI_ISL_2284009, EPI_ISL_2284012, EPI_ISL_2284013, EPI_ISL_2284015, EPI_ISL_2284016, EPI_ISL_2284017, EPI_ISL_2284018, EPI_ISL_2284019, EPI_ISL_2284021, EPI_ISL_2284023, EPI_ISL_2284024, EPI_ISL_2284025, EPI_ISL_2284026, EPI_ISL_2284027, EPI_ISL_2284029, EPI_ISL_2284030, EPI_ISL_2284031, EPI_ISL_2284032, EPI_ISL_2284033, EPI_ISL_2284034, EPI_ISL_2284035, EPI_ISL_2284036, EPI_ISL_2284037, EPI_ISL_2284038, EPI_ISL_2284039, EPI_ISL_2284041, EPI_ISL_2284042, EPI_ISL_2284043, EPI_ISL_2284044, EPI_ISL_2284045, EPI_ISL_2284046, EPI_ISL_2284047, EPI_ISL_2284048, EPI_ISL_2284050, EPI_ISL_2284051, EPI_ISL_2284054, EPI_ISL_2284055, EPI_ISL_2284056, EPI_ISL_2284057, EPI_ISL_2284058, EPI_ISL_2284059, EPI_ISL_2284060, EPI_ISL_2284061, EPI_ISL_2284062, EPI_ISL_2284066, EPI_ISL_2284067, EPI_ISL_2284179, EPI_ISL_2284195 | | | |
| see above | VA Connecticut Healthcare System | Yale Center for Genomic Analysis | Brooke Sullivan; Curt Scharfe; Irina Tikhonova; Kaya Bilguvar; Shrikant Mane |
| EPI_ISL_3370239 | Yale Clinical Virology Lab | Grubaugh Lab - Yale School of Public Health | Anderson Brito; Annie Watkins; Chaney Kalinich; Chantal Vogels; Isabel Ott; Jessica Rothman; Joseph Fauver; Kendall Billig; Mallery Breban; Marie L. Landry; Mary Petrone; Nathan Grubaugh; Tara Alpert; Tobias Koch |
| EPI_ISL_2159045 | Yale Clinical Virology Lab | Yale Center for Genomic Analysis | Brooke Sullivan; Curt Scharfe; Irina Tikhonova; Kaya Bilguvar; Shrikant Mane |
