## Supplementary material for "Comparative transmissibility of SARS-CoV-2 variants Delta and Alpha in New England, USA": DataS1-7: DataS5_New-Hampshire.pdf

We gratefully acknowledge the following Authors from the Originating laboratories responsible for obtaining the specimens, as well as the Submitting laboratories where the genome data were generated and shared via GISAID, on which this research is based.

All Submitters of data may be contacted directly via www.gisaid.org

Authors are sorted alphabetically.

| Accession ID | Originating Laboratory | Submitting Laboratory | Authors |
| --- | --- | --- | --- |
| EPI_ISL_1445451, EPI_ISL_1445571, EPI_ISL_1445603, EPI_ISL_1446016, EPI_ISL_1446019, EPI_ISL_1446094, EPI_ISL_1479562, EPI_ISL_1479821, EPI_ISL_1479822, EPI_ISL_1491802, EPI_ISL_1513230, EPI_ISL_1513485, EPI_ISL_1550633, EPI_ISL_1551125, EPI_ISL_1560538, EPI_ISL_1562476, EPI_ISL_1562875, EPI_ISL_1650086, EPI_ISL_1651011, EPI_ISL_1651019, EPI_ISL_1667534, EPI_ISL_1687918, EPI_ISL_1688207, EPI_ISL_1688208, EPI_ISL_1690011, EPI_ISL_1695496, EPI_ISL_1702802, EPI_ISL_1735907, EPI_ISL_1737848, EPI_ISL_1737851, EPI_ISL_1835936, EPI_ISL_1835959, EPI_ISL_1835963, EPI_ISL_1836076, EPI_ISL_1836341, EPI_ISL_1836491, EPI_ISL_1836881, EPI_ISL_1836882, EPI_ISL_1837027, EPI_ISL_1837028, EPI_ISL_1837029, EPI_ISL_1837304, EPI_ISL_1837393, EPI_ISL_1837433, EPI_ISL_1837434, EPI_ISL_1837859, EPI_ISL_1925279, EPI_ISL_1925419, EPI_ISL_1996102, EPI_ISL_1996433, EPI_ISL_1996434, EPI_ISL_1997606, EPI_ISL_1997978, EPI_ISL_1998128, EPI_ISL_1998222, EPI_ISL_1998229, EPI_ISL_1998864, EPI_ISL_2039897, EPI_ISL_2040034, EPI_ISL_2042884, EPI_ISL_2145941, EPI_ISL_2146652, EPI_ISL_2146653, EPI_ISL_2147454, EPI_ISL_2147460, EPI_ISL_2147923, EPI_ISL_2147924, EPI_ISL_2148238, EPI_ISL_2148276, EPI_ISL_2148955, EPI_ISL_2149409, EPI_ISL_2150293, EPI_ISL_2150734, EPI_ISL_2150890, EPI_ISL_2150892, EPI_ISL_2150895, EPI_ISL_2159455, EPI_ISL_2180786, EPI_ISL_2181498, EPI_ISL_2186210, EPI_ISL_2186230, EPI_ISL_2241928, EPI_ISL_2242085, EPI_ISL_2242762, EPI_ISL_2243009, EPI_ISL_2243031, EPI_ISL_2243076, EPI_ISL_2244502, EPI_ISL_2244542, EPI_ISL_2268068, EPI_ISL_2280733, EPI_ISL_2280775, EPI_ISL_2280922, EPI_ISL_2321259, EPI_ISL_2370251, EPI_ISL_2482412, EPI_ISL_2686986, EPI_ISL_3218315 | | | |
| see above | Aegis Sciences Corporation | Centers for Disease Control and Prevention Division of Viral Diseases, Pathogen Discovery | Adrian Paskey; Alec Vest; Benjamin Rambo-Martin; Christopher Gulvick; Clinton R. Paden; Cyndi Clark; Dakota Howard; Darlene Wagner; Dhwani Batra; Dillon Nall; Duncan MacCannell; Ethan Sanders; Holly Houdeshell; Jason Caravas; Kara Moser; Matthew Hardison; Matthew Schmerer; Ola Kvalvaag; Patrick Campbell; Peter W. Cook; Rob Case; Scott Sammons; Shatavia Morrison; Shaun Westlund; Vikramsinha Ghorpade; Yvette Unoarumhi |
| EPI_ISL_2991947, EPI_ISL_2991948, EPI_ISL_2991950, EPI_ISL_2991951, EPI_ISL_2991952, EPI_ISL_2991953, EPI_ISL_2991954, EPI_ISL_2991955, EPI_ISL_2991956, EPI_ISL_2991957, EPI_ISL_2991958, EPI_ISL_2991960, EPI_ISL_2991961 | | | |
| see above | Broad Institute Clinical Research Sequencing Platform | Infectious Disease Program, Broad Institute of Harvard and MIT | A.E.; Adams, G.; Anahtar, M.; B.L.; B.W.; Bauer, M.; Birren; Branda, J.; Carter, A.; Cerrato, F.; Chaluvadi, S.; Chapman; Cusick, C.; D.J.; DeRuff, K.; Flowers, K.; Gallagher, G.; Gladden-Young, A.; Gnirke, A.; Harris, J.; J.E.; K.J.; LaRocque, R.; Lagerborg, K.; Lemieux; Lin; Loreth, C.; MacInnis; Neumann, A.; Normandin, E.; P.C.; Park; Pierce, V.; Reilly, S.; Rosenberg, E.; Rudy, M.; Ryan, E.; S.B.; Sabeti; Shaw, B.; Siddle; Slater, D.; Smole, S.; Tomkins-Tinch, C.; Turbett, S. |
| EPI_ISL_1413157, EPI_ISL_1413163, EPI_ISL_1413165, EPI_ISL_1413168, EPI_ISL_1413179, EPI_ISL_1413226, EPI_ISL_1413232, EPI_ISL_1413238, EPI_ISL_1413249, EPI_ISL_1413311, EPI_ISL_1413322, EPI_ISL_1413325, EPI_ISL_1413336, EPI_ISL_1413339, EPI_ISL_1413374, EPI_ISL_1516446, EPI_ISL_1516447, EPI_ISL_1516448, EPI_ISL_1516449, EPI_ISL_1516450, EPI_ISL_1516451, EPI_ISL_1516453, EPI_ISL_1516456, EPI_ISL_1516458, EPI_ISL_1516459, EPI_ISL_1516462, EPI_ISL_1516463, EPI_ISL_1516483, EPI_ISL_1516519, EPI_ISL_1516566, EPI_ISL_1516573, EPI_ISL_1516581, EPI_ISL_1516582, EPI_ISL_1516583, EPI_ISL_1578241, EPI_ISL_1578283, EPI_ISL_1578303, EPI_ISL_1578335, EPI_ISL_1578348, EPI_ISL_1578383, EPI_ISL_1578397, EPI_ISL_1578434, EPI_ISL_1578440, EPI_ISL_1710053, EPI_ISL_1710066, EPI_ISL_1710070, EPI_ISL_1710071, EPI_ISL_1710072, EPI_ISL_1710133, EPI_ISL_1710137, EPI_ISL_1710139, EPI_ISL_1710141, EPI_ISL_1710143, EPI_ISL_1710147, EPI_ISL_1710148, EPI_ISL_1757042, EPI_ISL_1757081, EPI_ISL_1757083, EPI_ISL_1757084, EPI_ISL_1757085, EPI_ISL_1757086, EPI_ISL_1757087, EPI_ISL_1757088, EPI_ISL_1757267, EPI_ISL_1757276, EPI_ISL_1757282, EPI_ISL_1757288, EPI_ISL_1757289, EPI_ISL_1757293, EPI_ISL_1757297, EPI_ISL_1787242, EPI_ISL_1787243, EPI_ISL_1787244, EPI_ISL_1787245, EPI_ISL_1787246, EPI_ISL_1787247, EPI_ISL_1787248, EPI_ISL_1787249, EPI_ISL_1787250, EPI_ISL_1787255, EPI_ISL_1787257, EPI_ISL_1787258, EPI_ISL_1825197, EPI_ISL_1825232, EPI_ISL_1825240, EPI_ISL_1825455, EPI_ISL_1825456, EPI_ISL_1825458, EPI_ISL_1825465, EPI_ISL_1825467, EPI_ISL_1825473, EPI_ISL_1825529, EPI_ISL_1825588, EPI_ISL_1825590, EPI_ISL_1918954, EPI_ISL_1918957, EPI_ISL_1918959, EPI_ISL_1918960, EPI_ISL_1918961, EPI_ISL_1918963, EPI_ISL_1918964, EPI_ISL_1918966, EPI_ISL_1919070, EPI_ISL_1919072, EPI_ISL_1919075, EPI_ISL_1919078, EPI_ISL_1919079, EPI_ISL_1919080, EPI_ISL_1919082, EPI_ISL_1919083, EPI_ISL_1919085, EPI_ISL_1919088, EPI_ISL_1968484, EPI_ISL_1968485, EPI_ISL_1968486, EPI_ISL_1968487, EPI_ISL_1968488, EPI_ISL_1968490, EPI_ISL_1968491, EPI_ISL_1968492, EPI_ISL_1968494, EPI_ISL_1968495, EPI_ISL_1968497, EPI_ISL_1968498, EPI_ISL_1968501, EPI_ISL_1968502, EPI_ISL_1968503, EPI_ISL_1968504, EPI_ISL_1968507, EPI_ISL_1971990, EPI_ISL_1971991, EPI_ISL_1971994, EPI_ISL_1971995, EPI_ISL_1971996, EPI_ISL_1971998, EPI_ISL_1971999, EPI_ISL_1972000, EPI_ISL_1972001, EPI_ISL_1972003, EPI_ISL_1972006, EPI_ISL_1972008, EPI_ISL_1972009, EPI_ISL_1972010, EPI_ISL_1972011, EPI_ISL_1972012, EPI_ISL_2096681, EPI_ISL_2096682, EPI_ISL_2096683, EPI_ISL_2096684, EPI_ISL_2096685, EPI_ISL_2096686, EPI_ISL_2096687, EPI_ISL_2096688, EPI_ISL_2096689, EPI_ISL_2096690, EPI_ISL_2096691, EPI_ISL_2096692, EPI_ISL_2096693, EPI_ISL_2096694, EPI_ISL_2096695, EPI_ISL_2096696, EPI_ISL_2096697, EPI_ISL_2096698, EPI_ISL_2096699, EPI_ISL_2096700, EPI_ISL_2096701, EPI_ISL_2096702, EPI_ISL_2096704, EPI_ISL_2096705, EPI_ISL_2096706, EPI_ISL_2096707, EPI_ISL_2096708, EPI_ISL_2096709, EPI_ISL_2096710, EPI_ISL_2096711, EPI_ISL_2096712, EPI_ISL_2096713, EPI_ISL_2104301, EPI_ISL_2104302, EPI_ISL_2104303, EPI_ISL_2104306, EPI_ISL_2104307, EPI_ISL_2104308, EPI_ISL_2104309, EPI_ISL_2104310, EPI_ISL_2104311, EPI_ISL_2104312, EPI_ISL_2104313, EPI_ISL_2104315, EPI_ISL_2104317, EPI_ISL_2104318, EPI_ISL_2104319, EPI_ISL_2104320, EPI_ISL_2104321, EPI_ISL_2104322, EPI_ISL_2104323, EPI_ISL_2104325, EPI_ISL_2104327, EPI_ISL_2104329, EPI_ISL_2104330, EPI_ISL_2193115, EPI_ISL_2193116, EPI_ISL_2193117, EPI_ISL_2193118, EPI_ISL_2193119, EPI_ISL_2193121, EPI_ISL_2193122, EPI_ISL_2193123, EPI_ISL_2193124, EPI_ISL_2193125, EPI_ISL_2193126, EPI_ISL_2193128, EPI_ISL_2193130, EPI_ISL_2193132, EPI_ISL_2193133, EPI_ISL_2193134, EPI_ISL_2193136, EPI_ISL_2193137, EPI_ISL_2193139, EPI_ISL_2193141, EPI_ISL_2193144, EPI_ISL_2193146, EPI_ISL_2193148, EPI_ISL_2193149, EPI_ISL_2193150, EPI_ISL_2312422, EPI_ISL_2312423, EPI_ISL_2312424, EPI_ISL_2312425, EPI_ISL_2312426, EPI_ISL_2312427, EPI_ISL_2312428, EPI_ISL_2312429, EPI_ISL_2312430, EPI_ISL_2312431, EPI_ISL_2312432, EPI_ISL_2312433, EPI_ISL_2312434, EPI_ISL_2312435, EPI_ISL_2312436, EPI_ISL_2312438, EPI_ISL_2312439, EPI_ISL_2312440, EPI_ISL_2312441, EPI_ISL_2312442, EPI_ISL_3007461, EPI_ISL_3007462, EPI_ISL_3007464, EPI_ISL_3007466, EPI_ISL_3007467, EPI_ISL_3007468, EPI_ISL_3085049, EPI_ISL_3085051, EPI_ISL_3085053, EPI_ISL_3085055, EPI_ISL_3085105, EPI_ISL_3085106, EPI_ISL_3085107, EPI_ISL_3085108, EPI_ISL_3127389, EPI_ISL_3134536, EPI_ISL_3153176, EPI_ISL_3156017, EPI_ISL_3156018, EPI_ISL_3156019 | | | |
| see above | Broad Institute Clinical Research Sequencing Platform | Infectious Disease Program, Broad Institute of Harvard and MIT | Adams, G.; B.L.; B.W.; Bauer, M.; Birren; Blumenstiel, B.; Brown, C.; Carter, A.; Chaluvadi, S.; D.J.; DeFelice, M.; DeRuff, K.; Dodge, S.; Gabriel, S.; Gallagher, G.; Gladden-Young, A.; Granger, B.; J.E.; K.J.; Lagerborg, K.; Larkin, K.; Lee, M.; Lemieux; Lennon, N.; Loreth, C.; Madoff, L.; McGovern, S.; Meldrim, J.; Normandin, E.; P.C.; Park; Pearlman, L.; Reilly, S.; Rudy, M.; Sabeti; Siddle; Smole, S.; Tomkins-Tinch, C.; Vicente, G.; and MacInnis |
| EPI_ISL_1262080, EPI_ISL_1267799, EPI_ISL_1512038, EPI_ISL_1512120, EPI_ISL_1512578, EPI_ISL_1553498, EPI_ISL_1576125, EPI_ISL_1592469, EPI_ISL_1679458, EPI_ISL_1734604, EPI_ISL_1797404, EPI_ISL_1803685, EPI_ISL_1924064, EPI_ISL_2145677 | | | |
| see above | Helix/Illumina | Centers for Disease Control and Prevention Division of Viral Diseases, Pathogen Discovery | Adrian Paskey; Alexandre Bolze; Ary Ascencio; Ben L. Rambo-Martin; Benjamin Rambo-Martin; Brad Sickler; Charlotte Rivera-Garcia; Christine Tran; Christopher Gulvick; Clinton R. Paden; Dakota Howard; Darlene Wagner; David Becker; Dhwani Batra; Duncan MacCannell; Efren Sandoval; Eileen de Feo; Elizabeth Cirulli; Eric Allen; Geraint Levan; James Lu; Jan Antico; Jason Caravas; Jason Nguyen; Jimmy Ramirez; Jingtao Liu; Kara Moser; Kelly Schiabor Barrett; Kim Gietzen; Magnus Isaksson; Marc Laurent; Matthew Schmerer; Matthew Tolentino; Nicole L. Washington; Peter W. Cook; Phil Febbo; Ryan Cho; Scott Sammons; Shannon Wickline; Shatavia Morrison; Sherry Wang; Simon White; Summer Galloway; Suxiang Tong; Tyler Cassens; William Lee; Yvette Unoarumhi |
| EPI_ISL_850990, EPI_ISL_876708 | Helix/Illumina | Genomics and Discovery, Respiratory Viruses Branch, Division of Viral Diseases, Centers for Disease Control and Prevention | ; Alexandre Bolze; Ary Ascencio; Ben L. Rambo-Martin; Ben L. Rambo-Martin Eileen de Feo; Brad Sickler; Charlotte Rivera-Garcia; Christine Tran; Clinton R. Paden; David Becker; Dhwani Batra; Duncan MacCannell; Efren Sandoval; Eileen de Feo; Elizabeth Cirulli; Eric Allen; Geraint Levan; James Lu; Jan Antico; Jason Nguyen; Jimmy Ramirez; Jingtao Liu; Kelly Schiabor Barrett; Kim Gietzen; Magnus Isaksson; Marc Laurent; Matthew Tolentino; Nicole L. Washington; Peter W. Cook; Phil Febbo; Ryan Cho; Shannon Wickline; Sherry Wang; Simon White; Summer Galloway; Suxiang Tong; Tyler Cassens; William Lee |
| EPI_ISL_966529, EPI_ISL_966726, EPI_ISL_966919, EPI_ISL_967357, EPI_ISL_967841, EPI_ISL_978765 | Helix/Illumina | Respiratory Viruses Branch, Division of Viral Diseases, Centers for Disease Control and Prevention | ; Alexandre Bolze; Ary Ascencio; Ben L. Rambo-Martin; Brad Sickler; Charlotte Rivera-Garcia; Christine Tran; Clinton R. Paden; Dakota Howard; David Becker; Dhwani Batra; Duncan MacCannell; Efren Sandoval; Eileen de Feo; Elizabeth Cirulli; Eric Allen; Geraint Levan; James Lu; Jan Antico; Jason Nguyen; Jimmy Ramirez; Jingtao Liu; Kelly Schiabor Barrett; Kim Gietzen; Magnus Isaksson; Marc Laurent; Matthew Tolentino; Nicole L. Washington; Peter W. Cook; Phil Febbo; Ryan Cho; Shannon Wickline; Sherry Wang; Simon White; Summer Galloway; Suxiang Tong; Tyler Cassens; William Lee |
| EPI_ISL_886186, EPI_ISL_886227, EPI_ISL_886247, EPI_ISL_886318, EPI_ISL_886364 | Labcorp | Genomics and Discovery, Respiratory Viruses Branch, Division of Viral Diseases, Centers for Disease Control and Prevention | ; Amanda Douglas; Amanda Suchanek; Andrea Throop; Ayla Burns; Ben L. Rambo-Martin; Bobbi Croy; Brian Krueger; Brian Norvell; Christos Petropoulos; Clinton R. Paden; Craig Lukasik; Debbie Boles; Dhwani Batra; Duncan MacCannell; Eyad Almasri; Goran Stevovic; Howard Engler; Hrushikesh Deshmukh; Jake Humphrey; Jana Schroth; Joe Voshell; John Pruitt; Jonathan Meltzer; Jonathan Williams; Kimberly Wagner; Lax Iyer; Lyndon Tilson; Manoj Jain; Marcia Eisenberg; Mary Ann Cristobal; Mary Williamson; Michael Levandoski; Mike Sapeta; Mindy Nye; Minoo Agarwal; Mohan Kolli; Nuthawin Charoensri; Oren Cohen; Peter W. Cook; Prashant Gupta; Qian Zeng; Rama Ghatti; Scott Parker; Scott Ryan; Stanley Letovsky; Steven Ragan; Summer Galloway; Suresh Babu Selvaraju; Susan Countryman; Susan Hicks; Suxiang Tong; Suzanne Dale; Thomas Urban; Tim Kuphal; Tricia Zwiefelhofer; Vincent Drouillon |
| EPI_ISL_1193371, EPI_ISL_1220929, EPI_ISL_1220930, EPI_ISL_1220931, EPI_ISL_1220932, EPI_ISL_1220934, EPI_ISL_1221045, EPI_ISL_1221090, EPI_ISL_1221405, EPI_ISL_1221428, EPI_ISL_1221764, EPI_ISL_1221877, EPI_ISL_1221880, EPI_ISL_1221883, EPI_ISL_1221889, EPI_ISL_1221890, EPI_ISL_1222042, EPI_ISL_1222045, EPI_ISL_1222046, EPI_ISL_1298334, EPI_ISL_1319051, EPI_ISL_1319052, EPI_ISL_1320342, EPI_ISL_1320359, EPI_ISL_1320496, EPI_ISL_1320499, EPI_ISL_1320537, EPI_ISL_1320543, EPI_ISL_1320591, EPI_ISL_1320728, EPI_ISL_1320807, EPI_ISL_1320837, EPI_ISL_1320838, EPI_ISL_1320869, EPI_ISL_1320871, EPI_ISL_1320889, EPI_ISL_1320922, EPI_ISL_1320950, EPI_ISL_1320971, EPI_ISL_1320972, EPI_ISL_1320973, EPI_ISL_1320975, EPI_ISL_1320976, EPI_ISL_1320977, EPI_ISL_1320992, EPI_ISL_1320999, EPI_ISL_1321009, EPI_ISL_1338399, EPI_ISL_1338409, EPI_ISL_1338414, EPI_ISL_1338453, EPI_ISL_1338455, EPI_ISL_1338458, EPI_ISL_1338459, EPI_ISL_1338460, EPI_ISL_1338543, EPI_ISL_1338544, EPI_ISL_1338546, EPI_ISL_1338547, EPI_ISL_1338561, EPI_ISL_1338562, EPI_ISL_1338564, EPI_ISL_1338567, EPI_ISL_1338575, EPI_ISL_1338602, EPI_ISL_1338603, EPI_ISL_1422101, EPI_ISL_1422120, EPI_ISL_1422121, EPI_ISL_1422149, EPI_ISL_1422153, EPI_ISL_1422162, EPI_ISL_1462337, EPI_ISL_1462338, EPI_ISL_1462384, EPI_ISL_1462385, EPI_ISL_1462386, EPI_ISL_1462387, EPI_ISL_1462390, EPI_ISL_1462394, EPI_ISL_1462423, EPI_ISL_1462447, EPI_ISL_1462451, EPI_ISL_1462465, EPI_ISL_1462470, EPI_ISL_1462471, EPI_ISL_1462809, EPI_ISL_1462830, EPI_ISL_1462831, EPI_ISL_1462836, EPI_ISL_1462837, EPI_ISL_1462839, EPI_ISL_1462844, EPI_ISL_1463197, EPI_ISL_1463199, EPI_ISL_1463203, EPI_ISL_1463204, EPI_ISL_1463205, EPI_ISL_1463206, EPI_ISL_1463224, EPI_ISL_1463225, EPI_ISL_1463230, EPI_ISL_1463231, EPI_ISL_1463232, EPI_ISL_1463233, EPI_ISL_1463244, EPI_ISL_1463248, EPI_ISL_1463253, EPI_ISL_1463284, EPI_ISL_1463291, EPI_ISL_1463306, EPI_ISL_1463325, EPI_ISL_1463328, EPI_ISL_1463329, EPI_ISL_1463330, EPI_ISL_1463331, EPI_ISL_1463332, EPI_ISL_1463337, EPI_ISL_1463339, EPI_ISL_1463343, EPI_ISL_1463352, EPI_ISL_1463353, EPI_ISL_1463371, EPI_ISL_1463373, EPI_ISL_1463374, EPI_ISL_1463376, EPI_ISL_1463386, EPI_ISL_1463402, EPI_ISL_1463420, EPI_ISL_1463424, EPI_ISL_1463432, EPI_ISL_1463433, EPI_ISL_1481029, EPI_ISL_1513974, EPI_ISL_1514011, EPI_ISL_1548450, EPI_ISL_1548451, EPI_ISL_1548473, EPI_ISL_1548484, EPI_ISL_1548485, EPI_ISL_1548503, EPI_ISL_1548511, EPI_ISL_1548526, EPI_ISL_1548530, EPI_ISL_1548721, EPI_ISL_1549040, EPI_ISL_1549041, EPI_ISL_1549046, EPI_ISL_1549075, EPI_ISL_1549086, EPI_ISL_1549088, EPI_ISL_1549089, EPI_ISL_1549090, EPI_ISL_1549107, EPI_ISL_1549115, EPI_ISL_1549126, EPI_ISL_1549131, EPI_ISL_1549132, EPI_ISL_1549344, EPI_ISL_1549362, EPI_ISL_1549384, EPI_ISL_1610229, EPI_ISL_1610254, EPI_ISL_1610255, EPI_ISL_1610257, EPI_ISL_1610259, EPI_ISL_1610260, EPI_ISL_1610276, EPI_ISL_1610278, EPI_ISL_1610281, EPI_ISL_1610282, EPI_ISL_1610284, EPI_ISL_1610285, EPI_ISL_1610306, EPI_ISL_1610310, EPI_ISL_1610314, EPI_ISL_1610319, EPI_ISL_1610320, EPI_ISL_1610328, EPI_ISL_1610344, EPI_ISL_1610355, EPI_ISL_1610357, EPI_ISL_1610358, EPI_ISL_1610392, EPI_ISL_1610393, EPI_ISL_1610394, EPI_ISL_1610436, EPI_ISL_1610438, EPI_ISL_1610443, EPI_ISL_1610446, EPI_ISL_1610457, EPI_ISL_1610465, EPI_ISL_1610468, EPI_ISL_1610470, EPI_ISL_1610471, EPI_ISL_1610579, EPI_ISL_1610582, EPI_ISL_1610594, EPI_ISL_1610609, EPI_ISL_1610619, EPI_ISL_1610631, EPI_ISL_1610633, EPI_ISL_1610660, EPI_ISL_1610894, EPI_ISL_1610899, EPI_ISL_1610910, EPI_ISL_1610915, EPI_ISL_1610918, EPI_ISL_1610934, EPI_ISL_1610946, EPI_ISL_1610960, EPI_ISL_1610961, EPI_ISL_1610962, EPI_ISL_1610963, EPI_ISL_1610964, EPI_ISL_1610965, EPI_ISL_1610970, EPI_ISL_1610972, EPI_ISL_1610973, EPI_ISL_1610974, EPI_ISL_1611031, EPI_ISL_1611033, EPI_ISL_1611107, EPI_ISL_1611109, EPI_ISL_1611110, EPI_ISL_1680466, EPI_ISL_1680467, EPI_ISL_1680468, EPI_ISL_1680496, EPI_ISL_1680512, EPI_ISL_1680513, EPI_ISL_1680530, EPI_ISL_1680538, EPI_ISL_1680539, EPI_ISL_1680556, EPI_ISL_1680557, EPI_ISL_1680558, EPI_ISL_1680561, EPI_ISL_1680570, EPI_ISL_1680575, EPI_ISL_1680576, EPI_ISL_1680577, EPI_ISL_1680619, EPI_ISL_1680645, EPI_ISL_1680646, EPI_ISL_1680648, EPI_ISL_1680649, EPI_ISL_1680652, EPI_ISL_1680655, EPI_ISL_1680660, EPI_ISL_1680671, EPI_ISL_1680683, EPI_ISL_1680684, EPI_ISL_1680688, EPI_ISL_1680691, EPI_ISL_1680787, EPI_ISL_1680788, EPI_ISL_1680793, EPI_ISL_1680794, EPI_ISL_1680795, EPI_ISL_1680804, EPI_ISL_1680823, EPI_ISL_1680824, EPI_ISL_1680825, EPI_ISL_1680826, EPI_ISL_1680838, EPI_ISL_1680839, EPI_ISL_1680840, EPI_ISL_1680841, EPI_ISL_1680842, EPI_ISL_1680864, EPI_ISL_1681179, EPI_ISL_1681185, EPI_ISL_1681186, EPI_ISL_1681202, EPI_ISL_1681206, EPI_ISL_1681207, EPI_ISL_1681213, EPI_ISL_1681214, EPI_ISL_1681216, EPI_ISL_1681223, EPI_ISL_1681230, EPI_ISL_1681231, EPI_ISL_1681245, EPI_ISL_1681292, EPI_ISL_1681301, EPI_ISL_1681302, EPI_ISL_1681306, EPI_ISL_1681330, EPI_ISL_1681348, EPI_ISL_1681365, EPI_ISL_1681366, EPI_ISL_1681367, EPI_ISL_1681368, EPI_ISL_1681423, EPI_ISL_1681424, EPI_ISL_1681434, EPI_ISL_1681443, EPI_ISL_1681467, EPI_ISL_1681474, EPI_ISL_1681477, EPI_ISL_1681479, EPI_ISL_1681480, EPI_ISL_1681481, EPI_ISL_1681482, EPI_ISL_1681488, EPI_ISL_1681489, EPI_ISL_1681492, EPI_ISL_1681494, EPI_ISL_1681503, EPI_ISL_1681504, EPI_ISL_1681510, EPI_ISL_1681511, EPI_ISL_1681512, EPI_ISL_1681517, EPI_ISL_1681524, EPI_ISL_1681533, EPI_ISL_1681541, EPI_ISL_1681553, EPI_ISL_1681554, EPI_ISL_1681562, EPI_ISL_1681566, EPI_ISL_1681568, EPI_ISL_1681637, EPI_ISL_1683232, EPI_ISL_1683233, EPI_ISL_1694995, EPI_ISL_1800518, EPI_ISL_1800519, EPI_ISL_1800520, EPI_ISL_1801295, EPI_ISL_1801302, EPI_ISL_1801311, EPI_ISL_1801317, EPI_ISL_1801319, EPI_ISL_1801340, EPI_ISL_1801346, EPI_ISL_1801349, EPI_ISL_1801367, EPI_ISL_1801393, EPI_ISL_1801394, EPI_ISL_1801423, EPI_ISL_1801429, EPI_ISL_1801434, EPI_ISL_1801441, EPI_ISL_1801453, EPI_ISL_1801545, EPI_ISL_1801546, EPI_ISL_1801549, EPI_ISL_1801559, EPI_ISL_1801582, EPI_ISL_1801713, EPI_ISL_1801718, EPI_ISL_1801722, EPI_ISL_1801724, EPI_ISL_1801790, EPI_ISL_1801794, EPI_ISL_1801848, EPI_ISL_1801881, EPI_ISL_1801882, EPI_ISL_1801895, EPI_ISL_1801949, EPI_ISL_1801953, EPI_ISL_1801956, EPI_ISL_1801994, EPI_ISL_1802001, EPI_ISL_1925785, EPI_ISL_1926368, EPI_ISL_1926369, EPI_ISL_1926459, EPI_ISL_1926477, EPI_ISL_1926478, EPI_ISL_1926479, EPI_ISL_1926547, EPI_ISL_1926593, EPI_ISL_1926596, EPI_ISL_1926605, EPI_ISL_1928535, EPI_ISL_1928538, EPI_ISL_1929439, EPI_ISL_1929457, EPI_ISL_1929460, EPI_ISL_1929464, EPI_ISL_1929917, EPI_ISL_1929926, EPI_ISL_1930020, EPI_ISL_1930079, EPI_ISL_1930089, EPI_ISL_1930103, EPI_ISL_1930109, EPI_ISL_1930147, EPI_ISL_1930150, EPI_ISL_1930168, EPI_ISL_1930193, EPI_ISL_1930196, EPI_ISL_1930231, EPI_ISL_1930234, EPI_ISL_1930237, EPI_ISL_1930239, EPI_ISL_1930290, EPI_ISL_1930327, EPI_ISL_1930335, EPI_ISL_1930391, EPI_ISL_1930398, EPI_ISL_1930400, EPI_ISL_1930403, EPI_ISL_2044635, EPI_ISL_2044986, EPI_ISL_2045436, EPI_ISL_2045437, EPI_ISL_2045438, EPI_ISL_2045456, EPI_ISL_2045500, EPI_ISL_2045508, EPI_ISL_2045738, EPI_ISL_2045743, EPI_ISL_2045749, EPI_ISL_2045760, EPI_ISL_2045772, EPI_ISL_2045775, EPI_ISL_2045777, EPI_ISL_2045783, EPI_ISL_2045801, EPI_ISL_2045802, EPI_ISL_2045833, EPI_ISL_2045834, EPI_ISL_2045835, EPI_ISL_2045837, EPI_ISL_2045841, EPI_ISL_2045842, EPI_ISL_2045876, EPI_ISL_2045885, EPI_ISL_2045924, EPI_ISL_2181659, EPI_ISL_2181699, EPI_ISL_2181707, EPI_ISL_2181708, EPI_ISL_2181709, EPI_ISL_2181746, EPI_ISL_2181747, EPI_ISL_2181748, EPI_ISL_2182434, EPI_ISL_2182479, EPI_ISL_2182482, EPI_ISL_2182514, EPI_ISL_2182619, EPI_ISL_2182625, EPI_ISL_2182647, EPI_ISL_2182649, EPI_ISL_2182720, EPI_ISL_2182722, EPI_ISL_2182823, EPI_ISL_2182850, EPI_ISL_2182863, EPI_ISL_2182867, EPI_ISL_2182935, EPI_ISL_2182938, EPI_ISL_2183044, EPI_ISL_2184327, EPI_ISL_2184333, EPI_ISL_2306658, EPI_ISL_2306659, EPI_ISL_2306662, EPI_ISL_2306674, EPI_ISL_2306677, EPI_ISL_2306698, EPI_ISL_2306726, EPI_ISL_2306727, EPI_ISL_2306764, EPI_ISL_2306766, EPI_ISL_2306767, EPI_ISL_2306868, EPI_ISL_2306920, EPI_ISL_2306955, EPI_ISL_2306969, EPI_ISL_2306984, EPI_ISL_2398992, EPI_ISL_2398995, EPI_ISL_2398997, EPI_ISL_2398998, EPI_ISL_2399001, EPI_ISL_2399050, EPI_ISL_2399054, EPI_ISL_2399072, EPI_ISL_2399108, EPI_ISL_2399110, EPI_ISL_2399254, EPI_ISL_2399255, EPI_ISL_2399282, EPI_ISL_2399289, EPI_ISL_2399300, EPI_ISL_2399302, EPI_ISL_2399325, EPI_ISL_2399346, EPI_ISL_2399361, EPI_ISL_2399362, EPI_ISL_2480481, EPI_ISL_2480492, EPI_ISL_2480498, EPI_ISL_2480501, EPI_ISL_2480506, EPI_ISL_2480510, EPI_ISL_2480511, EPI_ISL_2480512, EPI_ISL_2480520, EPI_ISL_2480521, EPI_ISL_2480524, EPI_ISL_2611300, EPI_ISL_2611314, EPI_ISL_2611315, EPI_ISL_2611316, EPI_ISL_2611358, EPI_ISL_2872165, EPI_ISL_2872240, EPI_ISL_2926211, EPI_ISL_2926212, EPI_ISL_2926217, EPI_ISL_2926218, EPI_ISL_2926490, EPI_ISL_2926495, EPI_ISL_3016804, EPI_ISL_3111867, EPI_ISL_3111868, EPI_ISL_3111869, EPI_ISL_3111875, EPI_ISL_3111891, EPI_ISL_3111946, EPI_ISL_3328311, EPI_ISL_3330512, EPI_ISL_3330513, EPI_ISL_3330514, EPI_ISL_3330518, EPI_ISL_3330524, EPI_ISL_3330561, EPI_ISL_3330563, EPI_ISL_3330564, EPI_ISL_3330565 | | | |
| see above | Laboratory Corporation of America | Centers for Disease Control and Prevention Division of Viral Diseases, Pathogen Discovery | Adrian Paskey; Amanda Douglas; Amanda Suchanek; Andrea Throop; Ayla Burns; Ben L. Rambo-Martin; Benjamin Rambo-Martin; Bobbi Croy; Brian Krueger; Brian Norvell; Christopher Gulvick; Christos Petropoulos; Clinton R. Paden; Craig Lukasik; Dakota Howard; Darlene Wagner; Debbie Boles; Dhwani Batra; Duncan MacCannell; Eyad Almasri; Eyad Almasri Debbie Boles; Goran Stevovic; Howard Engler; Hrushikesh Deshmukh; Jake Humphrey; Jana Schroth; Jason Caravas; Joe Voshell; John Pruitt; Jonathan Meltzer; Jonathan Williams; Kara Moser; Kimberly Wagner; Lax Iyer; Lisa Pfefferle; Lyndon Tilson; Manoj Jain; Marcia Eisenberg; Mary Ann Cristobal; Mary Williamson; Matthew Robinson; Matthew Schmerer; Michael Levandoski; Mike Sapeta; Mindy Nye; Minoo Agarwal; Mohan Kolli; Nuthawin Charoensri; Oren Cohen; Peter W. Cook; Prashant Gupta; Qian Zeng; Rama Ghatti; Scott Parker; Scott Ryan; Scott Sammons; Shatavia Morrison; Stanley Letovsky; Steven Ragan; Suresh Babu Selvaraju; Susan Countryman; Susan Hicks; Suxiang Tong; Suzanne Dale; Thomas Urban; Tim Kuphal; Tricia Zwiefelhofer; Vincent Drouillon; Yvette Unoarumhi |
| EPI_ISL_1021248, EPI_ISL_1021401, EPI_ISL_1021426, EPI_ISL_1021569, EPI_ISL_1021590, EPI_ISL_1021658, EPI_ISL_1021667, EPI_ISL_1021763, EPI_ISL_1021807, EPI_ISL_1021809, EPI_ISL_1021820, EPI_ISL_1021831, EPI_ISL_1021864, EPI_ISL_1021872, EPI_ISL_1021887, EPI_ISL_1021889, EPI_ISL_1021891, EPI_ISL_1021893, EPI_ISL_1021909, EPI_ISL_1021944, EPI_ISL_1021949, EPI_ISL_1022044, EPI_ISL_1022049, EPI_ISL_1022053, EPI_ISL_1022081, EPI_ISL_1022102, EPI_ISL_1022111, EPI_ISL_1022113, EPI_ISL_1022148, EPI_ISL_1022197, EPI_ISL_1022199, EPI_ISL_1022216, EPI_ISL_1022277, EPI_ISL_1022310, EPI_ISL_1022364, EPI_ISL_1026028, EPI_ISL_1026651, EPI_ISL_1026694, EPI_ISL_1026734, EPI_ISL_1026744, EPI_ISL_1028638, EPI_ISL_1031326, EPI_ISL_1031854, EPI_ISL_1032361, EPI_ISL_1032540, EPI_ISL_1037013, EPI_ISL_1037136, EPI_ISL_1037189, EPI_ISL_1037699, EPI_ISL_1037706, EPI_ISL_1037725, EPI_ISL_1037843, EPI_ISL_1037895, EPI_ISL_1038208, EPI_ISL_1038761, EPI_ISL_1038774, EPI_ISL_1159889, EPI_ISL_1160052, EPI_ISL_1160066, EPI_ISL_1160072, EPI_ISL_1160086, EPI_ISL_1160126, EPI_ISL_1160271, EPI_ISL_1160412, EPI_ISL_1160442, EPI_ISL_1160454, EPI_ISL_1160589, EPI_ISL_1160617, EPI_ISL_1160621, EPI_ISL_1160630, EPI_ISL_1160719, EPI_ISL_1160731, EPI_ISL_1160774, EPI_ISL_1160790, EPI_ISL_1160821, EPI_ISL_1160843, EPI_ISL_1160946, EPI_ISL_1160978, EPI_ISL_1160983, EPI_ISL_1161147, EPI_ISL_1161152, EPI_ISL_1161156, EPI_ISL_1161364, EPI_ISL_1161369, EPI_ISL_1161370, EPI_ISL_1161489, EPI_ISL_1161503, EPI_ISL_1161504, EPI_ISL_1161516, EPI_ISL_1161517, EPI_ISL_1161540, EPI_ISL_1161575, EPI_ISL_1161605, EPI_ISL_1161606, EPI_ISL_1161630, EPI_ISL_1161742, EPI_ISL_1161749, EPI_ISL_1161770, EPI_ISL_1161803, EPI_ISL_1161805, EPI_ISL_1161859, EPI_ISL_1162360, EPI_ISL_1162375, EPI_ISL_1162401, EPI_ISL_1162433, EPI_ISL_1162447, EPI_ISL_1162461, EPI_ISL_1162597, EPI_ISL_1162622, EPI_ISL_1162628, EPI_ISL_1162649, EPI_ISL_1162688, EPI_ISL_1162852, EPI_ISL_1162905, EPI_ISL_1162908, EPI_ISL_1162924, EPI_ISL_1162943, EPI_ISL_1163007, EPI_ISL_1163008, EPI_ISL_1163016, EPI_ISL_1163055, EPI_ISL_1163130, EPI_ISL_1163131, EPI_ISL_1163199, EPI_ISL_1163224, EPI_ISL_1163253, EPI_ISL_1163254, EPI_ISL_1163395, EPI_ISL_1163411, EPI_ISL_1163443, EPI_ISL_1163482, EPI_ISL_1255391 | | | |
| see above | Laboratory Corporation of America | Respiratory Viruses Branch, Division of Viral Diseases, Centers for Disease Control and Prevention | Amanda Douglas; Amanda Suchanek; Andrea Throop; Ayla Burns; Ben L. Rambo-Martin; Bobbi Croy; Brian Krueger; Brian Norvell; Christos Petropoulos; Clinton R. Paden; Craig Lukasik; Dakota Howard; Dhwani Batra; Duncan MacCannell; Eyad Almasri Debbie Boles; Goran Stevovic; Howard Engler; Hrushikesh Deshmukh; Jake Humphrey; Jana Schroth; Joe Voshell; John Pruitt; Jonathan Meltzer; Jonathan Williams; Kimberly Wagner; Lax Iyer; Lyndon Tilson; Manoj Jain; Marcia Eisenberg; Mary Ann Cristobal; Mary Williamson; Michael Levandoski; Mike Sapeta; Mindy Nye; Minoo Agarwal; Mohan Kolli; Nuthawin Charoensri; Oren Cohen; Peter W. Cook; Prashant Gupta; Qian Zeng; Rama Ghatti; Scott Parker; Scott Ryan; Stanley Letovsky; Steven Ragan; Suresh Babu Selvaraju; Susan Countryman; Susan Hicks; Suxiang Tong; Suzanne Dale; Thomas Urban; Tim Kuphal; Tricia Zwiefelhofer; Vincent Drouillon |
| EPI_ISL_1273037, EPI_ISL_1273038, EPI_ISL_1273039, EPI_ISL_1273040, EPI_ISL_1273041, EPI_ISL_1273042, EPI_ISL_1318420, EPI_ISL_1318421, EPI_ISL_1318422, EPI_ISL_1318423, EPI_ISL_1447059, EPI_ISL_1528493, EPI_ISL_1528494, EPI_ISL_1528495, EPI_ISL_1528496, EPI_ISL_1678233, EPI_ISL_1678235, EPI_ISL_1678236, EPI_ISL_1678238, EPI_ISL_1678369, EPI_ISL_1678370, EPI_ISL_1678371, | | | |

| | | | |
|---|---|---|---|
| EPI_ISL_1678372, EPI_ISL_1678373, EPI_ISL_1678374, EPI_ISL_1678375, EPI_ISL_1823997, EPI_ISL_1823998, EPI_ISL_1823999, EPI_ISL_1824000, EPI_ISL_1824002, EPI_ISL_1824003, EPI_ISL_1824004, EPI_ISL_1824006, EPI_ISL_1824007, EPI_ISL_1937841, EPI_ISL_1937842, EPI_ISL_1937843, EPI_ISL_1937844, EPI_ISL_1937845, EPI_ISL_1937846, EPI_ISL_1937848, EPI_ISL_2105193, EPI_ISL_2105194, EPI_ISL_2105203, EPI_ISL_2105211, EPI_ISL_2105216, EPI_ISL_2229195, EPI_ISL_2229196, EPI_ISL_2229198, EPI_ISL_2384002, EPI_ISL_2384003, EPI_ISL_2384004, EPI_ISL_2384006, EPI_ISL_2384007, EPI_ISL_2384008, EPI_ISL_2452184, EPI_ISL_2452187, EPI_ISL_2452189, EPI_ISL_2452191, EPI_ISL_2452192, EPI_ISL_2452194, EPI_ISL_2452196, EPI_ISL_2452197, EPI_ISL_2645374, EPI_ISL_2787665, EPI_ISL_3354075, EPI_ISL_3354219, EPI_ISL_3354220, EPI_ISL_3354228, EPI_ISL_3354492, EPI_ISL_3354512, EPI_ISL_3354516 | | | |
| see above | NH Dept. of Health and Human Services Public Health Labs | Centers for Disease Control and Prevention Division of Viral Diseases, Pathogen Discovery | Alex Burgin; Alison Laufer Halpin; Anna Montmayeur; Anna Uehara; Ben L. Rambo-Martin; Ben Rambo-Martin; Clinton Paden; Clinton R. Paden; Dakota Howard; Darlene Wagner; Dave Wentworth; Dhwani Batra; Haibin Wang; Jasmine Padilla; Jing Zhang; Justin Lee; Katie Dillon; Krista Queen; Kristen Knipe; Kristine Lacek; Lori Rowe; Mark Burroughs; Matthew Schmerer; Meghan Bentz; Mili Sheth; Peter Cook; Peter W. Cook; Rachel Marine; Sam Shepard; Sarah Nobles; Shoshona Le; Suxiang Tong; Vivien Dugan; Yan Li; Ying Tao; Yvette Unoarumhi |
| EPI_ISL_1408892, EPI_ISL_1509930, EPI_ISL_1624196 | NH Dept. of Health and Human Services Public Health Labs | Genomics and Discovery, Respiratory Viruses Branch, Division of Viral Diseases, Centers for Disease Control and Prevention | Adam Retchless; Anna Kelleher; Anna Montmayeur; Anna Uehara; Brian Lynch; Clinton R. Paden; Haibin Wang; Han Jia Justin Ng; Jing Zhang; Justin Lee; Krista Queen; Mark Burroughs; Peter Cook; Rachel Marine; Suxiang Tong; Yan Li; Ying Tao |
| EPI_ISL_1759390, EPI_ISL_1759391, EPI_ISL_1759392, EPI_ISL_2042418 | NH Dept. of Health and Human Services Public Health Labs | Pathogen Discovery, Respiratory Viruses Branch, Division of Viral Diseases, Centers for Disease Control and Prevention | Adam Retchless; Anna Kelleher; Anna Montmayeur; Anna Uehara; Brian Lynch; Clinton R. Paden; Dhwani Batra; Haibin Wang; Han Jia Justin Ng; Jasmine Padilla; Jing Zhang; Justin Lee; Krista Queen; Mark Burroughs; Mili Sheth; Morgan Davis; Peter Cook; Rachel Marine; Sarah Nobles; Suxiang Tong; Tara Coalter; Yan Li; Ying Tao |
| EPI_ISL_854967, EPI_ISL_854968, EPI_ISL_854969, EPI_ISL_854970, EPI_ISL_854971, EPI_ISL_854972, EPI_ISL_855160, EPI_ISL_855161, EPI_ISL_855162, EPI_ISL_855163, EPI_ISL_855164, EPI_ISL_877196, EPI_ISL_877204, EPI_ISL_903551, EPI_ISL_906449, EPI_ISL_906450, EPI_ISL_906451, EPI_ISL_906452, EPI_ISL_906453, EPI_ISL_915333, EPI_ISL_915334 | | | |
| see above | Quest Diagnostics | Quest Diagnostics | Anderson, B.; Bernstein; D.F.; Gerasimova, A.; Hua, M.; I.A.; K.E.; Kagan; L.E.; Lacbawan, F.; Liu Y.; Livingston; Owen, R.; Perez, A.; R.M.; Rosenthal; S.H.; Shalhout; Shlyakhter; Tanpaiboon, P. |
| EPI_ISL_1193508, EPI_ISL_1193516, EPI_ISL_1193540, EPI_ISL_1193554, EPI_ISL_1193563, EPI_ISL_1193602, EPI_ISL_1193615, EPI_ISL_1193633, EPI_ISL_1193634, EPI_ISL_1193660, EPI_ISL_1193788, EPI_ISL_1193816, EPI_ISL_1193867, EPI_ISL_1193894, EPI_ISL_1193941, EPI_ISL_1193987, EPI_ISL_1193993, EPI_ISL_1194001, EPI_ISL_1194190, EPI_ISL_1194208, EPI_ISL_1194235, EPI_ISL_1194366, EPI_ISL_1194373, EPI_ISL_1194384, EPI_ISL_1220580, EPI_ISL_1252925, EPI_ISL_1252931, EPI_ISL_1252940, EPI_ISL_1252942, EPI_ISL_1252951, EPI_ISL_1252956, EPI_ISL_1252960, EPI_ISL_1252961, EPI_ISL_1252990, EPI_ISL_1253320, EPI_ISL_1253372, EPI_ISL_1253412, EPI_ISL_1253414, EPI_ISL_1253430, EPI_ISL_1253470, EPI_ISL_1253493, EPI_ISL_1253511, EPI_ISL_1254085, EPI_ISL_1254337, EPI_ISL_1254341, EPI_ISL_1254382, EPI_ISL_1254391, EPI_ISL_1254394, EPI_ISL_1266189, EPI_ISL_1266872, EPI_ISL_1266939, EPI_ISL_1267303, EPI_ISL_1267313, EPI_ISL_1267376, EPI_ISL_1291119, EPI_ISL_1291469, EPI_ISL_1291565, EPI_ISL_1291570, EPI_ISL_1297610, EPI_ISL_1297671, EPI_ISL_1297672, EPI_ISL_1297682, EPI_ISL_1297690, EPI_ISL_1297693, EPI_ISL_1297695, EPI_ISL_1297697, EPI_ISL_1297699, EPI_ISL_1297702, EPI_ISL_1297703, EPI_ISL_1297733, EPI_ISL_1297749, EPI_ISL_1297752, EPI_ISL_1297761, EPI_ISL_1298230, EPI_ISL_1314579, EPI_ISL_1314581, EPI_ISL_1314648, EPI_ISL_1314754, EPI_ISL_1314756, EPI_ISL_1314792, EPI_ISL_1314806, EPI_ISL_1314852, EPI_ISL_1314878, EPI_ISL_1314942, EPI_ISL_1335988, EPI_ISL_1335990, EPI_ISL_1335999, EPI_ISL_1336059, EPI_ISL_1336060, EPI_ISL_1336072, EPI_ISL_1336076, EPI_ISL_1336078, EPI_ISL_1339955, EPI_ISL_1339960, EPI_ISL_1340597, EPI_ISL_1340720, EPI_ISL_1340730, EPI_ISL_1367173, EPI_ISL_1367179, EPI_ISL_1367183, EPI_ISL_1367189, EPI_ISL_1367194, EPI_ISL_1367210, EPI_ISL_1367224, EPI_ISL_1367231, EPI_ISL_1367236, EPI_ISL_1367248, EPI_ISL_1367332, EPI_ISL_1367341, EPI_ISL_1367342, EPI_ISL_1367363, EPI_ISL_1367365, EPI_ISL_1367368, EPI_ISL_1367371, EPI_ISL_1367377, EPI_ISL_1367384, EPI_ISL_1367392, EPI_ISL_1367399, EPI_ISL_1367401, EPI_ISL_1367413, EPI_ISL_1391443, EPI_ISL_1391455, EPI_ISL_1391456, EPI_ISL_1391458, EPI_ISL_1391669, EPI_ISL_1391678, EPI_ISL_1391691, EPI_ISL_1392374, EPI_ISL_1422545, EPI_ISL_1422650, EPI_ISL_1422700, EPI_ISL_1422730, EPI_ISL_1422739, EPI_ISL_1422769, EPI_ISL_1422779, EPI_ISL_1422966, EPI_ISL_1422985, EPI_ISL_1444501, EPI_ISL_1444505, EPI_ISL_1444507, EPI_ISL_1444508, EPI_ISL_1444513, EPI_ISL_1444515, EPI_ISL_1444516, EPI_ISL_1444536, EPI_ISL_1444537, EPI_ISL_1444540, EPI_ISL_1479744, EPI_ISL_1479747, EPI_ISL_1479753, EPI_ISL_1479756, EPI_ISL_1479761, EPI_ISL_1479773, EPI_ISL_1479794, EPI_ISL_1479917, EPI_ISL_1479923, EPI_ISL_1479958, EPI_ISL_1480533, EPI_ISL_1480718, EPI_ISL_1480788, EPI_ISL_1480801, EPI_ISL_1480808, EPI_ISL_1480882, EPI_ISL_1480884, EPI_ISL_1480886, EPI_ISL_1480891, EPI_ISL_1480901, EPI_ISL_1480903, EPI_ISL_1480911, EPI_ISL_1480912, EPI_ISL_1493614, EPI_ISL_1493973, EPI_ISL_1494009, EPI_ISL_1551742, EPI_ISL_1551872, EPI_ISL_1552158, EPI_ISL_1552160, EPI_ISL_1552180, EPI_ISL_1552192, EPI_ISL_1552193, EPI_ISL_1552196, EPI_ISL_1552204, EPI_ISL_1552210, EPI_ISL_1552216, EPI_ISL_1552324, EPI_ISL_1552344, EPI_ISL_1552350, EPI_ISL_1552367, EPI_ISL_1552375, EPI_ISL_1552376, EPI_ISL_1552398, EPI_ISL_1552406, EPI_ISL_1648884, EPI_ISL_1668477, EPI_ISL_1738081, EPI_ISL_1738282, EPI_ISL_1753136, EPI_ISL_1798007, EPI_ISL_1818145, EPI_ISL_1818171, EPI_ISL_1818174, EPI_ISL_1818177, EPI_ISL_1818189, EPI_ISL_1818192, EPI_ISL_1818213, EPI_ISL_1818230, EPI_ISL_1818231, EPI_ISL_1818243, EPI_ISL_1818257, EPI_ISL_1818271, EPI_ISL_1818275, EPI_ISL_1818280, EPI_ISL_1818338, EPI_ISL_1818354, EPI_ISL_1834198, EPI_ISL_1834332, EPI_ISL_1834358, EPI_ISL_1839009, EPI_ISL_1839037, EPI_ISL_1840461, EPI_ISL_1840467, EPI_ISL_1840511, EPI_ISL_1900054, EPI_ISL_1900425, EPI_ISL_1900456, EPI_ISL_1906192, EPI_ISL_1906227, EPI_ISL_1906233, EPI_ISL_1906250, EPI_ISL_1906264, EPI_ISL_1906288, EPI_ISL_1906310, EPI_ISL_1906314, EPI_ISL_1906323, EPI_ISL_1906338, EPI_ISL_1906343, EPI_ISL_1906356, EPI_ISL_1906366, EPI_ISL_1906368, EPI_ISL_1906385, EPI_ISL_1906392, EPI_ISL_1906397, EPI_ISL_1906404, EPI_ISL_1906413, EPI_ISL_1906431, EPI_ISL_1906435, EPI_ISL_1906458, EPI_ISL_1906518, EPI_ISL_1906545, EPI_ISL_1924279, EPI_ISL_1924283, EPI_ISL_1924292, EPI_ISL_1924294, EPI_ISL_1924305, EPI_ISL_1924307, EPI_ISL_1924333, EPI_ISL_1924344, EPI_ISL_1924351, EPI_ISL_1924360, EPI_ISL_1924368, EPI_ISL_1924369, EPI_ISL_1924372, EPI_ISL_1924380, EPI_ISL_1924386, EPI_ISL_1924387, EPI_ISL_1924388, EPI_ISL_1924389, EPI_ISL_1924396, EPI_ISL_1924418, EPI_ISL_1924419, EPI_ISL_1924428, EPI_ISL_1924556, EPI_ISL_1924563, EPI_ISL_1935567, EPI_ISL_1935568, EPI_ISL_1990414, EPI_ISL_1990415, EPI_ISL_1990427, EPI_ISL_1990434, EPI_ISL_1990443, EPI_ISL_1990471, EPI_ISL_1990473, EPI_ISL_1990502, EPI_ISL_1990507, EPI_ISL_2011036, EPI_ISL_2011041, EPI_ISL_2011072, EPI_ISL_2011076, EPI_ISL_2011078, EPI_ISL_2011098, EPI_ISL_2011123, EPI_ISL_2011125, EPI_ISL_2011196, EPI_ISL_2011204, EPI_ISL_2011234, EPI_ISL_2035325, EPI_ISL_2035326, EPI_ISL_2047085, EPI_ISL_2133095, EPI_ISL_2133104, EPI_ISL_2133119, EPI_ISL_2133129, EPI_ISL_2133145, EPI_ISL_2133186, EPI_ISL_2133190, EPI_ISL_2133191, EPI_ISL_2133211, EPI_ISL_2133216, EPI_ISL_2133221, EPI_ISL_2133223, EPI_ISL_2133228, EPI_ISL_2133391, EPI_ISL_2133408, EPI_ISL_2133424, EPI_ISL_2133443, EPI_ISL_2133454, EPI_ISL_2133459, EPI_ISL_2133481, EPI_ISL_2133516, EPI_ISL_2133521, EPI_ISL_2133529, EPI_ISL_2133791, EPI_ISL_2140732, EPI_ISL_2140735, EPI_ISL_2140740, EPI_ISL_2140741, EPI_ISL_2140744, EPI_ISL_2140749, EPI_ISL_2140766, EPI_ISL_2142882, EPI_ISL_2142887, EPI_ISL_2142891, EPI_ISL_2142906, EPI_ISL_2142908, EPI_ISL_2142928, EPI_ISL_2142935, EPI_ISL_2142947, EPI_ISL_2142953, EPI_ISL_2142963, EPI_ISL_2142987, EPI_ISL_2143045, EPI_ISL_2143051, EPI_ISL_2143064, EPI_ISL_2143067, EPI_ISL_2143080, EPI_ISL_2143090, EPI_ISL_2143121, EPI_ISL_2143127, EPI_ISL_2143143, EPI_ISL_2143299, EPI_ISL_2143382, EPI_ISL_2143405, EPI_ISL_2143772, EPI_ISL_2143862, EPI_ISL_2143875, EPI_ISL_2143929, EPI_ISL_2143986, EPI_ISL_2143988, EPI_ISL_2179923, EPI_ISL_2179945, EPI_ISL_2180518, EPI_ISL_2187247, EPI_ISL_2187271, EPI_ISL_2187292, EPI_ISL_2187304, EPI_ISL_2187311, EPI_ISL_2268398, EPI_ISL_2268422, EPI_ISL_2268438, EPI_ISL_2268448, EPI_ISL_2268458, EPI_ISL_2268482, EPI_ISL_2268515, EPI_ISL_2268553, EPI_ISL_2268709, EPI_ISL_2268741, EPI_ISL_2268743, EPI_ISL_2268744, EPI_ISL_2268773, EPI_ISL_2268777, EPI_ISL_2268798, EPI_ISL_2268808, EPI_ISL_2268822, EPI_ISL_2268881, EPI_ISL_2268895, EPI_ISL_2268898, EPI_ISL_2268933, EPI_ISL_2269080, EPI_ISL_2269094, EPI_ISL_2269104, EPI_ISL_2269112, EPI_ISL_2269132, EPI_ISL_2269138, EPI_ISL_2269144, EPI_ISL_2269166, EPI_ISL_2269190, EPI_ISL_2269209, EPI_ISL_2280085, EPI_ISL_2280116, EPI_ISL_2280125, EPI_ISL_2280162, EPI_ISL_2367952, EPI_ISL_2367953, EPI_ISL_2367963, EPI_ISL_2367982, EPI_ISL_2368035, EPI_ISL_2368057, EPI_ISL_2368064, EPI_ISL_2368066, EPI_ISL_2371735, EPI_ISL_2371750, EPI_ISL_2371777, EPI_ISL_2371867, EPI_ISL_2371956, EPI_ISL_2371975, EPI_ISL_2372014, EPI_ISL_2372028, EPI_ISL_2372033, EPI_ISL_2372036, EPI_ISL_2372080, EPI_ISL_2372095, EPI_ISL_2372097, EPI_ISL_2372099, EPI_ISL_2372160, EPI_ISL_2372191, EPI_ISL_2372209, EPI_ISL_2372247, EPI_ISL_2372942, EPI_ISL_2397361, EPI_ISL_2397370, EPI_ISL_2397373, EPI_ISL_2397408, EPI_ISL_2397559, EPI_ISL_2397611, EPI_ISL_2397616, EPI_ISL_2397675, EPI_ISL_2397681, EPI_ISL_2397722, EPI_ISL_2397734, EPI_ISL_2397757, EPI_ISL_2399513, EPI_ISL_2399515, EPI_ISL_2439607, EPI_ISL_2439616, EPI_ISL_2439693, EPI_ISL_2439851, EPI_ISL_2440555, EPI_ISL_2440620, EPI_ISL_2440684, EPI_ISL_2440690, EPI_ISL_2440693, EPI_ISL_2440701, EPI_ISL_2440703, EPI_ISL_2440755, EPI_ISL_2440834, EPI_ISL_2440869, EPI_ISL_2451522, EPI_ISL_2451548, EPI_ISL_2463029, EPI_ISL_2463122, EPI_ISL_2488825, EPI_ISL_2488828, EPI_ISL_2527012, EPI_ISL_2529161, EPI_ISL_2529329, EPI_ISL_2534521, EPI_ISL_2534558, EPI_ISL_2534560, EPI_ISL_2534561, EPI_ISL_2599720, EPI_ISL_2611863, EPI_ISL_2652275, EPI_ISL_2686363, EPI_ISL_2868692, EPI_ISL_2869179, EPI_ISL_2869193, EPI_ISL_2870994, EPI_ISL_2871454, EPI_ISL_2871635, EPI_ISL_2874524, EPI_ISL_2876747, EPI_ISL_3113992, EPI_ISL_3114092, EPI_ISL_3114431, EPI_ISL_3331026 | | | |
| see above | Quest Diagnostics Incorporated | Centers for Disease Control and Prevention Division of Viral Diseases, Pathogen Discovery | A. Gerasimova; A. Perez; Adrian Paskey; B. Anderson; Ben L. Rambo-Martin; Benjamin Rambo-Martin; Christopher Gulvick; Clinton R. Paden; Dakota Howard; Darlene Wagner; Dhwani Batra; Duncan MacCannell; F. Lacbawan; I. A. Shlyakhter; Jason Caravas; K.E. Livingston; Kara Moser; L.E. Bernstein; M. Hua; Matthew Schmerer; P. Tanpaiboon; Peter W. Cook; R. M. Kagan; R. Owen; R. V. Rolando; S. H. Rosenthal; Scott Sammons; Shatavia Morrison; Suxiang Tong; Y. Liu; Yvette Unoarumhi |
| EPI_ISL_1086603, EPI_ISL_1086823, EPI_ISL_1086824, EPI_ISL_1086834, EPI_ISL_1086855, EPI_ISL_1086871, EPI_ISL_1086915, EPI_ISL_1086918, EPI_ISL_1086925, EPI_ISL_1086934, EPI_ISL_1087007, EPI_ISL_1087027, EPI_ISL_1087030, EPI_ISL_1087036, EPI_ISL_1087038, EPI_ISL_1087047, EPI_ISL_1087079, EPI_ISL_1087097, EPI_ISL_1087110, EPI_ISL_1087114, EPI_ISL_1087121, EPI_ISL_1087124, EPI_ISL_1087238, EPI_ISL_1087442, EPI_ISL_1087447, EPI_ISL_1087450, EPI_ISL_1087460, EPI_ISL_1087505, EPI_ISL_1087566, EPI_ISL_1087577, EPI_ISL_1087584, EPI_ISL_1087587, EPI_ISL_1087600, EPI_ISL_1087601, EPI_ISL_1087615, EPI_ISL_1087619, EPI_ISL_1087635, EPI_ISL_1087638, EPI_ISL_1087645, EPI_ISL_1087649, EPI_ISL_1087662, EPI_ISL_1087668, EPI_ISL_1087670, EPI_ISL_1087671, EPI_ISL_1087676, EPI_ISL_1087726, EPI_ISL_1087742, EPI_ISL_1087749, EPI_ISL_1087754, EPI_ISL_1087757, EPI_ISL_1087802, EPI_ISL_1087806, EPI_ISL_1087807, EPI_ISL_1088200, EPI_ISL_1090339, EPI_ISL_1090550, EPI_ISL_1090551, EPI_ISL_1090552, EPI_ISL_1090583, EPI_ISL_1090604, EPI_ISL_1090611, EPI_ISL_1090622, EPI_ISL_1090642, EPI_ISL_1090669, EPI_ISL_1090691, EPI_ISL_1090860, EPI_ISL_1090882, EPI_ISL_1090889, EPI_ISL_1090903, EPI_ISL_1090904, EPI_ISL_1090908, EPI_ISL_1090910, EPI_ISL_1090922, EPI_ISL_1090931, EPI_ISL_1090936, EPI_ISL_1090939, EPI_ISL_1090942, EPI_ISL_1113801, EPI_ISL_1113802, EPI_ISL_1113901, EPI_ISL_1113936, EPI_ISL_1113940, EPI_ISL_1113953, EPI_ISL_1113954, EPI_ISL_1113987, EPI_ISL_1113990, EPI_ISL_1114010, EPI_ISL_1114038, EPI_ISL_1139285, EPI_ISL_1139294, EPI_ISL_1139347, EPI_ISL_1139684, EPI_ISL_1139688, EPI_ISL_1139691, EPI_ISL_1139769, EPI_ISL_1159575, EPI_ISL_1159679 | | | |
| see above | Quest Diagnostics Incorporated | Respiratory Viruses Branch, Division of Viral Diseases, Centers for Disease Control and Prevention | A. Gerasimova; A. Perez; B. Anderson; Ben L. Rambo-Martin; Clinton R. Paden; Dakota Howard; Dhwani Batra; Duncan MacCannell; F. Lacbawan; I. A. Shlyakhter; K.E. Livingston; L.E. Bernstein; M. Hua; P. Tanpaiboon; Peter W. Cook; R. M. Kagan; R. Owen; R. V. Rolando; S. H. Rosenthal; Suxiang Tong; Y. Liu |
| EPI_ISL_884686, EPI_ISL_2254366, EPI_ISL_3091678, EPI_ISL_3091796, EPI_ISL_3091807, EPI_ISL_3091838, EPI_ISL_3091909, EPI_ISL_3092005, EPI_ISL_3092032 | | | |
| see above | Respiratory Viruses Branch, Centers for Disease Control and Prevention | Respiratory Viruses Branch, Centers for Disease Control and Prevention | Allen, E.; Anderson, B.; Antico, J.; Ascencio, A.; B.L.; Barret; Batra, D.; Becker, D.; Bernstein, L.; Bolze, A.; C.R.; Campbell, P.; Caravas, J.; Case, R.; Cassens, T.; Cho, R.; Cirulli, E.; Clark, C.; Cook; Febbo, P.; Galloway, S.; Gerasimova, A.; Ghorpade, V.; Gietzen, K.; Gulvick, C.; Hardison, M.; Houdeshell, H.; Howard, D.; Hua, M.; I.A.; Isaksson, M.; K.S.; Kagan; Kvalvaag, O.; Lacbawan, F.; Laurent, M.; Lee, W.; Levan, G.; Liu, J.; Liu, Y.; Livingston, K.; Lu, J.; MacCannell, D.; Morrison, S.; Moser, K.; N.L.; Nall, D.; Nguyen, J.; Owen, R.; P.W.; Paden; Paskey, A.; Perez, A.; R.M.; R.V.; Rambo-Martin; Rambo-Martin, B.; Ramirez, J.; Rivera-Garcia, C.; Rolando; Rosenthal; S.H.; Sammons, S.; Sanders, E.; Sandoval, E.; Schmerer, M.; Shlyakhter; Sickler, B.; Tanpaiboon, P.; Tolentino, M.; Tong, S.; Tran, C.; Unoarumhi, Y.; Vest, A.; Wagner, D.; Wang, S.; Washington; Westlund, S.; White, S.; Wickline, S.; de Feo, E. |
| EPI_ISL_1739434, EPI_ISL_1970658 | The Jackson Laboratory | The Jackson Laboratory | Adams M; Bergeron D; Kelly K; Li L; Long J; Omerza G; Renzette N |
| EPI_ISL_2283926, EPI_ISL_2284000, EPI_ISL_2284002, EPI_ISL_2284010, EPI_ISL_2284011, EPI_ISL_2284014, EPI_ISL_2284020, EPI_ISL_2284022, EPI_ISL_2284040, EPI_ISL_2284049, EPI_ISL_2284052, EPI_ISL_2284053, EPI_ISL_2284194 | | | |
| see above | VA Connecticut Healthcare System | Yale Center for Genomic Analysis | Brooke Sullivan; Curt Scharfe; Irina Tikhonova; Kaya Bilguvar; Shrikant Mane |
| EPI_ISL_2034860 | Yale Clinical Virology Lab | Yale Center for Genomic Analysis | Brooke Sullivan; Curt Scharfe; Irina Tikhonova; Kaya Bilguvar; Shrikant Mane |
