## Supplementary material for "Comparative transmissibility of SARS-CoV-2 variants Delta and Alpha in New England, USA": DataS1-7: DataS7_Vermont.pdf

We gratefully acknowledge the following Authors from the Originating laboratories responsible for obtaining the specimens, as well as the Submitting laboratories where the genome data were generated and shared via GISAID, on which this research is based.

All Submitters of data may be contacted directly via www.gisaid.org

Authors are sorted alphabetically.

| Accession ID | Originating Laboratory | Submitting Laboratory | Authors |
|---|---|---|---|
| EPI_ISL_1445581, EPI_ISL_1445734, EPI_ISL_1445753, EPI_ISL_1446129, EPI_ISL_1446175, EPI_ISL_1479480, EPI_ISL_1479526, EPI_ISL_1479553, EPI_ISL_1493506, EPI_ISL_1550813, EPI_ISL_1559915, EPI_ISL_1560678, EPI_ISL_1560874, EPI_ISL_1563275, EPI_ISL_1563311, EPI_ISL_1650851, EPI_ISL_1650983, EPI_ISL_1667799, EPI_ISL_1667800, EPI_ISL_1685468, EPI_ISL_1685469, EPI_ISL_1685507, EPI_ISL_1685870, EPI_ISL_1686003, EPI_ISL_1686717, EPI_ISL_1686852, EPI_ISL_1688360, EPI_ISL_1688363, EPI_ISL_1701951, EPI_ISL_1702052, EPI_ISL_1736856, EPI_ISL_1834872, EPI_ISL_1834883, EPI_ISL_1834973, EPI_ISL_1835275, EPI_ISL_1835786, EPI_ISL_1900233, EPI_ISL_1995987, EPI_ISL_1997016, EPI_ISL_1997459, EPI_ISL_1998632, EPI_ISL_1999603, EPI_ISL_2035172, EPI_ISL_2038985, EPI_ISL_2039406, EPI_ISL_2040287, EPI_ISL_2041519, EPI_ISL_2147058, EPI_ISL_2147655, EPI_ISL_2147656, EPI_ISL_2148030, EPI_ISL_2148507, EPI_ISL_2149610, EPI_ISL_2149929, EPI_ISL_2185828, EPI_ISL_2186127, EPI_ISL_2187438, EPI_ISL_2187535, EPI_ISL_2243824, EPI_ISL_2244168, EPI_ISL_2244644, EPI_ISL_2321308, EPI_ISL_2421413, EPI_ISL_2421437, EPI_ISL_2421583, EPI_ISL_2421766, EPI_ISL_2600171 | | | |
| see above | Aegis Sciences Corporation | Centers for Disease Control and Prevention Division of Viral Diseases, Pathogen Discovery | Adrian Paskey; Alec Vest; Benjamin Rambo-Martin; Christopher Gulvick; Clinton R. Paden; Cyndi Clark; Dakota Howard; Darlene Wagner; Dhwani Batra; Dillon Nall; Duncan MacCannell; Ethan Sanders; Holly Houdeshell; Jason Caravas; Kara Moser; Matthew Hardison; Matthew Schmerer; Ola Kvalvaag; Patrick Campbell; Peter W. Cook; Rob Case; Scott Sammons; Shatavia Morrison; Shaun Westlund; Vikramsinha Ghorpade; Yvette Unoarumhi |
| EPI_ISL_2991966, EPI_ISL_2991968, EPI_ISL_2991969, EPI_ISL_2991971, EPI_ISL_2991973, EPI_ISL_2991974, EPI_ISL_2991976, EPI_ISL_2991977, EPI_ISL_2991978, EPI_ISL_2991979, EPI_ISL_2991980, EPI_ISL_2991981, EPI_ISL_2991983, EPI_ISL_2991984, EPI_ISL_2991985, EPI_ISL_2991986, EPI_ISL_2991987, EPI_ISL_2991988, EPI_ISL_2991989, EPI_ISL_2991990, EPI_ISL_2991991, EPI_ISL_2991992, EPI_ISL_2991993, EPI_ISL_2991994 | | | |
| see above | Broad Institute Clinical Research Sequencing Platform | Infectious Disease Program, Broad Institute of Harvard and MIT | A.E.; Adams, G.; Anahtar, M.; B.L.; B.W.; Bauer, M.; Birren; Branda, J.; Carter, A.; Cerrato, F.; Chaluvadi, S.; Chapman; Cusick, C.; D.J.; DeRuff, K.; Flowers, K.; Gallagher, G.; Gladden-Young, A.; Gnirke, A.; Harris, J.; J.E.; K.J.; LaRocque, R.; Lagerborg, K.; Lemieux; Lin; Loreth, C.; MacInnis; Neumann, A.; Normandin, E.; P.C.; Park; Pierce, V.; Reilly, S.; Rosenberg, E.; Rudy, M.; Ryan, E.; S.B.; Sabeti; Shaw, B.; Siddle; Slater, D.; Smole, S.; Tomkins-Tinch, C.; Turbett, S. |
| EPI_ISL_1413171, EPI_ISL_1413173, EPI_ISL_1413181, EPI_ISL_1413184, EPI_ISL_1413187, EPI_ISL_1413191, EPI_ISL_1413196, EPI_ISL_1413199, EPI_ISL_1413202, EPI_ISL_1413204, EPI_ISL_1413207, EPI_ISL_1413257, EPI_ISL_1413266, EPI_ISL_1413272, EPI_ISL_1413277, EPI_ISL_1413280, EPI_ISL_1413282, EPI_ISL_1413287, EPI_ISL_1413290, EPI_ISL_1413293, EPI_ISL_1413296, EPI_ISL_1413304, EPI_ISL_1516464, EPI_ISL_1516465, EPI_ISL_1516466, EPI_ISL_1516467, EPI_ISL_1516468, EPI_ISL_1516469, EPI_ISL_1516470, EPI_ISL_1516534, EPI_ISL_1516547, EPI_ISL_1516548, EPI_ISL_1516549, EPI_ISL_1516550, EPI_ISL_1516551, EPI_ISL_1516552, EPI_ISL_1516553, EPI_ISL_1516555, EPI_ISL_1516556, EPI_ISL_1516557, EPI_ISL_1516558, EPI_ISL_1516559, EPI_ISL_1516560, EPI_ISL_1516561, EPI_ISL_1578159, EPI_ISL_1578182, EPI_ISL_1578183, EPI_ISL_1578186, EPI_ISL_1578194, EPI_ISL_1578195, EPI_ISL_1578269, EPI_ISL_1578270, EPI_ISL_1578271, EPI_ISL_1578273, EPI_ISL_1578276, EPI_ISL_1578281, EPI_ISL_1578315, EPI_ISL_1578317, EPI_ISL_1578332, EPI_ISL_1578402, EPI_ISL_1578433, EPI_ISL_1710052, EPI_ISL_1710056, EPI_ISL_1710082, EPI_ISL_1710083, EPI_ISL_1710084, EPI_ISL_1710088, EPI_ISL_1710089, EPI_ISL_1710090, EPI_ISL_1710091, EPI_ISL_1710106, EPI_ISL_1710110, EPI_ISL_1710111, EPI_ISL_1710112, EPI_ISL_1710113, EPI_ISL_1710116, EPI_ISL_1710117, EPI_ISL_1710118, EPI_ISL_1710119, EPI_ISL_1710120, EPI_ISL_1710361, EPI_ISL_1710366, EPI_ISL_1710371, EPI_ISL_1757004, EPI_ISL_1757023, EPI_ISL_1757055, EPI_ISL_1757056, EPI_ISL_1757057, EPI_ISL_1757058, EPI_ISL_1757059, EPI_ISL_1757060, EPI_ISL_1757061, EPI_ISL_1757062, EPI_ISL_1757253, EPI_ISL_1757254, EPI_ISL_1757255, EPI_ISL_1757256, EPI_ISL_1757257, EPI_ISL_1757287, EPI_ISL_1787276, EPI_ISL_1787277, EPI_ISL_1787278, EPI_ISL_1787279, EPI_ISL_1787280, EPI_ISL_1787281, EPI_ISL_1787282, EPI_ISL_1787283, EPI_ISL_1787284, EPI_ISL_1787285, EPI_ISL_1787286, EPI_ISL_1787287, EPI_ISL_1787288, EPI_ISL_1787289, EPI_ISL_1787290, EPI_ISL_1787291, EPI_ISL_1787292, EPI_ISL_1787293, EPI_ISL_1787294, EPI_ISL_1787295, EPI_ISL_1787296, EPI_ISL_1787297, EPI_ISL_1787298, EPI_ISL_1787299, EPI_ISL_1787300, EPI_ISL_1787301, EPI_ISL_1787302, EPI_ISL_1787303, EPI_ISL_1787304, EPI_ISL_1787306, EPI_ISL_1787307, EPI_ISL_1787308, EPI_ISL_1787309, EPI_ISL_1787310, EPI_ISL_1787311, EPI_ISL_1787312, EPI_ISL_1787313, EPI_ISL_1787314, EPI_ISL_1787315, EPI_ISL_1787316, EPI_ISL_1825237, EPI_ISL_1825263, EPI_ISL_1825268, EPI_ISL_1825269, EPI_ISL_1825270, EPI_ISL_1825271, EPI_ISL_1825275, EPI_ISL_1825276, EPI_ISL_1825277, EPI_ISL_1825282, EPI_ISL_1825283, EPI_ISL_1825284, EPI_ISL_1825285, EPI_ISL_1825286, EPI_ISL_1825287, EPI_ISL_1825295, EPI_ISL_1825314, EPI_ISL_1825474, EPI_ISL_1825475, EPI_ISL_1825476, EPI_ISL_1825478, EPI_ISL_1825485, EPI_ISL_1825486, EPI_ISL_1825491, EPI_ISL_1825495, EPI_ISL_1825496, EPI_ISL_1825498, EPI_ISL_1825505, EPI_ISL_1825506, EPI_ISL_1825511, EPI_ISL_1825512, EPI_ISL_1825514, EPI_ISL_1825519, EPI_ISL_1825528, EPI_ISL_1825540, EPI_ISL_1825547, EPI_ISL_1825564, EPI_ISL_1825571, EPI_ISL_1825572, EPI_ISL_1825573, EPI_ISL_1825574, EPI_ISL_1825578, EPI_ISL_1825580, EPI_ISL_1825584, EPI_ISL_1918984, EPI_ISL_1918985, EPI_ISL_1918986, EPI_ISL_1918987, EPI_ISL_1918988, EPI_ISL_1918989, EPI_ISL_1918990, EPI_ISL_1919250, EPI_ISL_1919251, EPI_ISL_1919252, EPI_ISL_1919253, EPI_ISL_1919254, EPI_ISL_1919255, EPI_ISL_1919256, EPI_ISL_1919257, EPI_ISL_1919258, EPI_ISL_1919259, EPI_ISL_1919260, EPI_ISL_1919261, EPI_ISL_1919262, EPI_ISL_1919263, EPI_ISL_1919264, EPI_ISL_1919265, EPI_ISL_1919266, EPI_ISL_1919267, EPI_ISL_1919268, EPI_ISL_1919269, EPI_ISL_1919270, EPI_ISL_1919271, EPI_ISL_1968524, EPI_ISL_1968525, EPI_ISL_1968526, EPI_ISL_1968527, EPI_ISL_1968528, EPI_ISL_1968529, EPI_ISL_1968530, EPI_ISL_1968531, EPI_ISL_1968532, EPI_ISL_1968534, EPI_ISL_1968535, EPI_ISL_1968536, EPI_ISL_1968537, EPI_ISL_1968538, EPI_ISL_1968539, EPI_ISL_1968540, EPI_ISL_1968541, EPI_ISL_1968542, EPI_ISL_1968543, EPI_ISL_1968544, EPI_ISL_1968545, EPI_ISL_1968546, EPI_ISL_1968547, EPI_ISL_1968548, EPI_ISL_1968549, EPI_ISL_1968550, EPI_ISL_1968551, EPI_ISL_1968552, EPI_ISL_1968553, EPI_ISL_1968554, EPI_ISL_1968555, EPI_ISL_1968556, EPI_ISL_1968557, EPI_ISL_1968559, EPI_ISL_1968560, EPI_ISL_1972039, EPI_ISL_1972040, EPI_ISL_1972041, EPI_ISL_1972042, EPI_ISL_1972043, EPI_ISL_1972045, EPI_ISL_1972046, EPI_ISL_1972047, EPI_ISL_1972048, EPI_ISL_1972049, EPI_ISL_1972051, EPI_ISL_1972052, EPI_ISL_1972053, EPI_ISL_1972054, EPI_ISL_1972055, EPI_ISL_1972056, EPI_ISL_1972057, EPI_ISL_1972058, EPI_ISL_1972059, EPI_ISL_1972060, EPI_ISL_1972061, EPI_ISL_1972062, EPI_ISL_1972063, EPI_ISL_1972064, EPI_ISL_1972065, EPI_ISL_1972066, EPI_ISL_1972067, EPI_ISL_1972068, EPI_ISL_1972069, EPI_ISL_1972070, EPI_ISL_1972071, EPI_ISL_1972072, EPI_ISL_1972073, EPI_ISL_1972074, EPI_ISL_1972075, EPI_ISL_1972076, EPI_ISL_1972077, EPI_ISL_1972079, EPI_ISL_1972080, EPI_ISL_1972081, EPI_ISL_1972082, EPI_ISL_1972083, EPI_ISL_1972084, EPI_ISL_1972085, EPI_ISL_1972087, EPI_ISL_1972088, EPI_ISL_1972089, EPI_ISL_1972090, EPI_ISL_1972091, EPI_ISL_1972092, EPI_ISL_1972093, EPI_ISL_1972094, EPI_ISL_1972095, EPI_ISL_1972096, EPI_ISL_1972097, EPI_ISL_1972098, EPI_ISL_1972099, EPI_ISL_1972100, EPI_ISL_1972101, EPI_ISL_2096732, EPI_ISL_2096734, EPI_ISL_2096735, EPI_ISL_2096736, EPI_ISL_2096737, EPI_ISL_2096738, EPI_ISL_2096739, EPI_ISL_2096740, EPI_ISL_2096741, EPI_ISL_2096742, EPI_ISL_2096743, EPI_ISL_2096744, EPI_ISL_2096745, EPI_ISL_2096746, EPI_ISL_2096747, EPI_ISL_2096748, EPI_ISL_2096750, EPI_ISL_2096751, EPI_ISL_2096753, EPI_ISL_2096754, EPI_ISL_2096755, EPI_ISL_2096756, EPI_ISL_2096757, EPI_ISL_2096758, EPI_ISL_2096759, EPI_ISL_2096760, EPI_ISL_2096761, EPI_ISL_2096763, EPI_ISL_2096764, EPI_ISL_2096766, EPI_ISL_2096767, EPI_ISL_2096768, EPI_ISL_2096769, EPI_ISL_2096770, EPI_ISL_2096771, EPI_ISL_2096772, EPI_ISL_2096773, EPI_ISL_2096774, EPI_ISL_2104339, EPI_ISL_2104340, EPI_ISL_2104341, EPI_ISL_2104342, EPI_ISL_2104343, EPI_ISL_2104344, EPI_ISL_2104345, EPI_ISL_2104346, EPI_ISL_2104347, EPI_ISL_2104350, EPI_ISL_2104351, EPI_ISL_2104353, EPI_ISL_2104354, EPI_ISL_2104355, EPI_ISL_2104357, EPI_ISL_2104358, EPI_ISL_2104359, EPI_ISL_2104360, EPI_ISL_2104361, EPI_ISL_2104363, EPI_ISL_2104365, EPI_ISL_2104366, EPI_ISL_2104368, EPI_ISL_2104369, EPI_ISL_2104371, EPI_ISL_2104372, EPI_ISL_2104373, EPI_ISL_2104374, EPI_ISL_2104376, EPI_ISL_2104377, EPI_ISL_2104378, EPI_ISL_2104379, EPI_ISL_2104380, EPI_ISL_2104381, EPI_ISL_2104382, EPI_ISL_2104383, EPI_ISL_2104385, EPI_ISL_2104386, EPI_ISL_2104387, EPI_ISL_2104388, EPI_ISL_2104390, EPI_ISL_2104392, EPI_ISL_2104393, EPI_ISL_2104394, EPI_ISL_2104395, EPI_ISL_2104399, EPI_ISL_2104400, EPI_ISL_2104401, EPI_ISL_2104402, EPI_ISL_2104405, EPI_ISL_2104406, EPI_ISL_2104409, EPI_ISL_2104410, EPI_ISL_2193157, EPI_ISL_2193158, EPI_ISL_2193160, EPI_ISL_2193162, EPI_ISL_2193168, EPI_ISL_2193169, EPI_ISL_2193170, EPI_ISL_2193171, EPI_ISL_2193172, EPI_ISL_2193174, EPI_ISL_2193175, EPI_ISL_2193176, EPI_ISL_2193177, EPI_ISL_2193179, EPI_ISL_2193180, EPI_ISL_2193181, EPI_ISL_2193183, EPI_ISL_2193184, EPI_ISL_2193185, EPI_ISL_2193186, EPI_ISL_2193189, EPI_ISL_2193191, EPI_ISL_2193193, EPI_ISL_2193195, EPI_ISL_2193196, EPI_ISL_2193197, EPI_ISL_2193198, EPI_ISL_2193199, EPI_ISL_2193201, EPI_ISL_2193204, EPI_ISL_2193205, EPI_ISL_2193206, EPI_ISL_2193207, EPI_ISL_2193208, EPI_ISL_2193210, EPI_ISL_2193211, EPI_ISL_2193212, EPI_ISL_2193213, EPI_ISL_2193216, EPI_ISL_2193217, EPI_ISL_2312447, EPI_ISL_2312449, EPI_ISL_2312450, EPI_ISL_2312452, EPI_ISL_2312454, EPI_ISL_2312459, EPI_ISL_2312461, EPI_ISL_2312463, EPI_ISL_2312464, EPI_ISL_2312465, EPI_ISL_2312468, EPI_ISL_2312469, EPI_ISL_2312470, EPI_ISL_2312471, EPI_ISL_2312474, EPI_ISL_2312475, EPI_ISL_2312477, EPI_ISL_2312479, EPI_ISL_2312480, EPI_ISL_2312481, EPI_ISL_2312482, EPI_ISL_2312484, EPI_ISL_2312485, EPI_ISL_2312486, EPI_ISL_2312487, EPI_ISL_2312488, EPI_ISL_2312489, EPI_ISL_2312490, EPI_ISL_2312491, EPI_ISL_3007473, EPI_ISL_3007474, EPI_ISL_3007475, EPI_ISL_3007477, EPI_ISL_3007478, EPI_ISL_3007479, EPI_ISL_3007480, EPI_ISL_3007481, EPI_ISL_3007482, EPI_ISL_3007483, EPI_ISL_3007484, EPI_ISL_3007485, EPI_ISL_3085063, EPI_ISL_3085065, EPI_ISL_3085067, EPI_ISL_3085069, EPI_ISL_3085071, EPI_ISL_3085073, EPI_ISL_3085075, EPI_ISL_3085077, EPI_ISL_3085079, EPI_ISL_3085081, EPI_ISL_3085083, EPI_ISL_3085085, EPI_ISL_3085087, EPI_ISL_3085089, EPI_ISL_3085091, EPI_ISL_3085093, EPI_ISL_3085095, EPI_ISL_3085097, EPI_ISL_3085099, EPI_ISL_3085101, EPI_ISL_3085113, EPI_ISL_3085114, EPI_ISL_3085115, EPI_ISL_3085116, EPI_ISL_3085117, EPI_ISL_3085118, EPI_ISL_3085119, EPI_ISL_3085120, EPI_ISL_3085121, EPI_ISL_3127395, EPI_ISL_3127396, EPI_ISL_3127397, EPI_ISL_3127398, EPI_ISL_3127399, EPI_ISL_3127400, EPI_ISL_3127401, EPI_ISL_3134543, EPI_ISL_3134544, EPI_ISL_3134545, EPI_ISL_3134546, EPI_ISL_3134547, EPI_ISL_3134548, EPI_ISL_3134549, EPI_ISL_3153181, EPI_ISL_3153182, EPI_ISL_3153183, EPI_ISL_3153184, EPI_ISL_3153185, EPI_ISL_3153186, EPI_ISL_3153187, EPI_ISL_3153188, EPI_ISL_3153189, EPI_ISL_3153190, EPI_ISL_3153191, EPI_ISL_3153192, EPI_ISL_3153193, EPI_ISL_3153194, EPI_ISL_3153195, EPI_ISL_3153196, EPI_ISL_3153197, EPI_ISL_3153198, EPI_ISL_3153199, EPI_ISL_3153200, EPI_ISL_3153201, EPI_ISL_3156022, EPI_ISL_3156023, EPI_ISL_3156024, EPI_ISL_3156025, EPI_ISL_3156026, EPI_ISL_3156027, EPI_ISL_3156028, EPI_ISL_3156029, EPI_ISL_3156030, EPI_ISL_3156031, EPI_ISL_3156032, EPI_ISL_3156033, EPI_ISL_3156034, EPI_ISL_3156035, EPI_ISL_3156036, EPI_ISL_3156037, EPI_ISL_3156038, EPI_ISL_3156039, EPI_ISL_3156040 | | | |
| see above | Broad Institute Clinical Research Sequencing Platform | Infectious Disease Program, Broad Institute of Harvard and MIT | Adams, G.; B.L.; B.W.; Bauer, M.; Birren; Blumenstiel, B.; Brown, C.; Carter, A.; Chaluvadi, S.; D.J.; DeFelice, M.; DeRuff, K.; Dodge, S.; Gabriel, S.; Gallagher, G.; Gladden-Young, A.; Granger, B.; J.E.; K.J.; Lagerborg, K.; Larkin, K.; Lee, M.; Lemieux; Lennon, N.; Loreth, C.; Madoff, L.; McGovern, S.; Meldrim, J.; Normandin, E.; P.C.; Park; Pearlman, L.; Reilly, S.; Rudy, M.; Sabeti; Siddle; Smole, S.; Tomkins-Tinch, C.; Vicente, G.; and MacInnis |
| EPI_ISL_1018070, EPI_ISL_2106208 | DPH, Massachusetts State Public Health Lab | DPH, Massachusetts State Public Health Lab | A.S.; Fink, T.; G.R.; Gallagher; Lang; S.C.; Smole |
| EPI_ISL_1283507, EPI_ISL_1421293, EPI_ISL_1480387 | Helix/Illumina | Centers for Disease Control and Prevention Division of Viral Diseases, Pathogen Discovery | Adrian Paskey; Alexandre Bolze; Ary Ascencio; Ben L. Rambo-Martin; Benjamin Rambo-Martin; Brad Sickler; Charlotte Rivera-Garcia; Christine Tran; Christopher Gulvick; Clinton R. Paden; Dakota Howard; Darlene Wagner; David Becker; Dhwani Batra; Duncan MacCannell; Efren Sandoval; Eileen de Feo; Elizabeth Cirulli; Eric Allen; Geraint Levan; James Lu; Jan Antico; Jason Caravas; Jason Nguyen; Jimmy Ramirez; Jingtao Liu; Kara Moser; Kelly Schiabor Barrett; Kim Gietzen; Magnus Isaksson; Marc Laurent; Matthew Schmerer; Matthew Tolentino; Nicole L. Washington; Peter W. Cook; Phil Febbo; Ryan Cho; Scott Sammons; Shannon Wickline; Shatavia Morrison; Sherry Wang; Simon White; Summer Galloway; Suxiang Tong; Tyler Cassens; William Lee; Yvette Unoarumhi |
| EPI_ISL_978754 | Helix/Illumina | Respiratory Viruses Branch, Division of Viral Diseases, Centers for Disease Control and Prevention | ; Alexandre Bolze; Ary Ascencio; Ben L. Rambo-Martin; Brad Sickler; Charlotte Rivera-Garcia; Christine Tran; Clinton R. Paden; Dakota Howard; David Becker; Dhwani Batra; Duncan MacCannell; Efren Sandoval; Eileen de Feo; Elizabeth Cirulli; Eric Allen; Geraint Levan; James Lu; Jan Antico; Jason Nguyen; Jimmy Ramirez; Jingtao Liu; Kelly Schiabor Barrett; Kim Gietzen; Magnus Isaksson; Marc Laurent; Matthew Tolentino; Nicole L. Washington; Peter W. Cook; Phil Febbo; Ryan Cho; Shannon Wickline; Sherry Wang; Simon White; Summer Galloway; Suxiang Tong; Tyler Cassens; William Lee |
| EPI_ISL_907074 | Infectious Diseases, Quest Diagnostics | Infectious Diseases, Quest Diagnostics | Anderson, B.; Bernstein; D.F.; Gerasimova, A.; Hua, M.; K.E.; Kagan; L.E.; Lacbawan, F.; Liu, Y.; Livingston; Owen, R.; R.M.; Rosenthal; S.H.; Shalhout |
| EPI_ISL_1692211 | Infinity Biologix | Centers for Disease Control and Prevention Division of Viral Diseases, Pathogen Discovery | Adrian Paskey; Benjamin Rambo-Martin; Chirayu Goswami; Christian Bixby; Christopher Gulvick; Clinton R. Paden; Dakota Howard; Darlene Wagner; Dhwani Batra; Duncan MacCannell; Jason Caravas; Jonathan Schultz; Kara Moser; Matthew Schmerer; Peter W. Cook; Robin Grimwood; Russ Hager; Scott Sammons; Shatavia Morrison; Yihe Wang; Yvette Unoarumhi |
| EPI_ISL_1193357, EPI_ISL_1220967, EPI_ISL_1221043, EPI_ISL_1221044, EPI_ISL_1222074, EPI_ISL_1222082, EPI_ISL_1338601, EPI_ISL_1463228, EPI_ISL_1549047, EPI_ISL_1549095, EPI_ISL_1549133, EPI_ISL_1549374, EPI_ISL_1610307, EPI_ISL_1610308, EPI_ISL_1610309, EPI_ISL_1610614, EPI_ISL_1610977, EPI_ISL_1610996, EPI_ISL_1611030, EPI_ISL_1680495, EPI_ISL_1680637, EPI_ISL_1681364, EPI_ISL_1681490, EPI_ISL_1800665, EPI_ISL_1801391, EPI_ISL_1801602, EPI_ISL_1926597, EPI_ISL_2045773, EPI_ISL_2181698, EPI_ISL_2184384, EPI_ISL_2305757, EPI_ISL_2306886, EPI_ISL_2397934, EPI_ISL_2399018, EPI_ISL_2399263, EPI_ISL_3330566 | | | |
| see above | Laboratory Corporation of America | Centers for Disease Control and Prevention Division of Viral Diseases, Pathogen Discovery | Adrian Paskey; Amanda Douglas; Amanda Suchanek; Andrea Throop; Ayla Burns; Ben L. Rambo-Martin; Benjamin Rambo-Martin; Bobbi Croy; Brian Krueger; Brian Norvell; Christopher Gulvick; Christos Petropoulos; Clinton R. Paden; Craig Lukasik; Dakota Howard; Darlene Wagner; Debbie Boles; Dhwani Batra; Duncan MacCannell; Eyad Almasri; Eyad Almasri Debbie Boles; Goran Stevovic; Howard Engler; Hrushikesh Deshmukh; Jake Humphrey; Jana Schroth; Jason Caravas; Joe Voshell; John Pruitt; Jonathan Meltzer; Jonathan Williams; Kara Moser; Kimberly Wagner; Lax Iyer; Lisa Pfefferle; Lyndon Tilson; Manoj Jain; Marcia Eisenberg; Mary Ann Cristobal; Mary Williamson; Matthew Robinson; Matthew Schmerer; Michael Levandoski; Mike Sapeta; Mindy Nye; Minoo Agarwal; Mohan Kolli; Nuthawin Charoensri; Oren Cohen; Peter W. Cook; Prashant Gupta; Qian Zeng; Rama Ghatti; Scott Parker; Scott Ryan; Scott Sammons; Shatavia Morrison; Stanley Letovsky; Steven Ragan; Suresh Babu Selvaraju; Susan Countryman; Susan Hicks; Suxiang Tong; Suzanne Dale; Thomas Urban; Tim Kuphal; Tricia Zwiefelhofer; Vincent Drouillon; Yvette Unoarumhi |
| EPI_ISL_1026609, EPI_ISL_1030586, EPI_ISL_1037673, EPI_ISL_1161149, EPI_ISL_1161887, EPI_ISL_1163047, EPI_ISL_1163171 | | | |
| see above | Laboratory Corporation of America | Respiratory Viruses Branch, Division of Viral Diseases, Centers for Disease Control and Prevention | Amanda Douglas; Amanda Suchanek; Andrea Throop; Ayla Burns; Ben L. Rambo-Martin; Bobbi Croy; Brian Krueger; Brian Norvell; Christos Petropoulos; Clinton R. Paden; Craig Lukasik; Dakota Howard; Dhwani Batra; Duncan MacCannell; Eyad Almasri Debbie Boles; Goran Stevovic; Howard Engler; Hrushikesh Deshmukh; Jake Humphrey; Jana Schroth; Joe Voshell; John Pruitt; Jonathan Meltzer; Jonathan Williams; Kimberly Wagner; Lax Iyer; Lyndon Tilson; Manoj Jain; Marcia Eisenberg; Mary Ann Cristobal; Mary Williamson; Michael Levandoski; Mike Sapeta; Mindy Nye; Minoo Agarwal; Mohan Kolli; Nuthawin Charoensri; Oren Cohen; Peter W. Cook; Prashant Gupta; Qian Zeng; Rama Ghatti; Scott Parker; Scott Ryan; Stanley Letovsky; Steven Ragan; Suresh Babu Selvaraju; Susan Countryman; Susan Hicks; Suxiang Tong; Suzanne Dale; Thomas Urban; Tim Kuphal; Tricia Zwiefelhofer; Vincent Drouillon |
| EPI_ISL_965855, EPI_ISL_965856, EPI_ISL_965857, EPI_ISL_965858, EPI_ISL_965859, EPI_ISL_965860, EPI_ISL_965861, EPI_ISL_965862, EPI_ISL_965863, EPI_ISL_1009793, EPI_ISL_1009794, EPI_ISL_1009795, EPI_ISL_1233294, EPI_ISL_1233295, EPI_ISL_1233296, EPI_ISL_1233297, EPI_ISL_1233298, EPI_ISL_1233299, EPI_ISL_1233300, EPI_ISL_1233301, EPI_ISL_1233302, EPI_ISL_1233348, EPI_ISL_1233349, EPI_ISL_1233350, EPI_ISL_1233351, EPI_ISL_1233352, EPI_ISL_1233353, EPI_ISL_1233354, EPI_ISL_1233355, EPI_ISL_1503481, EPI_ISL_1503482, EPI_ISL_1503483, EPI_ISL_1503484, EPI_ISL_1503485, EPI_ISL_1503486, EPI_ISL_1503487, EPI_ISL_1503488, EPI_ISL_1503489, EPI_ISL_1503490, EPI_ISL_1503491, EPI_ISL_1503492, EPI_ISL_1503493, EPI_ISL_1503494, EPI_ISL_1503495, EPI_ISL_1503496, EPI_ISL_1503550, EPI_ISL_1503551, EPI_ISL_1503552, EPI_ISL_1503553, EPI_ISL_1503554, EPI_ISL_1503555, EPI_ISL_1503556, EPI_ISL_1503557, EPI_ISL_1503558, EPI_ISL_1503559, EPI_ISL_1503560, EPI_ISL_1503652, EPI_ISL_1503653, EPI_ISL_1503654, EPI_ISL_1503655, EPI_ISL_1503750, EPI_ISL_1503751, EPI_ISL_1503752, EPI_ISL_1503753, EPI_ISL_1503754, EPI_ISL_1503755, EPI_ISL_1503772, EPI_ISL_1503773, EPI_ISL_1503774, EPI_ISL_1503775, EPI_ISL_1503776, EPI_ISL_1503777, EPI_ISL_1503778, EPI_ISL_1503779, EPI_ISL_1503780, EPI_ISL_1503781, EPI_ISL_1503782, EPI_ISL_1503783, EPI_ISL_1503784, EPI_ISL_1503785, EPI_ISL_1503786, EPI_ISL_1503787, EPI_ISL_1503788, EPI_ISL_1503789, EPI_ISL_1503790, EPI_ISL_1503791, EPI_ISL_1527092, EPI_ISL_1527095, EPI_ISL_1527096, EPI_ISL_1527102, EPI_ISL_1527103, EPI_ISL_1527104, EPI_ISL_1527105, EPI_ISL_1527111, EPI_ISL_1527117, EPI_ISL_1527151, EPI_ISL_1527156, EPI_ISL_1527157, EPI_ISL_1527161, EPI_ISL_1527193, EPI_ISL_1527194, EPI_ISL_1527195, EPI_ISL_1527196, EPI_ISL_1527197, EPI_ISL_1527198, EPI_ISL_1527199, EPI_ISL_1527200, EPI_ISL_1527202, EPI_ISL_1527206, EPI_ISL_1527207, EPI_ISL_1527208, EPI_ISL_1527212, EPI_ISL_1527215, EPI_ISL_1527229, EPI_ISL_1527230, EPI_ISL_1527231, EPI_ISL_1527232, EPI_ISL_1527233, EPI_ISL_1527234, EPI_ISL_1527235, EPI_ISL_1527236, EPI_ISL_1624516, EPI_ISL_1624536, EPI_ISL_1624544, EPI_ISL_1624635, EPI_ISL_1624636, EPI_ISL_1624658, EPI_ISL_1624659, EPI_ISL_1624660, EPI_ISL_1624675, EPI_ISL_1624678, EPI_ISL_1624682, EPI_ISL_1624693, EPI_ISL_1624695, EPI_ISL_1624696, EPI_ISL_1710604, EPI_ISL_1710613, EPI_ISL_1710615, EPI_ISL_1710617, EPI_ISL_1710619, EPI_ISL_1710620, EPI_ISL_1710623, EPI_ISL_1710625, EPI_ISL_1710633, EPI_ISL_1710634, EPI_ISL_1710635, EPI_ISL_1710636, EPI_ISL_1710647, EPI_ISL_1710651, EPI_ISL_1710661, EPI_ISL_2008120, EPI_ISL_2008123, EPI_ISL_2008126, EPI_ISL_2008145, EPI_ISL_2008156, EPI_ISL_2008165, EPI_ISL_2008171, EPI_ISL_2008200, EPI_ISL_2008255, EPI_ISL_2008288, EPI_ISL_2008289, EPI_ISL_2008290, EPI_ISL_2008291, EPI_ISL_2008292, EPI_ISL_2008311, EPI_ISL_2008320, EPI_ISL_2008324, EPI_ISL_2008328, EPI_ISL_2008334, EPI_ISL_2008387, EPI_ISL_2008397, EPI_ISL_2008398, EPI_ISL_2008400, EPI_ISL_2229787, EPI_ISL_2229788, EPI_ISL_2229790, EPI_ISL_2229791, EPI_ISL_2229795, EPI_ISL_2400253, EPI_ISL_2400254, EPI_ISL_2400255, EPI_ISL_2400256, EPI_ISL_2400257, EPI_ISL_2400258, EPI_ISL_2400259, EPI_ISL_2400260, EPI_ISL_2400261, EPI_ISL_2400262, EPI_ISL_2400263, EPI_ISL_2483774, EPI_ISL_2483775, EPI_ISL_2483776 | | | |
| see above | Massachusetts State Public Health Laboratory | Massachusetts State Public Health Lab | Andrew Lang; Glen Gallagher; Sandra Smole; Timelia Fink |
| EPI_ISL_854976, EPI_ISL_855185 | Quest Diagnostics | Quest Diagnostics | Anderson, B.; Bernstein; D.F.; Gerasimova, A.; Hua, M.; I.A.; K.E.; Kagan; L.E.; Lacbawan, F.; Liu Y.; Livingston; Owen, R.; Perez, A.; R.M.; Rosenthal; S.H.; Shalhout; Shlyakhter; Tanpaiboon, P. |
| EPI_ISL_1194110, EPI_ISL_1266137, EPI_ISL_1267090, EPI_ISL_1267316, EPI_ISL_1291104, EPI_ISL_1297621, EPI_ISL_1297649, EPI_ISL_1314758, EPI_ISL_1314893, EPI_ISL_1340666, EPI_ISL_1340696, EPI_ISL_1367201, EPI_ISL_1367222, EPI_ISL_1392255, EPI_ISL_1392296, EPI_ISL_1392321, EPI_ISL_1444517, EPI_ISL_1444539, EPI_ISL_1444564, EPI_ISL_1479920, EPI_ISL_1480793, EPI_ISL_1491974, EPI_ISL_1551830, EPI_ISL_1552168, EPI_ISL_1818245, EPI_ISL_1834245, EPI_ISL_1834331, EPI_ISL_1834382, EPI_ISL_1840469, EPI_ISL_1906225, EPI_ISL_1906424, EPI_ISL_1924304, EPI_ISL_1924331, EPI_ISL_1924373, EPI_ISL_1924392, EPI_ISL_1924411, EPI_ISL_1924560, EPI_ISL_2011053, EPI_ISL_2090512, EPI_ISL_2090538, EPI_ISL_2133099, EPI_ISL_2133234, EPI_ISL_2143087, EPI_ISL_2143152, EPI_ISL_2143163, EPI_ISL_2143780, EPI_ISL_2143921, EPI_ISL_2187248, EPI_ISL_2268480, EPI_ISL_2268564, EPI_ISL_2268800, EPI_ISL_2268815, EPI_ISL_2268934, EPI_ISL_2269173, EPI_ISL_2269205, EPI_ISL_2280170, EPI_ISL_2304643, EPI_ISL_2369197, EPI_ISL_2371747, EPI_ISL_2372143, EPI_ISL_2440543, EPI_ISL_2440787, EPI_ISL_2529265, EPI_ISL_2529320, EPI_ISL_2652330, EPI_ISL_2930061, EPI_ISL_2930271, EPI_ISL_3018175, EPI_ISL_3018374 | | | |
| see above | Quest Diagnostics Incorporated | Centers for Disease Control and Prevention Division of Viral Diseases, Pathogen Discovery | A. Gerasimova; A. Perez; Adrian Paskey; B. Anderson; Ben L. Rambo-Martin; Benjamin Rambo-Martin; Christopher Gulvick; Clinton R. Paden; Dakota Howard; Darlene Wagner; Dhwani Batra; Duncan MacCannell; F. Lacbawan; I. A. Shlyakhter; Jason Caravas; K.E. Livingston; Kara Moser; L.E. Bernstein; M. Hua; Matthew Schmerer; P. Tanpaiboon; Peter W. Cook; R. M. Kagan; R. Owen; R. V. Rolando; S. H. Rosenthal; Scott Sammons; Shatavia Morrison; Suxiang Tong; Y. Liu; Yvette Unoarumhi |
| EPI_ISL_1086486, EPI_ISL_1086489, EPI_ISL_1086496, EPI_ISL_1086509, EPI_ISL_1086515, EPI_ISL_1086520, EPI_ISL_1086536, EPI_ISL_1086691, EPI_ISL_1086818, EPI_ISL_1086849, EPI_ISL_1087035, EPI_ISL_1087099, EPI_ISL_1087464, EPI_ISL_1087489, EPI_ISL_1087580, EPI_ISL_1087822, EPI_ISL_1088128, EPI_ISL_1088144, EPI_ISL_1088150, EPI_ISL_1088168, EPI_ISL_1088181, EPI_ISL_1088218, EPI_ISL_1088221, EPI_ISL_1088227, EPI_ISL_1088234, EPI_ISL_1088237, EPI_ISL_1088326, EPI_ISL_1090572, EPI_ISL_1090892, EPI_ISL_1090921, EPI_ISL_1114129, EPI_ISL_1139696 | | | |
| see above | Quest Diagnostics Incorporated | Respiratory Viruses Branch, Division of Viral Diseases, | A. Gerasimova; A. Perez; B. Anderson; Ben L. Rambo-Martin; Clinton R. Paden; Dakota Howard; Dhwani Batra; Duncan MacCannell; F. Lacbawan; I. A. Shlyakhter; K.E. Livingston; L.E. Bernstein; M. Hua; P. Tanpaiboon; Peter W. Cook; R. M. Kagan; R. Owen; R. V. Rolando; S. H. Rosenthal; Suxiang Tong; Y. Liu |

| | | | |
|---|---|---|---|
| | | Centers for Disease Control and Prevention | |
| EPI_ISL_1379956, EPI_ISL_1379957, EPI_ISL_1380499 | University of Vermont Medical Center | UW Virology Lab | Alexander Greninger; Allison L. Unger; Beth D. Kirkpatrick; Emily A. Bruce; Hong Xie; Jessica W. Crothers; Keith R. Jerome; Lasata Shrestha; Margaret Mills; Marya P. Carmolli; Meei-Li Huang; Michelle Lin; Noah R. Baker; Pavitra Roychoudhury; Saraswathi Sathees; Sean Ellis; Shah Mohamed Bakhash |
| EPI_ISL_1272869, EPI_ISL_1272870, EPI_ISL_1272871, EPI_ISL_1272872, EPI_ISL_1446667, EPI_ISL_1446668, EPI_ISL_1446669, EPI_ISL_1446670, EPI_ISL_1446672, EPI_ISL_1446673, EPI_ISL_1446674, EPI_ISL_1446675, EPI_ISL_1528386, EPI_ISL_1711919, EPI_ISL_1711920, EPI_ISL_1711922, EPI_ISL_1937809, EPI_ISL_1937810, EPI_ISL_1937811, EPI_ISL_3354001, EPI_ISL_3354002, EPI_ISL_3354003 | | | |
| see above | VT Dept. of Health Laboratory | Centers for Disease Control and Prevention Division of Viral Diseases, Pathogen Discovery | Alex Burgin; Alison Laufer Halpin; Anna Montmayeur; Anna Uehara; Ben L. Rambo-Martin; Ben Rambo-Martin; Clinton Paden; Clinton R. Paden; Dakota Howard; Darlene Wagner; Dave Wentworth; Dhwani Batra; Haibin Wang; Jasmine Padilla; Jing Zhang; Justin Lee; Katie Dillon; Krista Queen; Kristen Knipe; Kristine Lacek; Lori Rowe; Mark Burroughs; Matthew Schmerer; Meghan Bentz; Mili Sheth; Peter Cook; Peter W. Cook; Rachel Marine; Sam Shepard; Sarah Nobles; Shoshona Le; Suxiang Tong; Vivien Dugan; Yan Li; Ying Tao; Yvette Unoarumhi |
| EPI_ISL_1225941, EPI_ISL_1225942, EPI_ISL_1225943, EPI_ISL_1225944, EPI_ISL_1225945, EPI_ISL_1226231, EPI_ISL_1226232, EPI_ISL_1226233, EPI_ISL_1226234, EPI_ISL_1226235, EPI_ISL_1226236 | | | |
| see above | VT Dept. of Health Laboratory | Genomics and Discovery, Respiratory Viruses Branch, Division of Viral Diseases, Centers for Disease Control and Prevention | Anna Montmayeur; Anna Uehara; Ben L. Rambo-Martin; Clinton R. Paden; Dhwani Batra; Haibin Wang; Jasmine Padilla; Jing Zhang; Justin Lee; Katie Dillon; Krista Queen; Kristen Knipe; Kristine Lacek; Lori Rowe; Mark Burroughs; Matthew Schmerer; Mili Sheth; Peter W. Cook; Rachel Marine; Sam Shepard; Sarah Nobles; Shoshona Le; Suxiang Tong; Yan Li; Ying Tao |
| EPI_ISL_1094200, EPI_ISL_1094245, EPI_ISL_1094266, EPI_ISL_1094267, EPI_ISL_1094556, EPI_ISL_1094557, EPI_ISL_1094558, EPI_ISL_1095187, EPI_ISL_1095188, EPI_ISL_1095189 | | | |
| see above | VT Dept. of Health Laboratory | Respiratory Viruses Branch, Division of Viral Diseases, Centers for Disease Control and Prevention | Anna Montmayeur; Anna Uehara; Ben L. Rambo-Martin; Clinton R. Paden; Dhwani Batra; Haibin Wang; Jasmine Padilla; Jing Zhang; Justin Lee; Krista Queen; Lori Rowe; Mark Burroughs; Mili Sheth; Peter W. Cook; Rachel Marine; Sarah Nobles; Suxiang Tong; Yan Li; Ying Tao |
